## Supplementary Information for "The unintended consequences of inconsistent pandemic control policies"

### Supplementary Information: The unintended consequences of inconsistent pandemic control policies

**This supplementary material presents:** (1) additional analysis of the mobility data on a state-by-state basis, and (2) additional mathematical analysis of the model introduced in the main text.

#### Data

##### SafeGraph movement data

SafeGraph is a data company that aggregates anonymized location data from numerous applications in order to provide insights about physical places. To enhance privacy, SafeGraph excludes census block group information if fewer than five devices visited an establishment in a month (two devices in a week) from a given census block group. SafeGraph has made public data on foot traffic and visits to places free for COVID-19 researchers. This data is built on a panel of 50 million devices that collect anonymous location data across the US and includes movements by census block group and weekly numbers of visitors for over 6 million places. Using these data we use counts of visits and unique visitors to these places as well as the distance traveled from 'home' (defined as the common nighttime location for the device over a 6 week period where nighttime is 6 pm to 7 am). We calculate the changes in visits over time by location and the distance traveled from each unique county. To get movement patterns for specific venues, we search the business name for 'church' not 'chicken' (to remove visits to the fast-food chain Church's Chicken), 'bar' or 'tavern', 'park', 'grocery', and 'gym'.

##### Google search data

Google searches for 'church + churches', 'bar + bars', 'park + parks', 'grocery', and 'gym + gyms' were monitored using Google Trends for all 50 US states, D.C., and nationally from January 1, 2010 to October 1, 2020 and were normalized by the number of searches per 10,000,000 searches over the time period. Data were downloaded using the Trends Application Programming Interface for health. To examine increases in searches for churches in the month following the declaration of emergency on March 13, 2020 we compare search volumes for Sundays from March 13 to April 13 to Sundays in March 13 to April 13 from 2010–2019. This approximates a counterfactual scenario to account for the Catholic observance of Lent that occurs around this period. To assess potential differences in search behaviors for the first six months of the epidemic, we calculate the coefficient of variation (search volume standard deviation divided by mean search volume) for January to September for all years.

#### SARS-CoV-2 case data

SARS-CoV-2 incidence data at the county level were downloaded from the COVID-19 Data Repository by the Center for Systems Science and Engineering at Johns Hopkins University at the county level beginning in February, 2020. We join SafeGraph movement data with case counts at the county level and to assess movement in response to cases, we distinguish a *focal county* to *visiting counties*. That is, for each venue type (church, bar, park, grocery, gym), and each unique county represented by that venue type (this is the focal county) we calculate the total number of cases in that county for that week. We keep this as well as total cases divided by the population of that county. Next we calculate the total cases for each represented visiting counties to venues represented. Finally, we calculate the mean number of unique visiting counties by week and county.

With these data we calculate two metrics, one is the proportion of visiting counties which have more cases than the focal county being visited, and the other is the raw difference in cases between the focal and visiting counties. For the latter we calculate the 5th, 25th, 50th, 75th, and 95th quantiles of numbers of cases. We can then compare these two metrics by state with observed incidence in that state. To quantify any associations between the proportion of visiting counties larger than focal counties, we take the cross-wavelet of the two time series and see where the power is significant and the relative phase angle between the two.

Included as supplementary materials are plots of proportions of visiting counties with more cases than the focal county and differences in cases between visiting and focal counties for all 50 US states and D.C., and cross-wavelet plots for churches, parks, gyms, groceries, and bars and cases for all 50 US states and D.C..

#### Mobility data correlates

We used the social, demographic, and economical variables compiled by White & Laurent Hébert-Dufresne[1] to examine potential correlates for the mobility data. We found that none of these variables strongly correlated with either percentage decreases in visits or percent change in distance traveled (results not shown). However, increases in state-level “tightness" was correlated with larger decreases in church visits and farther distanced traveled (Fig. S1). Tight cultures are typically defined as those with strong social norms and little tolerance for deviance[2].

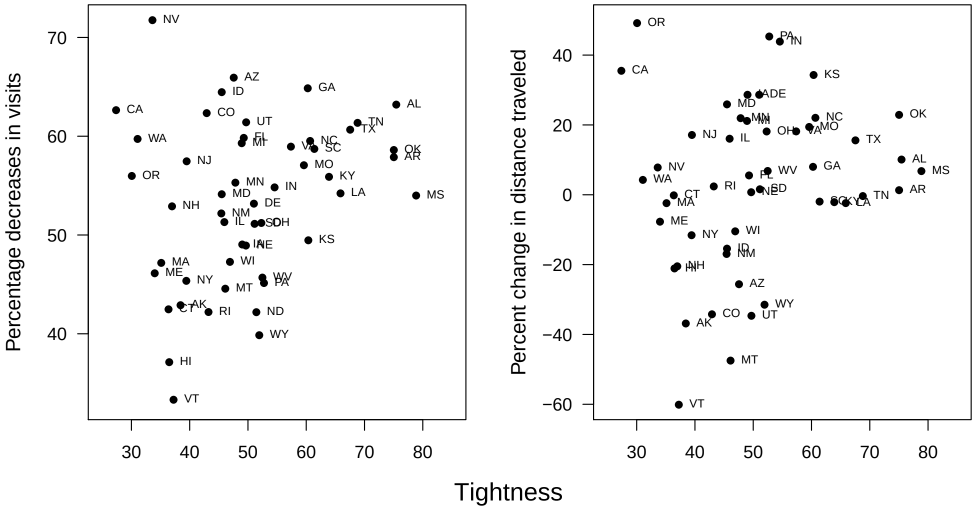


**Figure S1.** (a) Percent decreases in numbers of unique visitors to churches versus tightness by state. (b) Percent change in distances traveled to churches versus tightness by state.

#### Mathematical analysis of final outbreak size

The mathematical simplicity of the classic SIR model, on which our model is based, allows for a number of more detailed analyses of the role of $X$ and $Y$ on the final outbreak size. In general, the final outbreak size for a given $\lambda$, $X$ and $Y$ in our model is given by

$$R(\infty) = (1-X)R_o(\infty) + XR_c(\infty) \tag{S1}$$

We assume here that $t_c = 0$, and that $S_o(0) \approx 1$, $I_o(0) \ll 1$, and $R_o(0) = 0$. These assumptions serve as a natural motivating example while allowing for a less cumbersome mathematical analysis. In this case, $R_o(t_c)$ becomes 0 and so Eq. S1 simplifies to $(1-X)R_o(\infty)$. therefore for notational convenience we simply write $R$ and $S$ to denote the open compartments, since closed compartments will always be empty.

Note that after redistribution, the population sizes for open compartments are no longer normalized to 1. Therefore to help prevent confusion we let $r/s(t)$ be the proportion of recovered/susceptible individuals. After redistribution the population size in open compartments is $1+\frac{XY}{1-X} =: P$, so $s(t) = S(t)/P$ and $r(t) = R(t)/P$.

By Eq. (4) in Ma & Earn (2006)[3], for open compartments we then have

$$\begin{aligned} r(\infty) &= 1 - s(\infty) \\ &= 1 - s(0)\exp(-R_0(r(\infty) - r(0)) \\ &= 1 - \exp(-R_0 r(\infty))), \end{aligned} \tag{S2}$$

where we have used that $s(0) = 1-\varepsilon$ and $r(0) = 0$. Note the reproductive number $R_0$ here is defined $\lambda P$. This transcendental equation can then be solved for $r(\infty)$ with respect to a particular set of parameters though numerical means or using the Lambert W function. Following Appendix A of Ma & Earn (2006)[3] and elsewhere, $s(\infty) = -\frac{1}{R_0}W(-R_0 e^{-R_0})$, where $W$ is the principal branch of the Lambert W function. Therefore we may write (S2) in closed-form, which in turn gives

$$\begin{aligned} R(\infty) &= (1-X)P\left(1+\frac{W(-R_0e^{-R_0})}{R_0}\right) \\ &= (1-X+XY)\left(1+\frac{W(-R_0e^{-R_0})}{R_0}\right) \end{aligned} \tag{S3}$$

##### Finding critical $Y$ for a given $\lambda$

We first show how Eq. S2 can be used to find the value of $Y$ past which, for any $\lambda$, any choice of $X > 0$ will cause a worse final outbreak than compared to $X = 0$; i.e. critical Y.

Let $\beta$ be the initial infectiousness $\lambda(1+\frac{XY}{1-X})$, and as above we use $R$ to denote the open compartment. The total outbreak size in open churches is:

$$R(\infty) = 1 - e^{-\beta R(\infty)} \tag{S4}$$

To find critical $Y$ as defined, we want to solve for the following stationary point:

$$\frac{\partial}{\partial X}\left(R(\infty)(1-X)(1+\frac{XY}{1-X})\right)|_{X=0} = 0 \tag{S5}$$

Taking the partial derivative of S4 implicitly with respect to $X$ yields:

$$\partial_X(R(\infty)) = (\partial_X(\beta)R(\infty) + \beta\partial_X(R(\infty)))e^{-\beta R(\infty)}, \tag{S6}$$

which leads to the following system, which is solvable for $Y$ given any $\lambda$.

$$\begin{aligned} R(\infty) - 1 + e^{-\lambda R(\infty)} &= 0 \\ \partial_X R(\infty) - R(\infty) + R(\infty)Y &= 0 \\ \partial_X R(\infty) - e^{-\lambda R(\infty)}(\lambda Y R(\infty) + \lambda\partial_X R(\infty)) &= 0 \end{aligned}$$

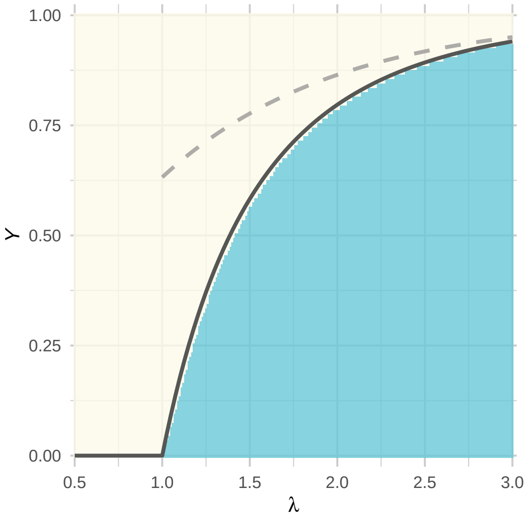


**Figure S2.** Value of $X$ giving the minimum value of Eq. (S3), as a function of $Y$ and $\lambda$, based on numerical simulation of the clorSIR model. A clear transition from $X = 0$ (yellow) to $X = 1$ (blue) is seen, with no intermediary values. The solid black line corresponds to the theoretical closed-form solution from Eq. (S7), while the dashed grey line corresponds to the rough approximation $Y > 1 - e^{-\lambda}$ past which it is best to not have any closures.

#### Finding Optimal Closure Percentage

We now turn to finding the value of $X$ which minimizes (S1) for a given $Y$ and $\lambda \geq 1$ (when $\lambda < 1$, $X = 0$ is clearly as optimal as anything else). When $\lambda \geq 1$, one can see from Figure 4 in the main text that $R(\infty)$ as a function of $X$ has either a single intermediate peak higher values of $Y$, or is monotone decreasing for lower values of $Y$. This pervasive downward parabolic shape arises from the fact that $R(\infty)$ is the product of the linearly decreasing, positive function $f(X) = 1 - X + XY$, and the sigmoidal, positive function $g(X) = 1 + (W(-R_0 e^{-R_0}))/R_0$, where $\frac{dg}{dX}$ approaches 0 as $X$ approaches 1. This guarantees that $R(\infty)$ is maximized at one of the extreme values $X = 0$ or $X = 1$.

While Eq. (S3) is not defined at $X = 1$, we can obtain a right-hand limit. Using that $\lim_{X \to 1+} -R_0 e^{-R_0} = 0$ and $W(x) \approx x$ for $x$ small, we have as $X \to 1$ that

$$\begin{aligned} R(\infty) &= (1 + e^{-R_0}) P(1 - X) \\ &= 1 - X + XY + e^{-R_0}(1 - X + XY) \\ &\to Y. \end{aligned}$$

This result makes sense, since we would expect that $r(\infty)$ be equal to 1 when $R_0 \to \infty$, so plugging this into Eq. (S2) and simplifying gives $R(\infty) = Y$ for $X = 1$.

This leads to the section's main result, which is summarized in Figure S2.

$$\arg\min_X R(\infty) = \begin{cases} 0 & \text{if } Y > \left(1 + \frac{W(-\lambda e^{-\lambda})}{\lambda}\right) \\ 1 & \text{otherwise.} \end{cases} \tag{S7}$$

While this is in closed form, $W$ cannot be expressed with elementary functions and hence poses similar interpretability issues to practitioners as implicit solutions or numerical approximations. Thankfully, a number of useful approximations for $W$ exist. For example, here we can use the crude estimate $W(x) < x$ for $-1/e \leq x < 0$ to obtain the bound $Y > 1 - e^{-\lambda}$, which serves as sufficient criteria to be certain that no closure is the best option.
