## Supplementary material for "The unintended consequences of inconsistent pandemic control policies": Case difference by state with incidence

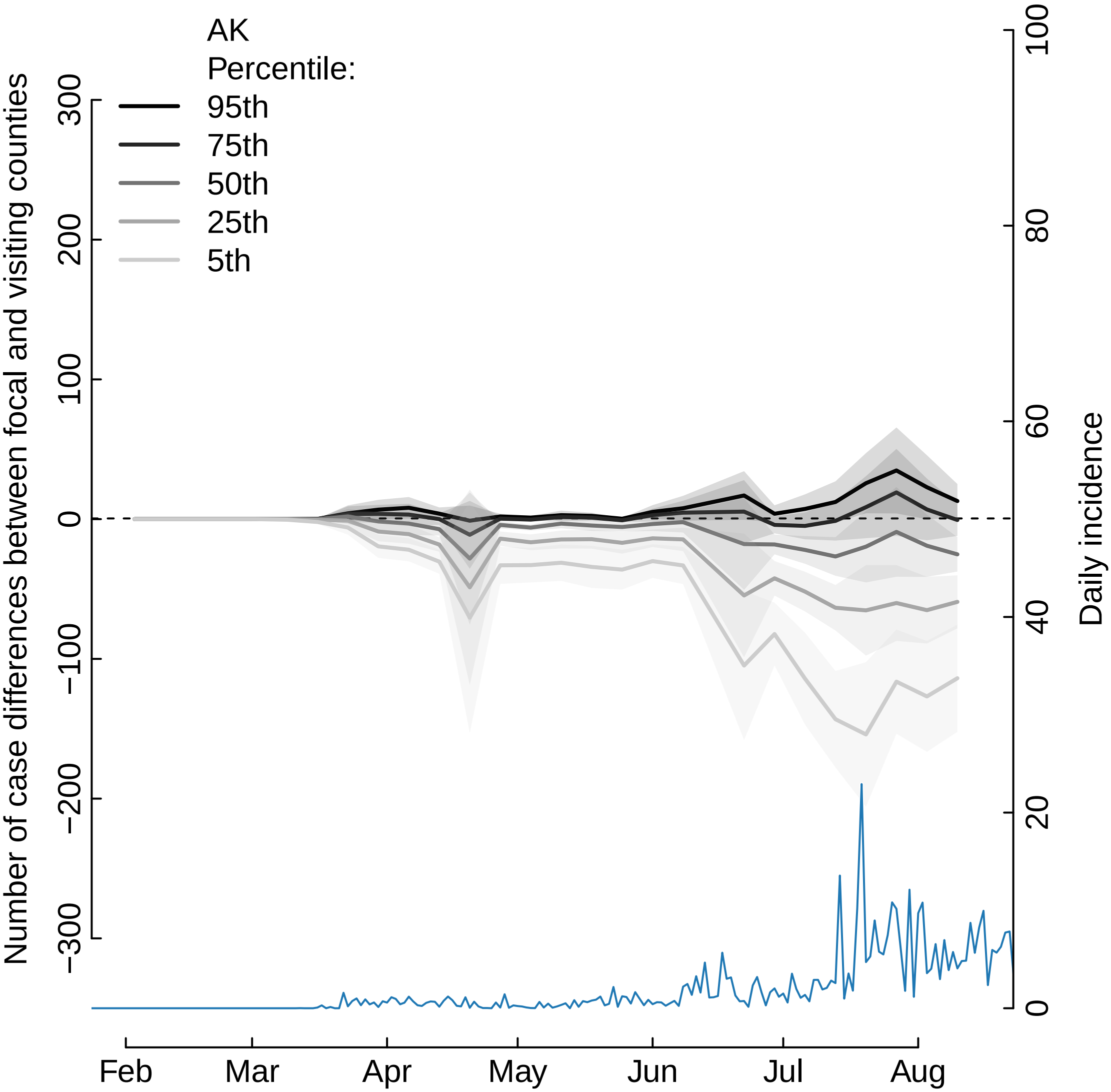
AK
Percentile:
95th
75th
50th
25th
5th
Number of case differences between focal and visiting counties
300
200
100
0
−100
−200
−300
Daily incidence
100
80
60
40
20
0
Feb
Mar
Apr
May
Jun
Jul
Aug

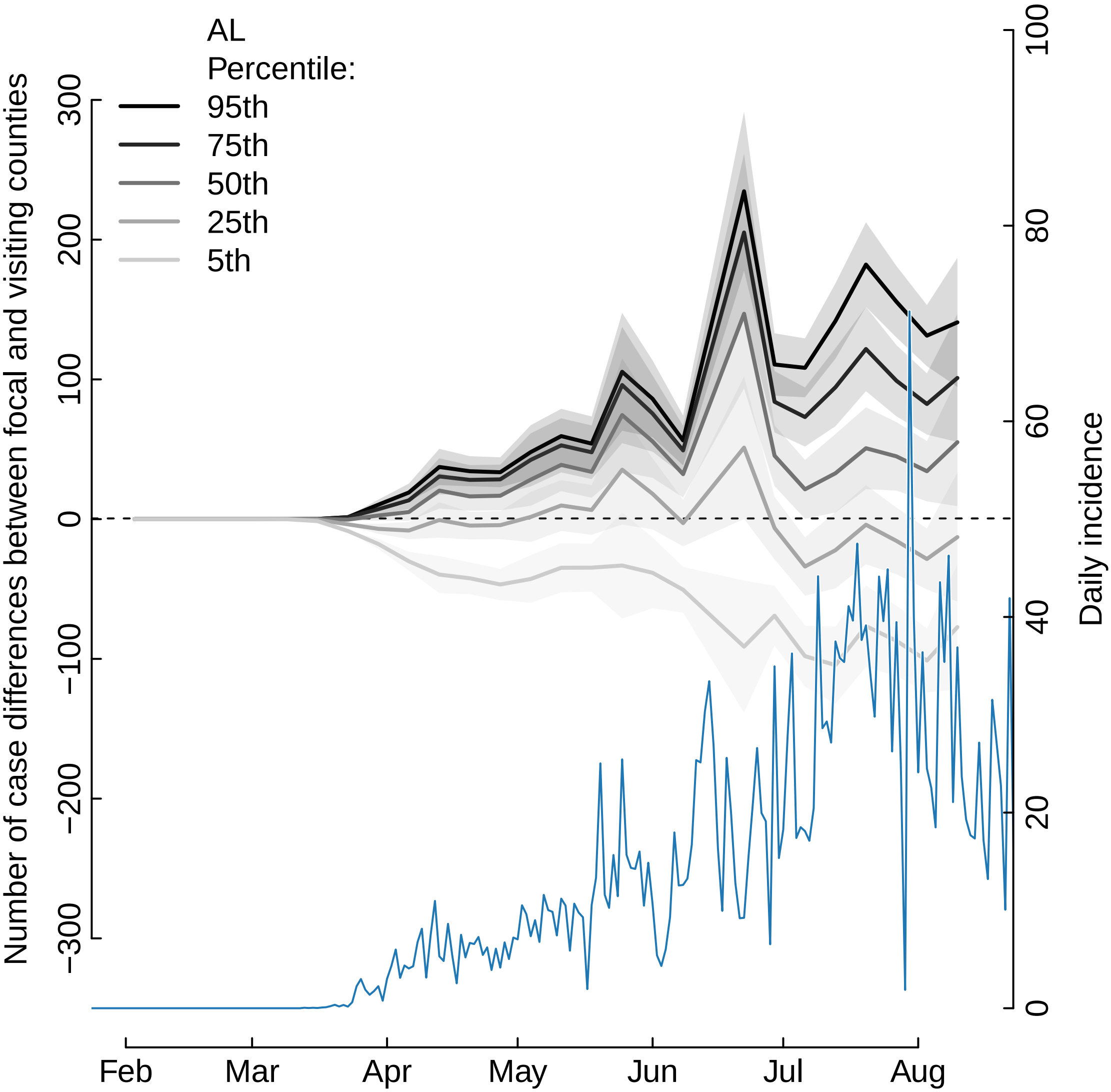
AL
Percentile:
95th
75th
50th
25th
5th
Number of case differences between focal and visiting counties
300
200
100
0
−100
−200
−300
Daily incidence
100
80
60
40
20
0
Feb
Mar
Apr
May
Jun
Jul
Aug

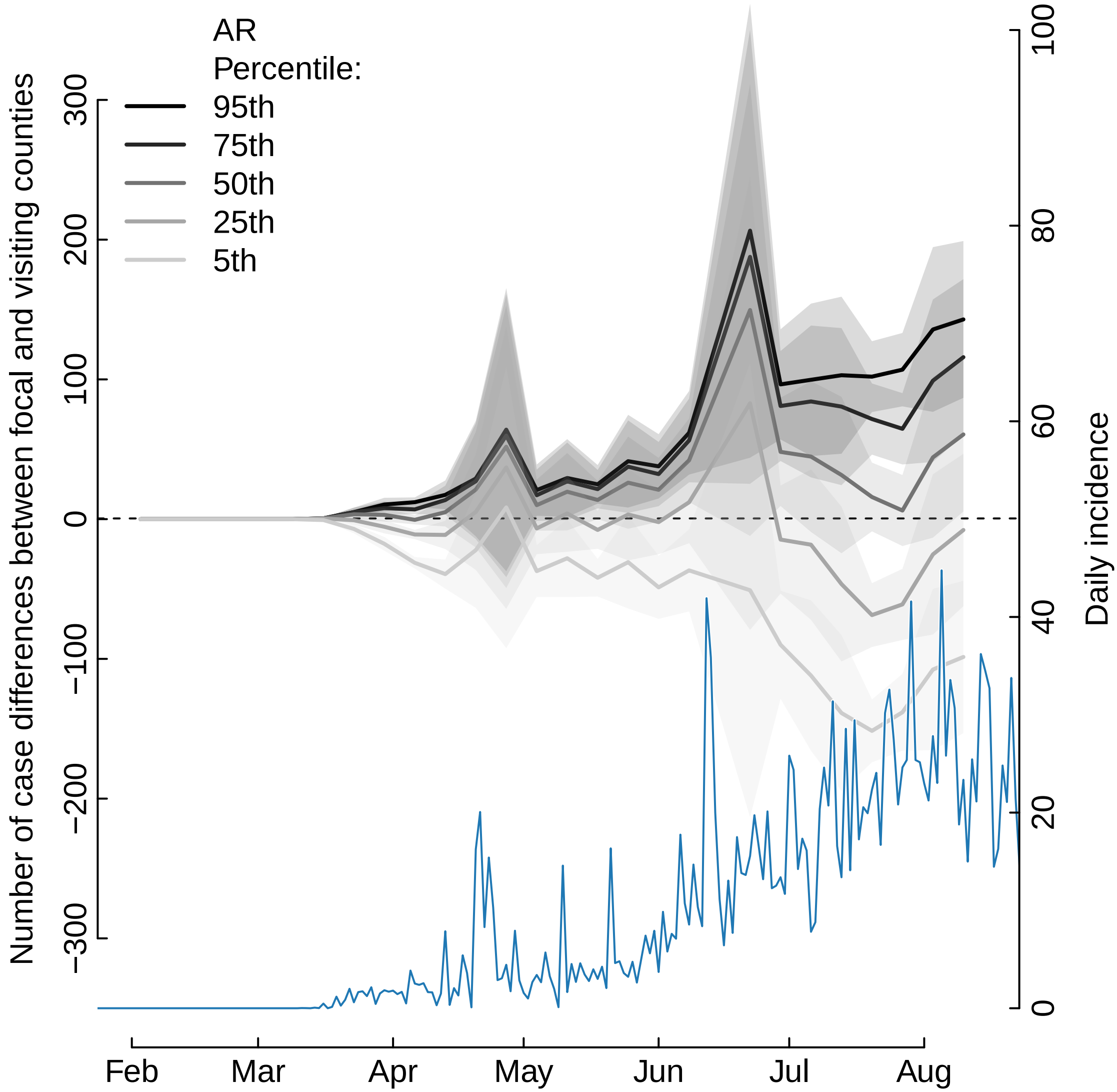

AR
Percentile:
95th
75th
50th
25th
5th
Number of case differences between focal and visiting counties
300
200
100
0
−100
−200
−300
Daily incidence
100
80
60
40
20
0
Feb
Mar
Apr
May
Jun
Jul
Aug

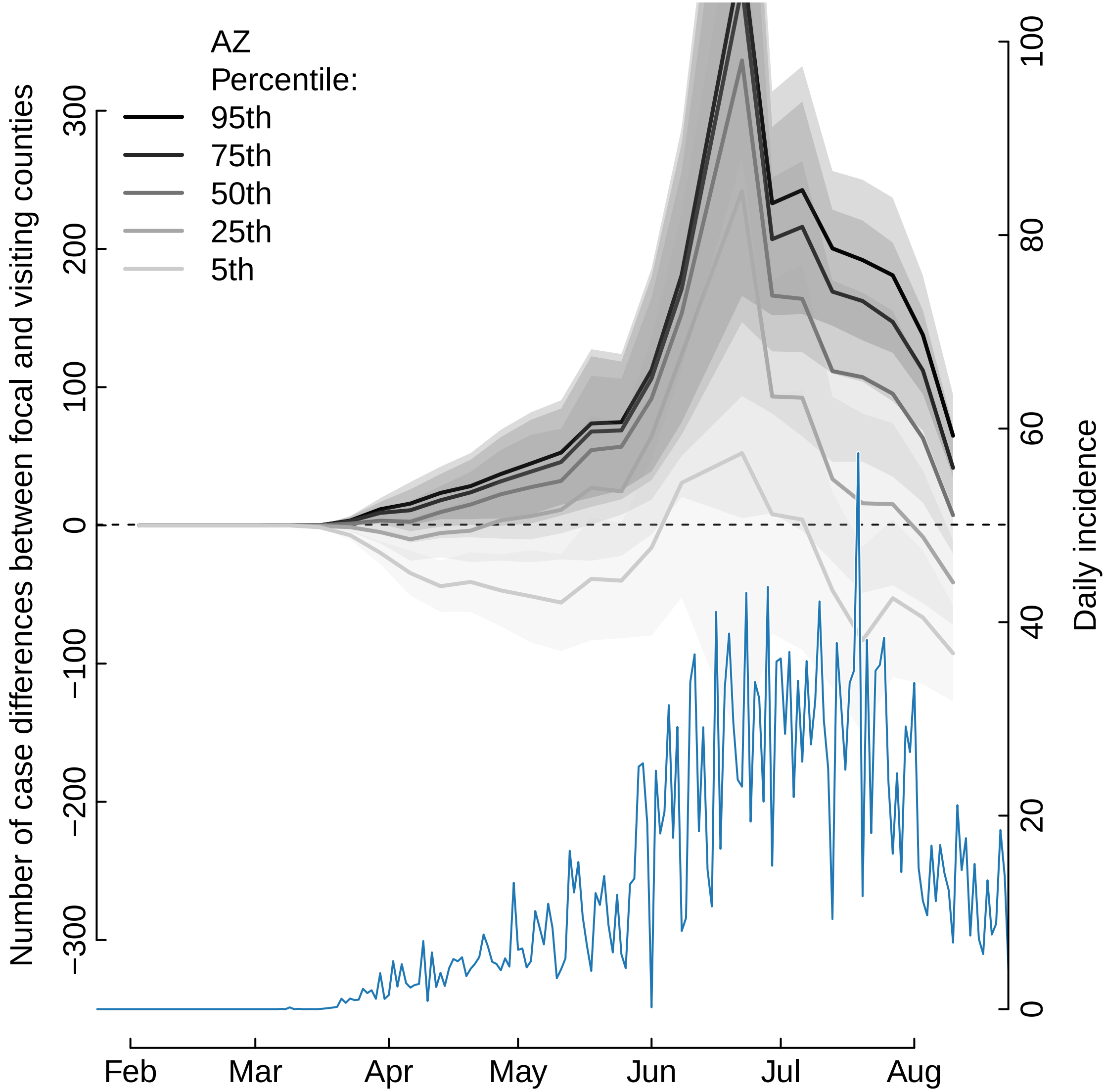

AZ
Percentile:
95th
75th
50th
25th
5th
Number of case differences between focal and visiting counties
300
200
100
0
−100
−200
−300
Daily incidence
100
80
60
40
20
0
Feb
Mar
Apr
May
Jun
Jul
Aug

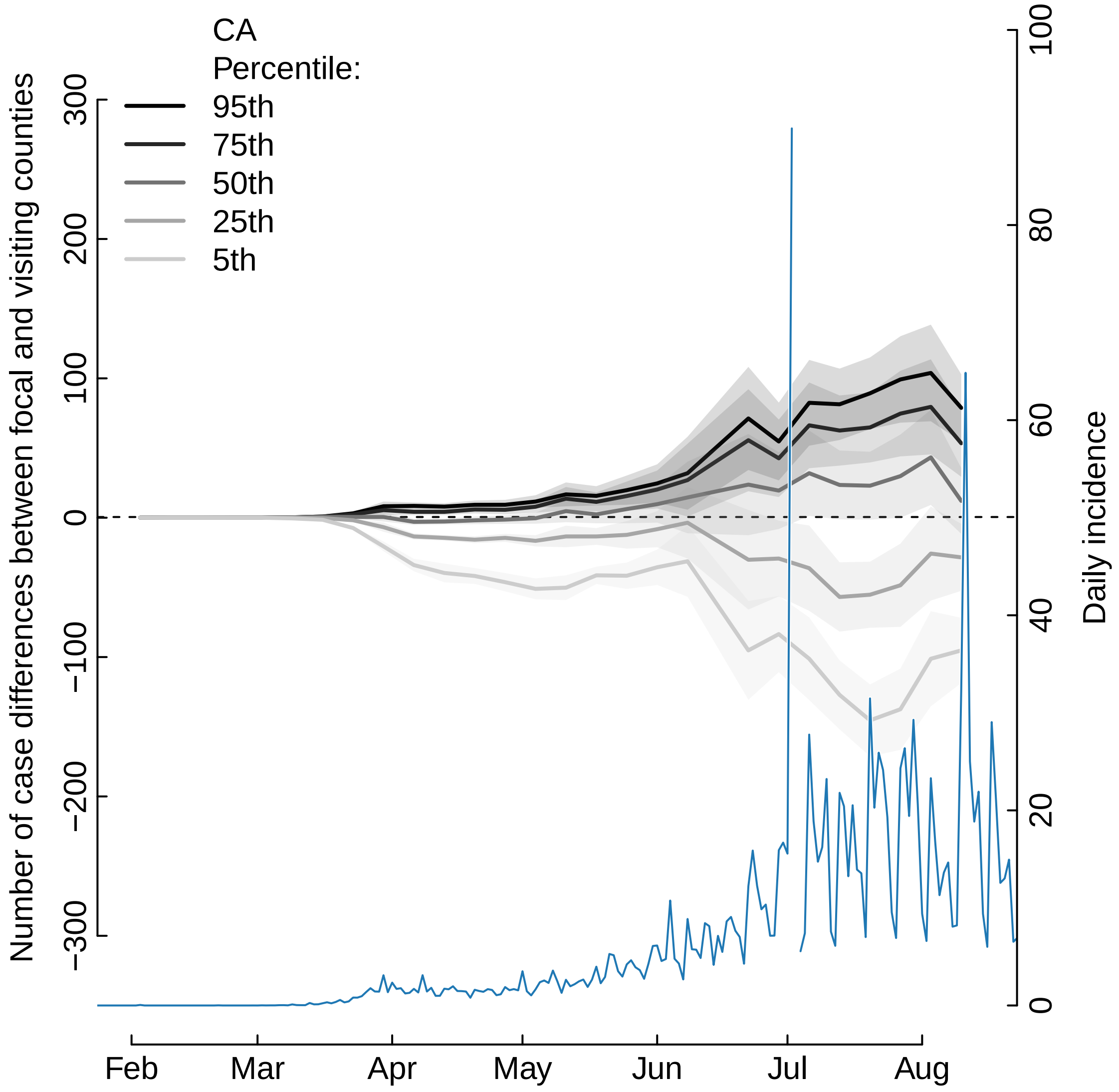

CA
Percentile:
95th
75th
50th
25th
5th
Number of case differences between focal and visiting counties
300
200
100
0
−100
−200
−300
Daily incidence
100
80
60
40
20
0
Feb
Mar
Apr
May
Jun
Jul
Aug

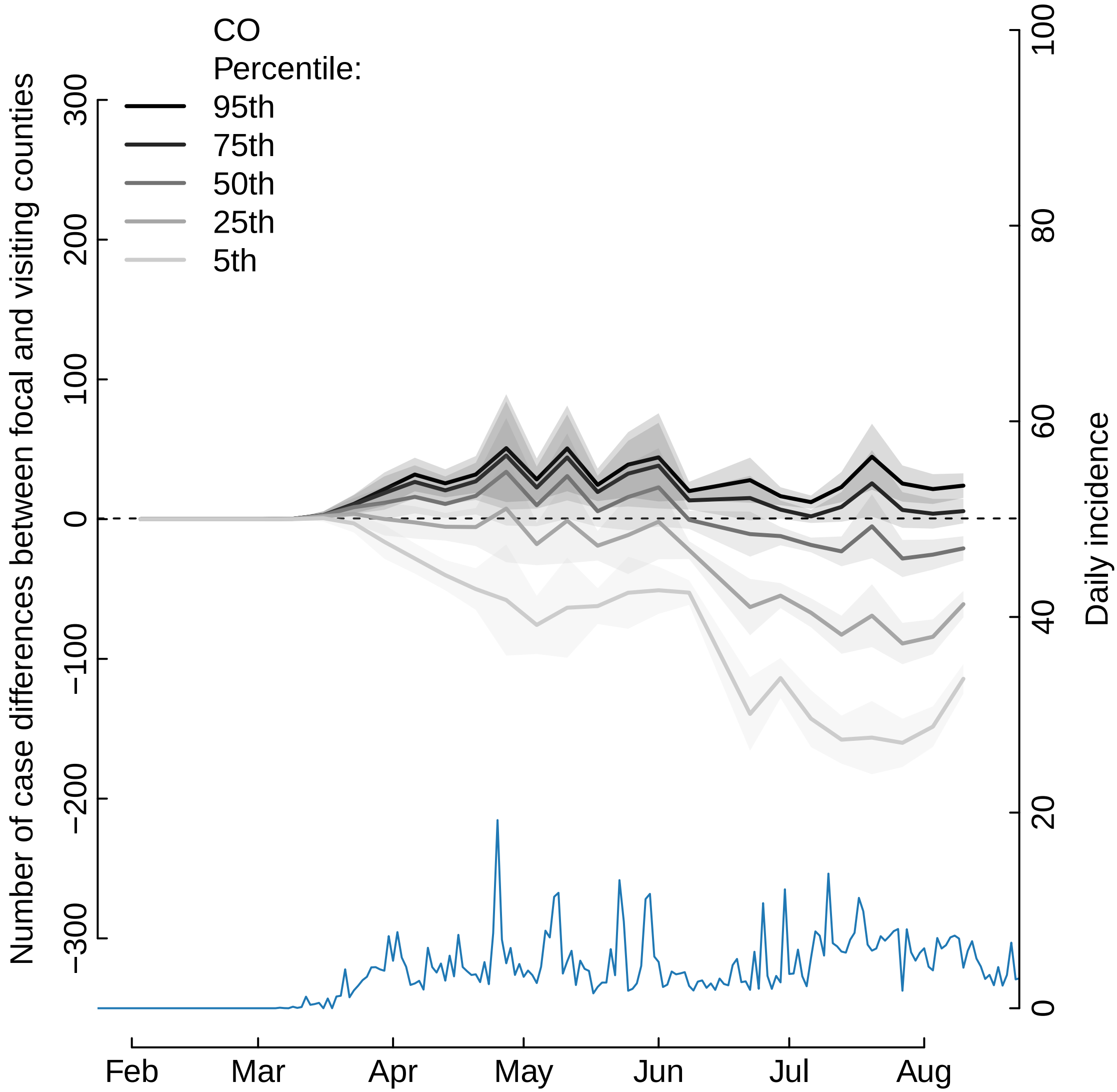
CO
Percentile:
95th
75th
50th
25th
5th
Number of case differences between focal and visiting counties
300
200
100
0
−100
−200
−300
Daily incidence
100
80
60
40
20
0
Feb
Mar
Apr
May
Jun
Jul
Aug

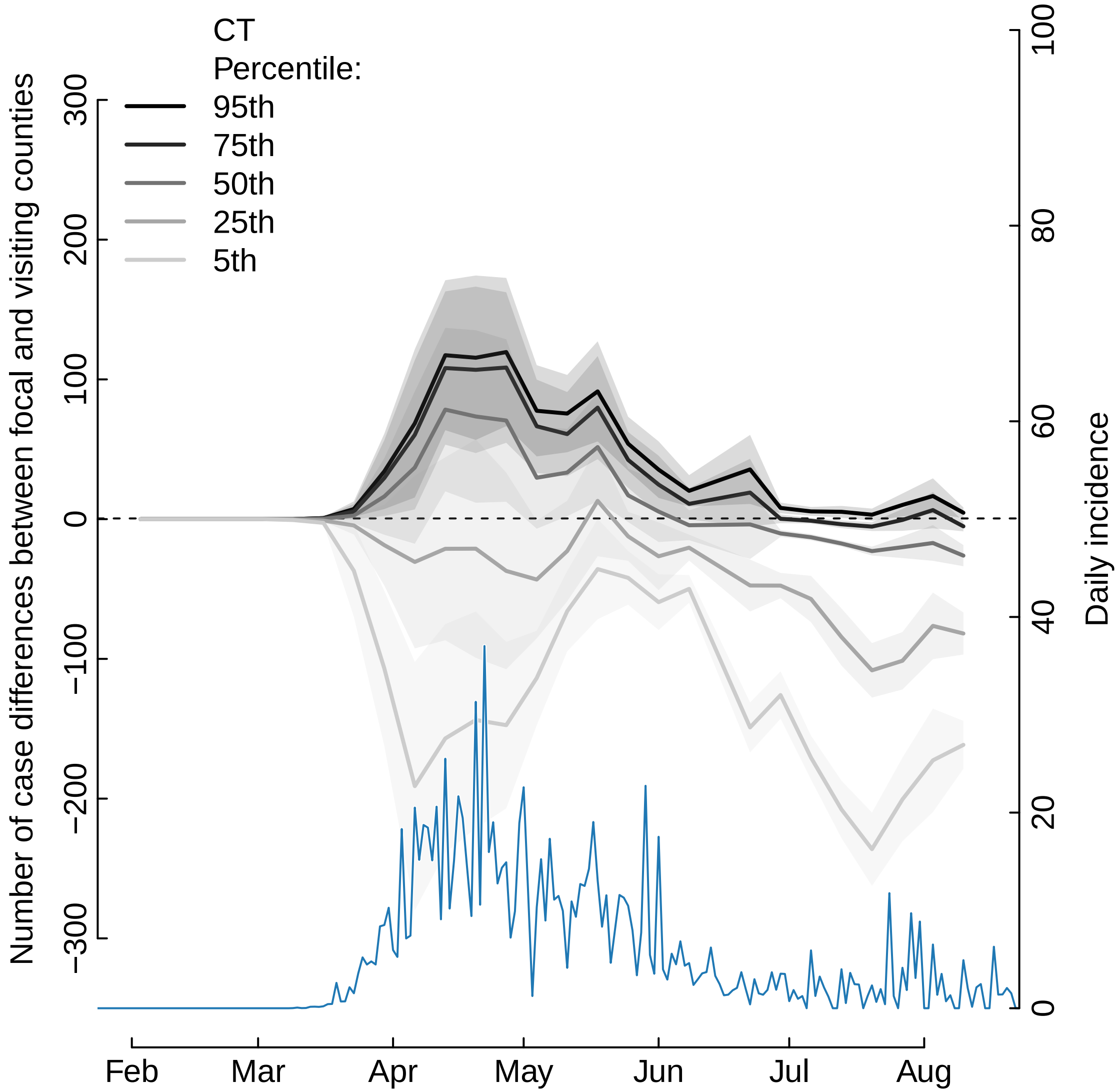

CT
Percentile:
95th
75th
50th
25th
5th
Number of case differences between focal and visiting counties
300
200
100
0
−100
−200
−300
Daily incidence
100
80
60
40
20
0
Feb
Mar
Apr
May
Jun
Jul
Aug

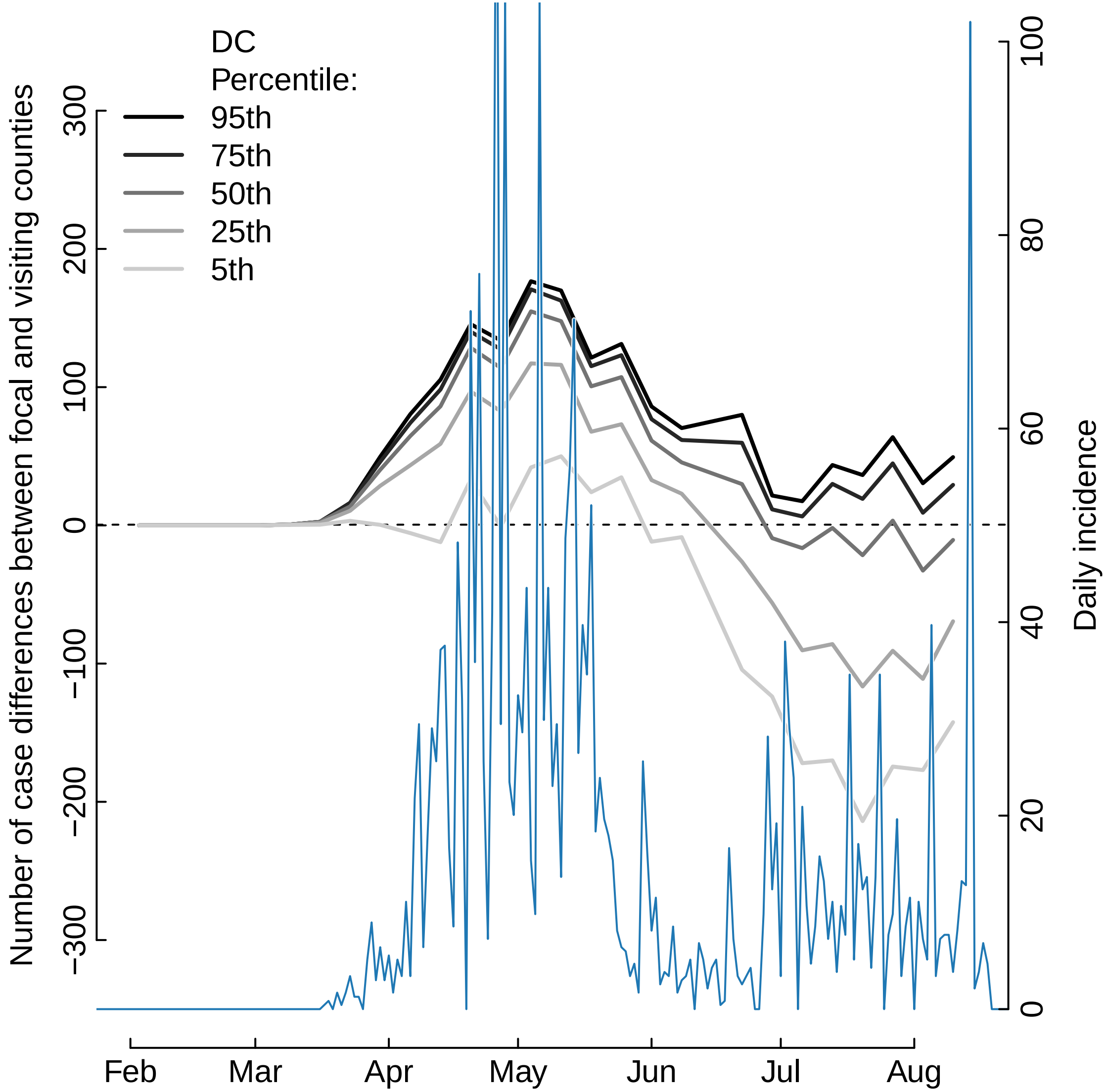
DC
Percentile:
95th
75th
50th
25th
5th
Number of case differences between focal and visiting counties
300
200
100
0
−100
−200
−300
Daily incidence
100
80
60
40
20
0
Feb
Mar
Apr
May
Jun
Jul
Aug

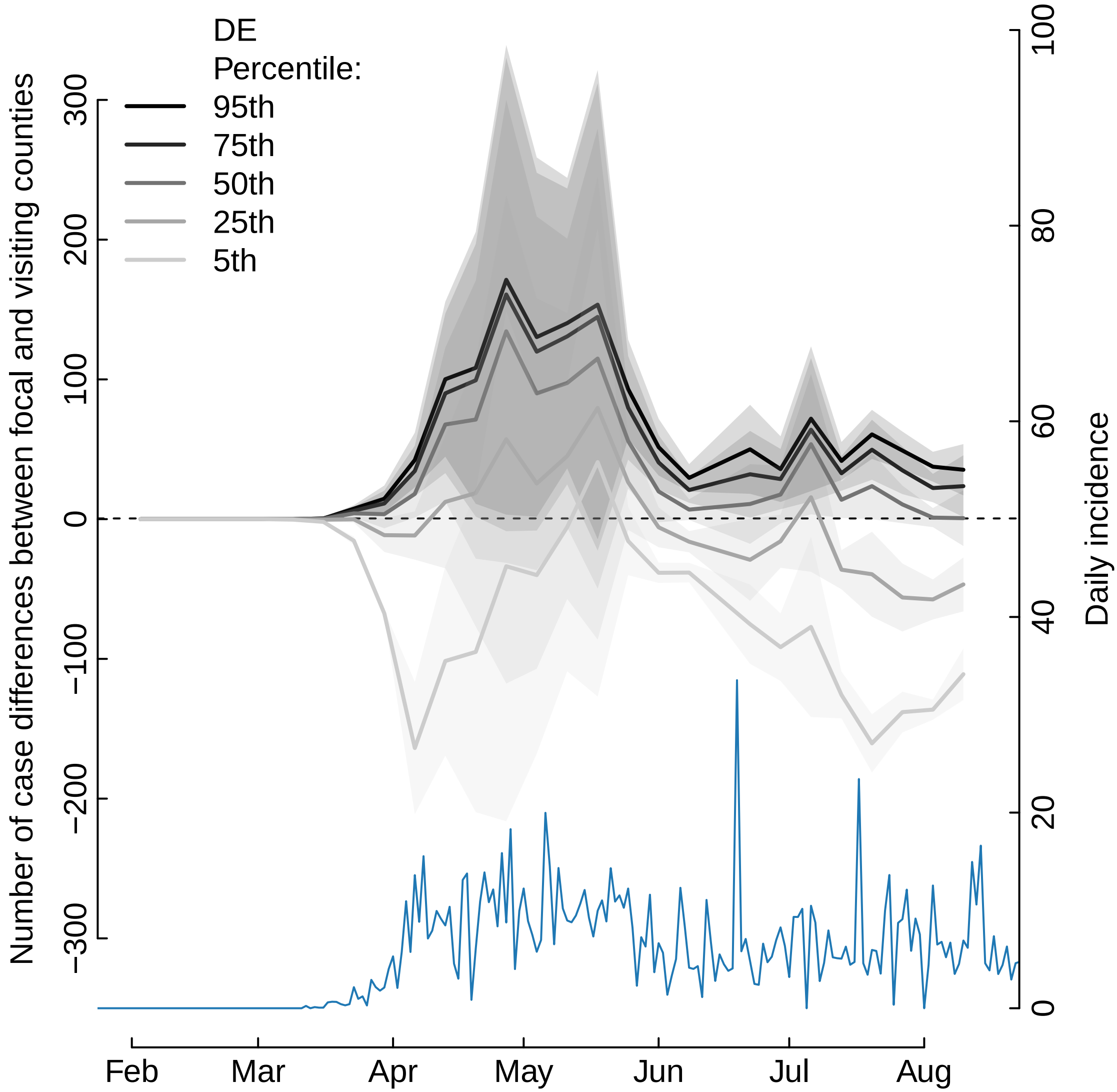
DE
Percentile:
95th
75th
50th
25th
5th
Number of case differences between focal and visiting counties
300
200
100
0
−100
−200
−300
Daily incidence
100
80
60
40
20
0
Feb
Mar
Apr
May
Jun
Jul
Aug

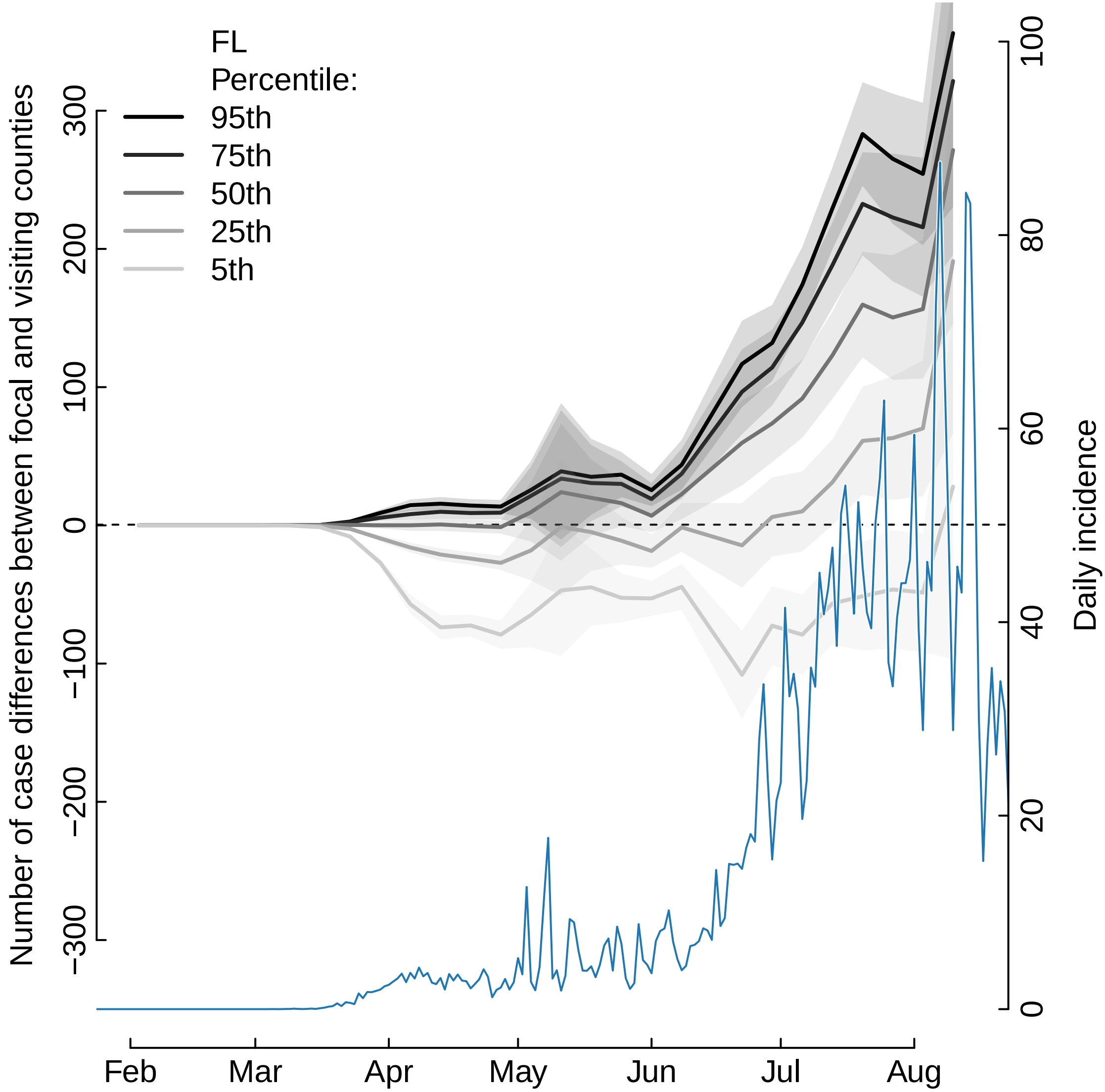

FL
Percentile:
95th
75th
50th
25th
5th
Number of case differences between focal and visiting counties
300
200
100
0
−100
−200
−300
Daily incidence
100
80
60
40
20
0
Feb
Mar
Apr
May
Jun
Jul
Aug

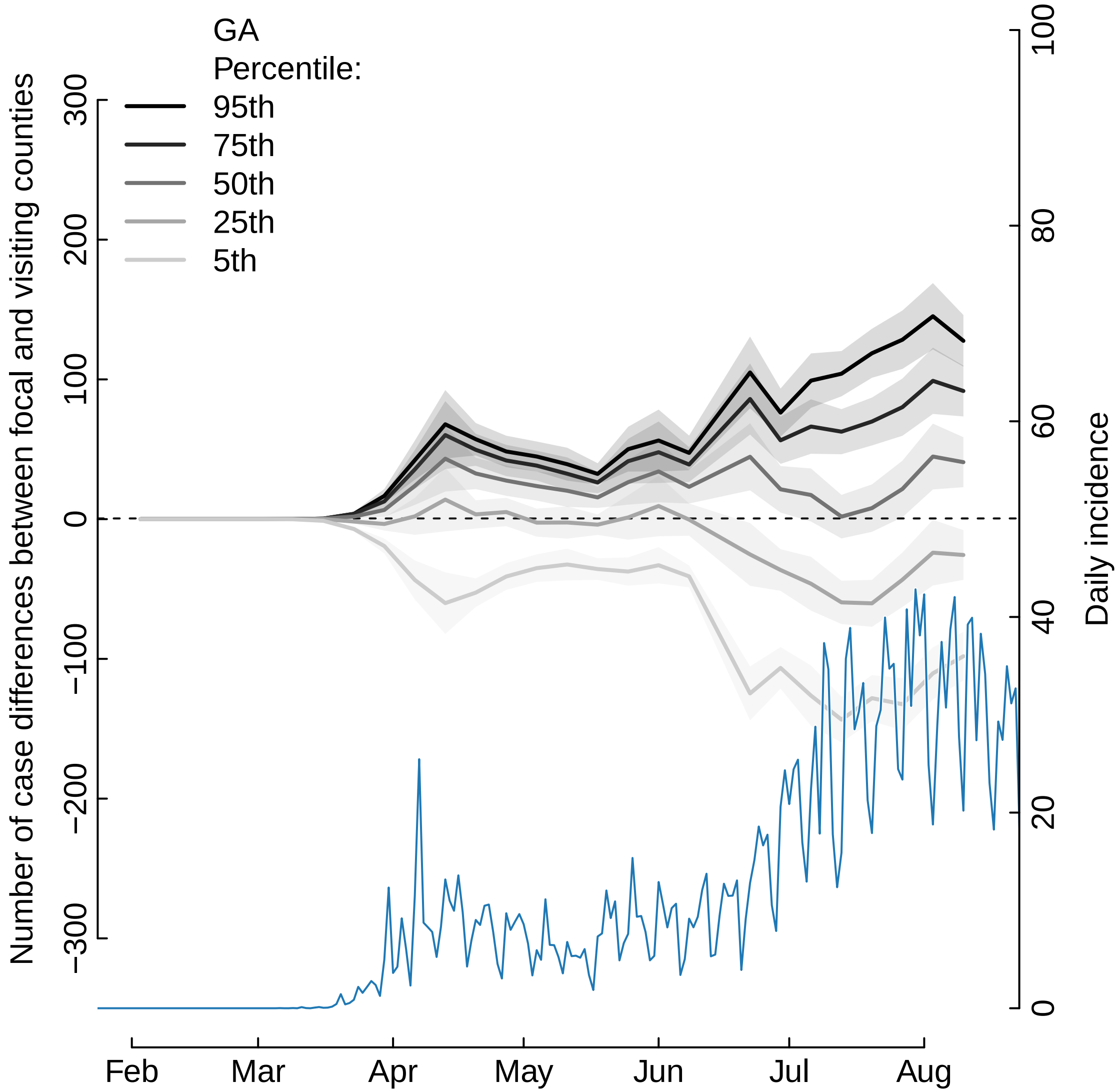
GA
Percentile:
95th
75th
50th
25th
5th
Number of case differences between focal and visiting counties
Daily incidence
300
200
100
0
−100
−200
−300
100
80
60
40
20
0
Feb
Mar
Apr
May
Jun
Jul
Aug

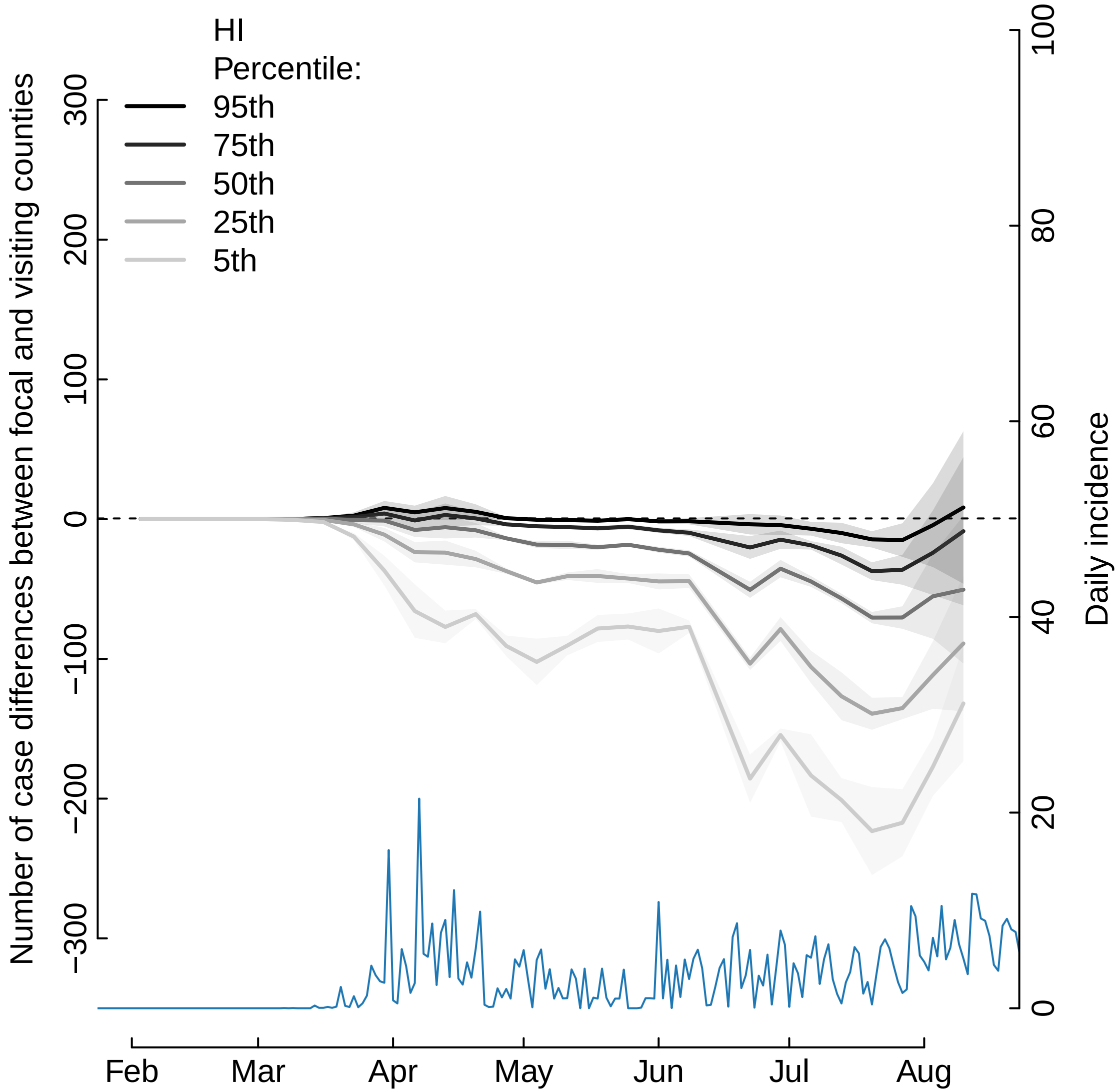

HI
Percentile:
95th
75th
50th
25th
5th
Number of case differences between focal and visiting counties
300
200
100
0
−100
−200
−300
Daily incidence
100
80
60
40
20
0
Feb
Mar
Apr
May
Jun
Jul
Aug

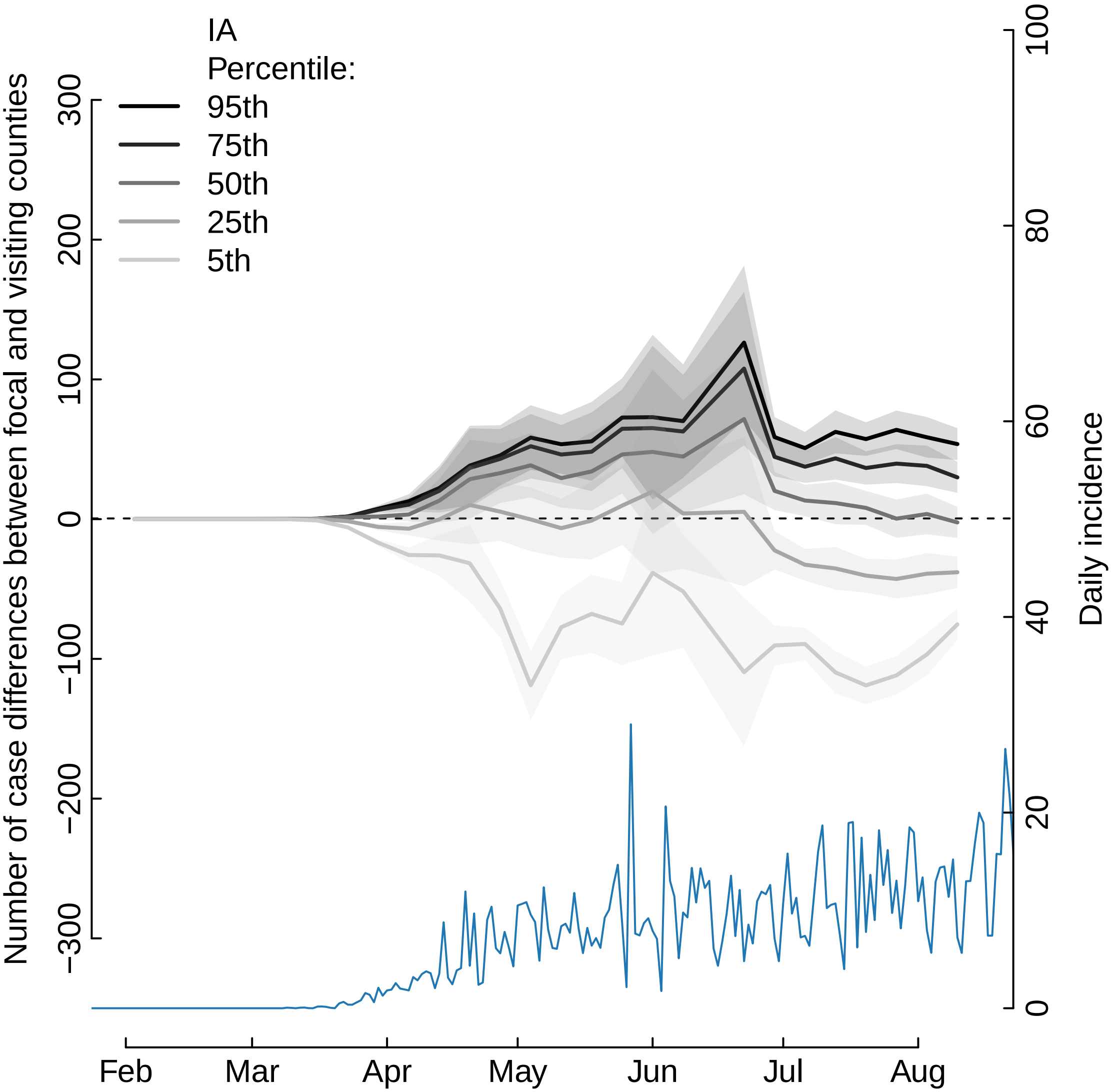

IA
Percentile:
95th
75th
50th
25th
5th
Number of case differences between focal and visiting counties
300
200
100
0
−100
−200
−300
Daily incidence
100
80
60
40
20
0
Feb
Mar
Apr
May
Jun
Jul
Aug

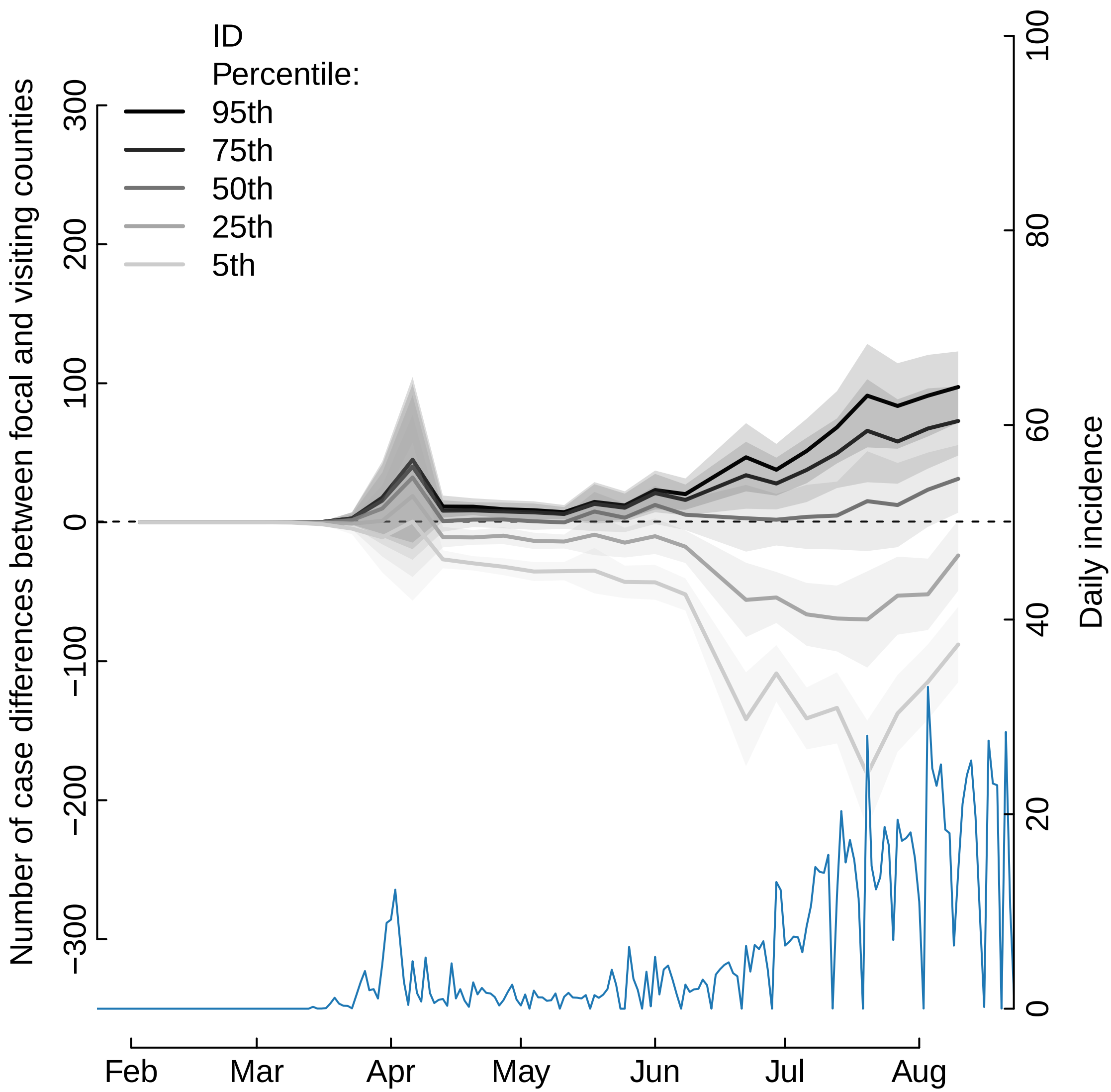
ID
Percentile:
95th
75th
50th
25th
5th
Number of case differences between focal and visiting counties
300
200
100
0
−100
−200
−300
Daily incidence
100
80
60
40
20
0
Feb
Mar
Apr
May
Jun
Jul
Aug

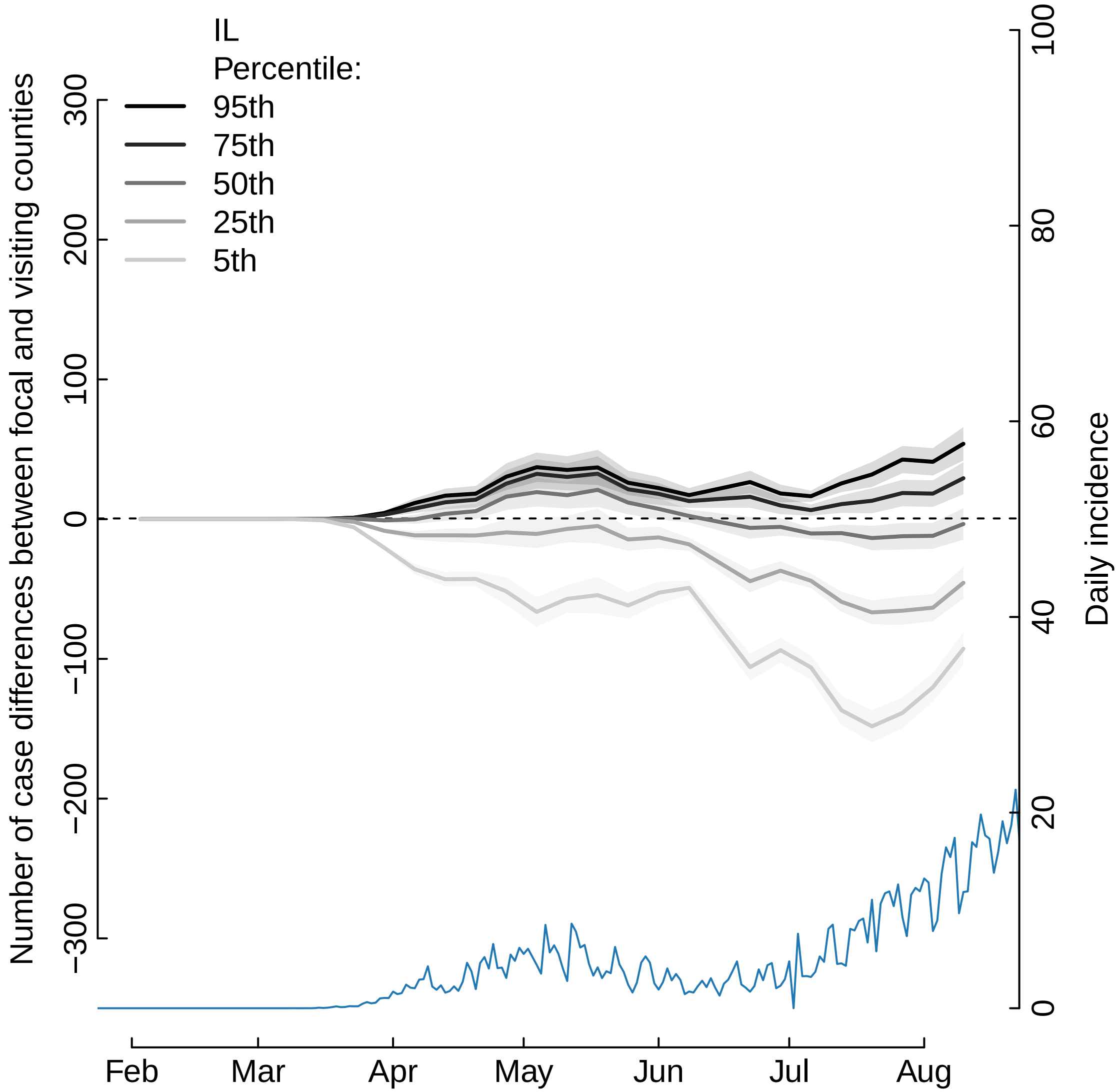

IL
Percentile:
95th
75th
50th
25th
5th
Number of case differences between focal and visiting counties
300
200
100
0
−100
−200
−300
Daily incidence
100
80
60
40
20
0
Feb
Mar
Apr
May
Jun
Jul
Aug

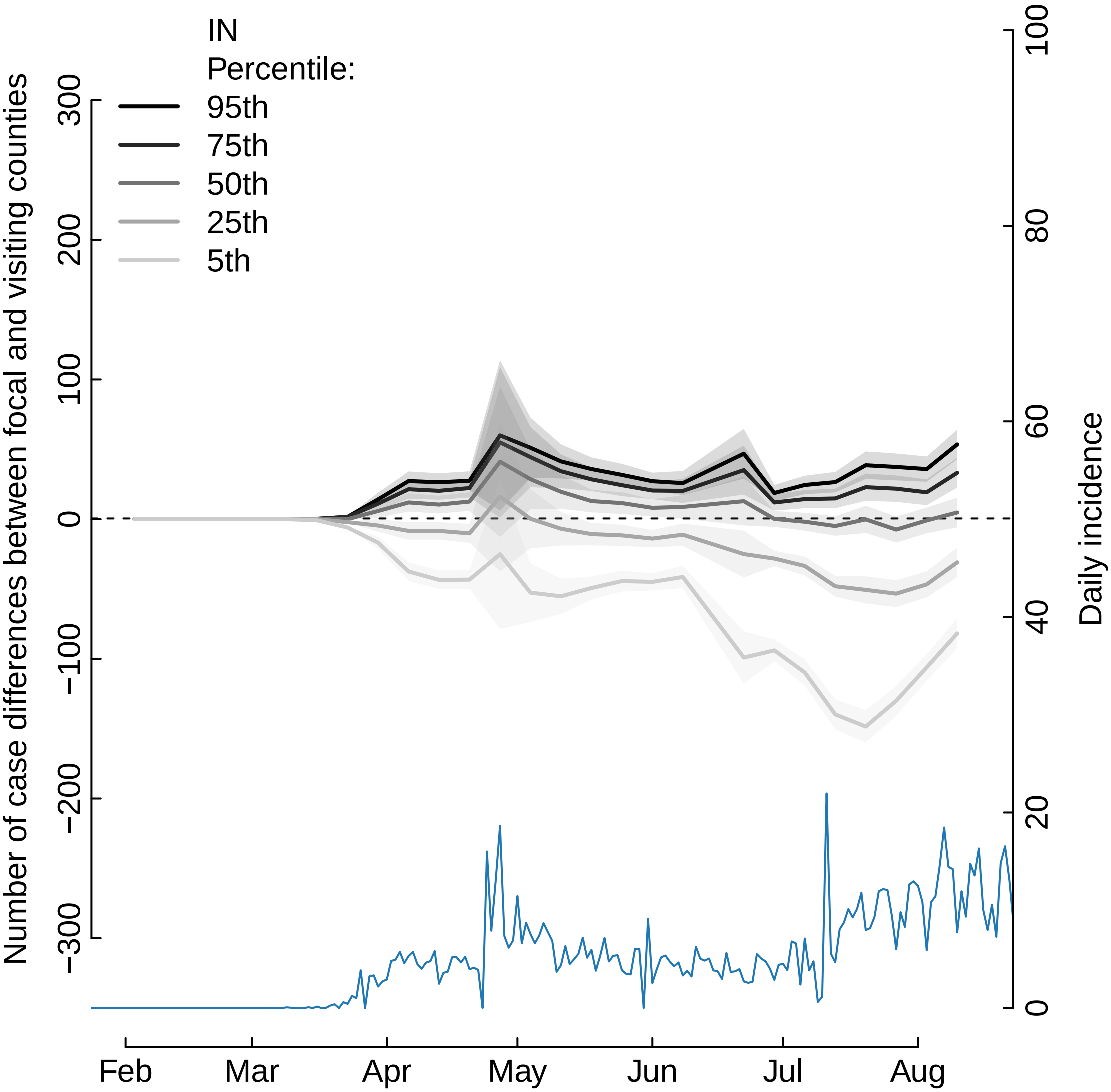
IN
Percentile:
95th
75th
50th
25th
5th
Number of case differences between focal and visiting counties
300
200
100
0
−100
−200
−300
Daily incidence
100
80
60
40
20
0
Feb
Mar
Apr
May
Jun
Jul
Aug

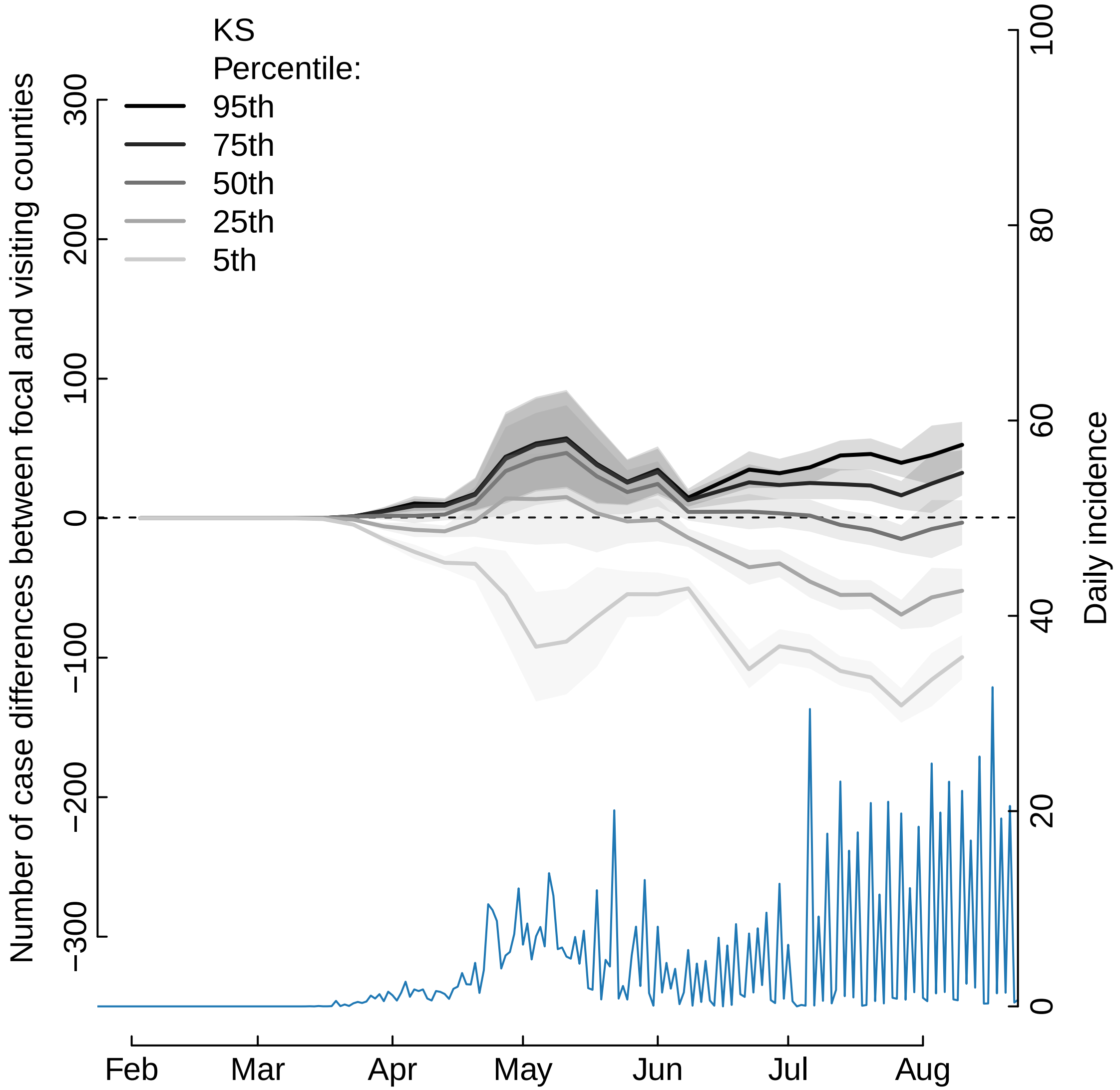

KS
Percentile:
95th
75th
50th
25th
5th
Number of case differences between focal and visiting counties
300
200
100
0
−100
−200
−300
Daily incidence
100
80
60
40
20
0
Feb
Mar
Apr
May
Jun
Jul
Aug

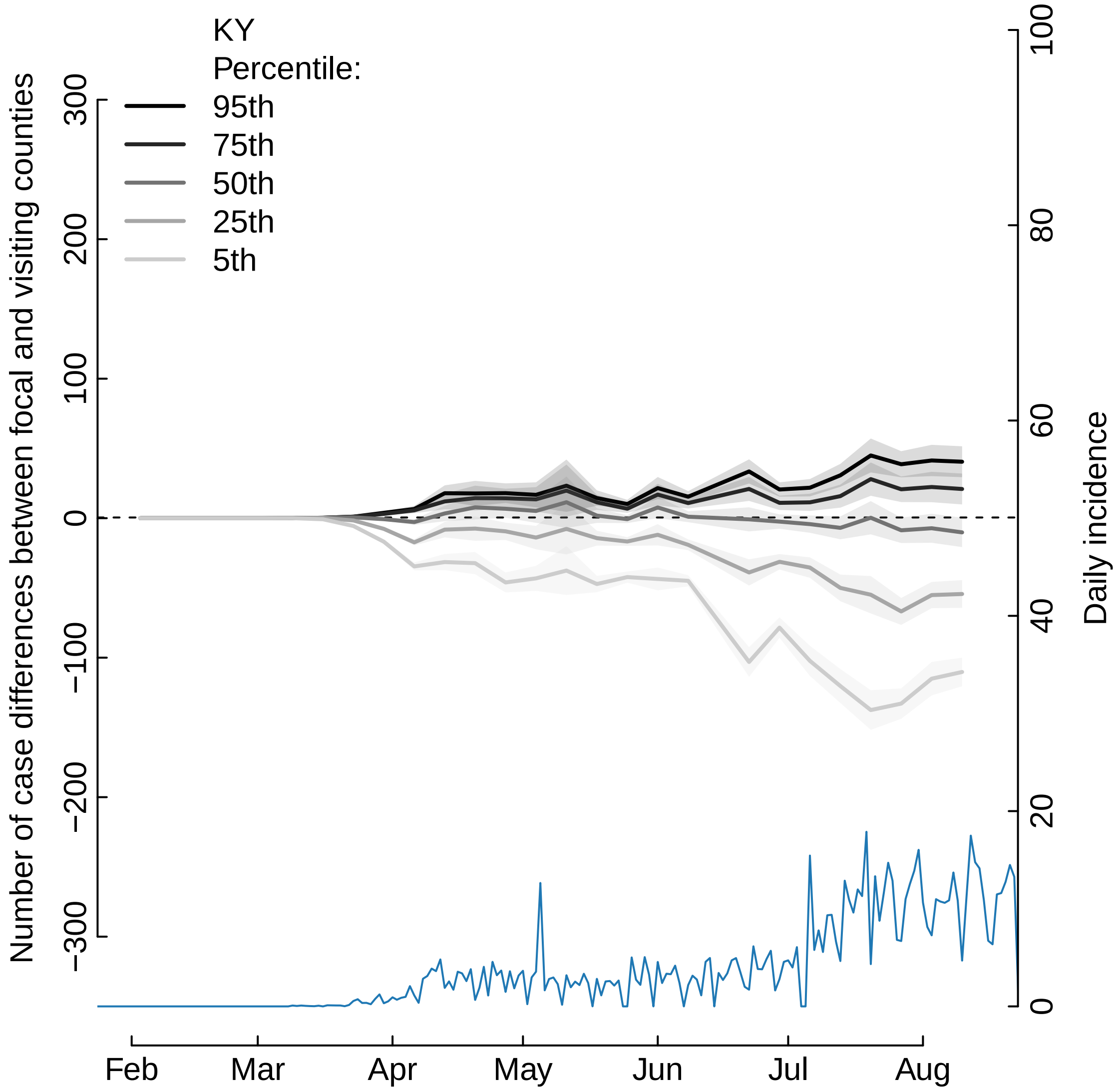
KY
Percentile:
95th
75th
50th
25th
5th
Number of case differences between focal and visiting counties
Daily incidence
300
200
100
0
−100
−200
−300
100
80
60
40
20
0
Feb
Mar
Apr
May
Jun
Jul
Aug

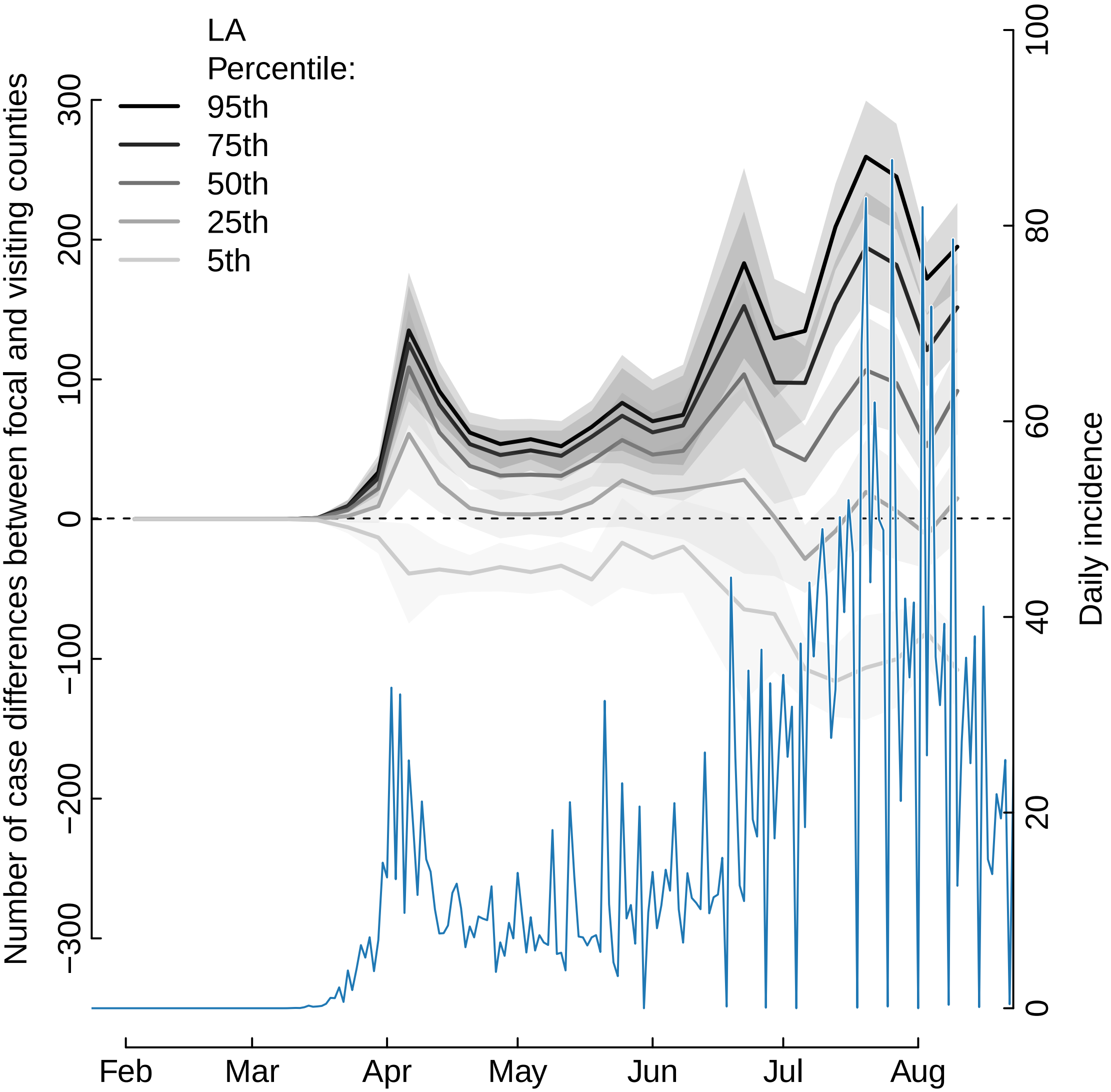
LA
Percentile:
95th
75th
50th
25th
5th
Number of case differences between focal and visiting counties
300
200
100
0
−100
−200
−300
Daily incidence
100
80
60
40
20
0
Feb
Mar
Apr
May
Jun
Jul
Aug

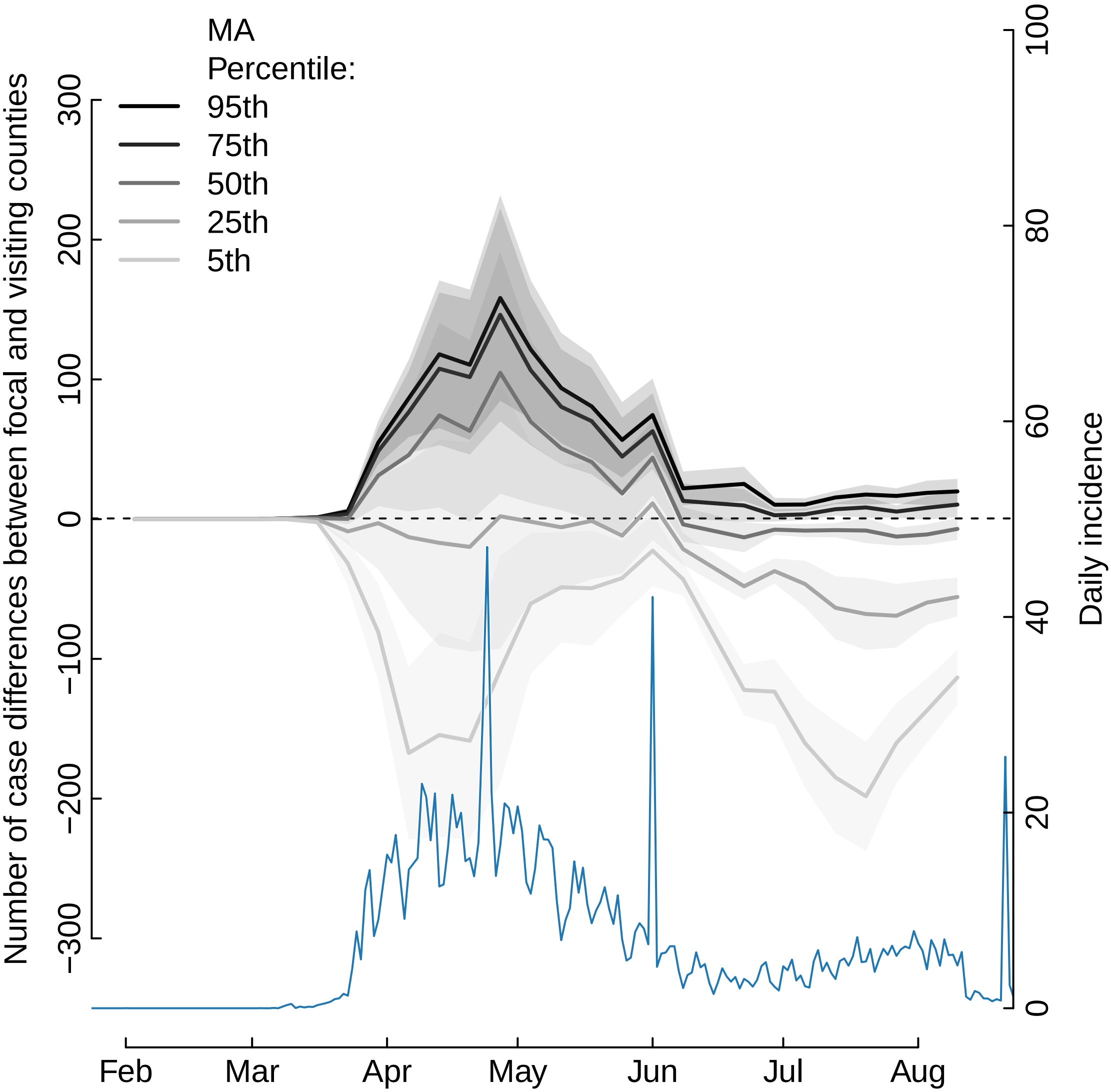

MA
Percentile:
95th
75th
50th
25th
5th
Number of case differences between focal and visiting counties
300
200
100
0
−100
−200
−300
Daily incidence
100
80
60
40
20
0
Feb
Mar
Apr
May
Jun
Jul
Aug

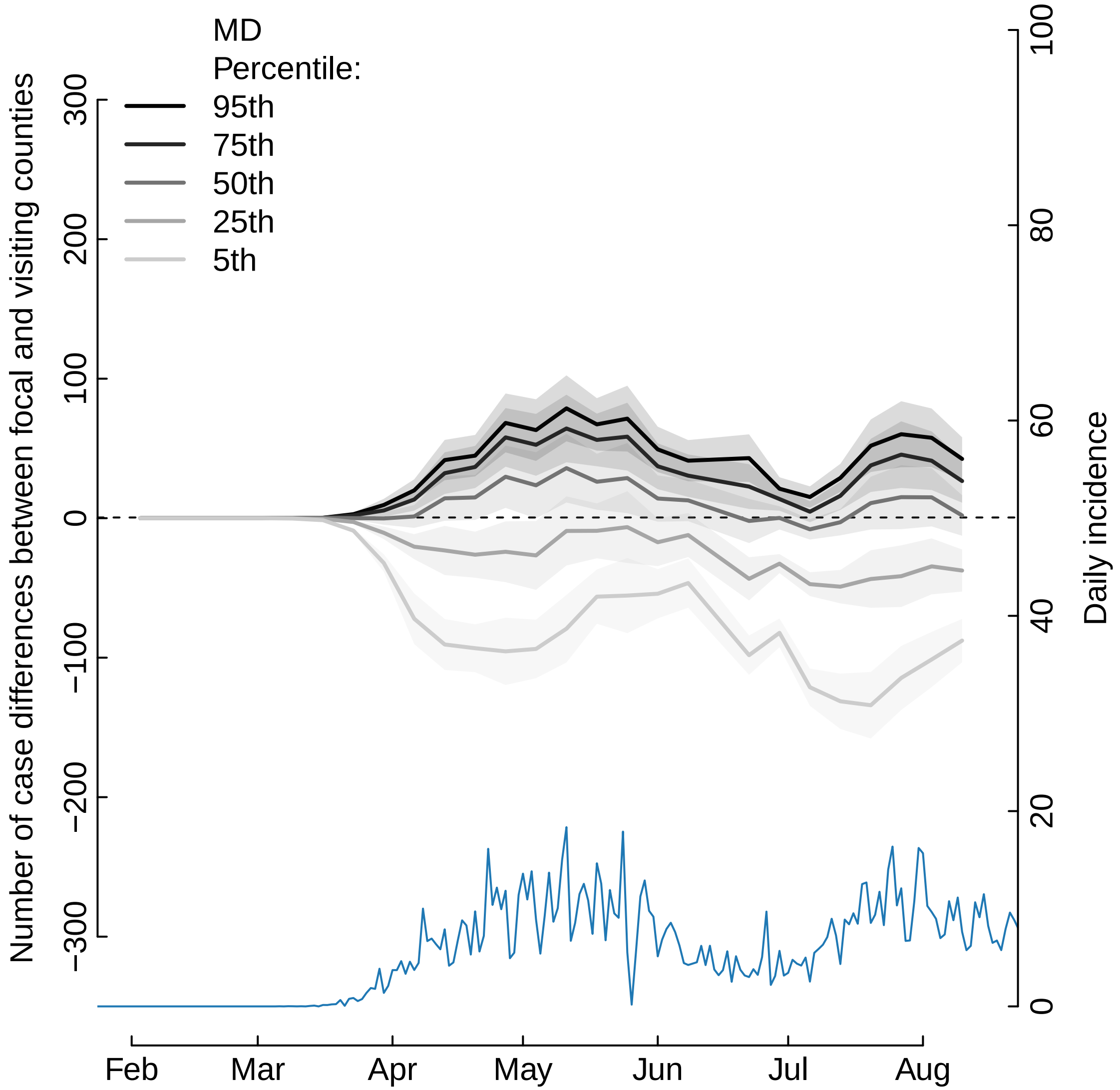

MD
Percentile:
95th
75th
50th
25th
5th
Number of case differences between focal and visiting counties
300
200
100
0
−100
−200
−300
Daily incidence
100
80
60
40
20
0
Feb
Mar
Apr
May
Jun
Jul
Aug

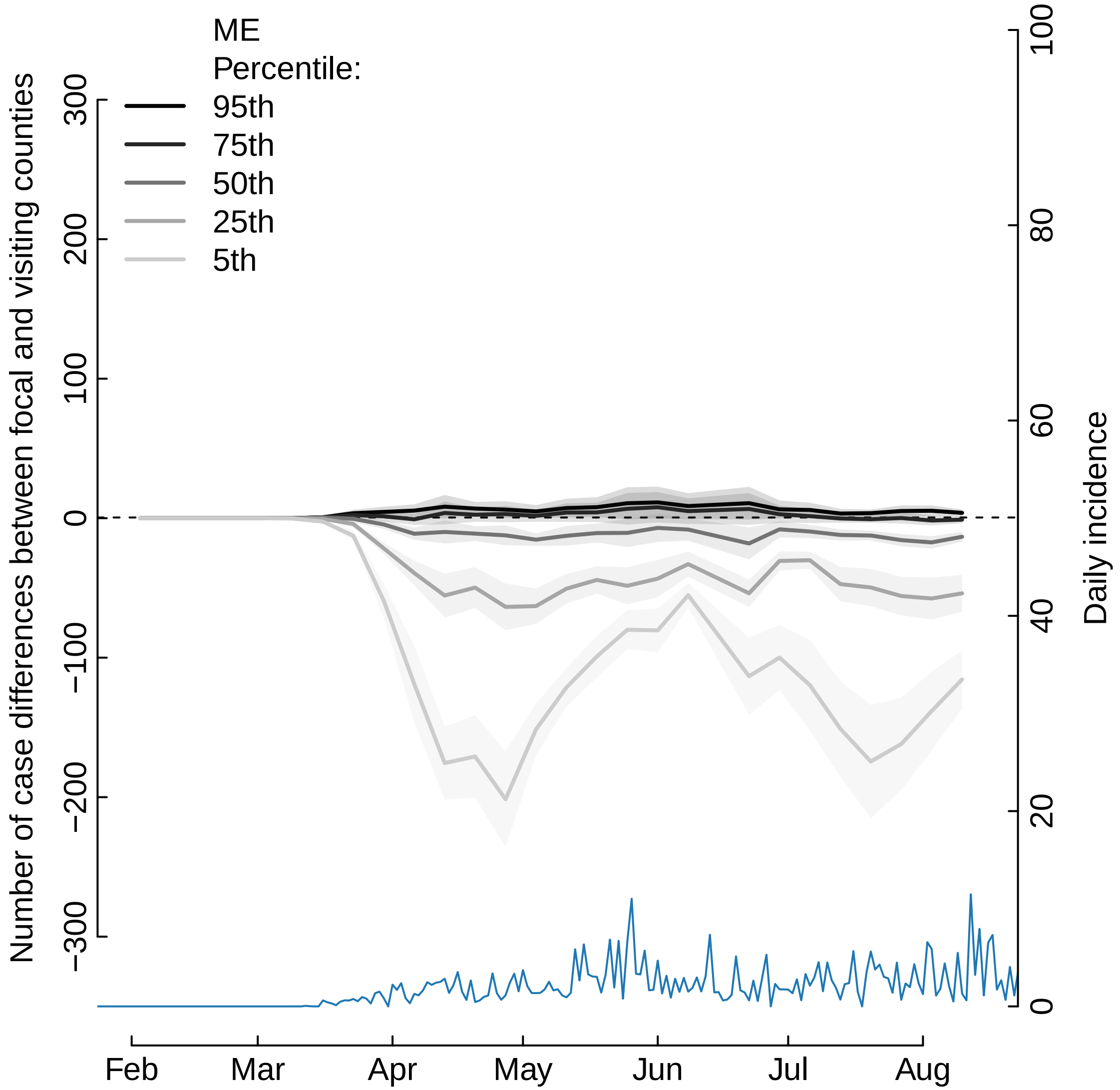

ME
Percentile:
95th
75th
50th
25th
5th
Number of case differences between focal and visiting counties
300
200
100
0
−100
−200
−300
Daily incidence
100
80
60
40
20
0
Feb
Mar
Apr
May
Jun
Jul
Aug

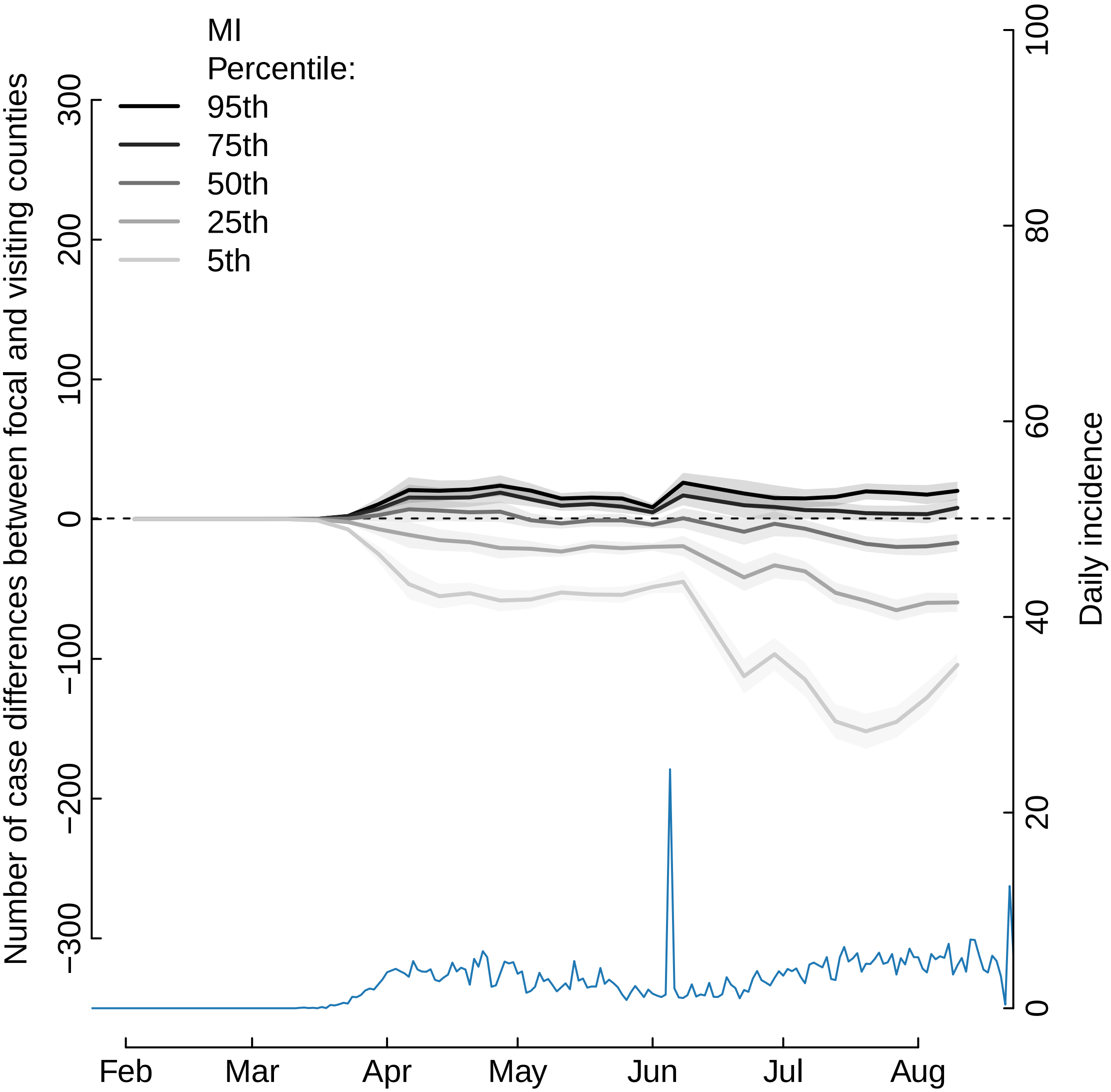

MI
Percentile:
95th
75th
50th
25th
5th
Number of case differences between focal and visiting counties
300
200
100
0
−100
−200
−300
Daily incidence
100
80
60
40
20
0
Feb
Mar
Apr
May
Jun
Jul
Aug

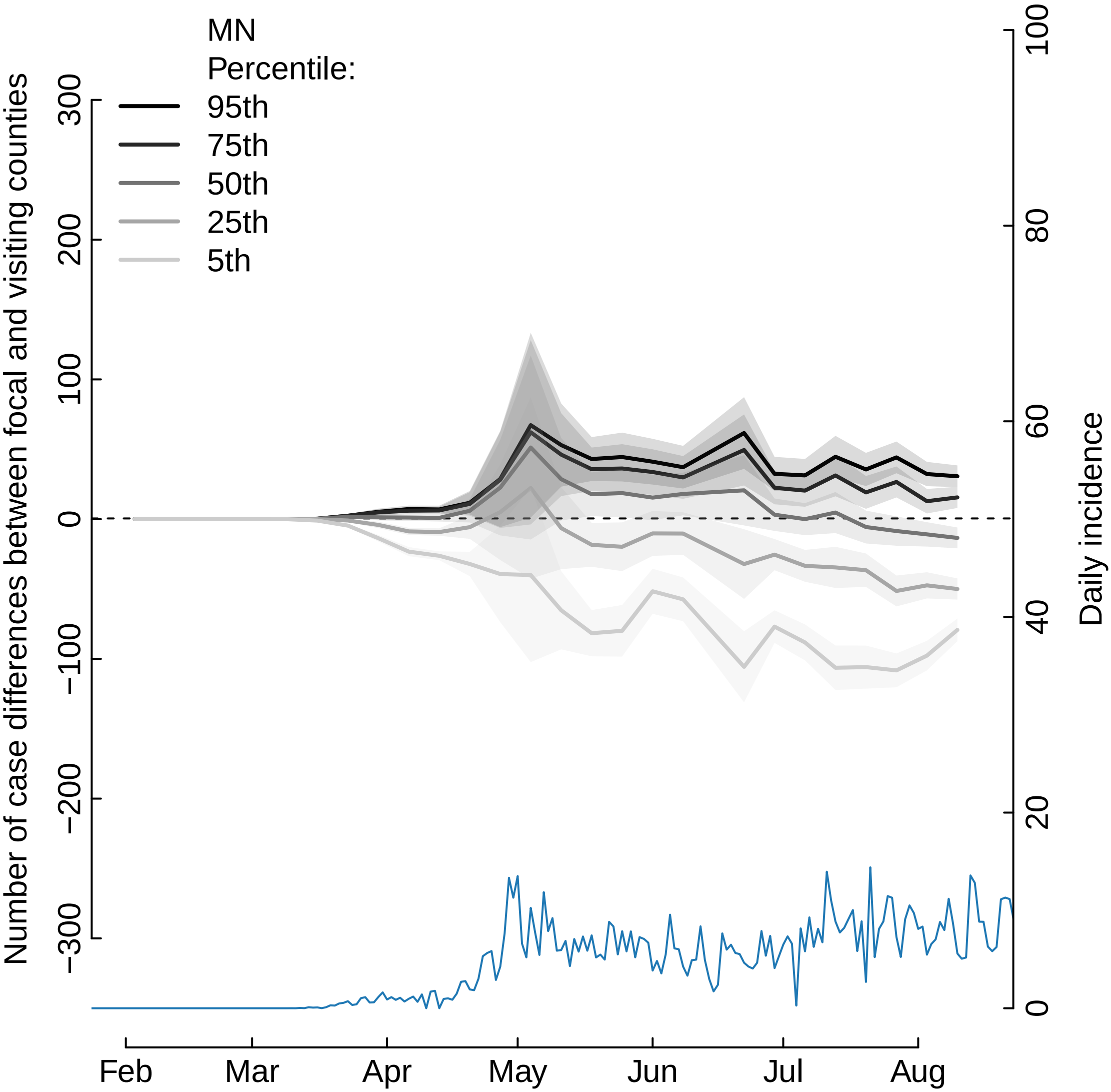
MN
Percentile:
95th
75th
50th
25th
5th
Number of case differences between focal and visiting counties
300
200
100
0
−100
−200
−300
Daily incidence
100
80
60
40
20
0
Feb
Mar
Apr
May
Jun
Jul
Aug

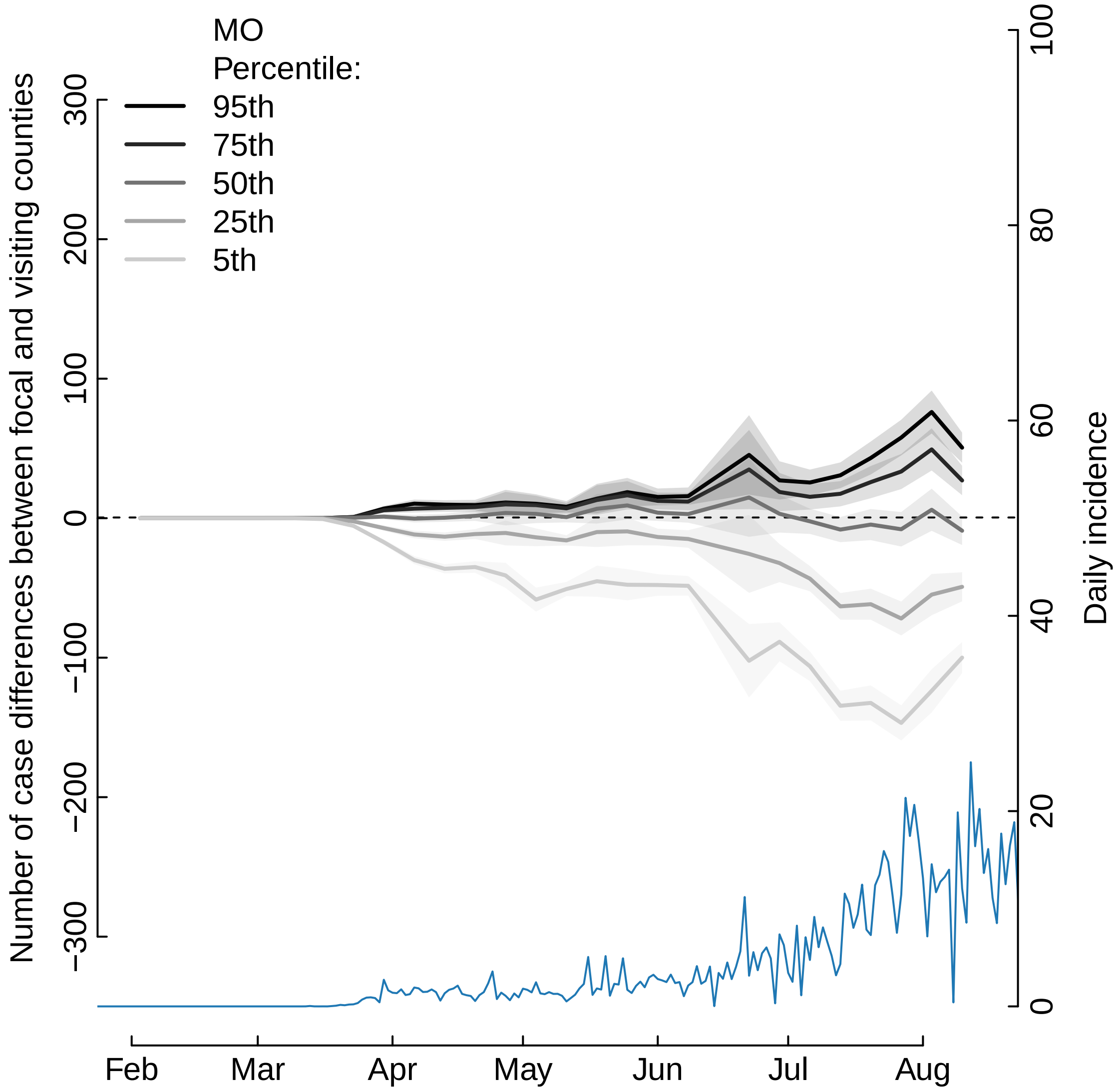
MO
Percentile:
95th
75th
50th
25th
5th
Number of case differences between focal and visiting counties
300
200
100
0
−100
−200
−300
Daily incidence
100
80
60
40
20
0
Feb
Mar
Apr
May
Jun
Jul
Aug

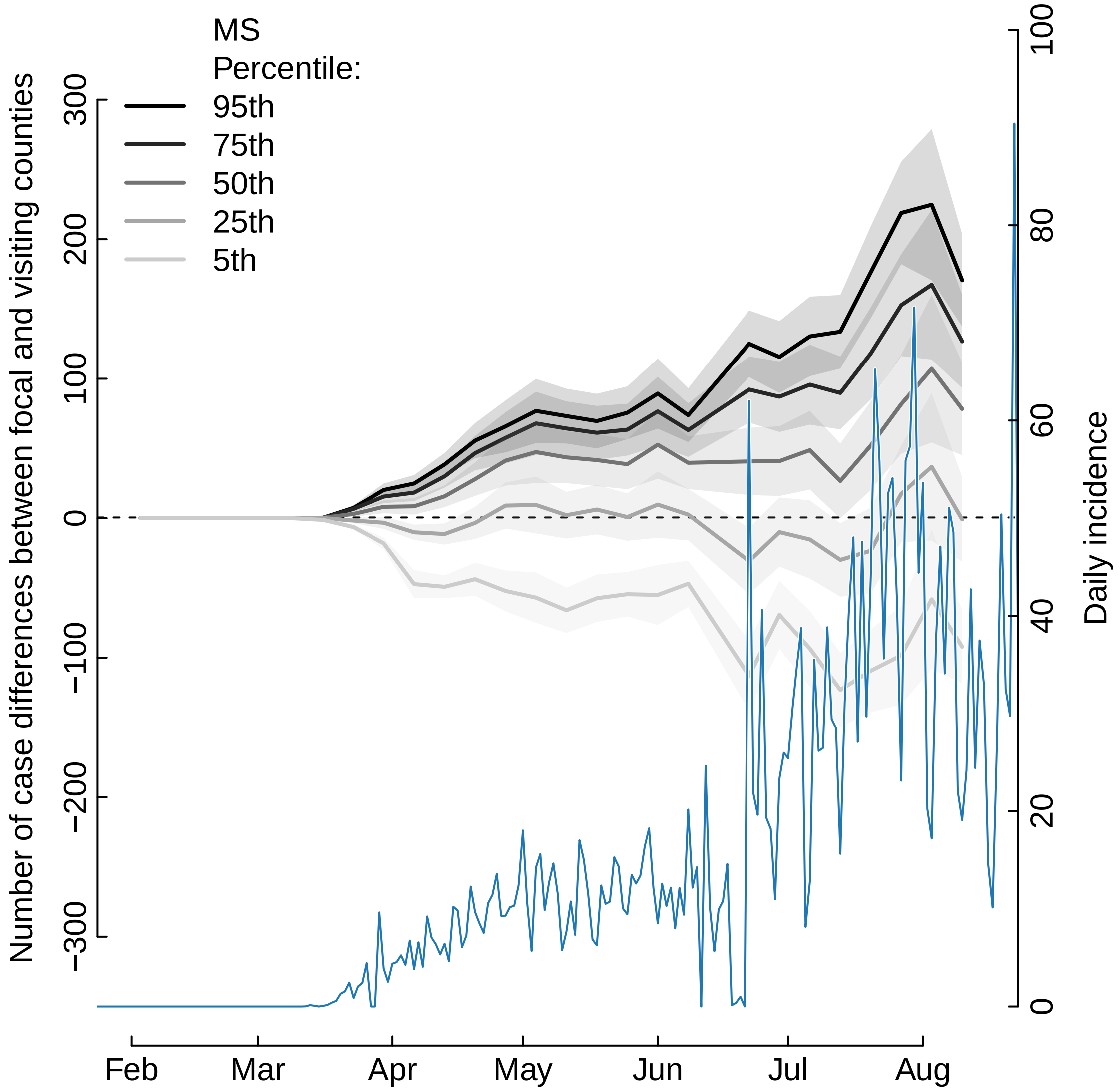

MS
Percentile:
95th
75th
50th
25th
5th
Number of case differences between focal and visiting counties
300
200
100
0
−100
−200
−300
Daily incidence
100
80
60
40
20
0
Feb
Mar
Apr
May
Jun
Jul
Aug

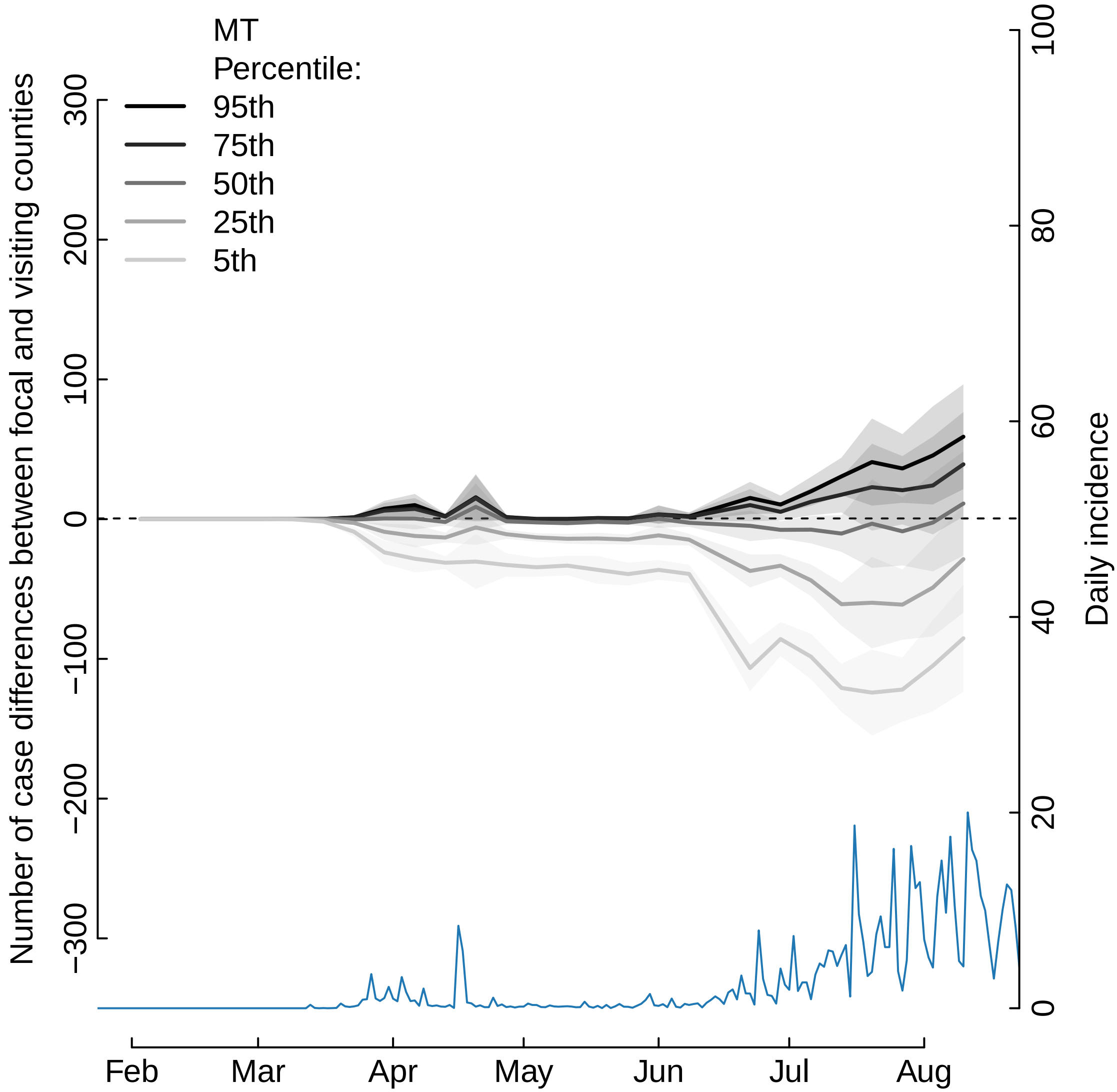

MT
Percentile:
95th
75th
50th
25th
5th
Number of case differences between focal and visiting counties
300
200
100
0
−100
−200
−300
Daily incidence
100
80
60
40
20
0
Feb
Mar
Apr
May
Jun
Jul
Aug

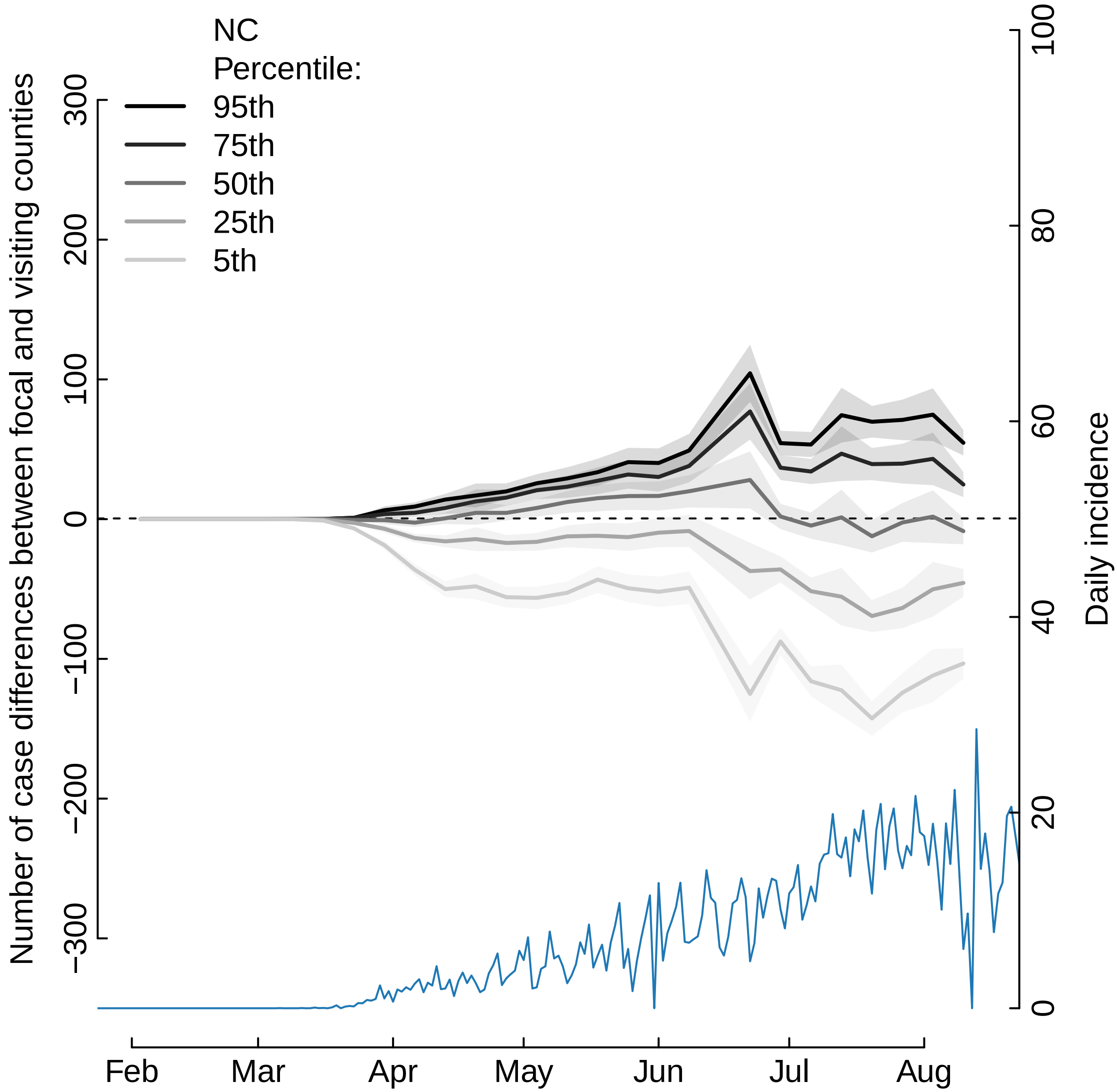
NC
Percentile:
95th
75th
50th
25th
5th
Number of case differences between focal and visiting counties
300
200
100
0
−100
−200
−300
Daily incidence
100
80
60
40
20
0
Feb
Mar
Apr
May
Jun
Jul
Aug

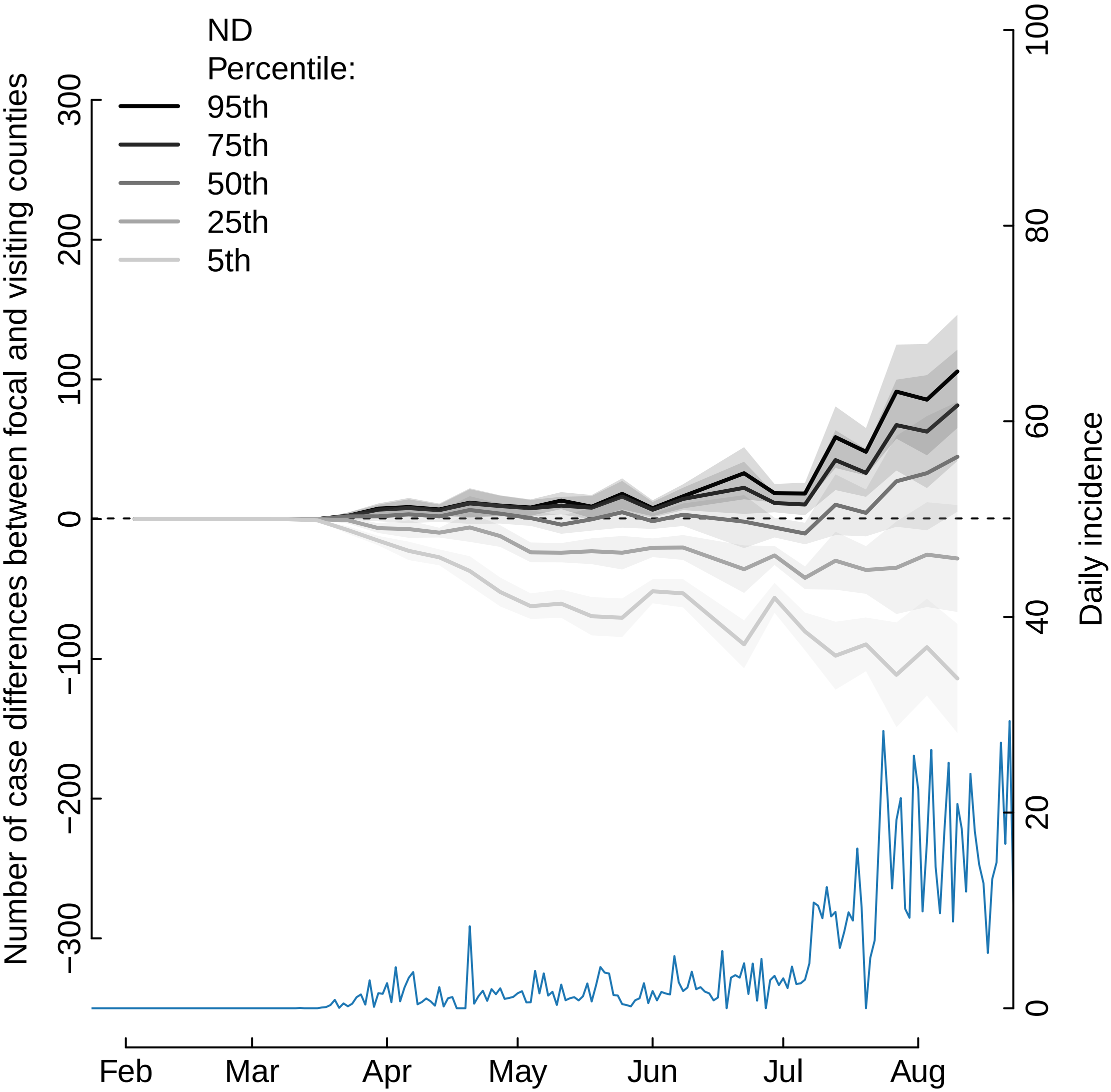

ND
Percentile:
95th
75th
50th
25th
5th
Number of case differences between focal and visiting counties
300
200
100
0
−100
−200
−300
Daily incidence
100
80
60
40
20
0
Feb
Mar
Apr
May
Jun
Jul
Aug

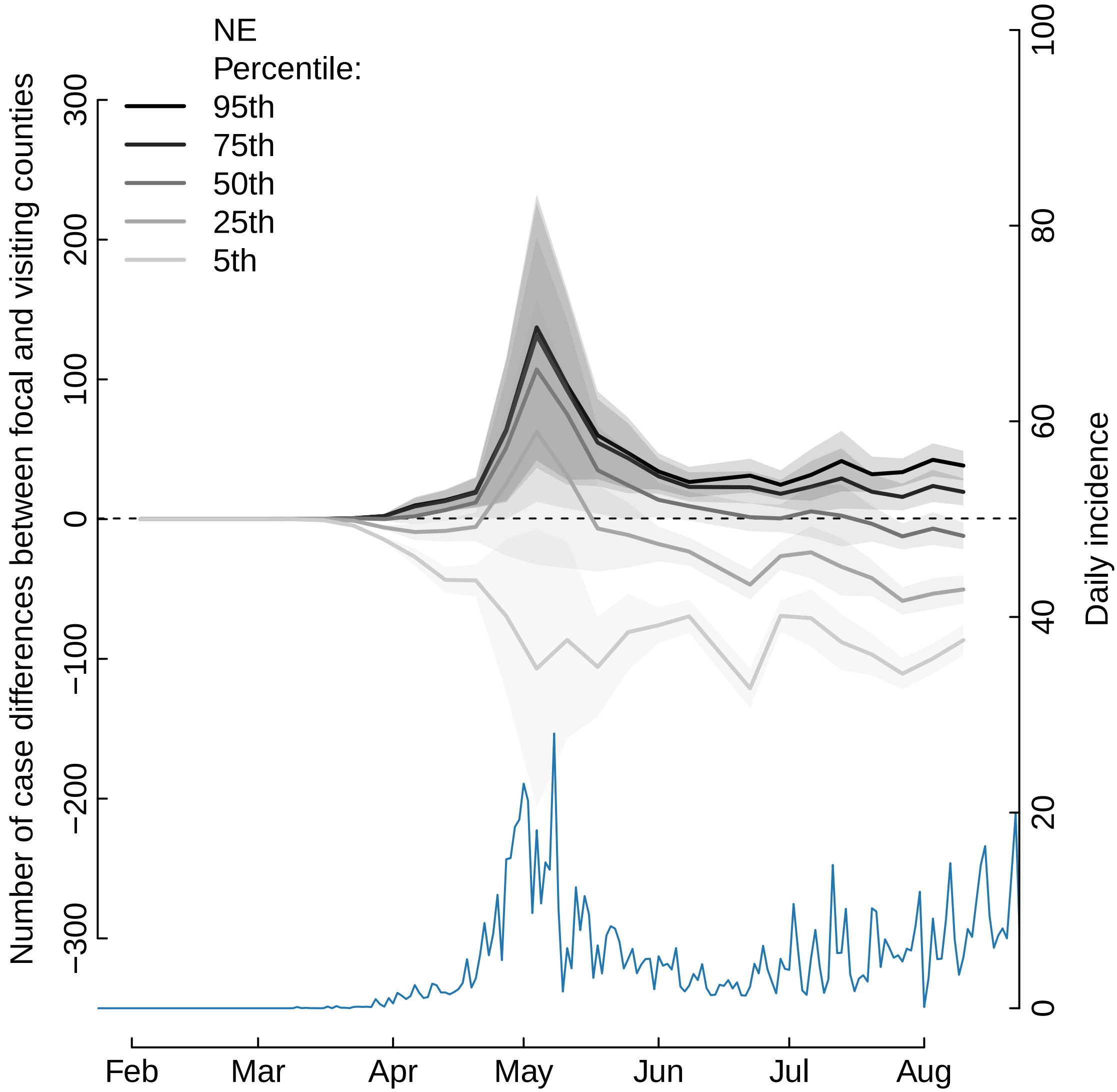

NE
Percentile:
95th
75th
50th
25th
5th
Number of case differences between focal and visiting counties
300
200
100
0
−100
−200
−300
Daily incidence
100
80
60
40
20
0
Feb
Mar
Apr
May
Jun
Jul
Aug

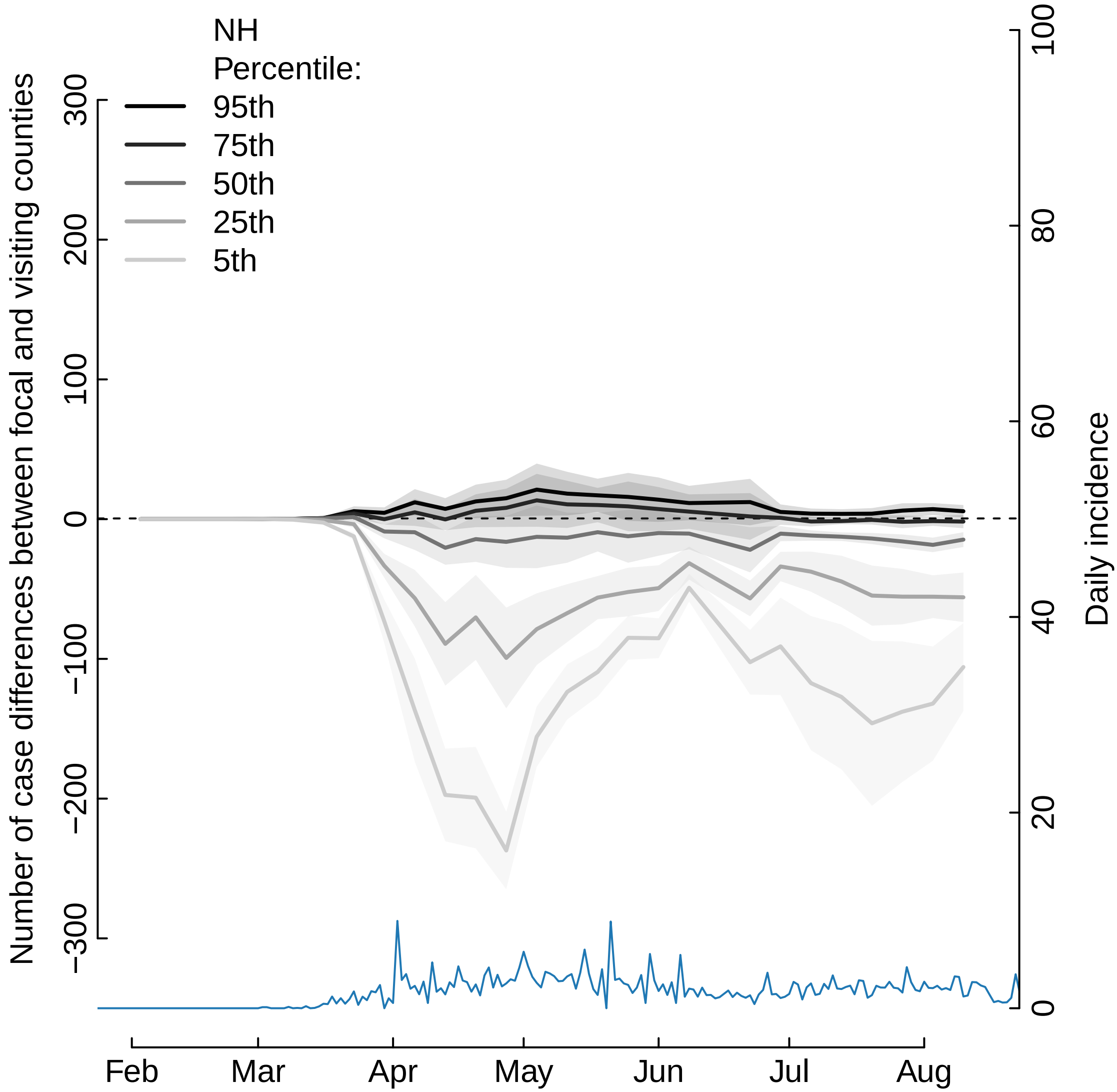
NH
Percentile:
95th
75th
50th
25th
5th
Number of case differences between focal and visiting counties
300
200
100
0
−100
−200
−300
Daily incidence
100
80
60
40
20
0
Feb
Mar
Apr
May
Jun
Jul
Aug

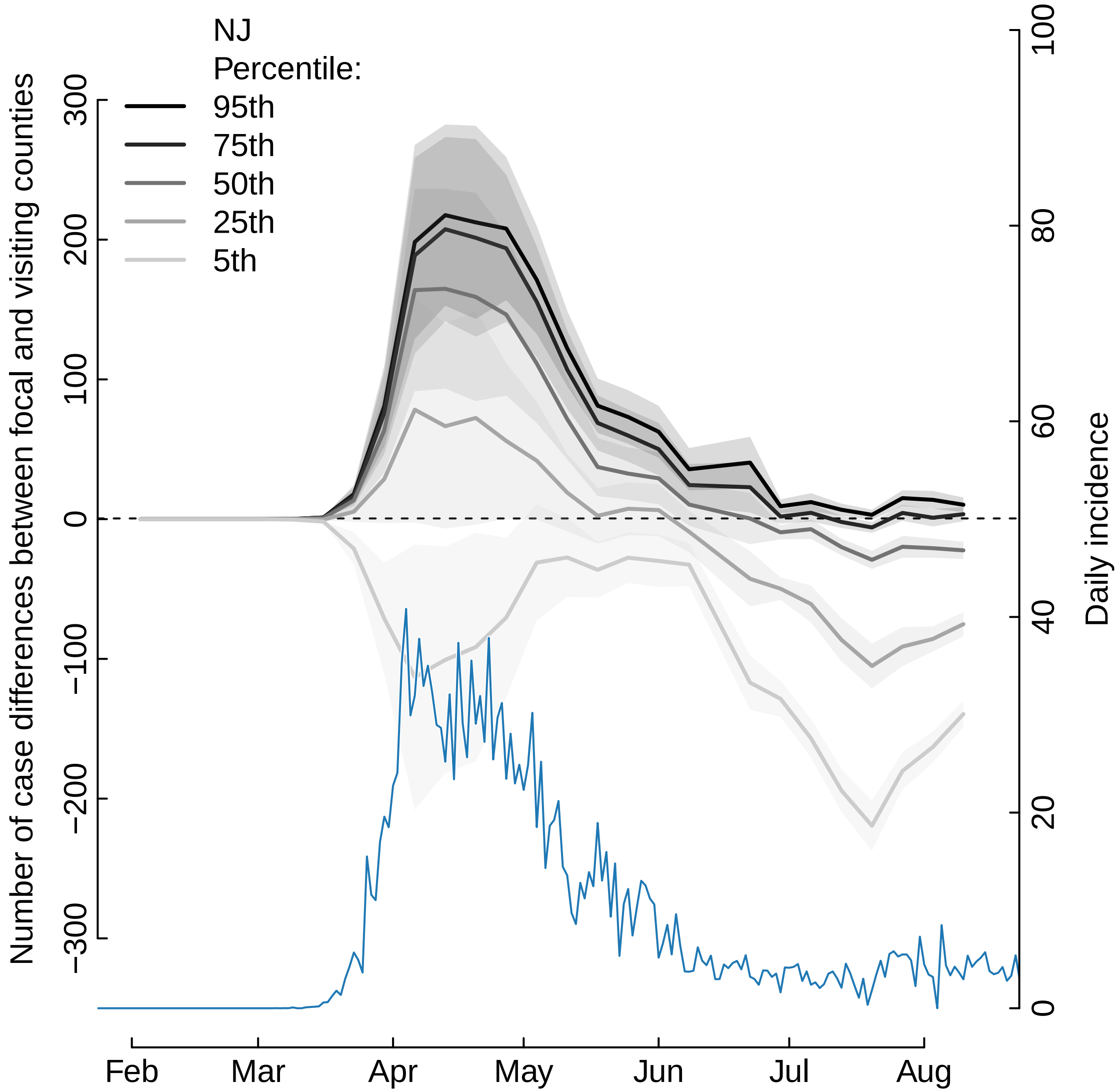

NJ
Percentile:
95th
75th
50th
25th
5th
Number of case differences between focal and visiting counties
300
200
100
0
−100
−200
−300
Daily incidence
100
80
60
40
20
0
Feb
Mar
Apr
May
Jun
Jul
Aug

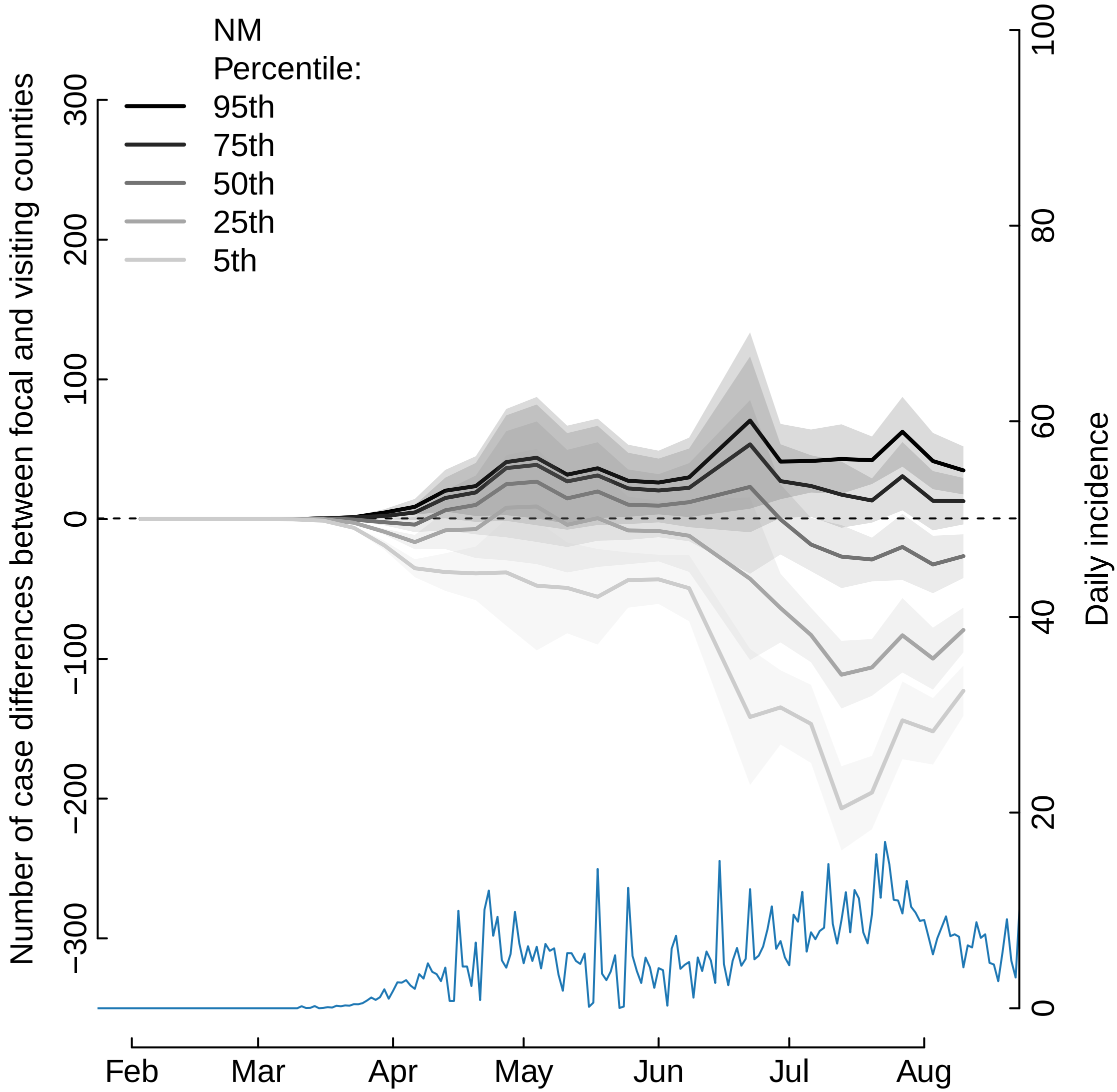
NM
Percentile:
95th
75th
50th
25th
5th
Number of case differences between focal and visiting counties
300
200
100
0
−100
−200
−300
Daily incidence
100
80
60
40
20
0
Feb
Mar
Apr
May
Jun
Jul
Aug

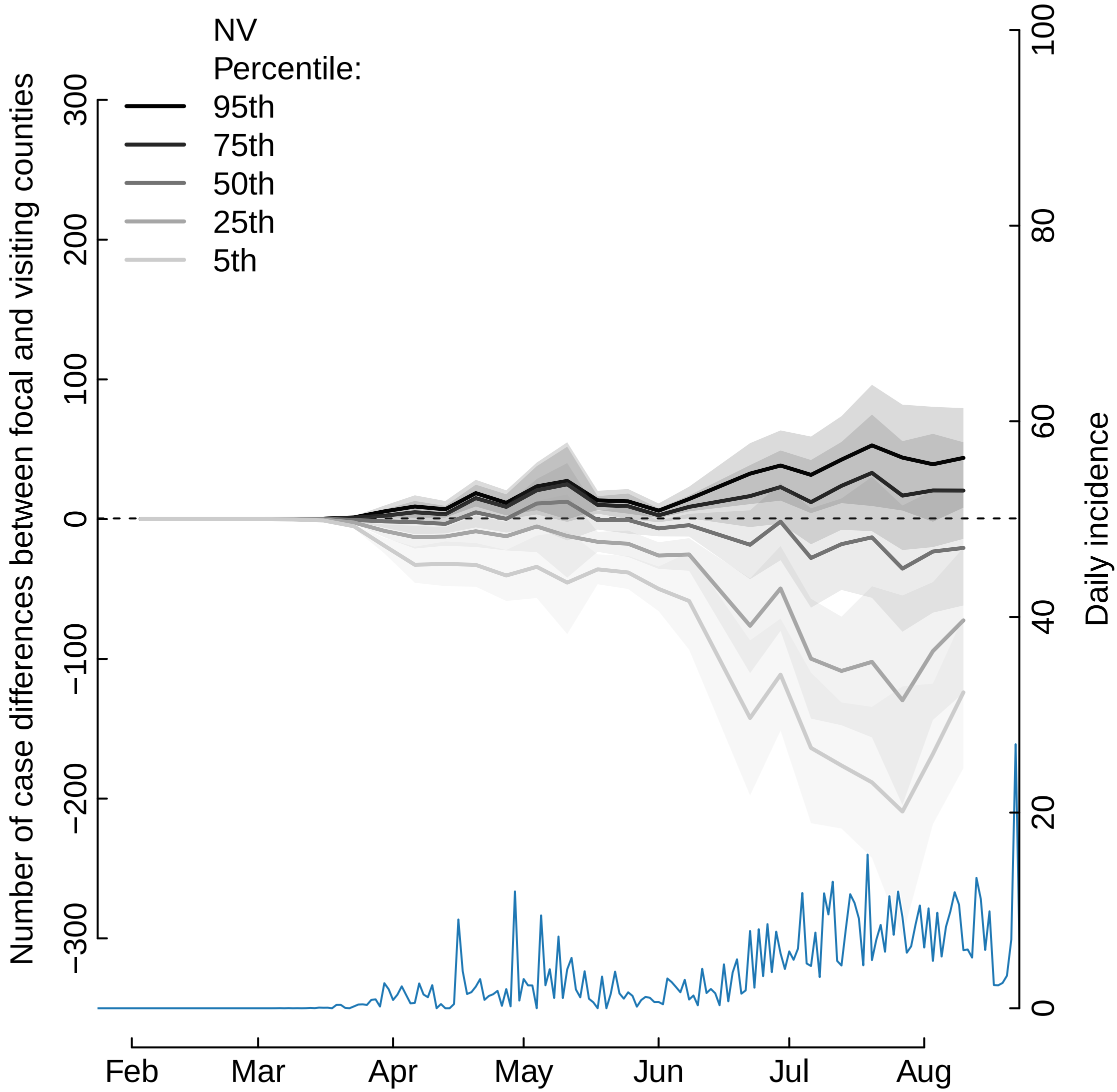
NV
Percentile:
95th
75th
50th
25th
5th
Number of case differences between focal and visiting counties
300
200
100
0
−100
−200
−300
Daily incidence
100
80
60
40
20
0
Feb
Mar
Apr
May
Jun
Jul
Aug

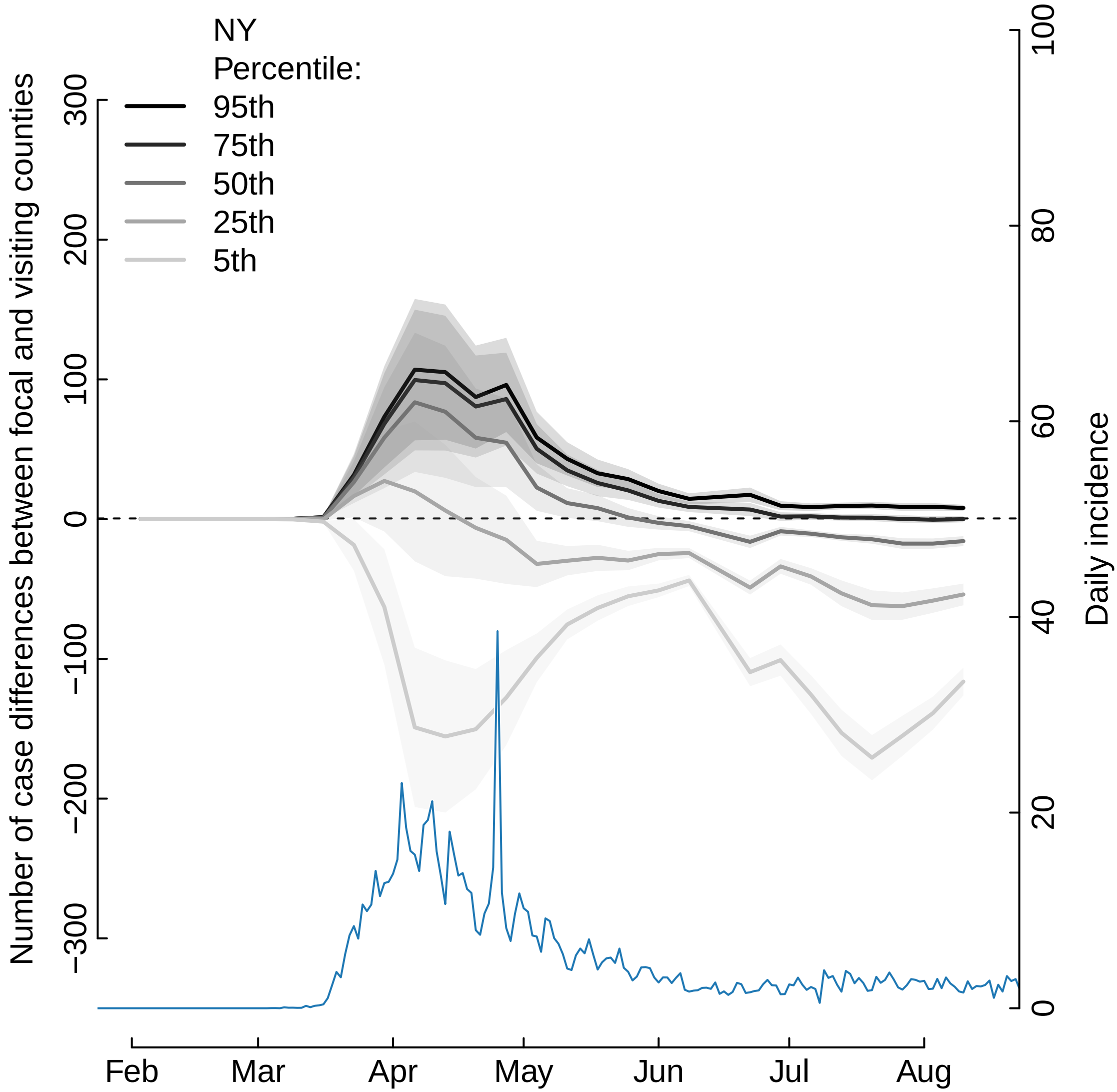

NY
Percentile:
95th
75th
50th
25th
5th
Number of case differences between focal and visiting counties
300
200
100
0
−100
−200
−300
Daily incidence
100
80
60
40
20
0
Feb
Mar
Apr
May
Jun
Jul
Aug

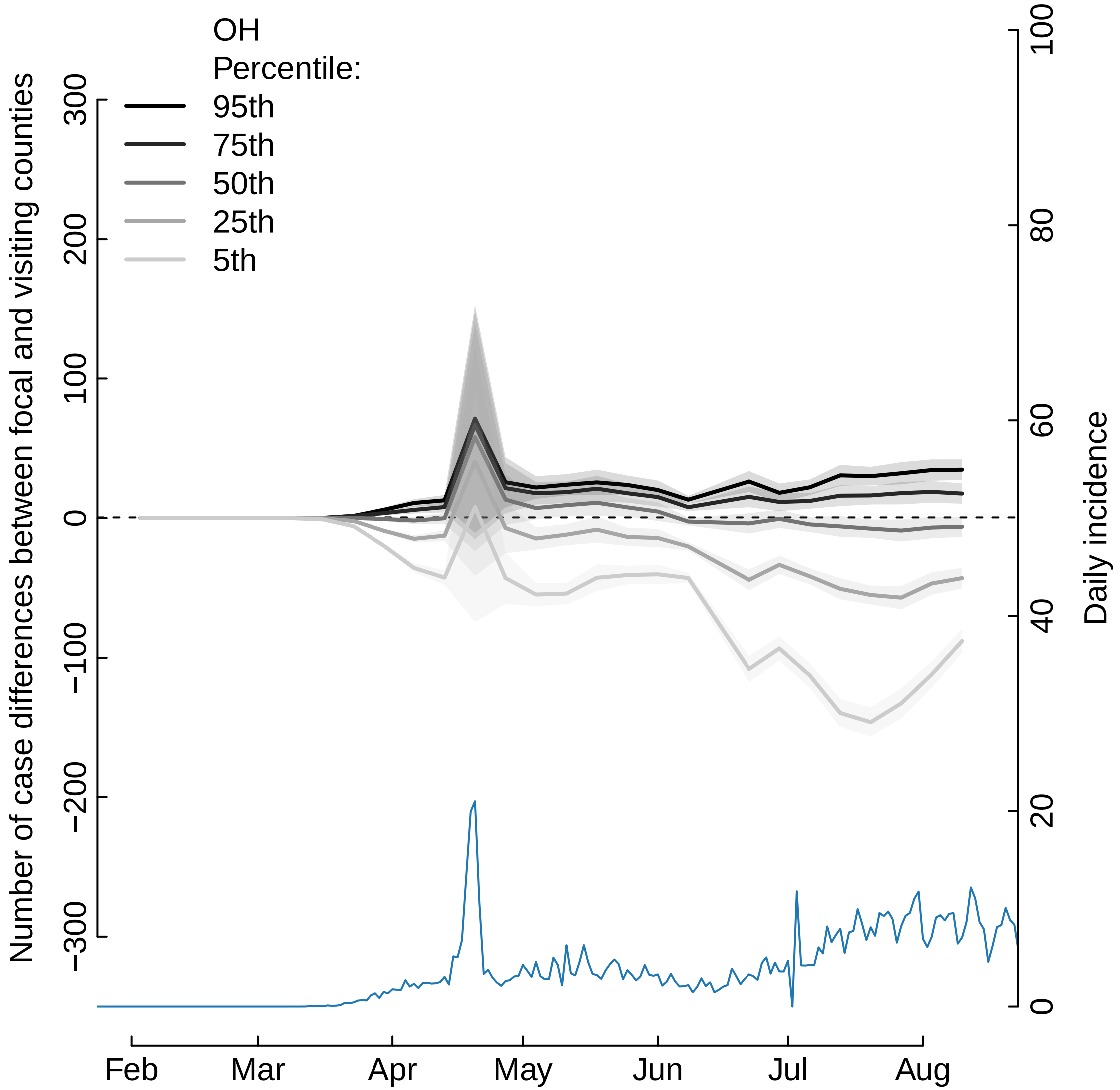
OH
Percentile:
95th
75th
50th
25th
5th
Number of case differences between focal and visiting counties
300
200
100
0
−100
−200
−300
Daily incidence
100
80
60
40
20
0
Feb
Mar
Apr
May
Jun
Jul
Aug

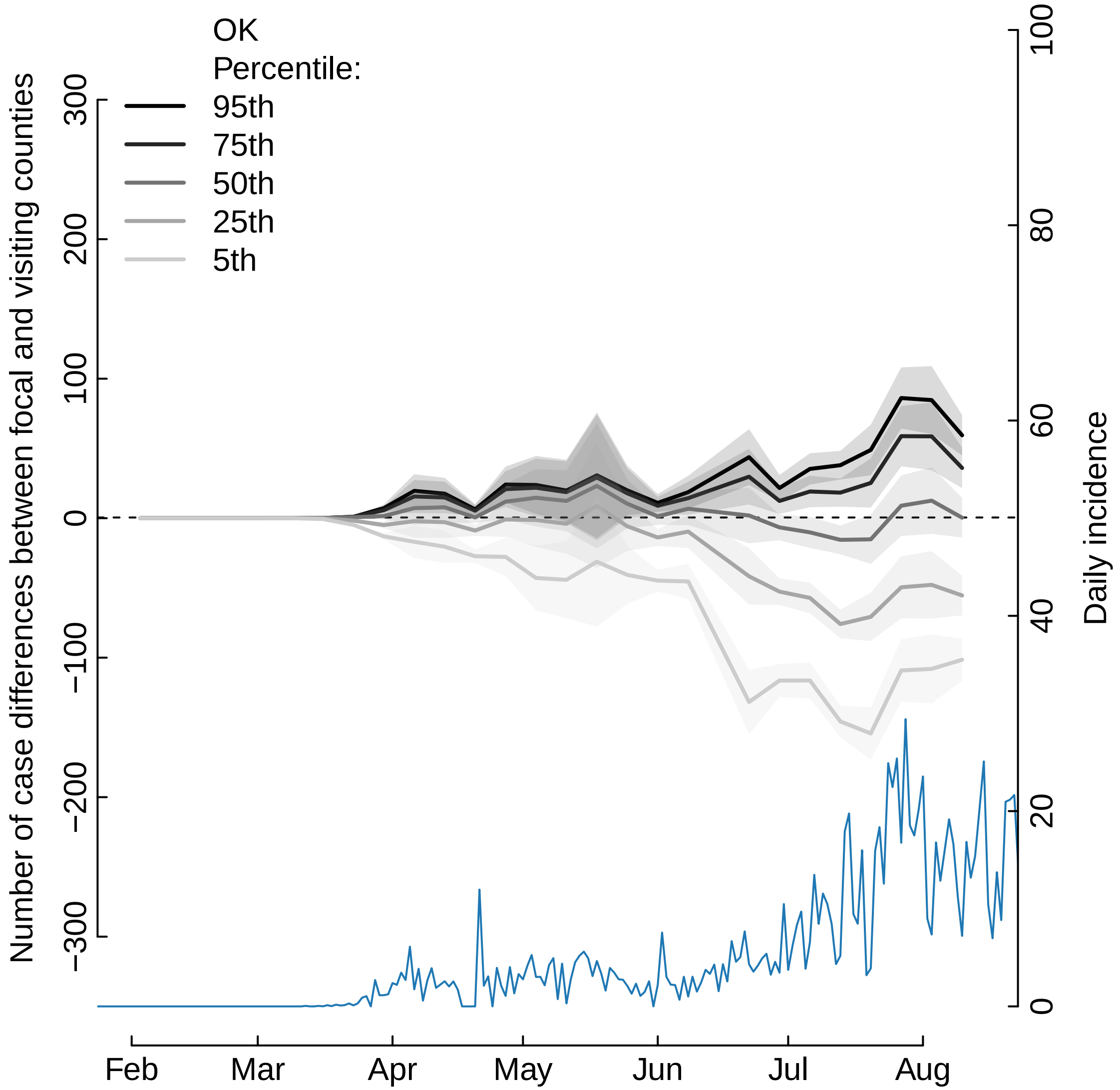

OK
Percentile:
95th
75th
50th
25th
5th
Number of case differences between focal and visiting counties
Daily incidence
300
200
100
0
−100
−200
−300
100
80
60
40
20
0
Feb
Mar
Apr
May
Jun
Jul
Aug

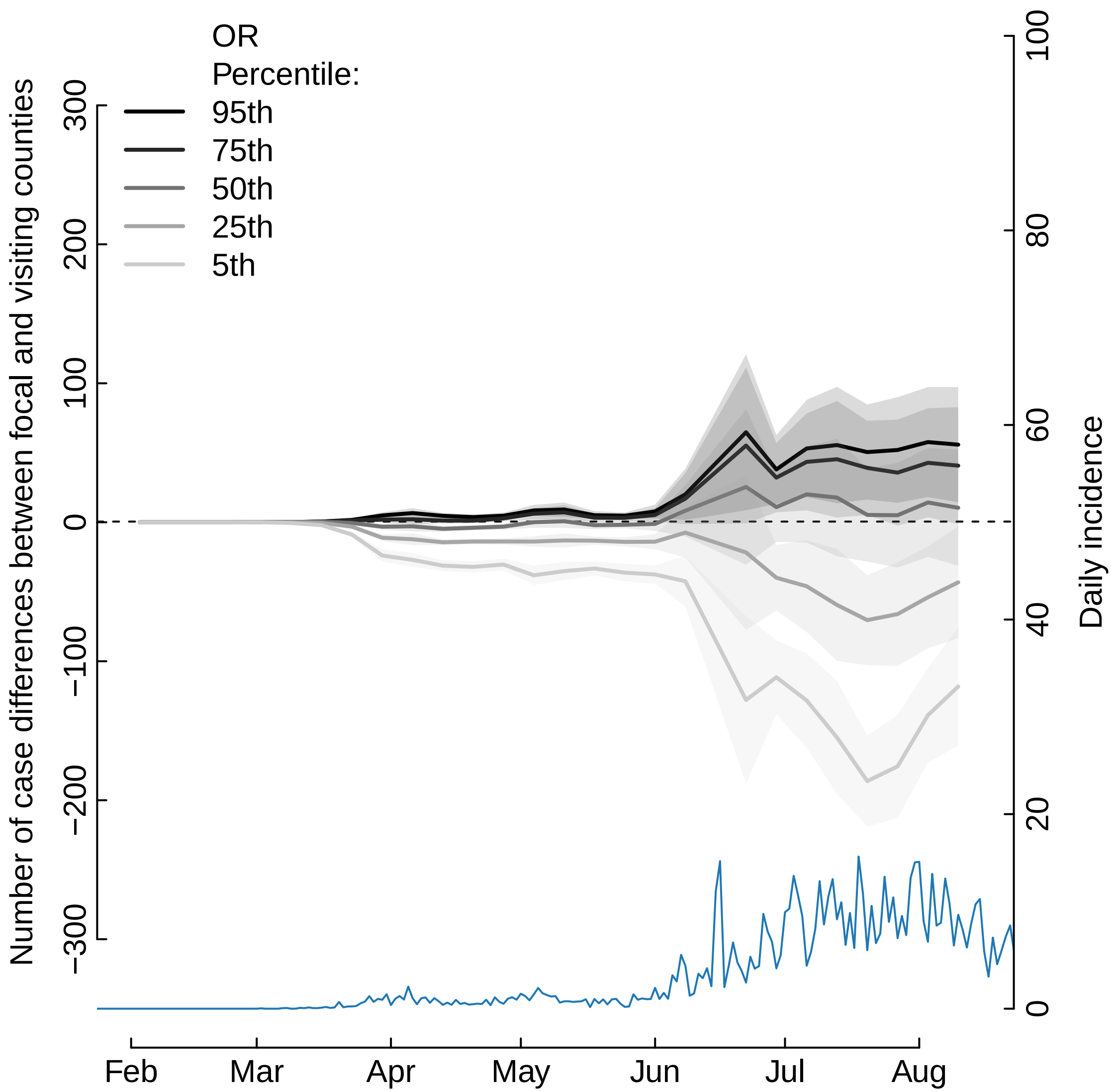
OR
Percentile:
95th
75th
50th
25th
5th
Number of case differences between focal and visiting counties
300
200
100
0
−100
−200
−300
Daily incidence
100
80
60
40
20
0
Feb
Mar
Apr
May
Jun
Jul
Aug

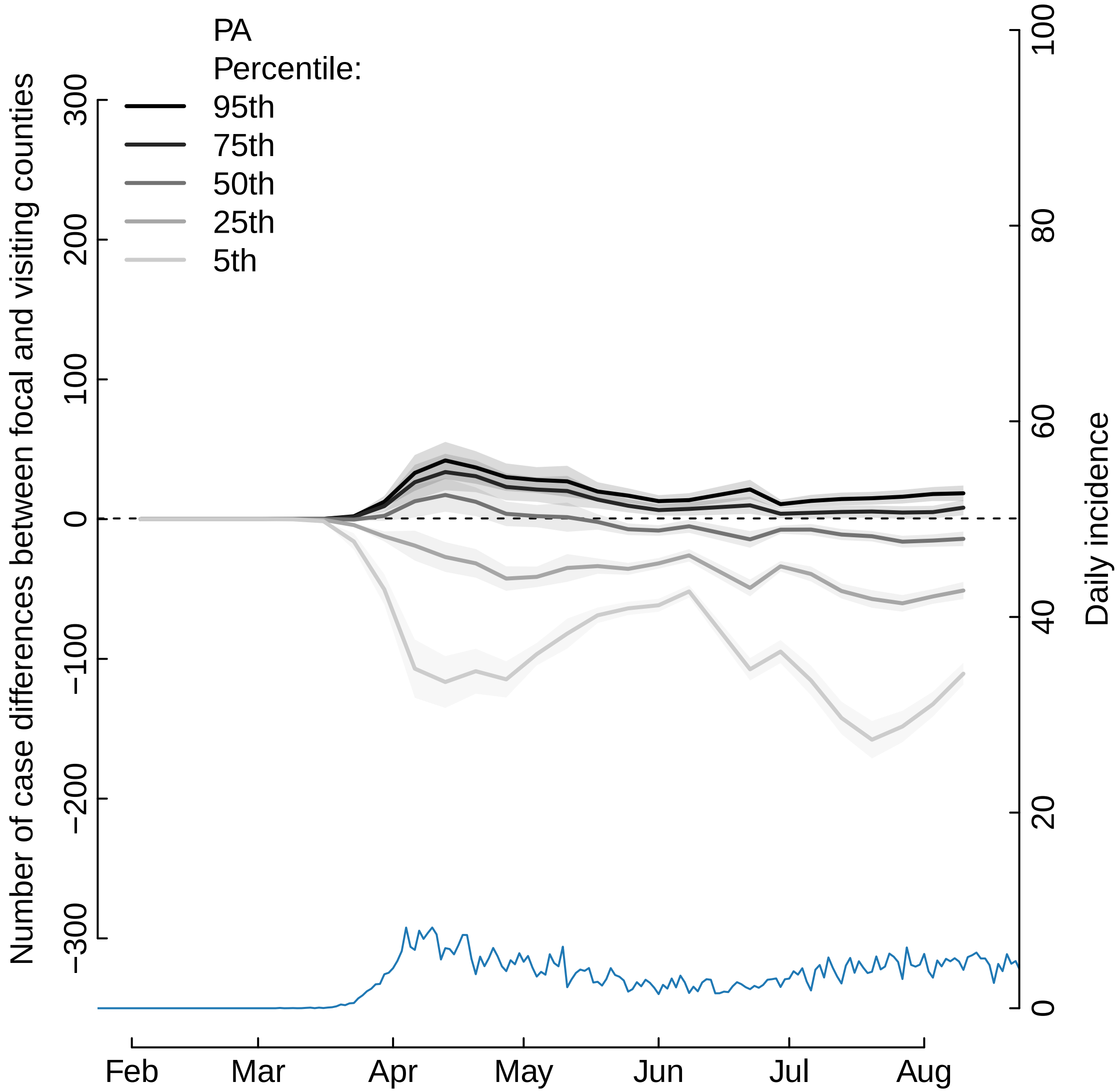

PA
Percentile:
95th
75th
50th
25th
5th
Number of case differences between focal and visiting counties
300
200
100
0
−100
−200
−300
Daily incidence
100
80
60
40
20
0
Feb
Mar
Apr
May
Jun
Jul
Aug

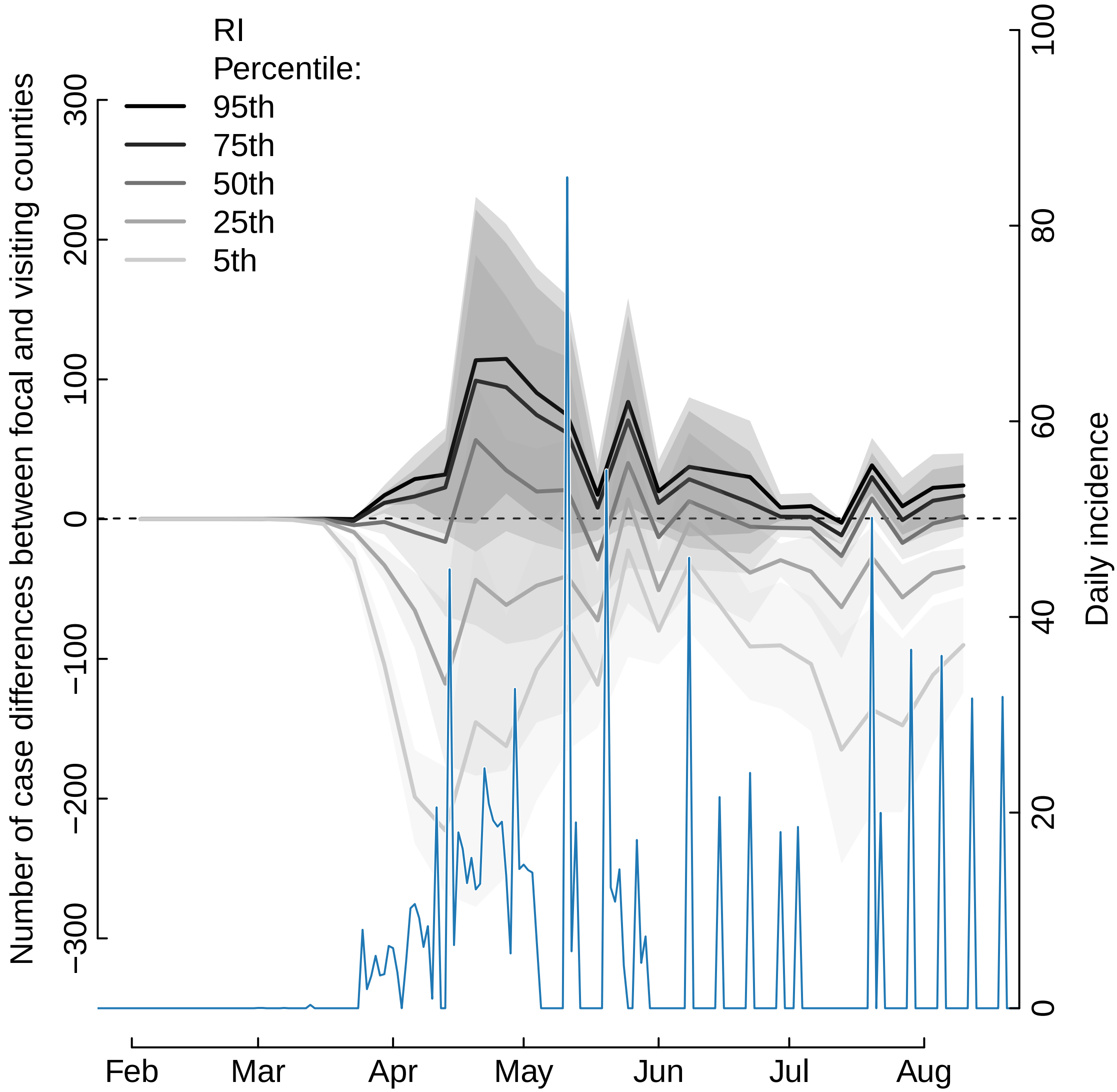

RI
Percentile:
95th
75th
50th
25th
5th
Number of case differences between focal and visiting counties
300
200
100
0
−100
−200
−300
Daily incidence
100
80
60
40
20
0
Feb
Mar
Apr
May
Jun
Jul
Aug

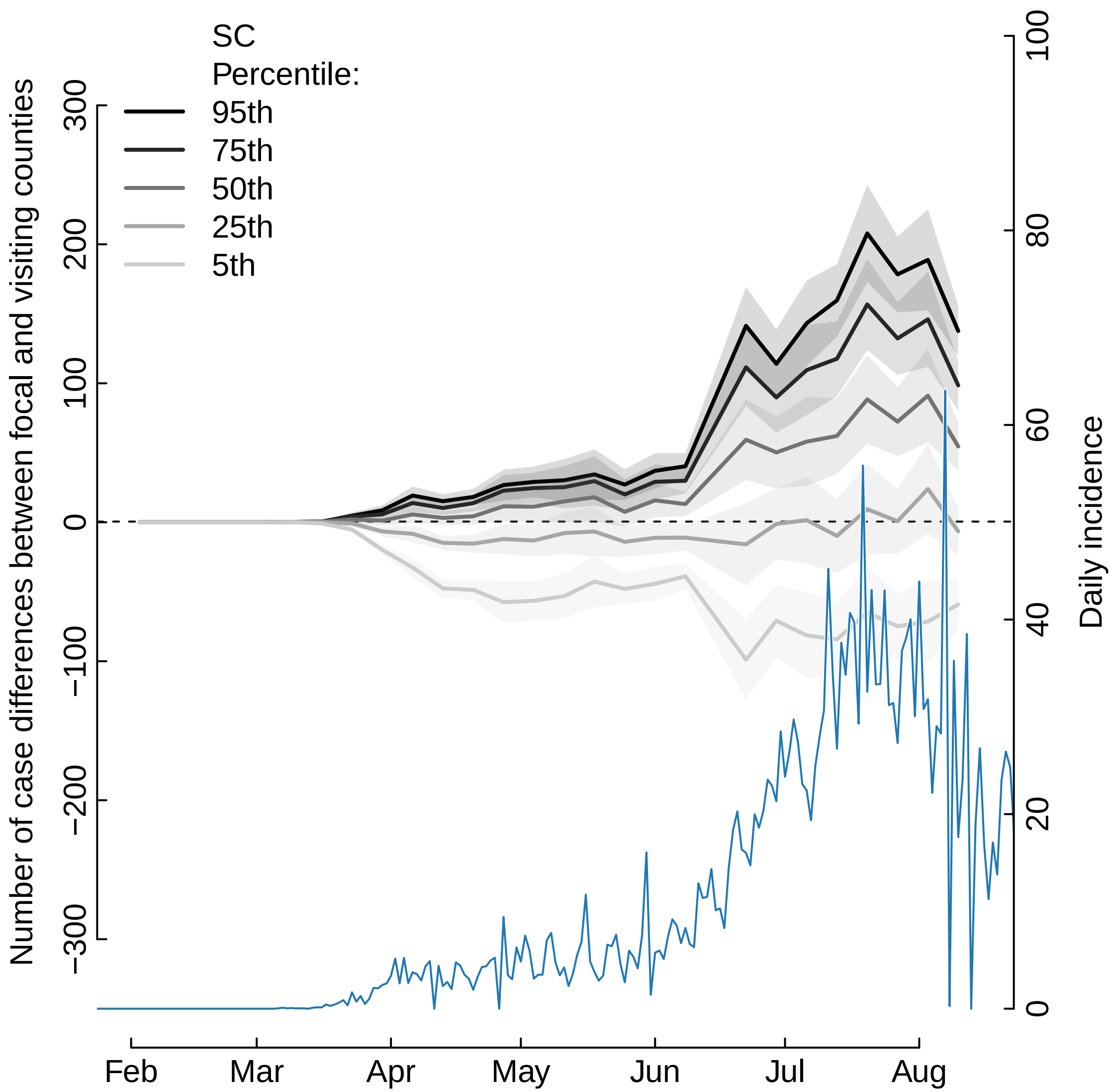

SC
Percentile:
95th
75th
50th
25th
5th
Number of case differences between focal and visiting counties
300
200
100
0
−100
−200
−300
Daily incidence
100
80
60
40
20
0
Feb
Mar
Apr
May
Jun
Jul
Aug

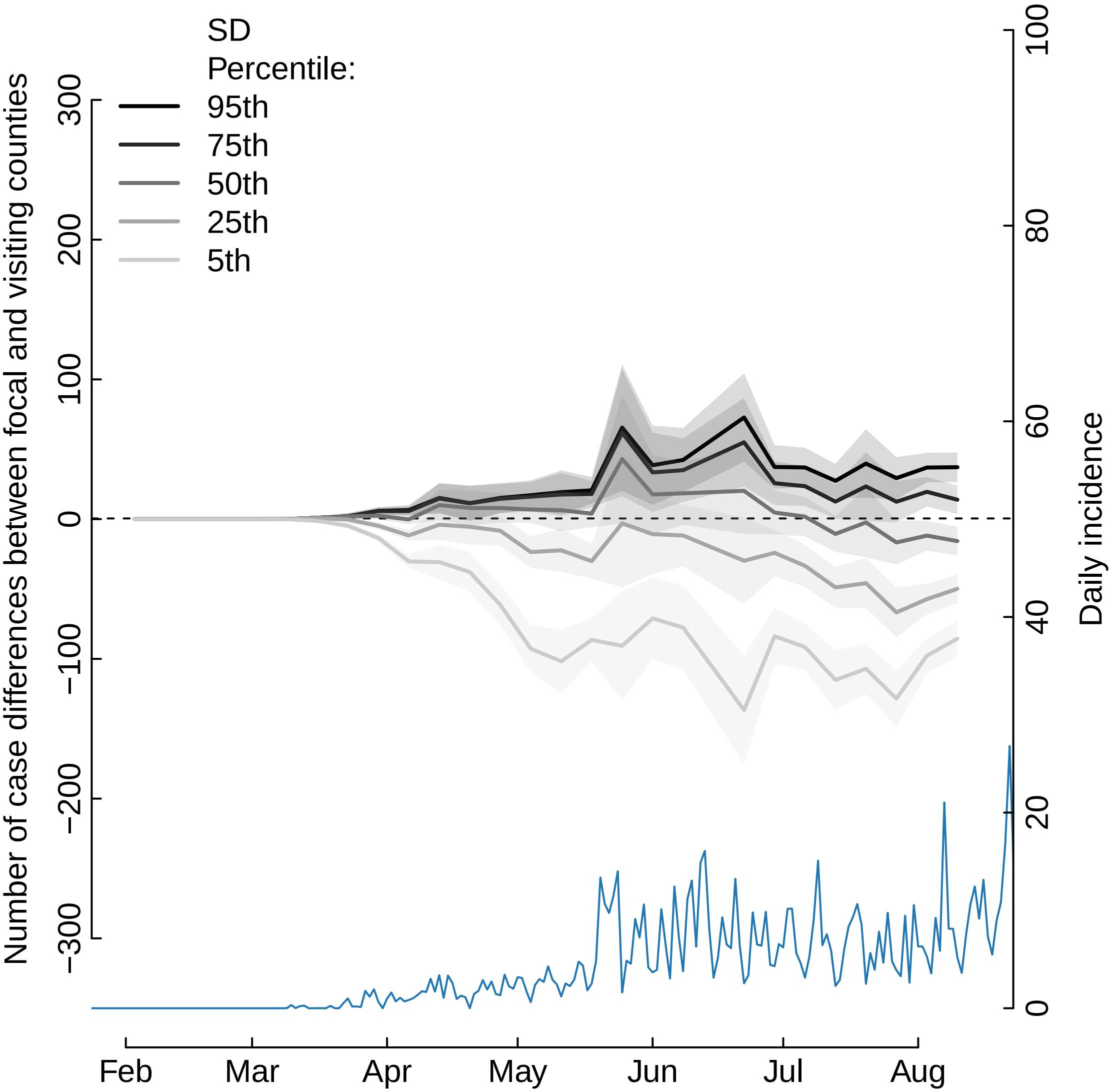

SD
Percentile:
95th
75th
50th
25th
5th
Number of case differences between focal and visiting counties
300
200
100
0
−100
−200
−300
Daily incidence
100
80
60
40
20
0
Feb
Mar
Apr
May
Jun
Jul
Aug

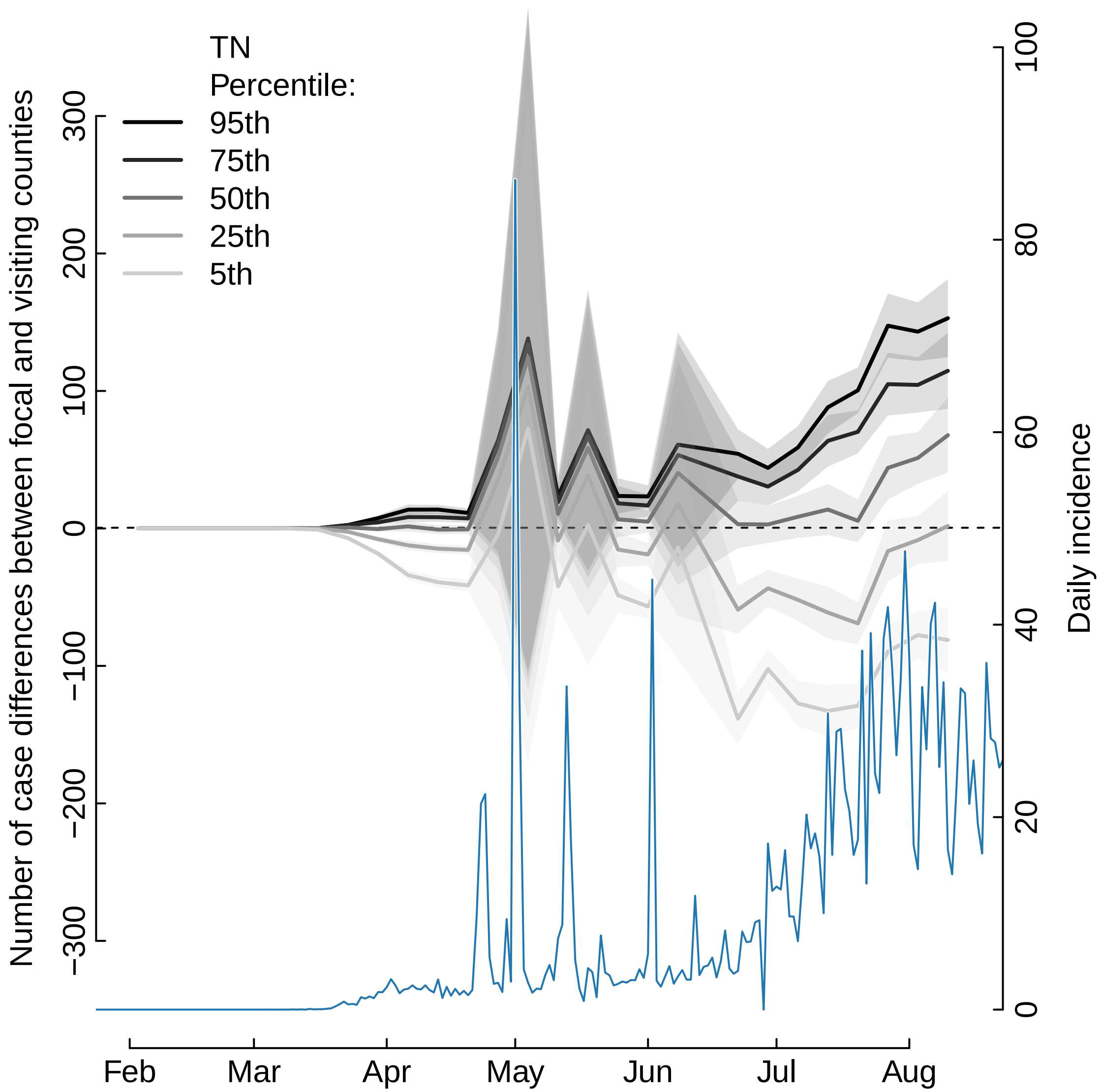

TN
Percentile:
95th
75th
50th
25th
5th
Number of case differences between focal and visiting counties
300
200
100
0
−100
−200
−300
Daily incidence
100
80
60
40
20
0
Feb
Mar
Apr
May
Jun
Jul
Aug

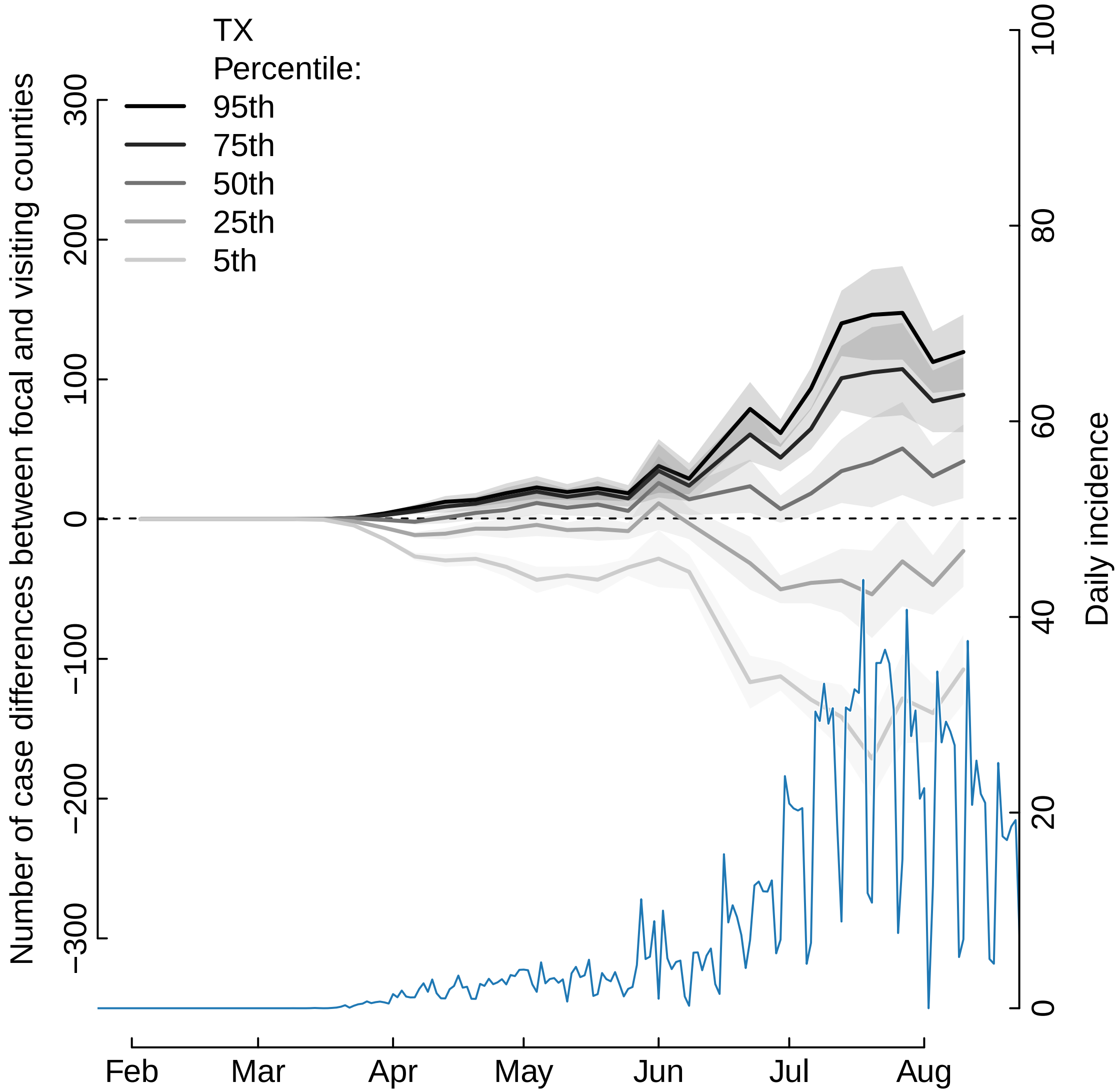
TX
Percentile:
95th
75th
50th
25th
5th
Number of case differences between focal and visiting counties
300
200
100
0
−100
−200
−300
Daily incidence
100
80
60
40
20
0
Feb
Mar
Apr
May
Jun
Jul
Aug

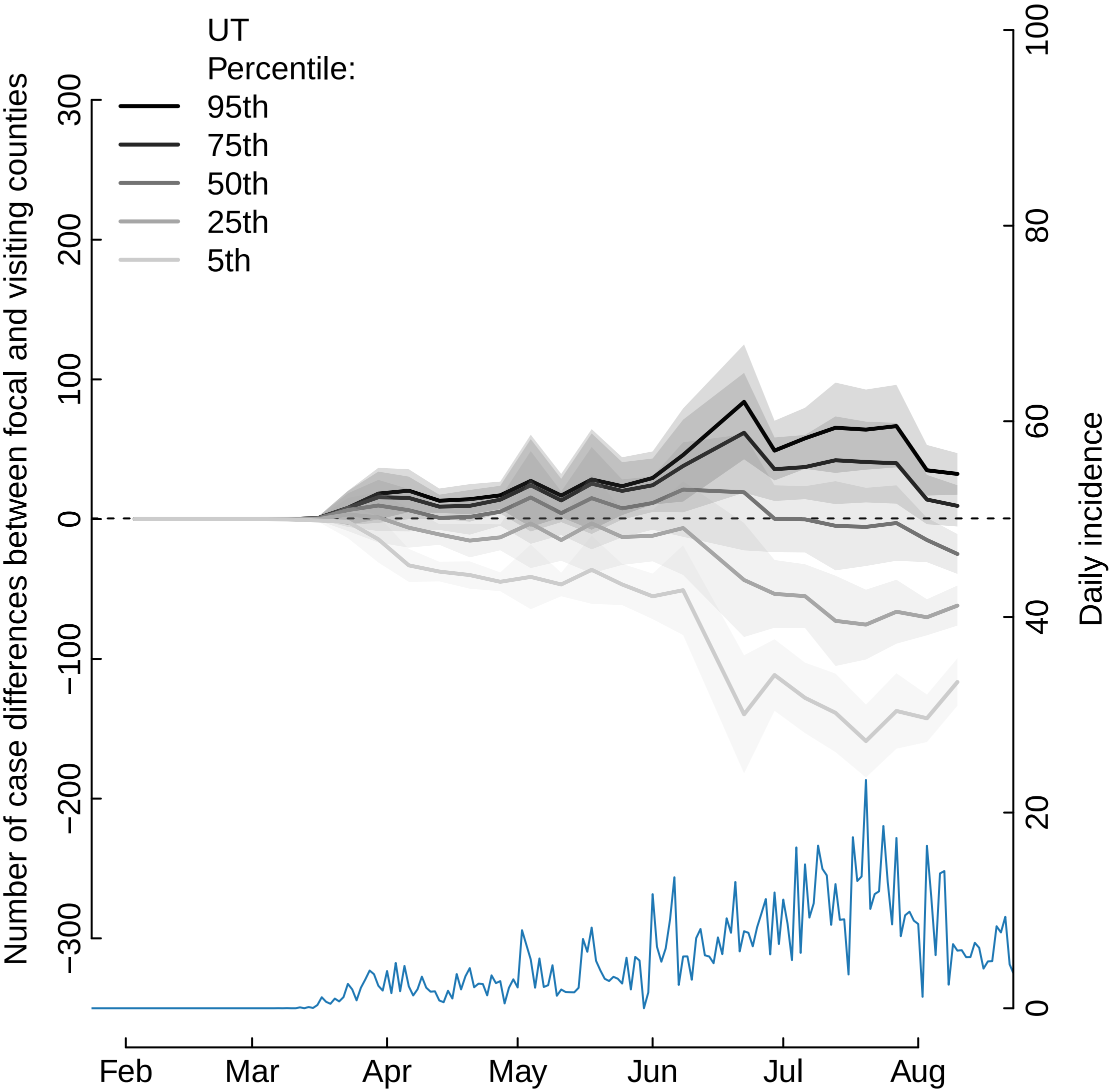

UT
Percentile:
95th
75th
50th
25th
5th
Number of case differences between focal and visiting counties
300
200
100
0
−100
−200
−300
Daily incidence
100
80
60
40
20
0
Feb
Mar
Apr
May
Jun
Jul
Aug

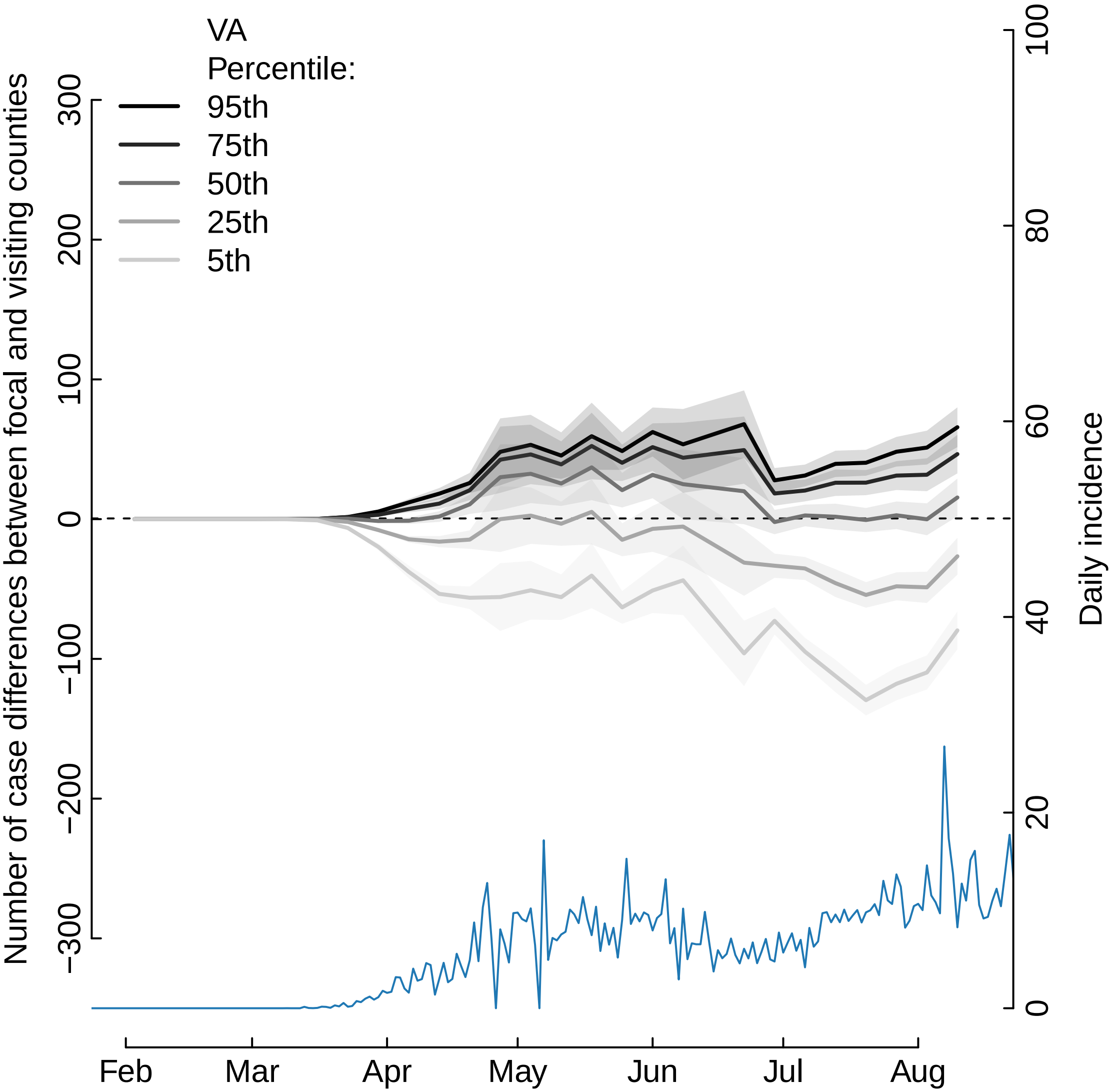
VA
Percentile:
95th
75th
50th
25th
5th
Number of case differences between focal and visiting counties
300
200
100
0
−100
−200
−300
Daily incidence
100
80
60
40
20
0
Feb
Mar
Apr
May
Jun
Jul
Aug

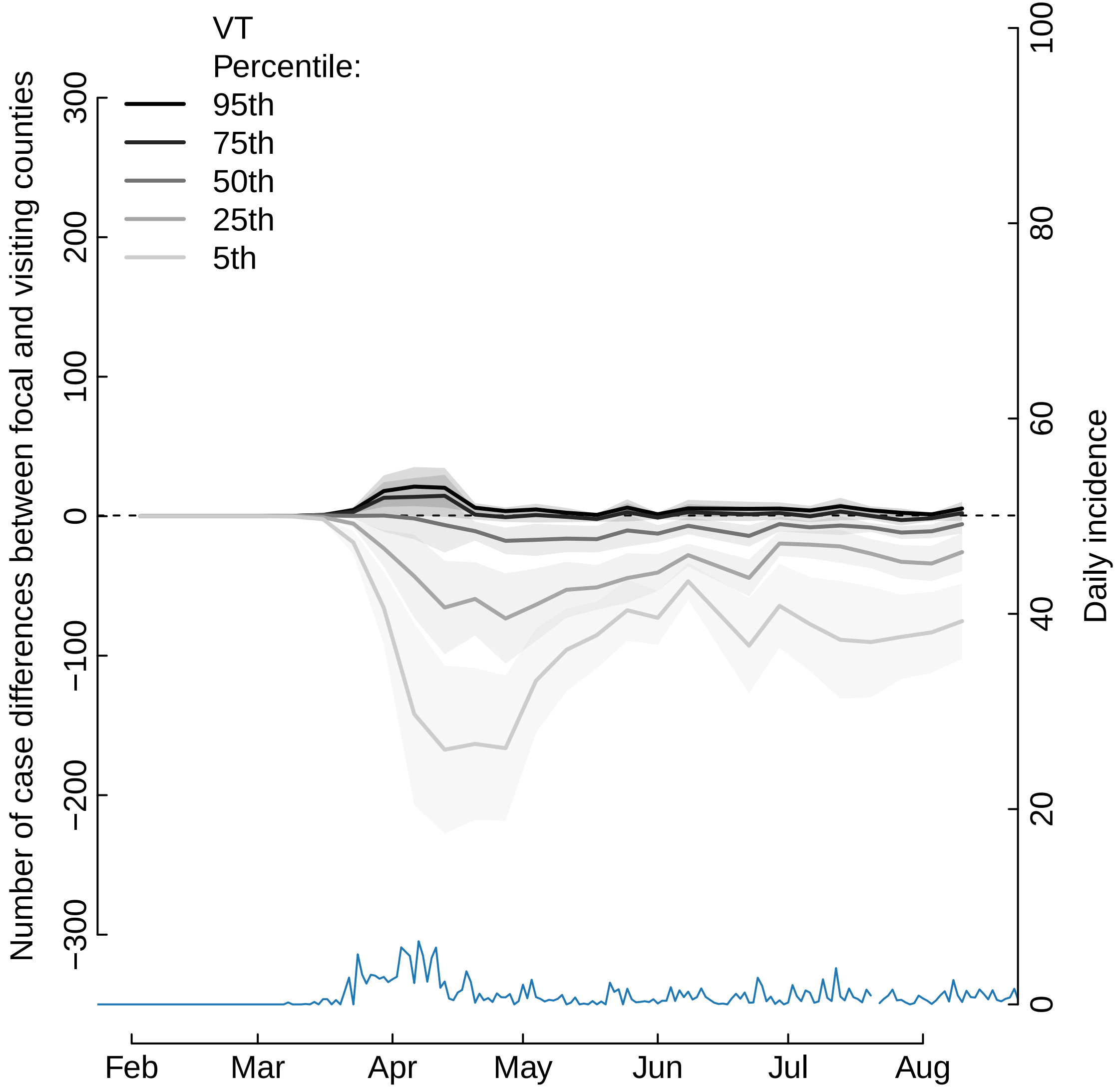

VT
Percentile:
95th
75th
50th
25th
5th
Number of case differences between focal and visiting counties
300
200
100
0
−100
−200
−300
Daily incidence
100
80
60
40
20
0
Feb
Mar
Apr
May
Jun
Jul
Aug

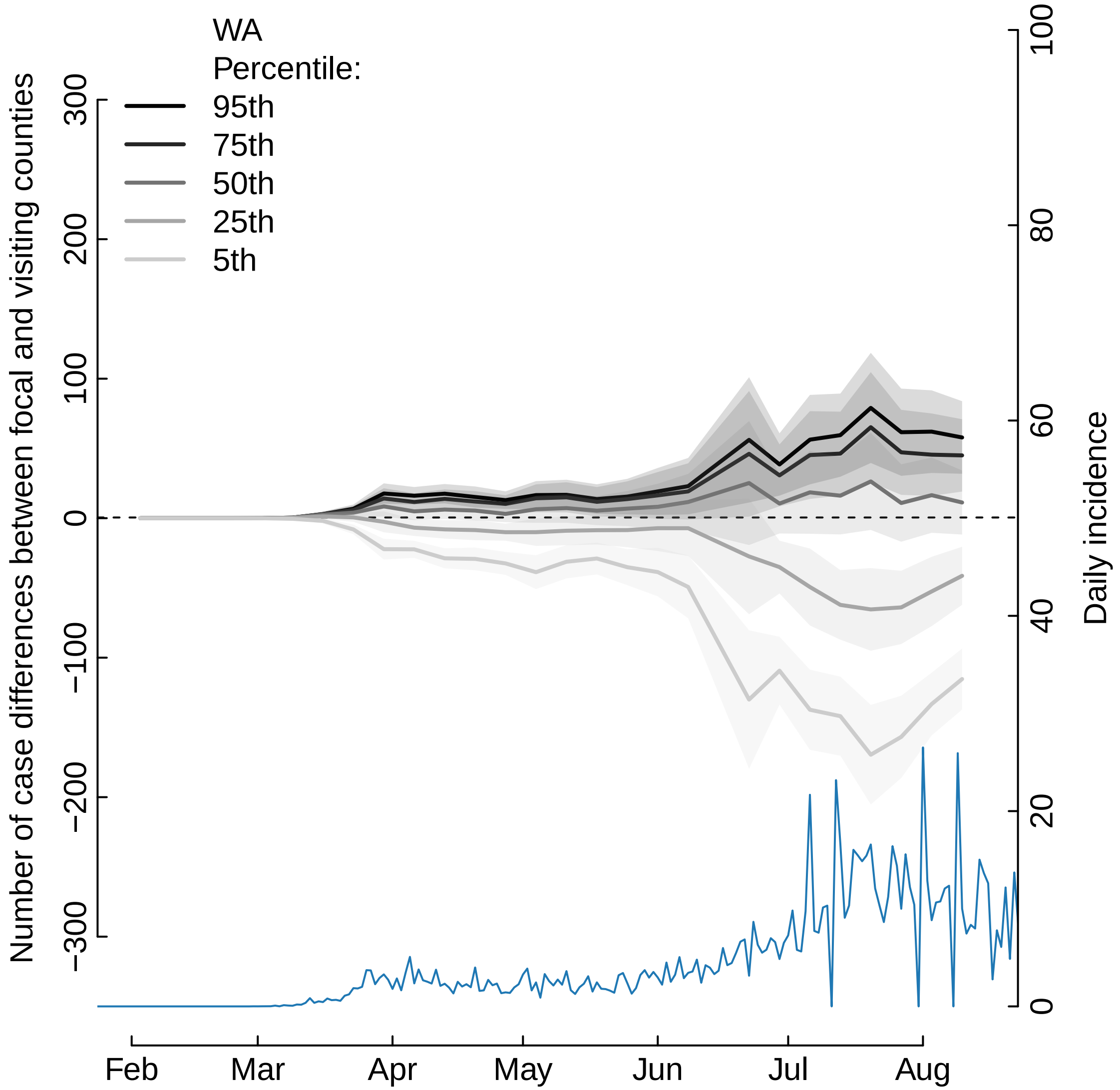

WA
Percentile:
95th
75th
50th
25th
5th
Number of case differences between focal and visiting counties
300
200
100
0
−100
−200
−300
Daily incidence
100
80
60
40
20
0
Feb
Mar
Apr
May
Jun
Jul
Aug

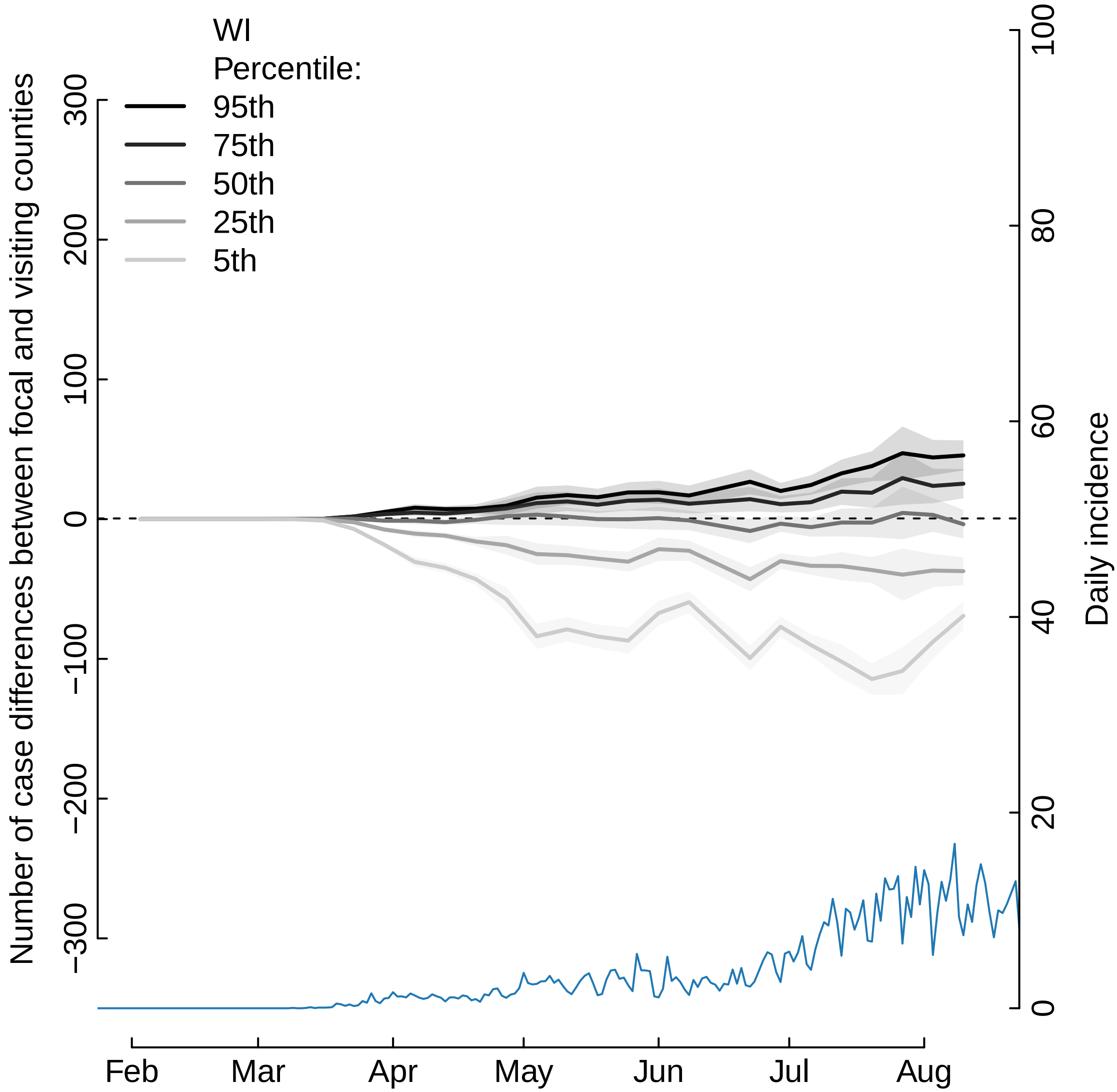
WI
Percentile:
95th
75th
50th
25th
5th
Number of case differences between focal and visiting counties
300
200
100
0
−100
−200
−300
Daily incidence
100
80
60
40
20
0
Feb
Mar
Apr
May
Jun
Jul
Aug

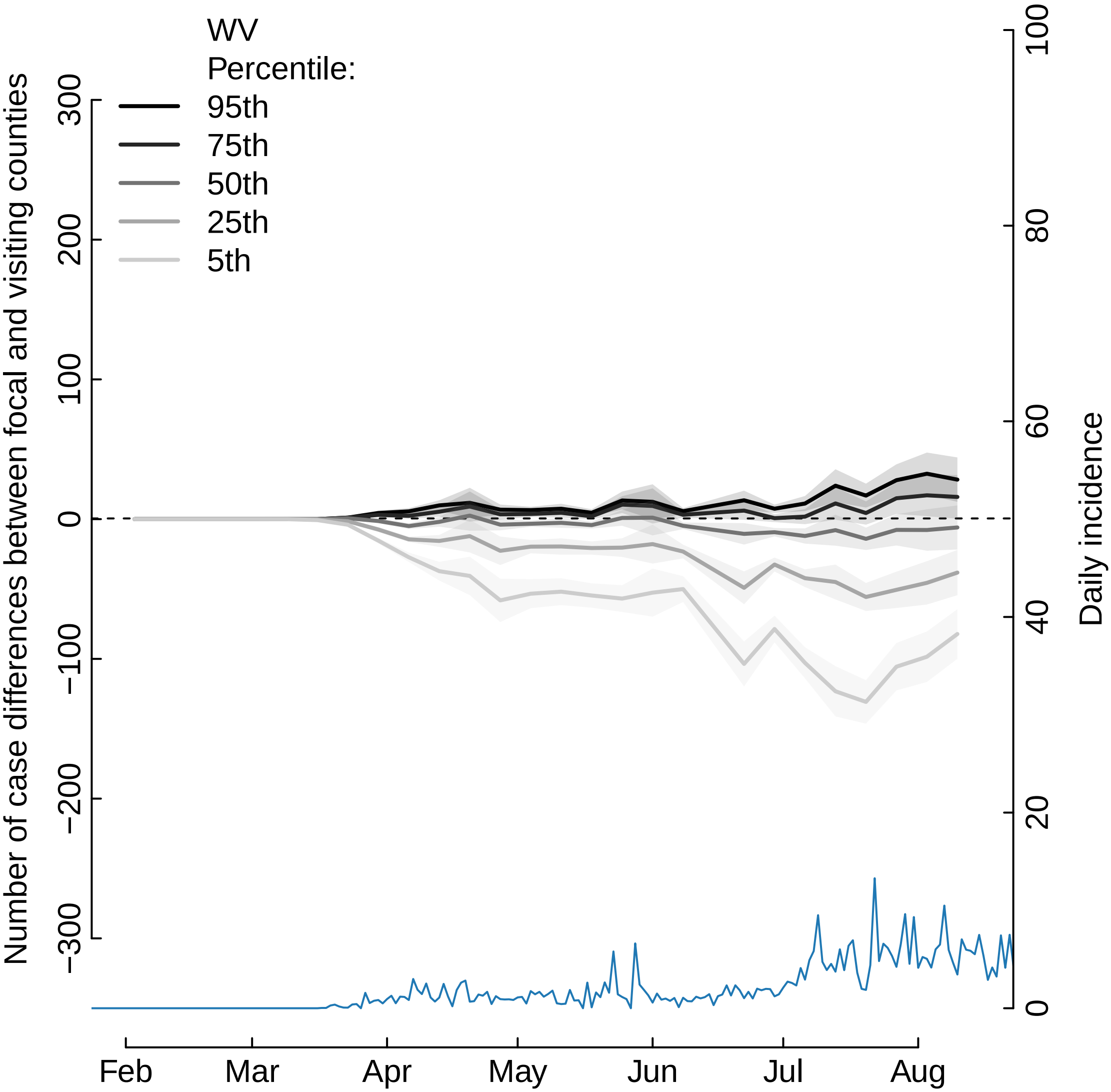

WV
Percentile:
95th
75th
50th
25th
5th
Number of case differences between focal and visiting counties
300
200
100
0
−100
−200
−300
Daily incidence
100
80
60
40
20
0
Feb
Mar
Apr
May
Jun
Jul
Aug

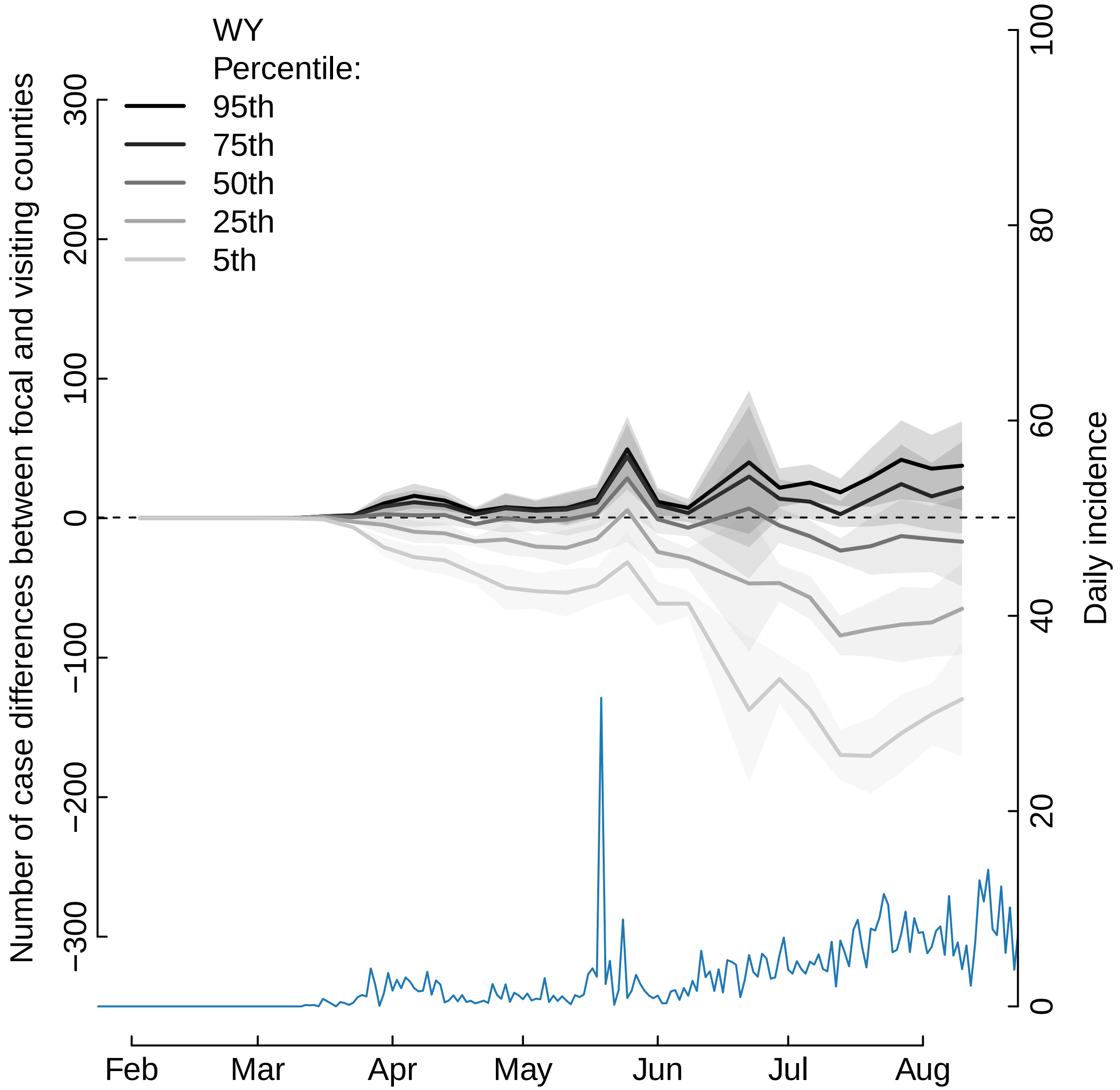

WY
Percentile:
95th
75th
50th
25th
5th
Number of case differences between focal and visiting counties
300
200
100
0
−100
−200
−300
Daily incidence
100
80
60
40
20
0
Feb
Mar
Apr
May
Jun
Jul
Aug
