## Supplementary material for "The unintended consequences of inconsistent pandemic control policies": Prop visiting more than focal and incidence

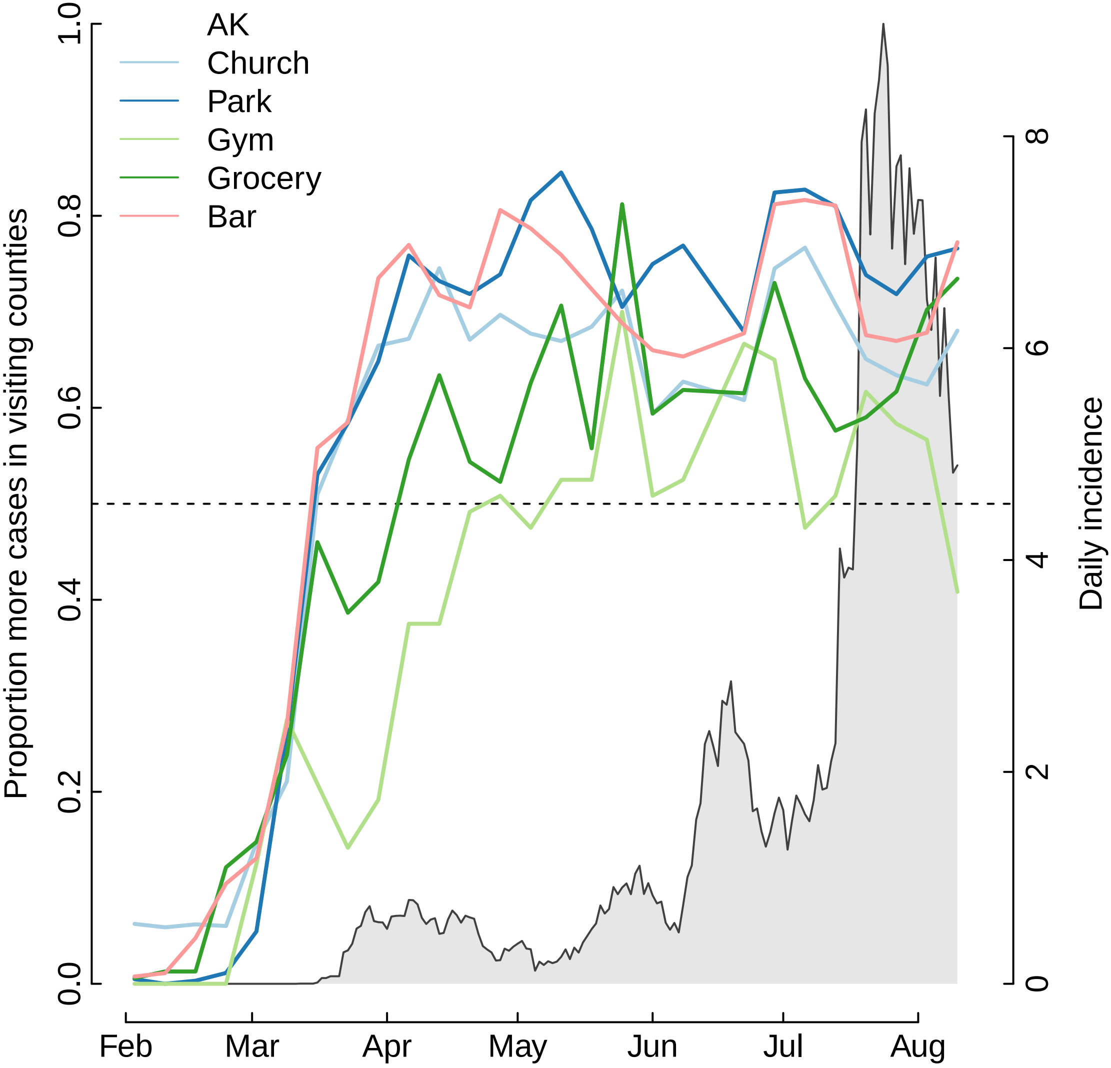

AK
Church
Park
Gym
Grocery
Bar
Proportion more cases in visiting counties
Daily incidence
0.0
0.2
0.4
0.6
0.8
1.0
0
2
4
6
8
Feb
Mar
Apr
May
Jun
Jul
Aug

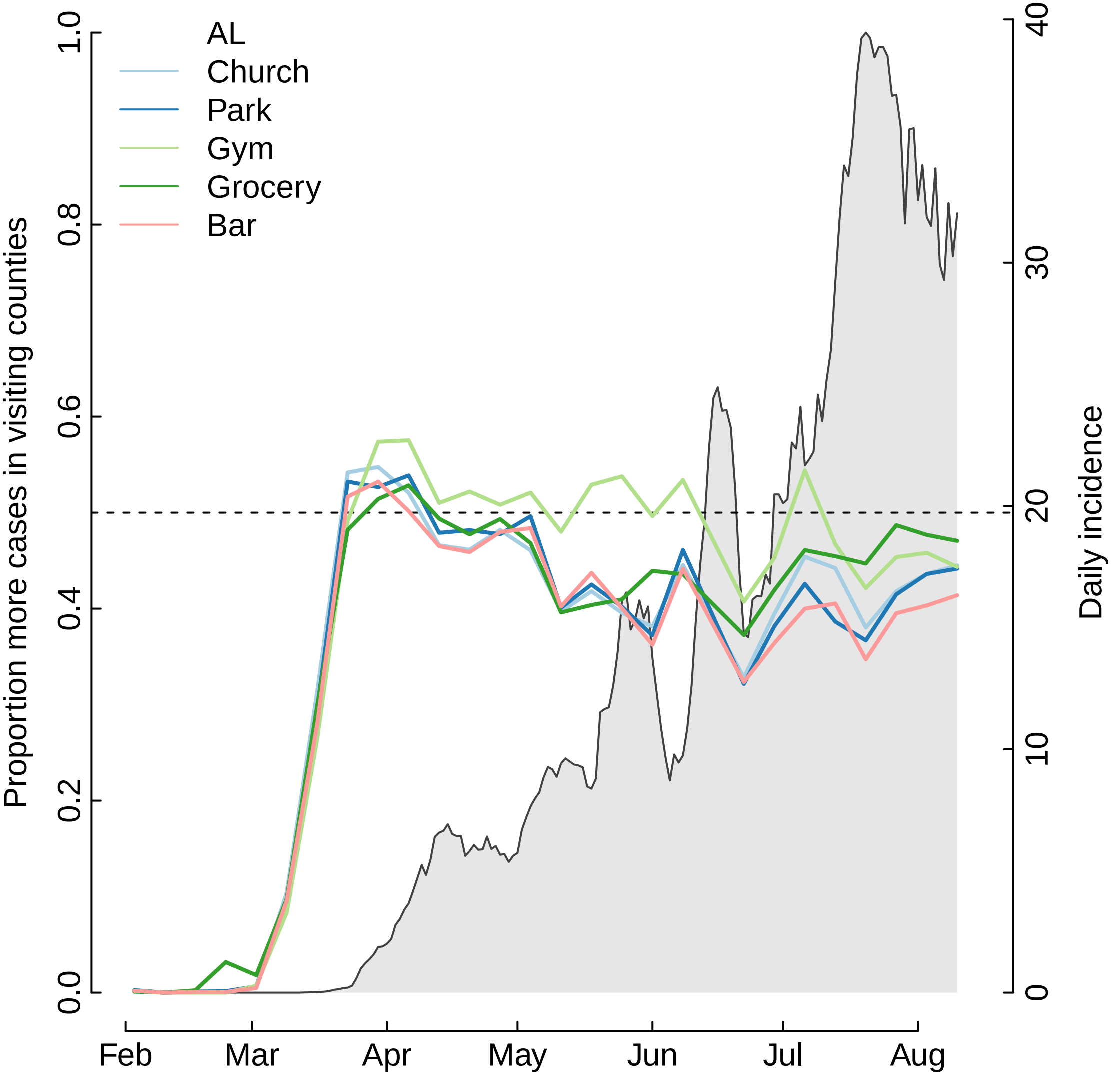

AL
Church
Park
Gym
Grocery
Bar
Proportion more cases in visiting counties
Daily incidence
0.0
0.2
0.4
0.6
0.8
1.0
0
10
20
30
40
Feb
Mar
Apr
May
Jun
Jul
Aug

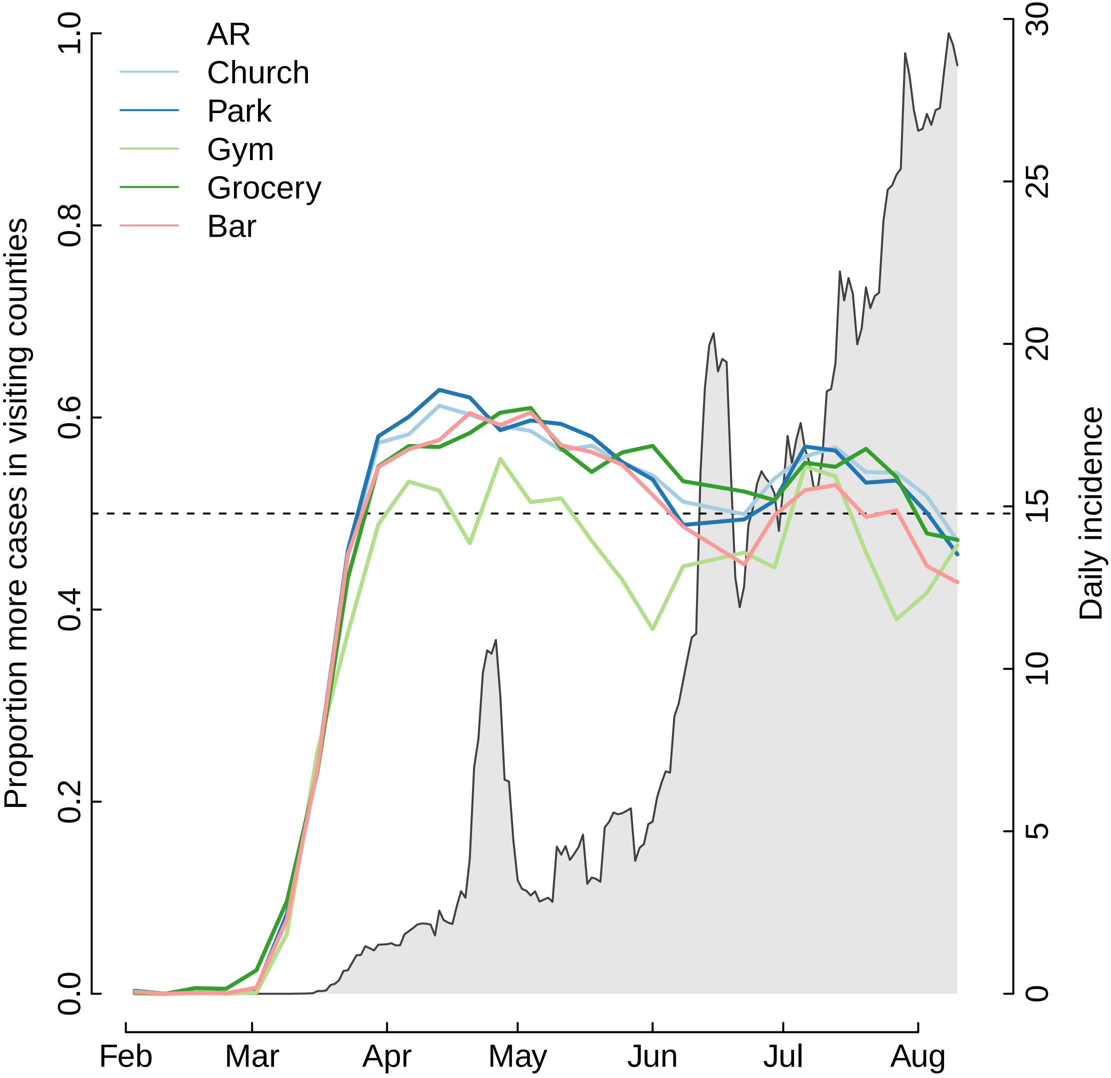

AR
Church
Park
Gym
Grocery
Bar
Proportion more cases in visiting counties
Daily incidence
0.0
0.2
0.4
0.6
0.8
1.0
0
5
10
15
20
25
30
Feb
Mar
Apr
May
Jun
Jul
Aug

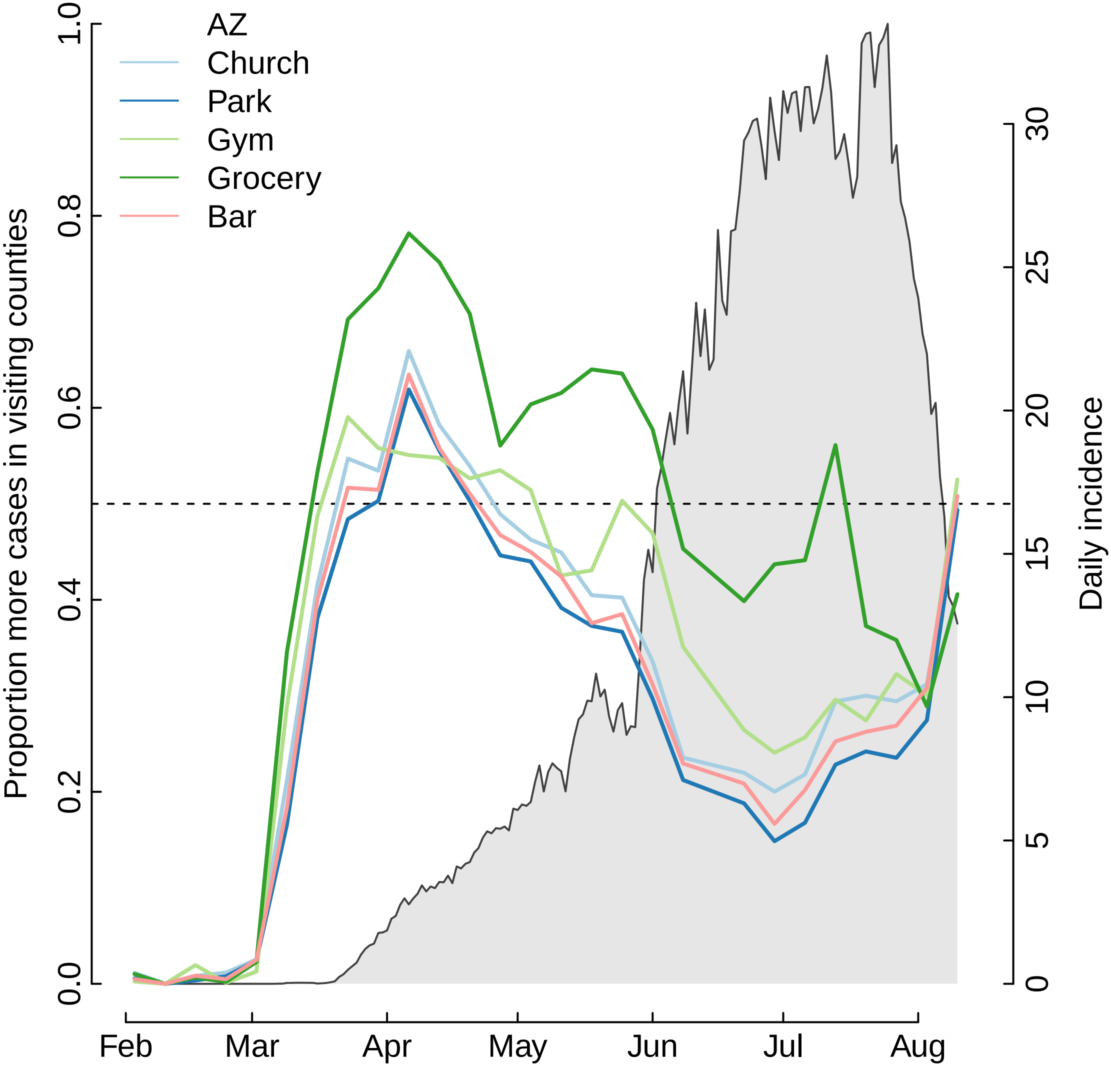

AZ
Church
Park
Gym
Grocery
Bar
Proportion more cases in visiting counties
Daily incidence
0.0
0.2
0.4
0.6
0.8
1.0
0
5
10
15
20
25
30
Feb
Mar
Apr
May
Jun
Jul
Aug

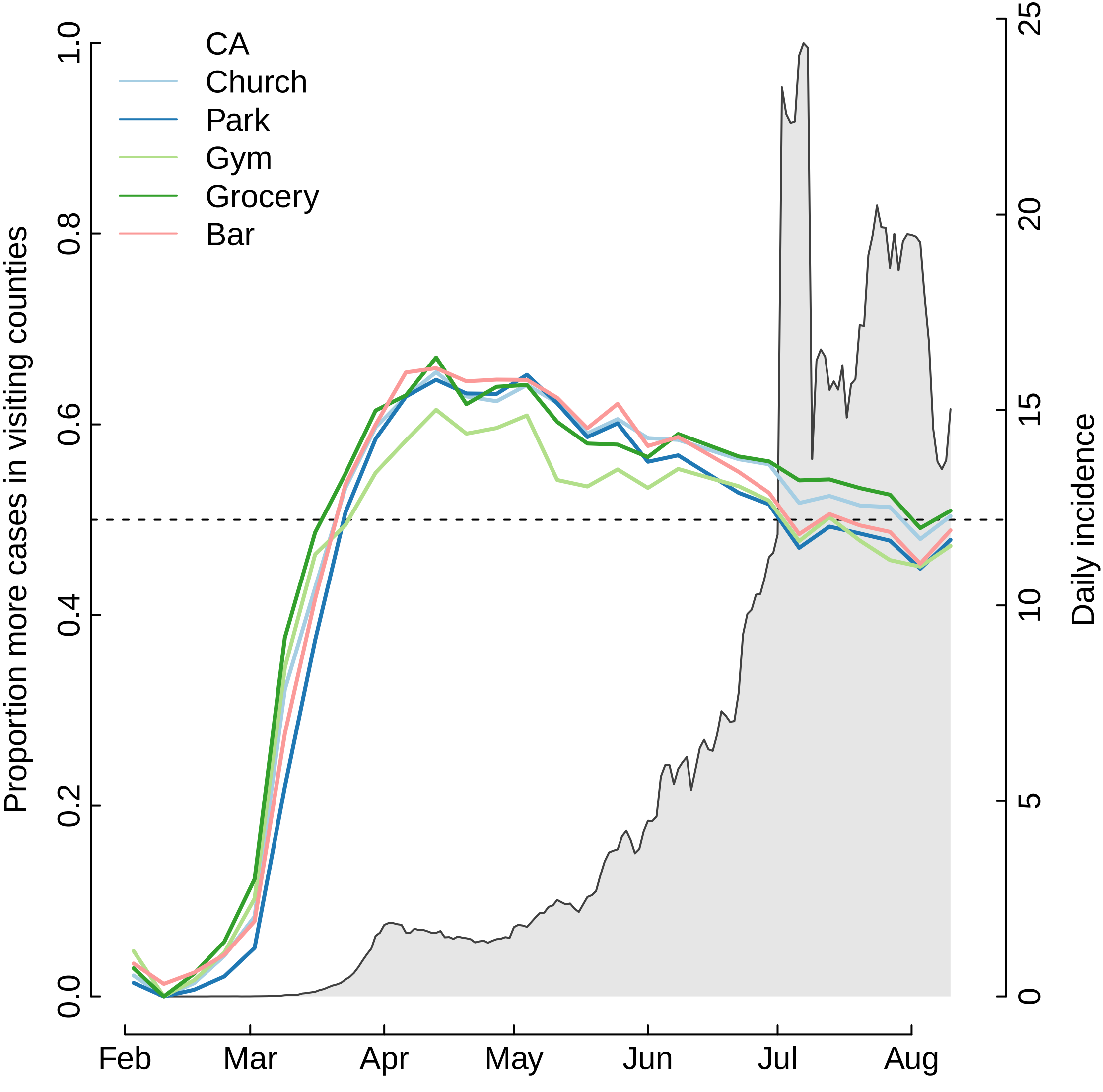
CA
Church
Park
Gym
Grocery
Bar
Proportion more cases in visiting counties
Daily incidence
0.0
0.2
0.4
0.6
0.8
1.0
0
5
10
15
20
25
Feb
Mar
Apr
May
Jun
Jul
Aug

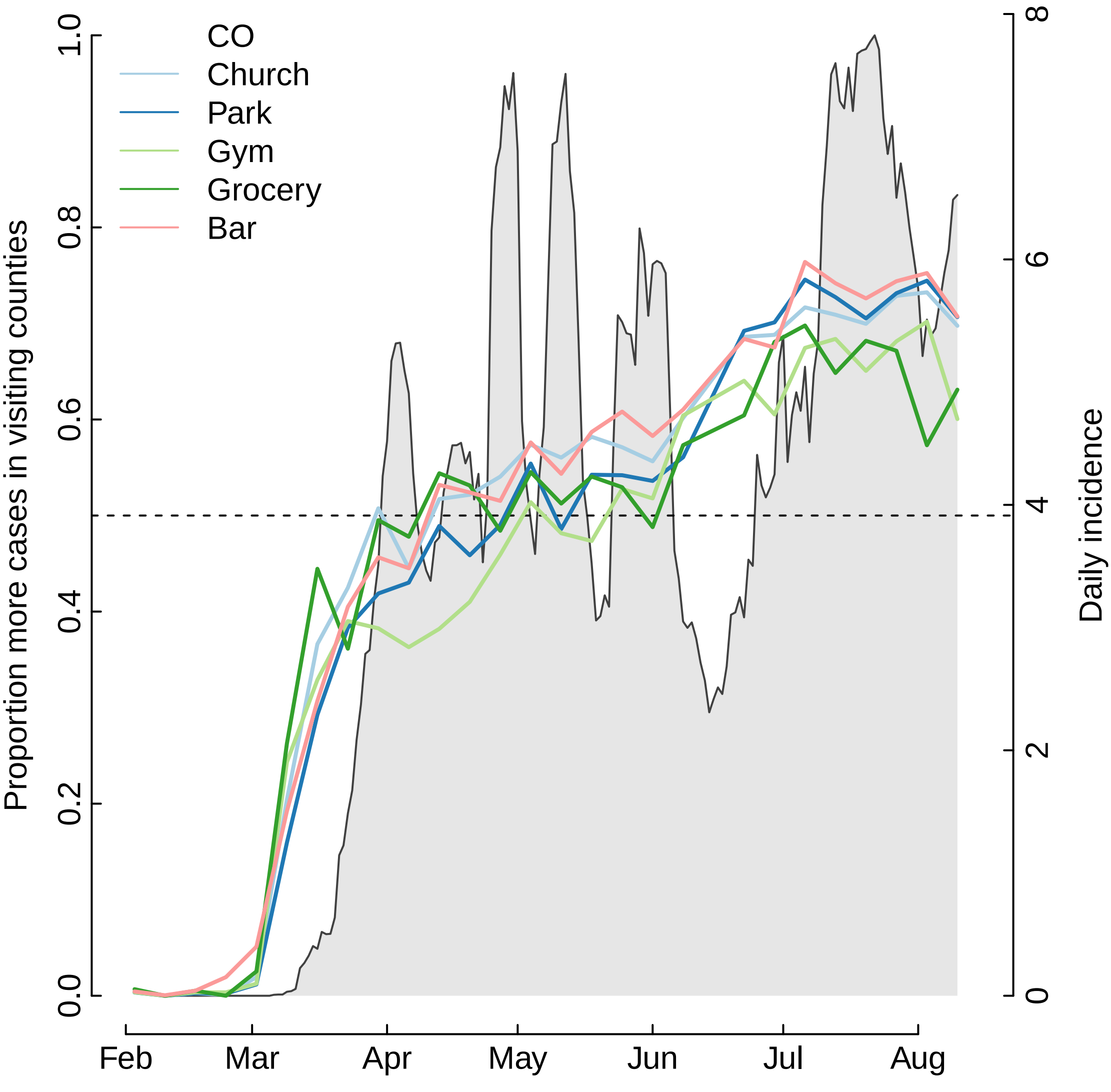

CO
Church
Park
Gym
Grocery
Bar
Proportion more cases in visiting counties
Daily incidence
0.0
0.2
0.4
0.6
0.8
1.0
0
2
4
6
8
Feb
Mar
Apr
May
Jun
Jul
Aug

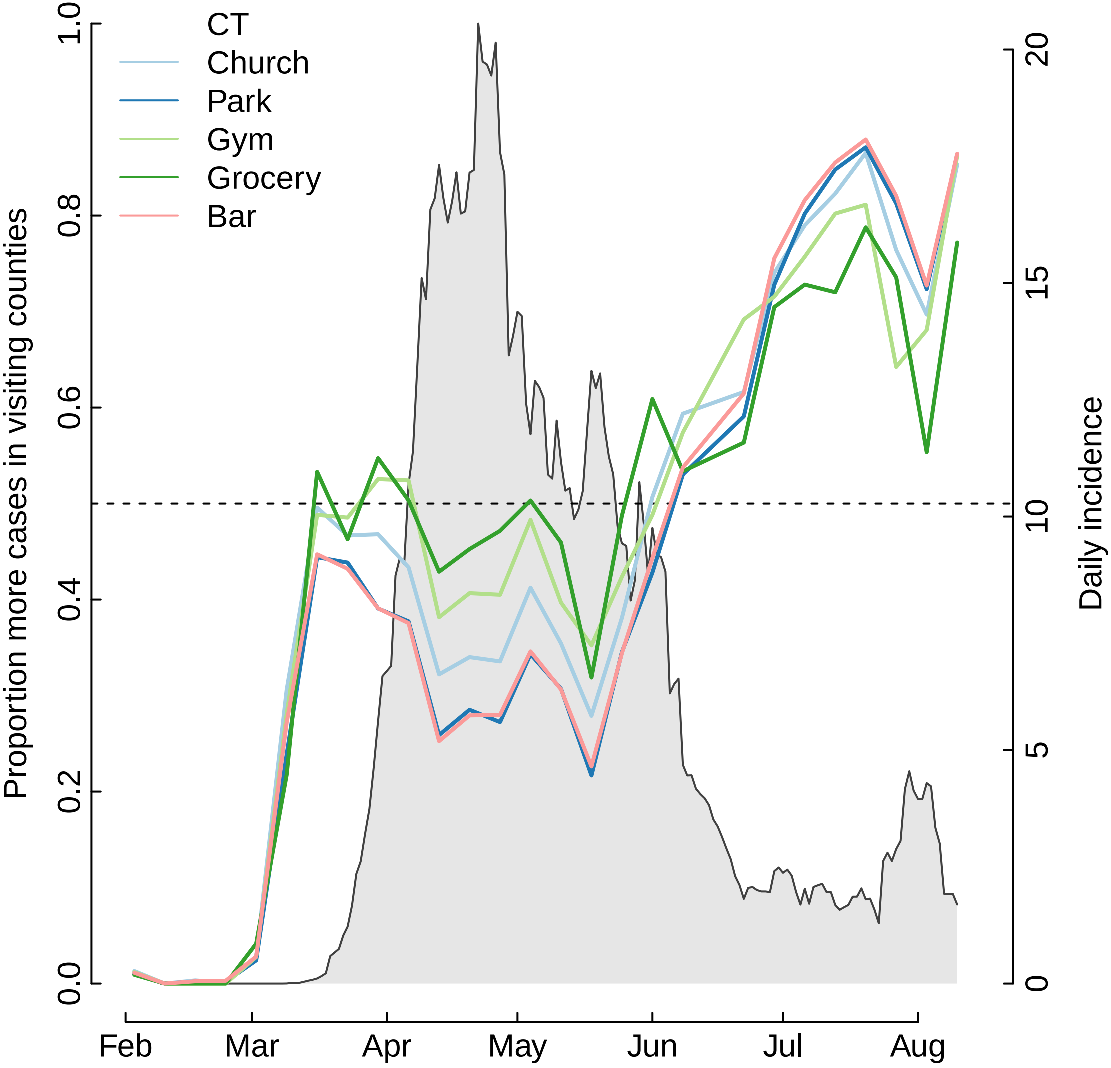
CT
Church
Park
Gym
Grocery
Bar
Proportion more cases in visiting counties
Daily incidence
0.0
0.2
0.4
0.6
0.8
1.0
0
5
10
15
20
Feb
Mar
Apr
May
Jun
Jul
Aug

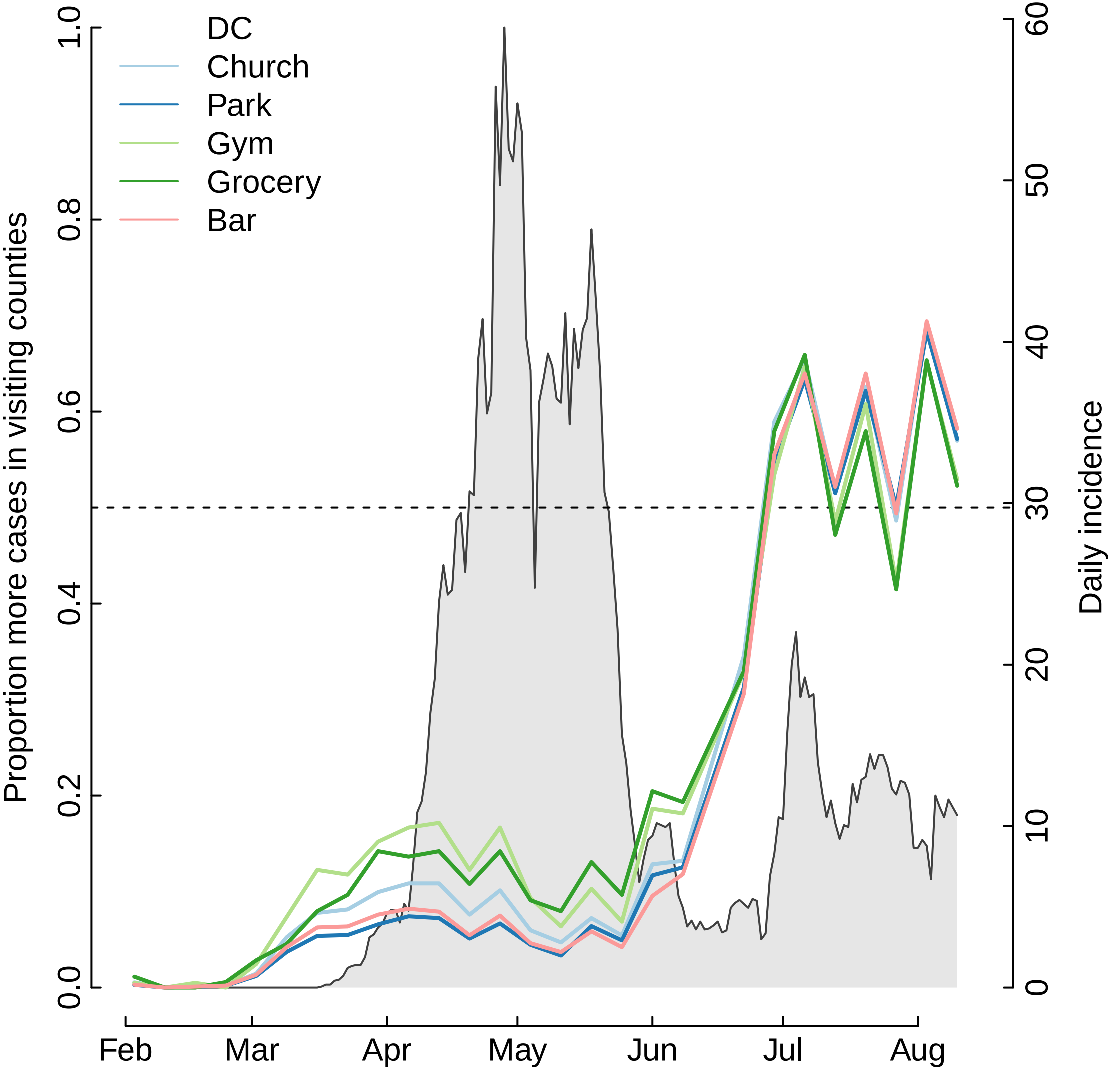

DC
Church
Park
Gym
Grocery
Bar
Proportion more cases in visiting counties
Daily incidence
0.0
0.2
0.4
0.6
0.8
1.0
0
10
20
30
40
50
60
Feb
Mar
Apr
May
Jun
Jul
Aug

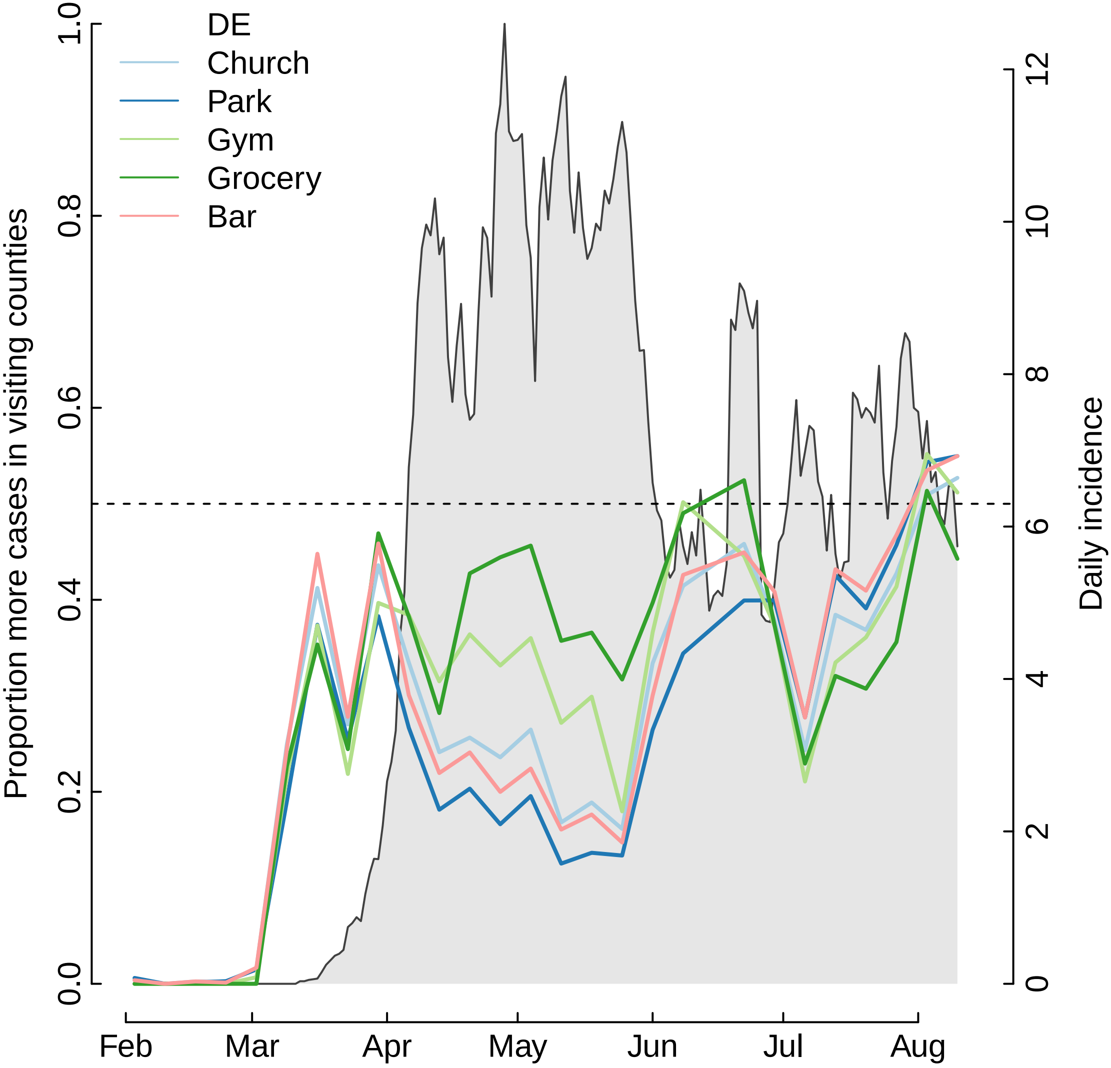

DE
Church
Park
Gym
Grocery
Bar
Proportion more cases in visiting counties
Daily incidence
0.0
0.2
0.4
0.6
0.8
1.0
0
2
4
6
8
10
12
Feb
Mar
Apr
May
Jun
Jul
Aug

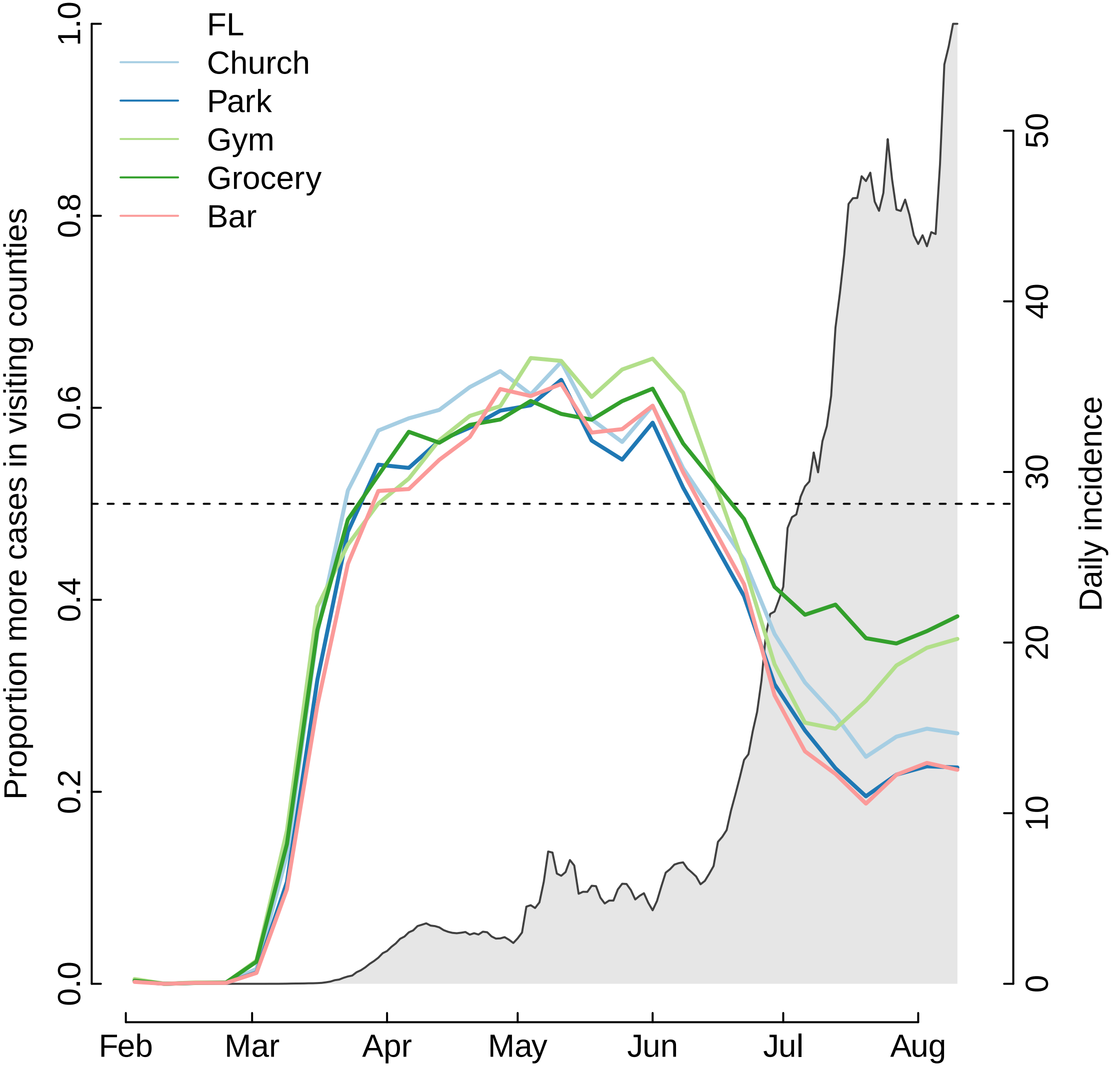
FL
Church
Park
Gym
Grocery
Bar
Proportion more cases in visiting counties
Daily incidence
0.0
0.2
0.4
0.6
0.8
1.0
0
10
20
30
40
50
Feb
Mar
Apr
May
Jun
Jul
Aug

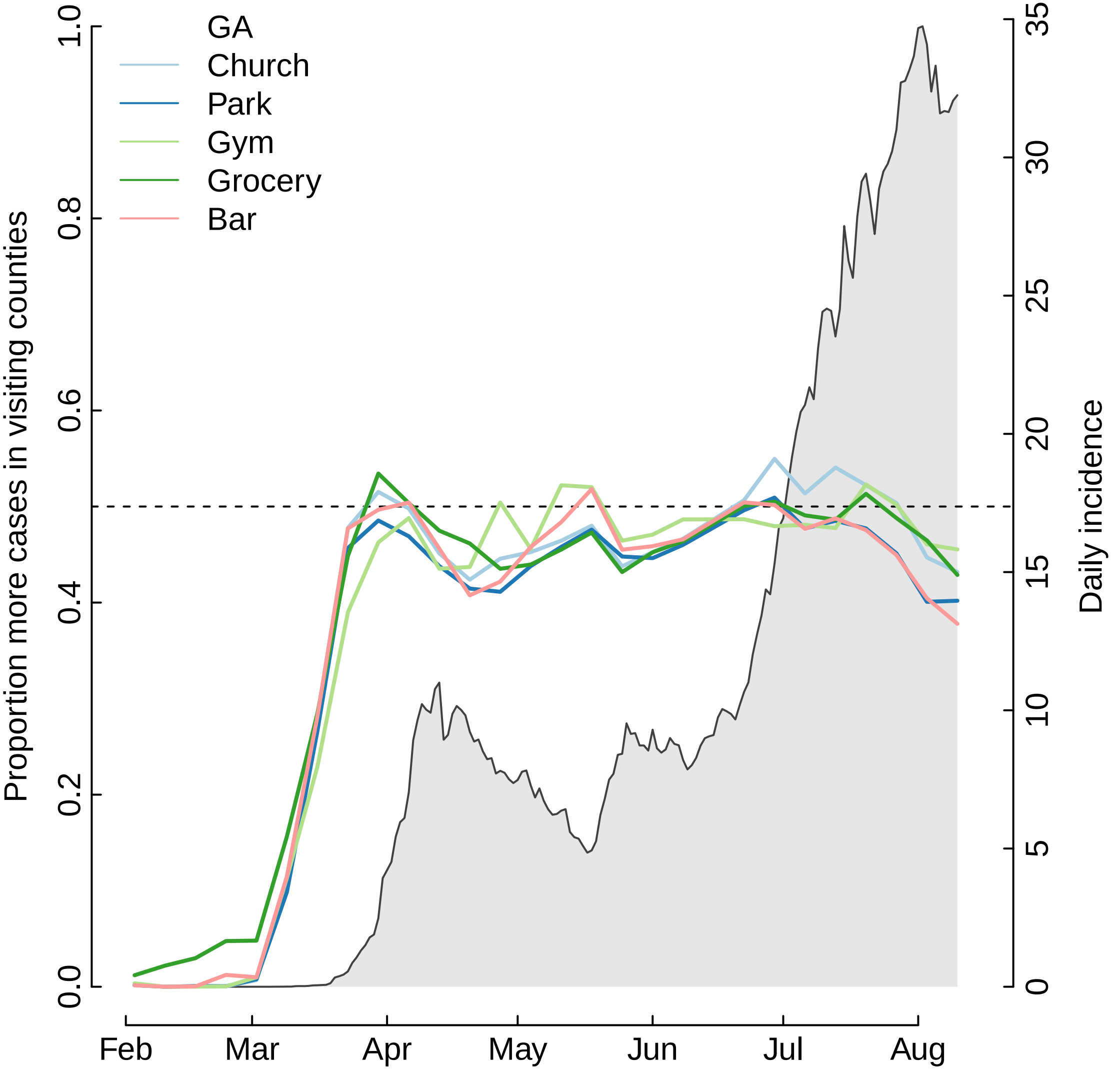
GA
Church
Park
Gym
Grocery
Bar
Proportion more cases in visiting counties
Daily incidence
0.0
0.2
0.4
0.6
0.8
1.0
0
5
10
15
20
25
30
35
Feb
Mar
Apr
May
Jun
Jul
Aug

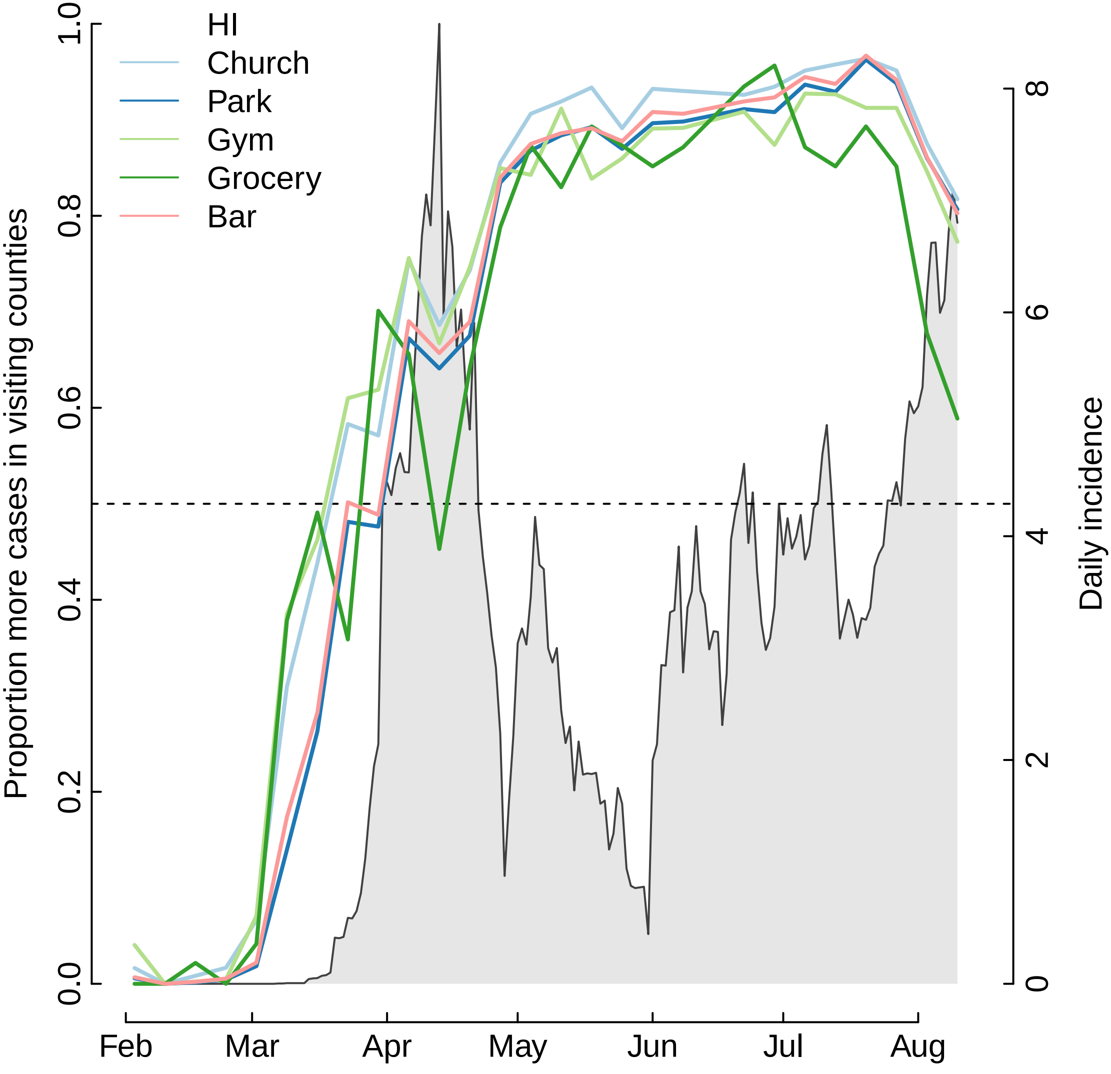
HI
Church
Park
Gym
Grocery
Bar
Proportion more cases in visiting counties
Daily incidence
0.0
0.2
0.4
0.6
0.8
1.0
0
2
4
6
8
Feb
Mar
Apr
May
Jun
Jul
Aug

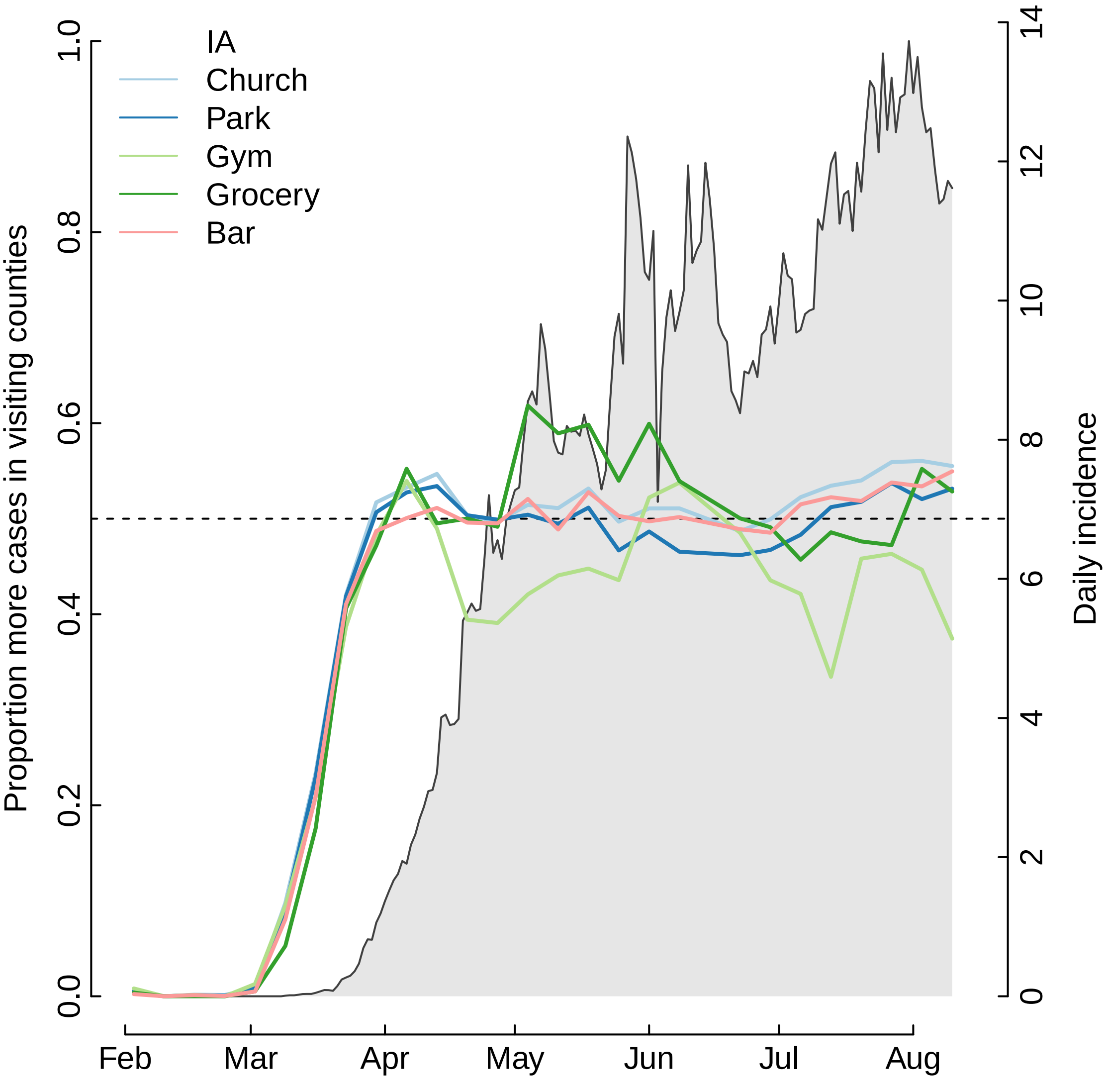

IA
Church
Park
Gym
Grocery
Bar
Proportion more cases in visiting counties
Daily incidence
0.0
0.2
0.4
0.6
0.8
1.0
0
2
4
6
8
10
12
14
Feb
Mar
Apr
May
Jun
Jul
Aug

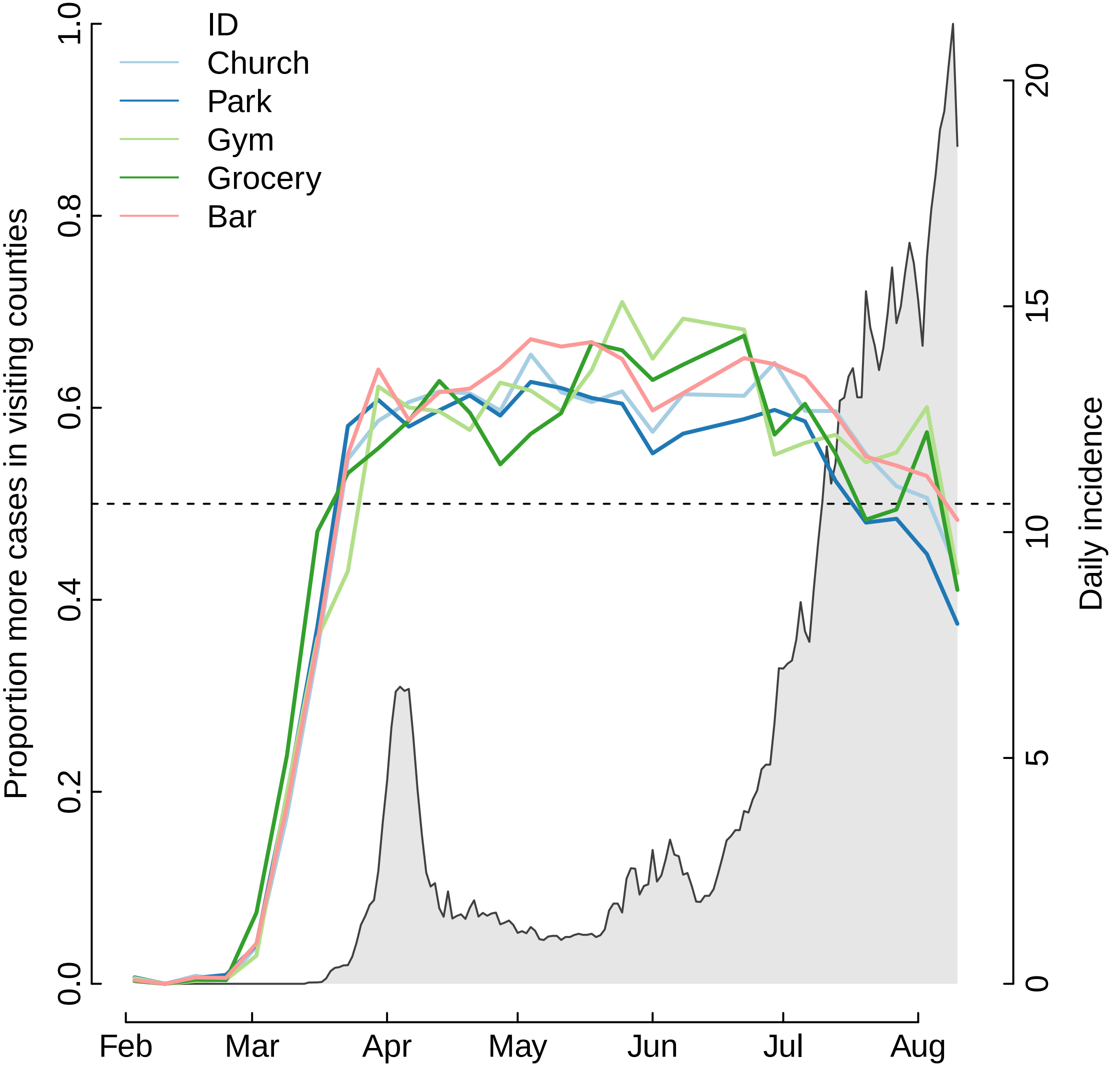
ID
Church
Park
Gym
Grocery
Bar
Proportion more cases in visiting counties
Daily incidence
0.0
0.2
0.4
0.6
0.8
1.0
0
5
10
15
20
Feb
Mar
Apr
May
Jun
Jul
Aug

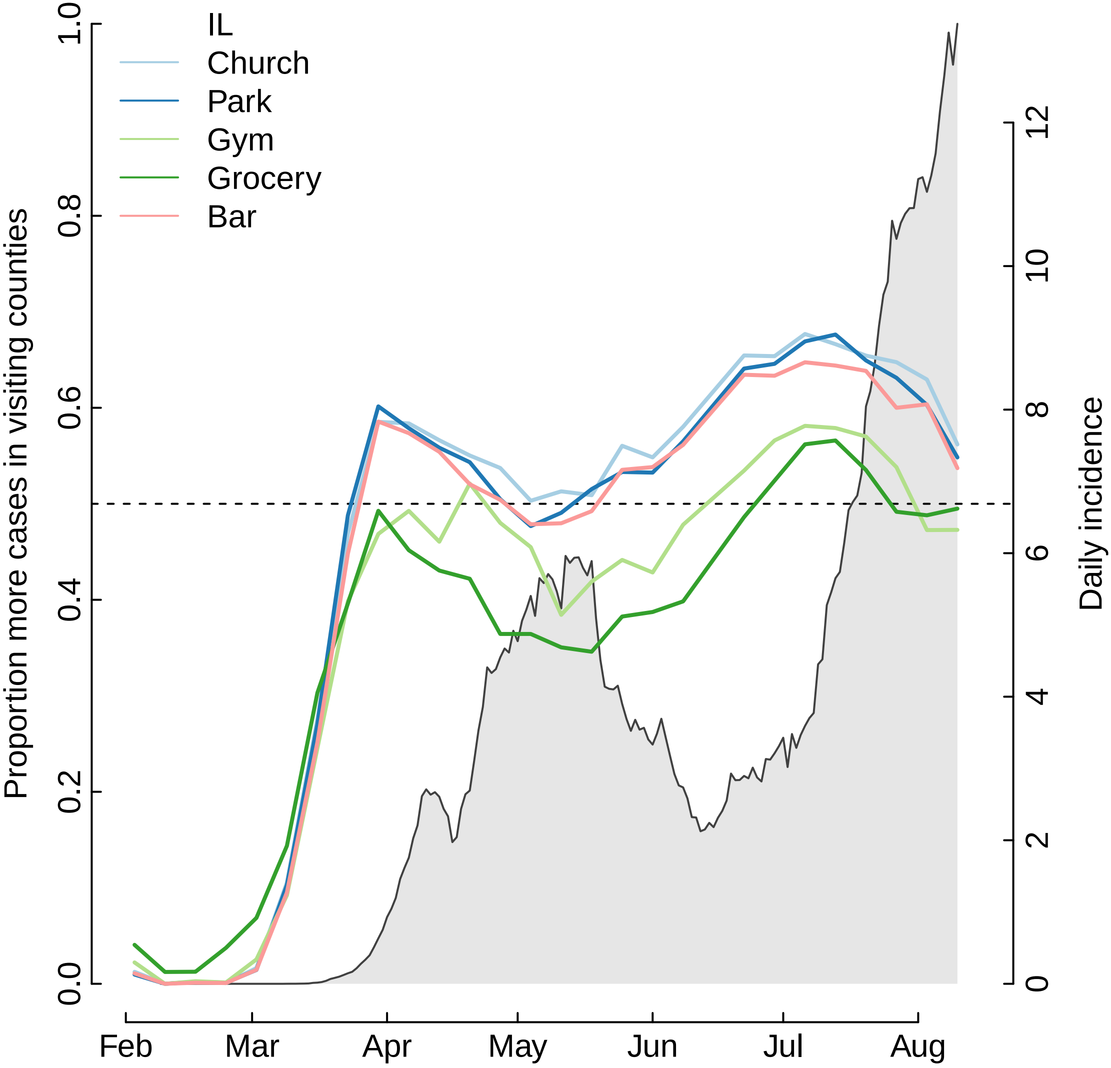

IL
Church
Park
Gym
Grocery
Bar
Proportion more cases in visiting counties
Daily incidence
0.0
0.2
0.4
0.6
0.8
1.0
0
2
4
6
8
10
12
Feb
Mar
Apr
May
Jun
Jul
Aug

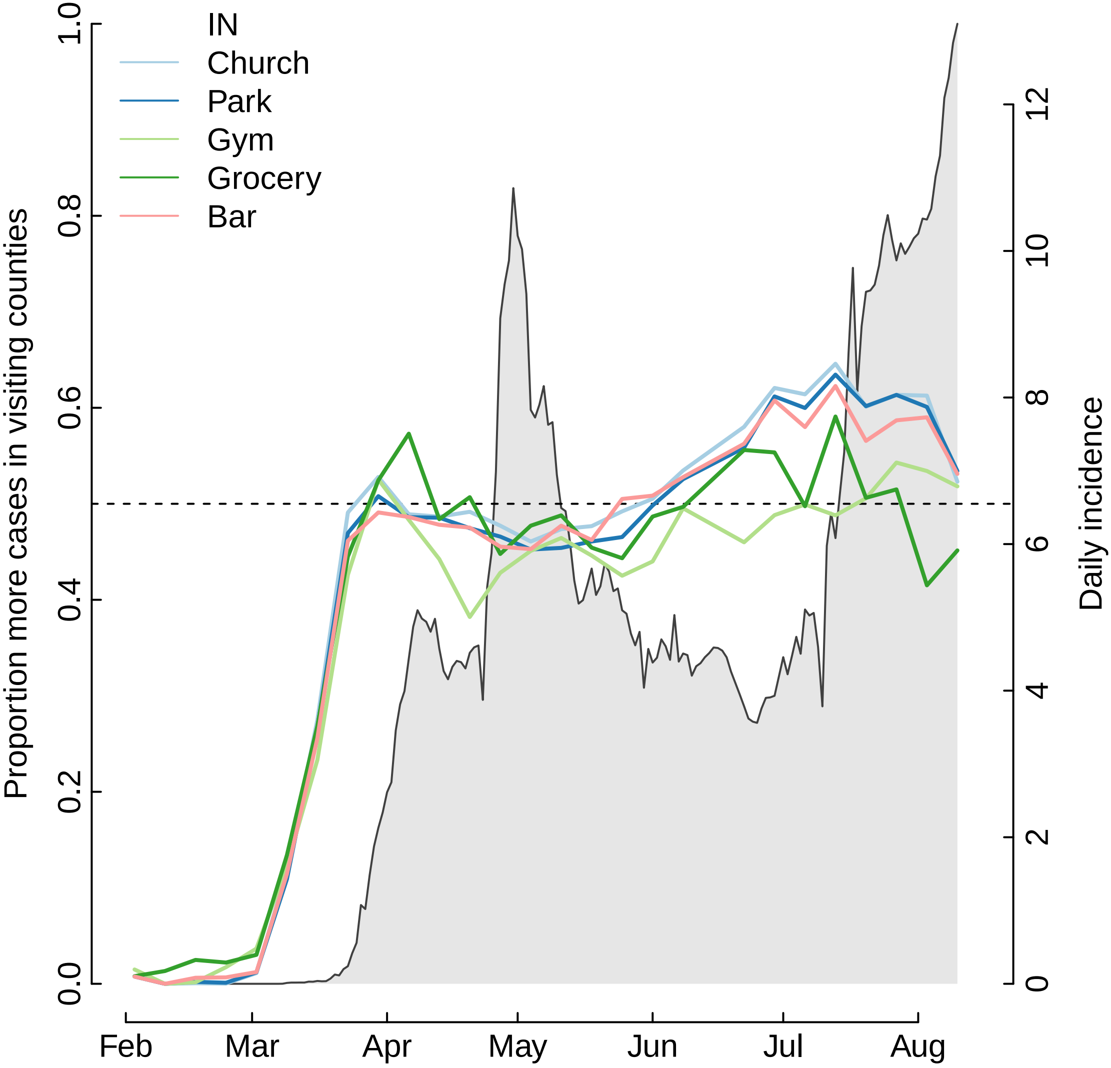
IN
Church
Park
Gym
Grocery
Bar
Proportion more cases in visiting counties
Daily incidence
0.0
0.2
0.4
0.6
0.8
1.0
0
2
4
6
8
10
12
Feb
Mar
Apr
May
Jun
Jul
Aug

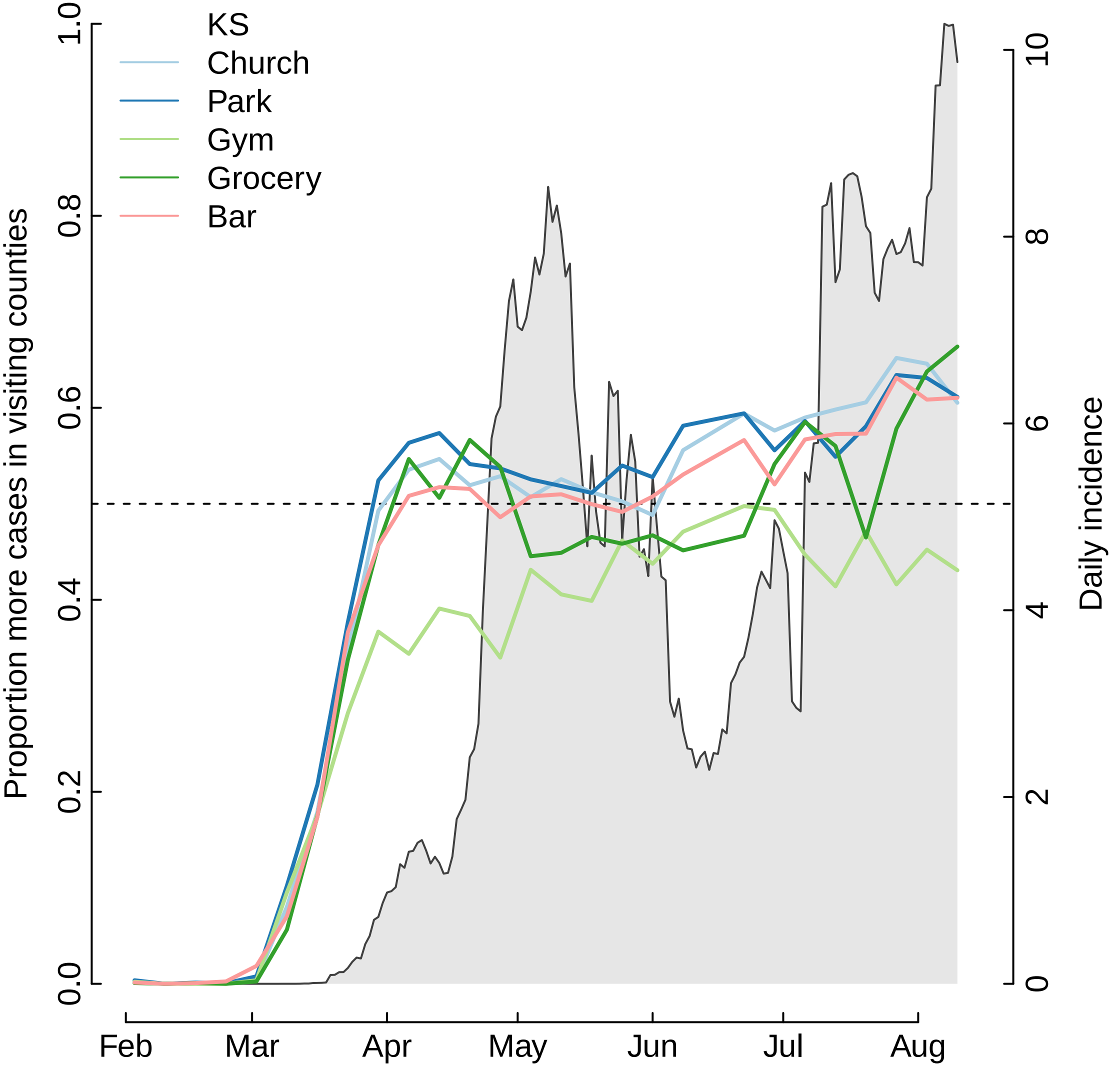

KS
Church
Park
Gym
Grocery
Bar
Proportion more cases in visiting counties
Daily incidence
0.0
0.2
0.4
0.6
0.8
1.0
0
2
4
6
8
10
Feb
Mar
Apr
May
Jun
Jul
Aug

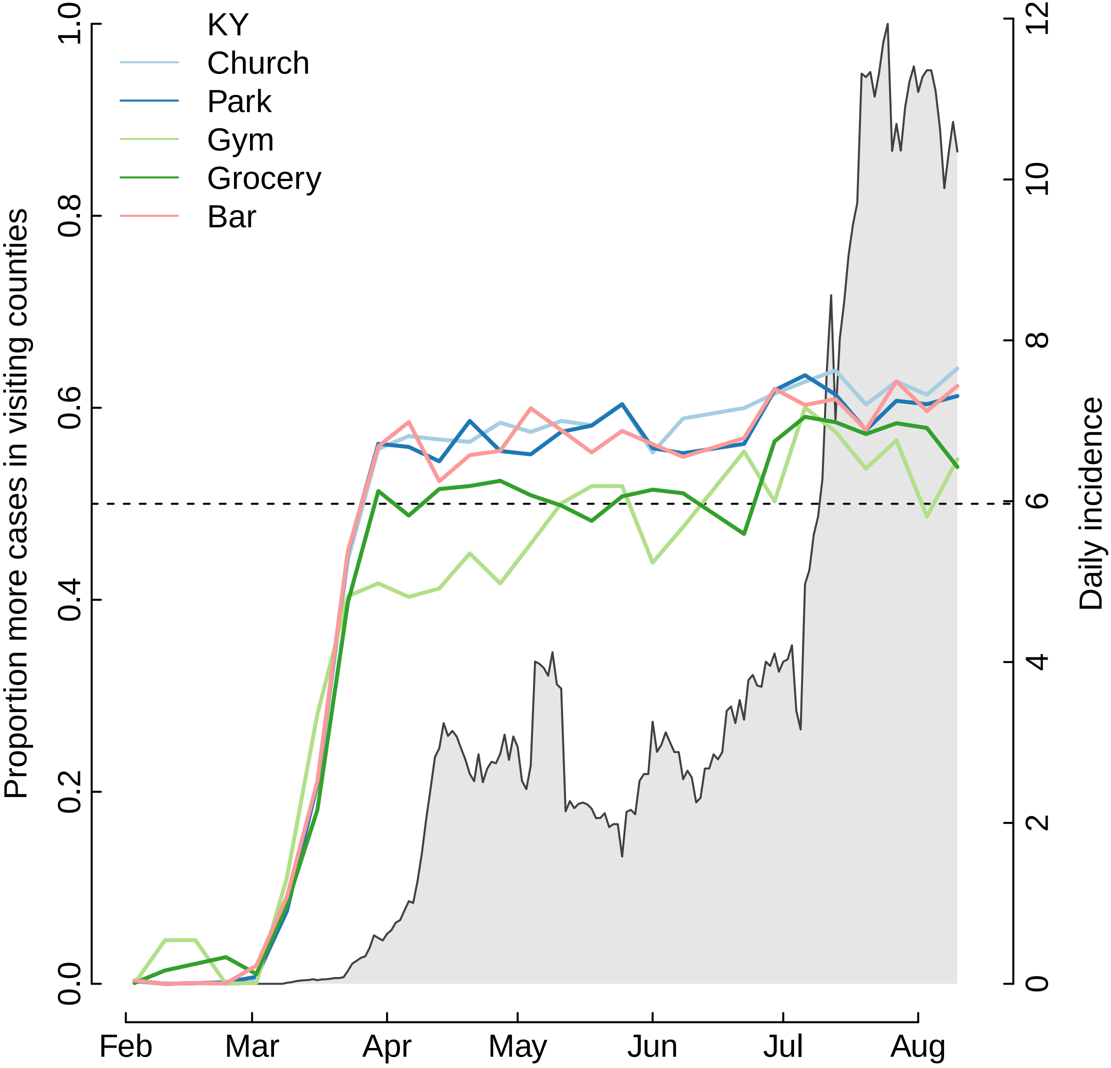

KY
Church
Park
Gym
Grocery
Bar
Proportion more cases in visiting counties
Daily incidence
0.0
0.2
0.4
0.6
0.8
1.0
0
2
4
6
8
10
12
Feb
Mar
Apr
May
Jun
Jul
Aug

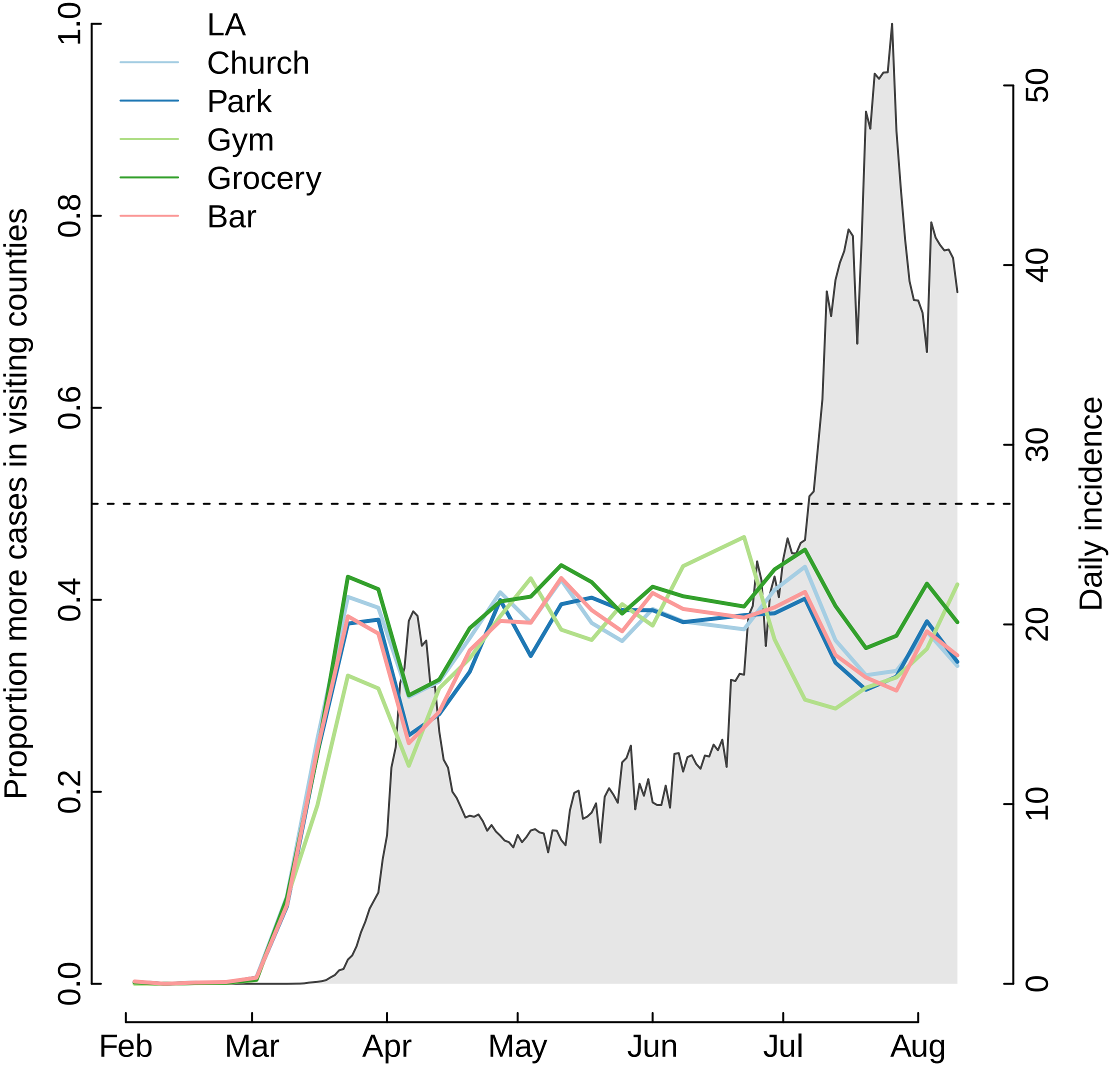
LA
Church
Park
Gym
Grocery
Bar
Proportion more cases in visiting counties
Daily incidence
0.0
0.2
0.4
0.6
0.8
1.0
0
10
20
30
40
50
Feb
Mar
Apr
May
Jun
Jul
Aug

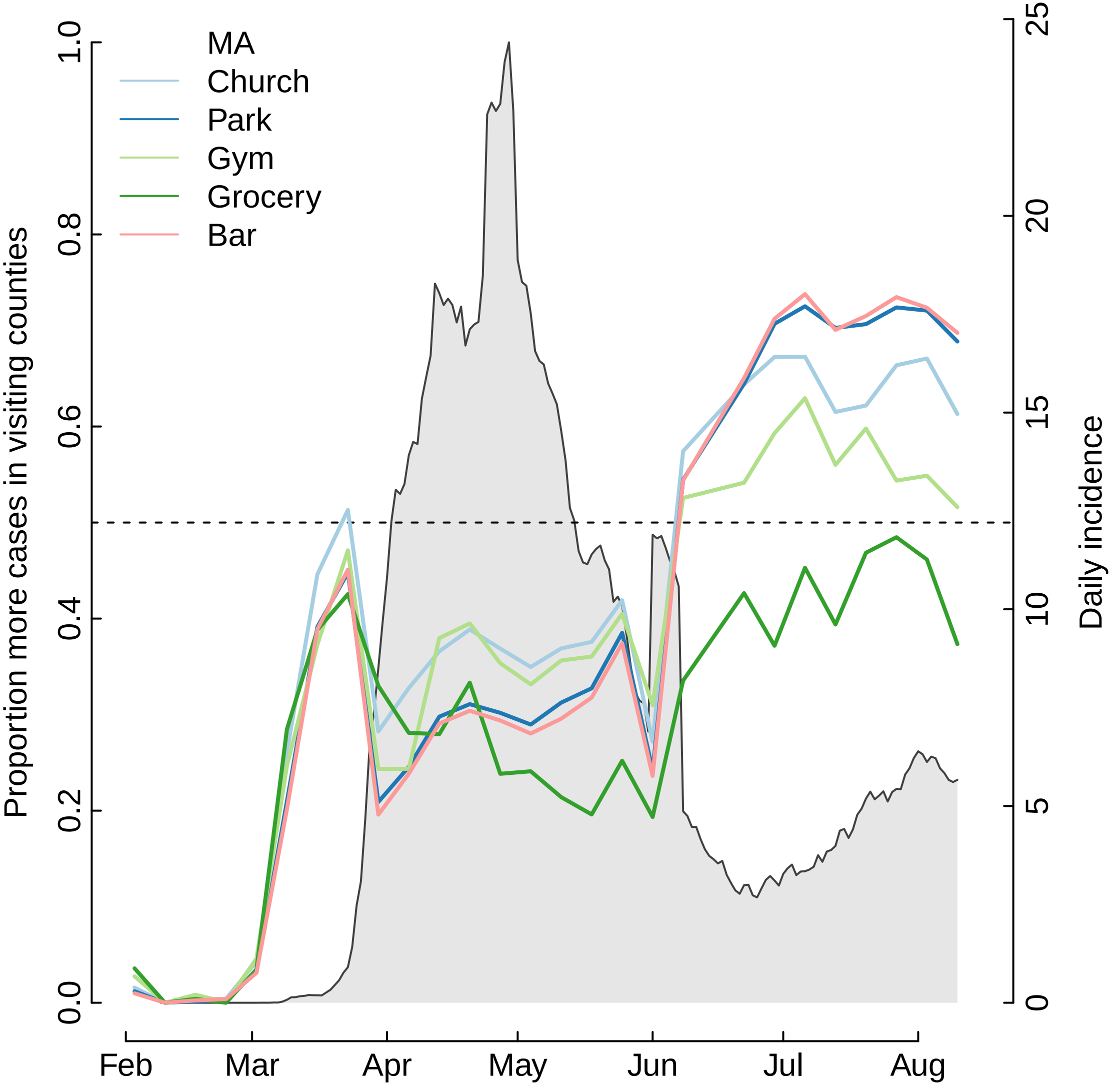

MA
Church
Park
Gym
Grocery
Bar
Proportion more cases in visiting counties
Daily incidence
0.0
0.2
0.4
0.6
0.8
1.0
0
5
10
15
20
25
Feb
Mar
Apr
May
Jun
Jul
Aug

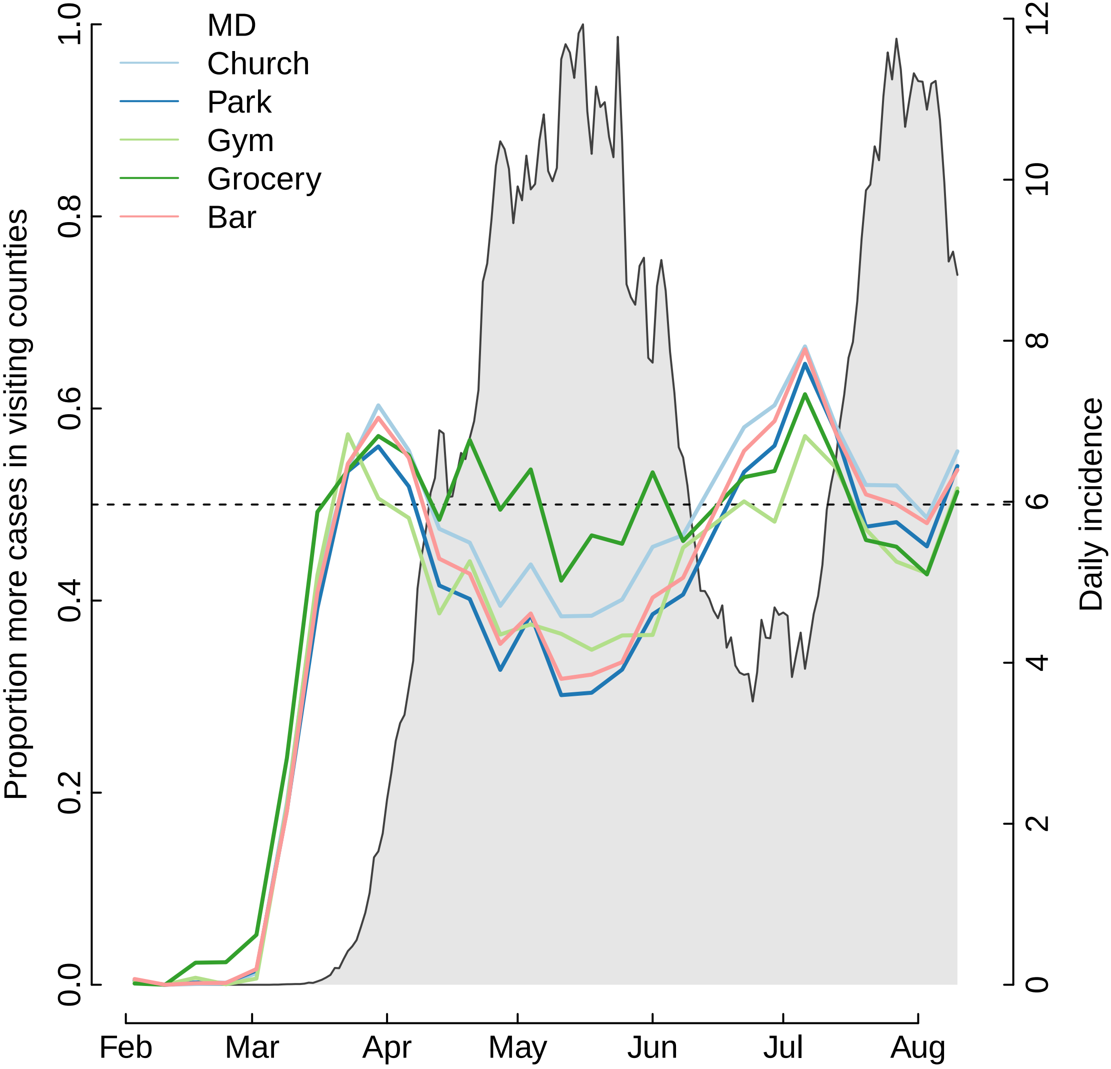
MD
Church
Park
Gym
Grocery
Bar
Proportion more cases in visiting counties
Daily incidence
0.0
0.2
0.4
0.6
0.8
1.0
0
2
4
6
8
10
12
Feb
Mar
Apr
May
Jun
Jul
Aug

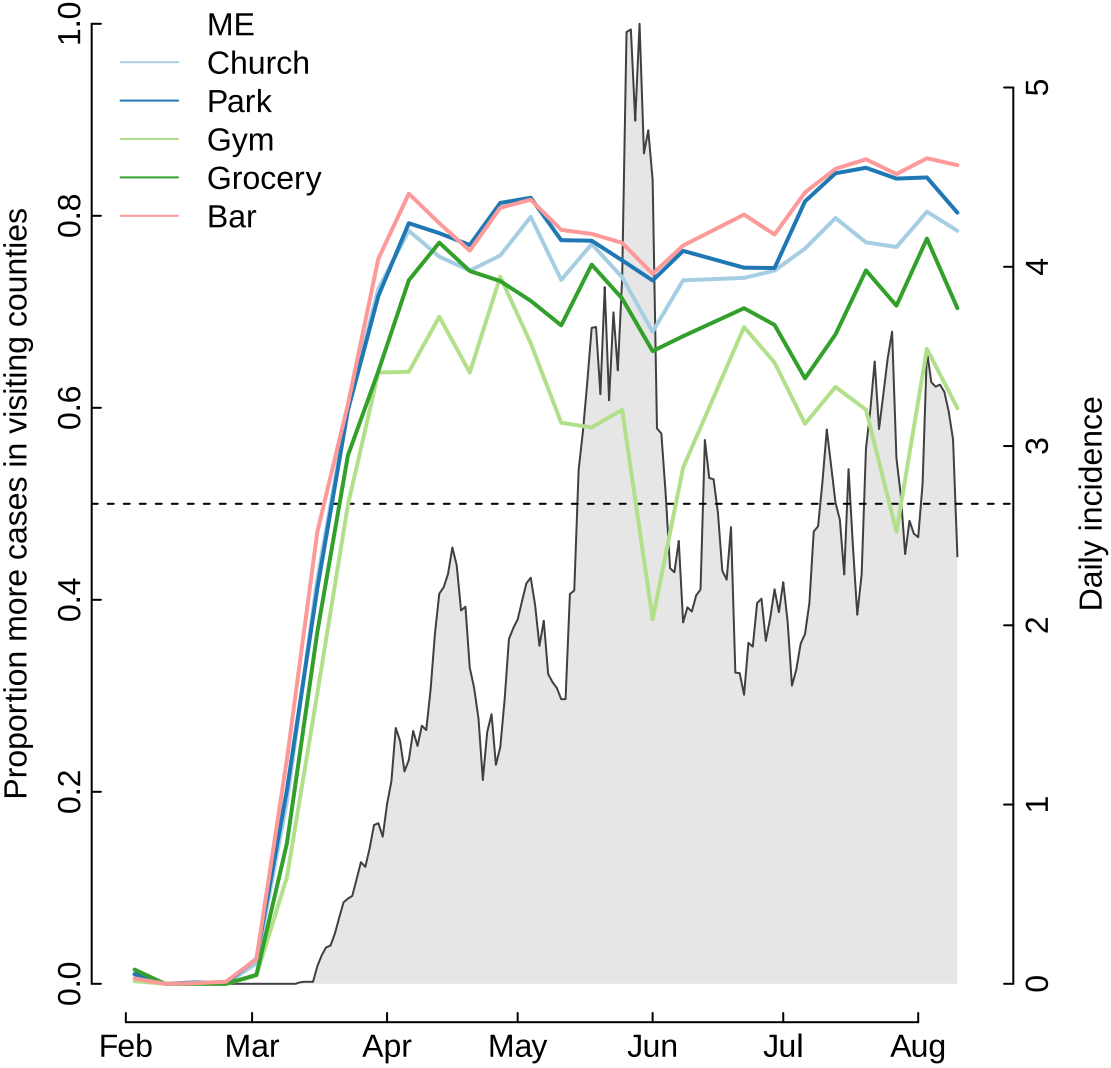
ME
Church
Park
Gym
Grocery
Bar
Proportion more cases in visiting counties
Daily incidence
0.0
0.2
0.4
0.6
0.8
1.0
0
1
2
3
4
5
Feb
Mar
Apr
May
Jun
Jul
Aug

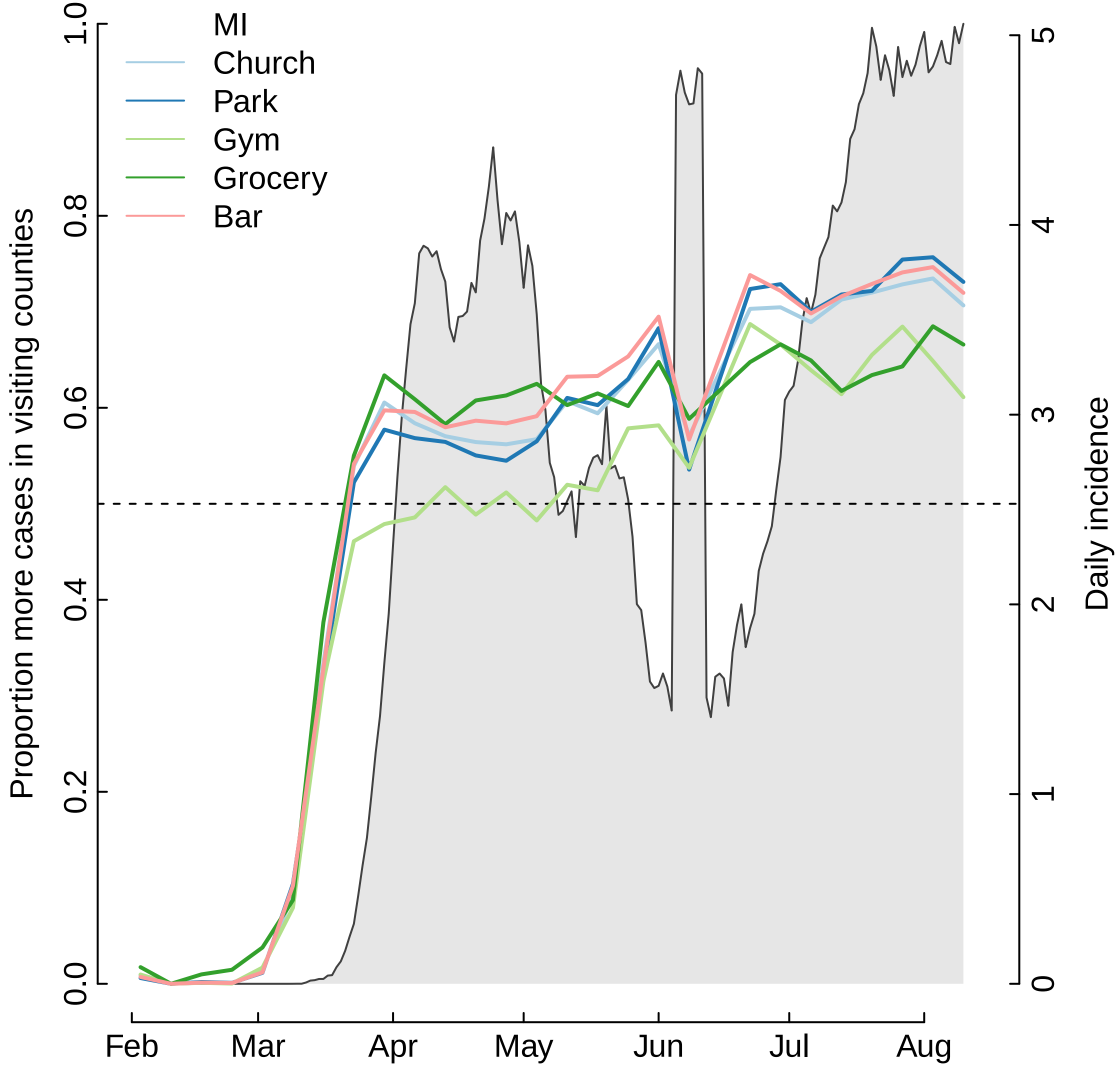

MI
Church
Park
Gym
Grocery
Bar
Proportion more cases in visiting counties
Daily incidence
0.0
0.2
0.4
0.6
0.8
1.0
0
1
2
3
4
5
Feb
Mar
Apr
May
Jun
Jul
Aug

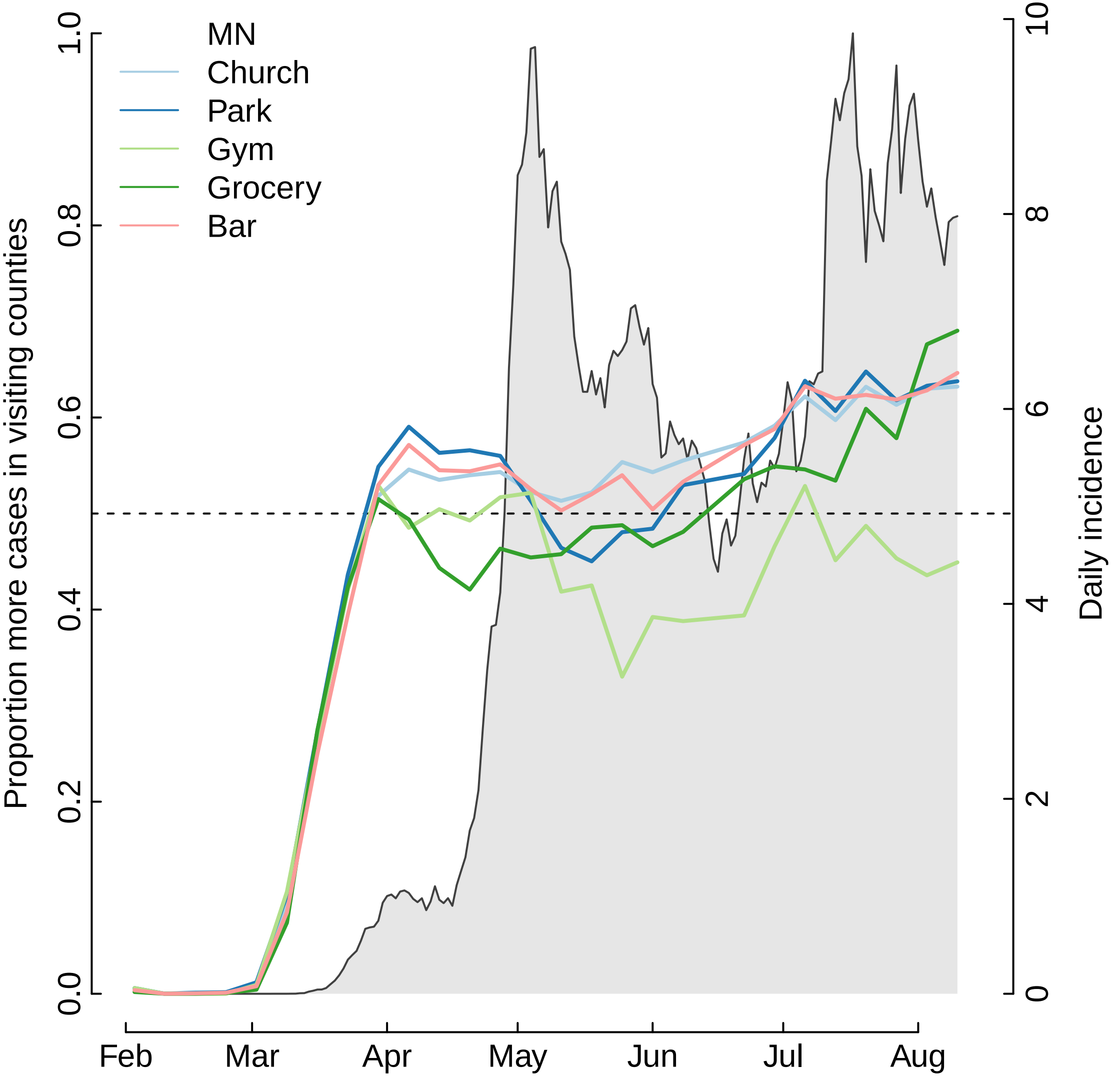

MN
Church
Park
Gym
Grocery
Bar
Proportion more cases in visiting counties
Daily incidence
0.0
0.2
0.4
0.6
0.8
1.0
0
2
4
6
8
10
Feb
Mar
Apr
May
Jun
Jul
Aug

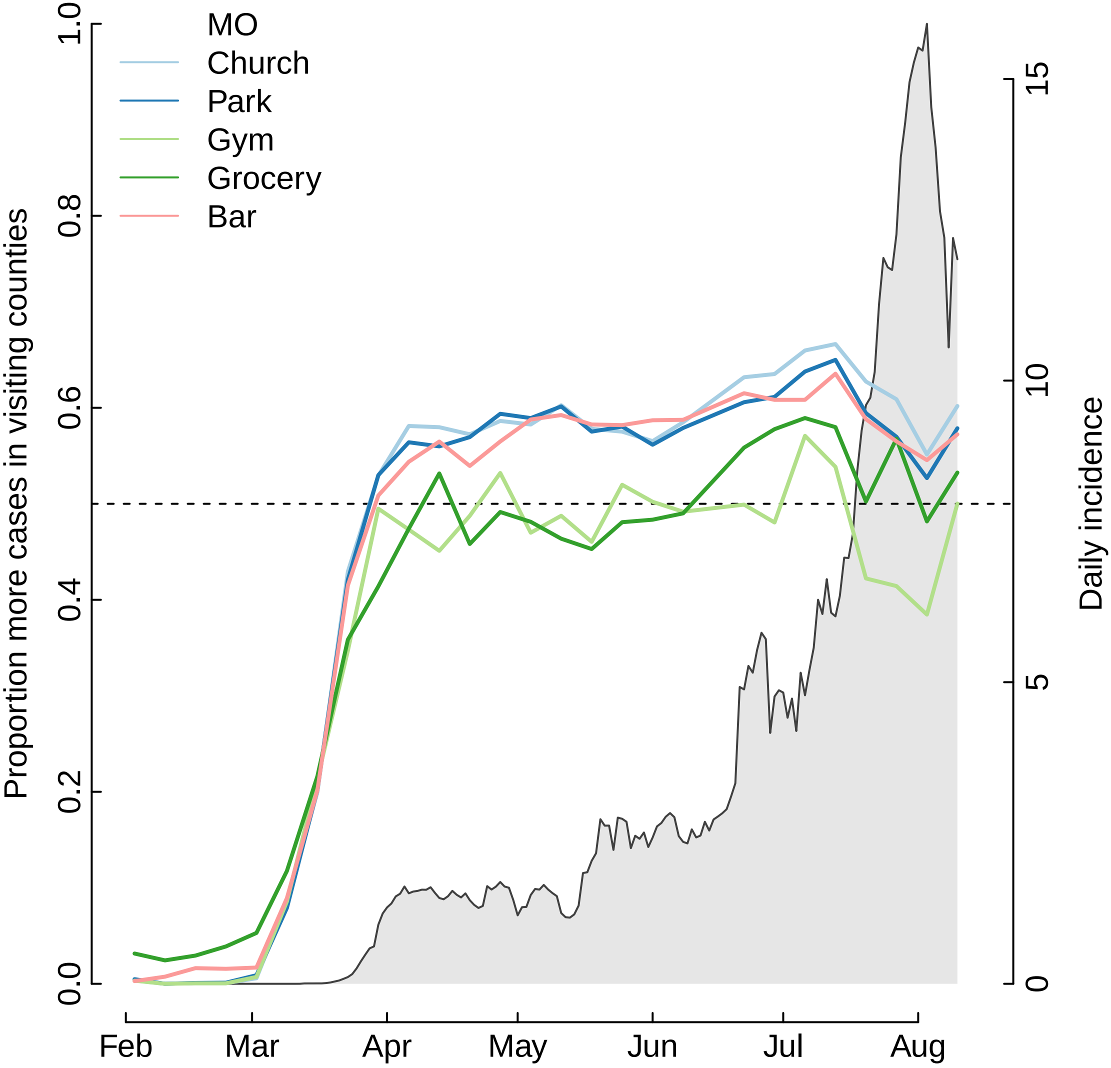

MO
Church
Park
Gym
Grocery
Bar
Proportion more cases in visiting counties
Daily incidence
0.0
0.2
0.4
0.6
0.8
1.0
0
5
10
15
Feb
Mar
Apr
May
Jun
Jul
Aug

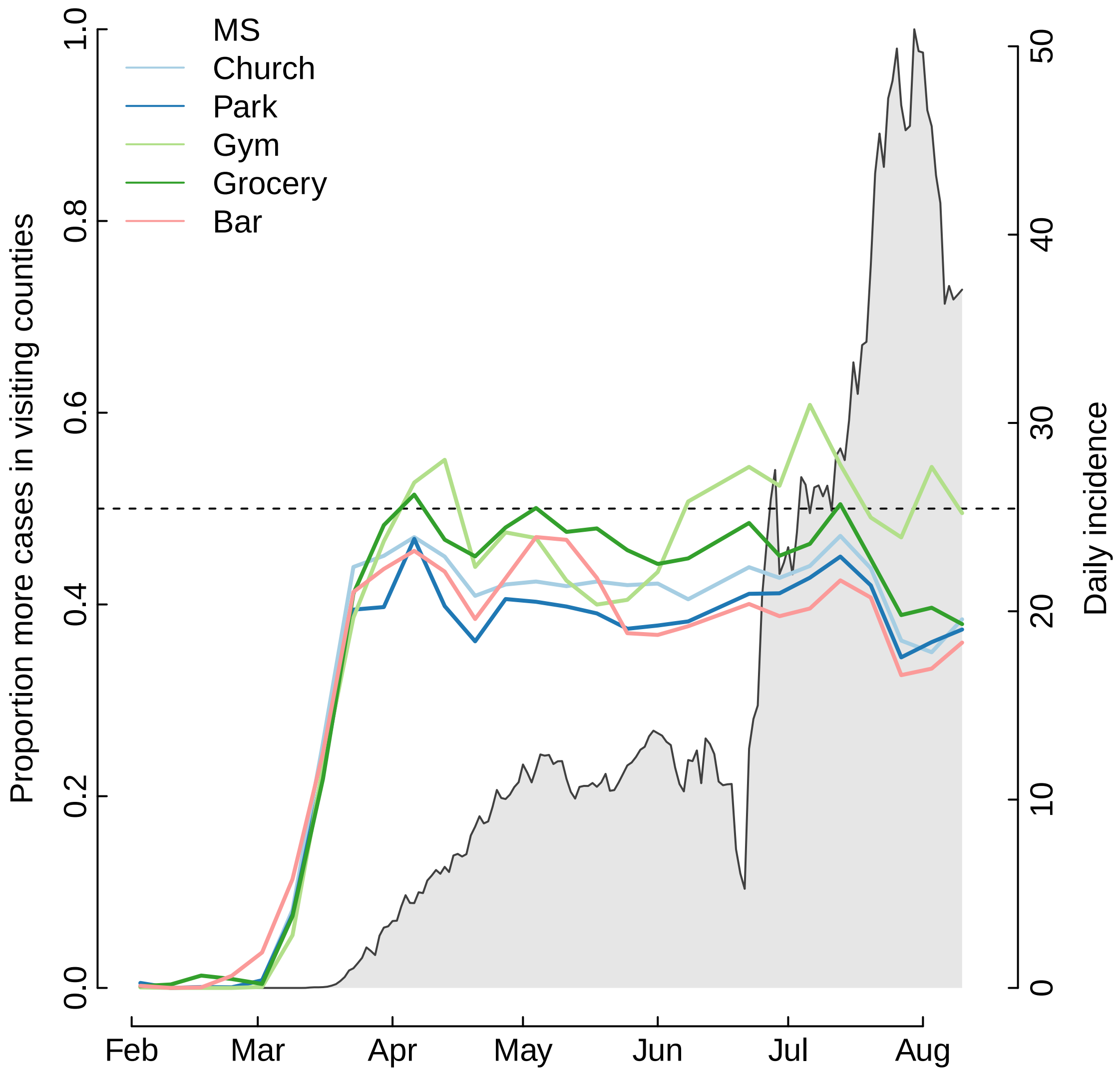

MS
Church
Park
Gym
Grocery
Bar
Proportion more cases in visiting counties
Daily incidence
0.0
0.2
0.4
0.6
0.8
1.0
0
10
20
30
40
50
Feb
Mar
Apr
May
Jun
Jul
Aug

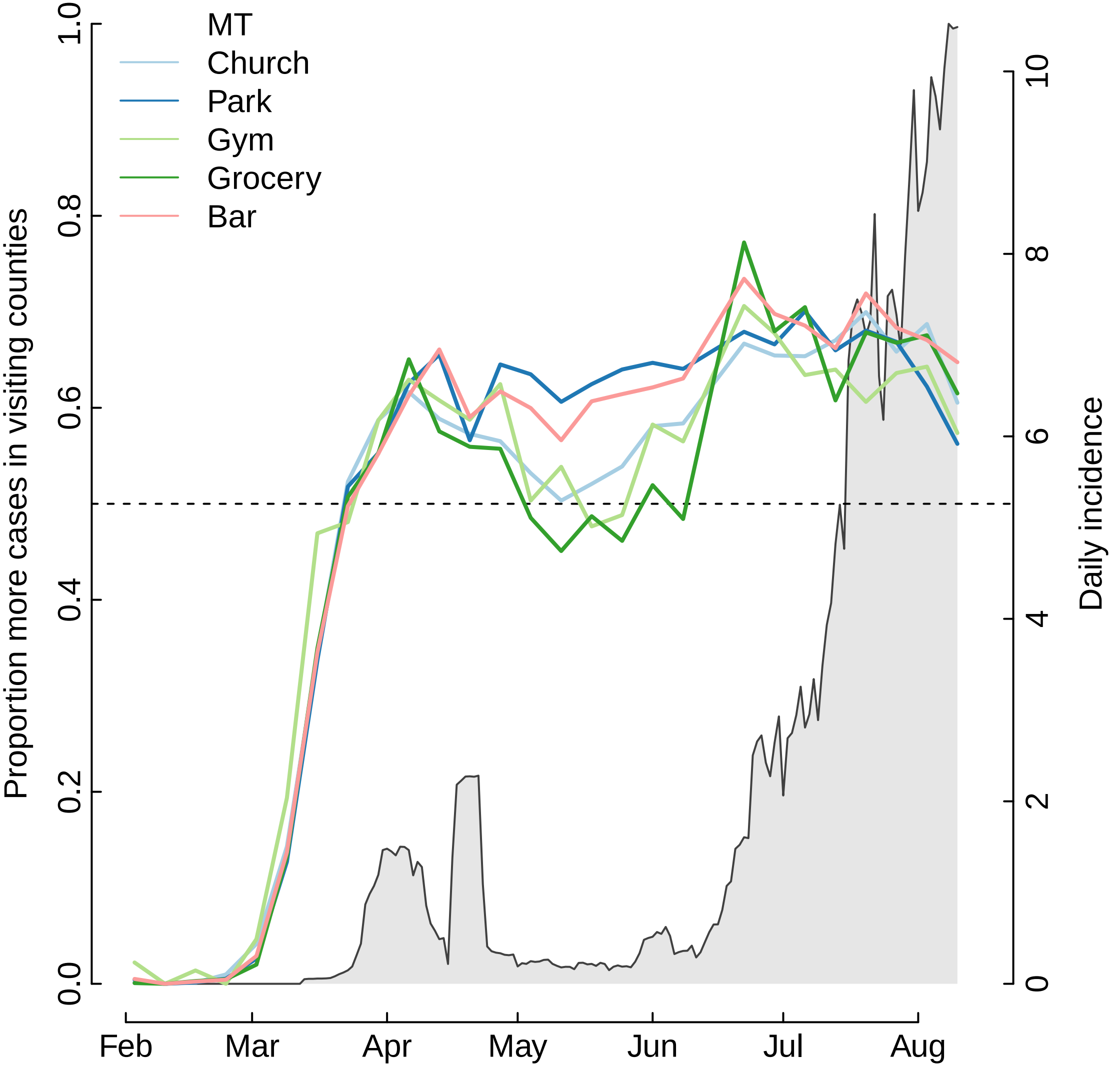

MT
Church
Park
Gym
Grocery
Bar
Proportion more cases in visiting counties
Daily incidence
0.0
0.2
0.4
0.6
0.8
1.0
0
2
4
6
8
10
Feb
Mar
Apr
May
Jun
Jul
Aug

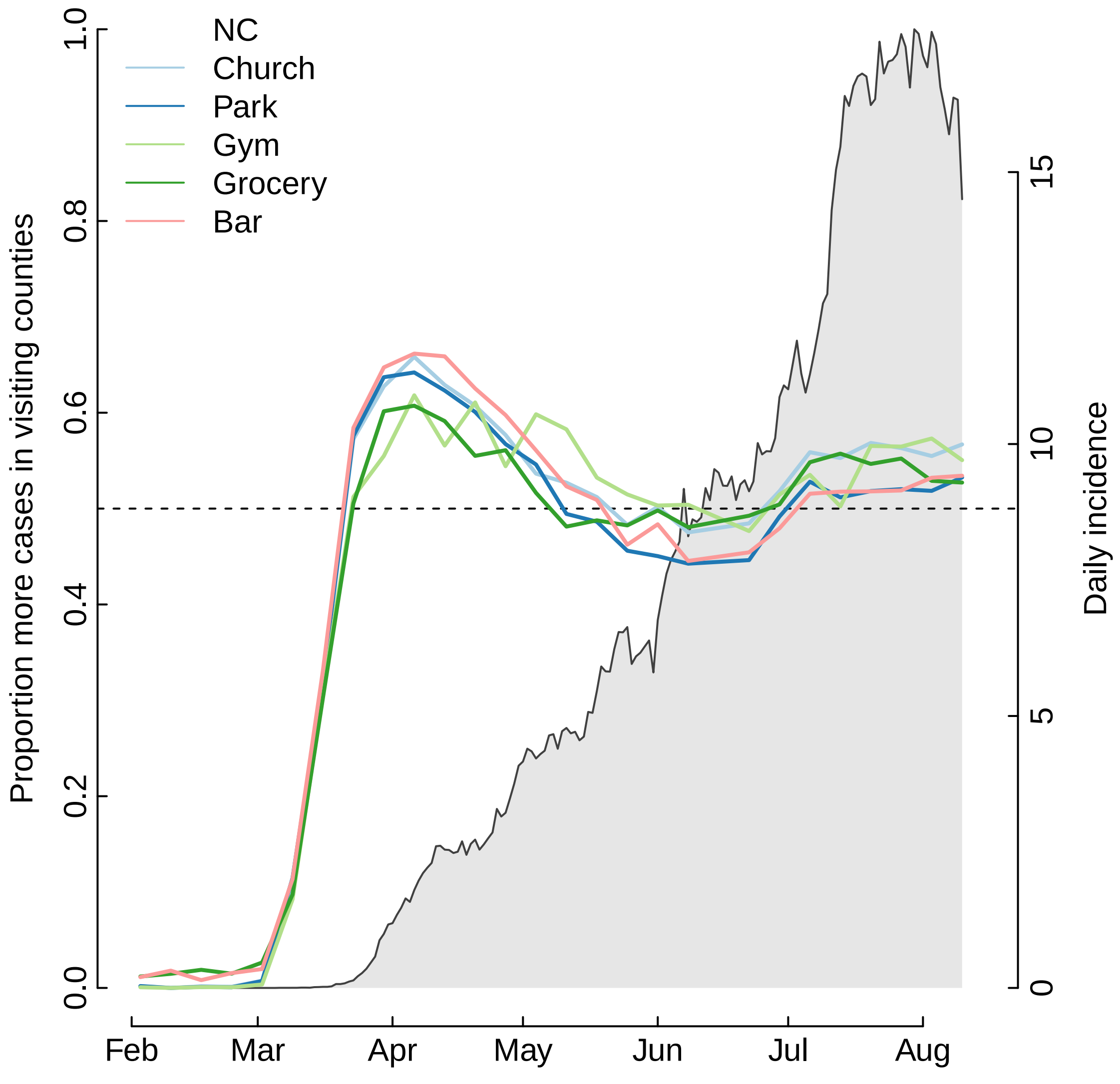

NC
Church
Park
Gym
Grocery
Bar
Proportion more cases in visiting counties
Daily incidence
0.0
0.2
0.4
0.6
0.8
1.0
0
5
10
15
Feb
Mar
Apr
May
Jun
Jul
Aug

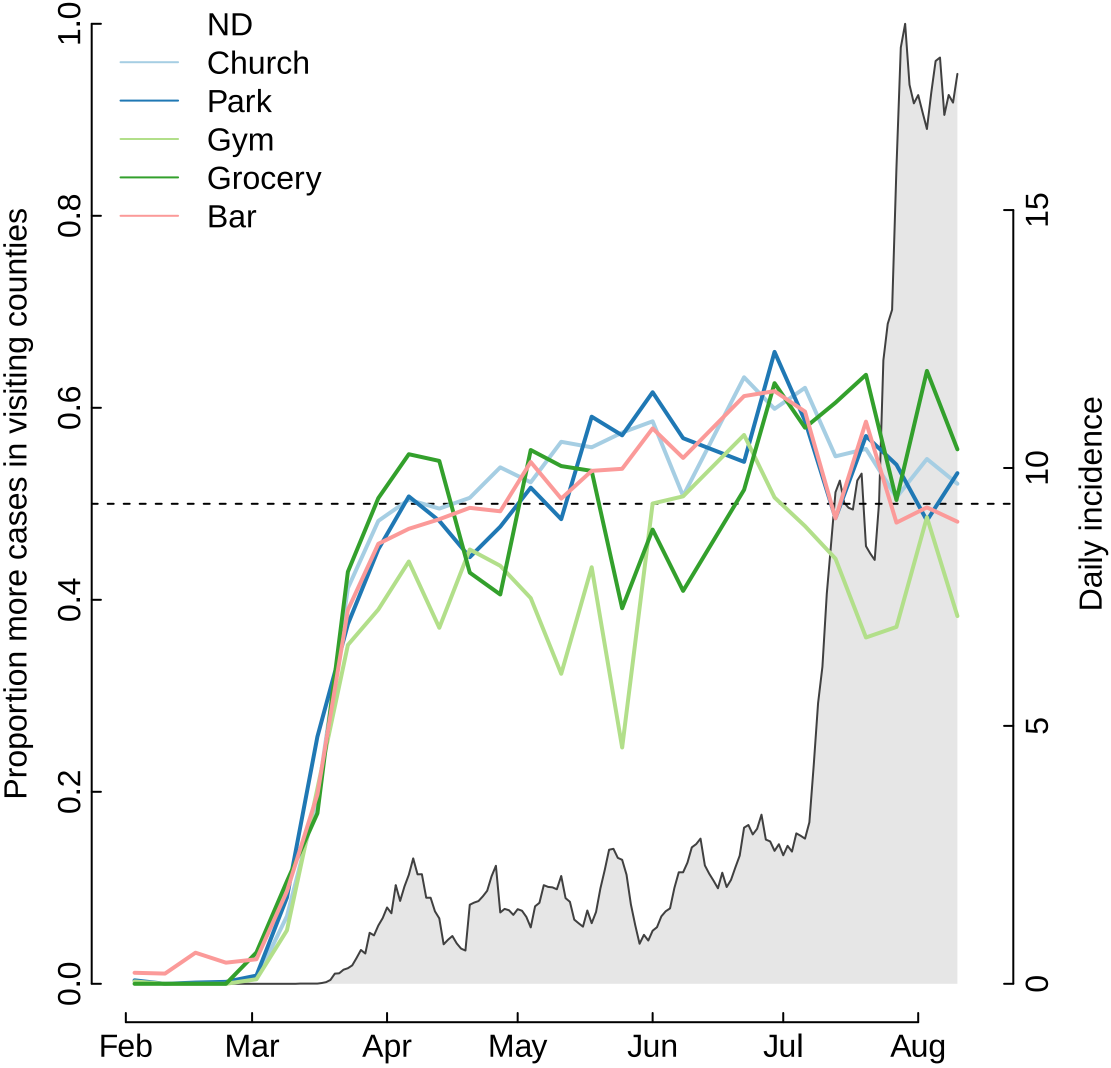

ND
Church
Park
Gym
Grocery
Bar
Proportion more cases in visiting counties
Daily incidence
0.0
0.2
0.4
0.6
0.8
1.0
0
5
10
15
Feb
Mar
Apr
May
Jun
Jul
Aug

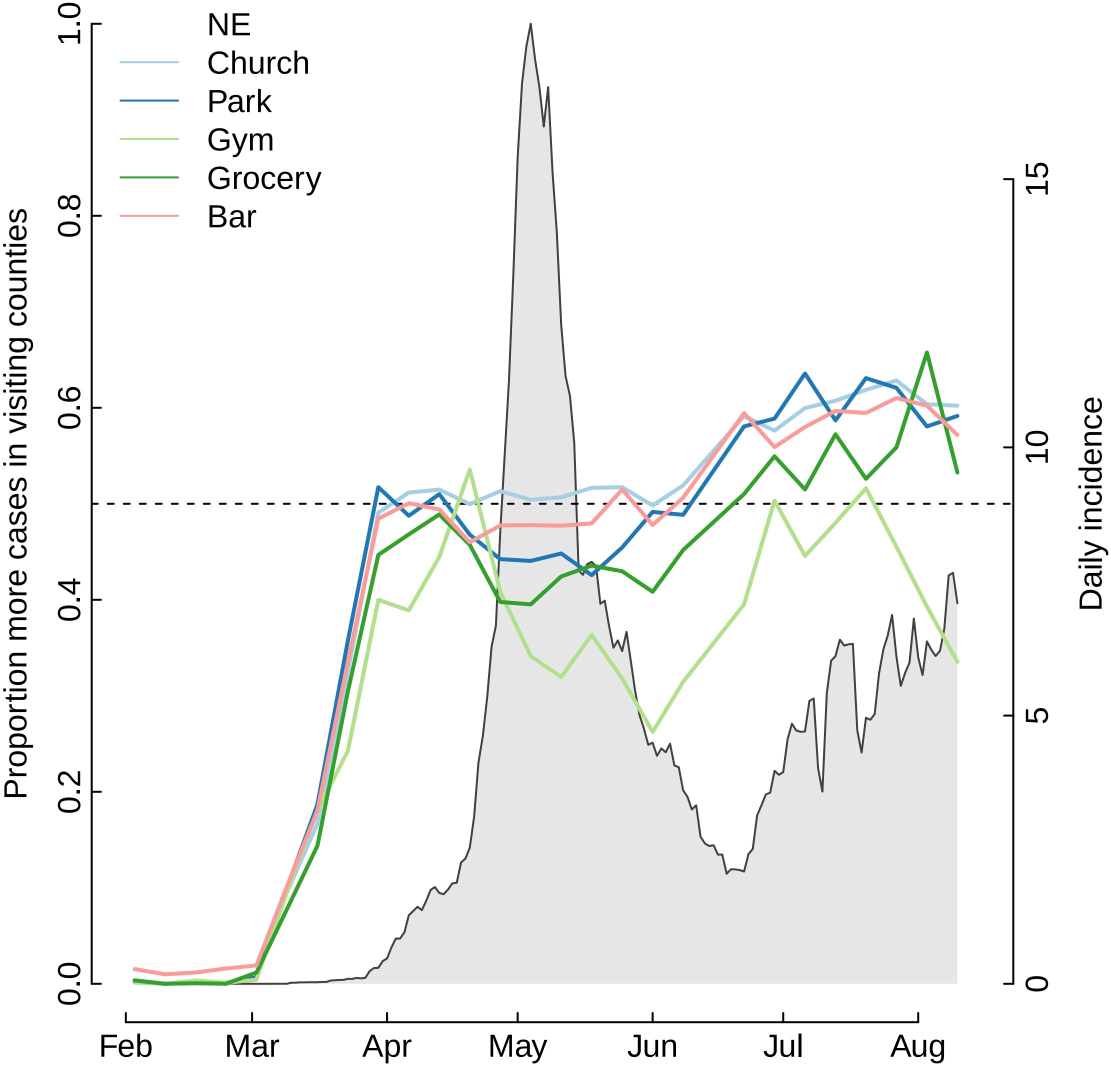
NE
Church
Park
Gym
Grocery
Bar
Proportion more cases in visiting counties
Daily incidence
0.0
0.2
0.4
0.6
0.8
1.0
0
5
10
15
Feb
Mar
Apr
May
Jun
Jul
Aug

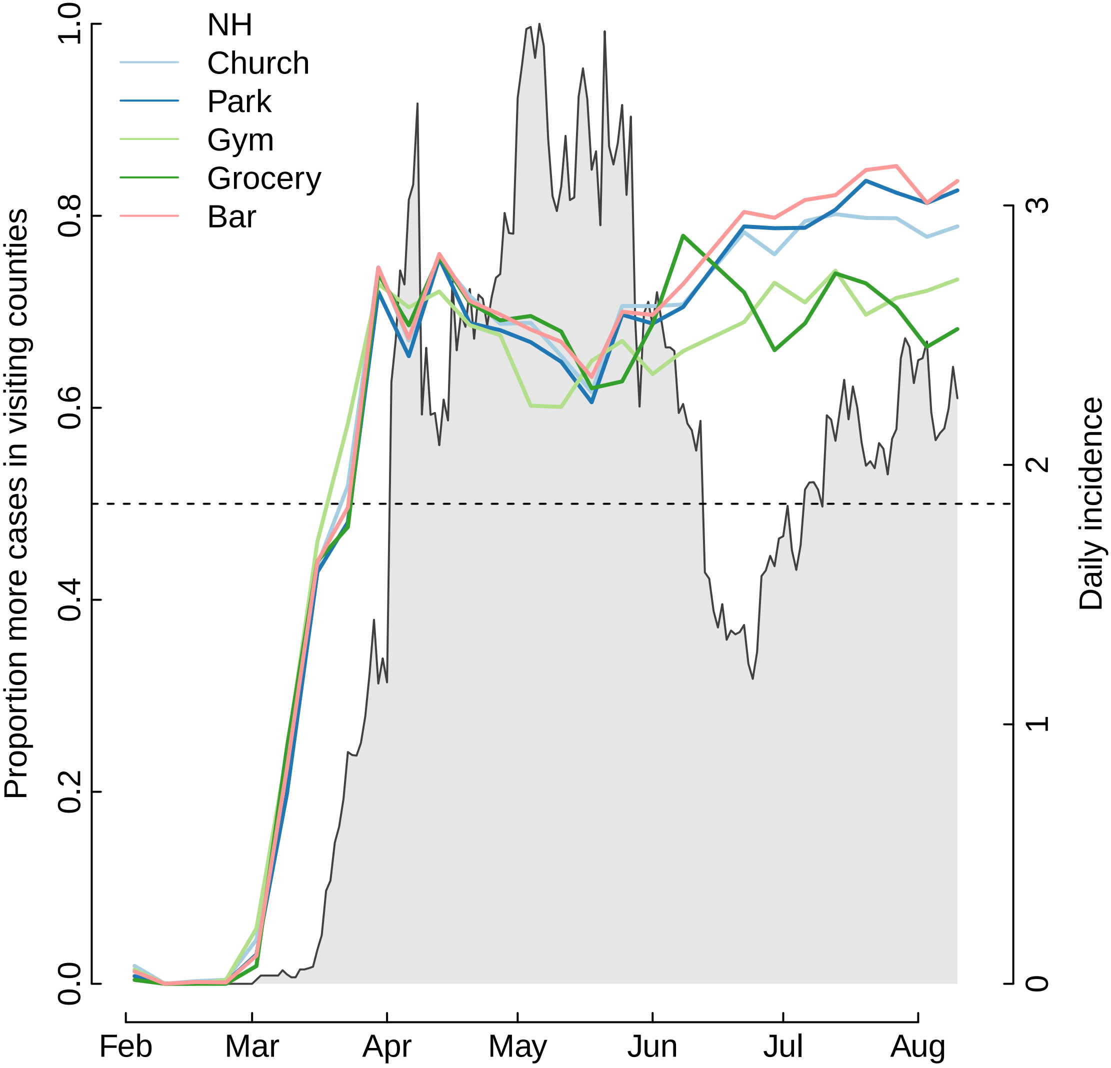

NH
Church
Park
Gym
Grocery
Bar
Proportion more cases in visiting counties
Daily incidence
0.0
0.2
0.4
0.6
0.8
1.0
0
1
2
3
Feb
Mar
Apr
May
Jun
Jul
Aug

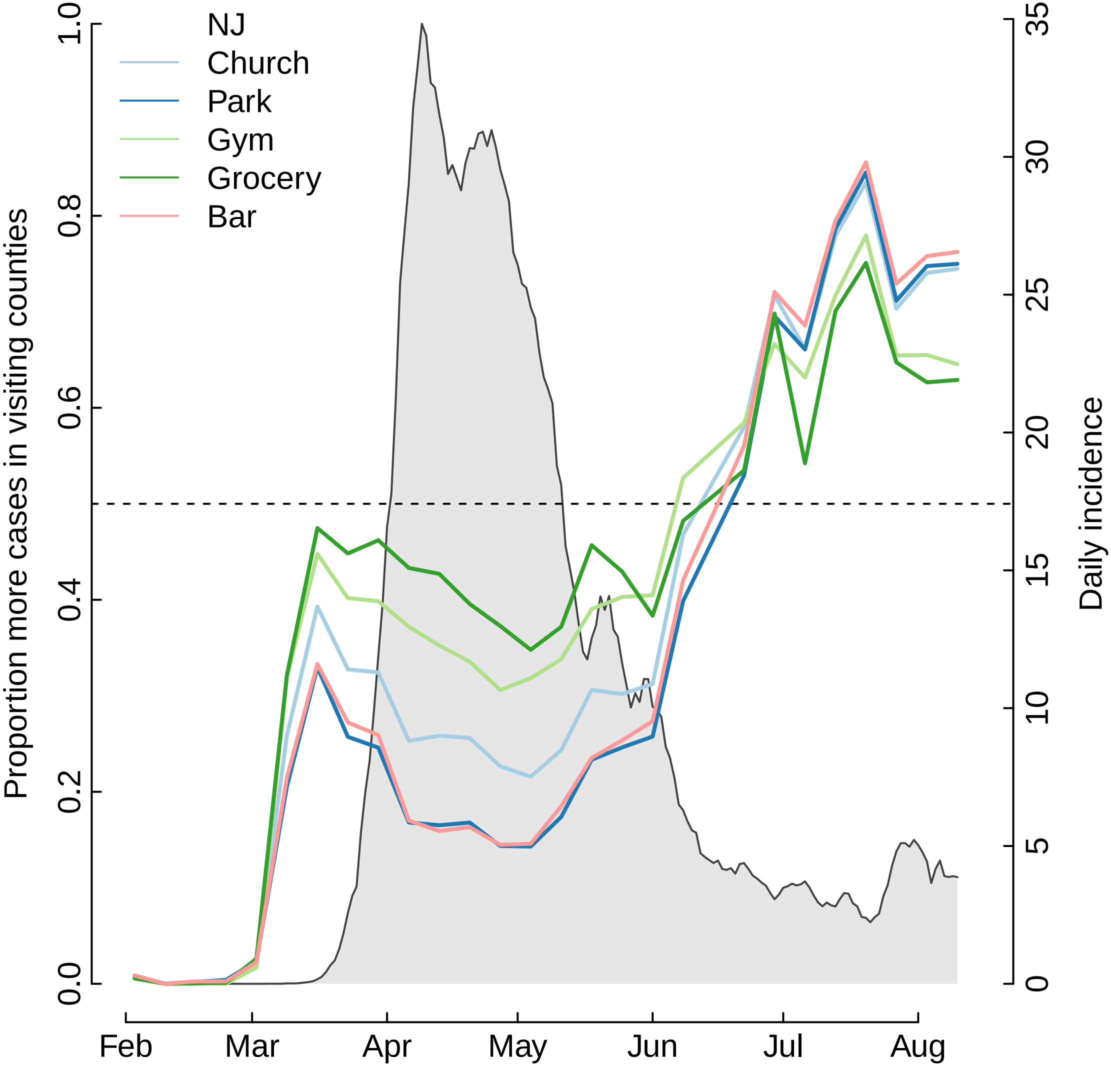

NJ
Church
Park
Gym
Grocery
Bar
Proportion more cases in visiting counties
Daily incidence
0.0
0.2
0.4
0.6
0.8
1.0
0
5
10
15
20
25
30
35
Feb
Mar
Apr
May
Jun
Jul
Aug

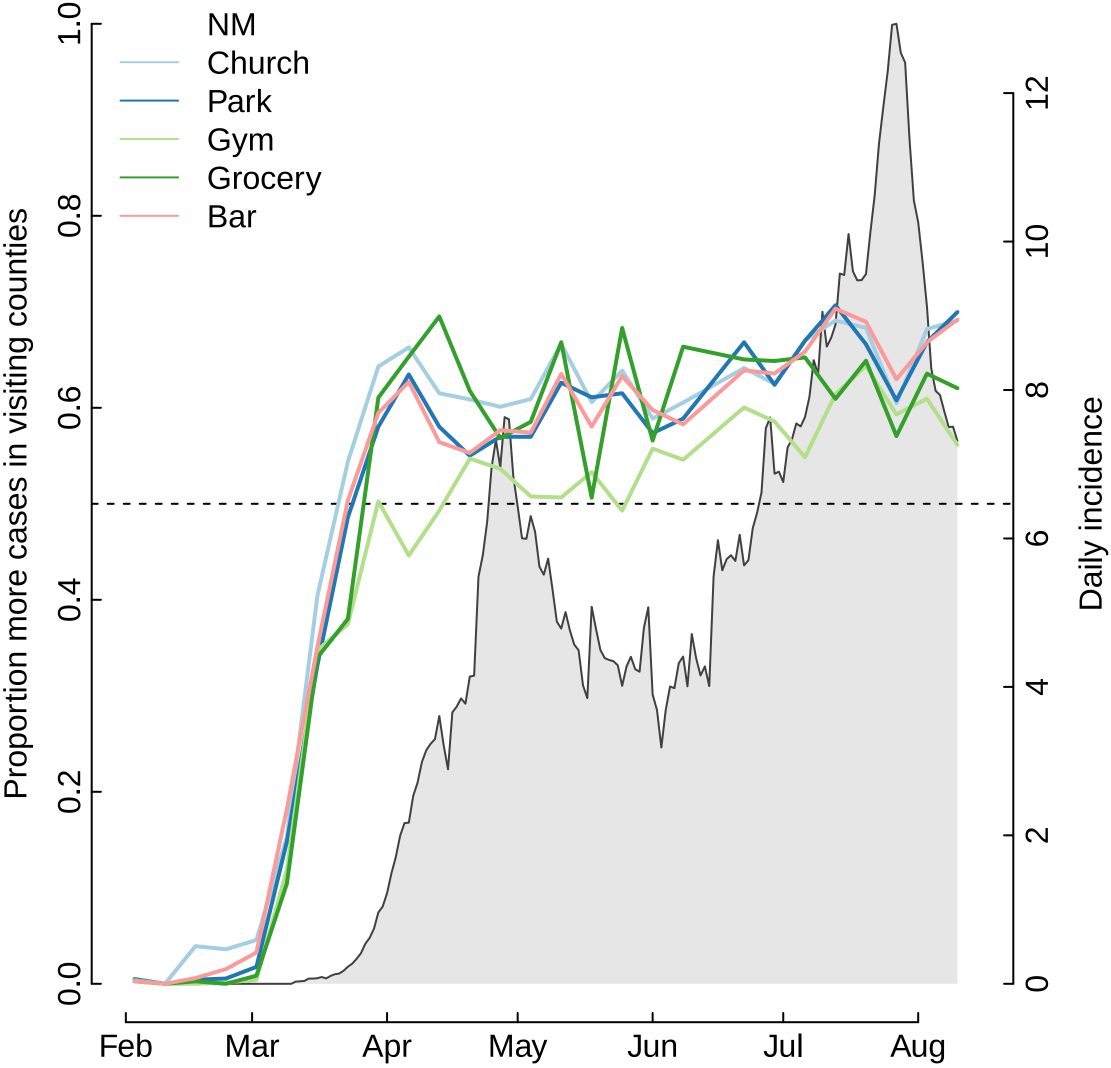

NM
Church
Park
Gym
Grocery
Bar
Proportion more cases in visiting counties
Daily incidence
0.0
0.2
0.4
0.6
0.8
1.0
0
2
4
6
8
10
12
Feb
Mar
Apr
May
Jun
Jul
Aug

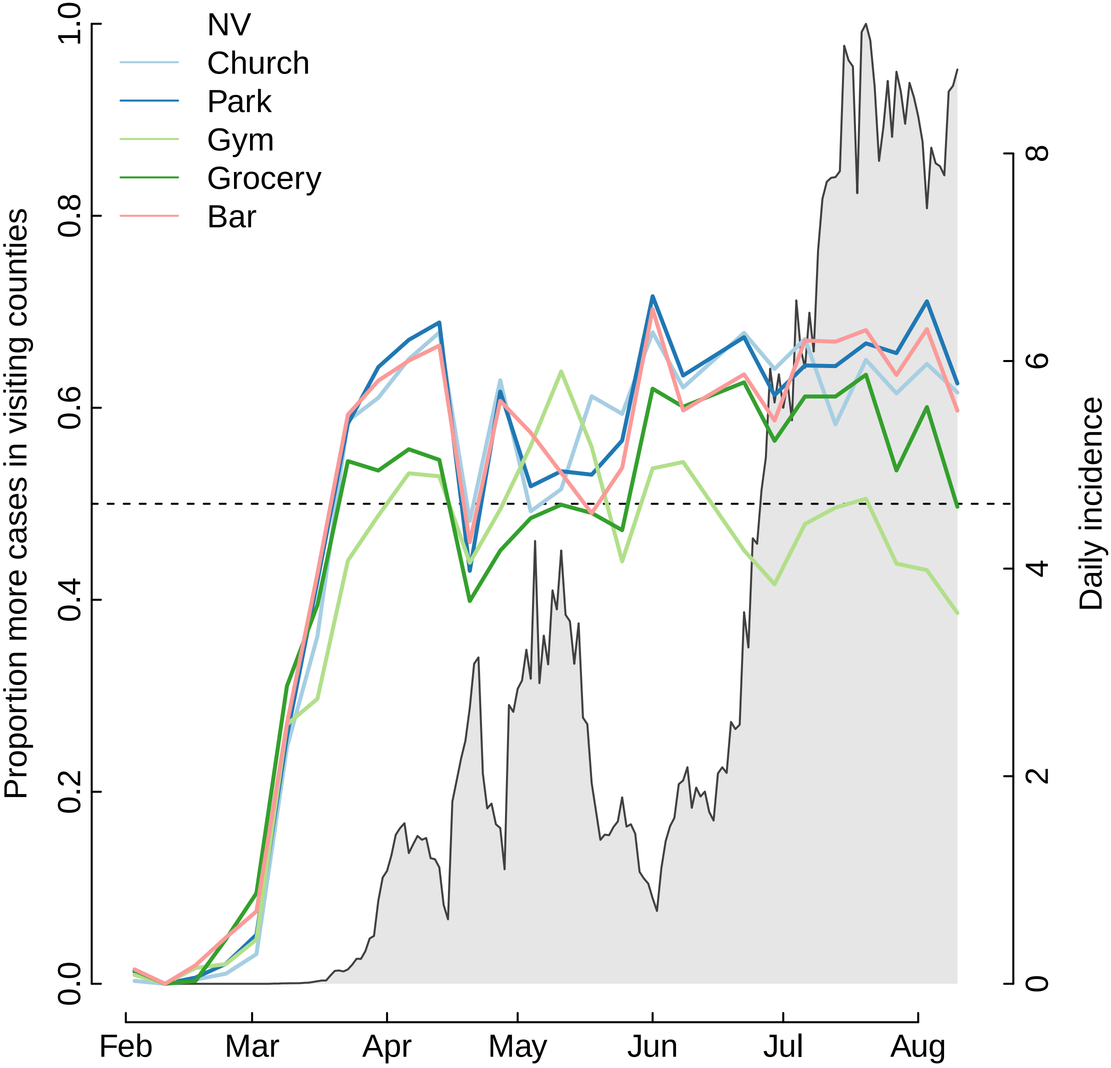

NV
Church
Park
Gym
Grocery
Bar
Proportion more cases in visiting counties
Daily incidence
0.0
0.2
0.4
0.6
0.8
1.0
0
2
4
6
8
Feb
Mar
Apr
May
Jun
Jul
Aug

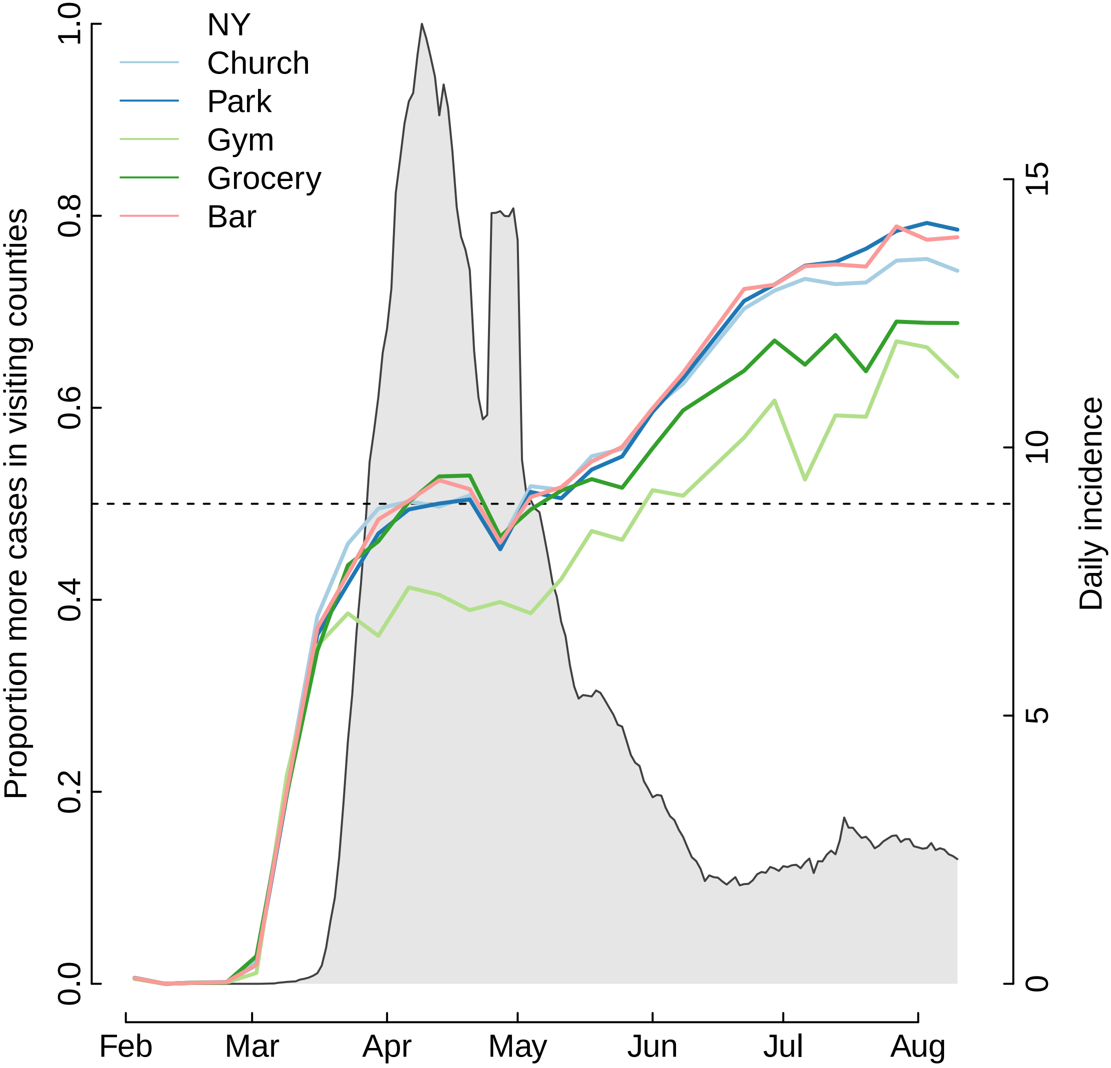

NY
Church
Park
Gym
Grocery
Bar
Proportion more cases in visiting counties
Daily incidence
0.0
0.2
0.4
0.6
0.8
1.0
0
5
10
15
Feb
Mar
Apr
May
Jun
Jul
Aug

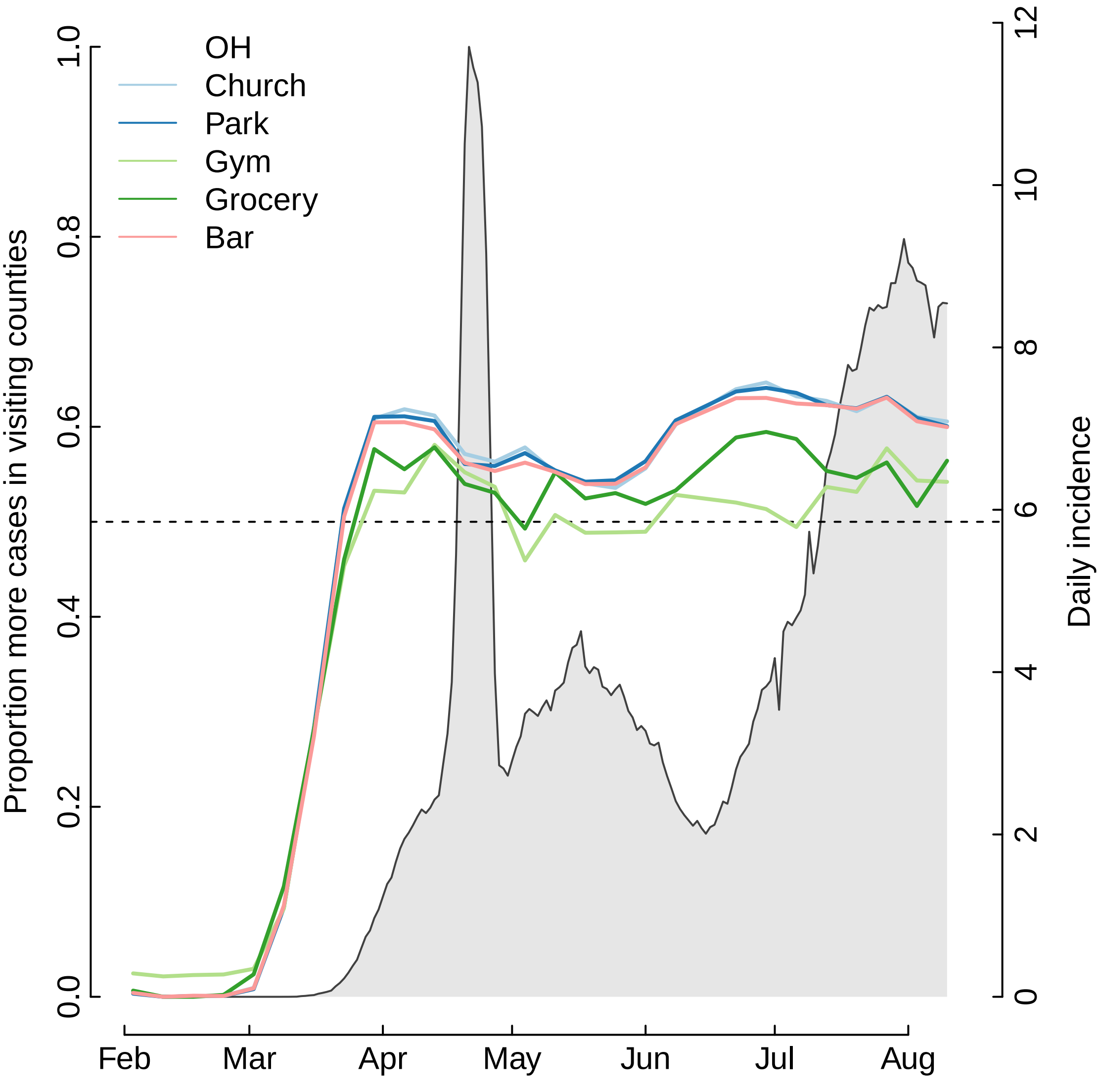

OH
Church
Park
Gym
Grocery
Bar
Proportion more cases in visiting counties
Daily incidence
0.0
0.2
0.4
0.6
0.8
1.0
0
2
4
6
8
10
12
Feb
Mar
Apr
May
Jun
Jul
Aug

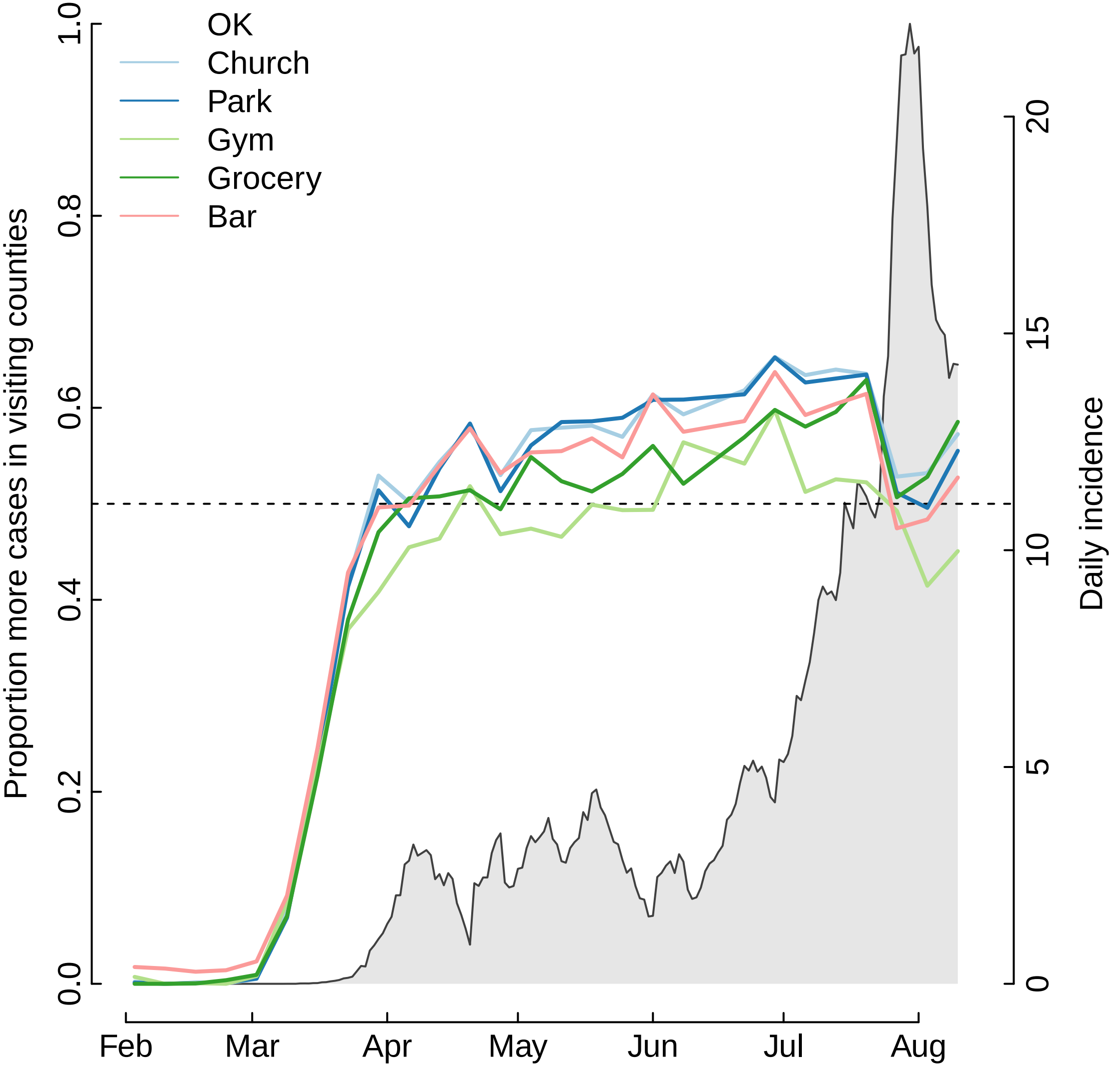

OK
Church
Park
Gym
Grocery
Bar
Proportion more cases in visiting counties
Daily incidence
0.0
0.2
0.4
0.6
0.8
1.0
0
5
10
15
20
Feb
Mar
Apr
May
Jun
Jul
Aug

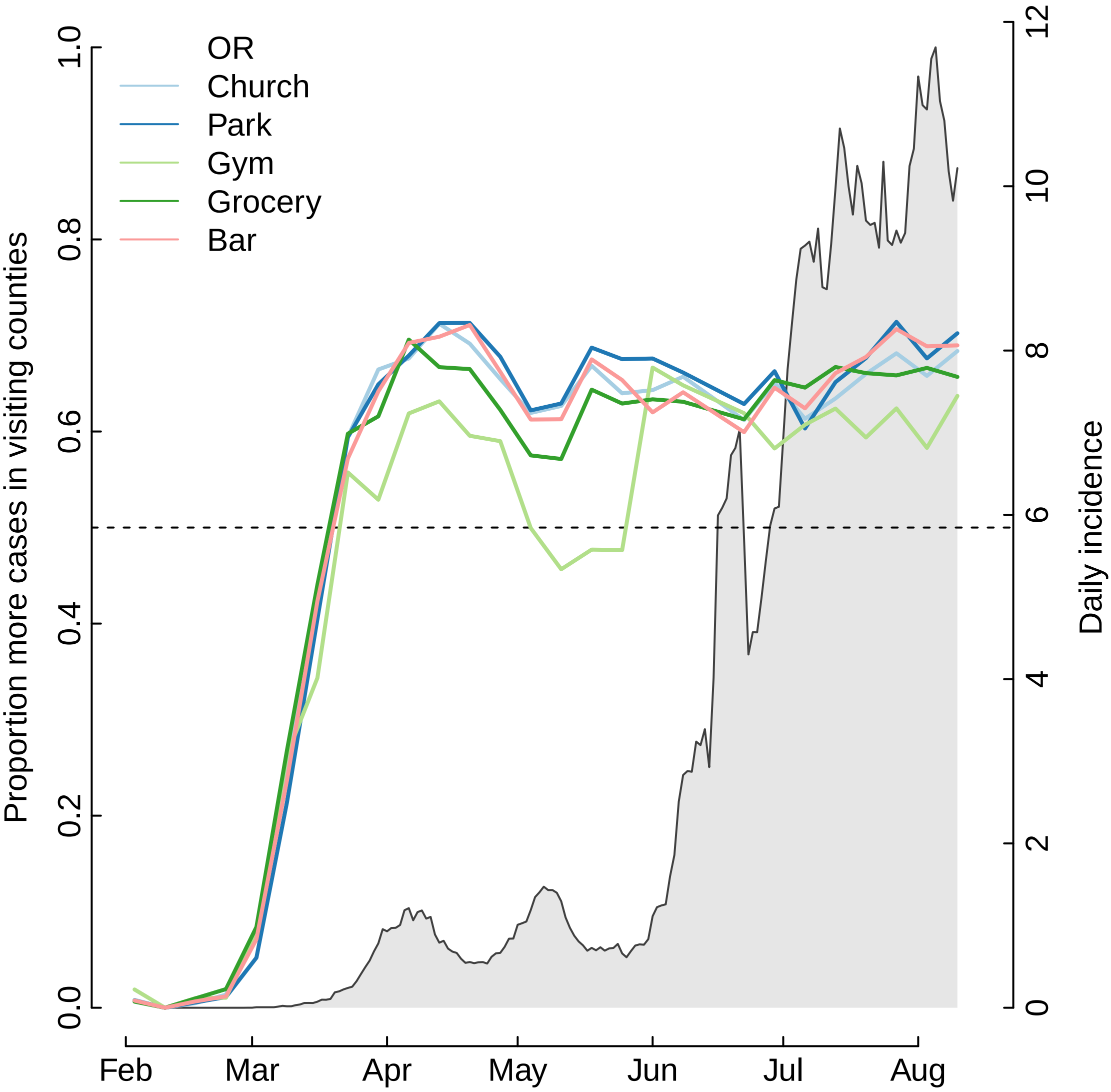

OR
Church
Park
Gym
Grocery
Bar
Proportion more cases in visiting counties
Daily incidence
0.0
0.2
0.4
0.6
0.8
1.0
0
2
4
6
8
10
12
Feb
Mar
Apr
May
Jun
Jul
Aug

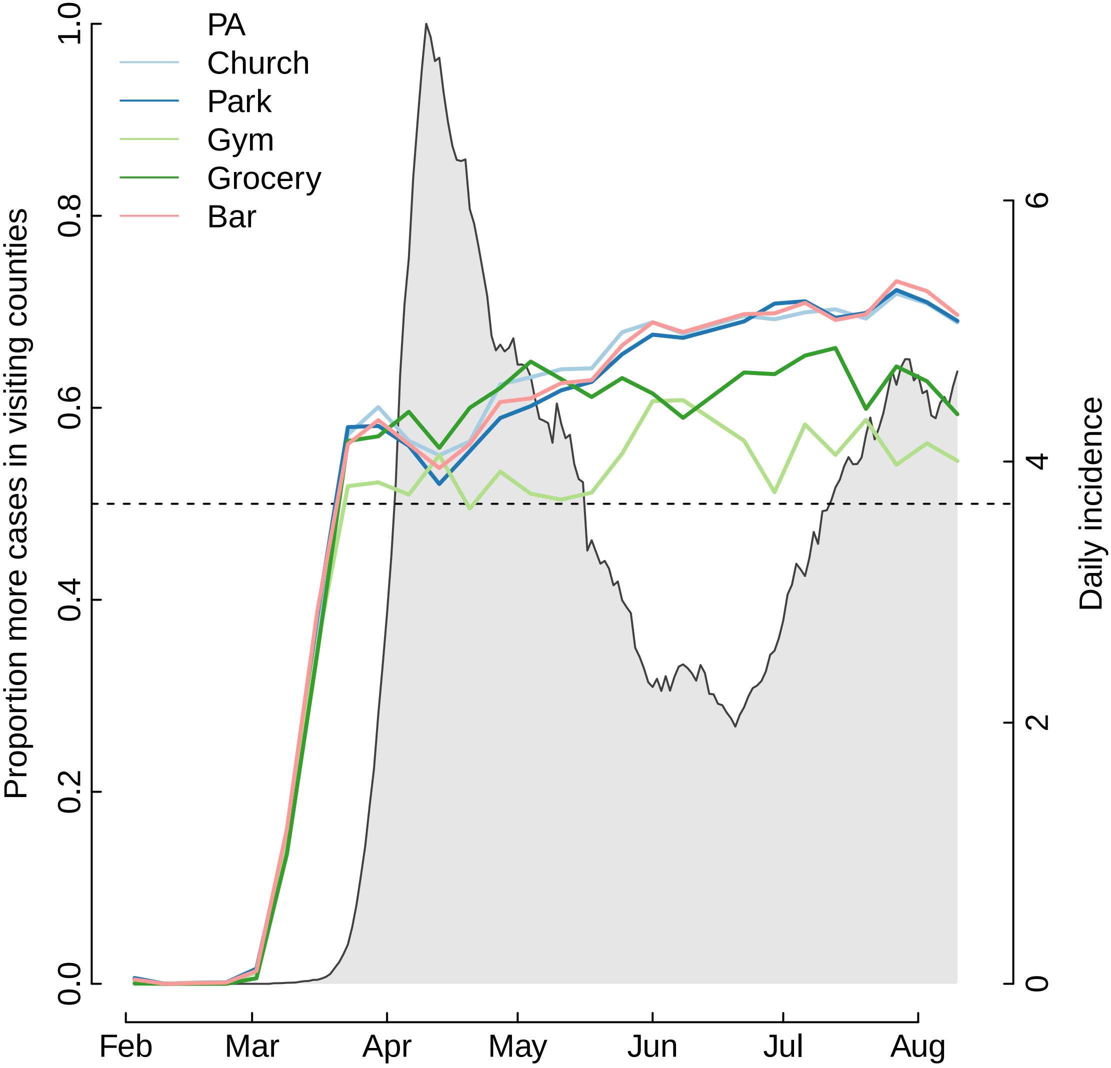

PA
Church
Park
Gym
Grocery
Bar
Proportion more cases in visiting counties
Daily incidence
0.0
0.2
0.4
0.6
0.8
1.0
0
2
4
6
Feb
Mar
Apr
May
Jun
Jul
Aug

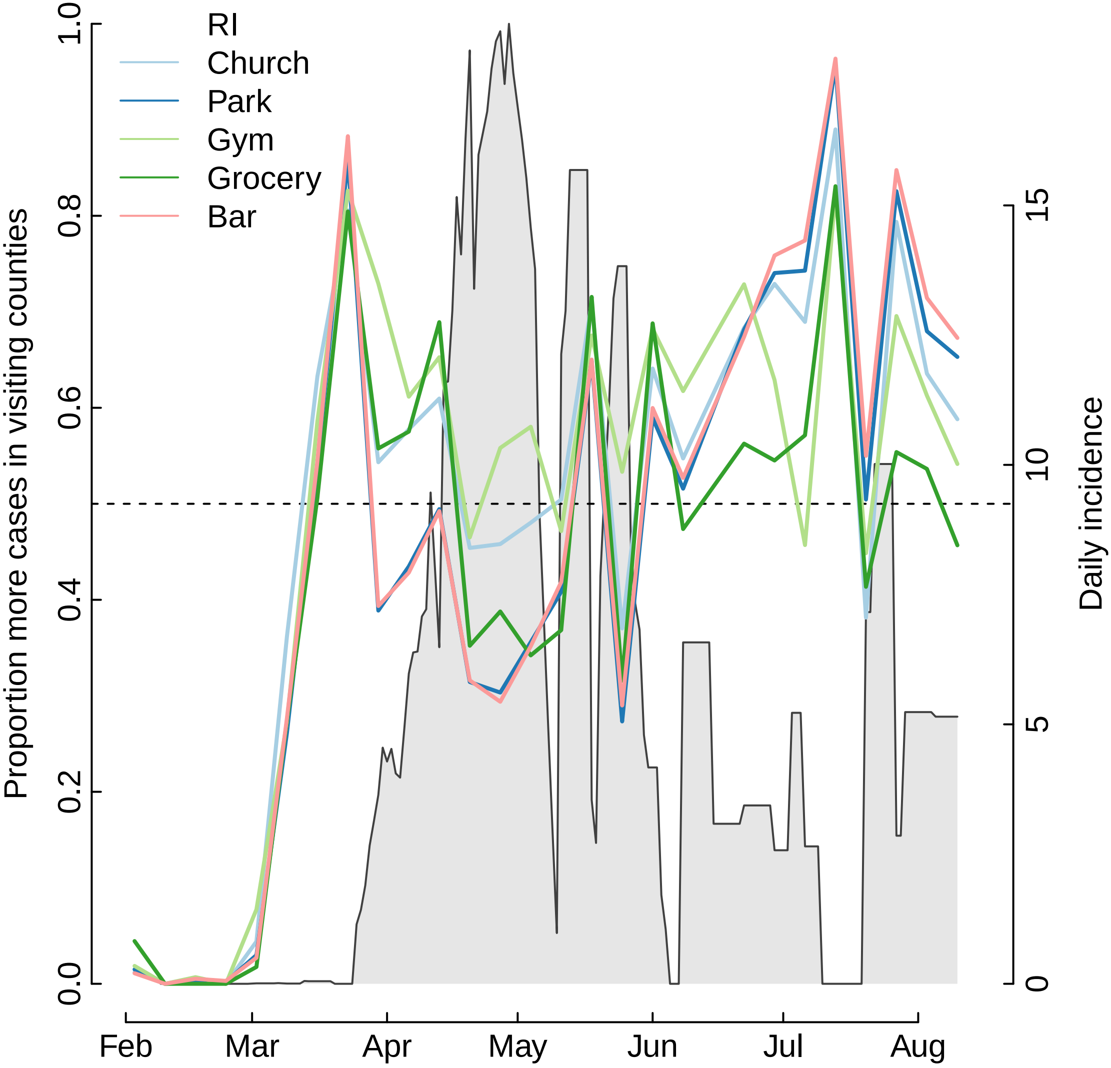
RI
Church
Park
Gym
Grocery
Bar
Proportion more cases in visiting counties
Daily incidence
0.0
0.2
0.4
0.6
0.8
1.0
0
5
10
15
Feb
Mar
Apr
May
Jun
Jul
Aug

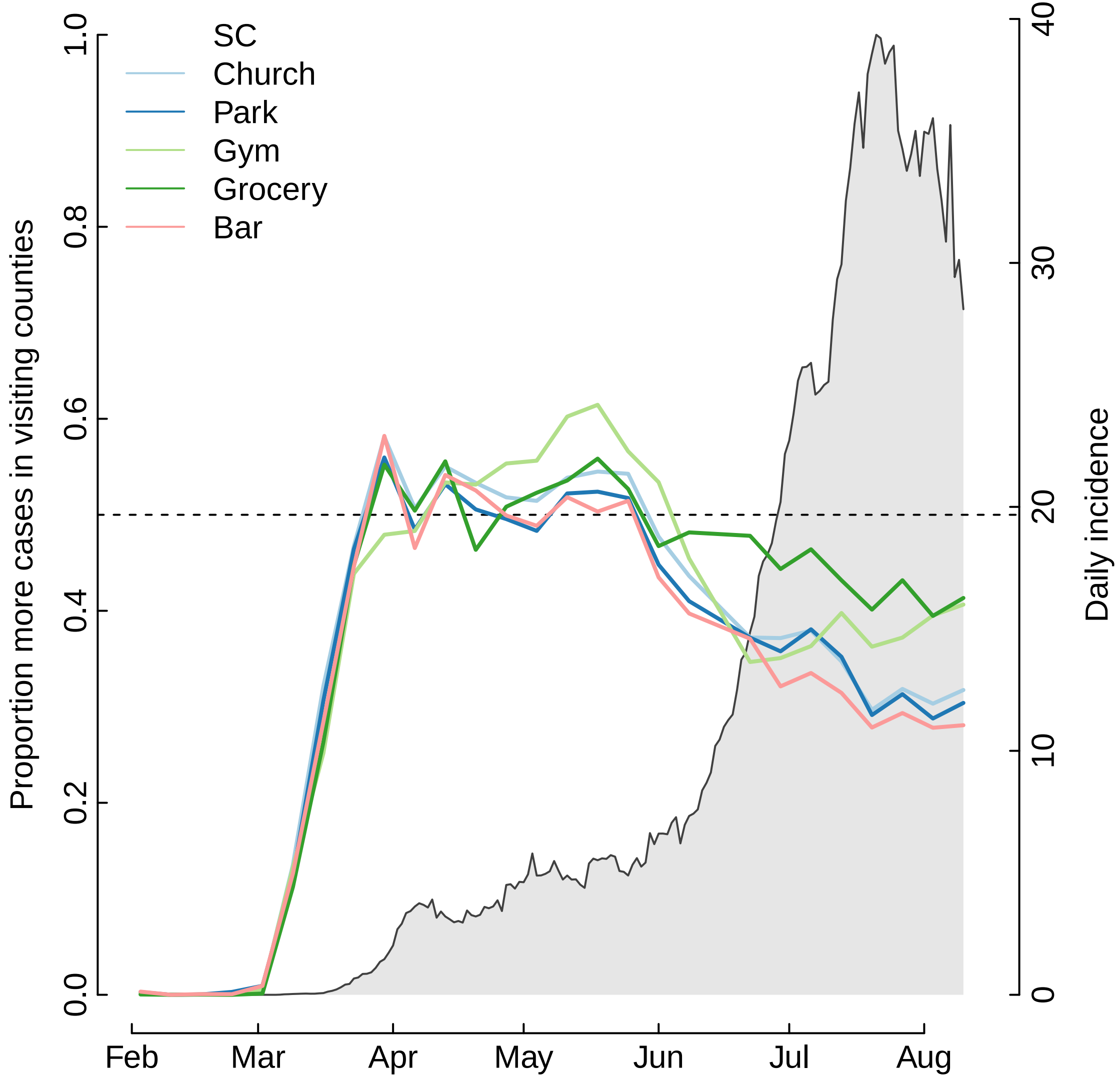

SC
Church
Park
Gym
Grocery
Bar
Proportion more cases in visiting counties
Daily incidence
0.0
0.2
0.4
0.6
0.8
1.0
0
10
20
30
40
Feb
Mar
Apr
May
Jun
Jul
Aug

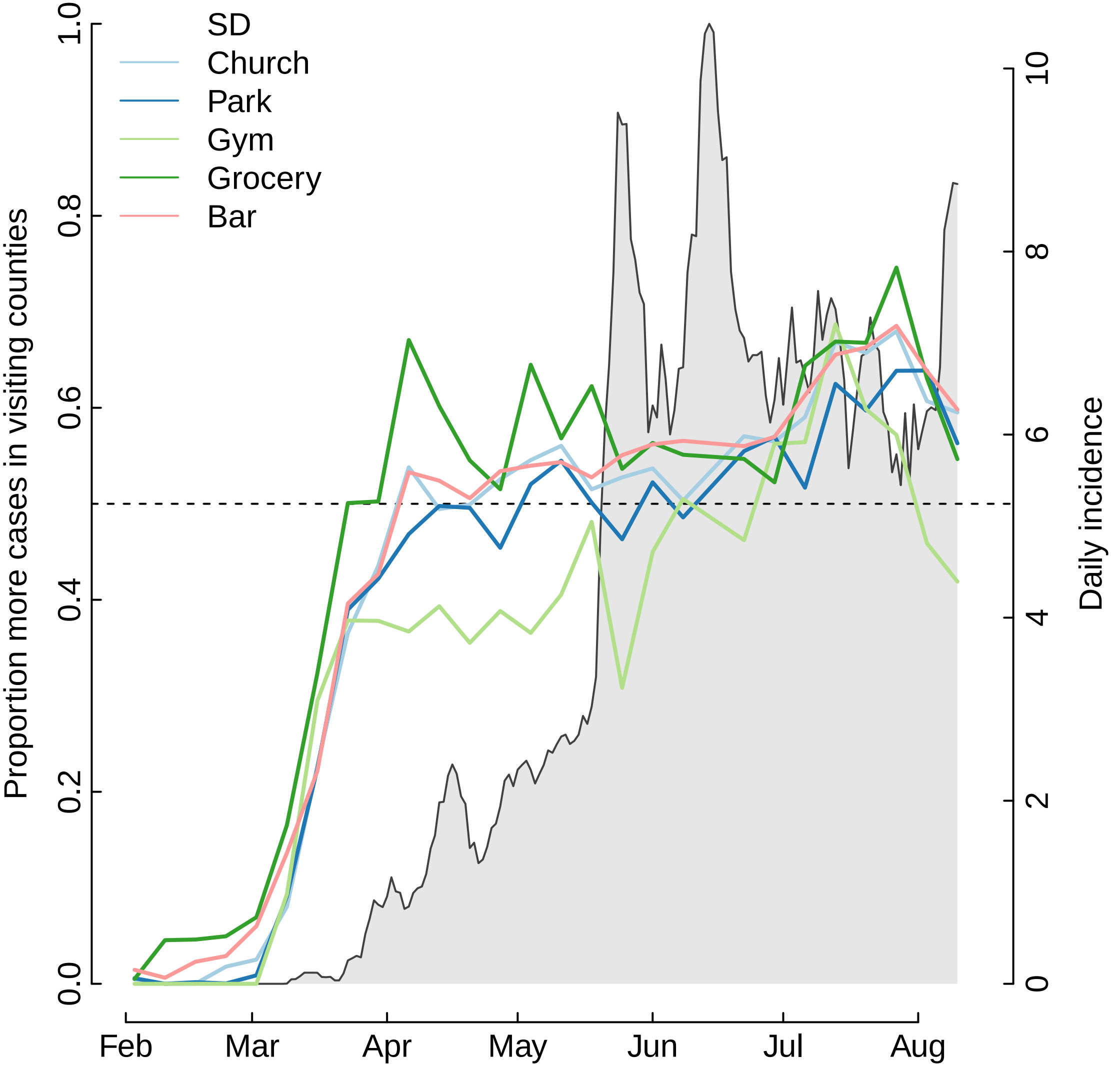
SD
Church
Park
Gym
Grocery
Bar
Proportion more cases in visiting counties
Daily incidence
0.0
0.2
0.4
0.6
0.8
1.0
0
2
4
6
8
10
Feb
Mar
Apr
May
Jun
Jul
Aug

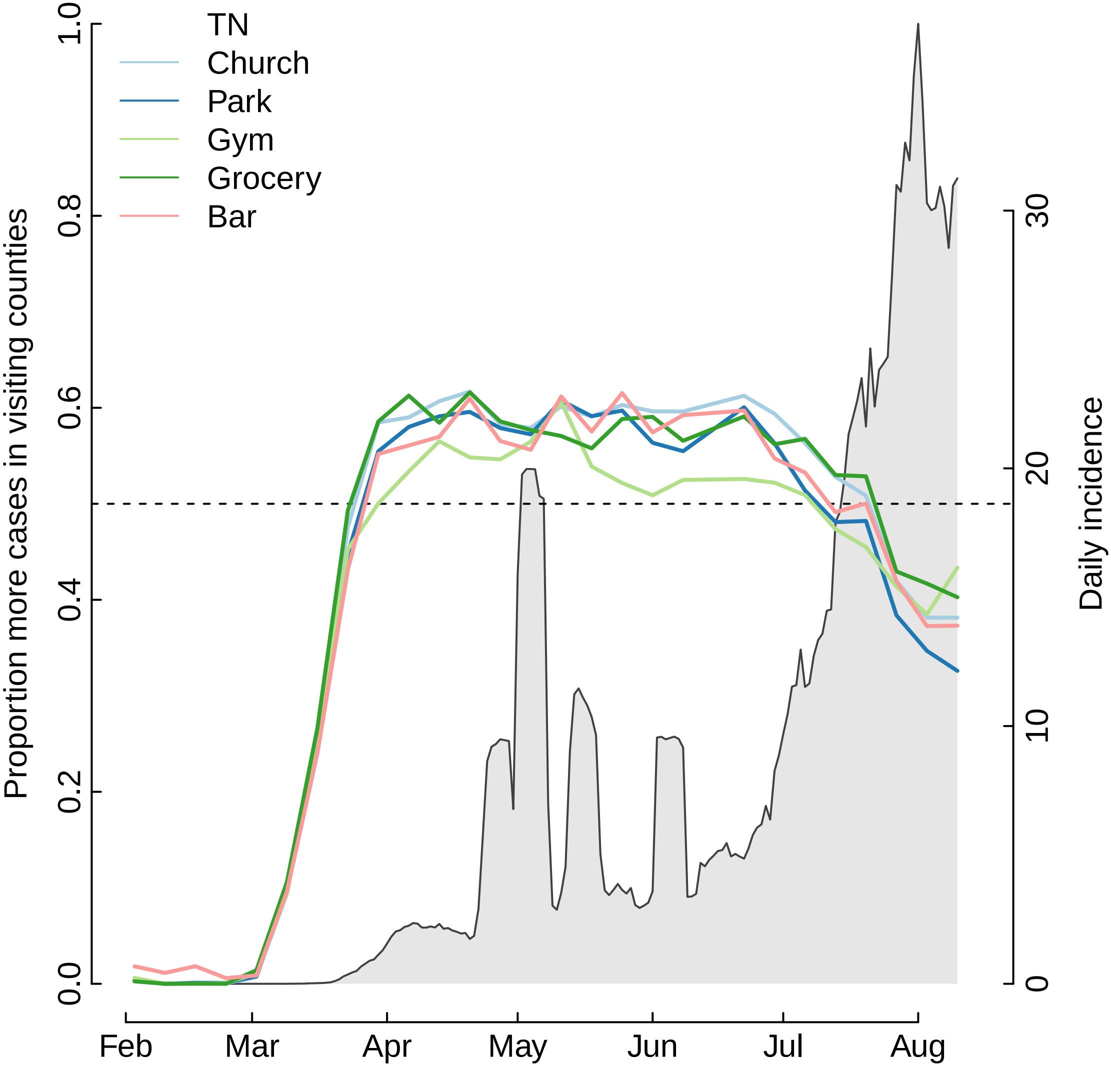
TN
Church
Park
Gym
Grocery
Bar
Proportion more cases in visiting counties
Daily incidence
0.0
0.2
0.4
0.6
0.8
1.0
0
10
20
30
Feb
Mar
Apr
May
Jun
Jul
Aug

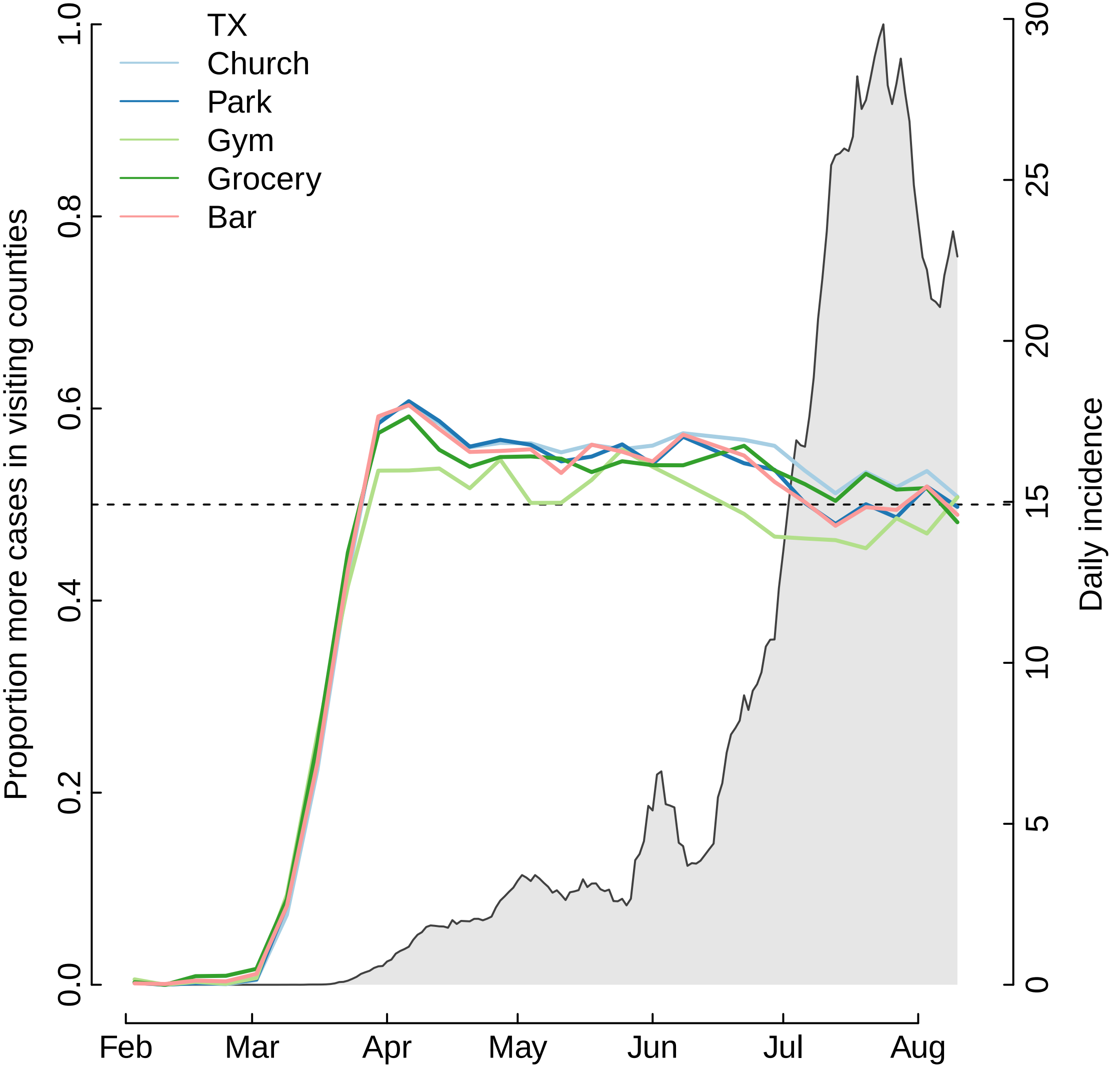

TX
Church
Park
Gym
Grocery
Bar
Proportion more cases in visiting counties
Daily incidence
0.0
0.2
0.4
0.6
0.8
1.0
0
5
10
15
20
25
30
Feb
Mar
Apr
May
Jun
Jul
Aug

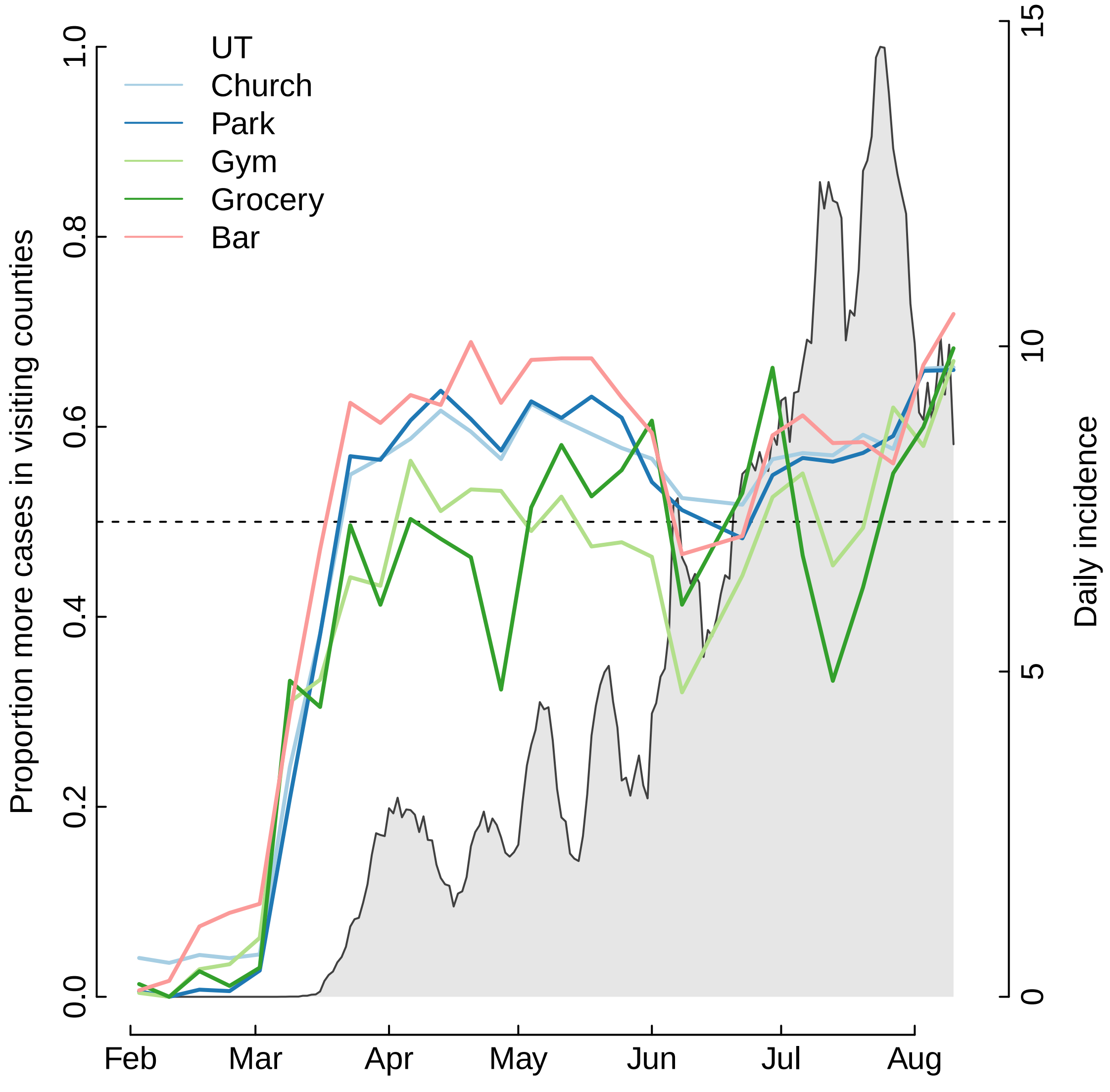

UT
Church
Park
Gym
Grocery
Bar
Proportion more cases in visiting counties
Daily incidence
0.0
0.2
0.4
0.6
0.8
1.0
0
5
10
15
Feb
Mar
Apr
May
Jun
Jul
Aug

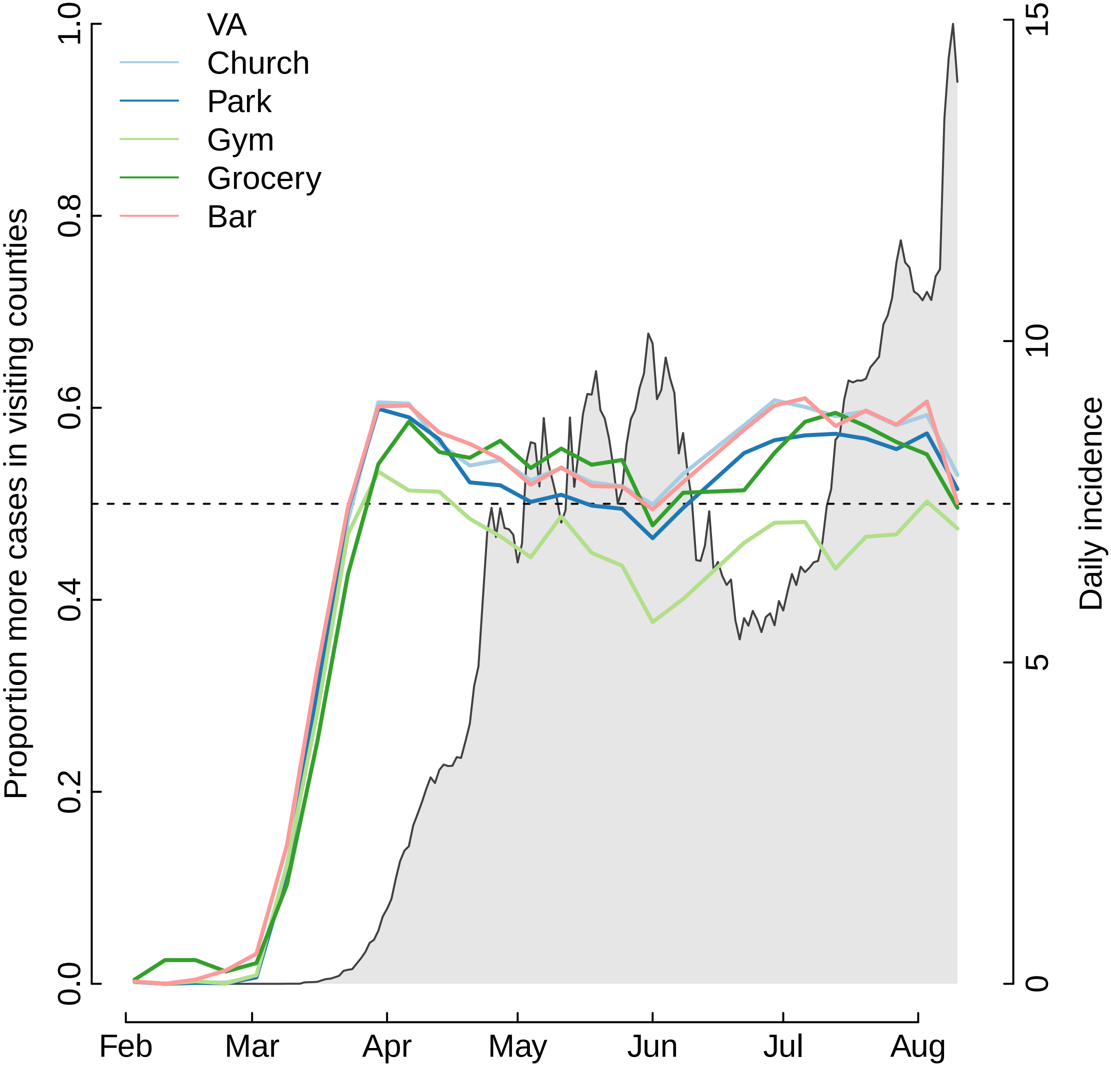

VA
Church
Park
Gym
Grocery
Bar
Proportion more cases in visiting counties
Daily incidence
0.0
0.2
0.4
0.6
0.8
1.0
0
5
10
15
Feb
Mar
Apr
May
Jun
Jul
Aug

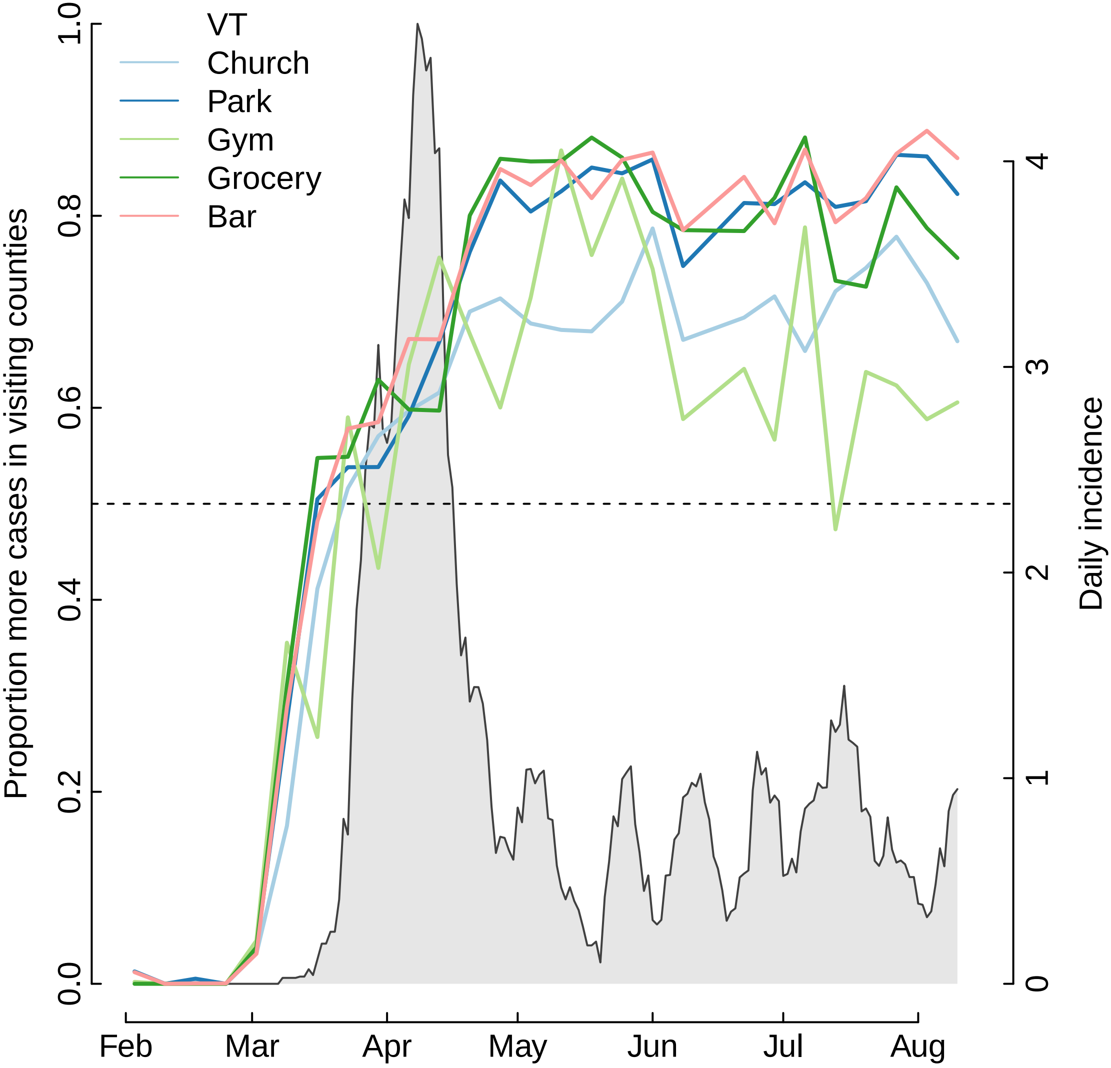

VT
Church
Park
Gym
Grocery
Bar
Proportion more cases in visiting counties
Daily incidence
0.0
0.2
0.4
0.6
0.8
1.0
0
1
2
3
4
Feb
Mar
Apr
May
Jun
Jul
Aug

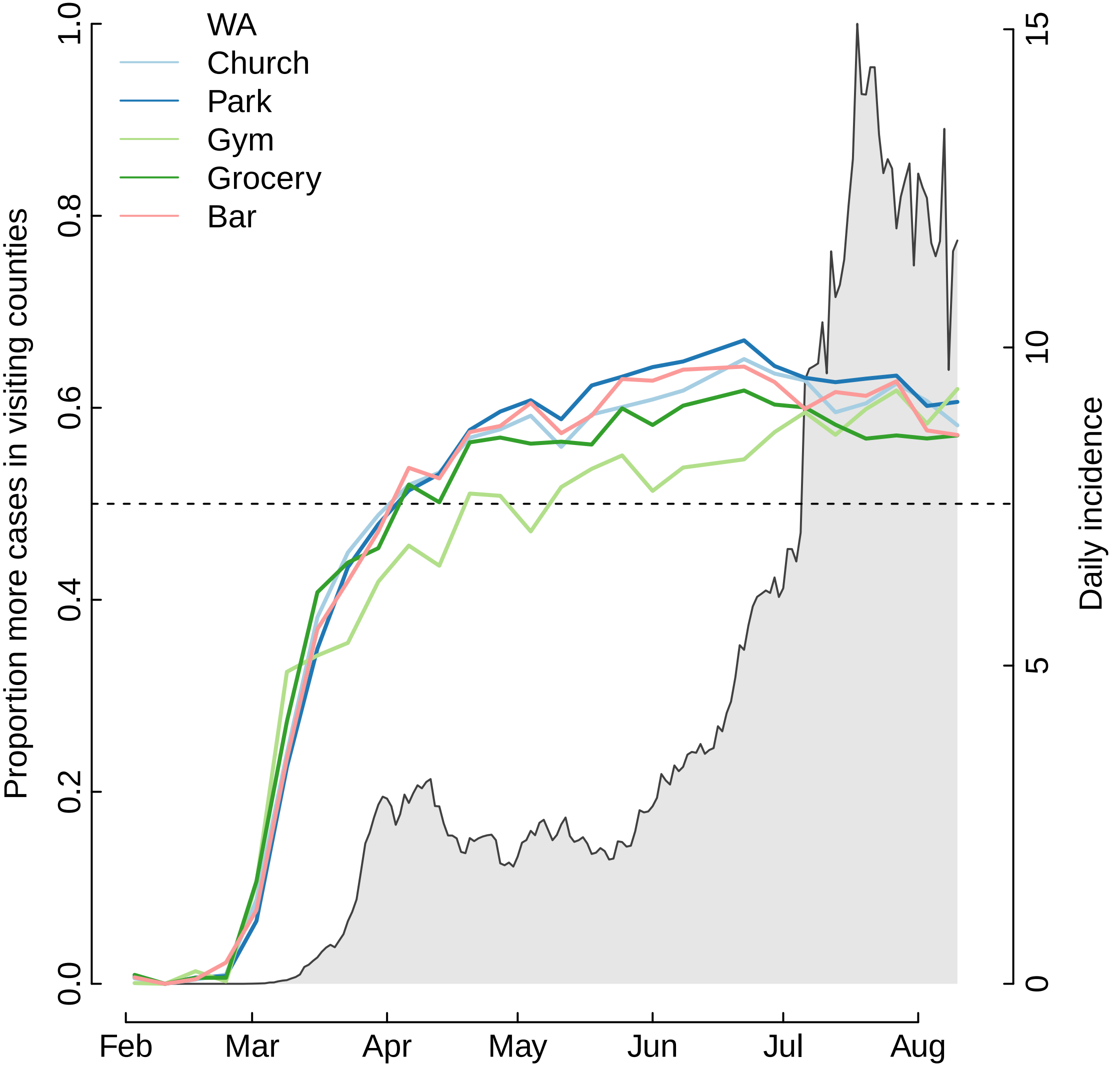

WA
Church
Park
Gym
Grocery
Bar
Proportion more cases in visiting counties
Daily incidence
0.0
0.2
0.4
0.6
0.8
1.0
0
5
10
15
Feb
Mar
Apr
May
Jun
Jul
Aug

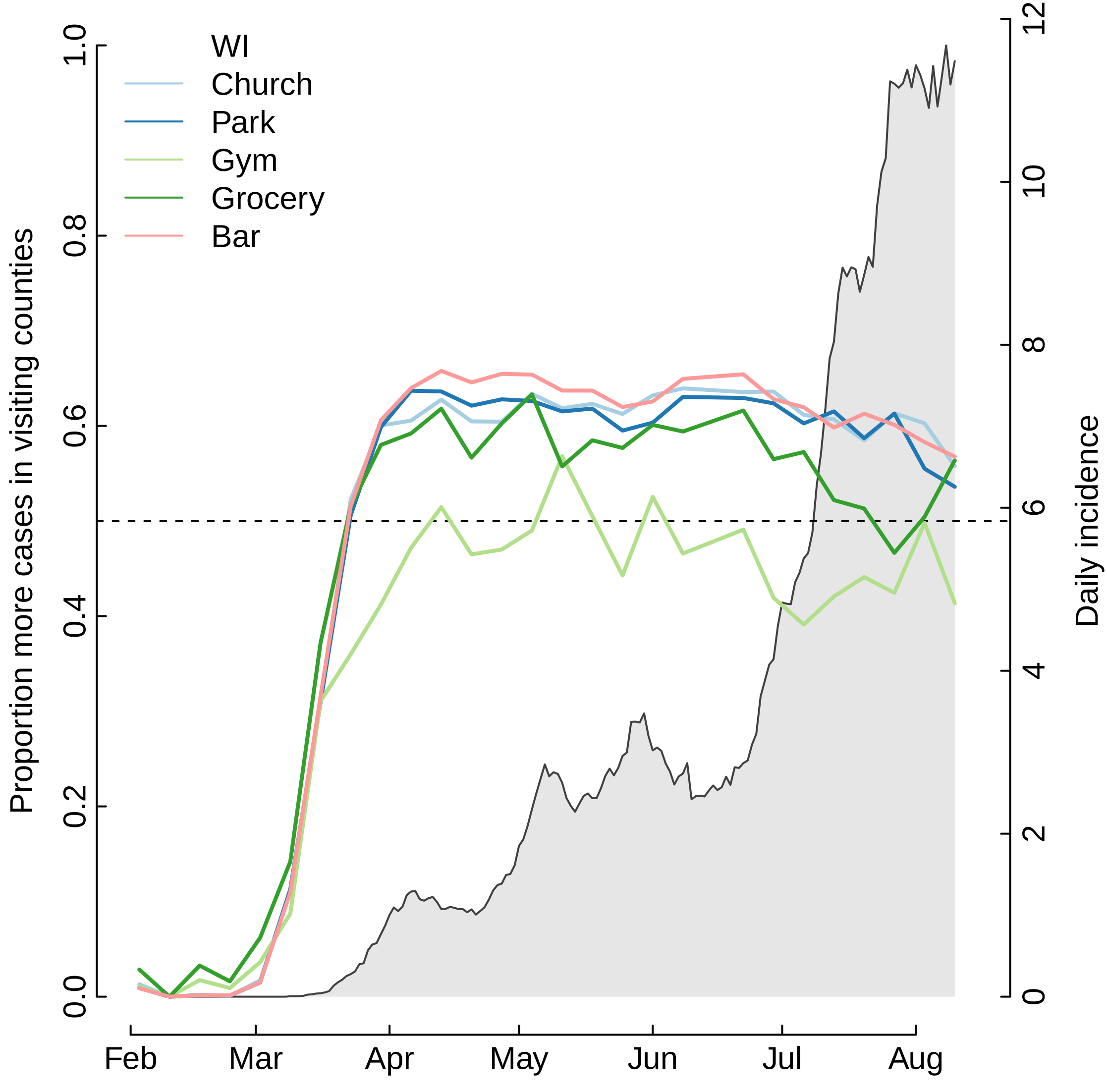

WI
Church
Park
Gym
Grocery
Bar
Proportion more cases in visiting counties
Daily incidence
0.0
0.2
0.4
0.6
0.8
1.0
0
2
4
6
8
10
12
Feb
Mar
Apr
May
Jun
Jul
Aug

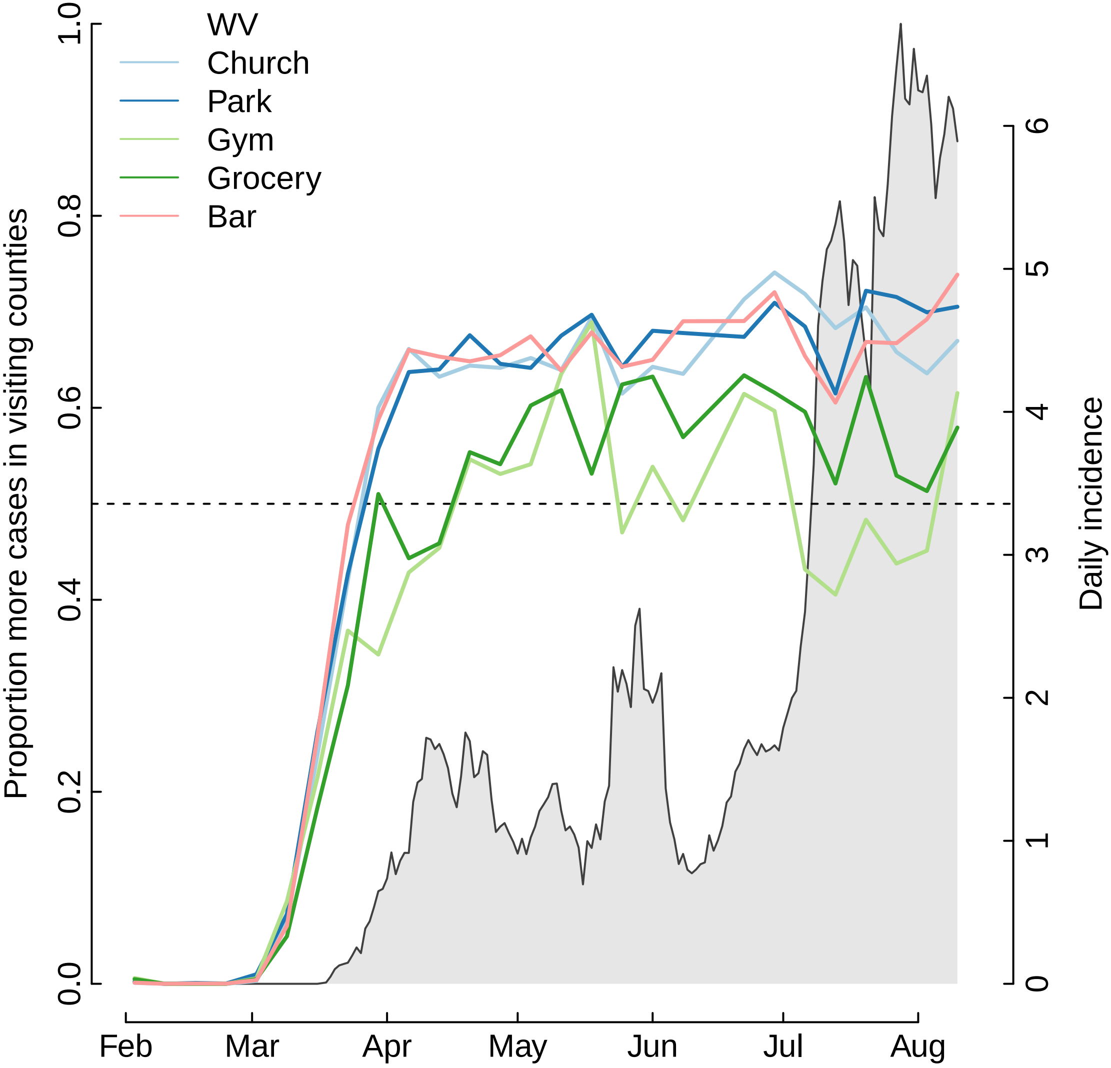

WV
Church
Park
Gym
Grocery
Bar
Proportion more cases in visiting counties
Daily incidence
0.0
0.2
0.4
0.6
0.8
1.0
0
1
2
3
4
5
6
Feb
Mar
Apr
May
Jun
Jul
Aug

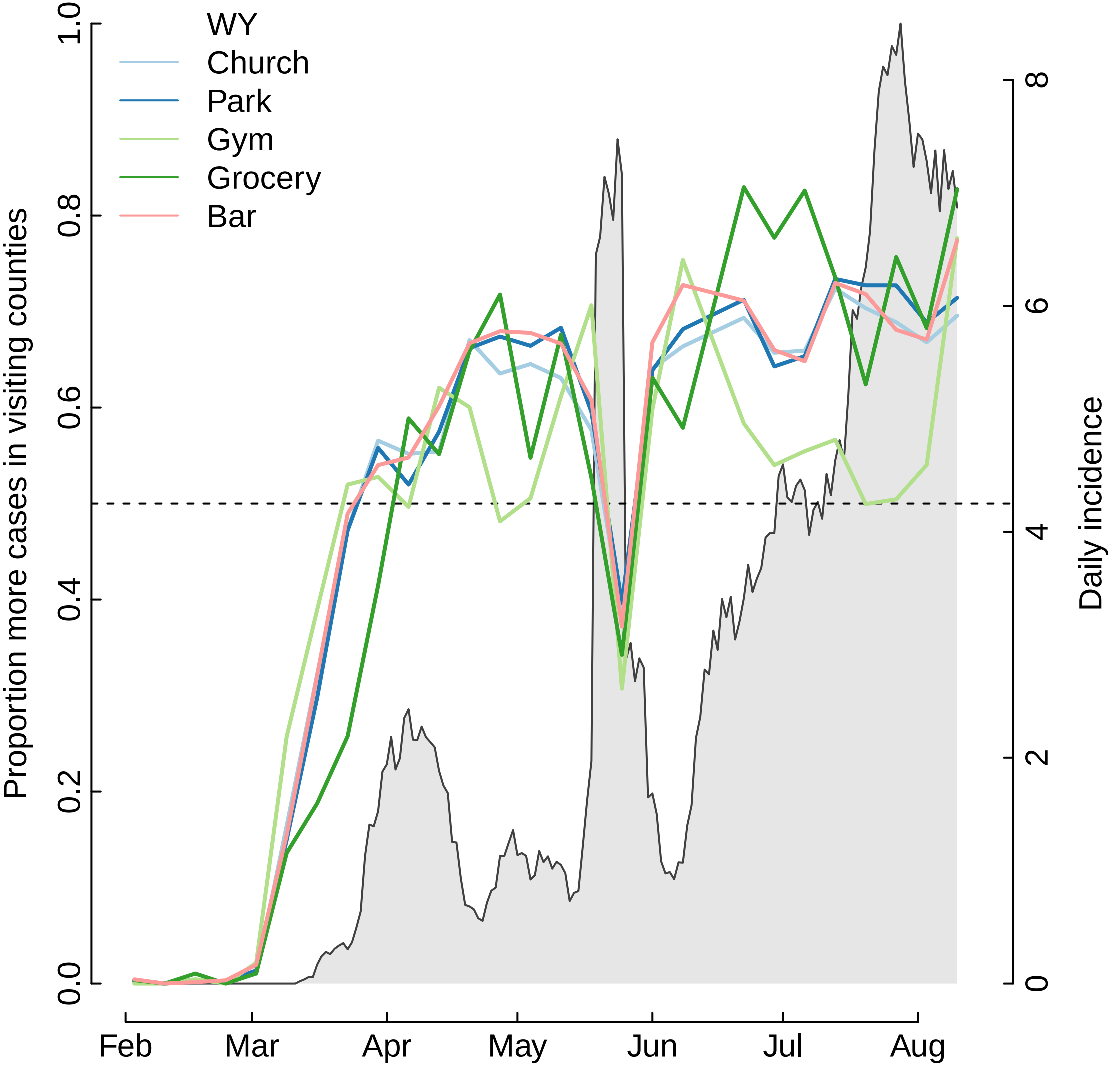

WY
Church
Park
Gym
Grocery
Bar
Proportion more cases in visiting counties
Daily incidence
0.0
0.2
0.4
0.6
0.8
1.0
0
2
4
6
8
Feb
Mar
Apr
May
Jun
Jul
Aug
