## Supplementary material for "The unintended consequences of inconsistent pandemic control policies": Cross-wavelets - groceries

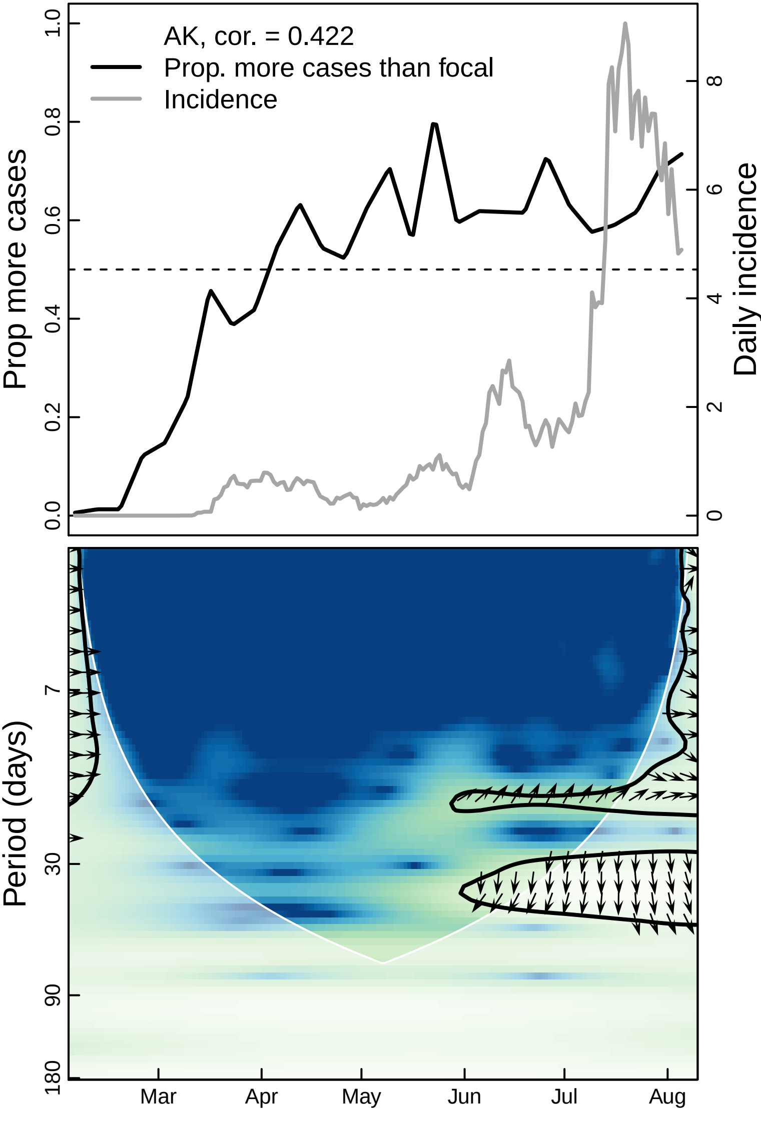

AK, cor. = 0.422
Prop. more cases than focal
Incidence
Prop more cases
Daily incidence
0.0
0.2
0.4
0.6
0.8
1.0
0
2
4
6
8
Period (days)
7
30
90
180
Mar
Apr
May
Jun
Jul
Aug

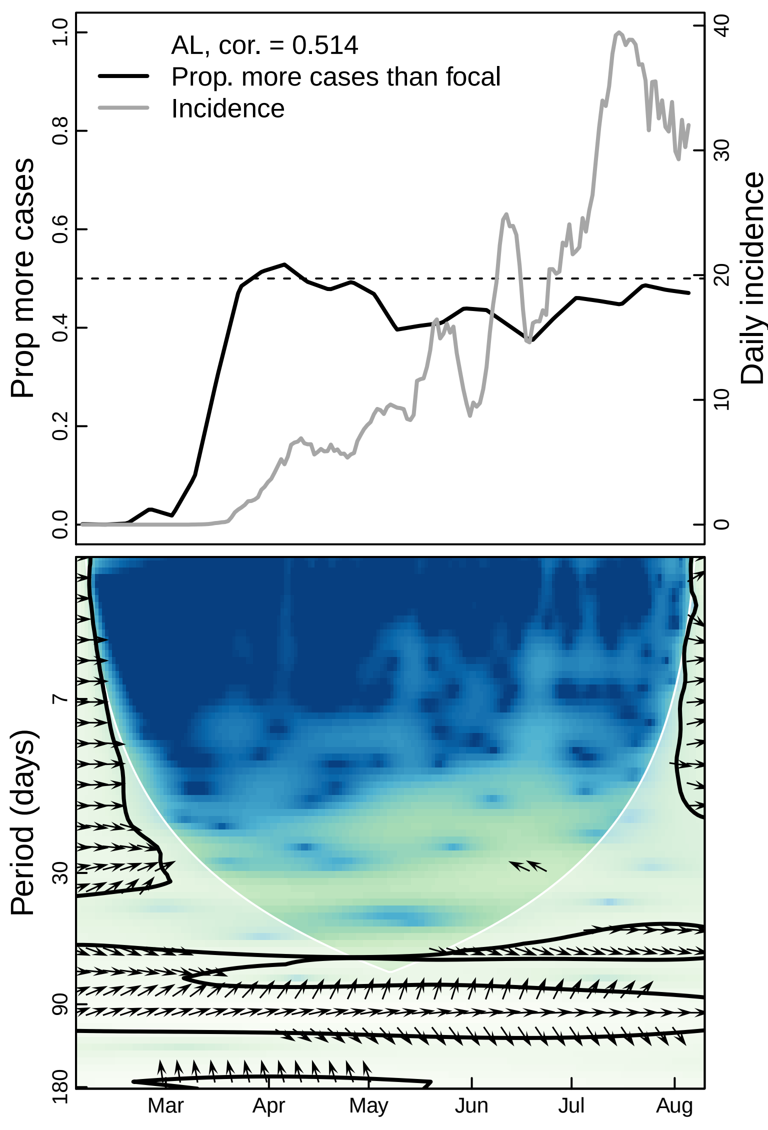
AL, cor. = 0.514
Prop. more cases than focal
Incidence
Prop more cases
Daily incidence
1.0
0.8
0.6
0.4
0.2
0.0
40
30
20
10
0
Period (days)
7
30
90
180
Mar
Apr
May
Jun
Jul
Aug

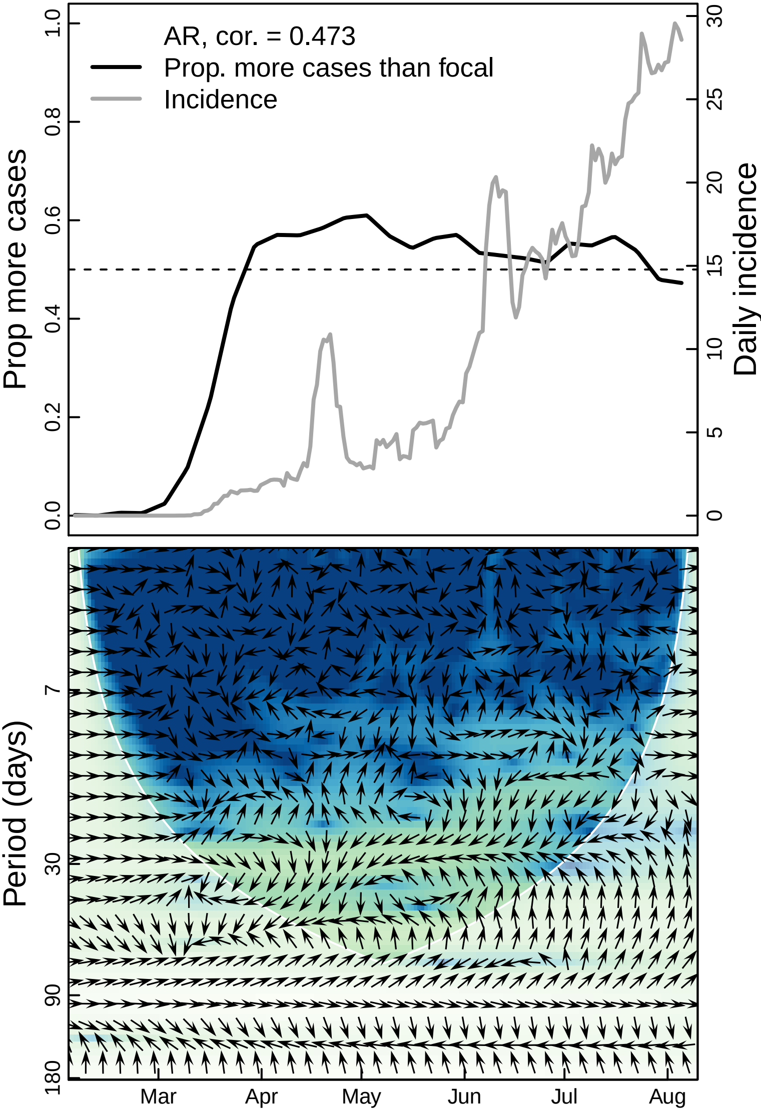
AR, cor. = 0.473
Prop. more cases than focal
Incidence
Prop more cases
Daily incidence
Period (days)
Mar
Apr
May
Jun
Jul
Aug

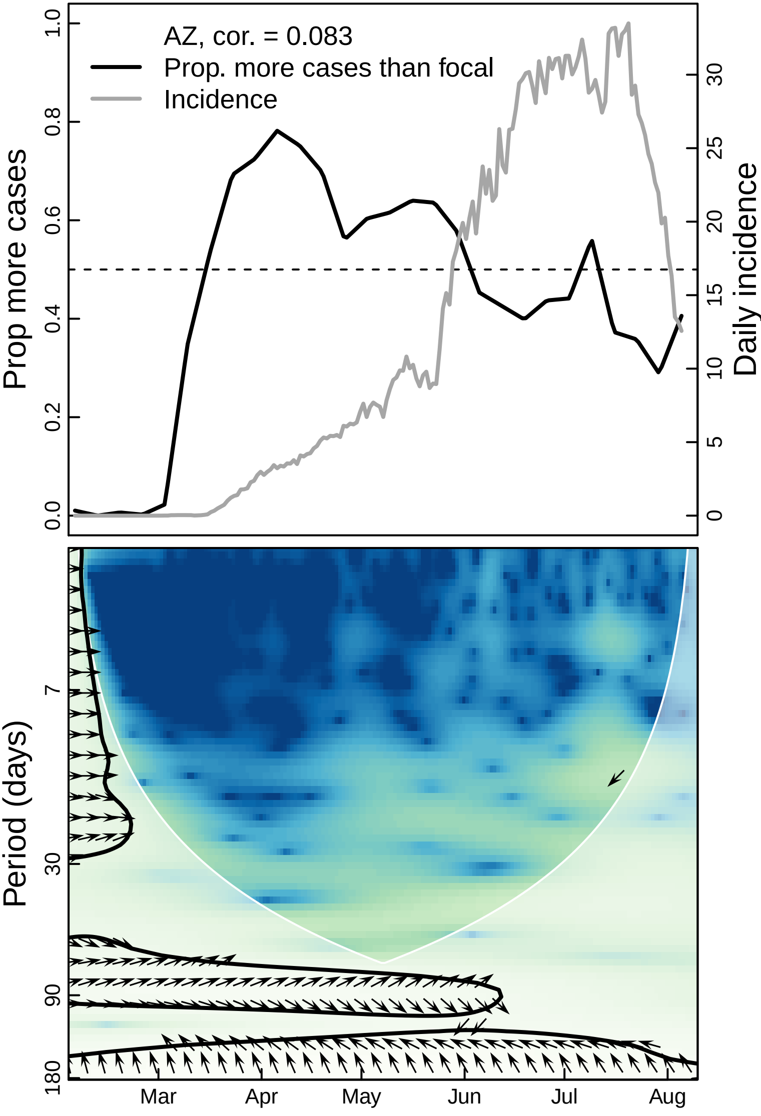

AZ, cor. = 0.083
Prop. more cases than focal
Incidence
Prop more cases
Daily incidence
0.0
0.2
0.4
0.6
0.8
1.0
0
5
10
15
20
25
30
Period (days)
7
30
90
180
Mar
Apr
May
Jun
Jul
Aug

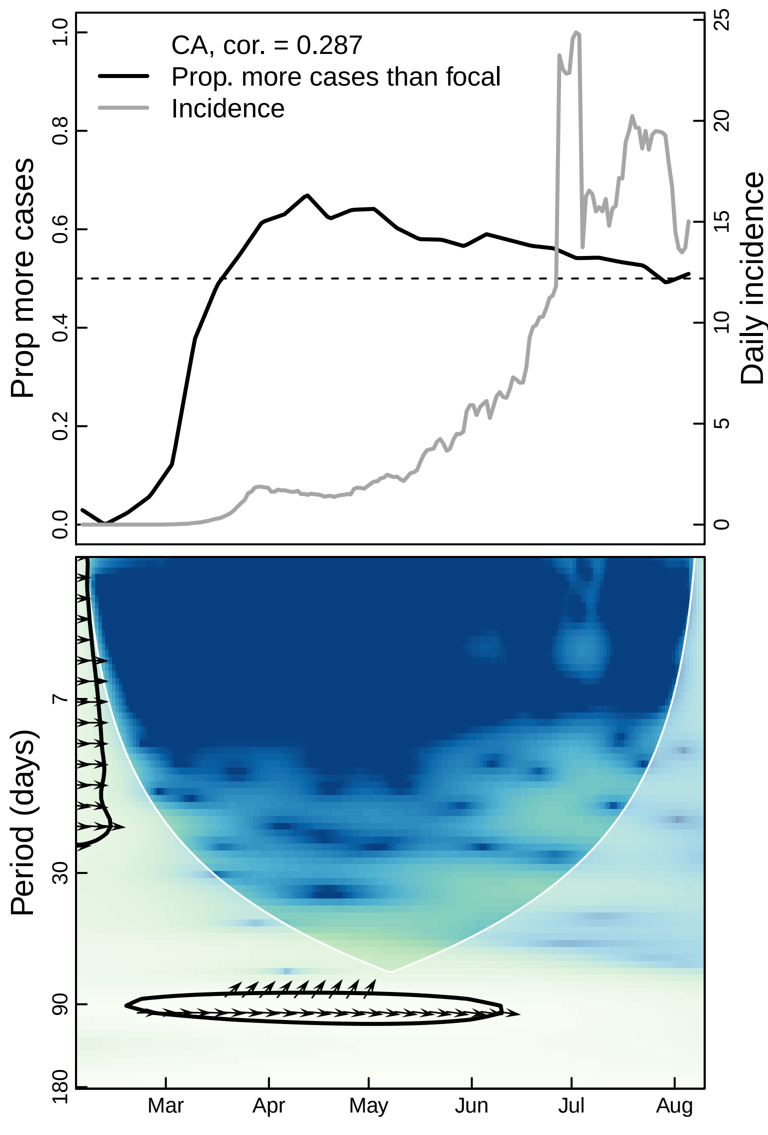
CA, cor. = 0.287
Prop. more cases than focal
Incidence
Prop more cases
Daily incidence
0.0
0.2
0.4
0.6
0.8
1.0
0
5
10
15
20
25
Period (days)
7
30
90
180
Mar
Apr
May
Jun
Jul
Aug

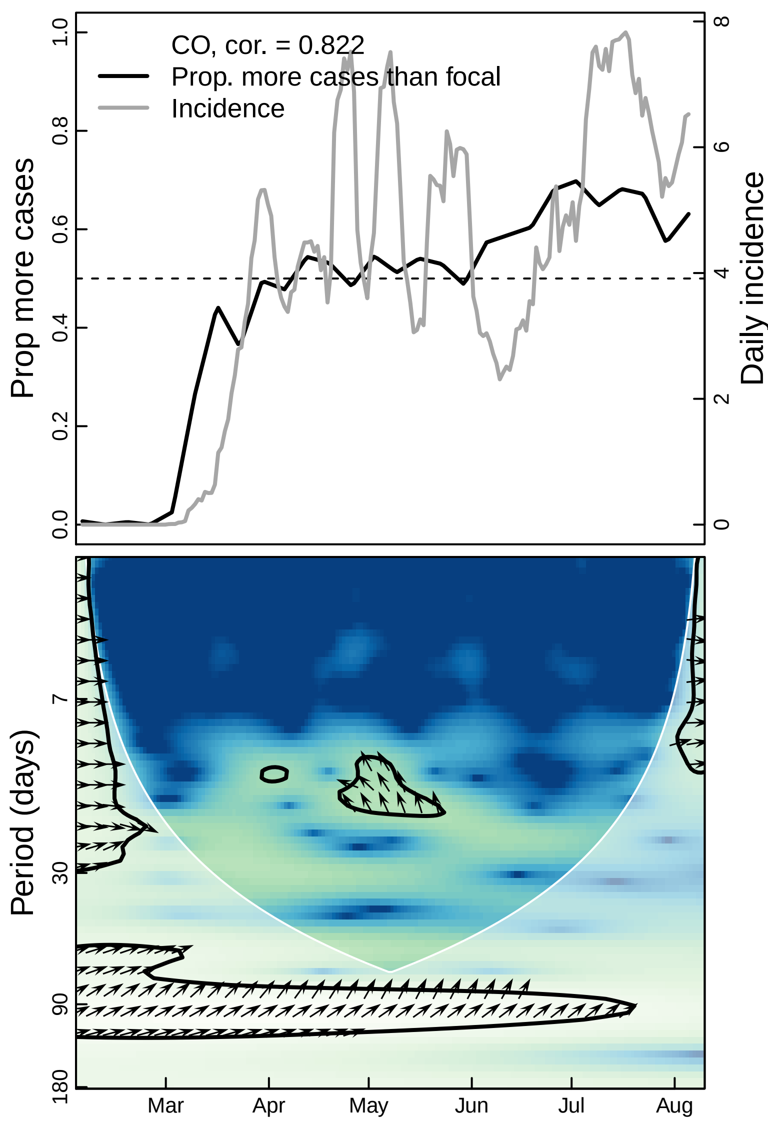
CO, cor. = 0.822
Prop. more cases than focal
Incidence
Prop more cases
Daily incidence
Period (days)
0.0
0.2
0.4
0.6
0.8
1.0
0
2
4
6
8
7
30
90
180
Mar
Apr
May
Jun
Jul
Aug

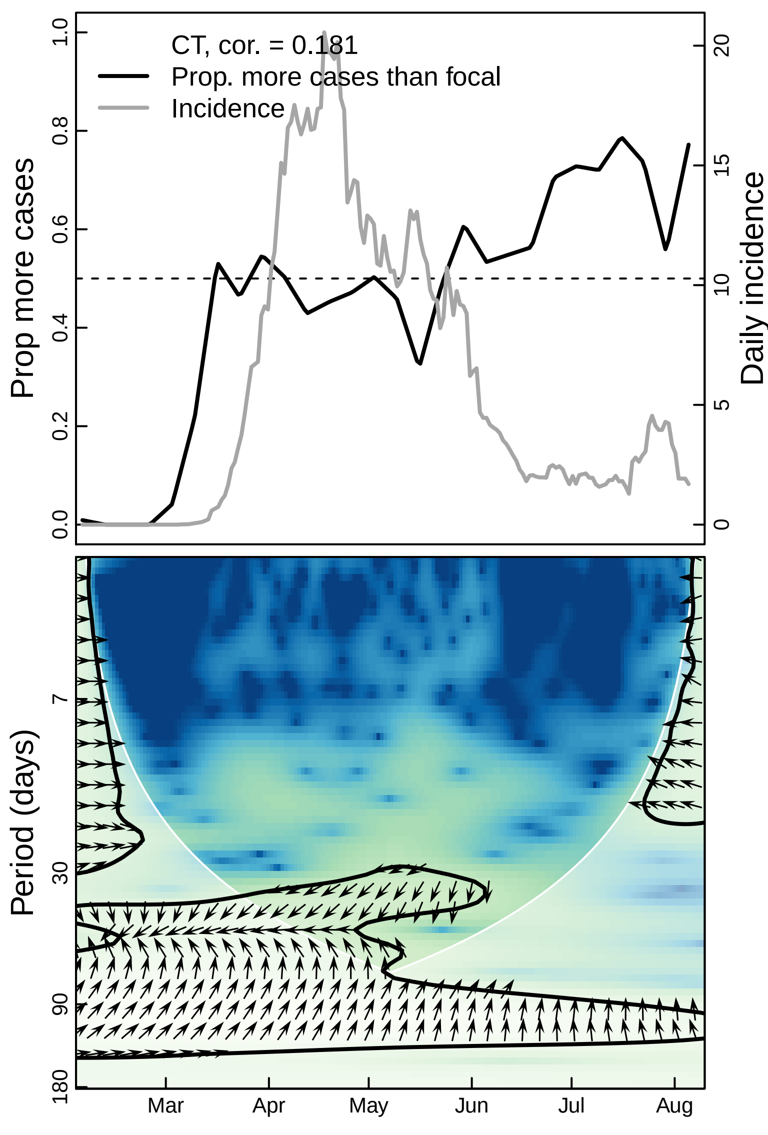

CT, cor. = 0.181
Prop. more cases than focal
Incidence
Prop more cases
Daily incidence
0.0
0.2
0.4
0.6
0.8
1.0
0
5
10
15
20
Period (days)
7
30
90
180
Mar
Apr
May
Jun
Jul
Aug

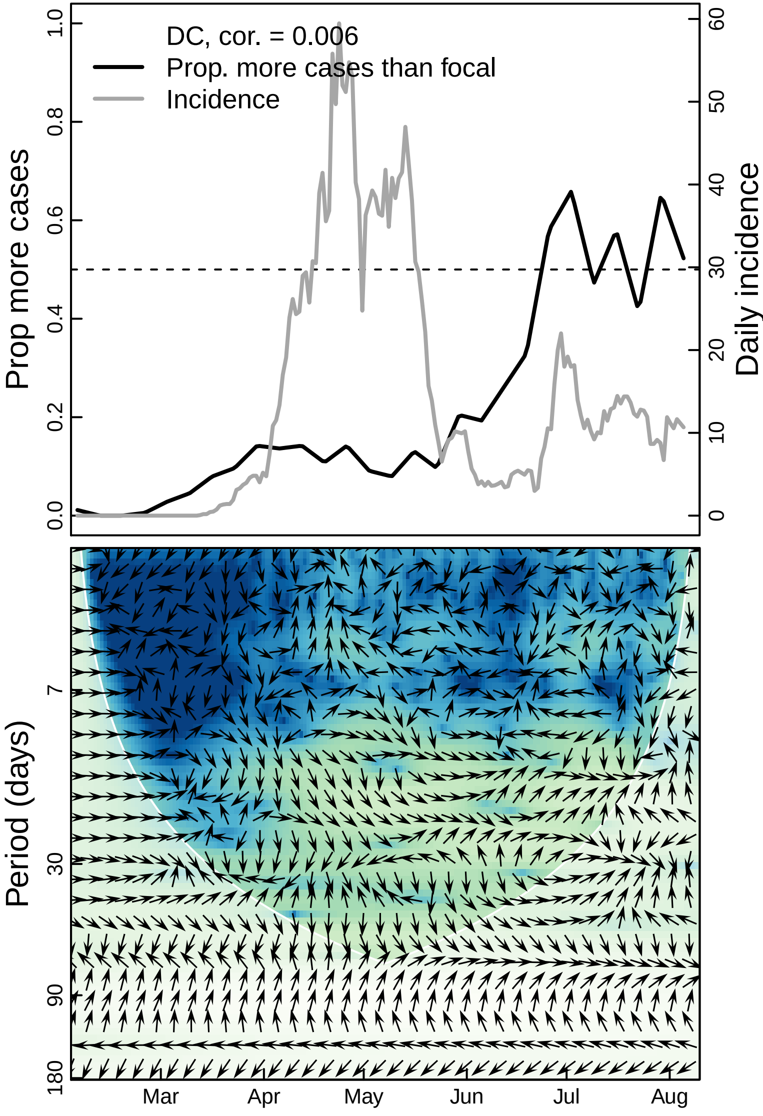

DC, cor. = 0.006
Prop. more cases than focal
Incidence
Prop more cases
Daily incidence
0.0
0.2
0.4
0.6
0.8
1.0
0
10
20
30
40
50
60
Period (days)
7
30
90
180
Mar
Apr
May
Jun
Jul
Aug

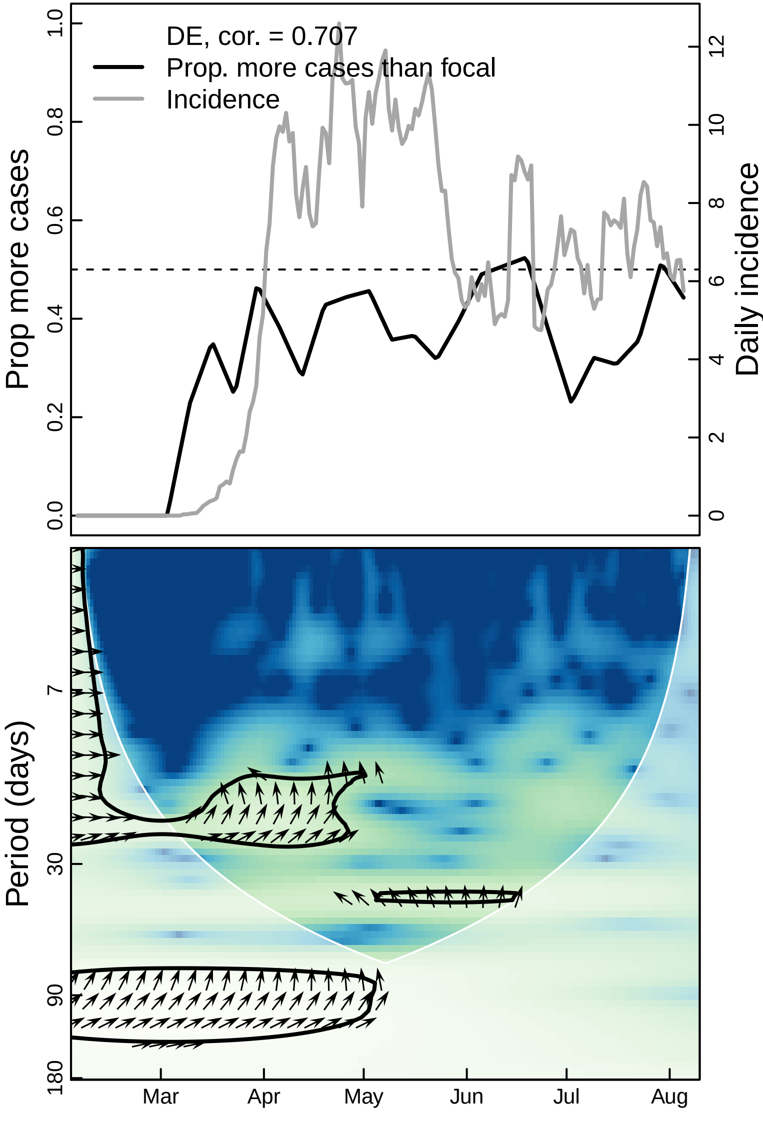
DE, cor. = 0.707
Prop. more cases than focal
Incidence
Prop more cases
Daily incidence
0.0
0.2
0.4
0.6
0.8
1.0
0
2
4
6
8
10
12
Period (days)
7
30
90
180
Mar
Apr
May
Jun
Jul
Aug

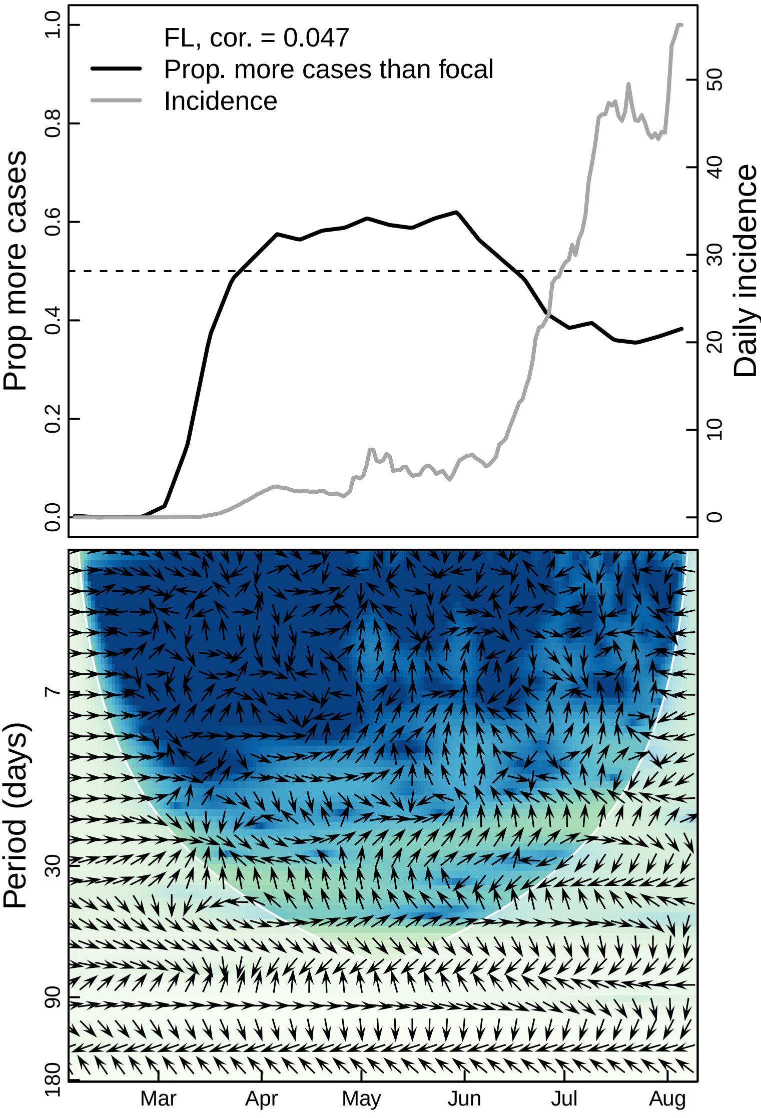
FL, cor. = 0.047
Prop. more cases than focal
Incidence
Prop more cases
Daily incidence
0.0
0.2
0.4
0.6
0.8
1.0
0
10
20
30
40
50
Period (days)
7
30
90
180
Mar
Apr
May
Jun
Jul
Aug

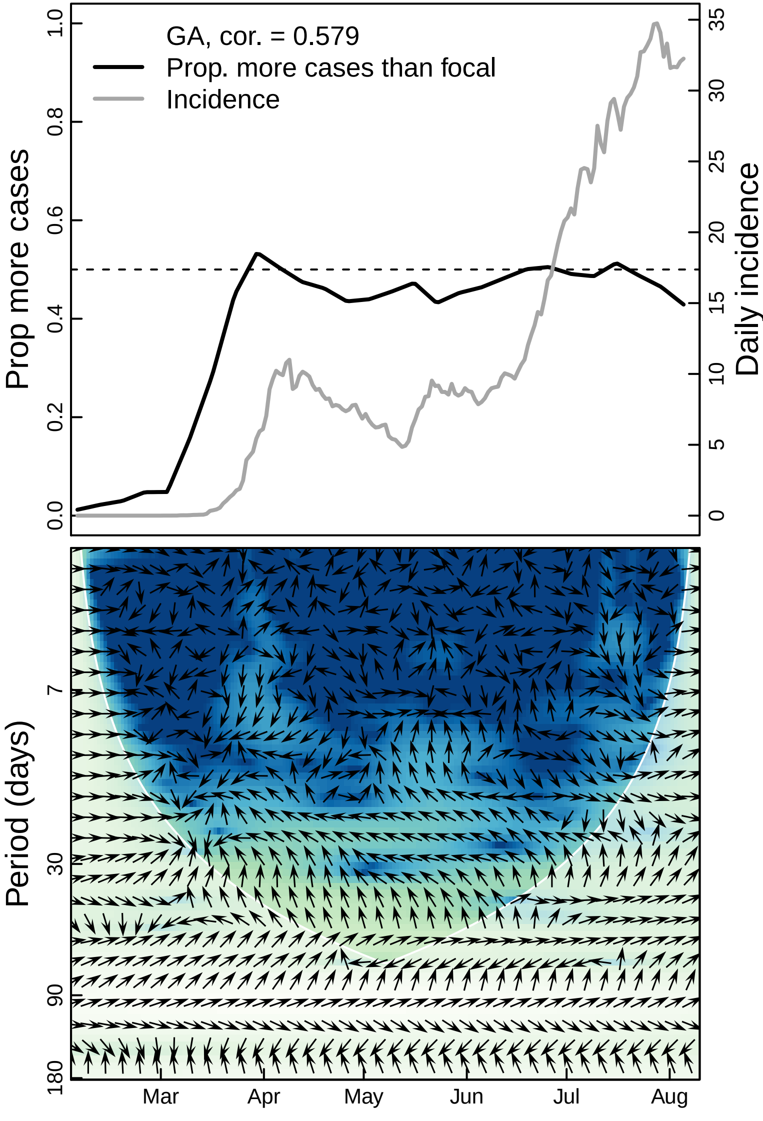

GA, cor. = 0.579
Prop. more cases than focal
Incidence
Prop more cases
Daily incidence
0.0
0.2
0.4
0.6
0.8
1.0
0
5
10
15
20
25
30
35
Period (days)
7
30
90
180
Mar
Apr
May
Jun
Jul
Aug

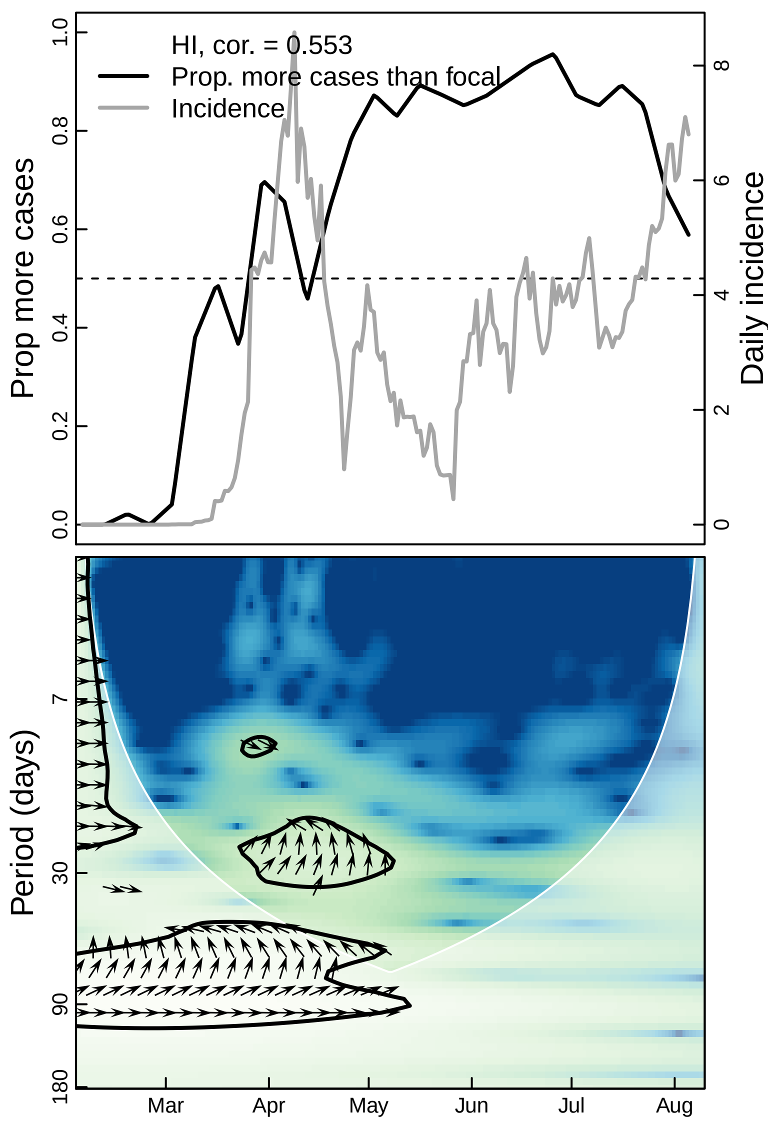

HI, cor. = 0.553
Prop. more cases than focal
Incidence
Prop more cases
Daily incidence
0.0
0.2
0.4
0.6
0.8
1.0
0
2
4
6
8
Period (days)
7
30
90
180
Mar
Apr
May
Jun
Jul
Aug

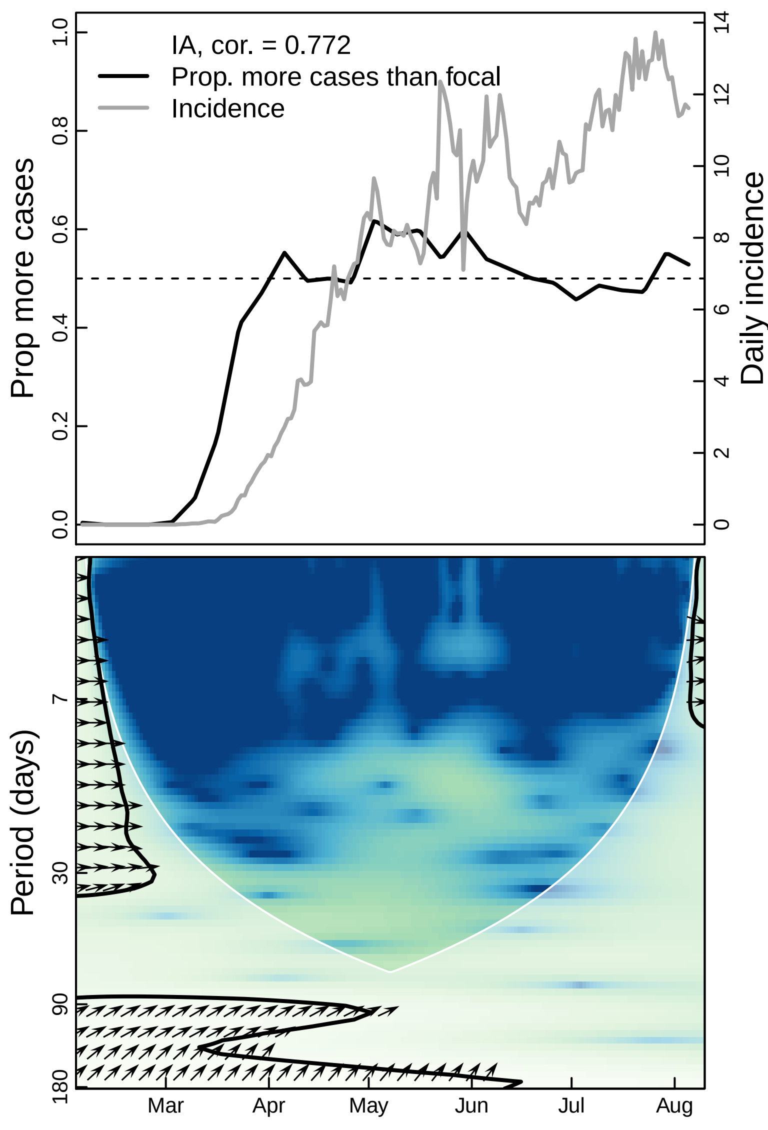

IA, cor. = 0.772
Prop. more cases than focal
Incidence
Prop more cases
Daily incidence
0.0
0.2
0.4
0.6
0.8
1.0
0
2
4
6
8
10
12
14
Period (days)
7
30
90
180
Mar
Apr
May
Jun
Jul
Aug

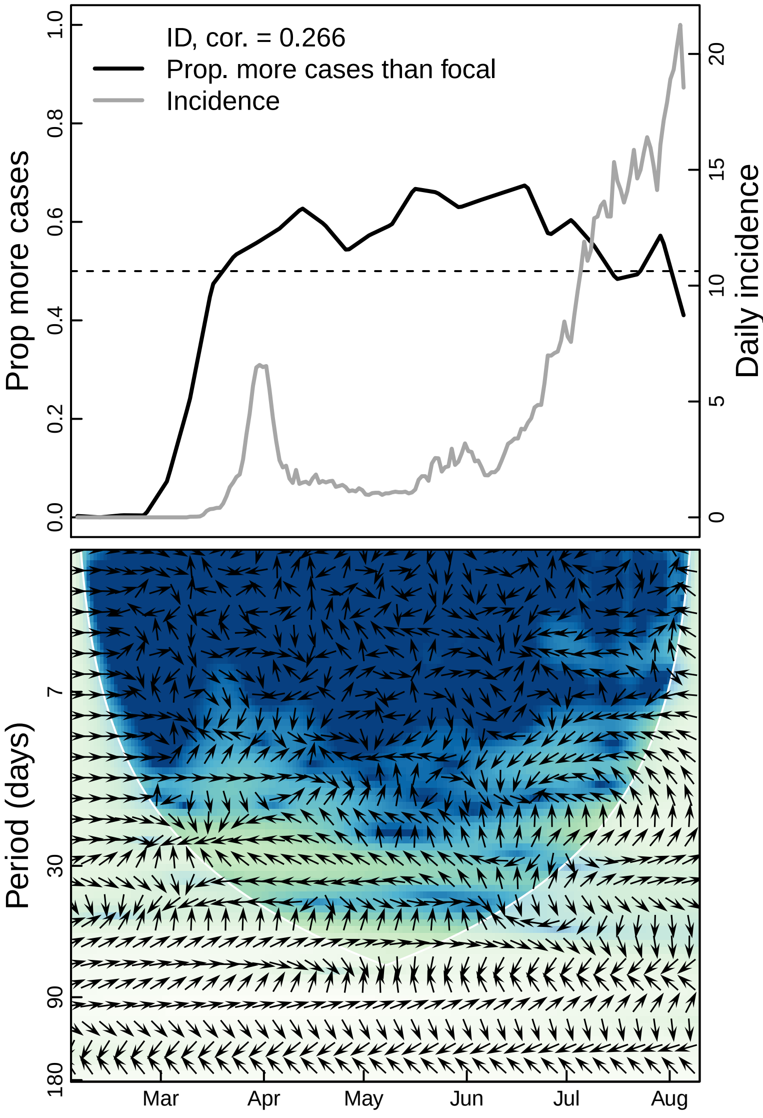
ID, cor. = 0.266
Prop. more cases than focal
Incidence
Prop more cases
Daily incidence
0.0
0.2
0.4
0.6
0.8
1.0
0
5
10
15
20
Period (days)
7
30
90
180
Mar
Apr
May
Jun
Jul
Aug

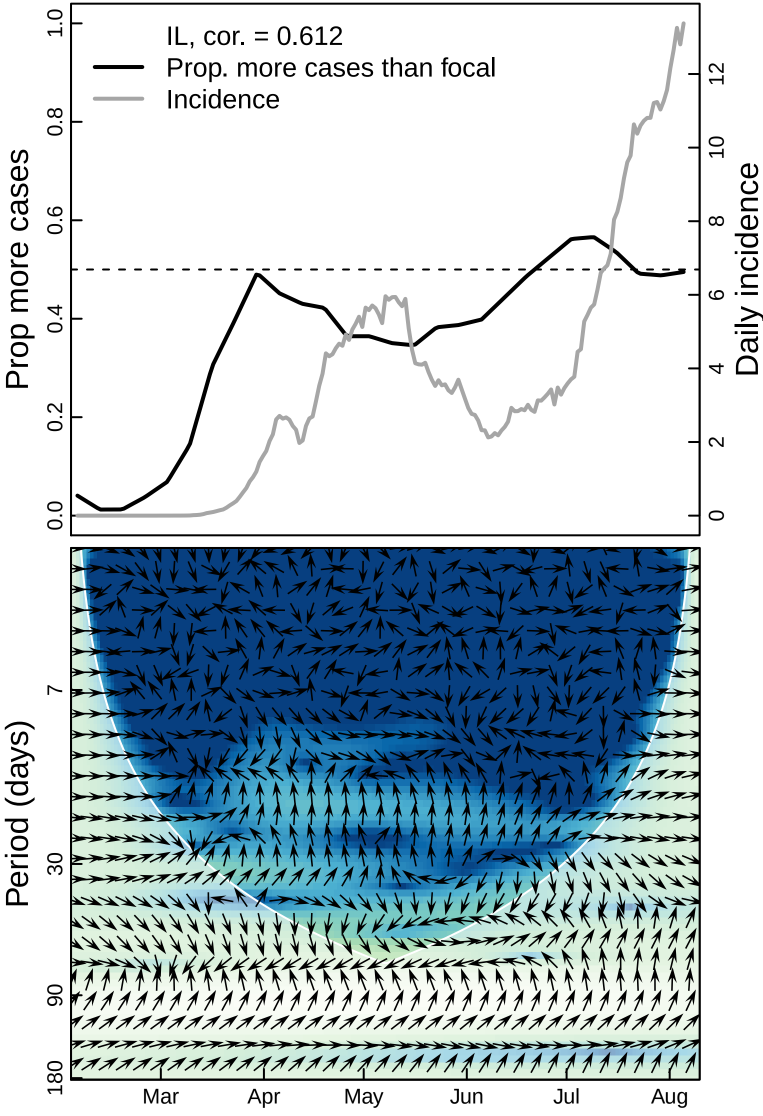

IL, cor. = 0.612
Prop. more cases than focal
Incidence
Prop more cases
Daily incidence
0.0
0.2
0.4
0.6
0.8
1.0
0
2
4
6
8
10
12
Period (days)
7
30
90
180
Mar
Apr
May
Jun
Jul
Aug

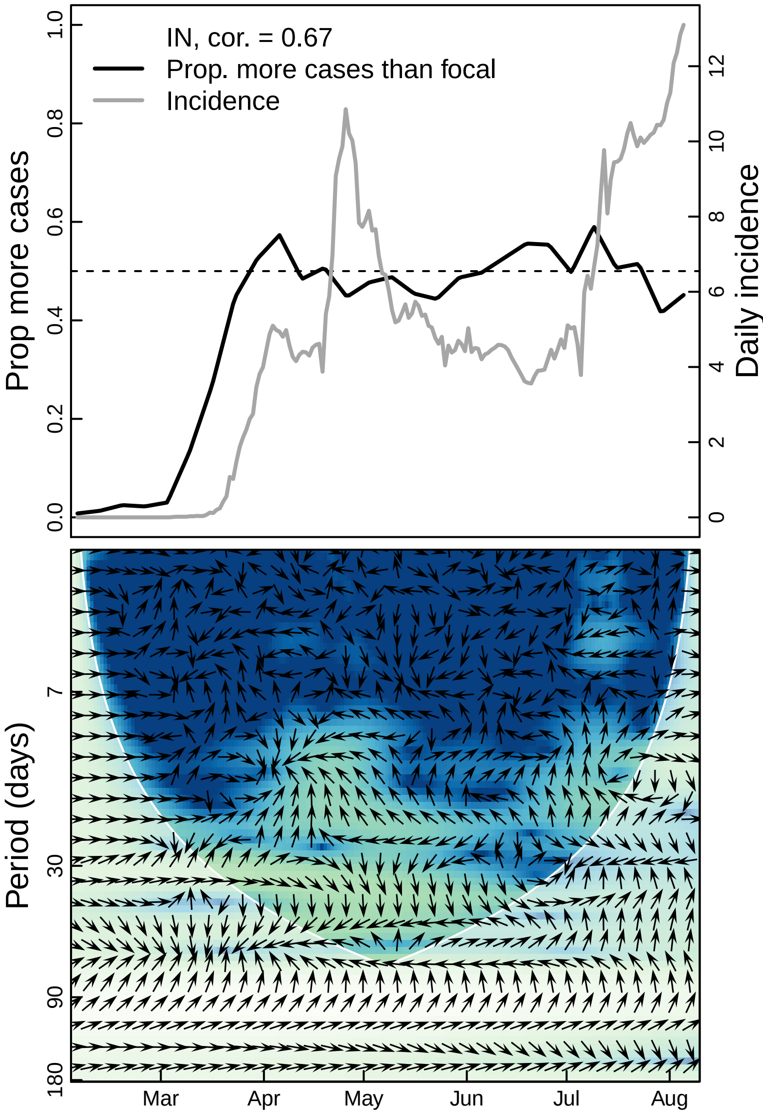

IN, cor. = 0.67
Prop. more cases than focal
Incidence
Prop more cases
Daily incidence
0.0
0.2
0.4
0.6
0.8
1.0
0
2
4
6
8
10
12
Period (days)
7
30
90
180
Mar
Apr
May
Jun
Jul
Aug

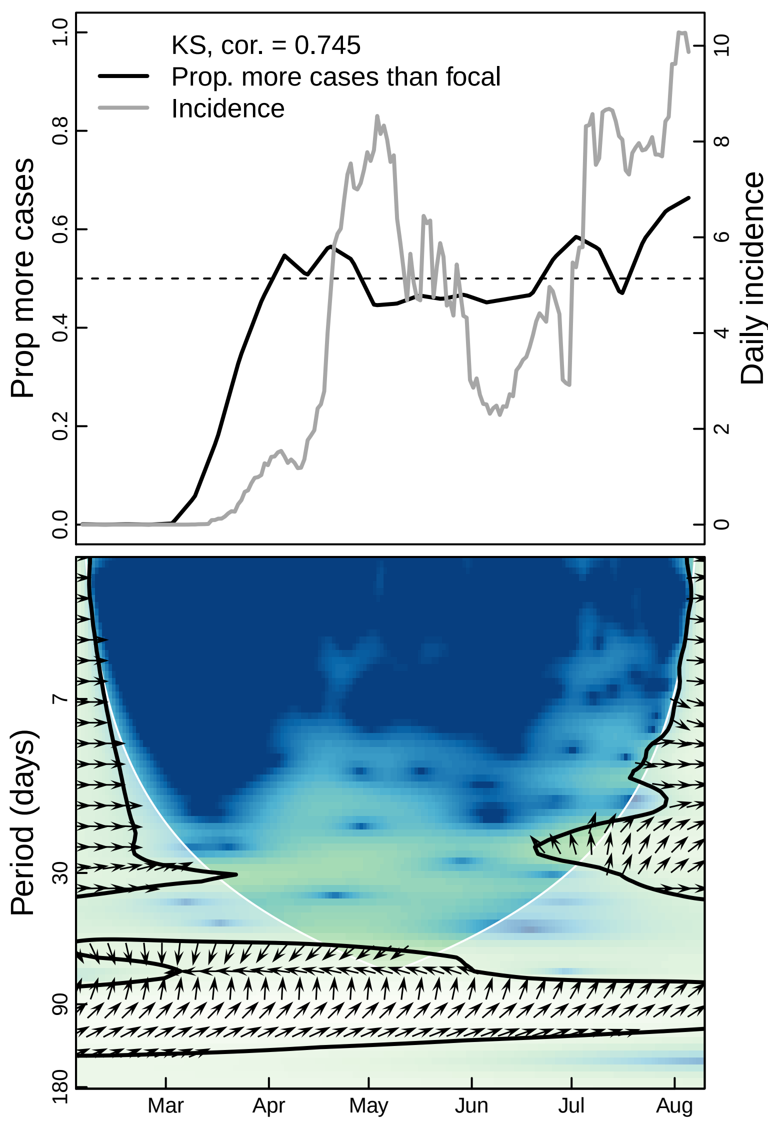

KS, cor. = 0.745
Prop. more cases than focal
Incidence
Prop more cases
Daily incidence
0.0
0.2
0.4
0.6
0.8
1.0
0
2
4
6
8
10
Period (days)
7
30
90
180
Mar
Apr
May
Jun
Jul
Aug

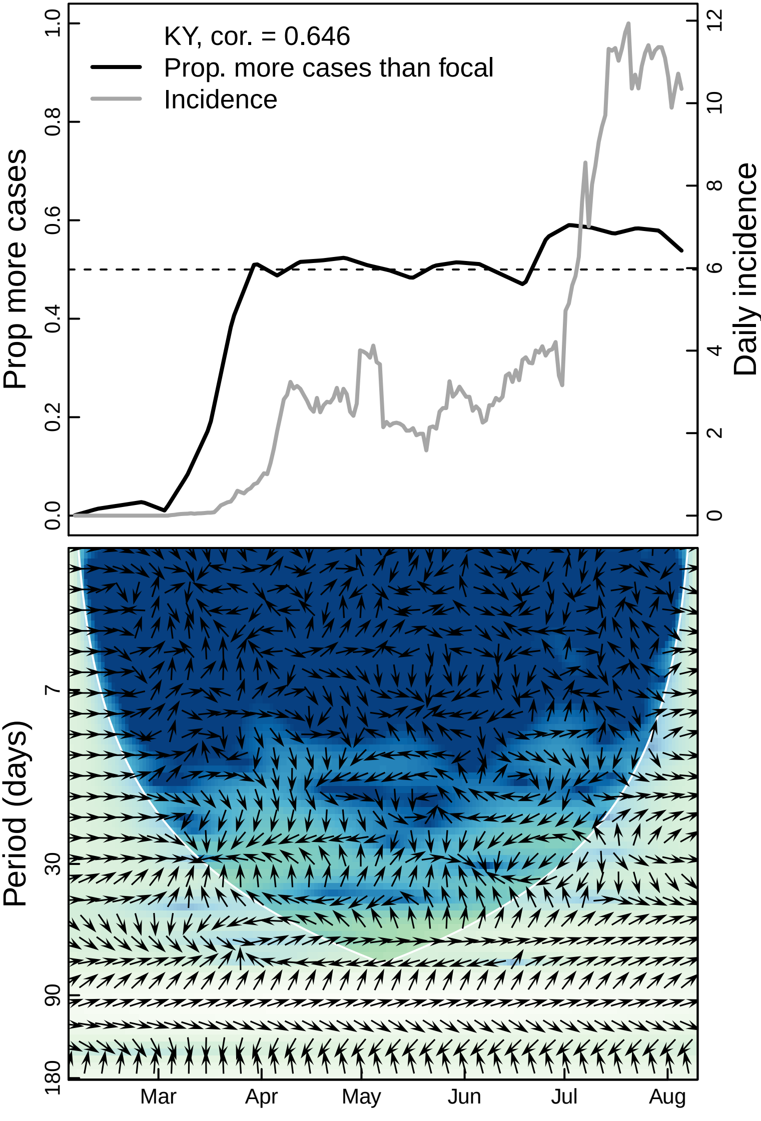

KY, cor. = 0.646
Prop. more cases than focal
Incidence
Prop more cases
Daily incidence
0.0
0.2
0.4
0.6
0.8
1.0
0
2
4
6
8
10
12
Period (days)
7
30
90
180
Mar
Apr
May
Jun
Jul
Aug

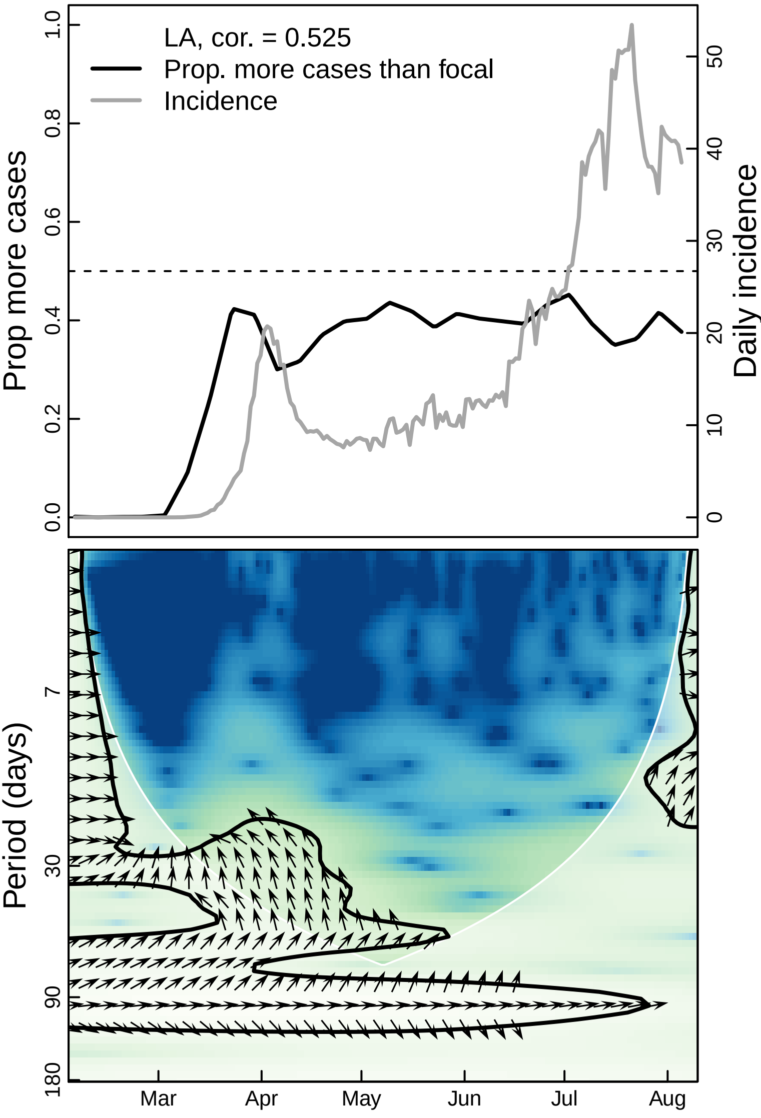

LA, cor. = 0.525
Prop. more cases than focal
Incidence
Prop more cases
Daily incidence
0.0
0.2
0.4
0.6
0.8
1.0
0
10
20
30
40
50
Period (days)
7
30
90
180
Mar
Apr
May
Jun
Jul
Aug

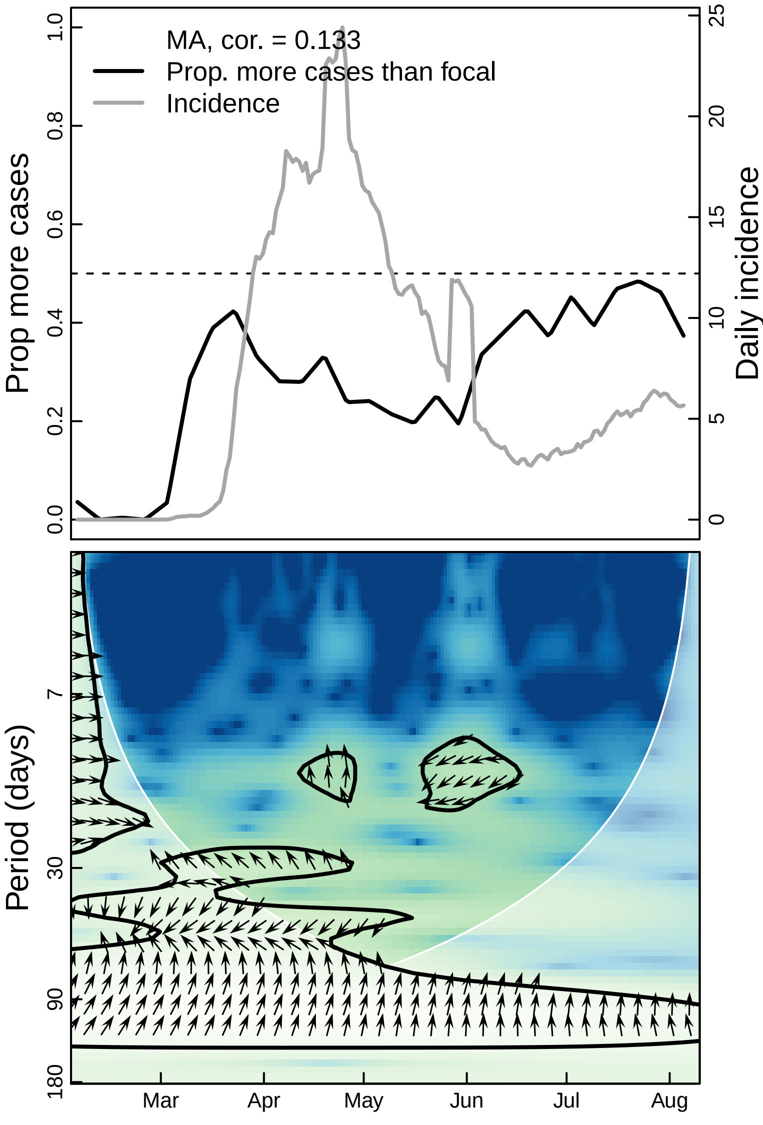

MA, cor. = 0.133
Prop. more cases than focal
Incidence
Prop more cases
0.0
0.2
0.4
0.6
0.8
1.0
Daily incidence
0
5
10
15
20
25
Period (days)
7
30
90
180
Mar
Apr
May
Jun
Jul
Aug

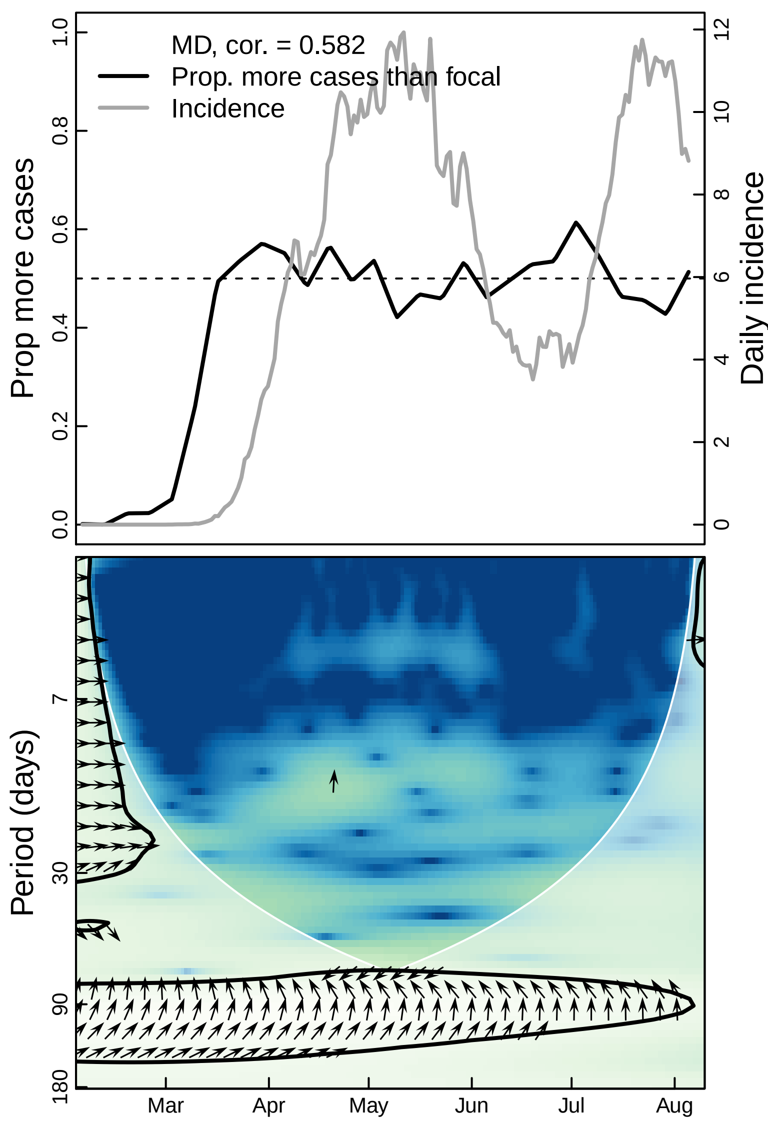
MD, cor. = 0.582
Prop. more cases than focal
Incidence
Prop more cases
Daily incidence
0.0
0.2
0.4
0.6
0.8
1.0
0
2
4
6
8
10
12
Period (days)
7
30
90
180
Mar
Apr
May
Jun
Jul
Aug

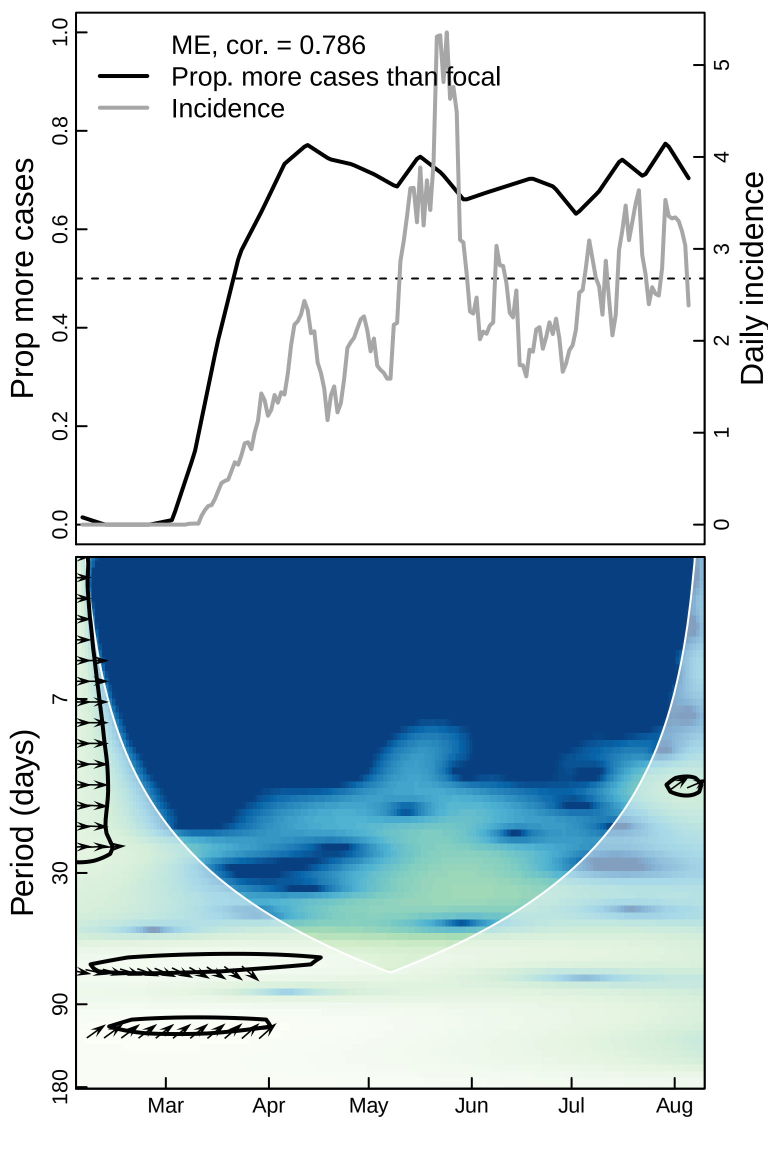
ME, cor. = 0.786
Prop. more cases than focal
Incidence
Prop more cases
Daily incidence
Period (days)
Mar
Apr
May
Jun
Jul
Aug

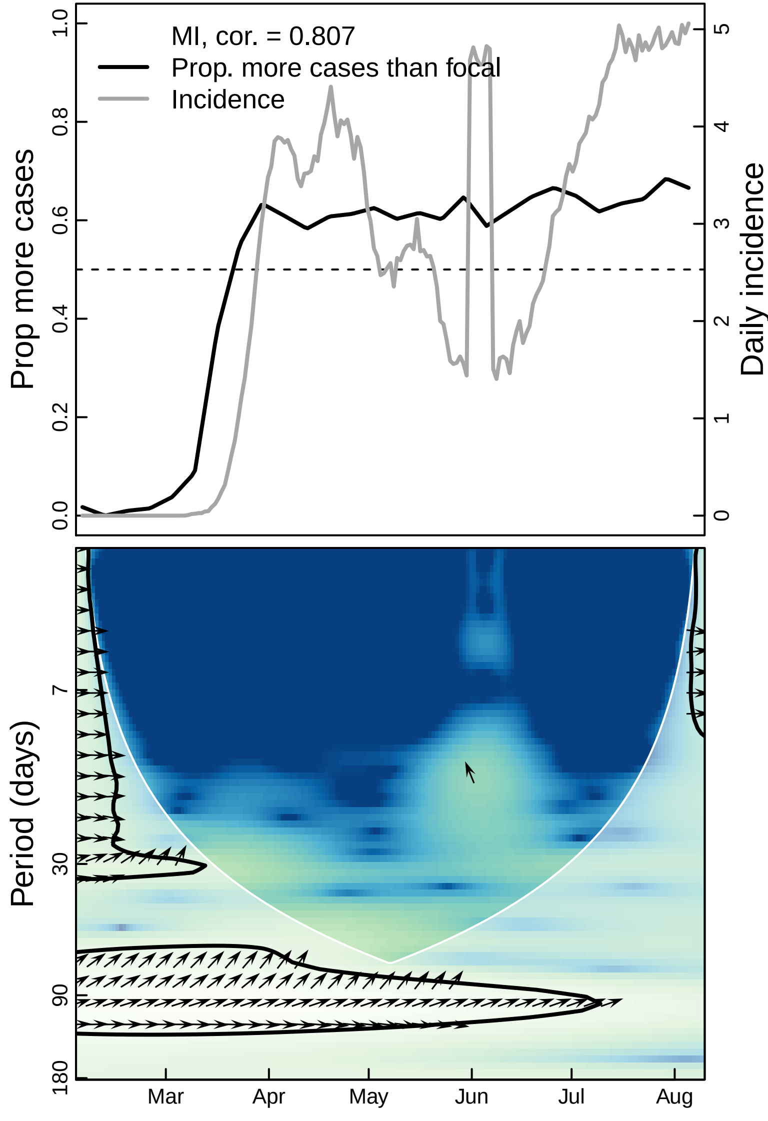
MI, cor. = 0.807
Prop. more cases than focal
Incidence
Prop more cases
Daily incidence
1.0
0.8
0.6
0.4
0.2
0.0
5
4
3
2
1
0
Period (days)
7
30
90
180
Mar
Apr
May
Jun
Jul
Aug

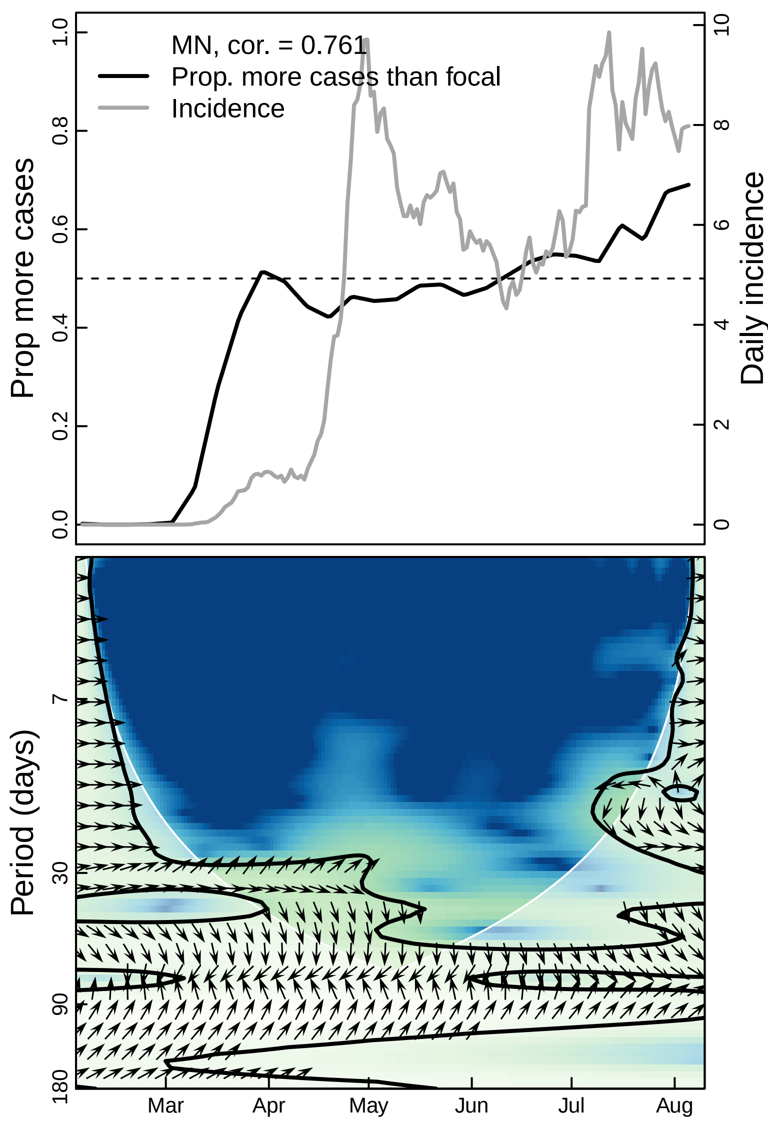
MN, cor. = 0.761
Prop. more cases than focal
Incidence
Prop more cases
Daily incidence
0.0
0.2
0.4
0.6
0.8
1.0
0
2
4
6
8
10
Period (days)
7
30
90
180
Mar
Apr
May
Jun
Jul
Aug

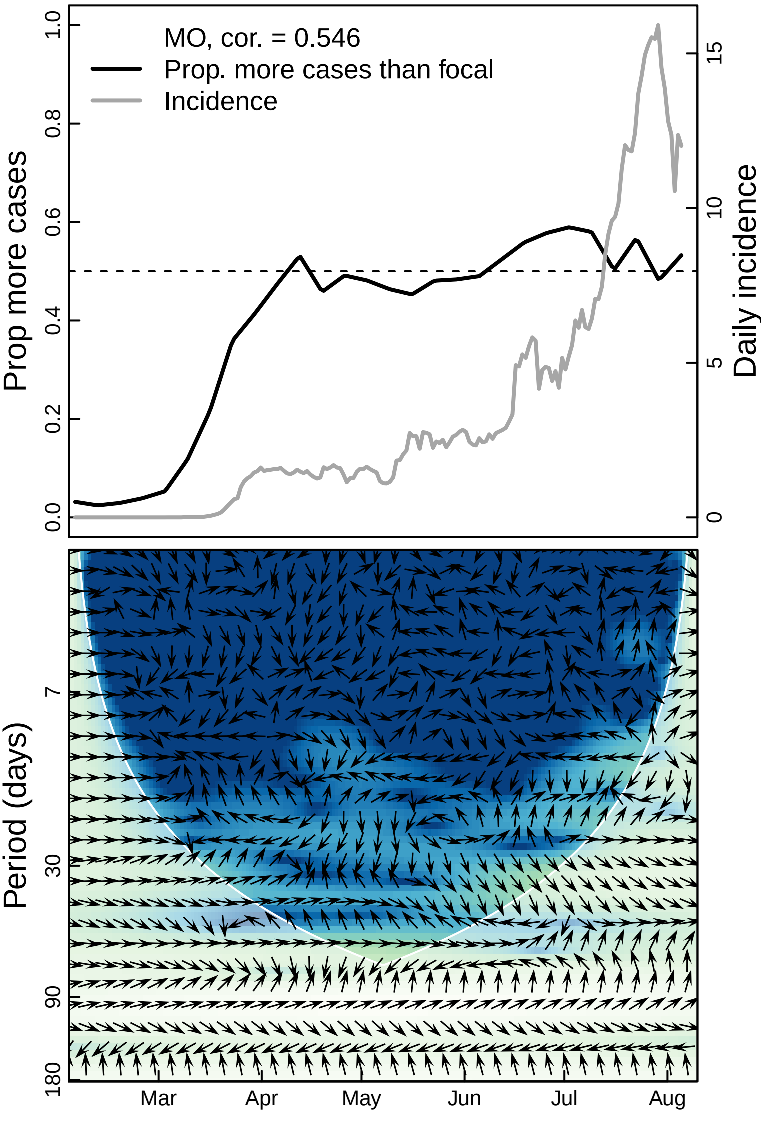

MO, cor. = 0.546
Prop. more cases than focal
Incidence
Prop more cases
Daily incidence
0.0
0.2
0.4
0.6
0.8
1.0
0
5
10
15
Period (days)
7
30
90
180
Mar
Apr
May
Jun
Jul
Aug

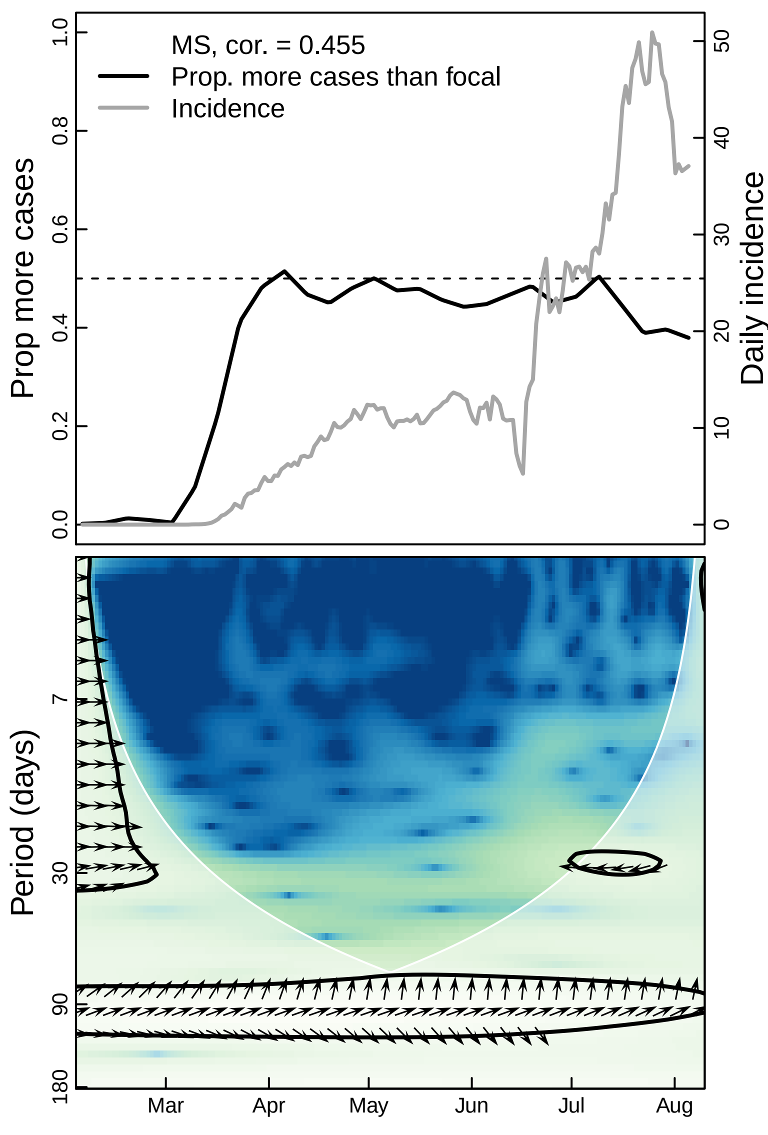
MS, cor. = 0.455
Prop. more cases than focal
Incidence
Prop more cases
Daily incidence
0.0
0.2
0.4
0.6
0.8
1.0
0
10
20
30
40
50
Period (days)
7
30
90
180
Mar
Apr
May
Jun
Jul
Aug

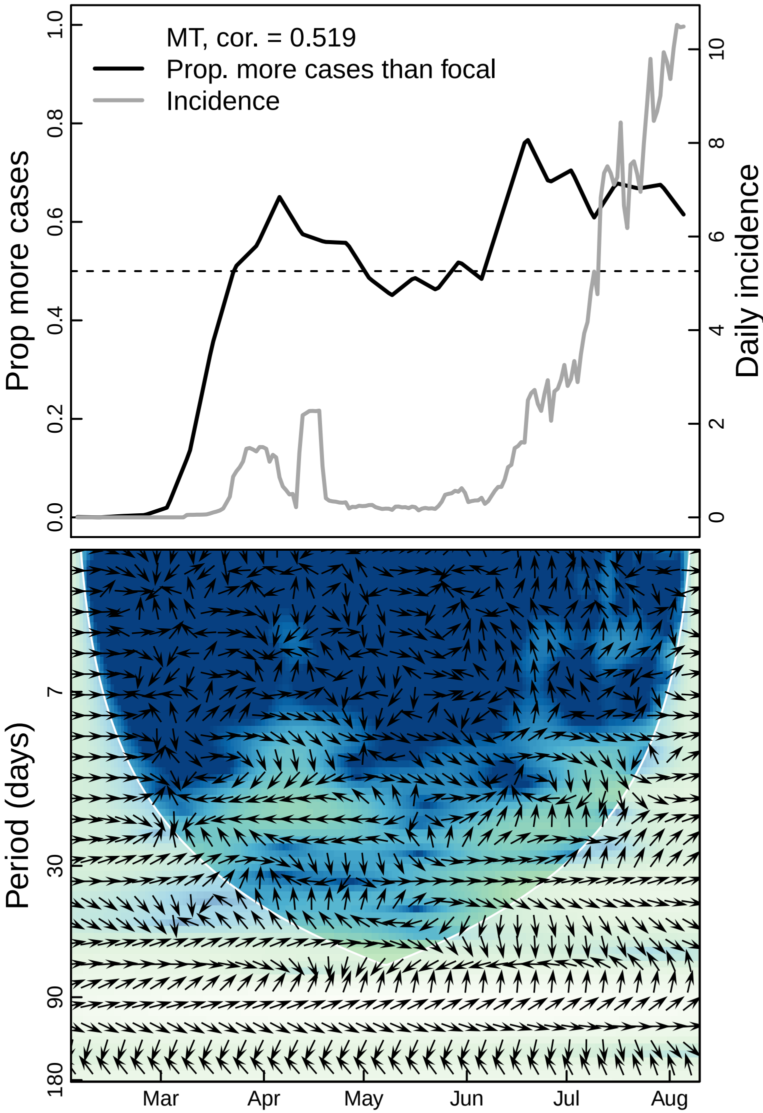

MT, cor. = 0.519
Prop. more cases than focal
Incidence
Prop more cases
0.0
0.2
0.4
0.6
0.8
1.0
Daily incidence
0
2
4
6
8
10
Period (days)
7
30
90
180
Mar
Apr
May
Jun
Jul
Aug

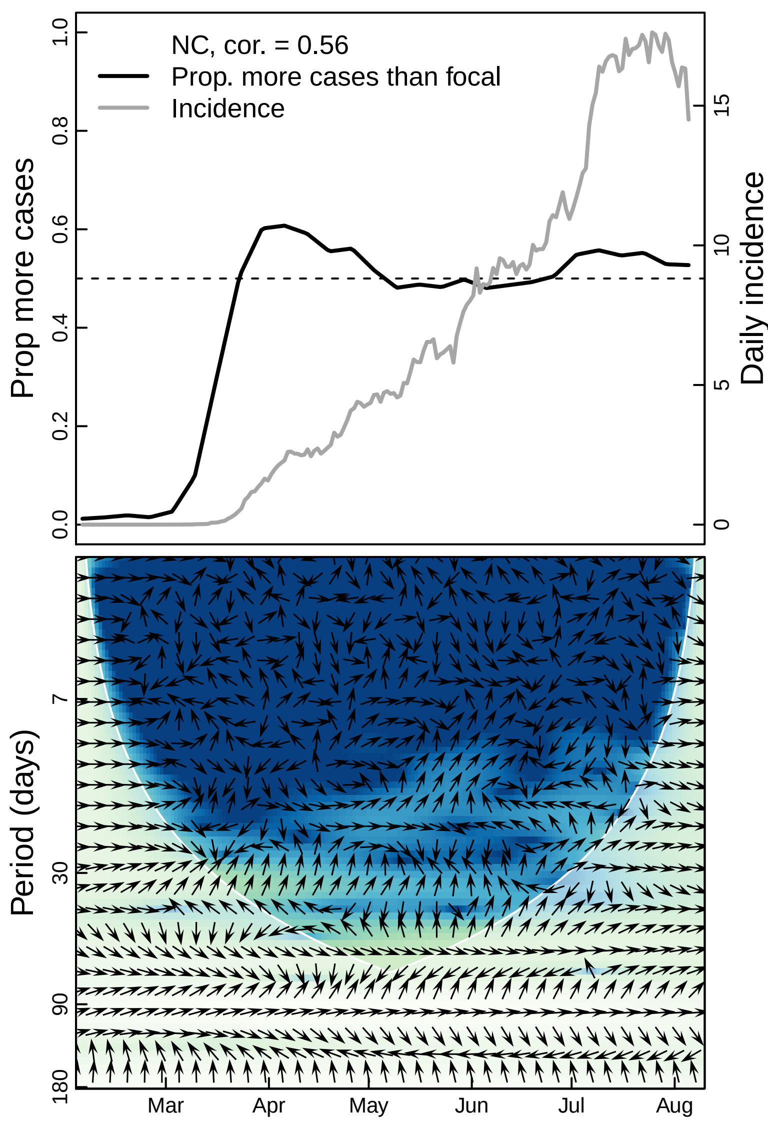

NC, cor. = 0.56
Prop. more cases than focal
Incidence
Prop more cases
Daily incidence
0.0
0.2
0.4
0.6
0.8
1.0
0
5
10
15
Period (days)
7
30
90
180
Mar
Apr
May
Jun
Jul
Aug

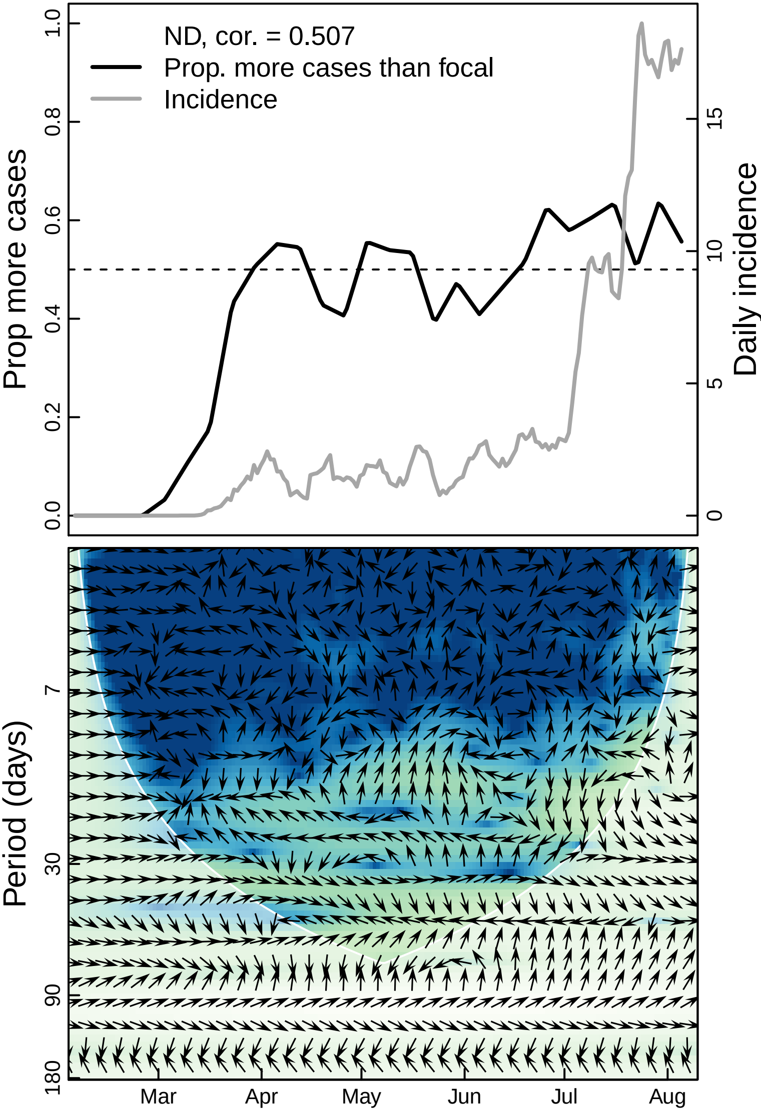

ND, cor. = 0.507
Prop. more cases than focal
Incidence
Prop more cases
Daily incidence
0.0
0.2
0.4
0.6
0.8
1.0
0
5
10
15
Period (days)
7
30
90
180
Mar
Apr
May
Jun
Jul
Aug

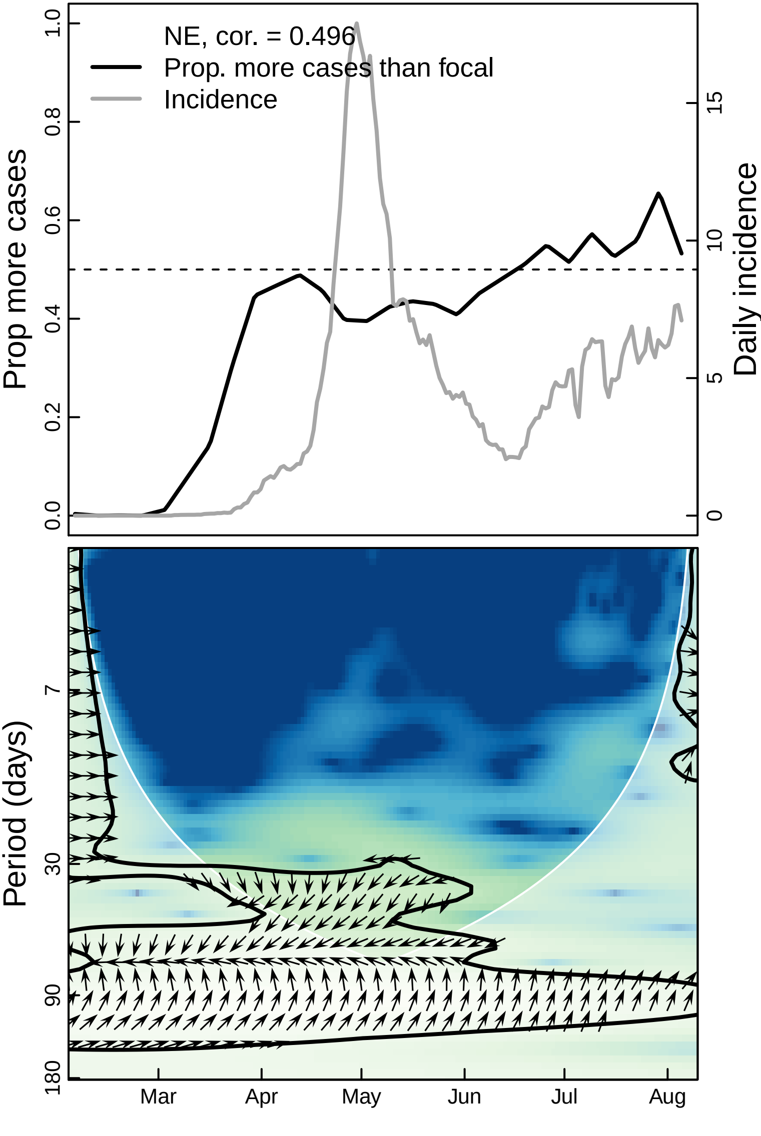
NE, cor. = 0.496
Prop. more cases than focal
Incidence
Prop more cases
Daily incidence
0.0
0.2
0.4
0.6
0.8
1.0
0
5
10
15
Period (days)
7
30
90
180
Mar
Apr
May
Jun
Jul
Aug

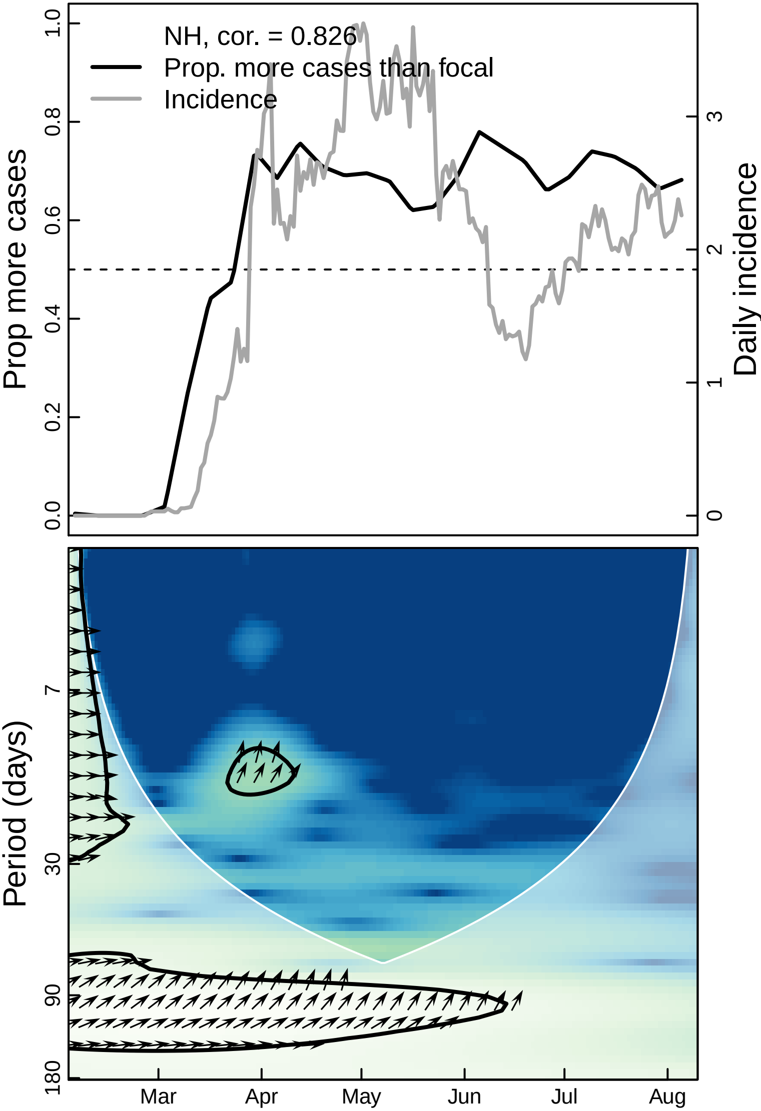
NH, cor. = 0.826
Prop. more cases than focal
Incidence
Prop more cases
0.0
0.2
0.4
0.6
0.8
1.0
Daily incidence
0
1
2
3
Period (days)
7
30
90
180
Mar
Apr
May
Jun
Jul
Aug

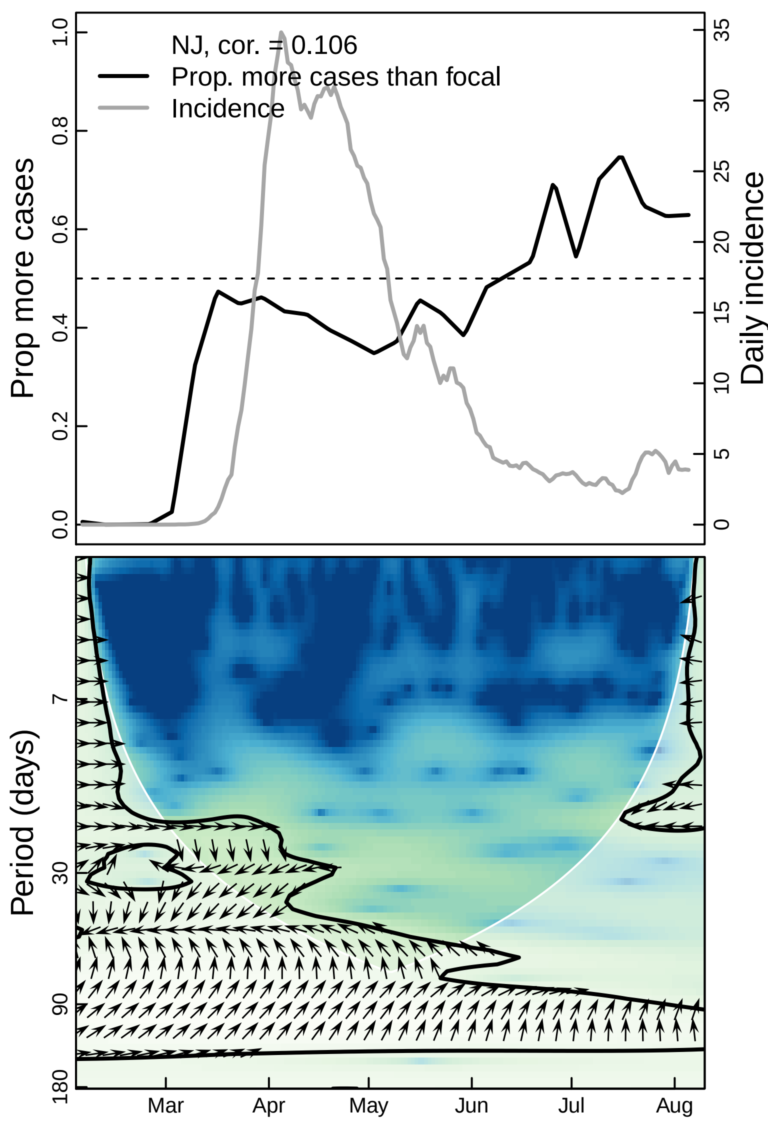

NJ, cor. = 0.106
Prop. more cases than focal
Incidence
Prop more cases
Daily incidence
0.0
0.2
0.4
0.6
0.8
1.0
0
5
10
15
20
25
30
35
Period (days)
7
30
90
180
Mar
Apr
May
Jun
Jul
Aug

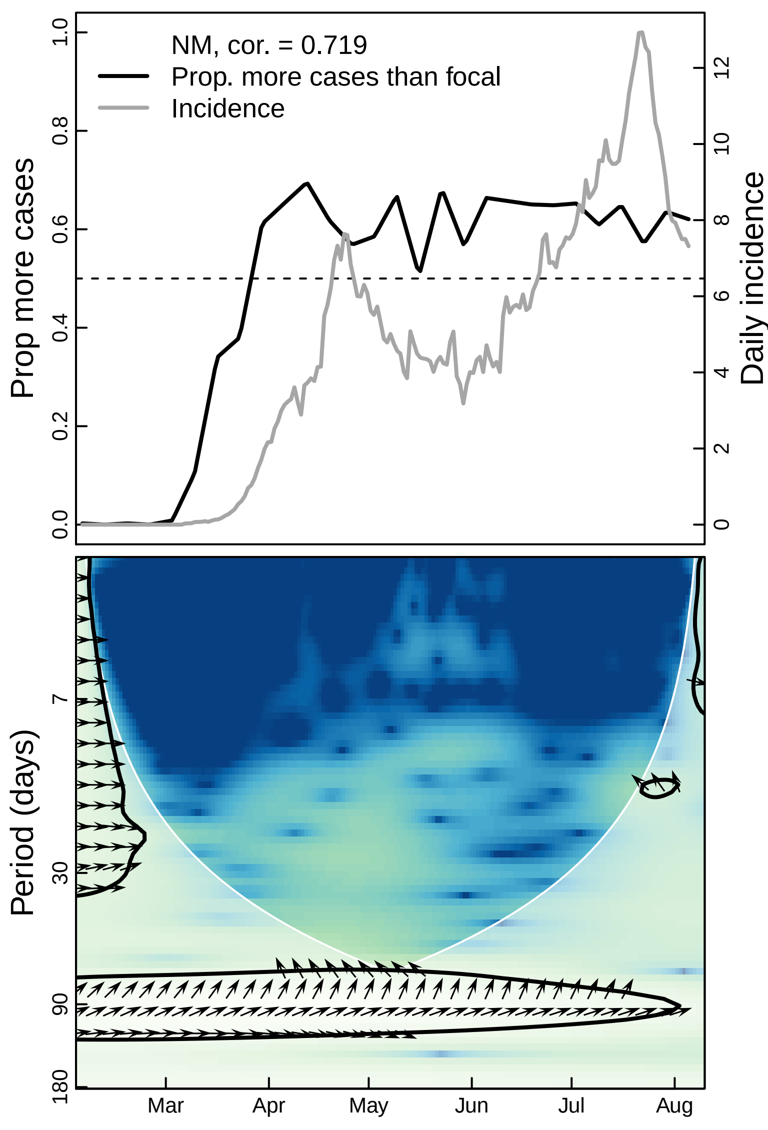
NM, cor. = 0.719
Prop. more cases than focal
Incidence
Prop more cases
Daily incidence
0.0
0.2
0.4
0.6
0.8
1.0
0
2
4
6
8
10
12
Period (days)
7
30
90
180
Mar
Apr
May
Jun
Jul
Aug

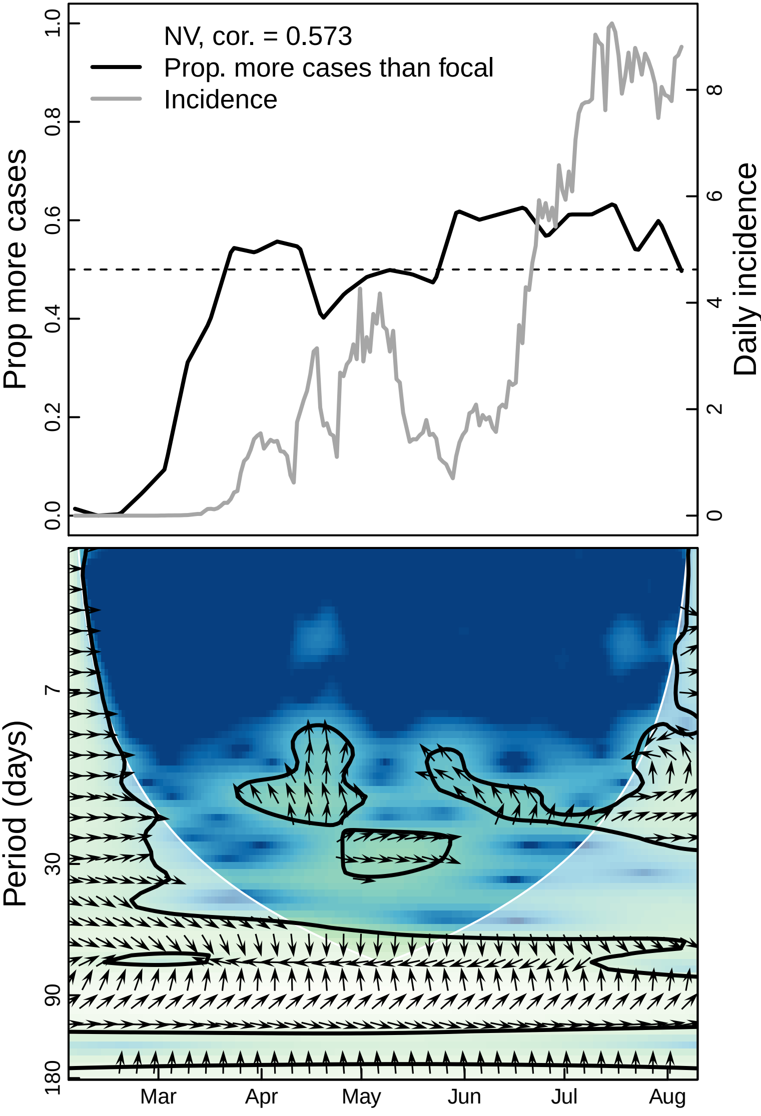

NV, cor. = 0.573
Prop. more cases than focal
Incidence
Prop more cases
Daily incidence
0.0
0.2
0.4
0.6
0.8
1.0
0
2
4
6
8
Period (days)
7
30
90
180
Mar
Apr
May
Jun
Jul
Aug

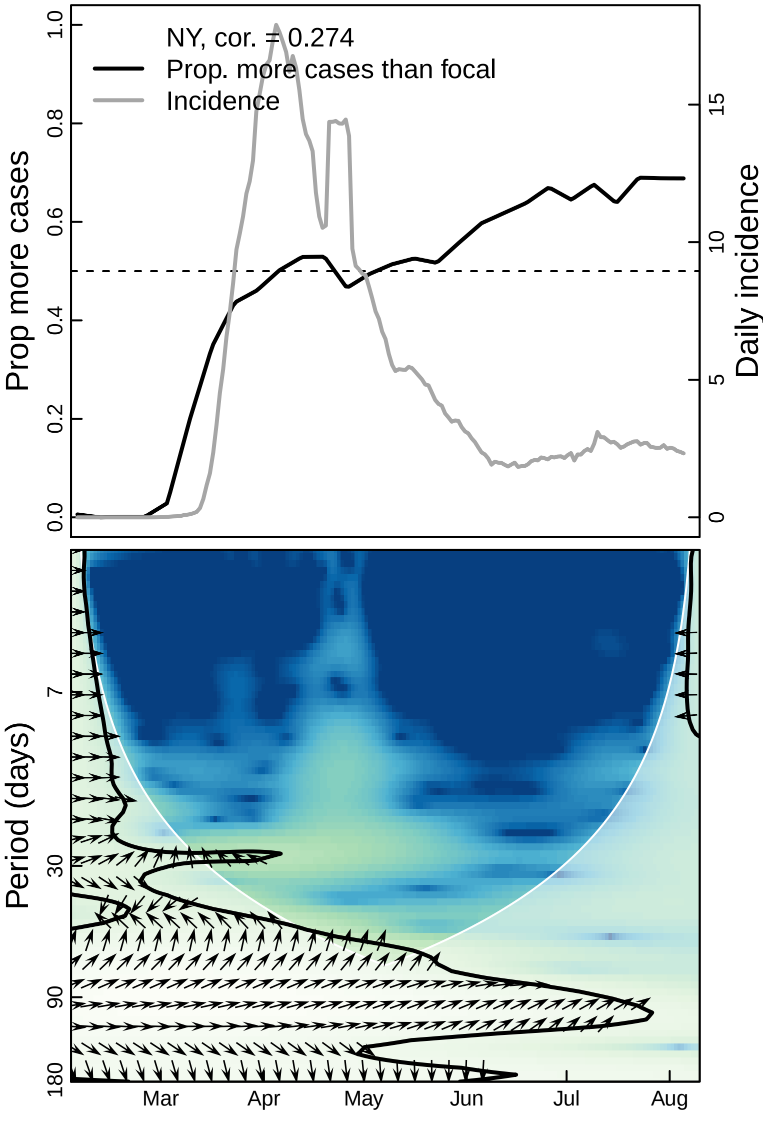

NY, cor. = 0.274
Prop. more cases than focal
Incidence
Prop more cases
Daily incidence
0.0
0.2
0.4
0.6
0.8
1.0
0
5
10
15
Period (days)
7
30
90
180
Mar
Apr
May
Jun
Jul
Aug

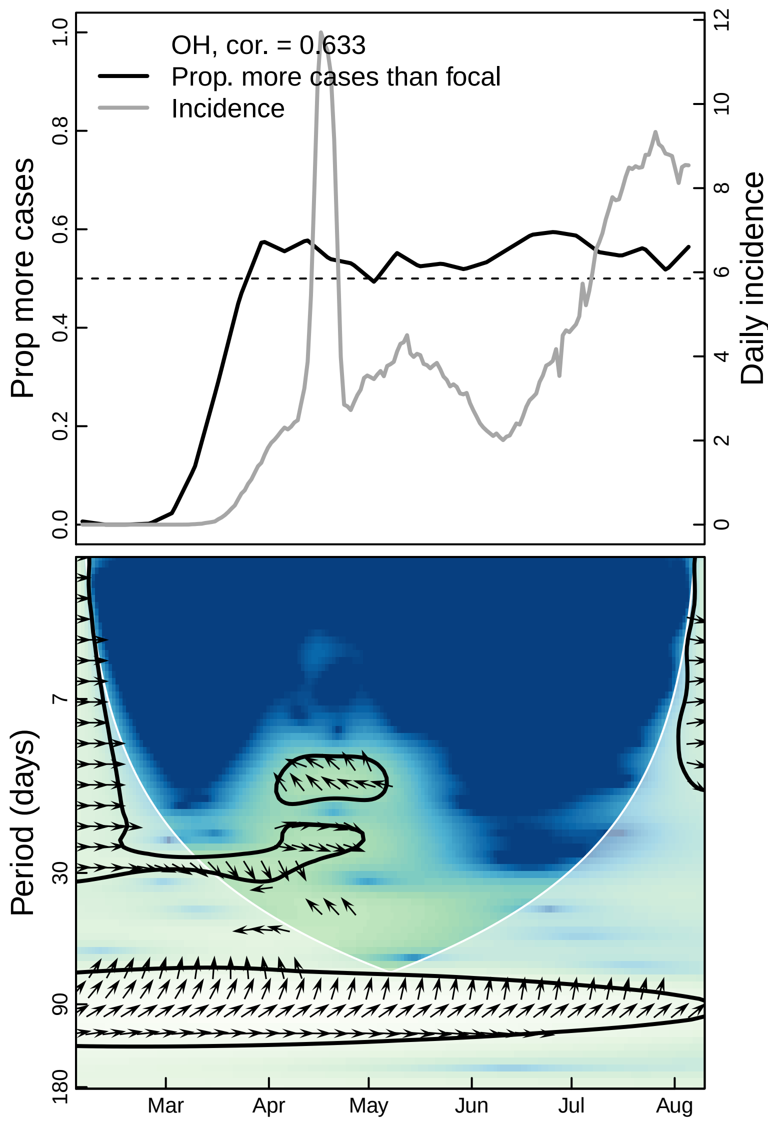
OH, cor. = 0.633
Prop. more cases than focal
Incidence
Prop more cases
Daily incidence
0.0
0.2
0.4
0.6
0.8
1.0
0
2
4
6
8
10
12
Period (days)
7
30
90
180
Mar
Apr
May
Jun
Jul
Aug

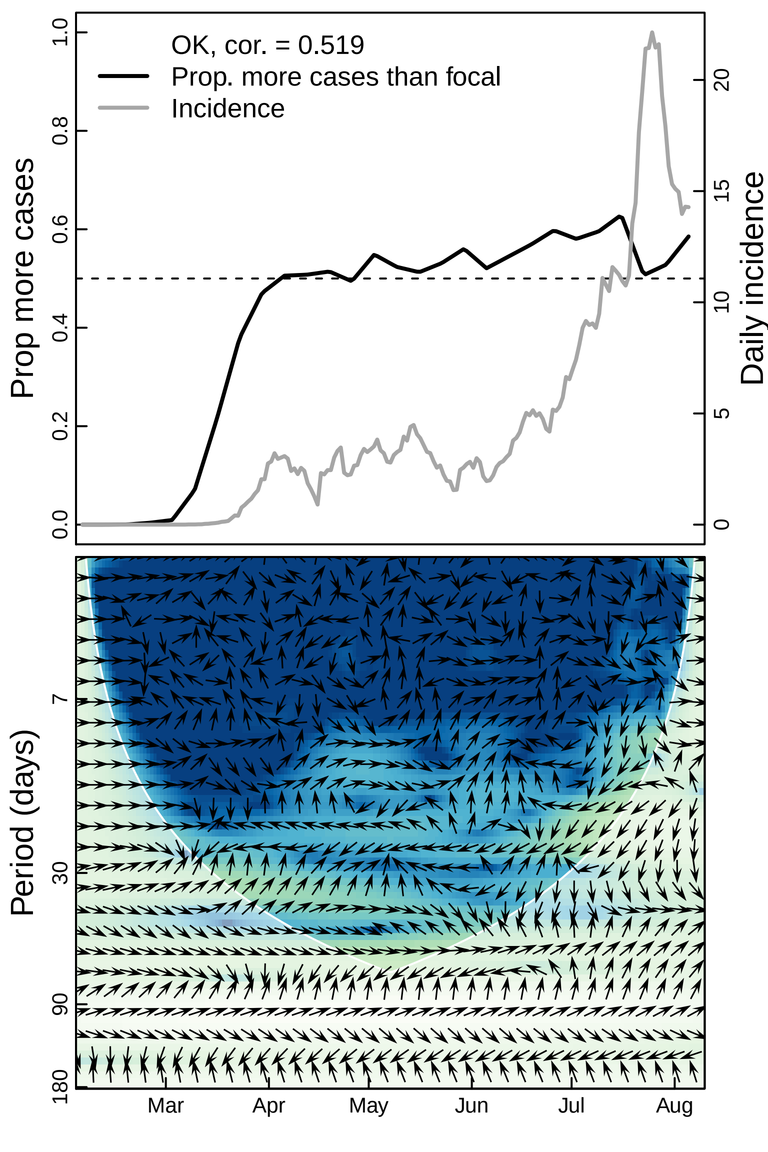

OK, cor. = 0.519
Prop. more cases than focal
Incidence
Prop more cases
Daily incidence
0.0
0.2
0.4
0.6
0.8
1.0
0
5
10
15
20
Period (days)
7
30
90
180
Mar
Apr
May
Jun
Jul
Aug

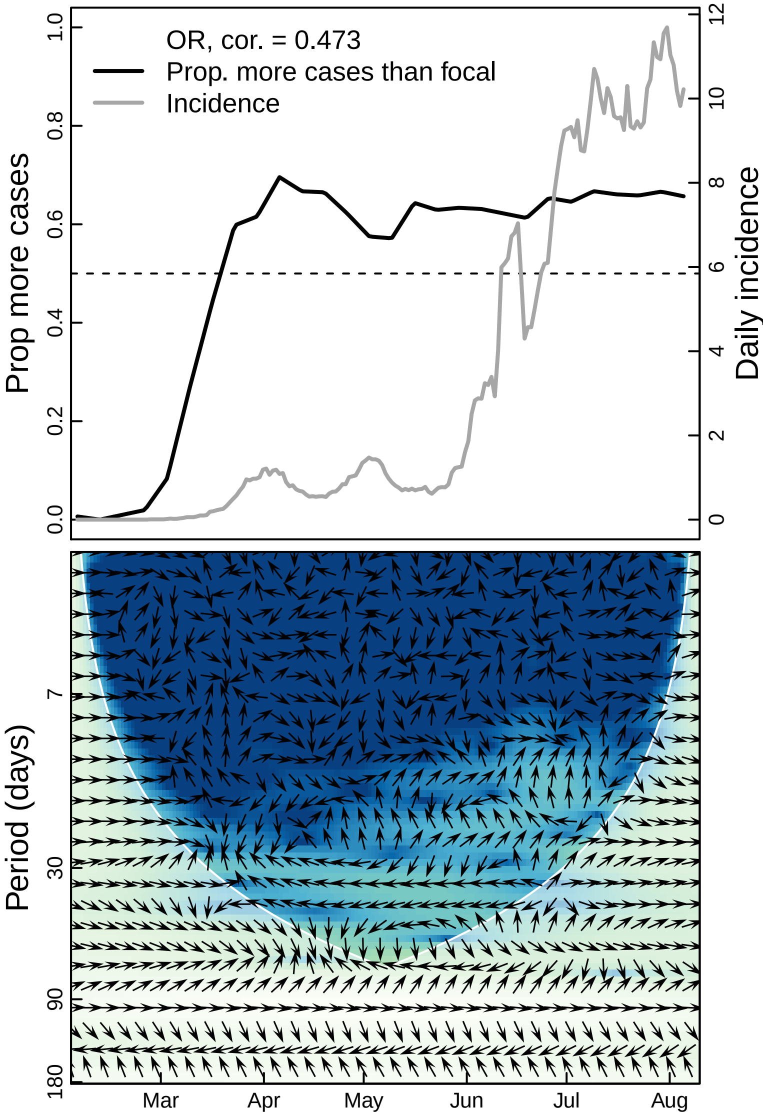

OR, cor. = 0.473
Prop. more cases than focal
Incidence
Prop more cases
Daily incidence
0.0
0.2
0.4
0.6
0.8
1.0
0
2
4
6
8
10
12
Period (days)
7
30
90
180
Mar
Apr
May
Jun
Jul
Aug

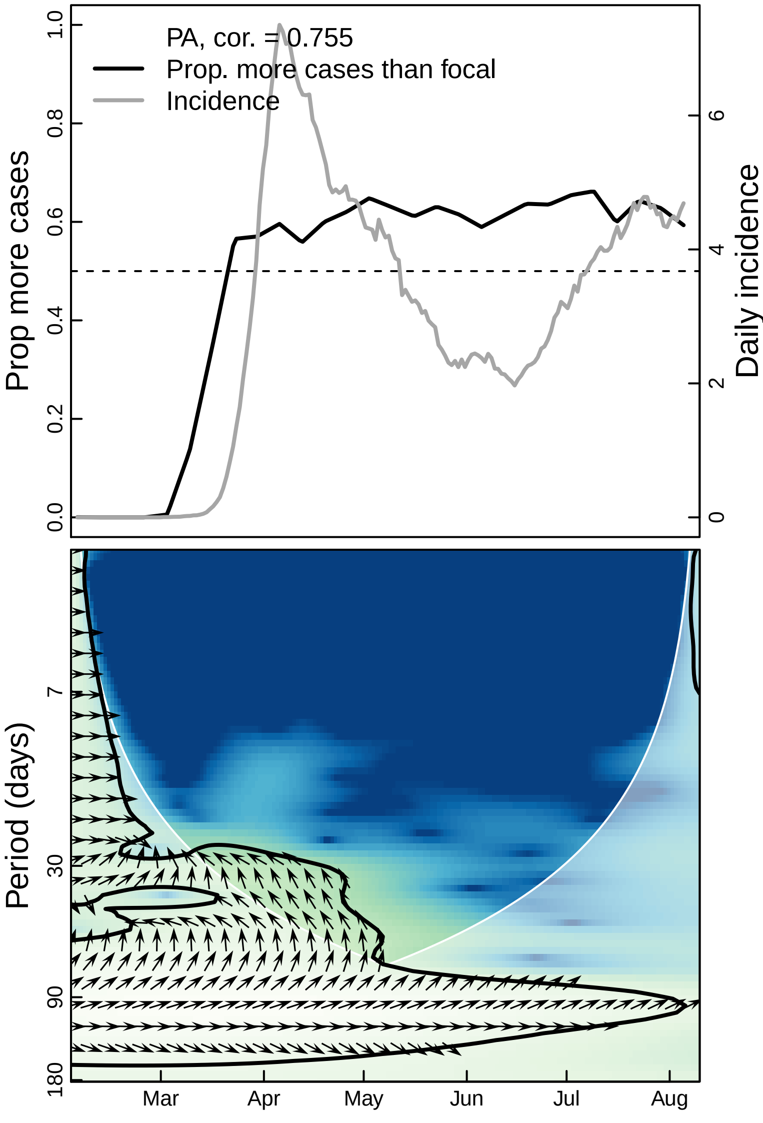

PA, cor. = 0.755
Prop. more cases than focal
Incidence
Prop more cases
Daily incidence
0.0
0.2
0.4
0.6
0.8
1.0
0
2
4
6
Period (days)
7
30
90
180
Mar
Apr
May
Jun
Jul
Aug

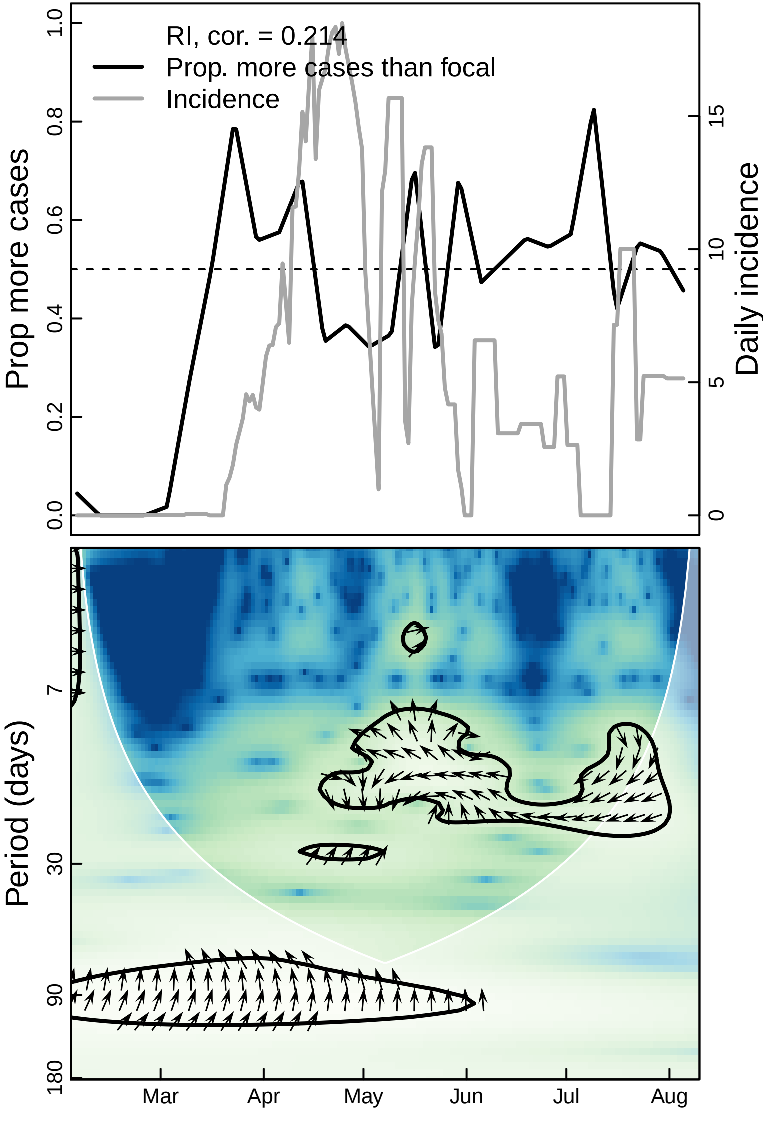

RI, cor. = 0.214
Prop. more cases than focal
Incidence
Prop more cases
Daily incidence
0.0
0.2
0.4
0.6
0.8
1.0
0
5
10
15
Period (days)
7
30
90
180
Mar
Apr
May
Jun
Jul
Aug

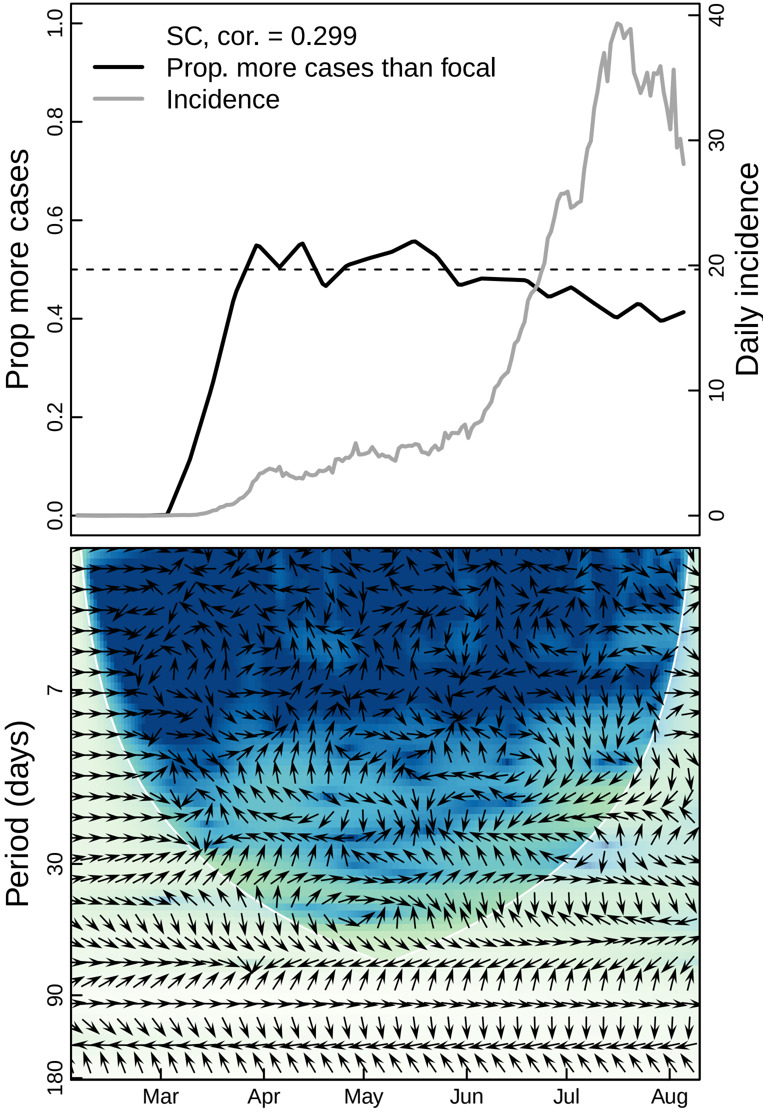

SC, cor. = 0.299
Prop. more cases than focal
Incidence
Prop more cases
Daily incidence
0.0
0.2
0.4
0.6
0.8
1.0
0
10
20
30
40
Period (days)
7
30
90
180
Mar
Apr
May
Jun
Jul
Aug

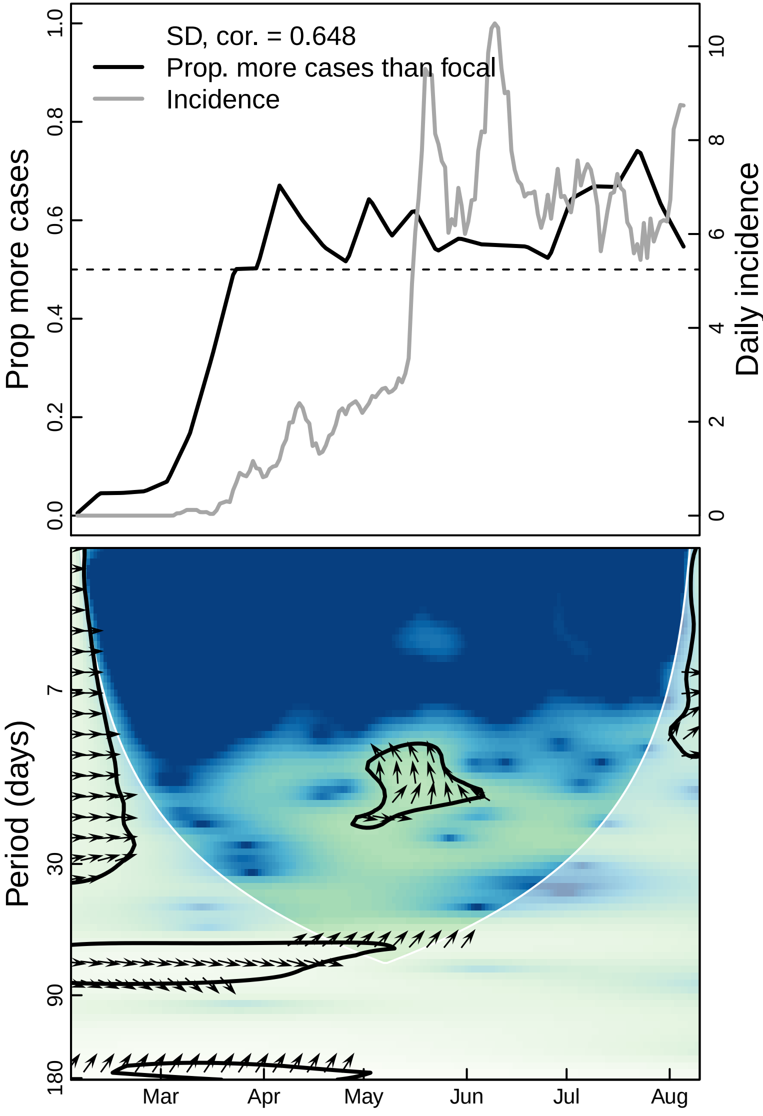
SD, cor. = 0.648
Prop. more cases than focal
Incidence
Prop more cases
Daily incidence
0.0
0.2
0.4
0.6
0.8
1.0
0
2
4
6
8
10
Period (days)
7
30
90
180
Mar
Apr
May
Jun
Jul
Aug

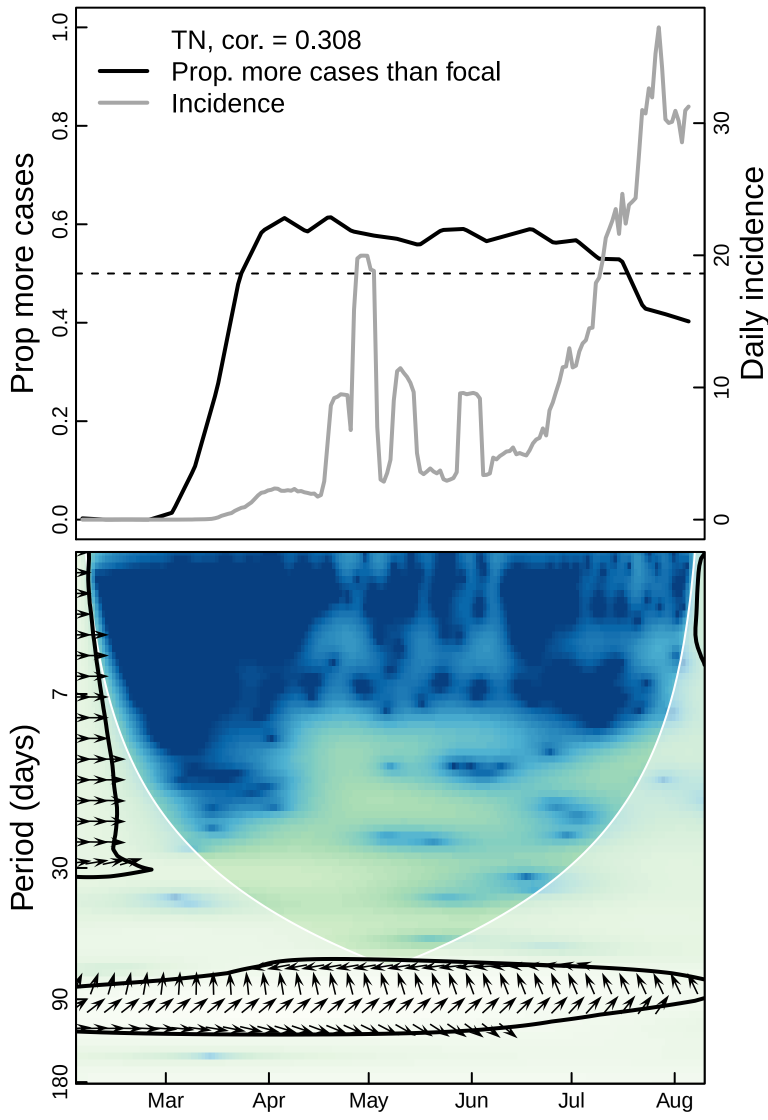

TN, cor. = 0.308
Prop. more cases than focal
Incidence
Prop more cases
Daily incidence
0.0
0.2
0.4
0.6
0.8
1.0
0
10
20
30
Period (days)
7
30
90
180
Mar
Apr
May
Jun
Jul
Aug

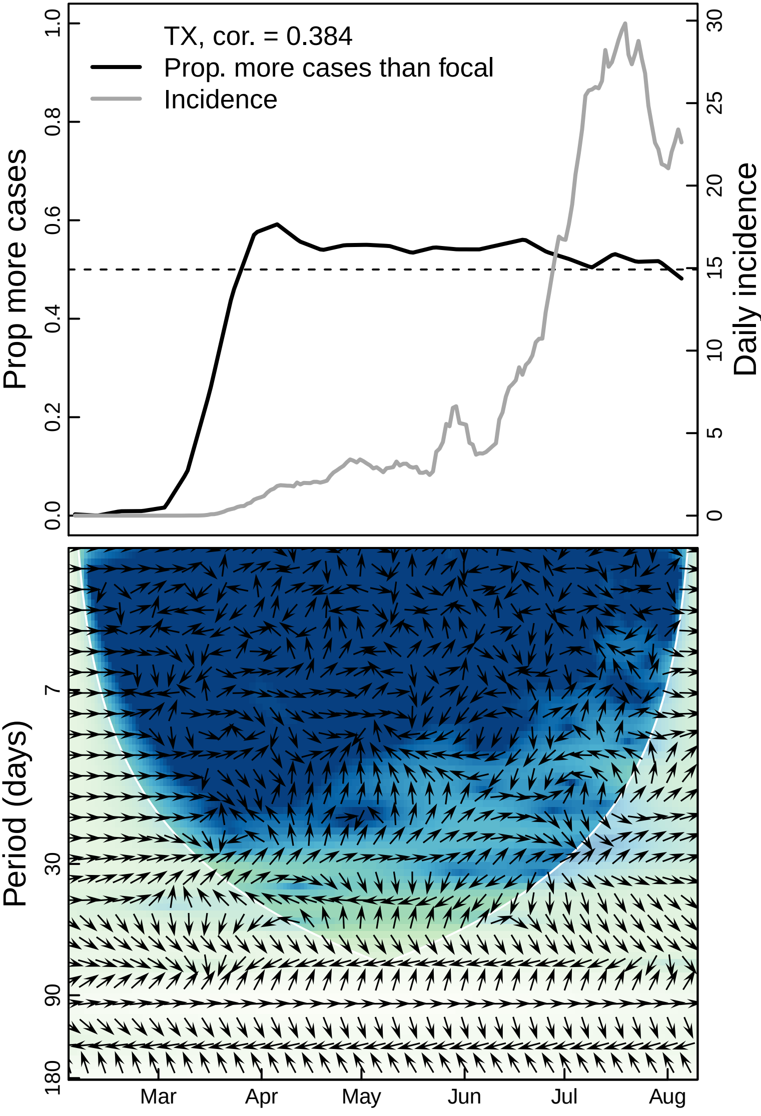
TX, cor. = 0.384
Prop. more cases than focal
Incidence
Prop more cases
Daily incidence
0.0
0.2
0.4
0.6
0.8
1.0
0
5
10
15
20
25
30
Period (days)
7
30
90
180
Mar
Apr
May
Jun
Jul
Aug

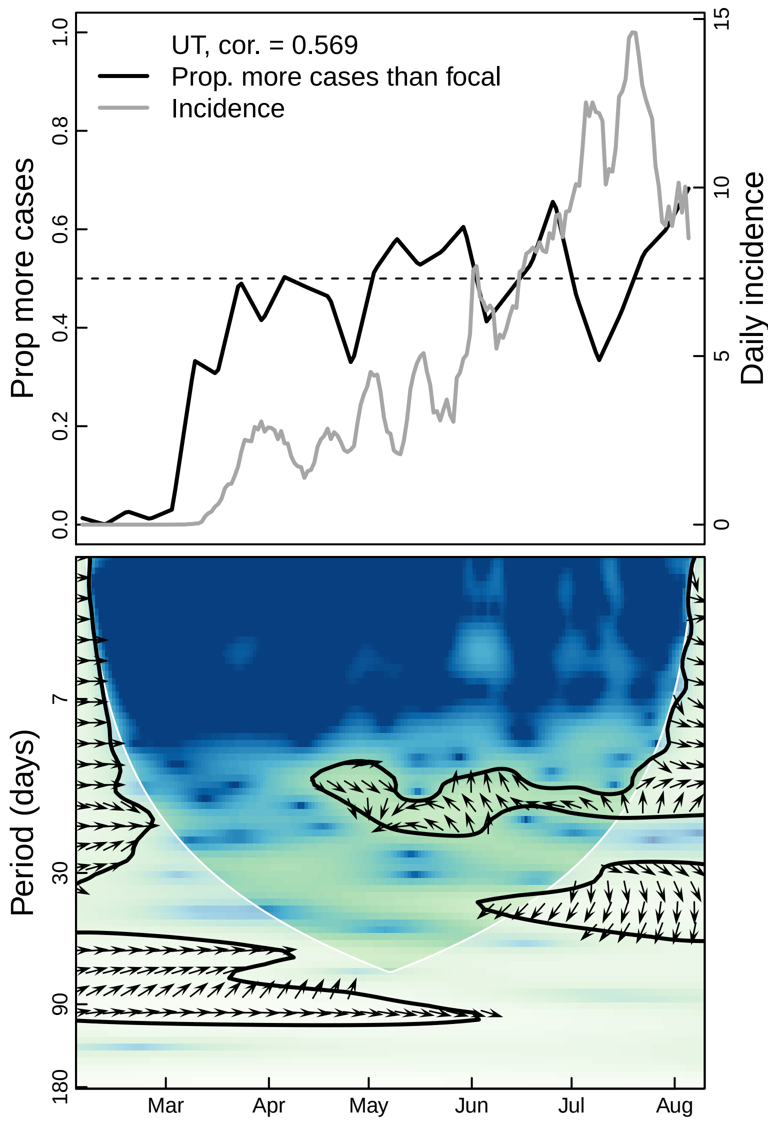
UT, cor. = 0.569
Prop. more cases than focal
Incidence
Prop more cases
Daily incidence
0.0
0.2
0.4
0.6
0.8
1.0
0
5
10
15
Period (days)
7
30
90
180
Mar
Apr
May
Jun
Jul
Aug

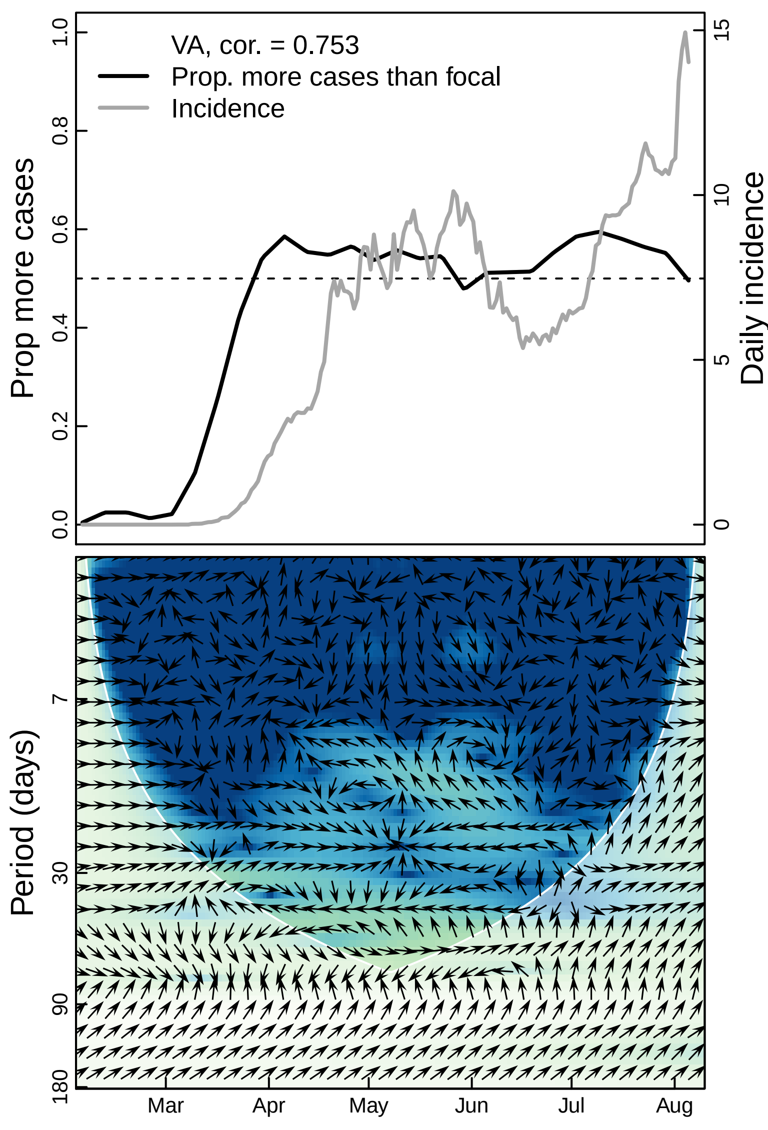
VA, cor. = 0.753
Prop. more cases than focal
Incidence
Prop more cases
Daily incidence
0.0
0.2
0.4
0.6
0.8
1.0
0
5
10
15
Period (days)
7
30
90
180
Mar
Apr
May
Jun
Jul
Aug

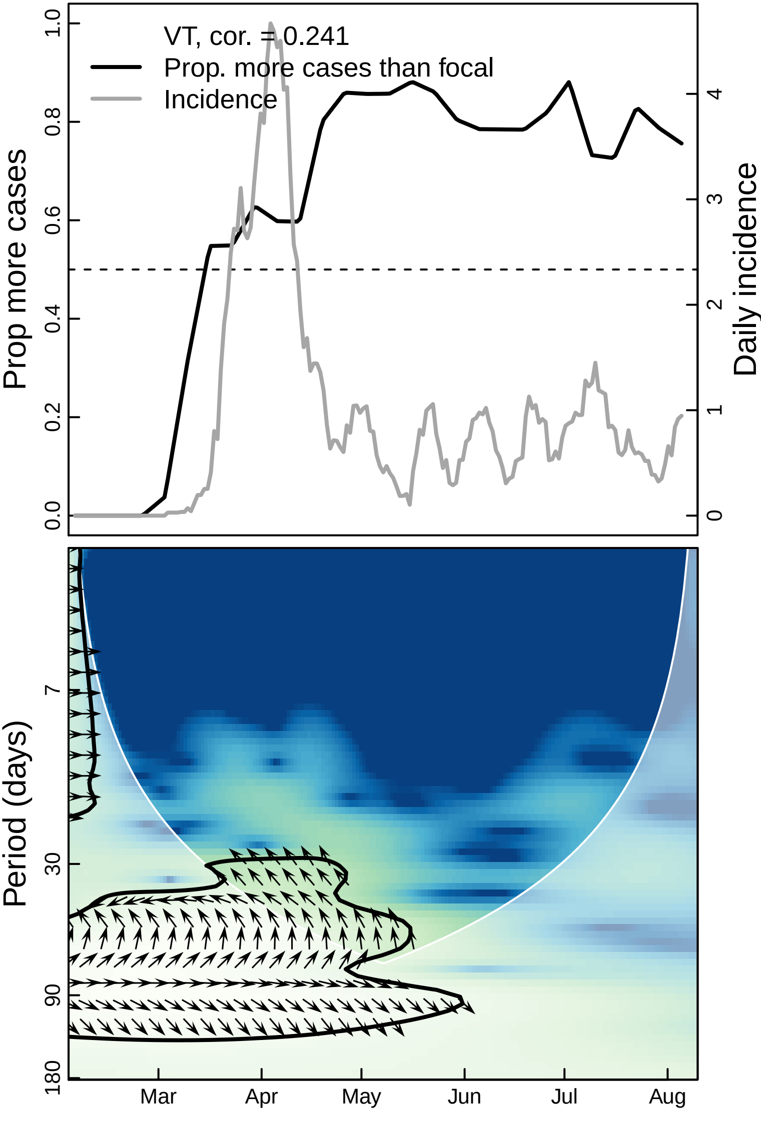

VT, cor. = 0.241
Prop. more cases than focal
Incidence
Prop more cases
Daily incidence
0.0
0.2
0.4
0.6
0.8
1.0
0
1
2
3
4
Period (days)
7
30
90
180
Mar
Apr
May
Jun
Jul
Aug

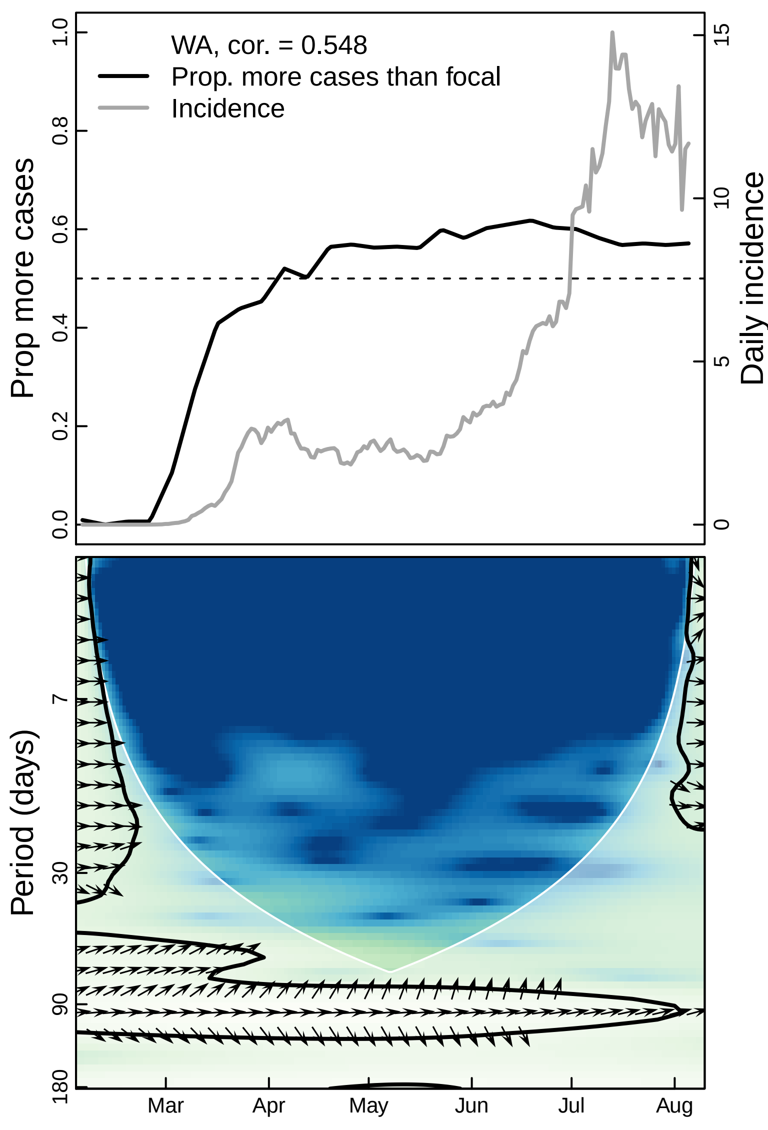

WA, cor. = 0.548
Prop. more cases than focal
Incidence
Prop more cases
Daily incidence
0.0
0.2
0.4
0.6
0.8
1.0
0
5
10
15
Period (days)
7
30
90
180
Mar
Apr
May
Jun
Jul
Aug

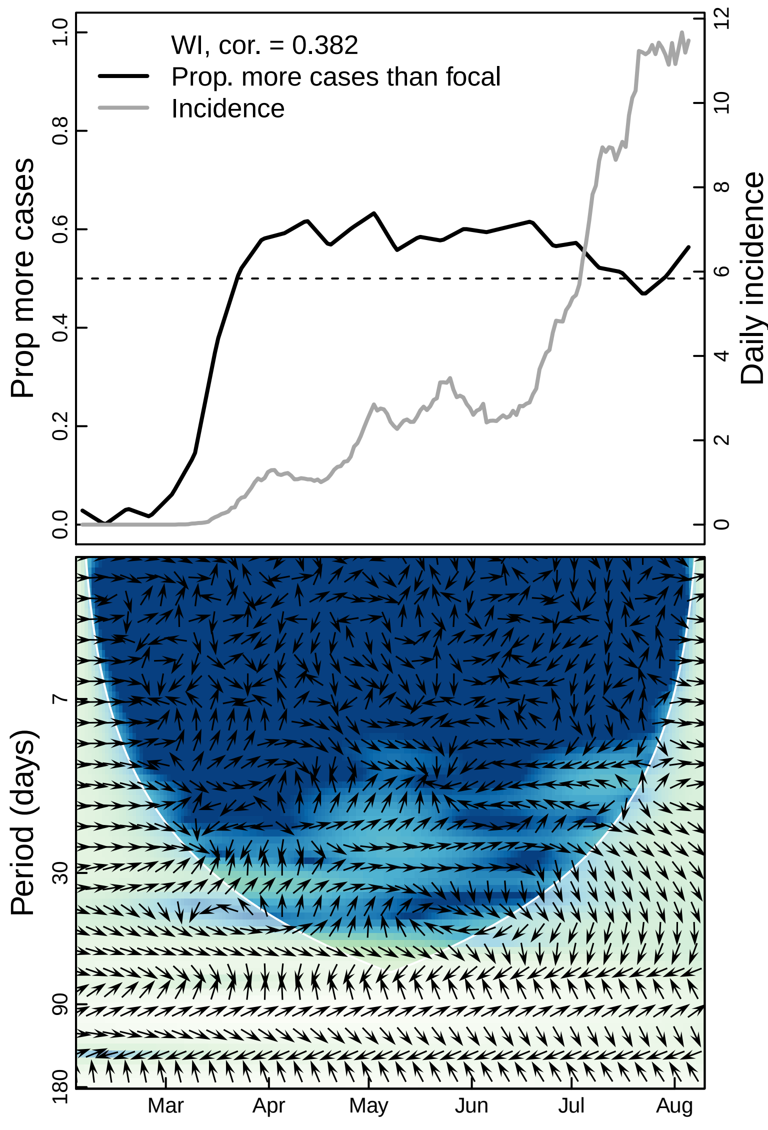

WI, cor. = 0.382
Prop. more cases than focal
Incidence
Prop more cases
Daily incidence
0.0
0.2
0.4
0.6
0.8
1.0
0
2
4
6
8
10
12
Period (days)
7
30
90
180
Mar
Apr
May
Jun
Jul
Aug

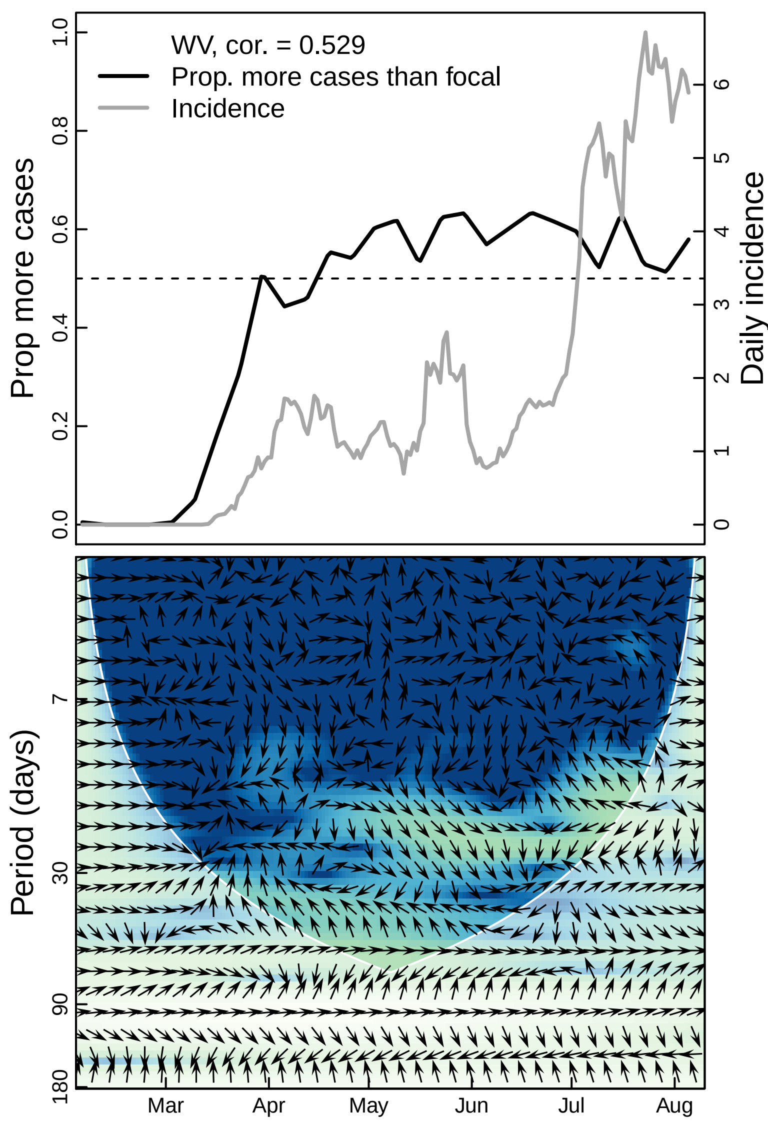
WV, cor. = 0.529
Prop. more cases than focal
Incidence
Prop more cases
Daily incidence
0.0
0.2
0.4
0.6
0.8
1.0
0
1
2
3
4
5
6
Period (days)
7
30
90
180
Mar
Apr
May
Jun
Jul
Aug

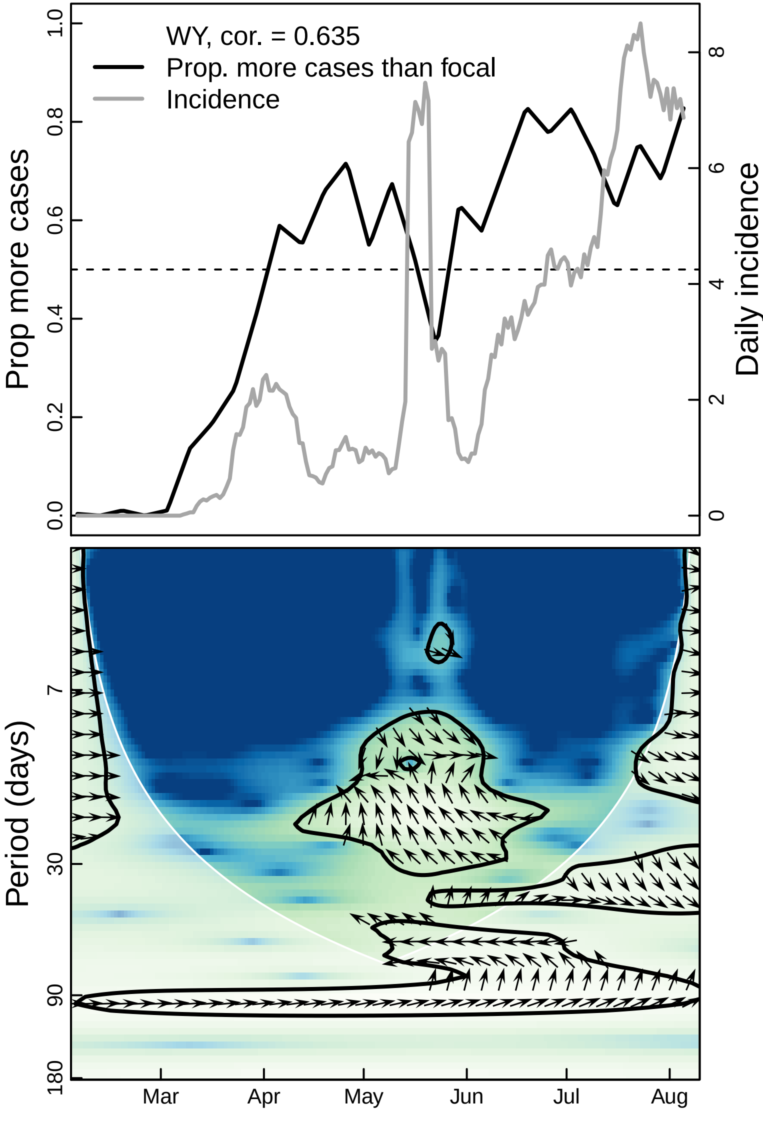

WY, cor. = 0.635
Prop. more cases than focal
Incidence
Prop more cases
Daily incidence
0.0
0.2
0.4
0.6
0.8
1.0
0
2
4
6
8
Period (days)
7
30
90
180
Mar
Apr
May
Jun
Jul
Aug
