## Supplementary material for "The unintended consequences of inconsistent pandemic control policies": Cross-wavelets - parks

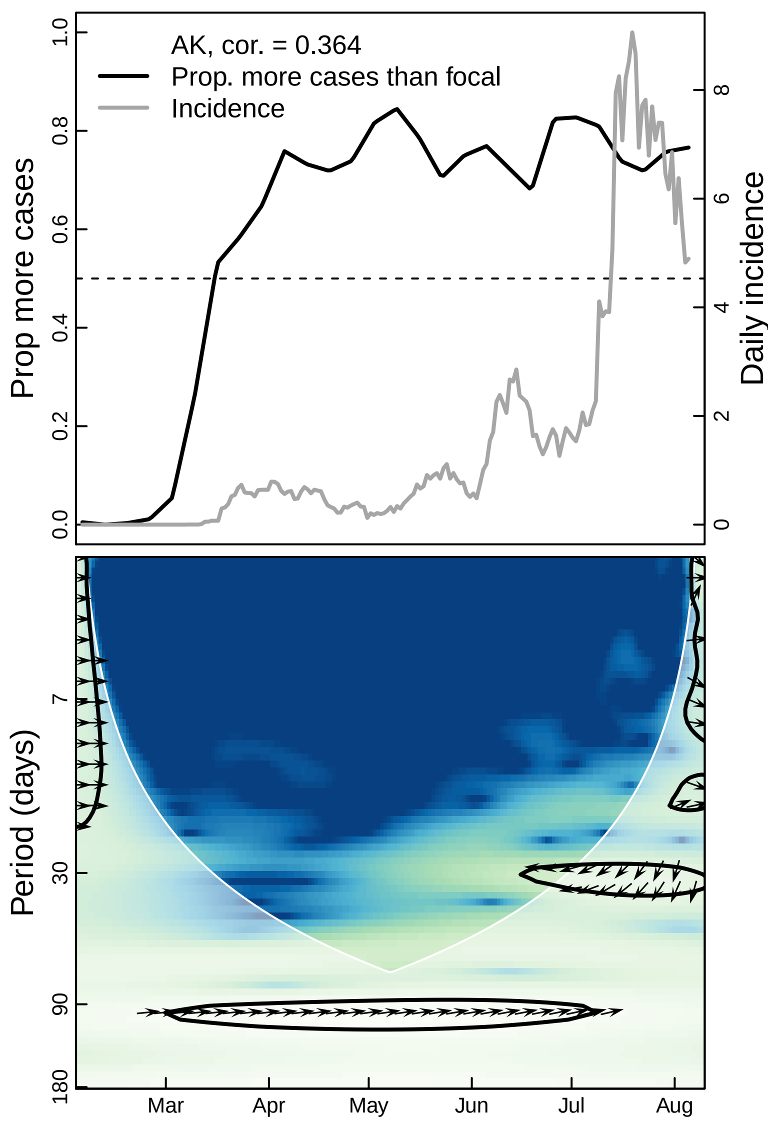
AK, cor. = 0.364
Prop. more cases than focal
Incidence
Prop more cases
Daily incidence
0.0
0.2
0.4
0.6
0.8
1.0
0
2
4
6
8
Period (days)
7
30
90
180
Mar
Apr
May
Jun
Jul
Aug

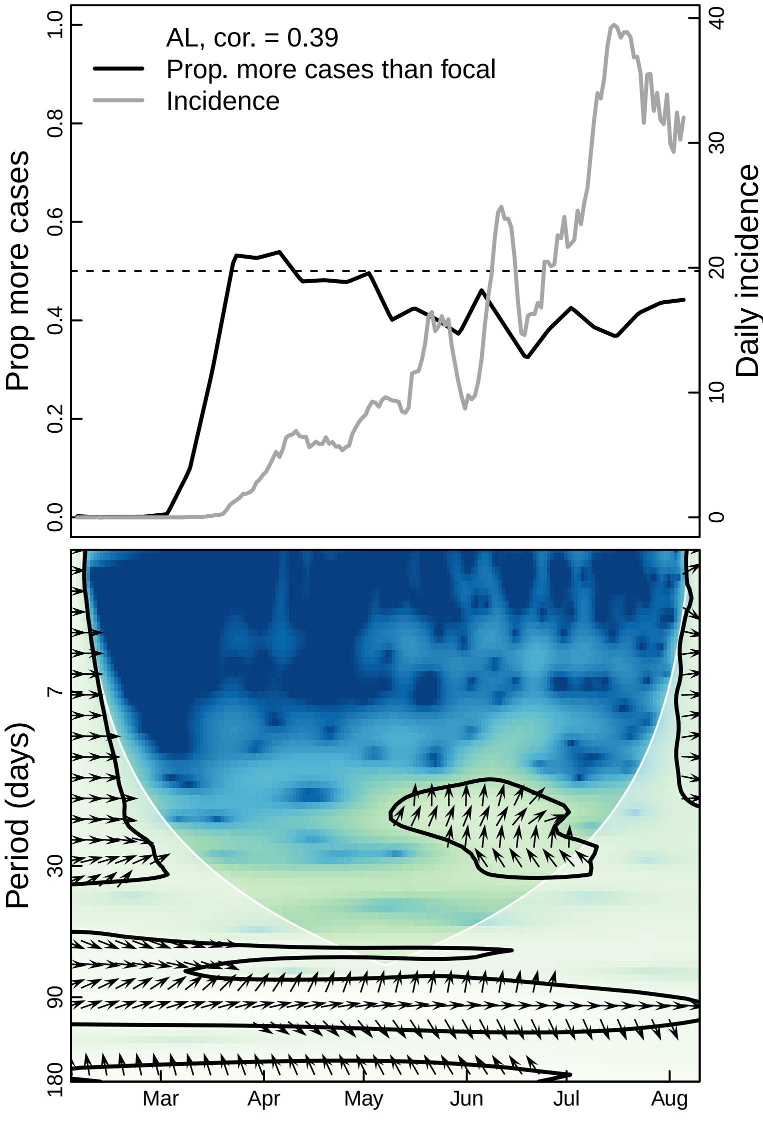

AL, cor. = 0.39
Prop. more cases than focal
Incidence
Prop more cases
Daily incidence
1.0
0.8
0.6
0.4
0.2
0.0
40
30
20
10
0
Period (days)
7
30
90
180
Mar
Apr
May
Jun
Jul
Aug

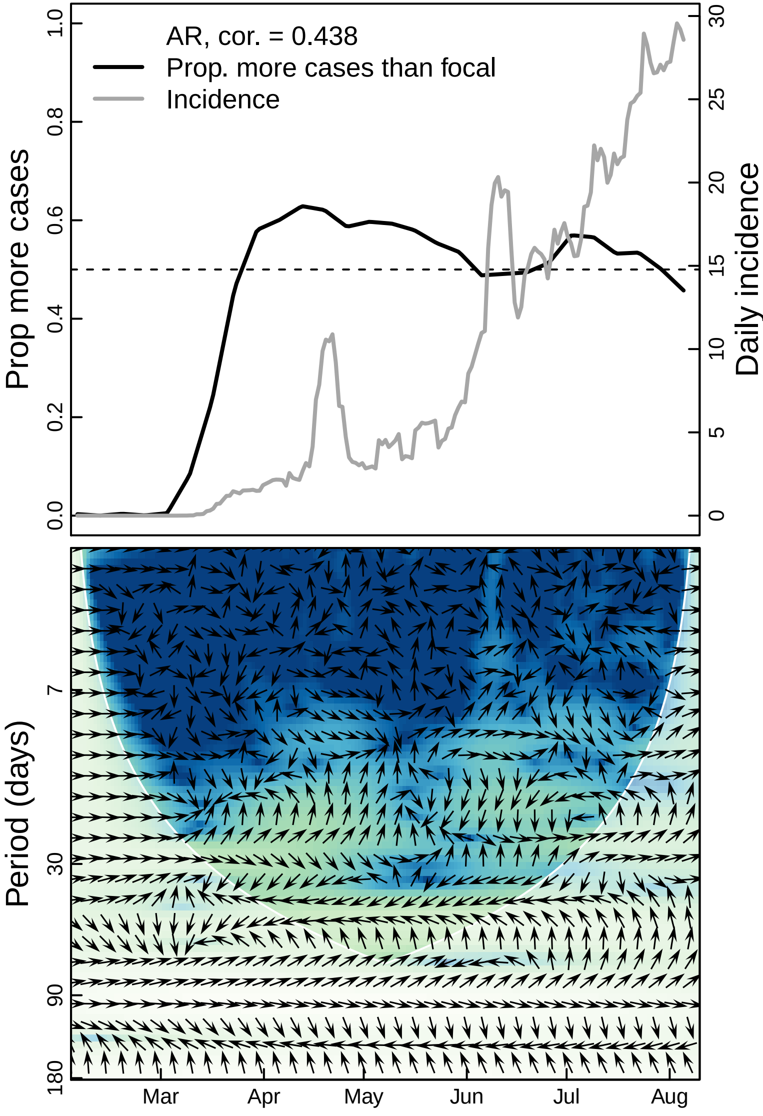

AR, cor. = 0.438
Prop. more cases than focal
Incidence
Prop more cases
Daily incidence
0.0
0.2
0.4
0.6
0.8
1.0
0
5
10
15
20
25
30
Period (days)
7
30
90
180
Mar
Apr
May
Jun
Jul
Aug

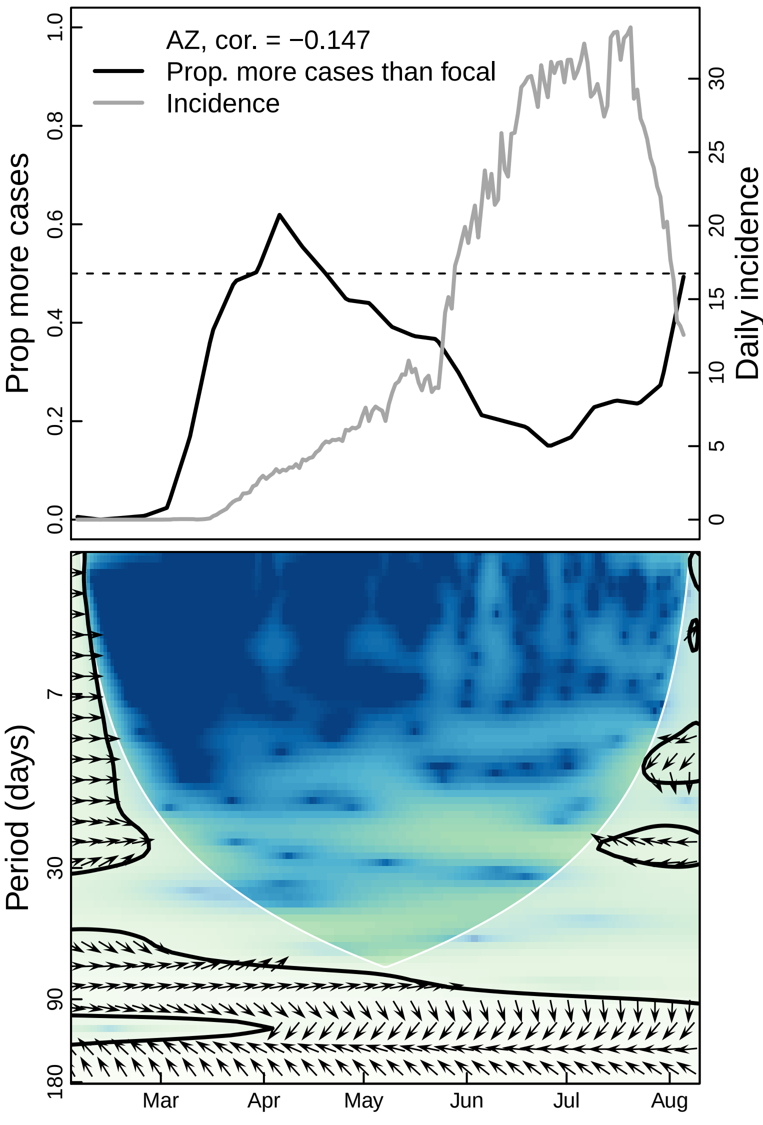

AZ, cor. = −0.147
Prop. more cases than focal
Incidence
Prop more cases
Daily incidence
0.0
0.2
0.4
0.6
0.8
1.0
0
5
10
15
20
25
30
Period (days)
7
30
90
180
Mar
Apr
May
Jun
Jul
Aug

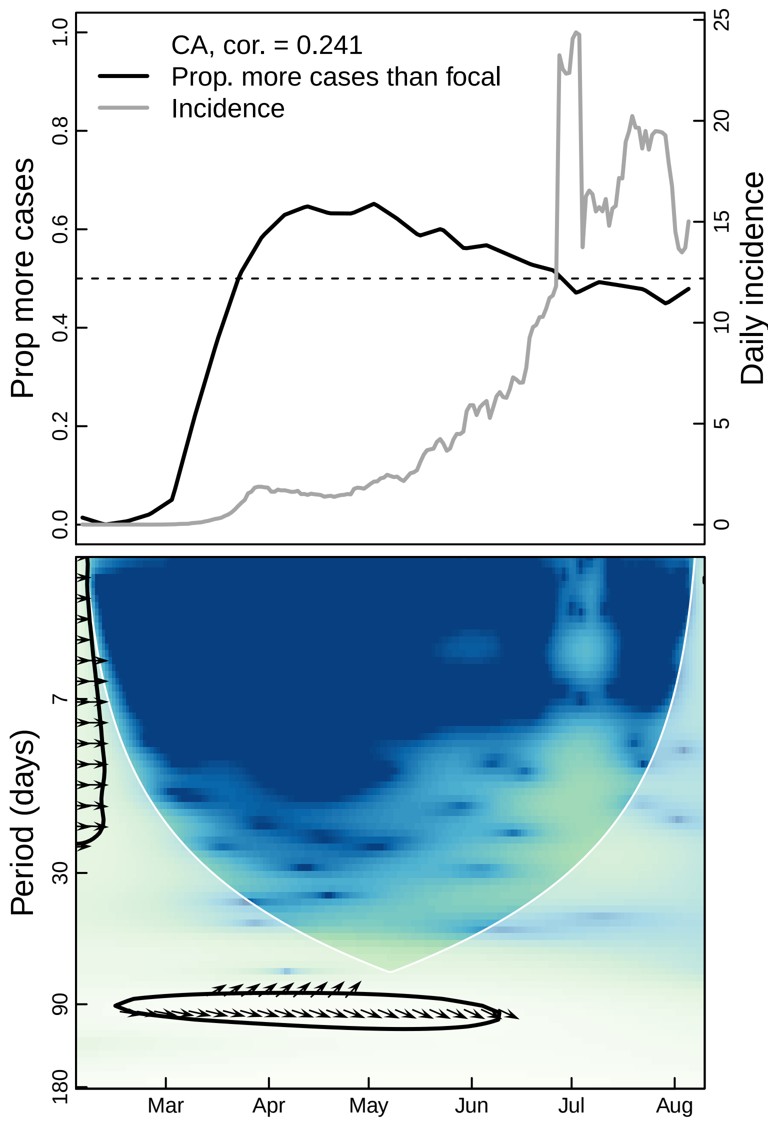

CA, cor. = 0.241
Prop. more cases than focal
Incidence
Prop more cases
Daily incidence
0.0
0.2
0.4
0.6
0.8
1.0
0
5
10
15
20
25
Period (days)
7
30
90
180
Mar
Apr
May
Jun
Jul
Aug

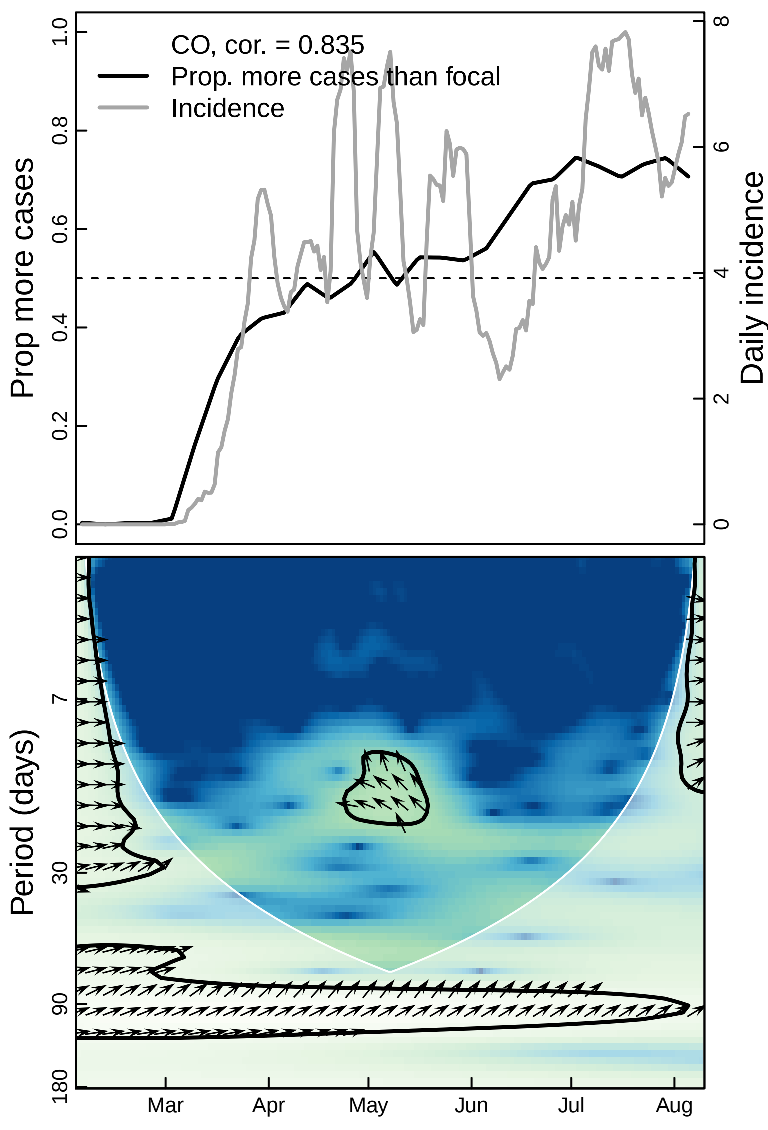
CO, cor. = 0.835
Prop. more cases than focal
Incidence
Prop more cases
Daily incidence
0.0
0.2
0.4
0.6
0.8
1.0
0
2
4
6
8
Period (days)
7
30
90
180
Mar
Apr
May
Jun
Jul
Aug

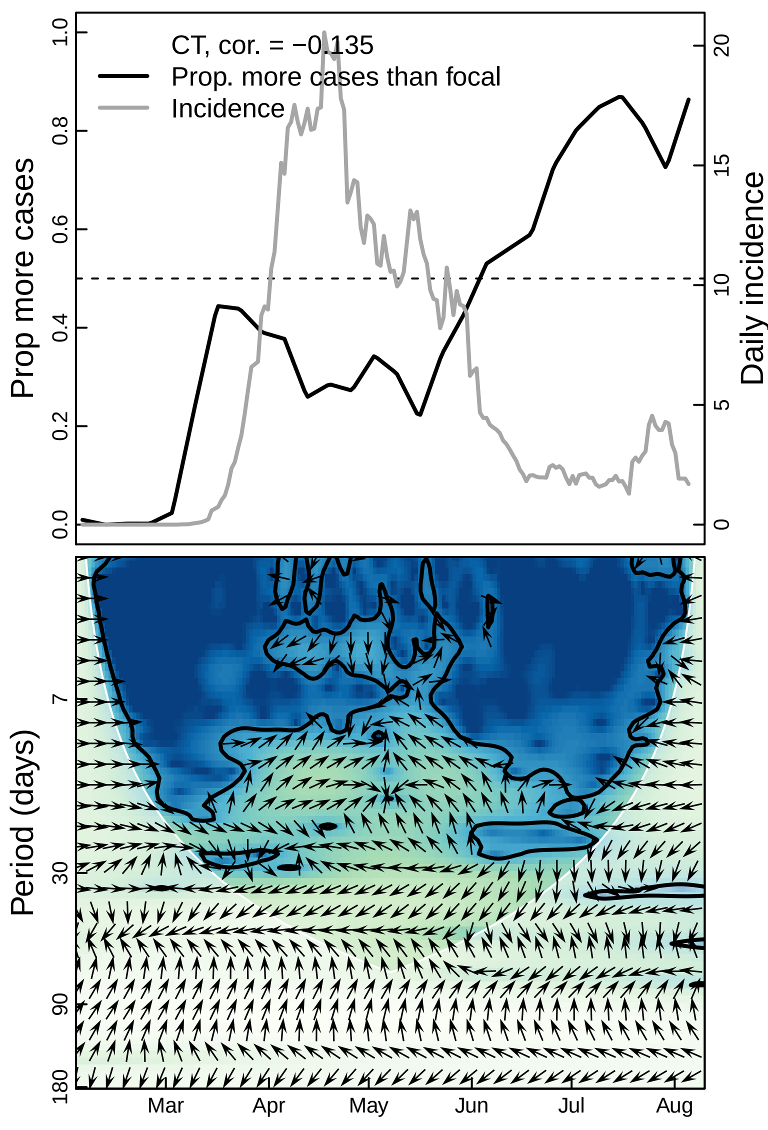

CT, cor. = −0.135
Prop. more cases than focal
Incidence
Prop more cases
Daily incidence
0.0
0.2
0.4
0.6
0.8
1.0
0
5
10
15
20
Period (days)
7
30
90
180
Mar
Apr
May
Jun
Jul
Aug

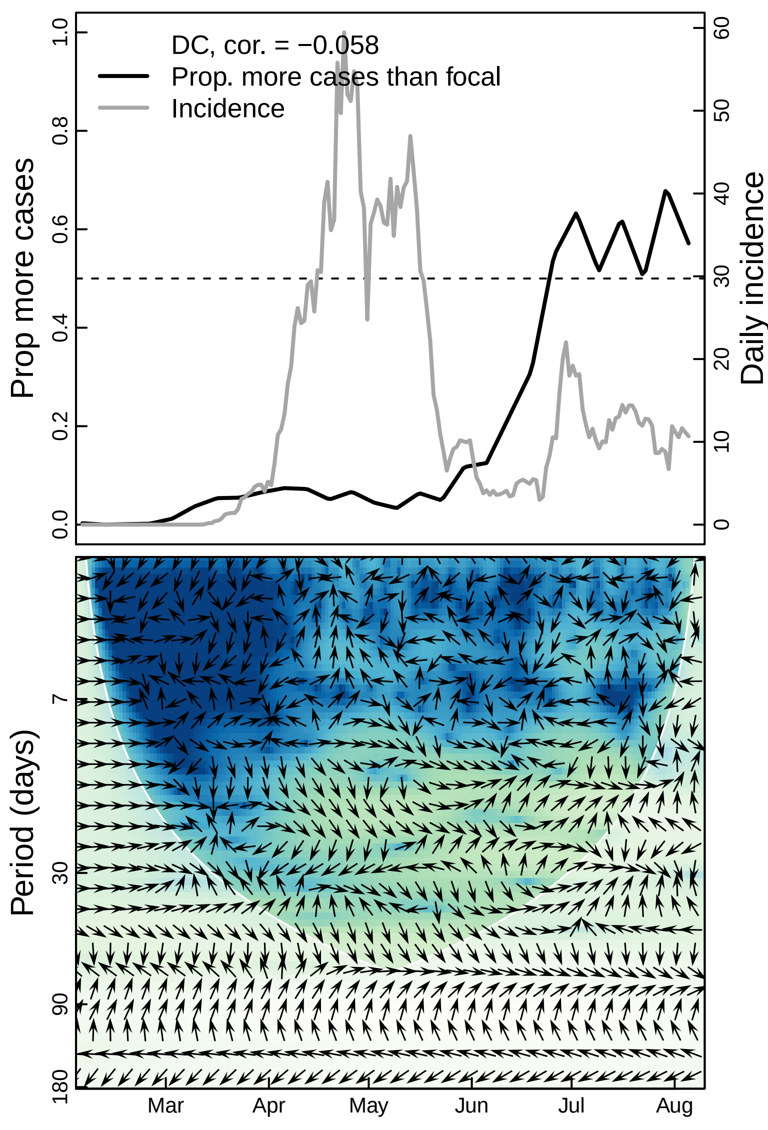

DC, cor. = −0.058
Prop. more cases than focal
Incidence
Prop more cases
Daily incidence
0.0
0.2
0.4
0.6
0.8
1.0
0
10
20
30
40
50
60
Period (days)
7
30
90
180
Mar
Apr
May
Jun
Jul
Aug

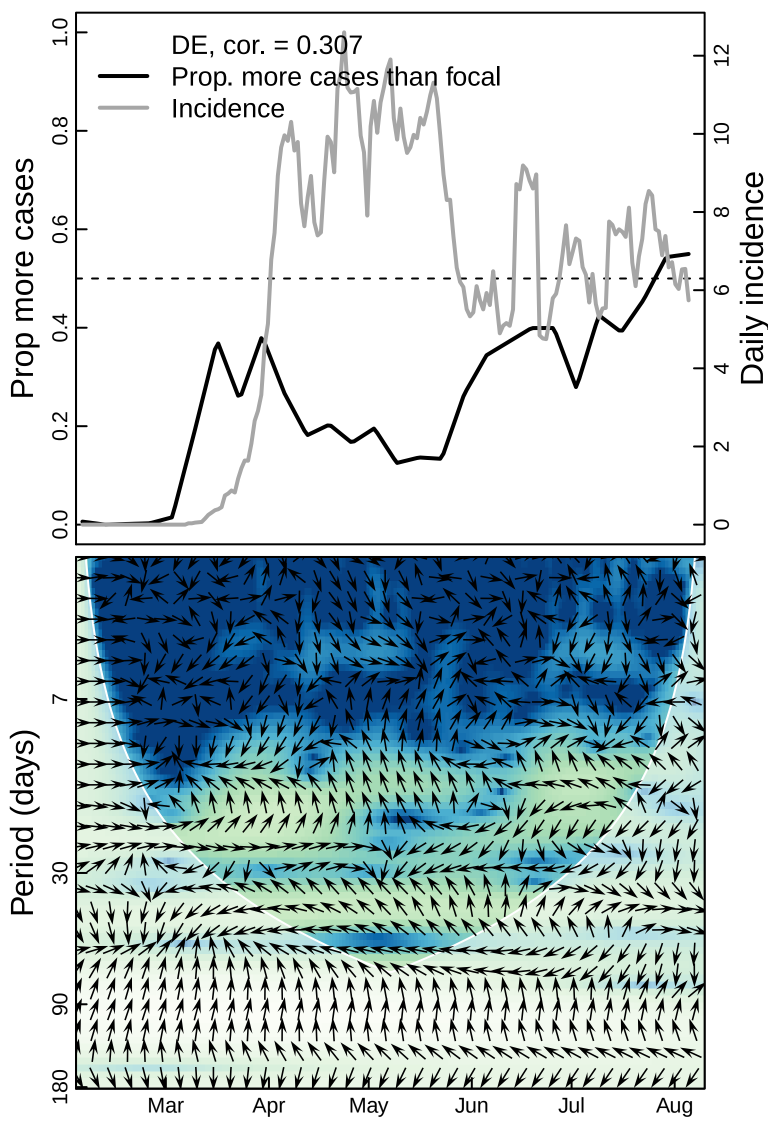

DE, cor. = 0.307
Prop. more cases than focal
Incidence
Prop more cases
Daily incidence
0.0
0.2
0.4
0.6
0.8
1.0
0
2
4
6
8
10
12
Period (days)
7
30
90
180
Mar
Apr
May
Jun
Jul
Aug

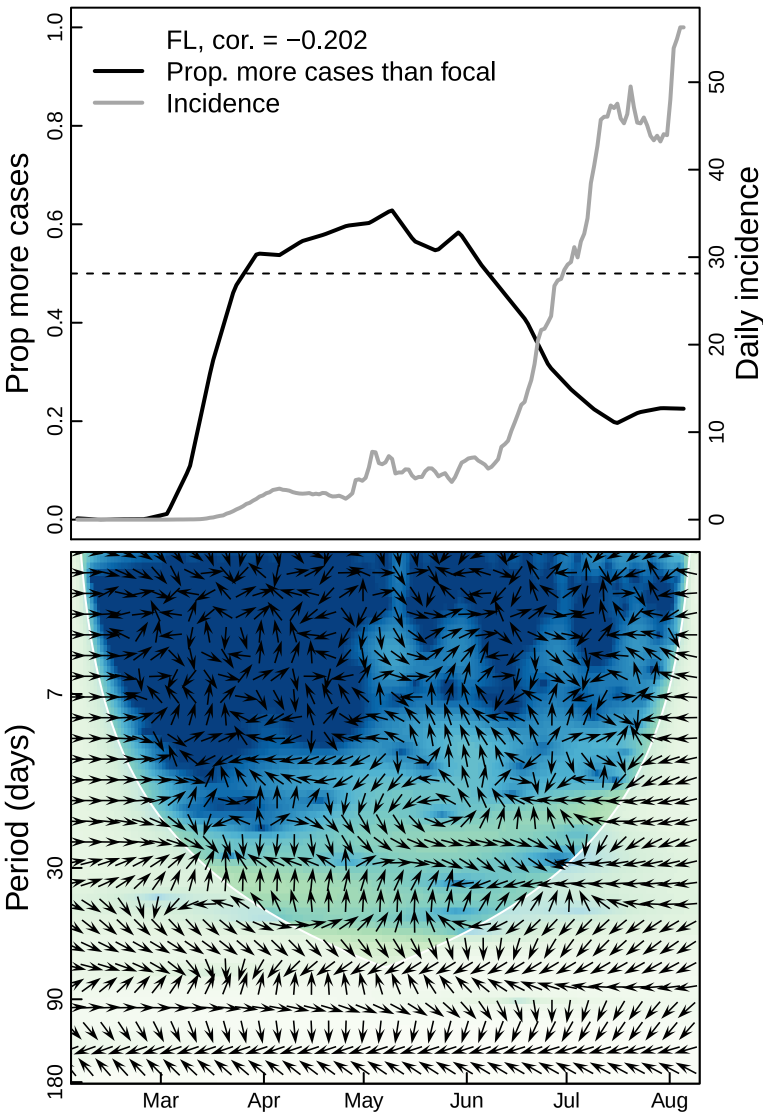
FL, cor. = −0.202
Prop. more cases than focal
Incidence
Prop more cases
Daily incidence
0.0
0.2
0.4
0.6
0.8
1.0
0
10
20
30
40
50
Period (days)
7
30
90
180
Mar
Apr
May
Jun
Jul
Aug

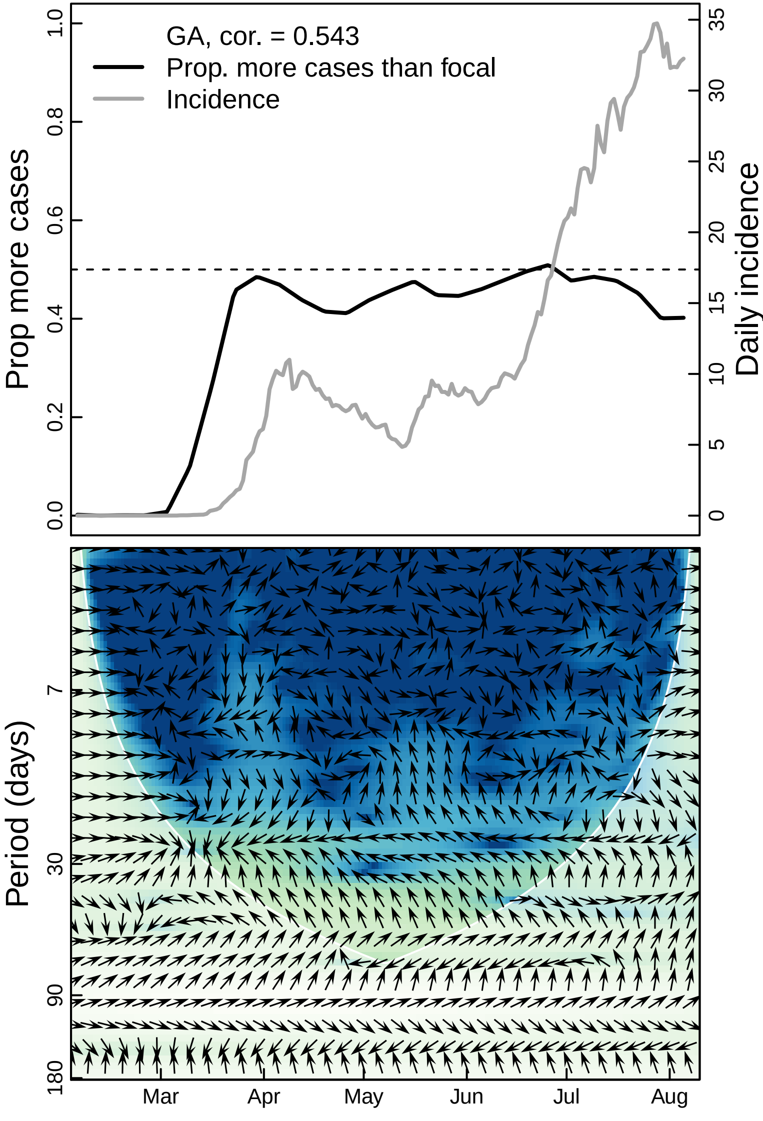

GA, cor. = 0.543
Prop. more cases than focal
Incidence
Prop more cases
Daily incidence
0.0
0.2
0.4
0.6
0.8
1.0
0
5
10
15
20
25
30
35
Period (days)
7
30
90
180
Mar
Apr
May
Jun
Jul
Aug

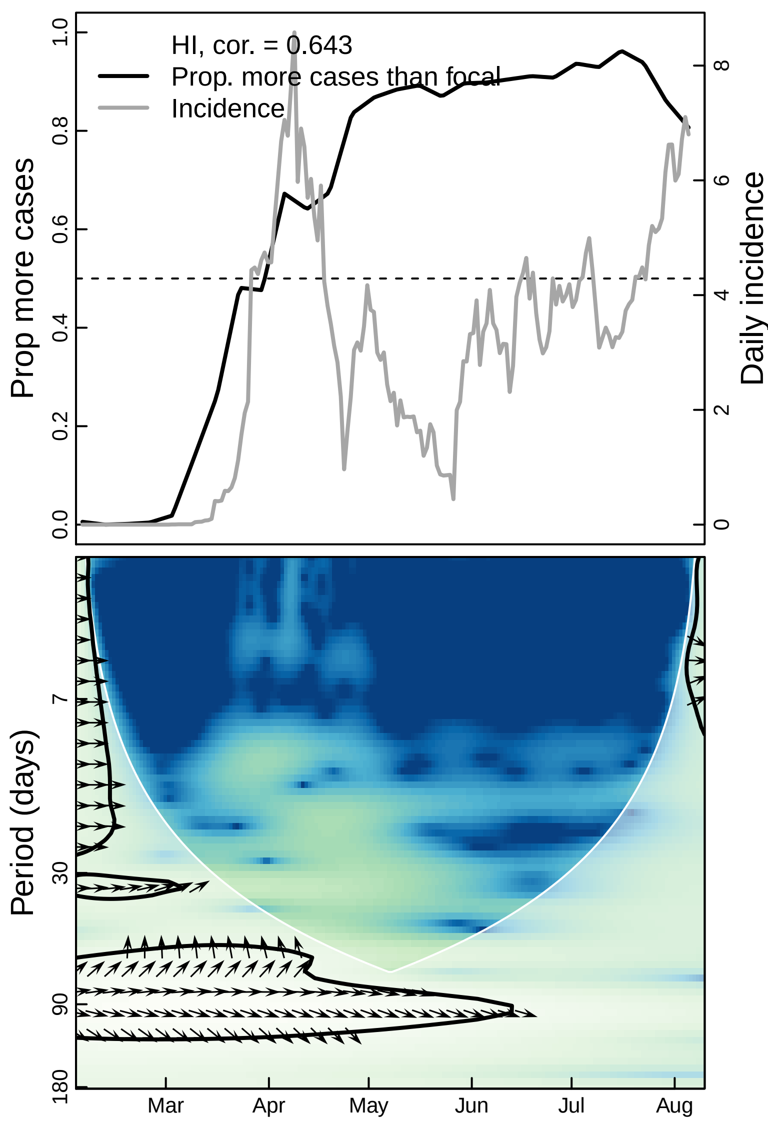

HI, cor. = 0.643
Prop. more cases than focal
Incidence
Prop more cases
Daily incidence
0.0
0.2
0.4
0.6
0.8
1.0
0
2
4
6
8
Period (days)
7
30
90
180
Mar
Apr
May
Jun
Jul
Aug

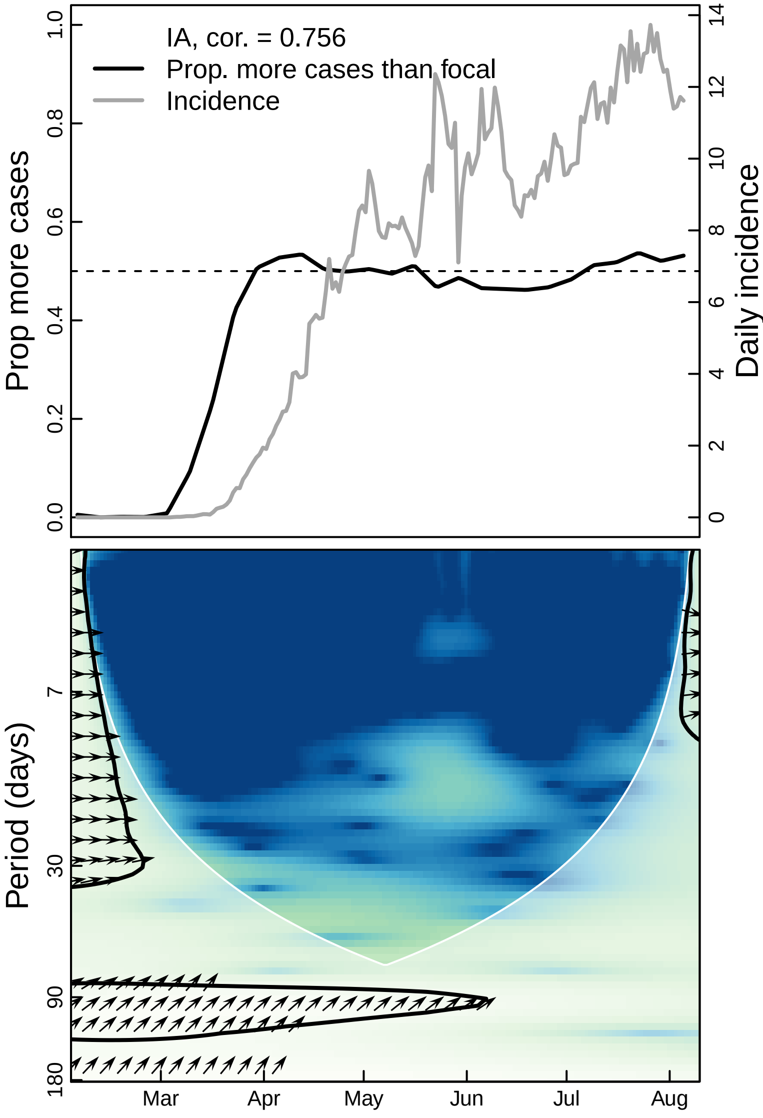

IA, cor. = 0.756
Prop. more cases than focal
Incidence
Prop more cases
Daily incidence
0.0
0.2
0.4
0.6
0.8
1.0
0
2
4
6
8
10
12
14
Period (days)
7
30
90
180
Mar
Apr
May
Jun
Jul
Aug

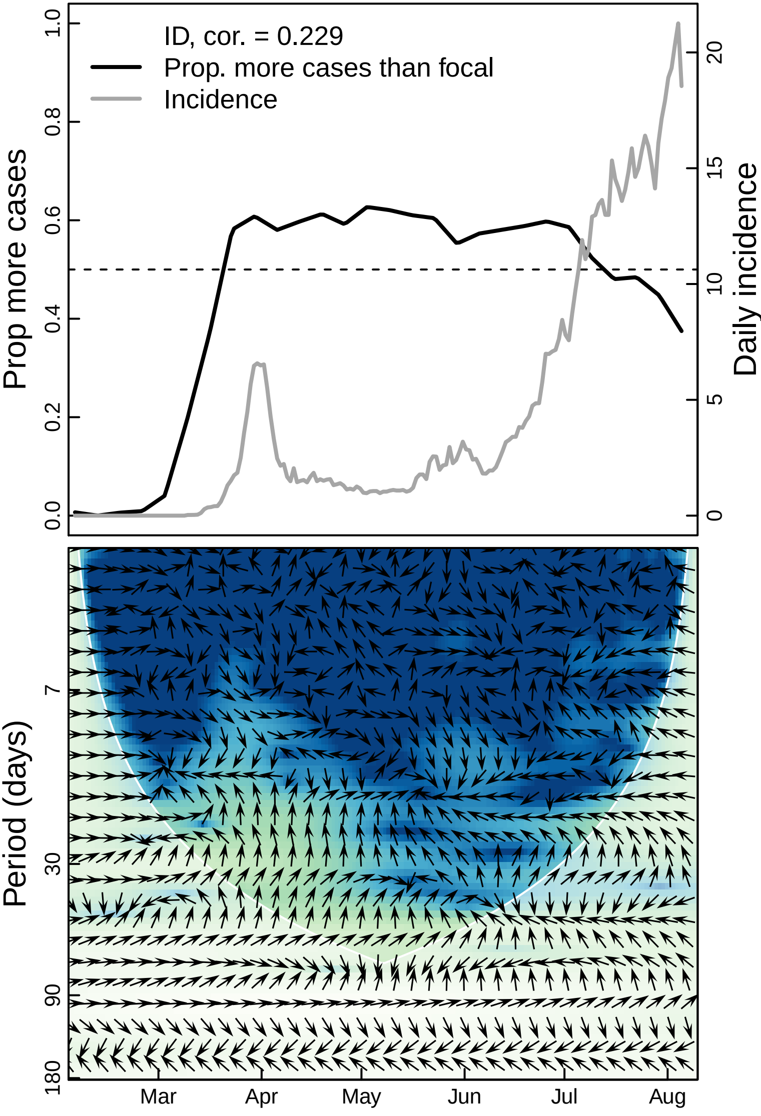

ID, cor. = 0.229
Prop. more cases than focal
Incidence
Prop more cases
Daily incidence
0.0
0.2
0.4
0.6
0.8
1.0
0
5
10
15
20
Period (days)
7
30
90
180
Mar
Apr
May
Jun
Jul
Aug

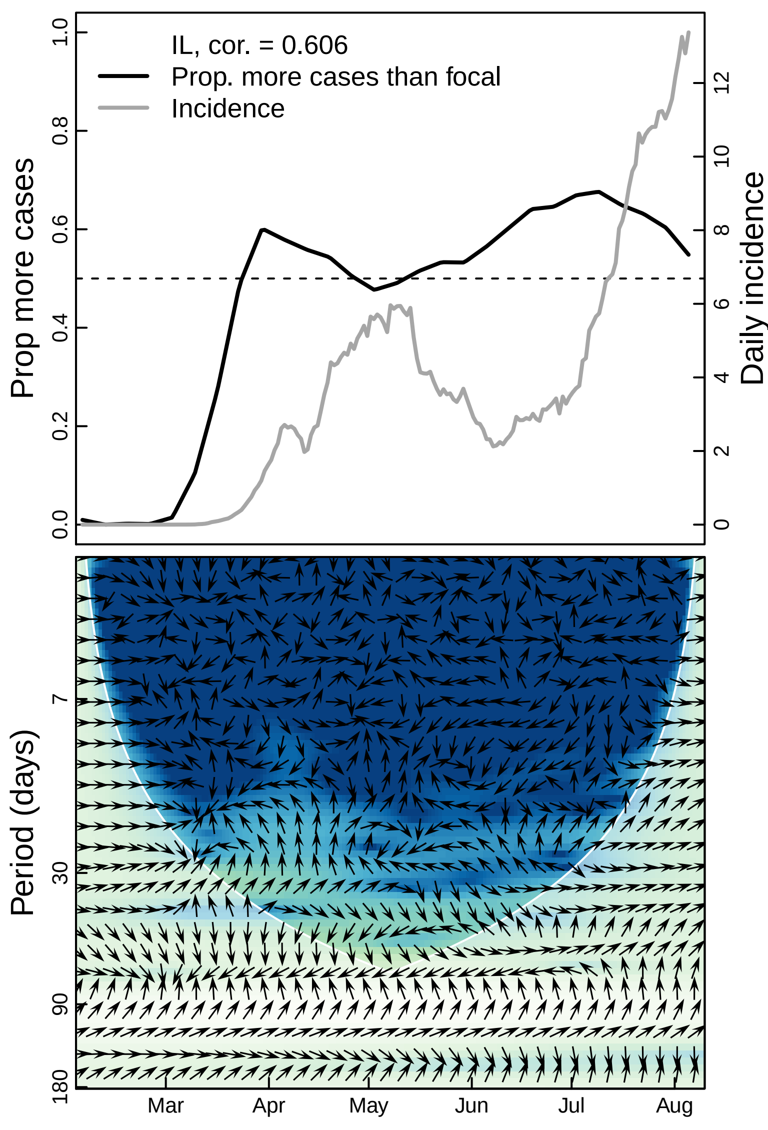

IL, cor. = 0.606
Prop. more cases than focal
Incidence
Prop more cases
Daily incidence
0.0
0.2
0.4
0.6
0.8
1.0
0
2
4
6
8
10
12
Period (days)
7
30
90
180
Mar
Apr
May
Jun
Jul
Aug

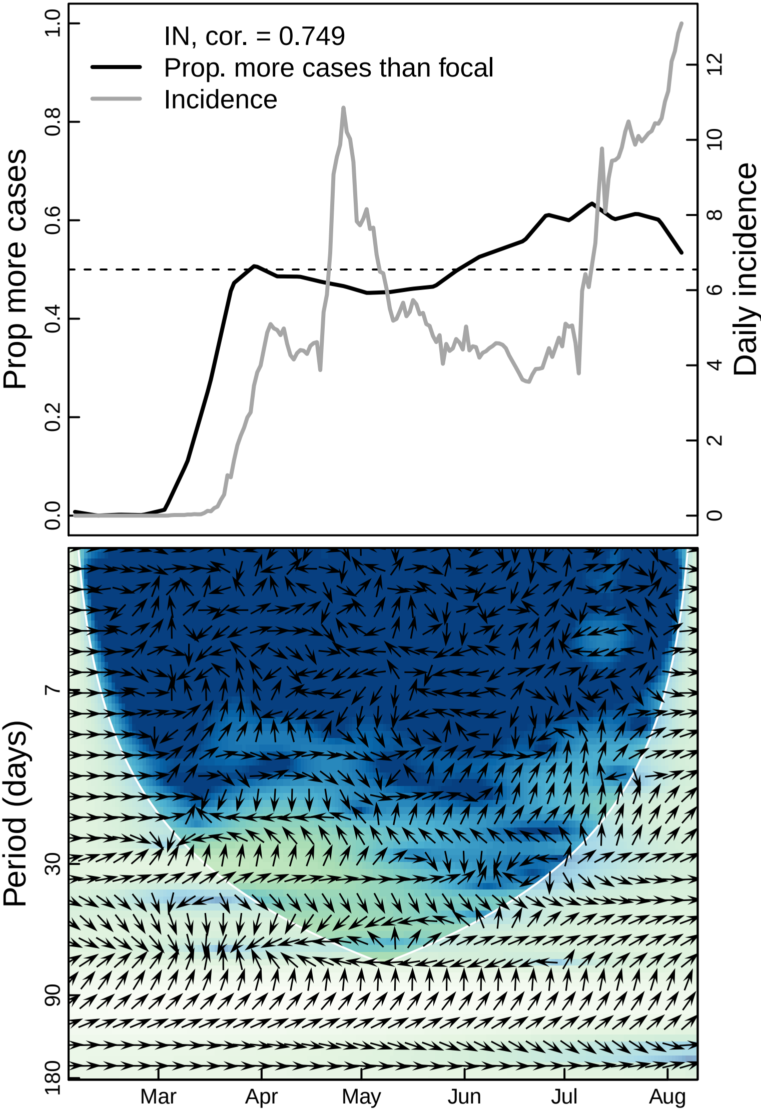

IN, cor. = 0.749
Prop. more cases than focal
Incidence
Prop more cases
Daily incidence
0.0
0.2
0.4
0.6
0.8
1.0
0
2
4
6
8
10
12
Period (days)
7
30
90
180
Mar
Apr
May
Jun
Jul
Aug

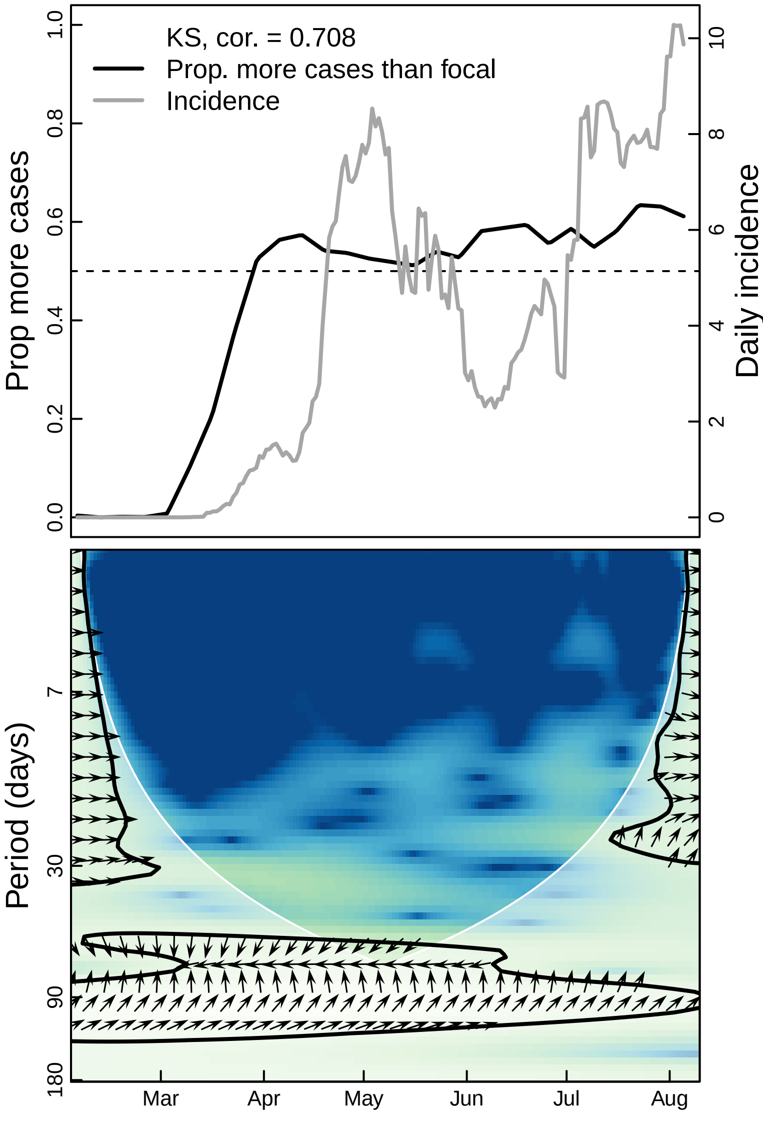

KS, cor. = 0.708
Prop. more cases than focal
Incidence
Prop more cases
Daily incidence
0.0
0.2
0.4
0.6
0.8
1.0
0
2
4
6
8
10
Period (days)
7
30
90
180
Mar
Apr
May
Jun
Jul
Aug

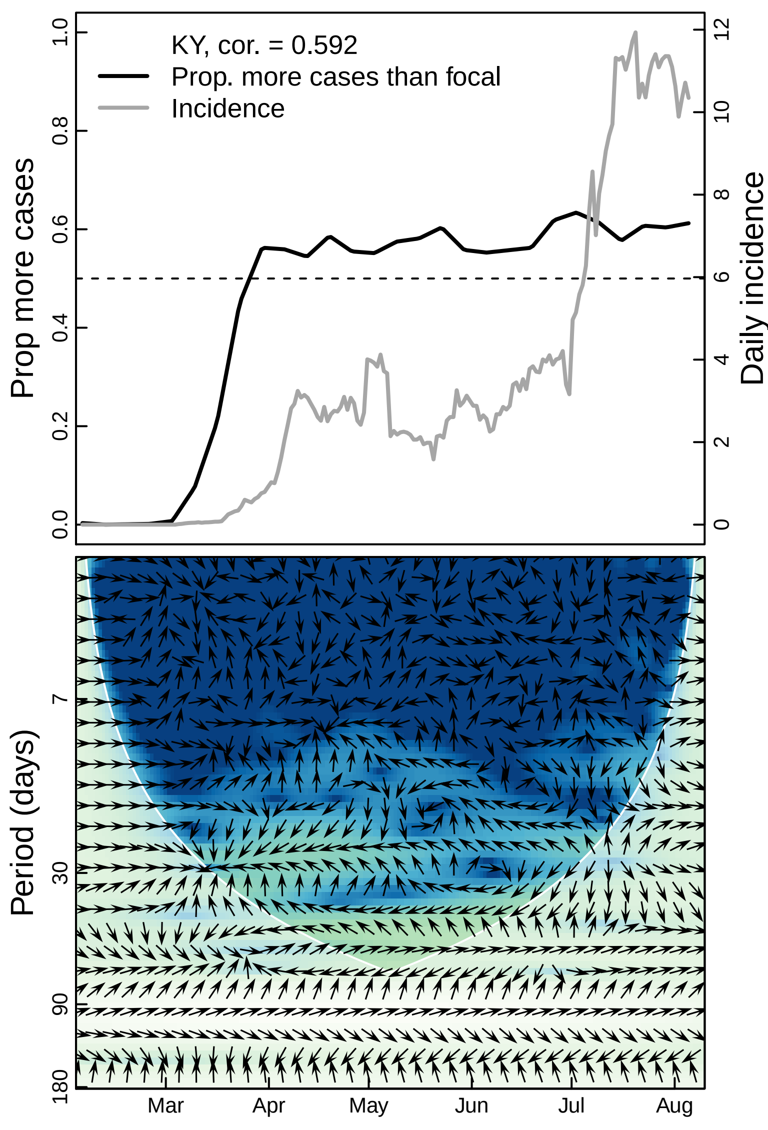

KY, cor. = 0.592
Prop. more cases than focal
Incidence
Prop more cases
Daily incidence
0.0
0.2
0.4
0.6
0.8
1.0
0
2
4
6
8
10
12
Period (days)
7
30
90
180
Mar
Apr
May
Jun
Jul
Aug

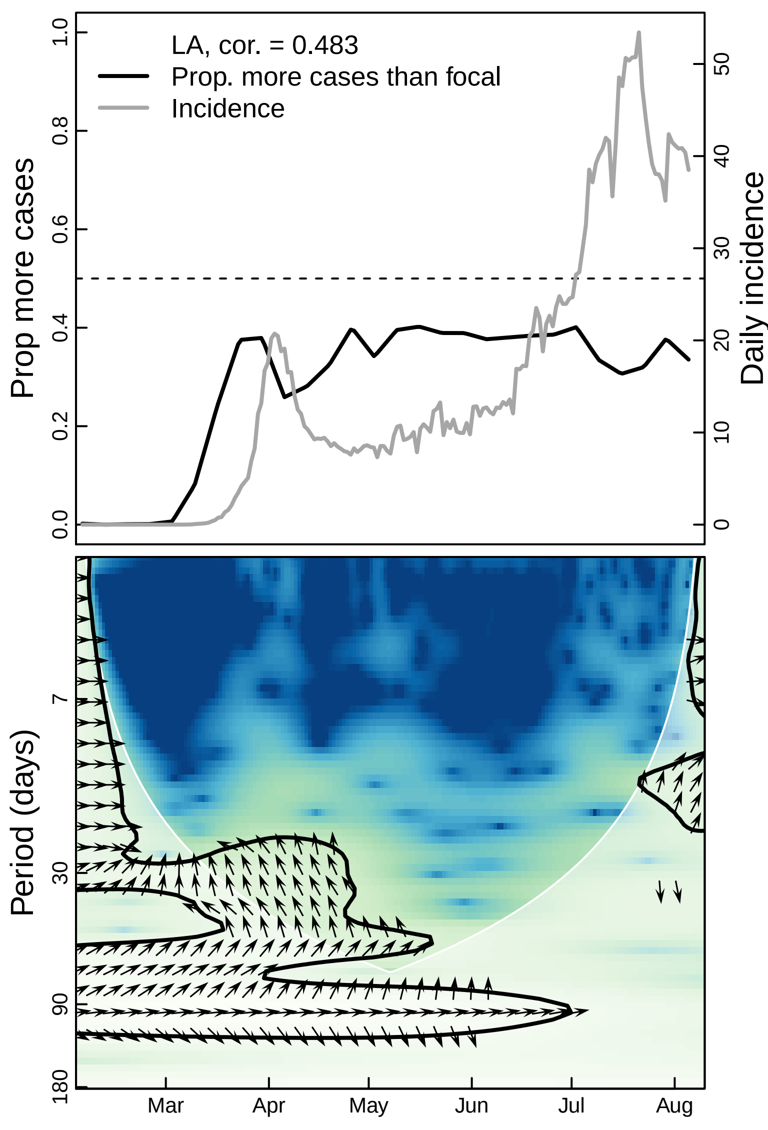

LA, cor. = 0.483
Prop. more cases than focal
Incidence
Prop more cases
Daily incidence
0.0
0.2
0.4
0.6
0.8
1.0
0
10
20
30
40
50
Period (days)
7
30
90
180
Mar
Apr
May
Jun
Jul
Aug

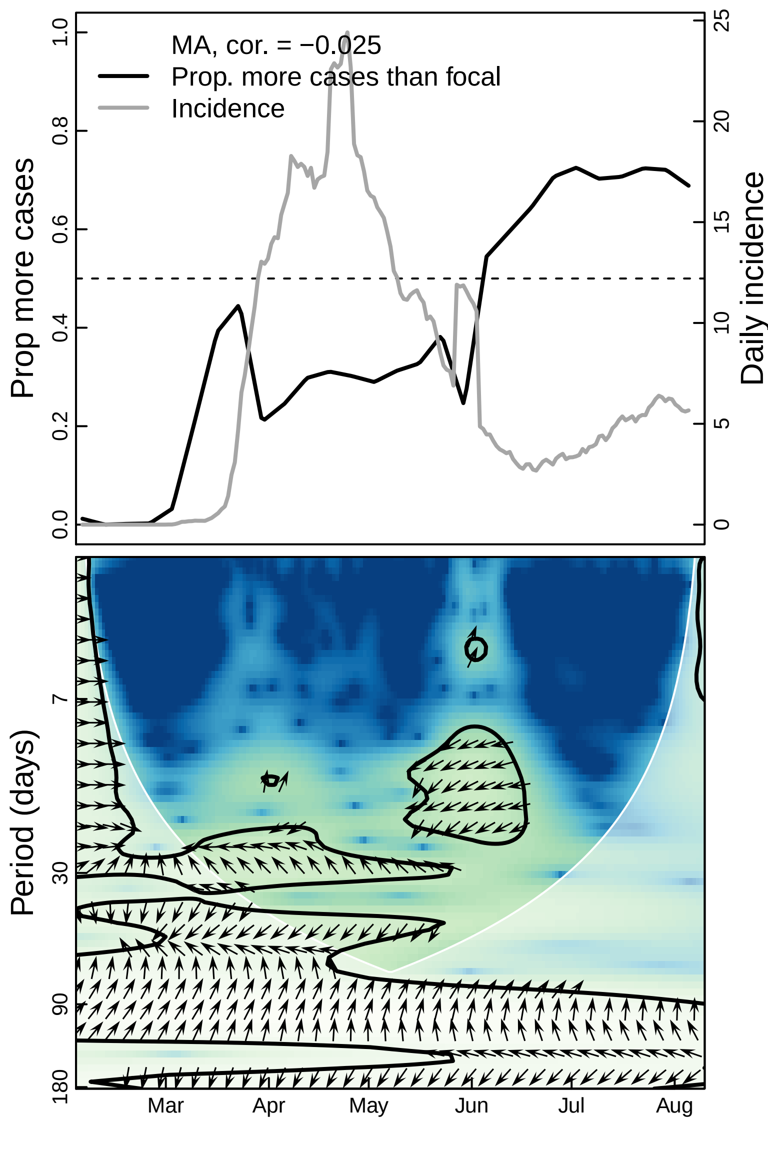

MA, cor. = −0.025
Prop. more cases than focal
Incidence
Prop more cases
Daily incidence
0.0
0.2
0.4
0.6
0.8
1.0
0
5
10
15
20
25
Period (days)
7
30
90
180
Mar
Apr
May
Jun
Jul
Aug

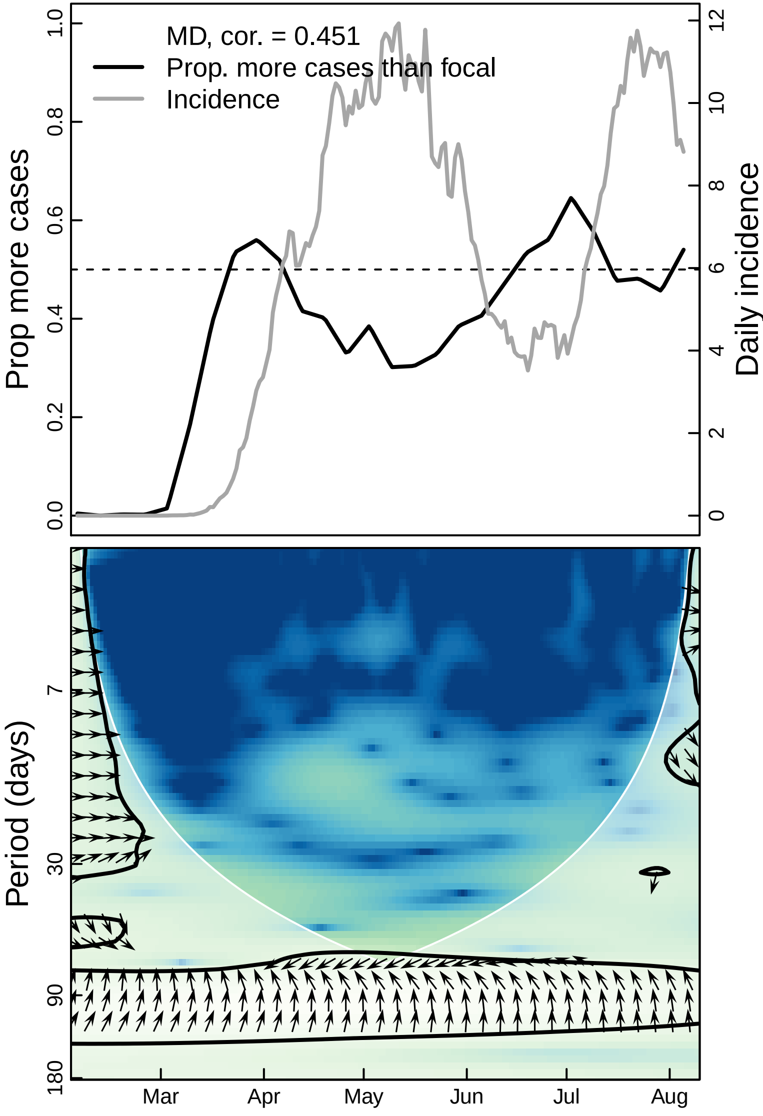

MD, cor. = 0.451
Prop. more cases than focal
Incidence
Prop more cases
Daily incidence
0.0
0.2
0.4
0.6
0.8
1.0
0
2
4
6
8
10
12
Period (days)
7
30
90
180
Mar
Apr
May
Jun
Jul
Aug

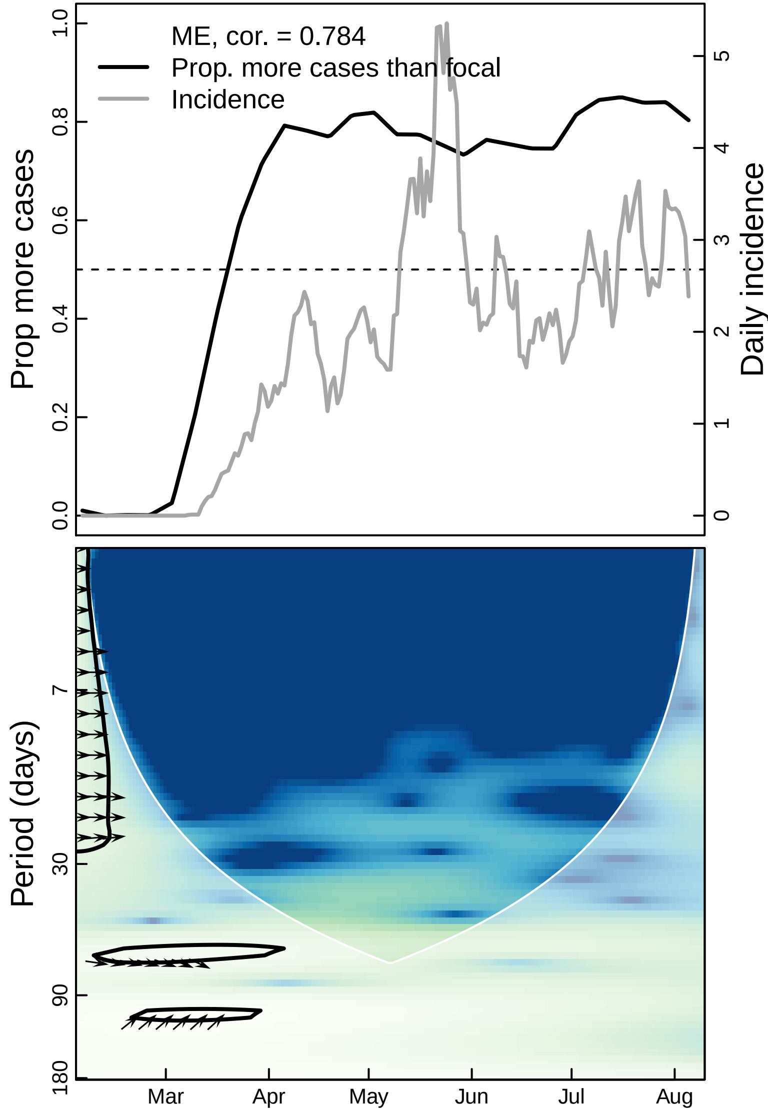

ME, cor. = 0.784
Prop. more cases than focal
Incidence
Prop more cases
Daily incidence
0.0
0.2
0.4
0.6
0.8
1.0
0
1
2
3
4
5
Period (days)
7
30
90
180
Mar
Apr
May
Jun
Jul
Aug

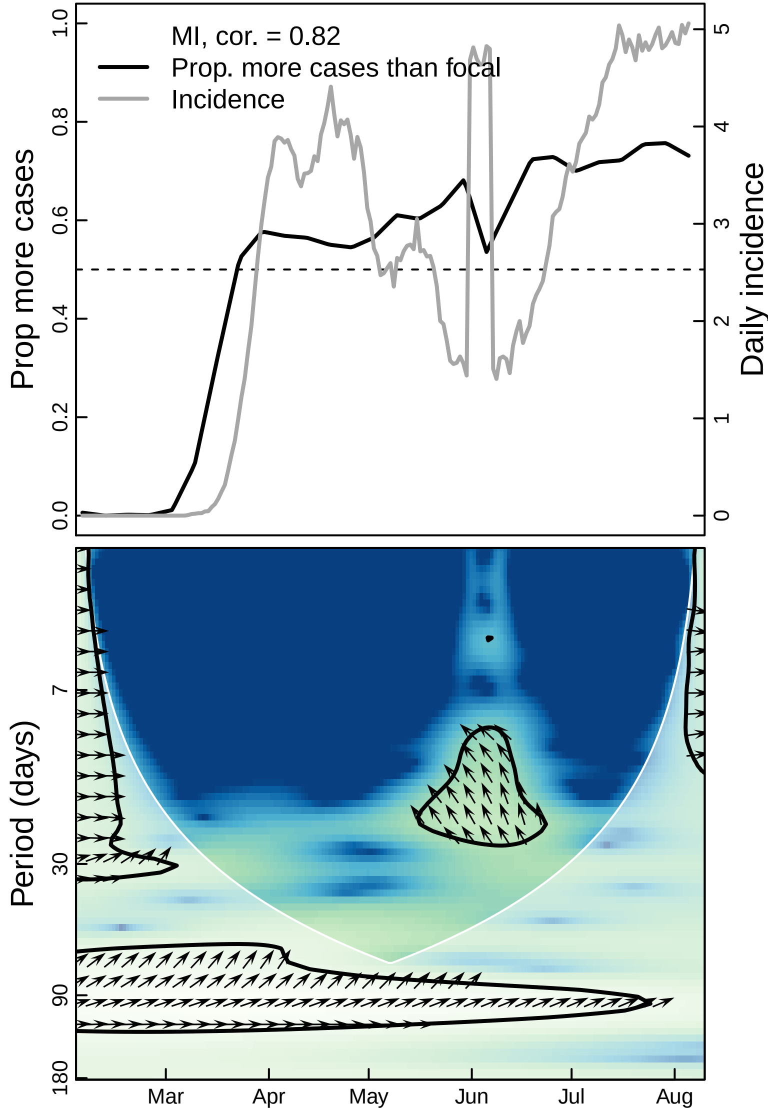

MI, cor. = 0.82
Prop. more cases than focal
Incidence
Prop more cases
Daily incidence
0.0
0.2
0.4
0.6
0.8
1.0
0
1
2
3
4
5
Period (days)
7
30
90
180
Mar
Apr
May
Jun
Jul
Aug

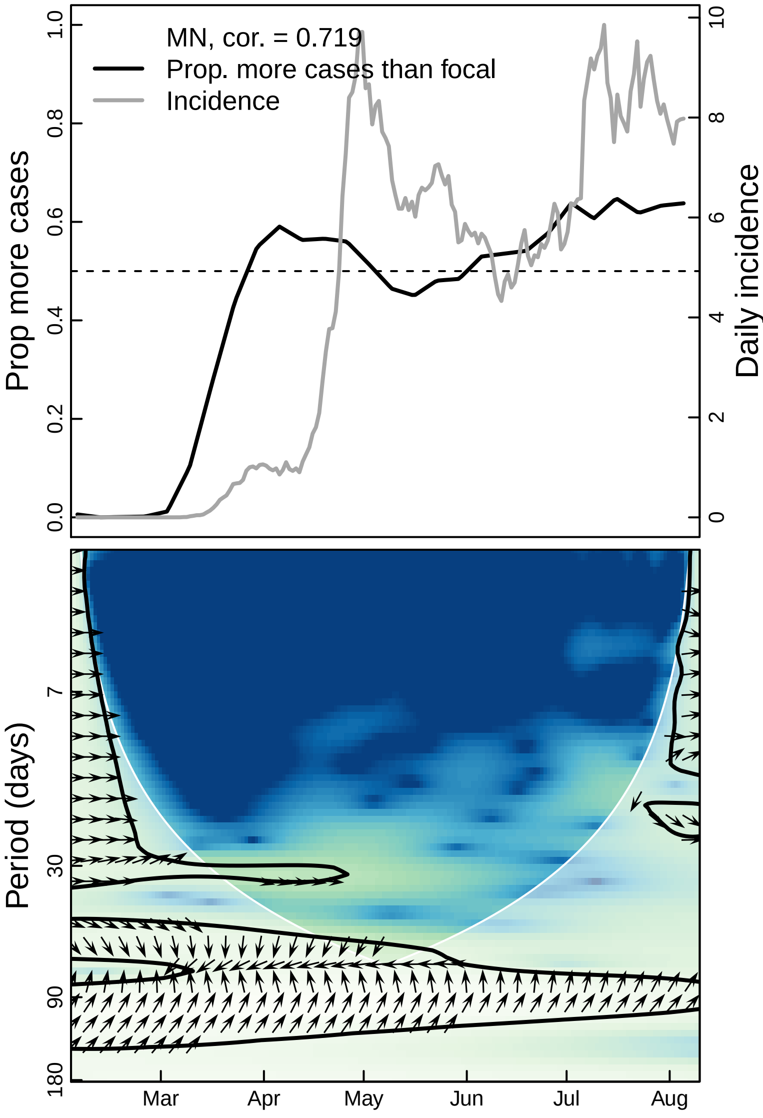

MN, cor. = 0.719
Prop. more cases than focal
Incidence
Prop more cases
Daily incidence
0.0
0.2
0.4
0.6
0.8
1.0
0
2
4
6
8
10
Period (days)
7
30
90
180
Mar
Apr
May
Jun
Jul
Aug

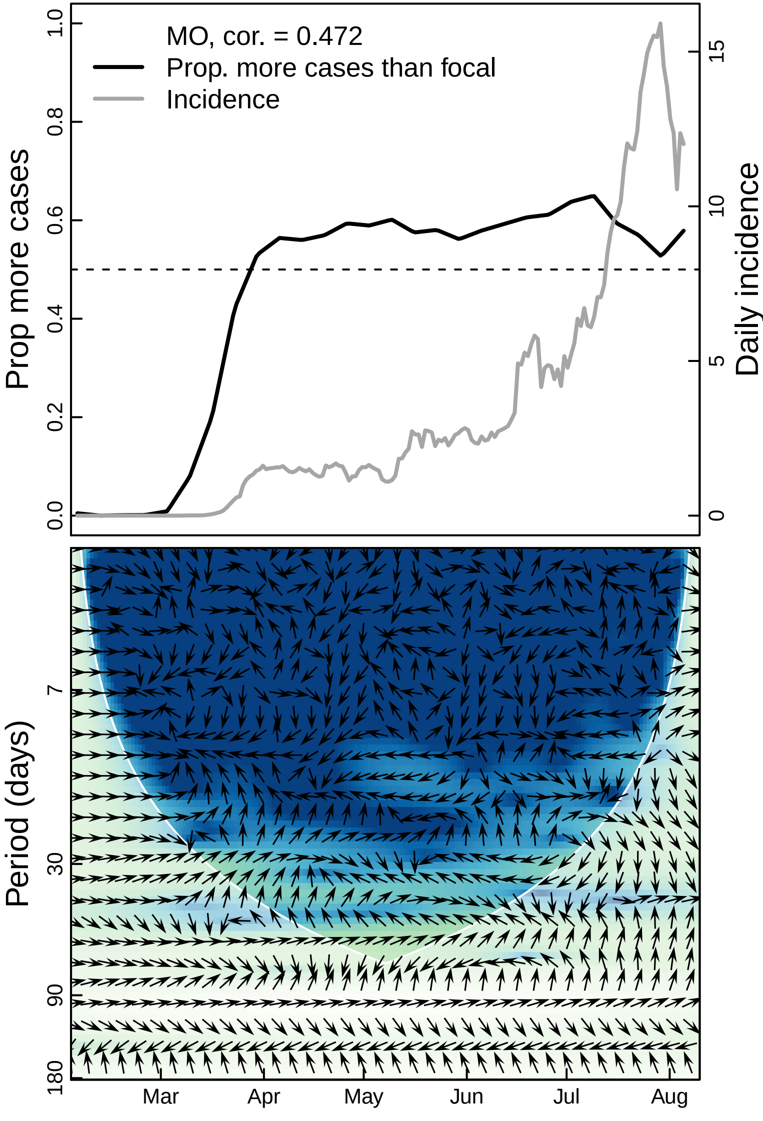

MO, cor. = 0.472
Prop. more cases than focal
Incidence
Prop more cases
Daily incidence
0.0
0.2
0.4
0.6
0.8
1.0
0
5
10
15
Period (days)
7
30
90
180
Mar
Apr
May
Jun
Jul
Aug

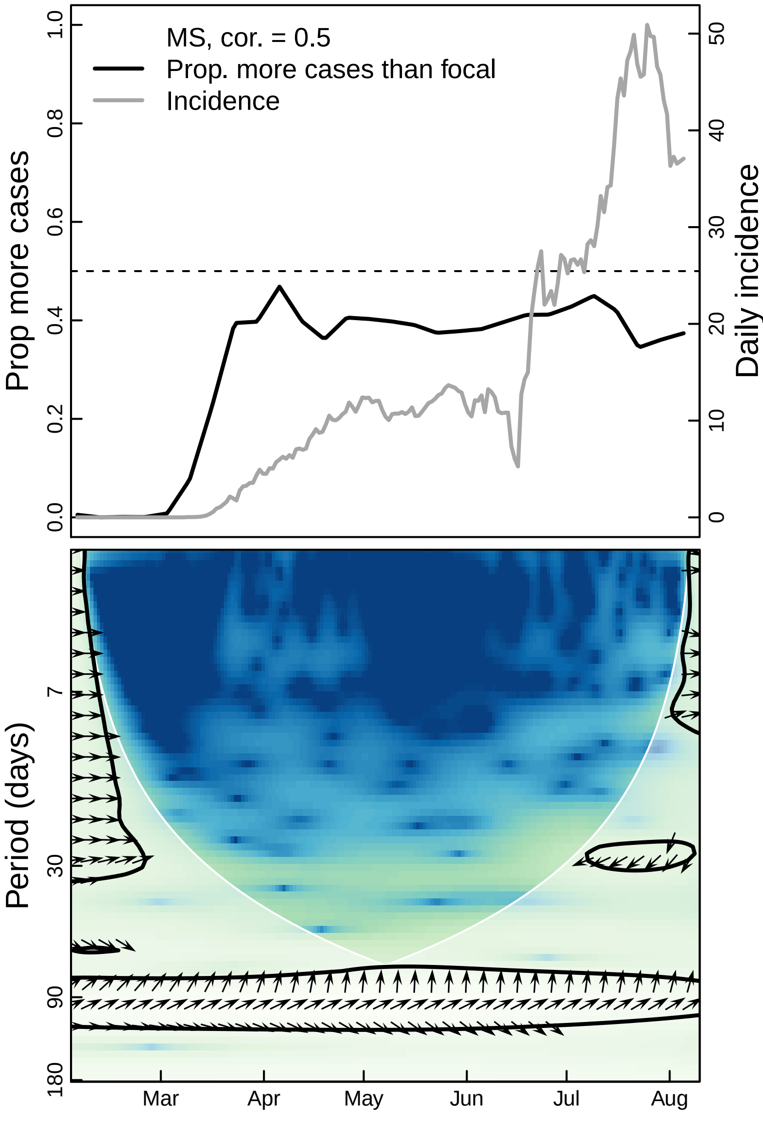

MS, cor. = 0.5
Prop. more cases than focal
Incidence
Prop more cases
Daily incidence
0.0
0.2
0.4
0.6
0.8
1.0
0
10
20
30
40
50
Period (days)
7
30
90
180
Mar
Apr
May
Jun
Jul
Aug

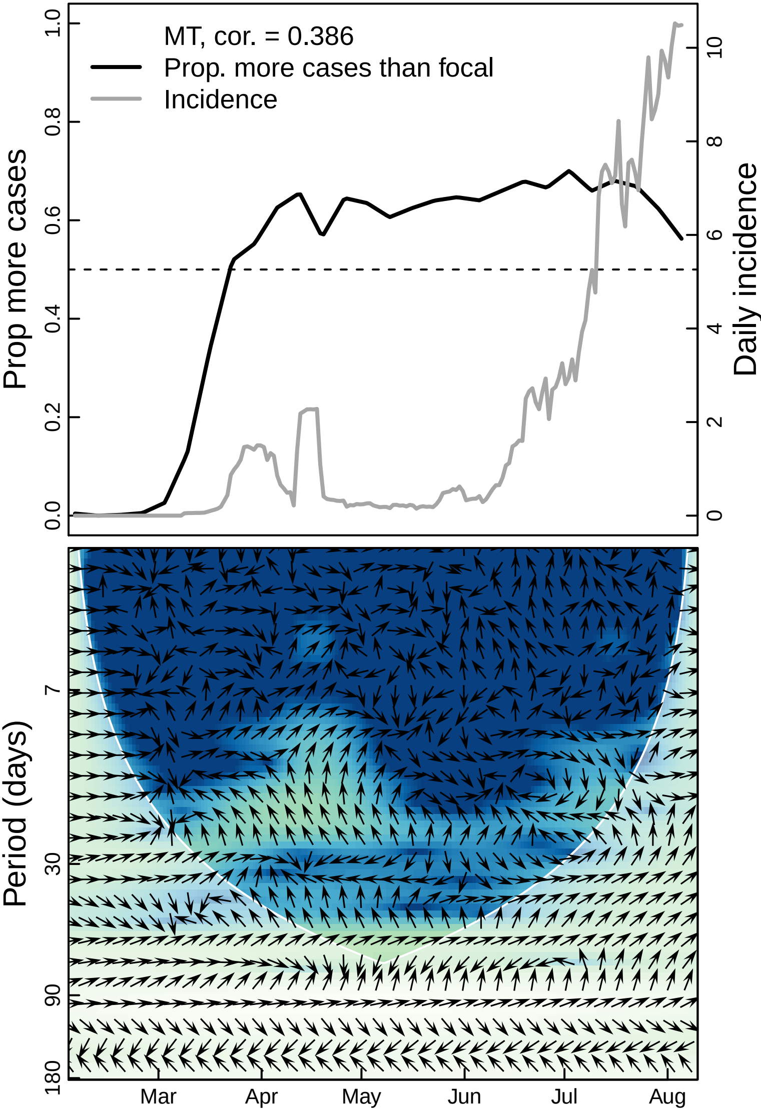

MT, cor. = 0.386
Prop. more cases than focal
Incidence
Prop more cases
Daily incidence
0.0
0.2
0.4
0.6
0.8
1.0
0
2
4
6
8
10
Period (days)
7
30
90
180
Mar
Apr
May
Jun
Jul
Aug

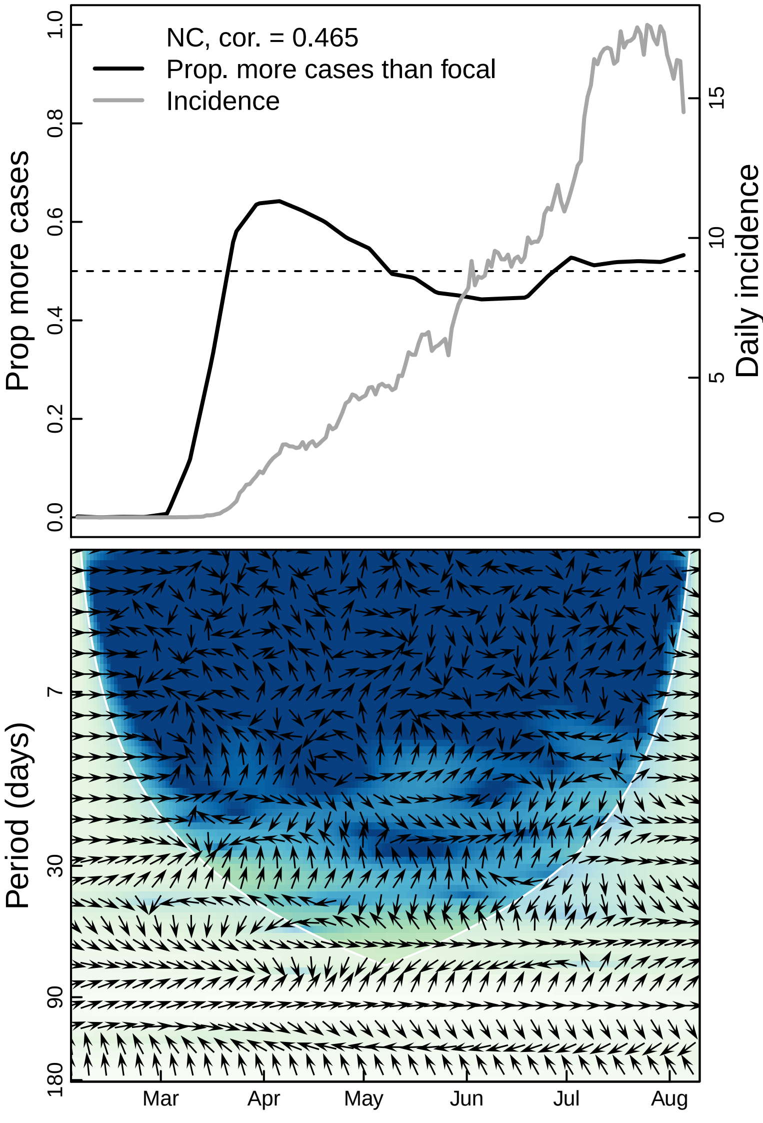

NC, cor. = 0.465
Prop. more cases than focal
Incidence
Prop more cases
Daily incidence
0.0
0.2
0.4
0.6
0.8
1.0
0
5
10
15
Period (days)
7
30
90
180
Mar
Apr
May
Jun
Jul
Aug

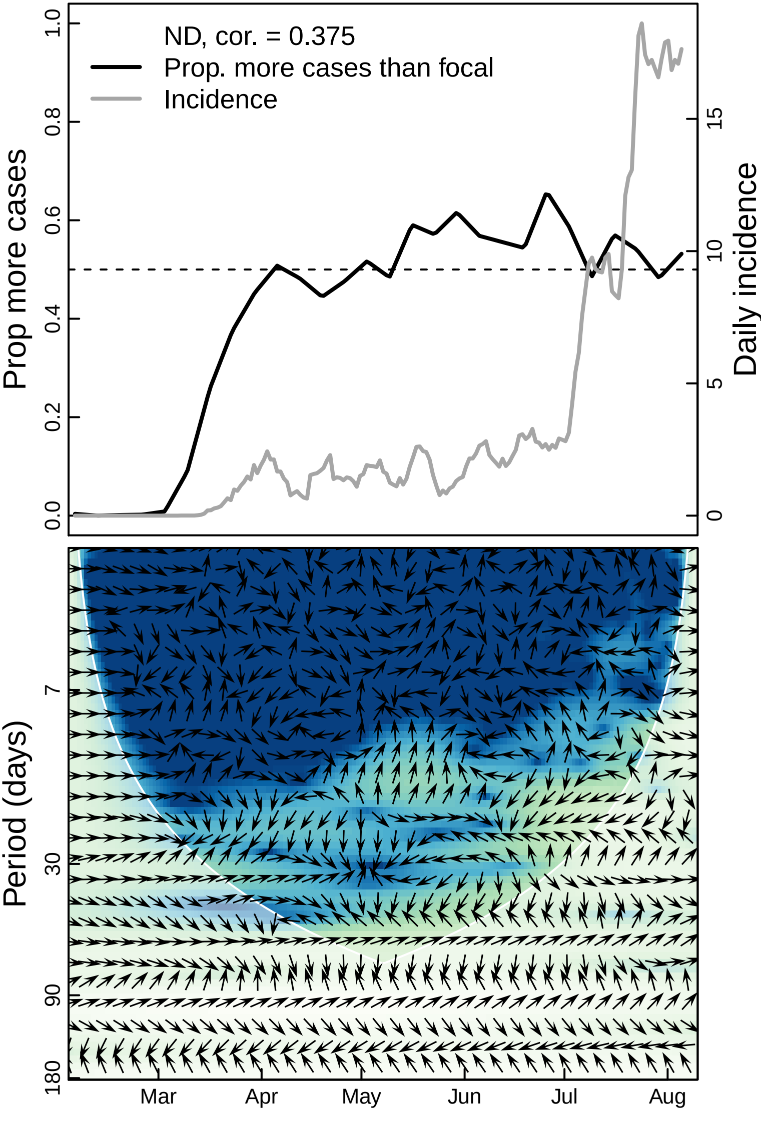

ND, cor. = 0.375
Prop. more cases than focal
Incidence
Prop more cases
Daily incidence
0.0
0.2
0.4
0.6
0.8
1.0
0
5
10
15
Period (days)
7
30
90
180
Mar
Apr
May
Jun
Jul
Aug

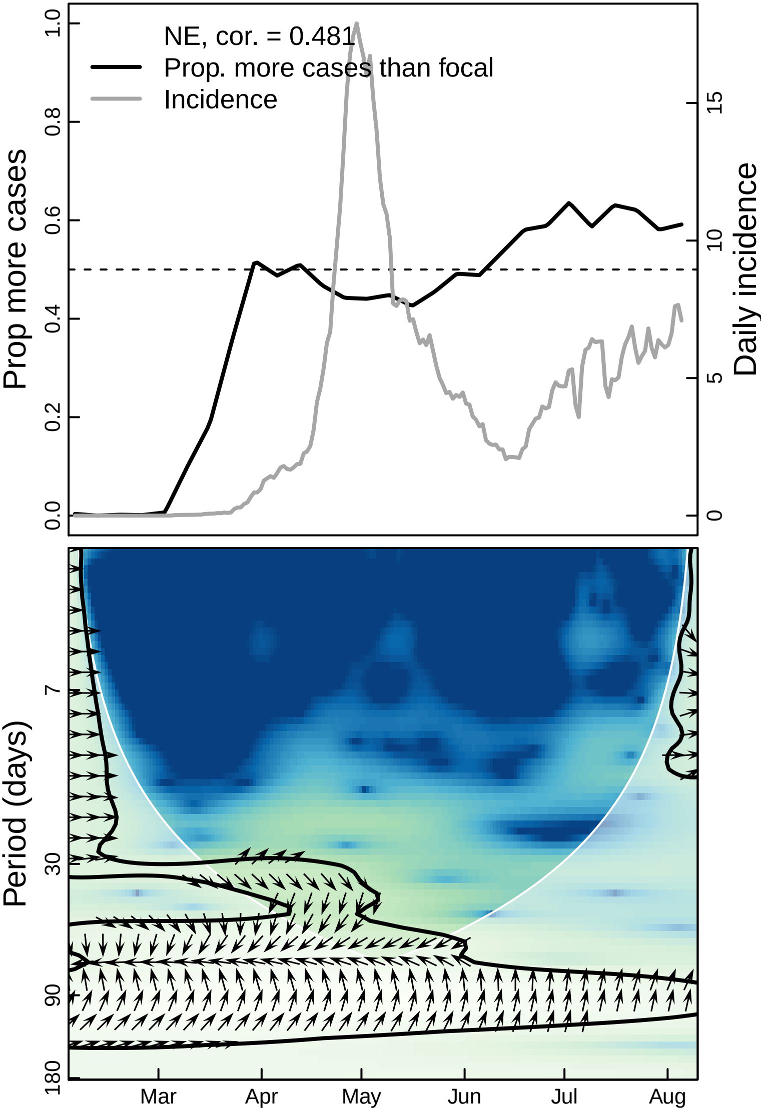

NE, cor. = 0.481
Prop. more cases than focal
Incidence
Prop more cases
Daily incidence
0.0
0.2
0.4
0.6
0.8
1.0
0
5
10
15
Period (days)
7
30
90
180
Mar
Apr
May
Jun
Jul
Aug

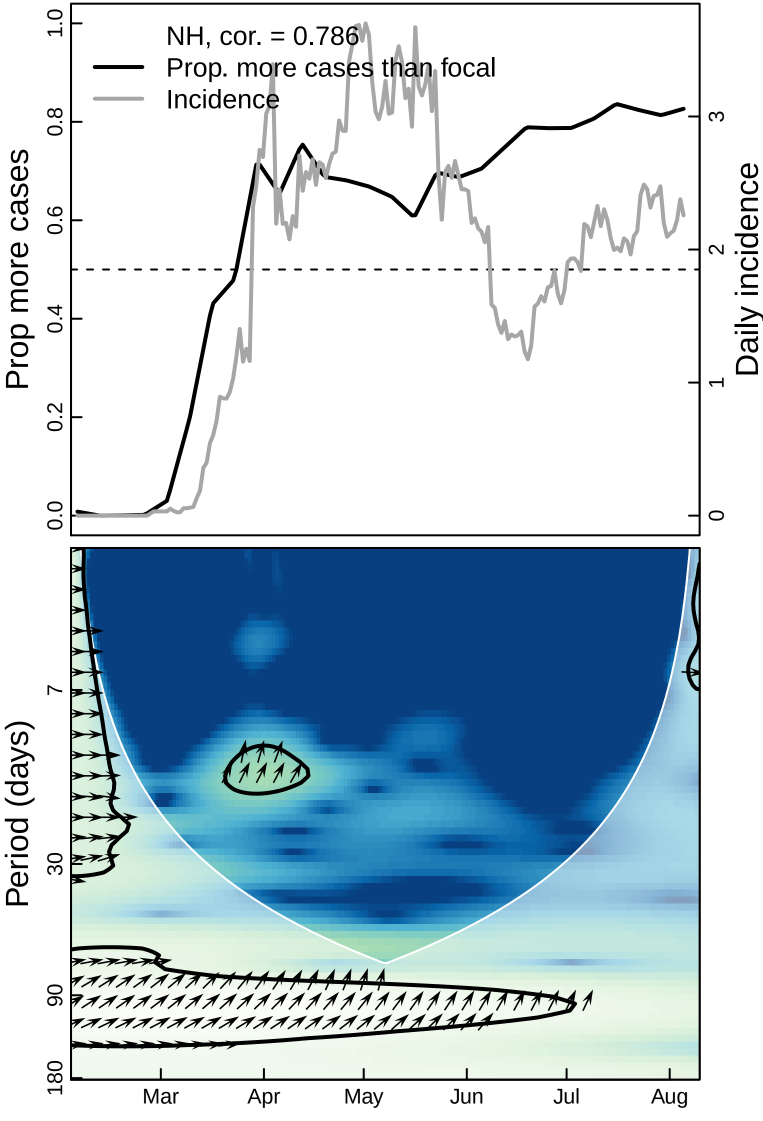

NH, cor. = 0.786
Prop. more cases than focal
Incidence
Prop more cases
Daily incidence
0.0
0.2
0.4
0.6
0.8
1.0
0
1
2
3
Period (days)
7
30
90
180
Mar
Apr
May
Jun
Jul
Aug

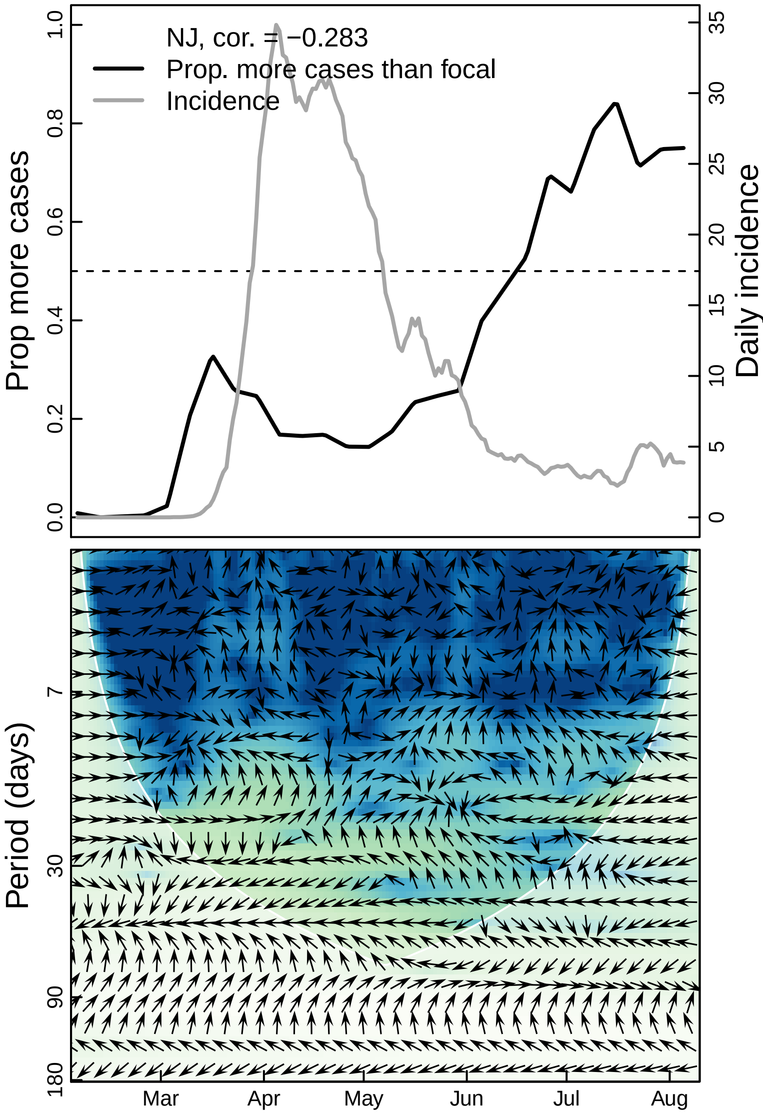

NJ, cor. = −0.283
Prop. more cases than focal
Incidence
Prop more cases
Daily incidence
0.0
0.2
0.4
0.6
0.8
1.0
0
5
10
15
20
25
30
35
Period (days)
7
30
90
180
Mar
Apr
May
Jun
Jul
Aug

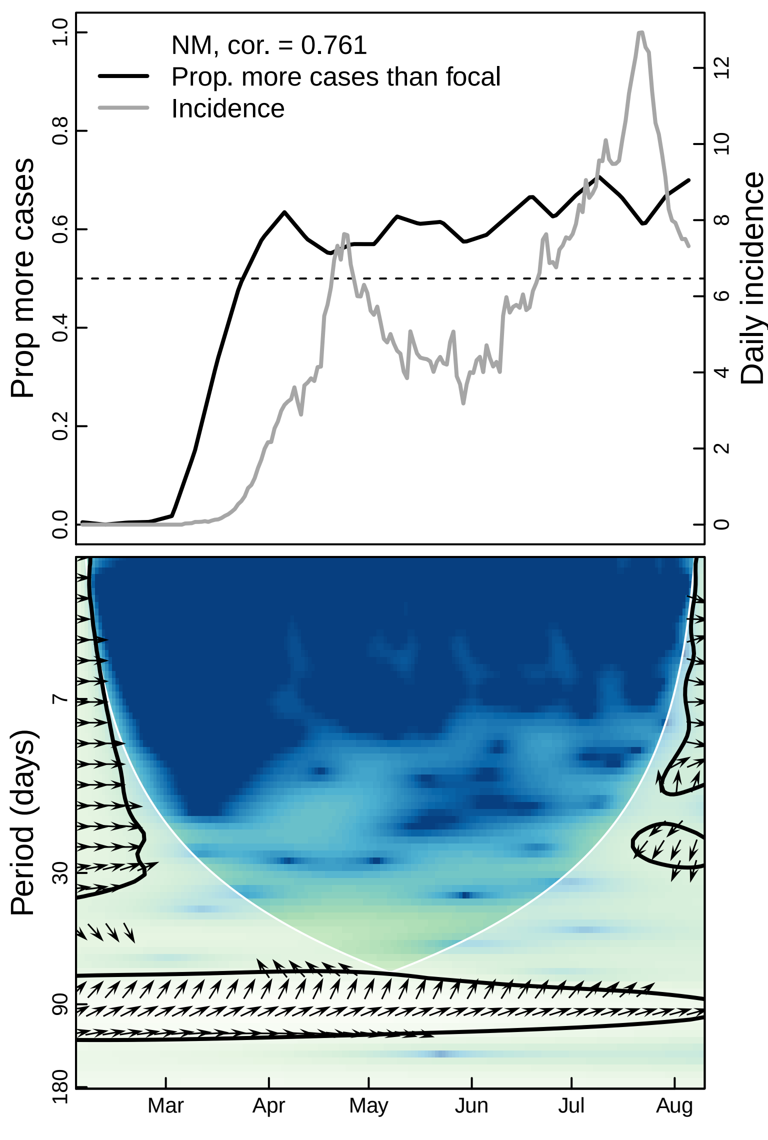

NM, cor. = 0.761
Prop. more cases than focal
Incidence
Prop more cases
Daily incidence
Period (days)
Mar
Apr
May
Jun
Jul
Aug

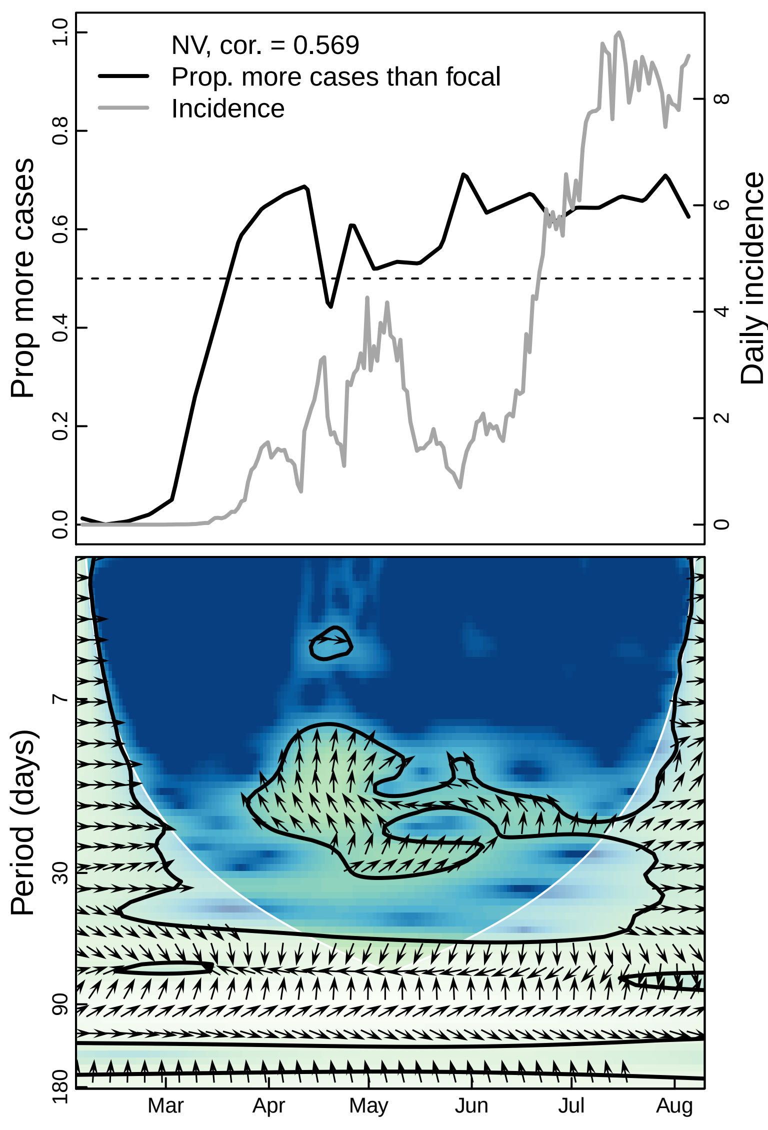
NV, cor. = 0.569
Prop. more cases than focal
Incidence
Prop more cases
Daily incidence
0.0
0.2
0.4
0.6
0.8
1.0
0
2
4
6
8
Period (days)
7
30
90
180
Mar
Apr
May
Jun
Jul
Aug

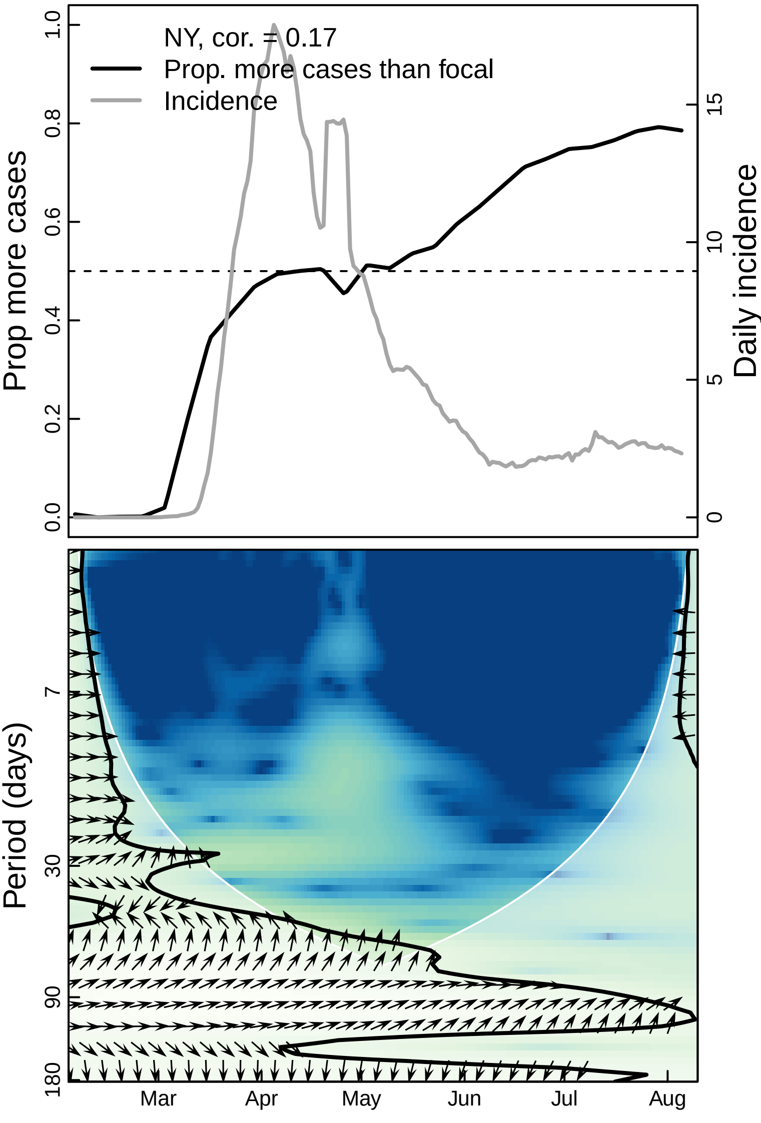

NY, cor. = 0.17
Prop. more cases than focal
Incidence
Prop more cases
Daily incidence
0.0
0.2
0.4
0.6
0.8
1.0
0
5
10
15
Period (days)
7
30
90
180
Mar
Apr
May
Jun
Jul
Aug

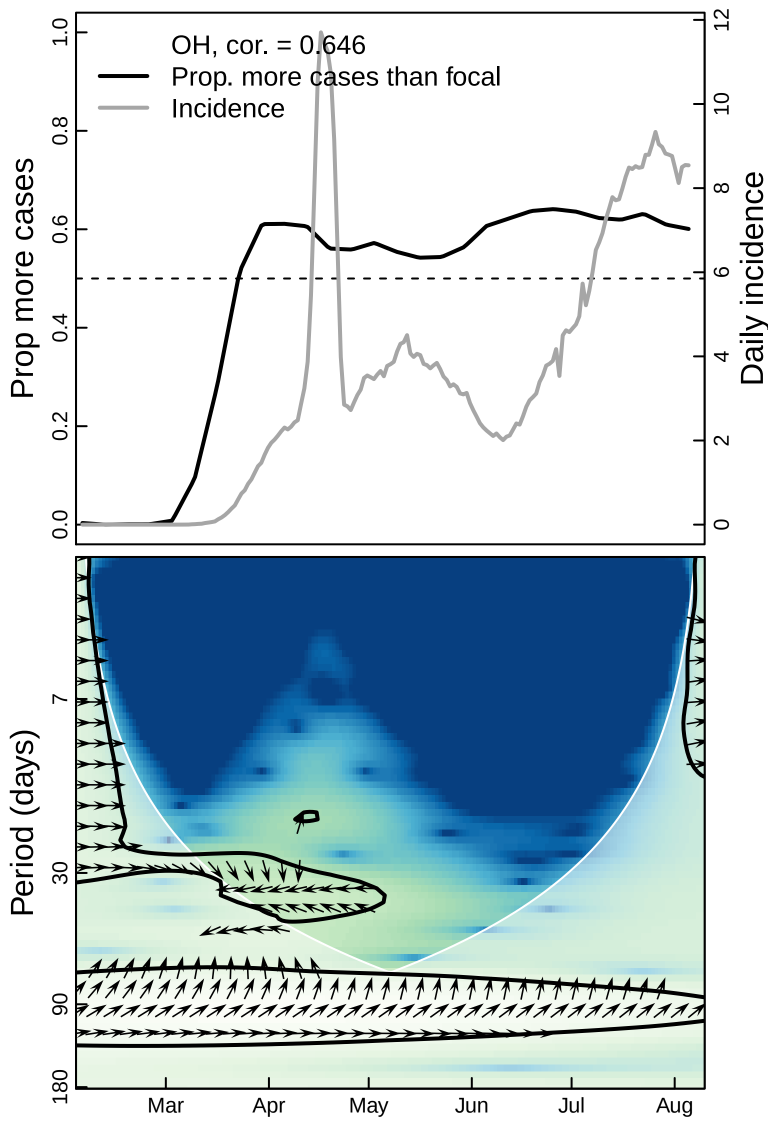

OH, cor. = 0.646
Prop. more cases than focal
Incidence
Prop more cases
Daily incidence
0.0
0.2
0.4
0.6
0.8
1.0
0
2
4
6
8
10
12
Period (days)
7
30
90
180
Mar
Apr
May
Jun
Jul
Aug

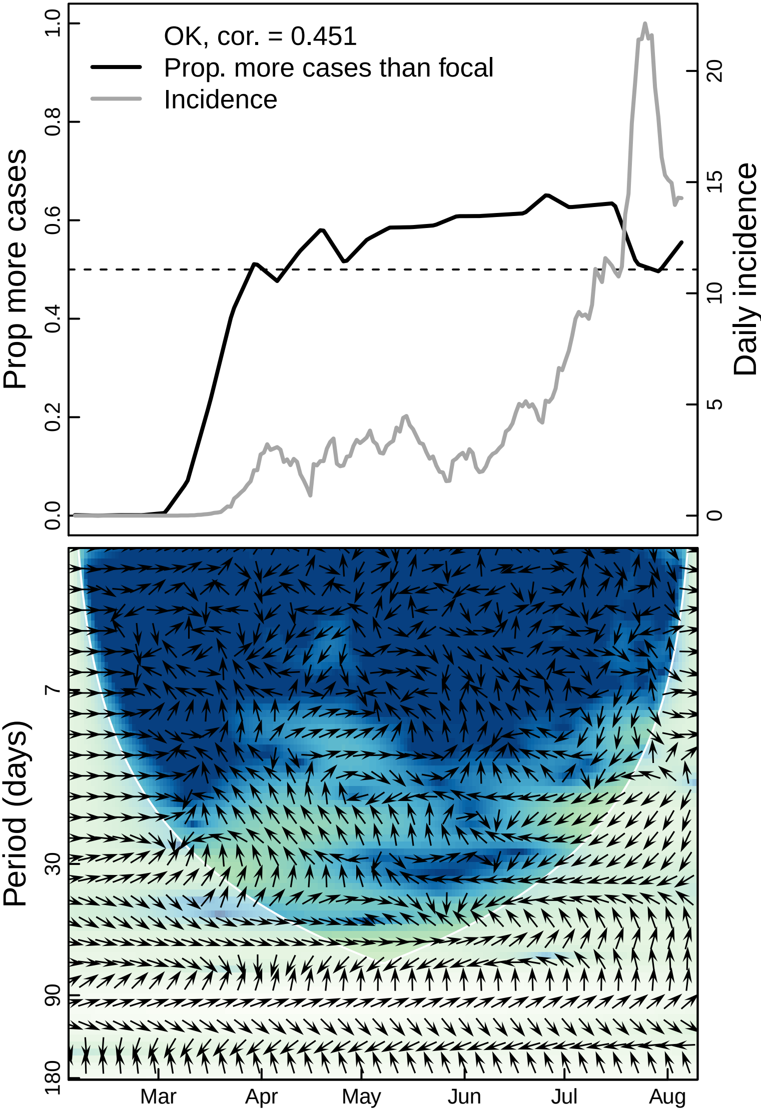

OK, cor. = 0.451
Prop. more cases than focal
Incidence
Prop more cases
Daily incidence
0.0
0.2
0.4
0.6
0.8
1.0
0
5
10
15
20
Period (days)
7
30
90
180
Mar
Apr
May
Jun
Jul
Aug

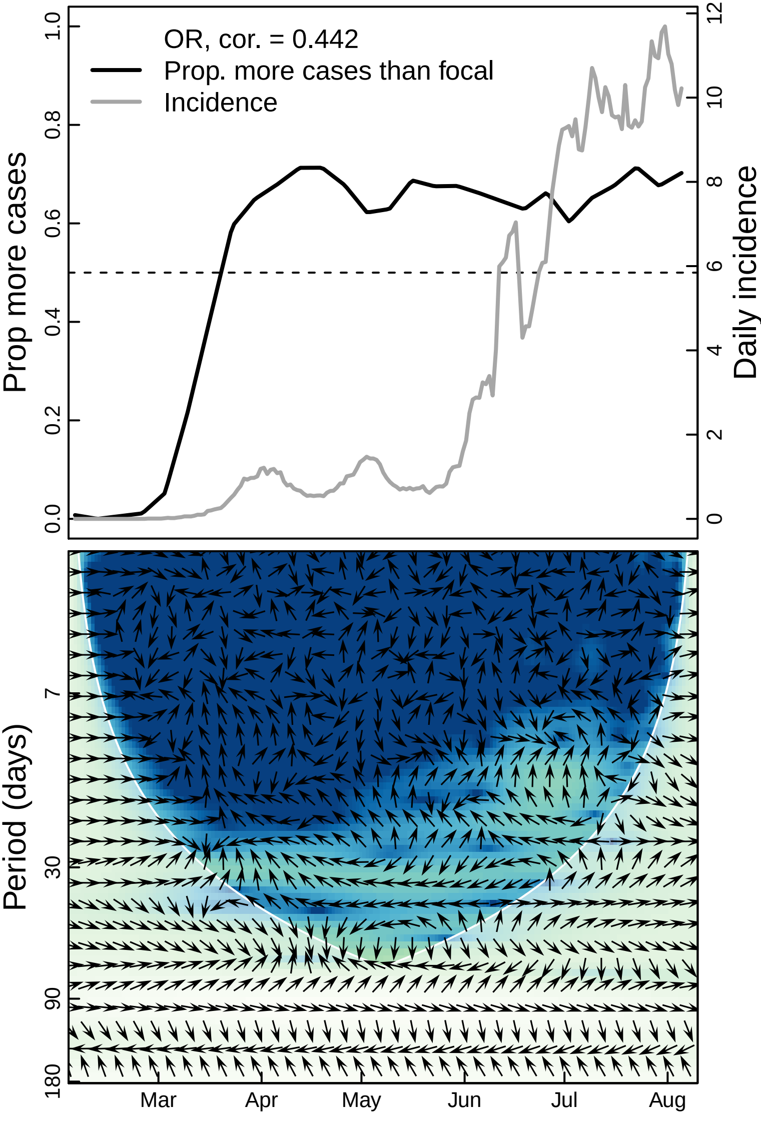

OR, cor. = 0.442
Prop. more cases than focal
Incidence
Prop more cases
Daily incidence
0.0
0.2
0.4
0.6
0.8
1.0
0
2
4
6
8
10
12
Period (days)
7
30
90
180
Mar
Apr
May
Jun
Jul
Aug

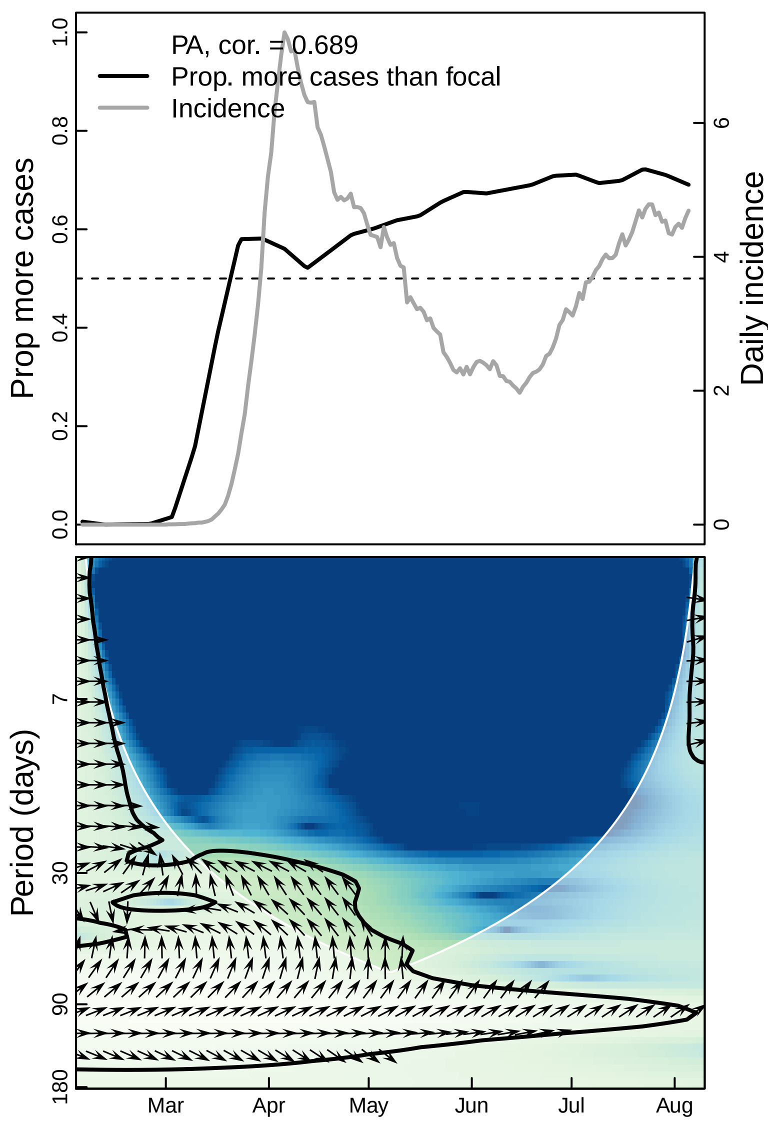

PA, cor. = 0.689
Prop. more cases than focal
Incidence
Prop more cases
Daily incidence
0.0
0.2
0.4
0.6
0.8
1.0
0
2
4
6
Period (days)
7
30
90
180
Mar
Apr
May
Jun
Jul
Aug

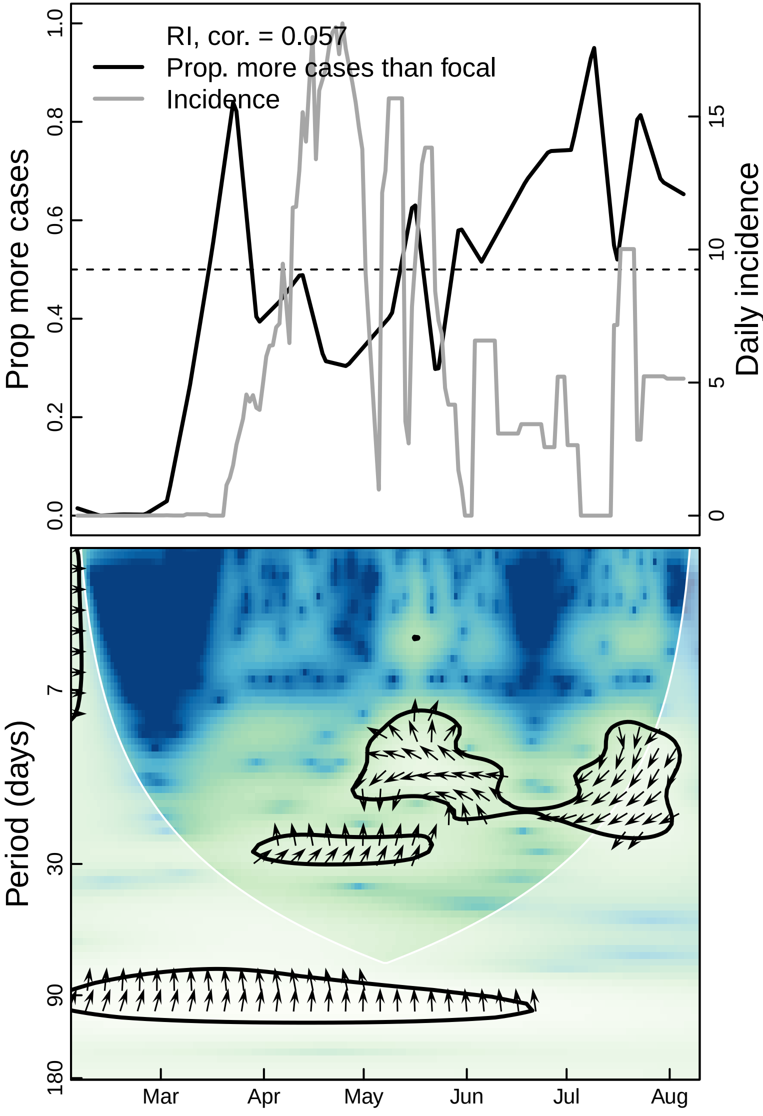

RI, cor. = 0.057
Prop. more cases than focal
Incidence
Prop more cases
0.0
0.2
0.4
0.6
0.8
1.0
Daily incidence
0
5
10
15
Period (days)
7
30
90
180
Mar
Apr
May
Jun
Jul
Aug

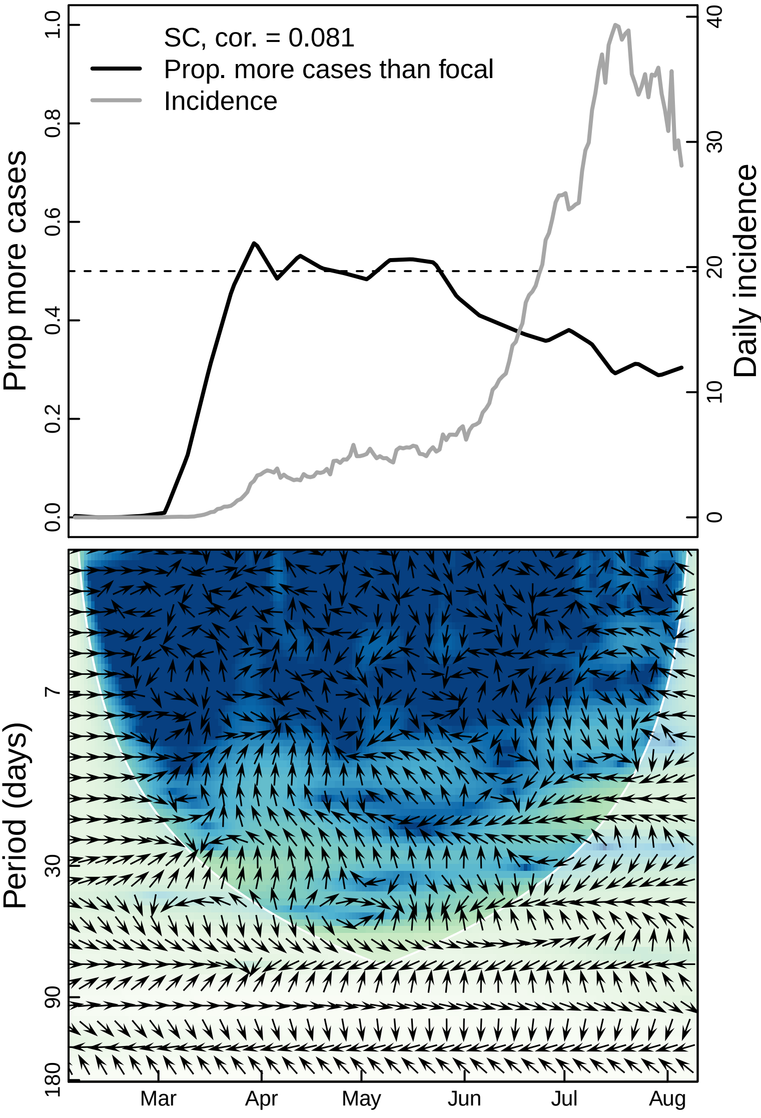
SC, cor. = 0.081
Prop. more cases than focal
Incidence
Prop more cases
Daily incidence
0.0
0.2
0.4
0.6
0.8
1.0
0
10
20
30
40
Period (days)
7
30
90
180
Mar
Apr
May
Jun
Jul
Aug

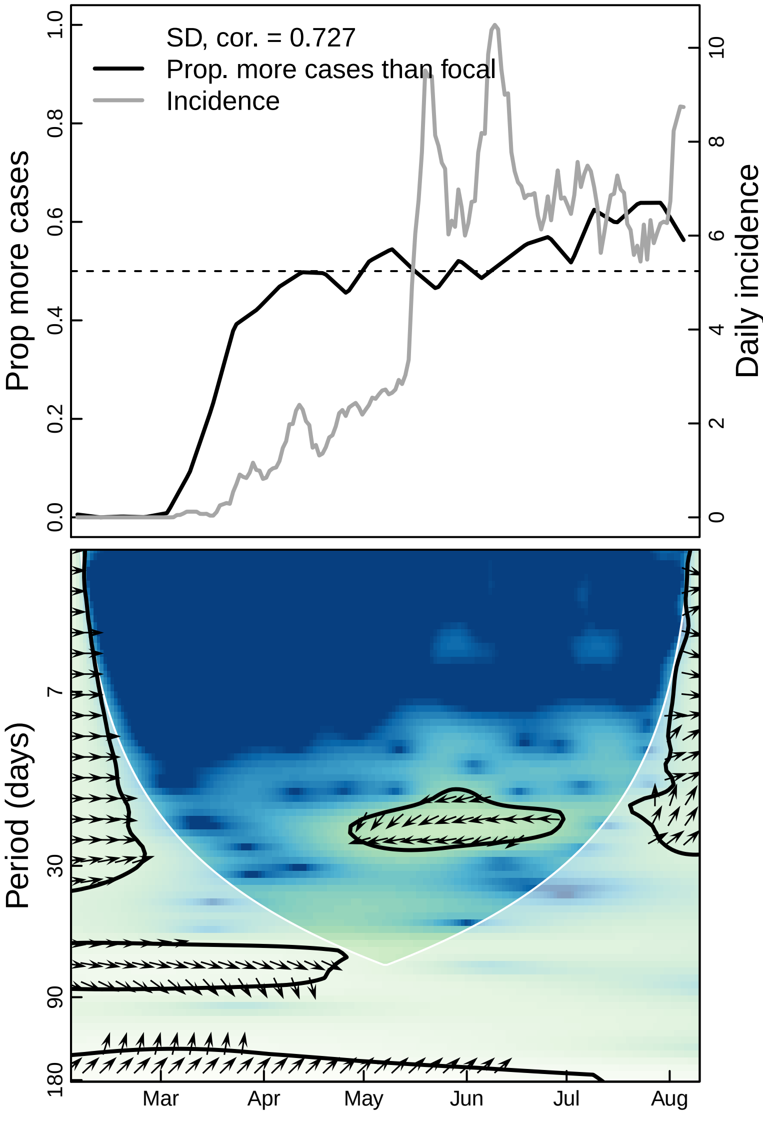

SD, cor. = 0.727
Prop. more cases than focal
Incidence
Prop more cases
Daily incidence
0.0
0.2
0.4
0.6
0.8
1.0
0
2
4
6
8
10
Period (days)
7
30
90
180
Mar
Apr
May
Jun
Jul
Aug

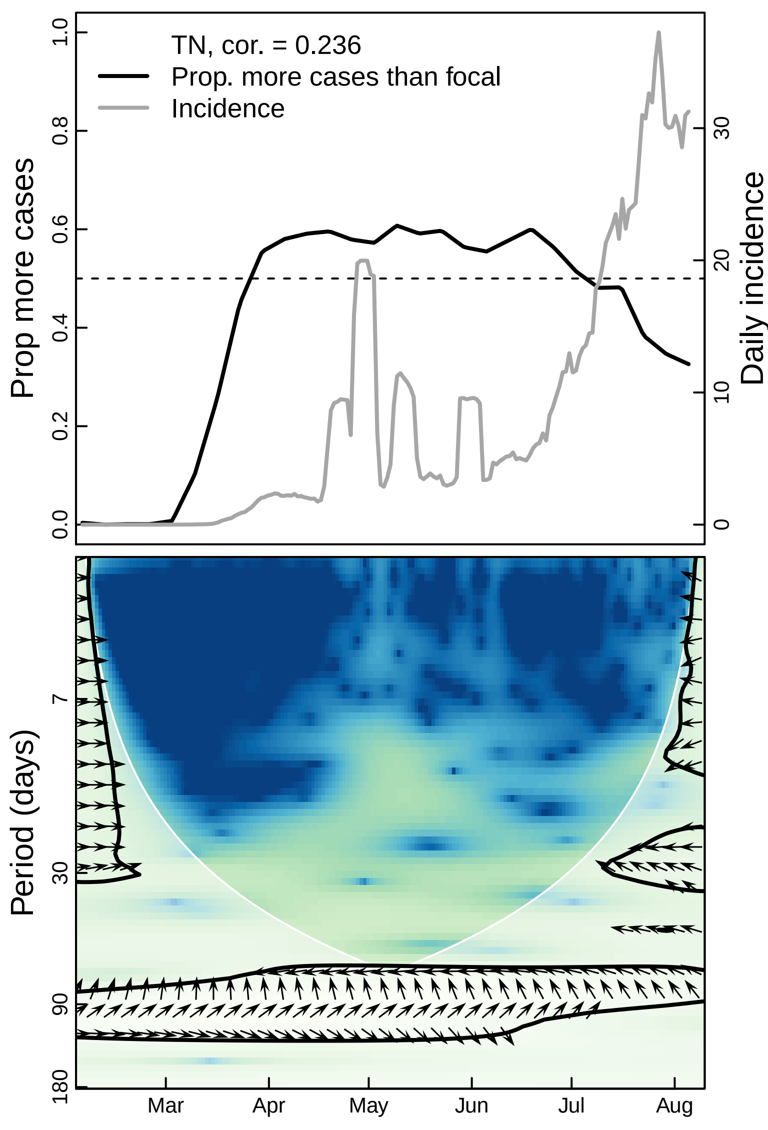

TN, cor. = 0.236
Prop. more cases than focal
Incidence
Prop more cases
Daily incidence
0.0
0.2
0.4
0.6
0.8
1.0
0
10
20
30
Period (days)
7
30
90
180
Mar
Apr
May
Jun
Jul
Aug

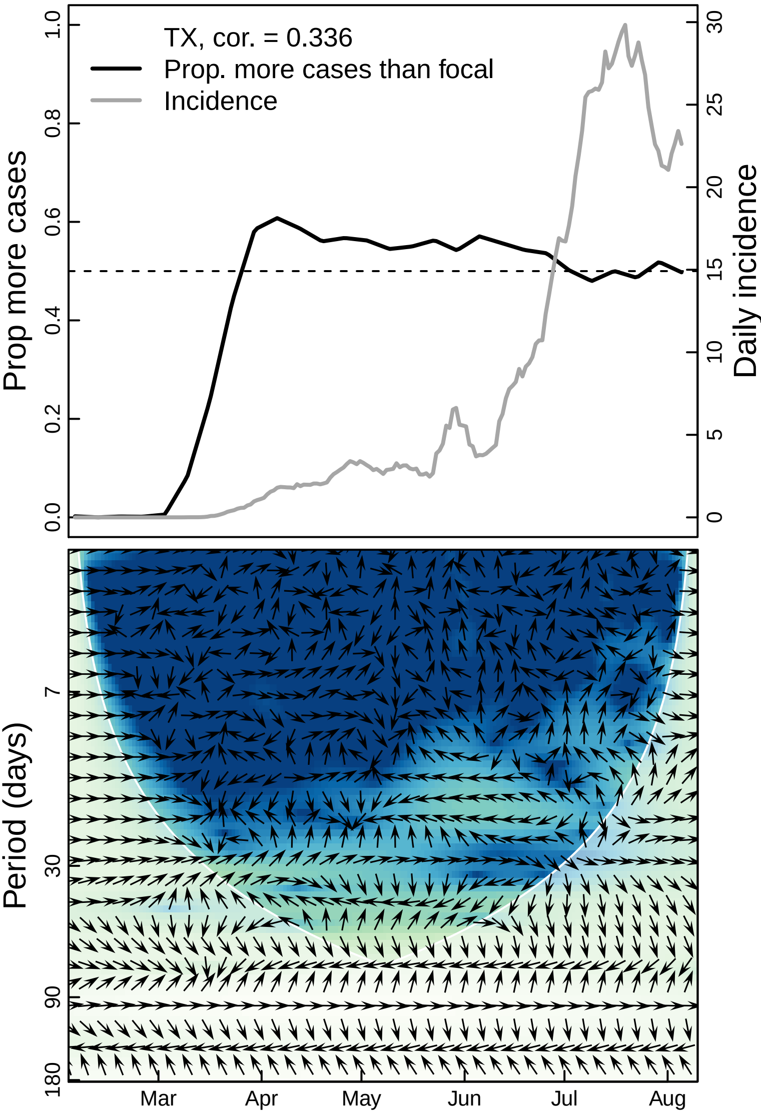

TX, cor. = 0.336
Prop. more cases than focal
Incidence
Prop more cases
Daily incidence
0.0
0.2
0.4
0.6
0.8
1.0
0
5
10
15
20
25
30
Period (days)
7
30
90
180
Mar
Apr
May
Jun
Jul
Aug

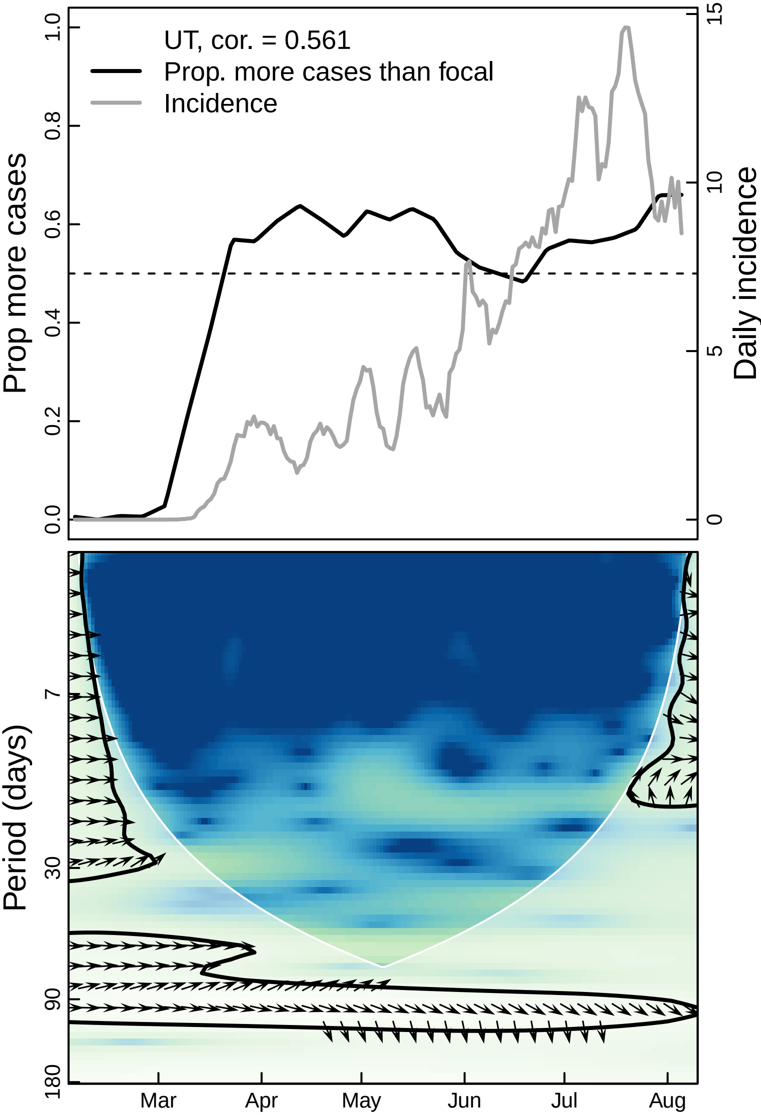

UT, cor. = 0.561
Prop. more cases than focal
Incidence
Prop more cases
Daily incidence
0.0
0.2
0.4
0.6
0.8
1.0
0
5
10
15
Period (days)
7
30
90
180
Mar
Apr
May
Jun
Jul
Aug

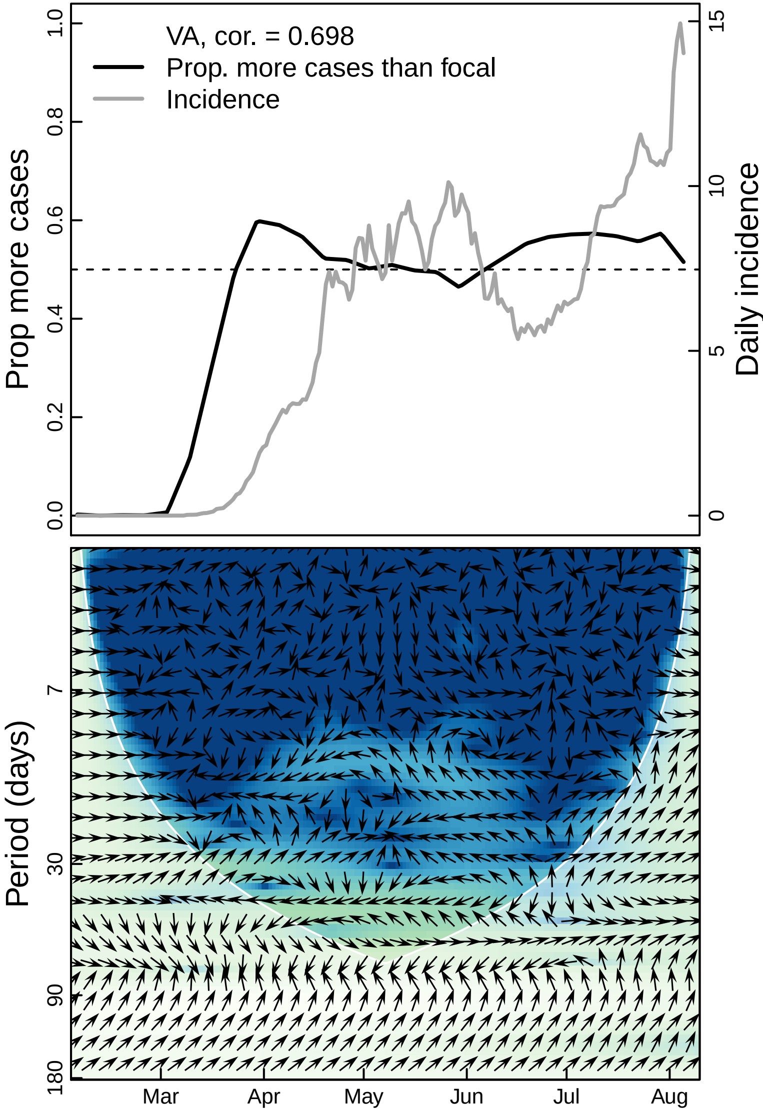
VA, cor. = 0.698
Prop. more cases than focal
Incidence
Prop more cases
Daily incidence
0.0
0.2
0.4
0.6
0.8
1.0
0
5
10
15
Period (days)
7
30
90
180
Mar
Apr
May
Jun
Jul
Aug

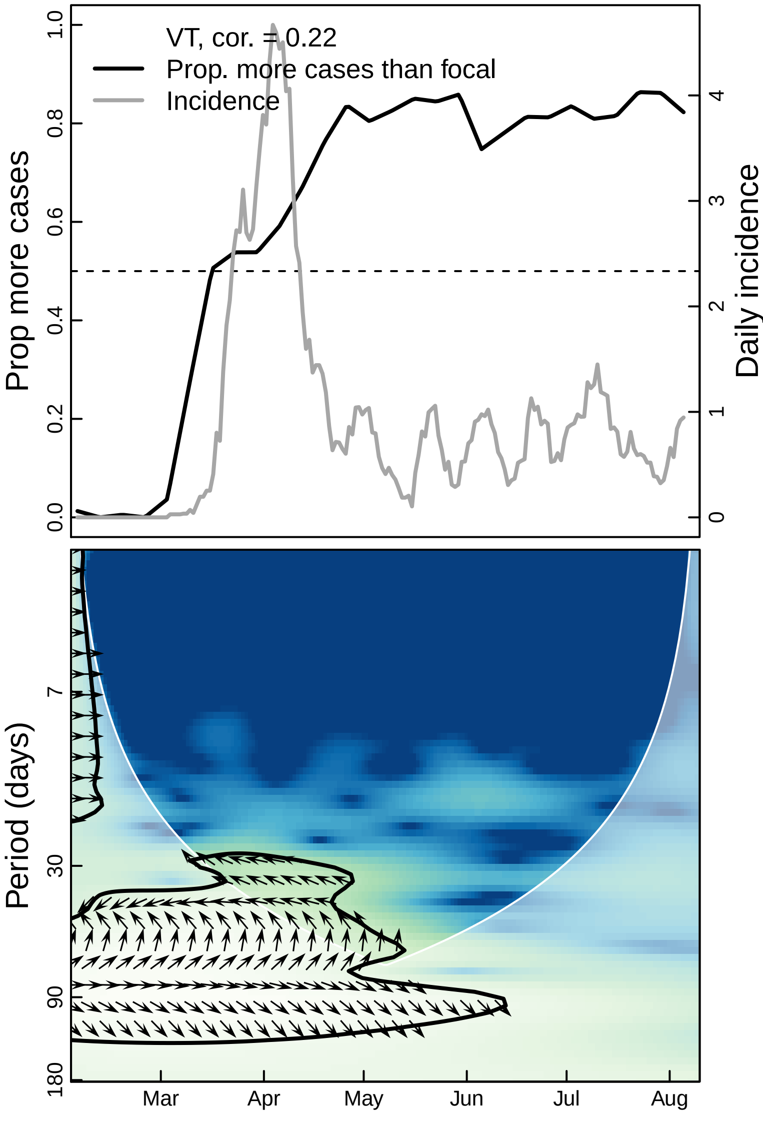

VT, cor. = 0.22
Prop. more cases than focal
Incidence
Prop more cases
Daily incidence
0.0
0.2
0.4
0.6
0.8
1.0
0
1
2
3
4
Period (days)
7
30
90
180
Mar
Apr
May
Jun
Jul
Aug

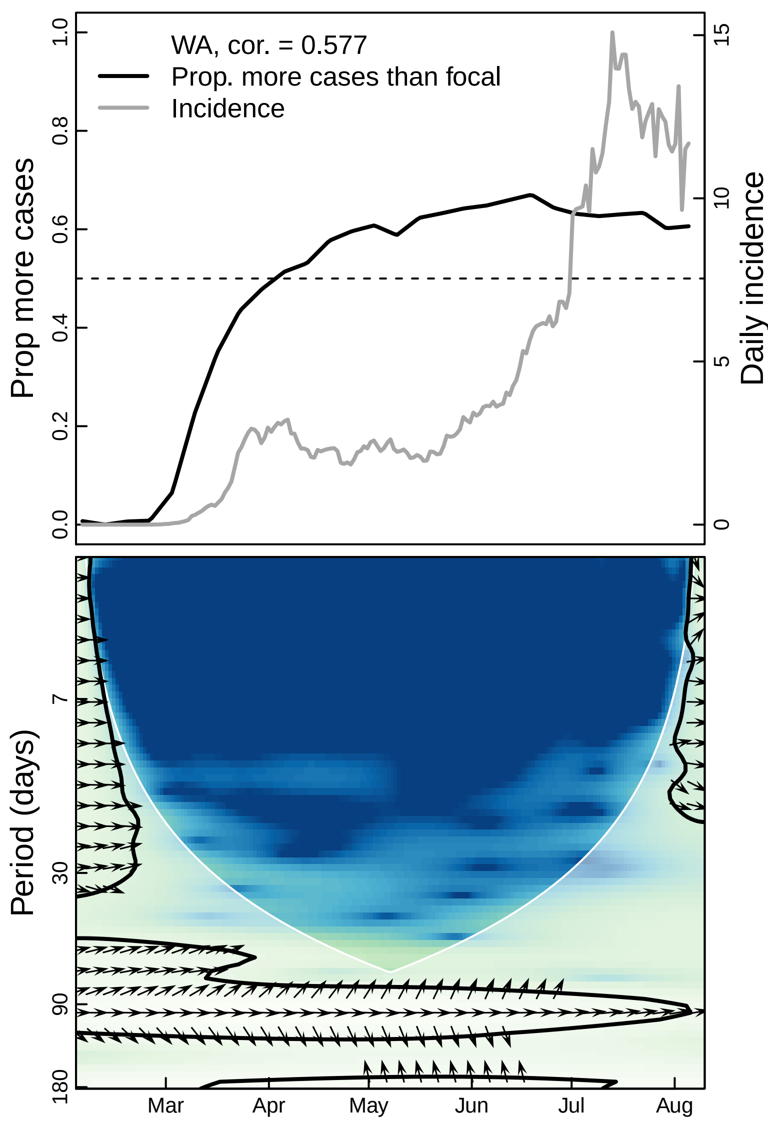

WA, cor. = 0.577
Prop. more cases than focal
Incidence
Prop more cases
Daily incidence
0.0
0.2
0.4
0.6
0.8
1.0
0
5
10
15
Period (days)
7
30
90
180
Mar
Apr
May
Jun
Jul
Aug

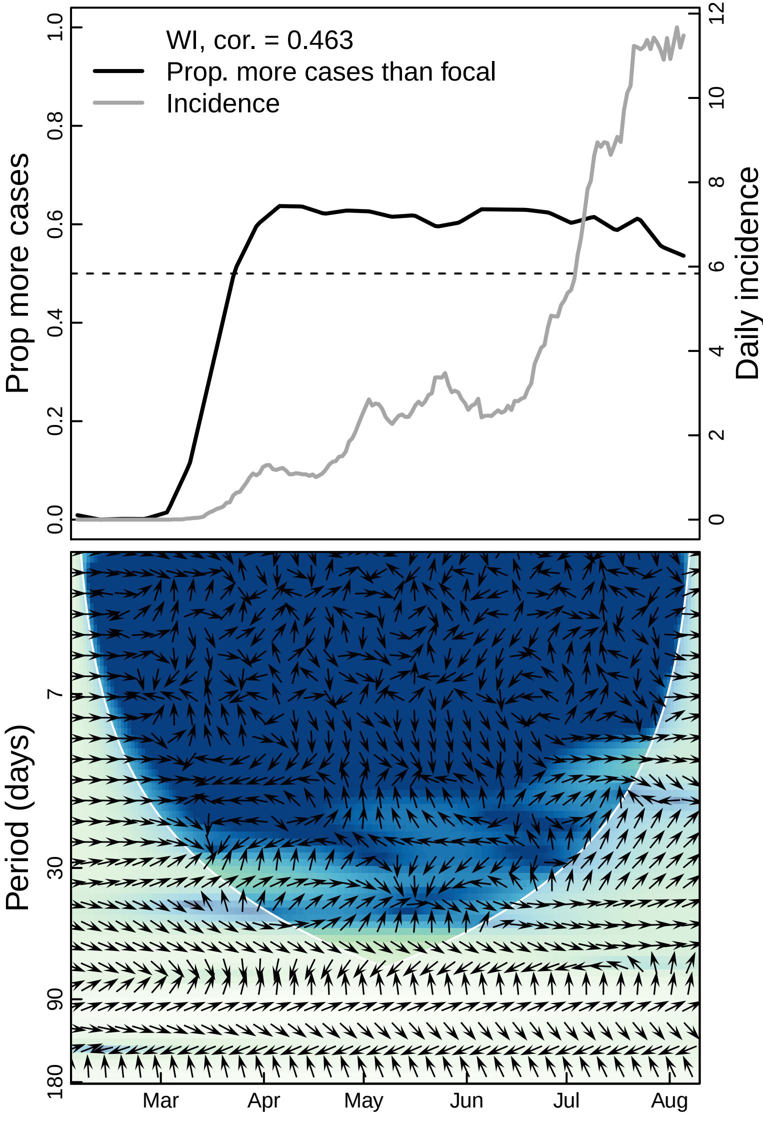

WI, cor. = 0.463
Prop. more cases than focal
Incidence
Prop more cases
Daily incidence
0.0
0.2
0.4
0.6
0.8
1.0
0
2
4
6
8
10
12
Period (days)
7
30
90
180
Mar
Apr
May
Jun
Jul
Aug

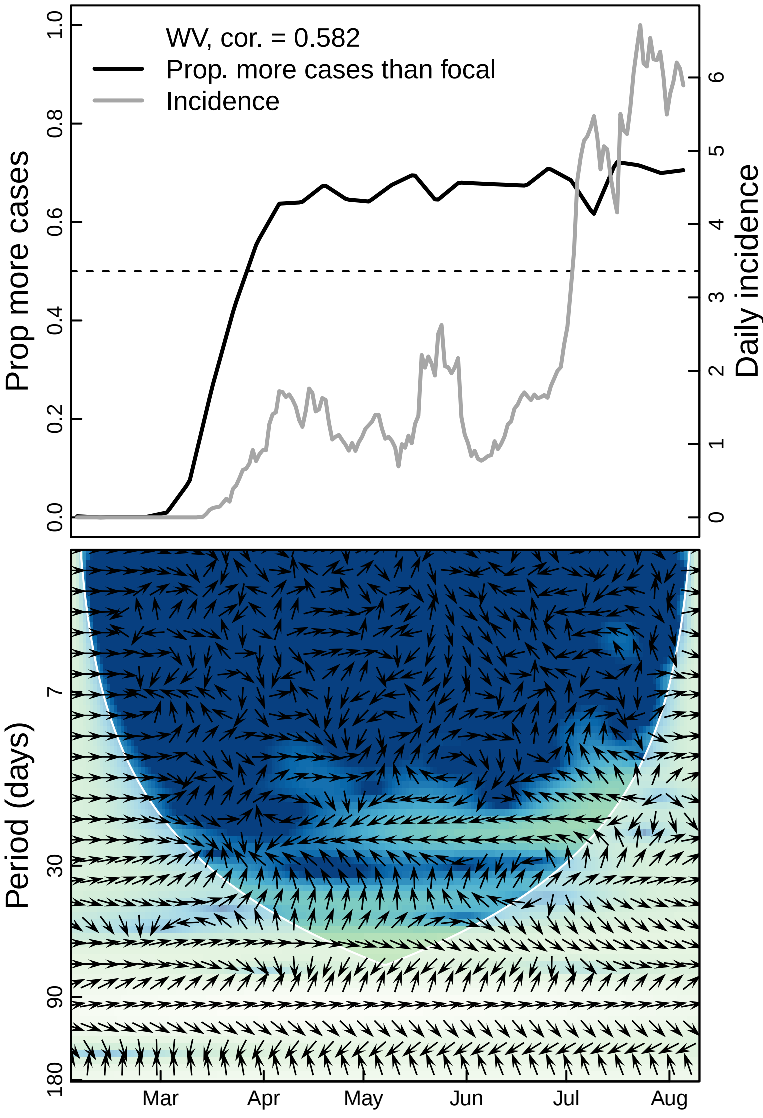

WV, cor. = 0.582
Prop. more cases than focal
Incidence
Prop more cases
Daily incidence
0.0
0.2
0.4
0.6
0.8
1.0
0
1
2
3
4
5
6
Period (days)
7
30
90
180
Mar
Apr
May
Jun
Jul
Aug

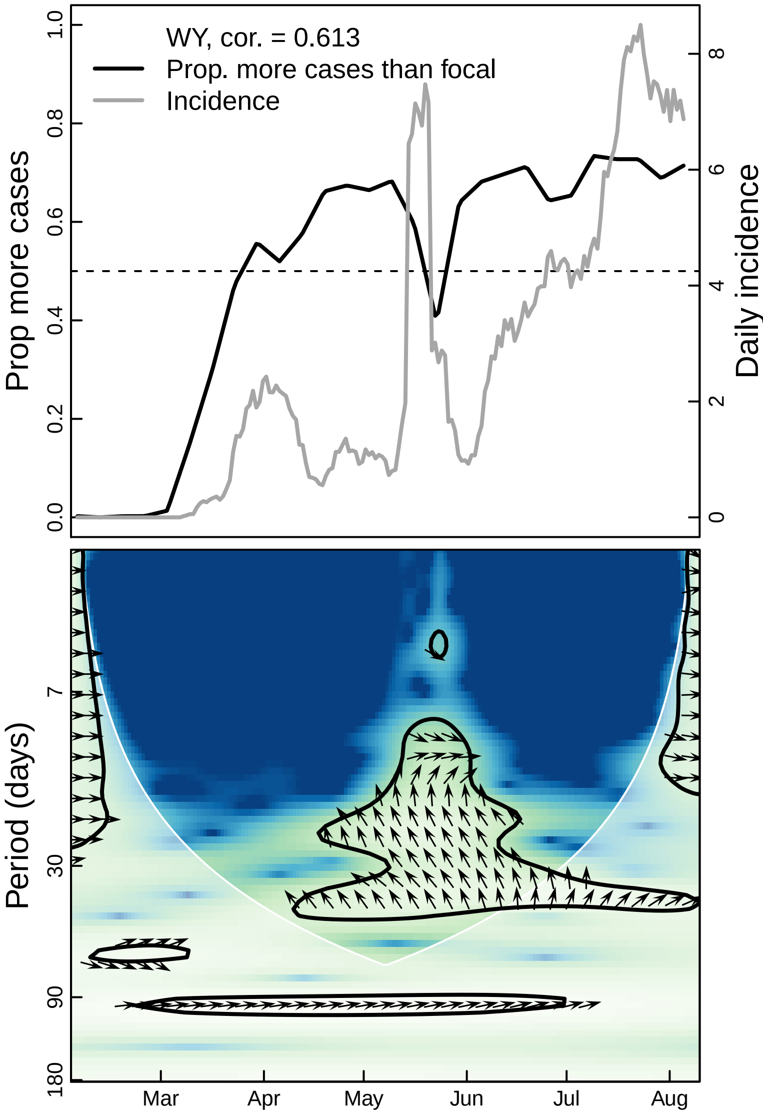

WY, cor. = 0.613
Prop. more cases than focal
Incidence
Prop more cases
Daily incidence
0.0
0.2
0.4
0.6
0.8
1.0
0
2
4
6
8
Period (days)
7
30
90
180
Mar
Apr
May
Jun
Jul
Aug
