## Supplementary material for "The unintended consequences of inconsistent pandemic control policies": Cross-wavelets - gyms

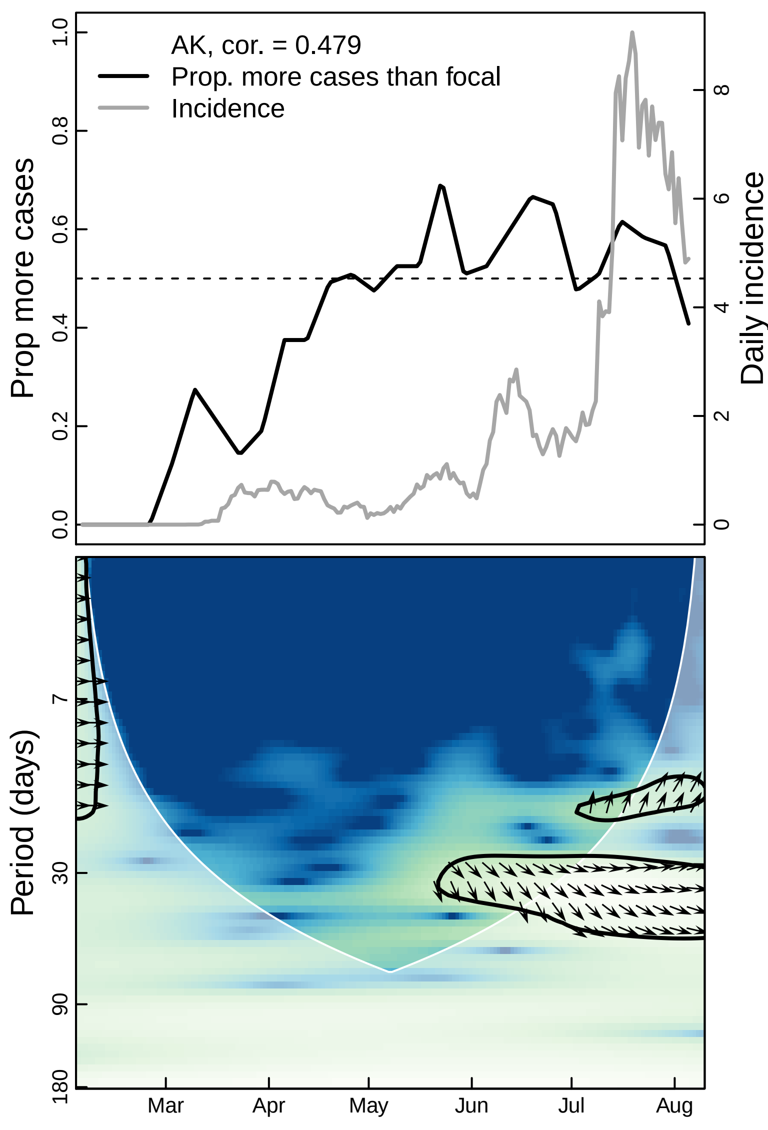
AK, cor. = 0.479
Prop. more cases than focal
Incidence
Prop more cases
Daily incidence
0.0
0.2
0.4
0.6
0.8
1.0
0
2
4
6
8
Period (days)
7
30
90
180
Mar
Apr
May
Jun
Jul
Aug

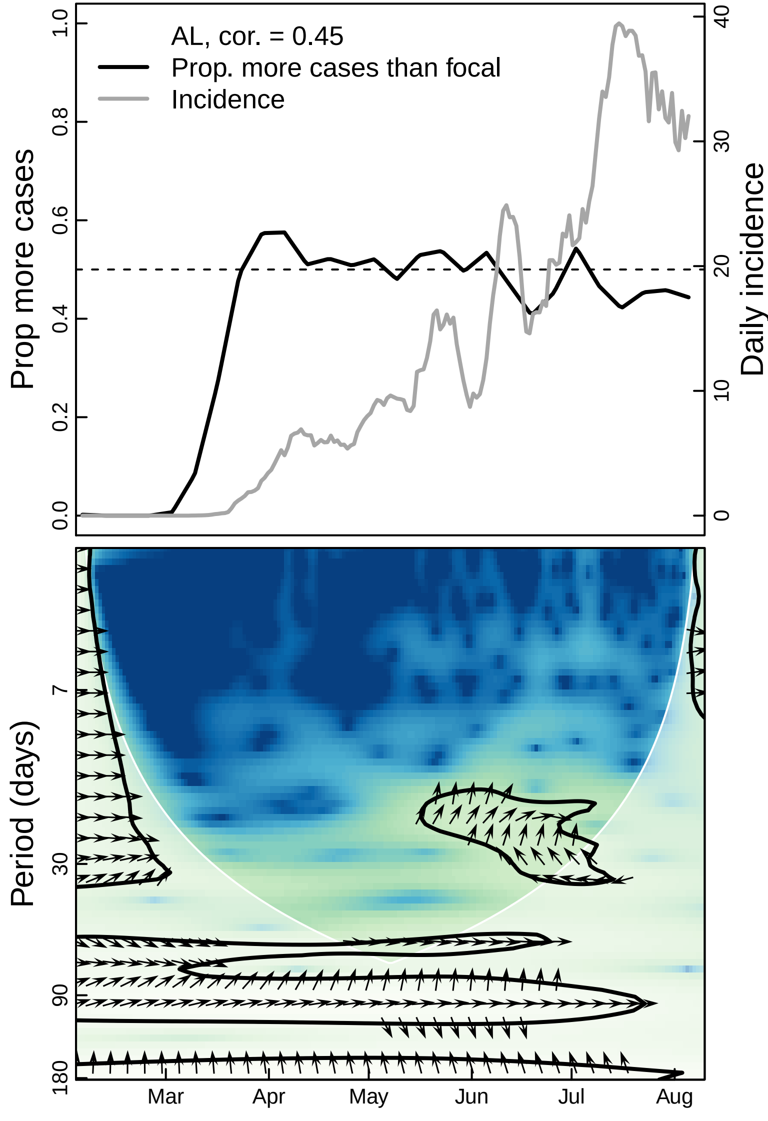

AL, cor. = 0.45
Prop. more cases than focal
Incidence
Prop more cases
Daily incidence
0.0
0.2
0.4
0.6
0.8
1.0
0
10
20
30
40
Period (days)
7
30
90
180
Mar
Apr
May
Jun
Jul
Aug

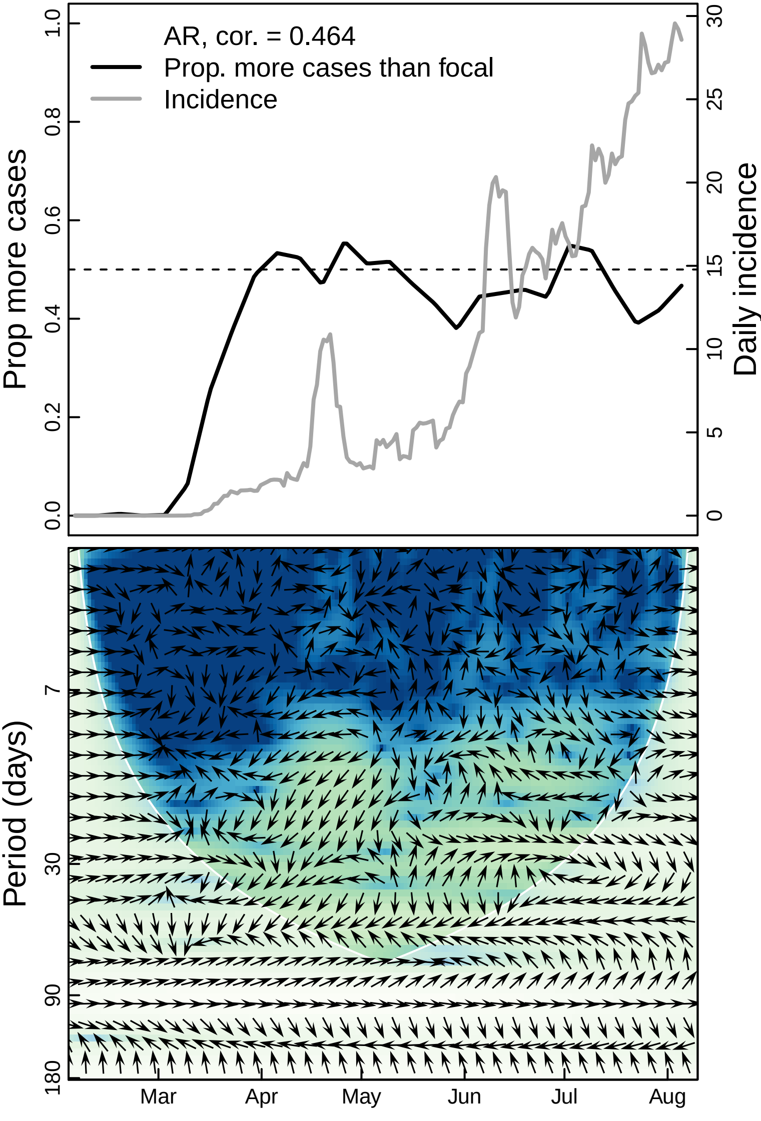

AR, cor. = 0.464
Prop. more cases than focal
Incidence
Prop more cases
Daily incidence
0.0
0.2
0.4
0.6
0.8
1.0
0
5
10
15
20
25
30
Period (days)
7
30
90
180
Mar
Apr
May
Jun
Jul
Aug

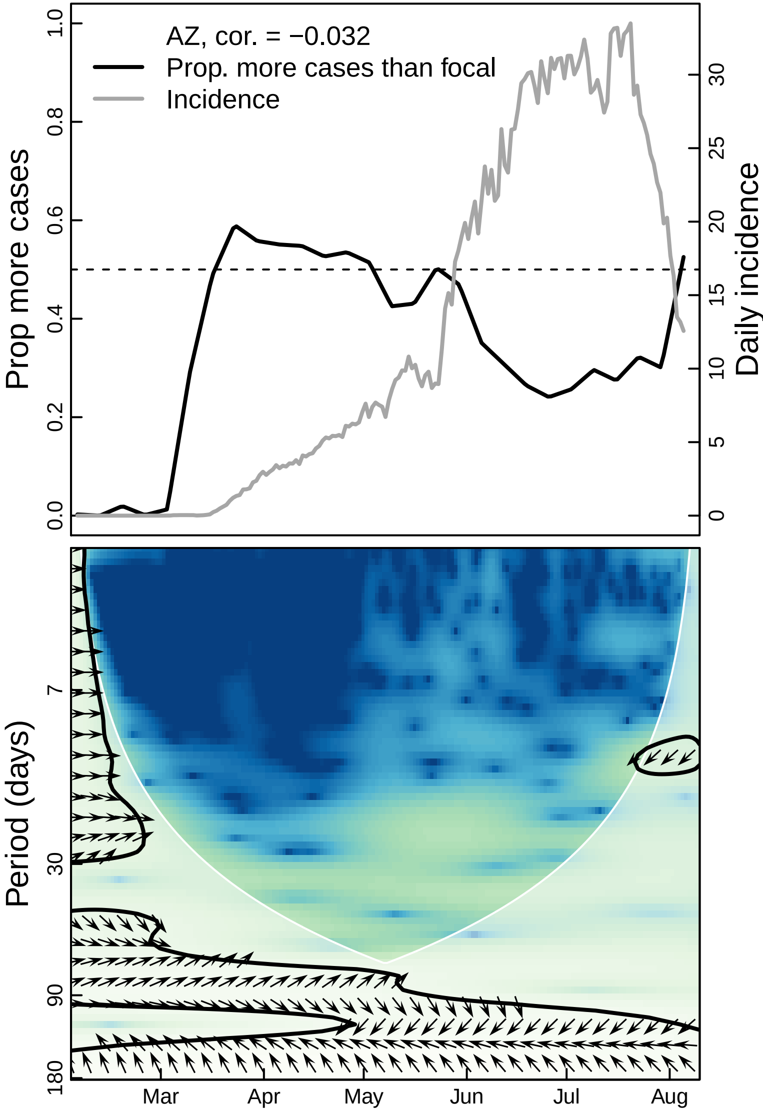
AZ, cor. = −0.032
Prop. more cases than focal
Incidence
Prop more cases
Daily incidence
0.0
0.2
0.4
0.6
0.8
1.0
0
5
10
15
20
25
30
Period (days)
7
30
90
180
Mar
Apr
May
Jun
Jul
Aug

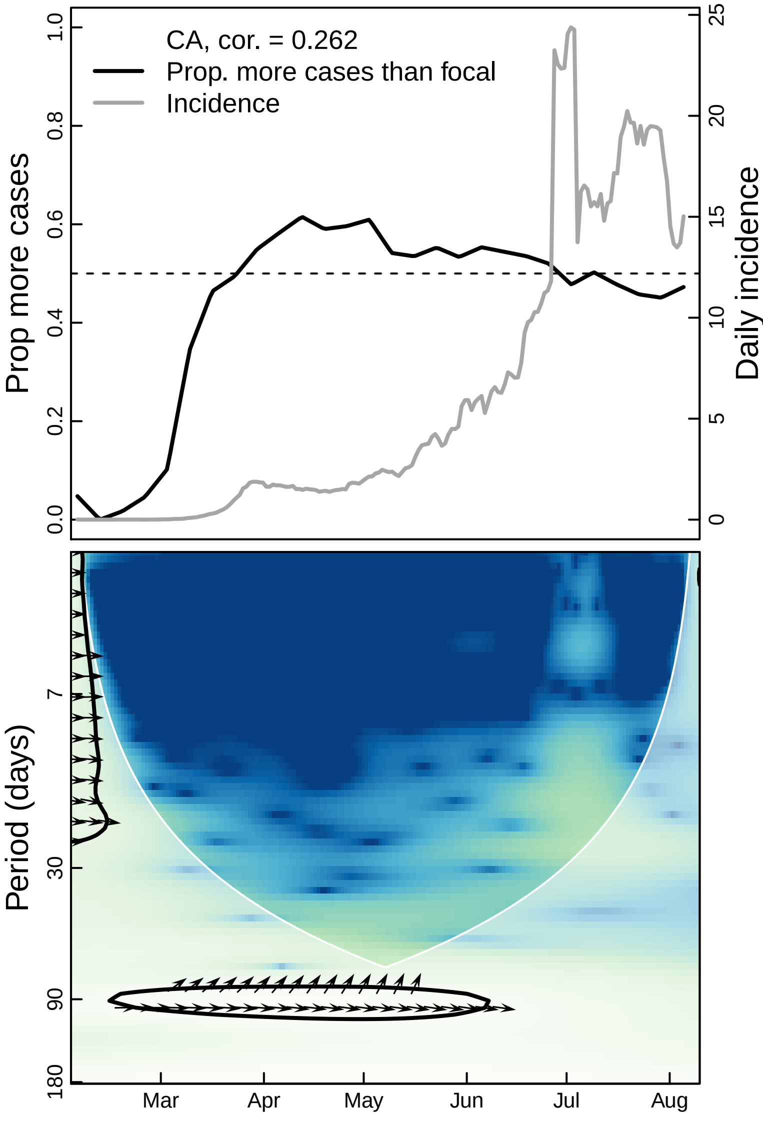

CA, cor. = 0.262
Prop. more cases than focal
Incidence
Prop more cases
Daily incidence
Period (days)
Mar
Apr
May
Jun
Jul
Aug

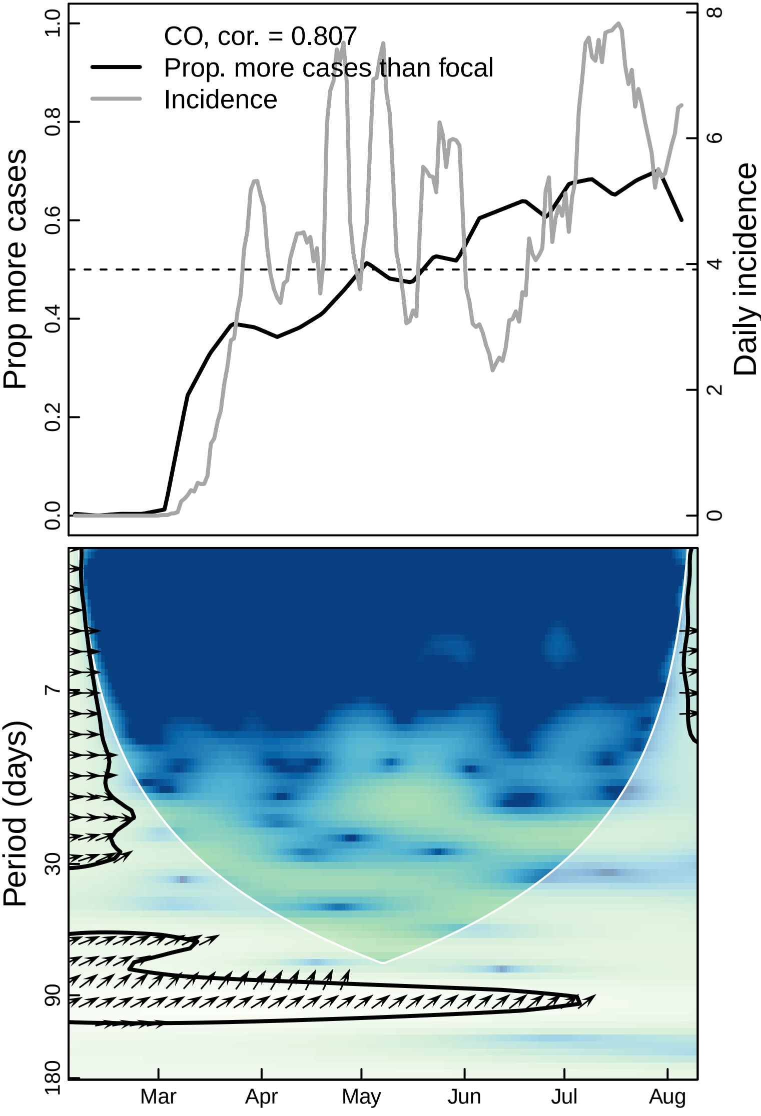

CO, cor. = 0.807
Prop. more cases than focal
Incidence
Prop more cases
Daily incidence
0.0
0.2
0.4
0.6
0.8
1.0
0
2
4
6
8
Period (days)
7
30
90
180
Mar
Apr
May
Jun
Jul
Aug

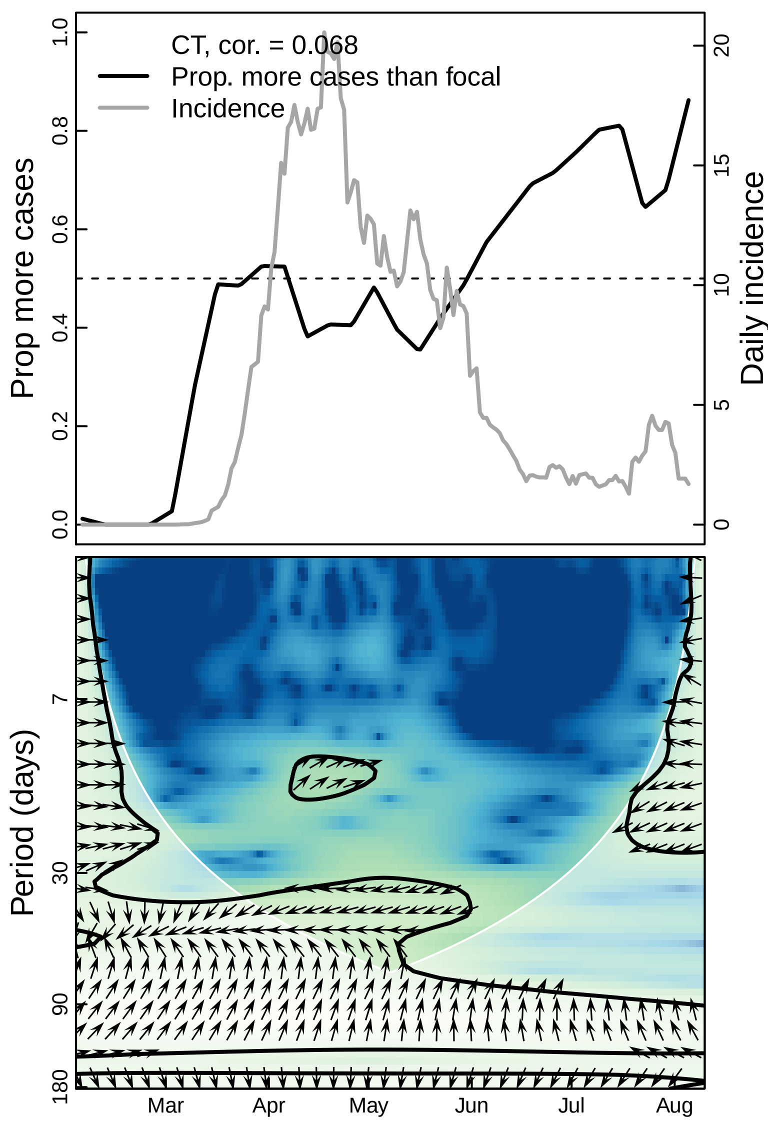

CT, cor. = 0.068
Prop. more cases than focal
Incidence
Prop more cases
Daily incidence
0.0
0.2
0.4
0.6
0.8
1.0
0
5
10
15
20
Period (days)
7
30
90
180
Mar
Apr
May
Jun
Jul
Aug

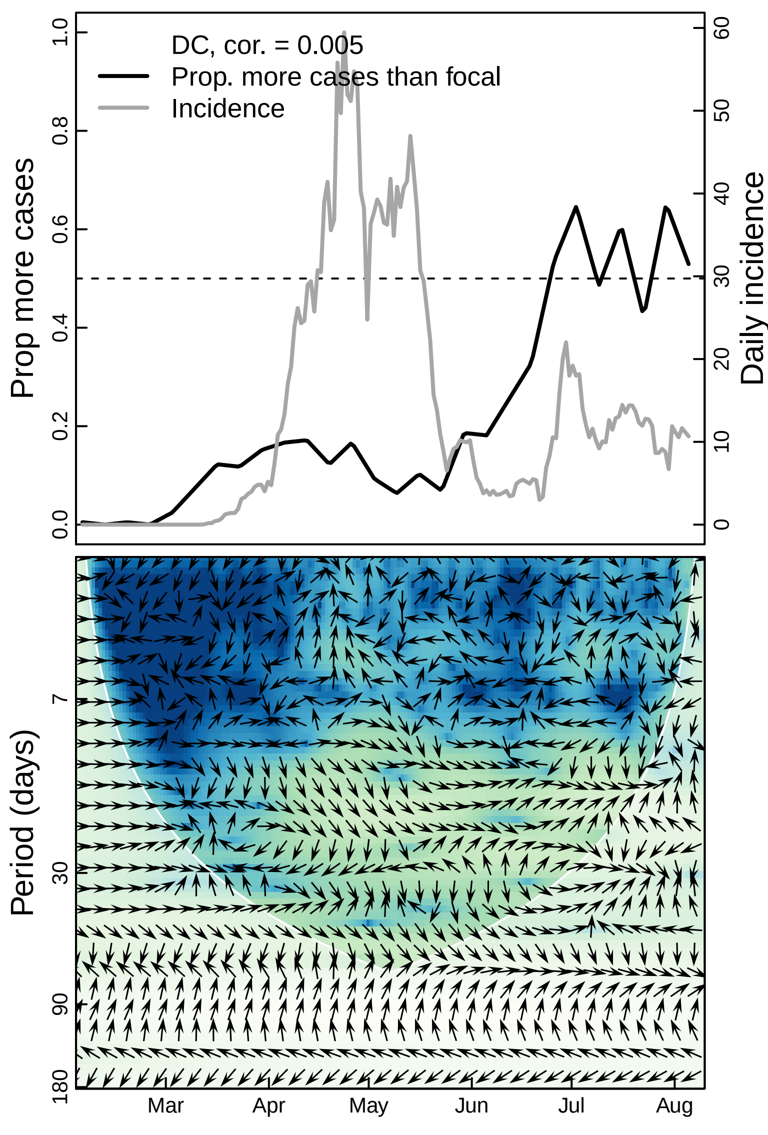

DC, cor. = 0.005
Prop. more cases than focal
Incidence
Prop more cases
Daily incidence
0.0
0.2
0.4
0.6
0.8
1.0
0
10
20
30
40
50
60
Period (days)
7
30
90
180
Mar
Apr
May
Jun
Jul
Aug

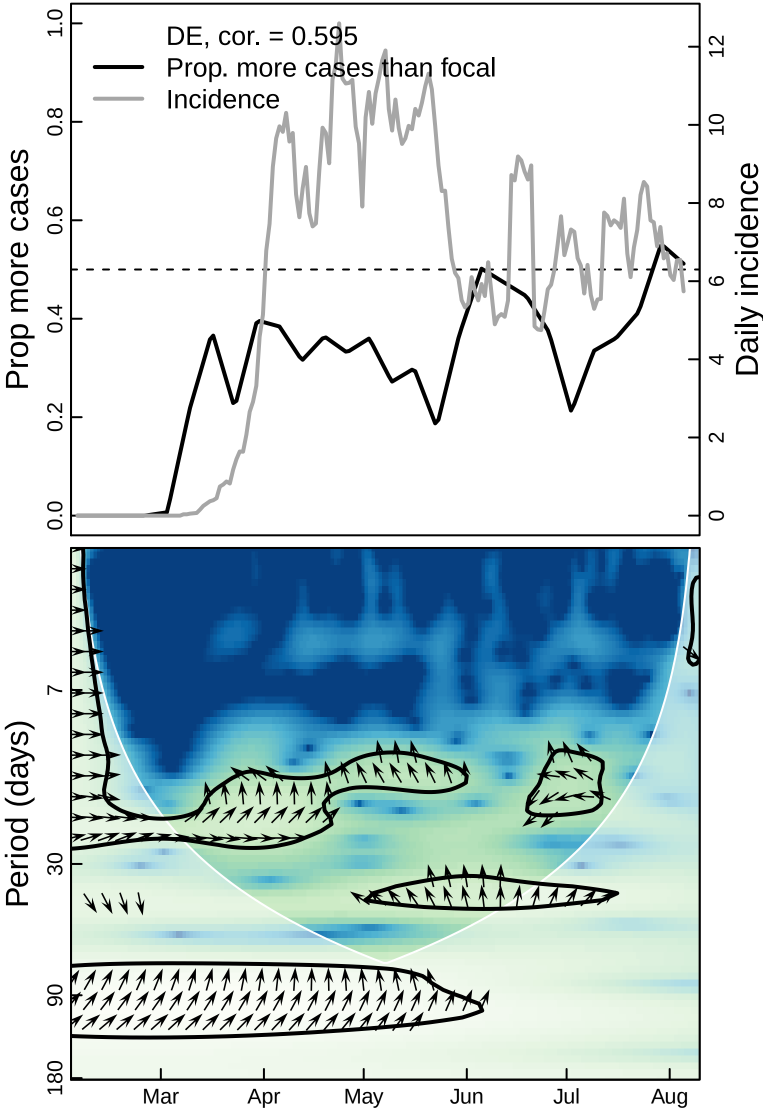
DE, cor. = 0.595
Prop. more cases than focal
Incidence
Prop more cases
Daily incidence
0.0
0.2
0.4
0.6
0.8
1.0
0
2
4
6
8
10
12
Period (days)
7
30
90
180
Mar
Apr
May
Jun
Jul
Aug

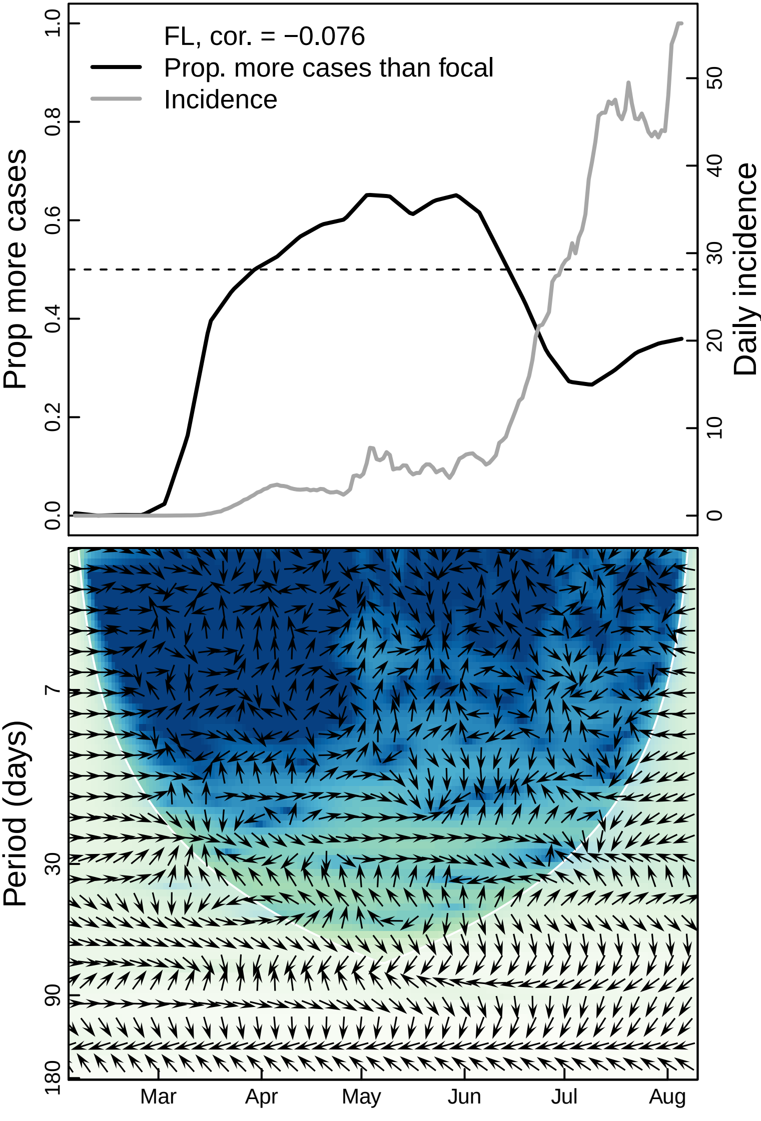

FL, cor. = −0.076
Prop. more cases than focal
Incidence
Prop more cases
Daily incidence
0.0
0.2
0.4
0.6
0.8
1.0
0
10
20
30
40
50
Period (days)
7
30
90
180
Mar
Apr
May
Jun
Jul
Aug

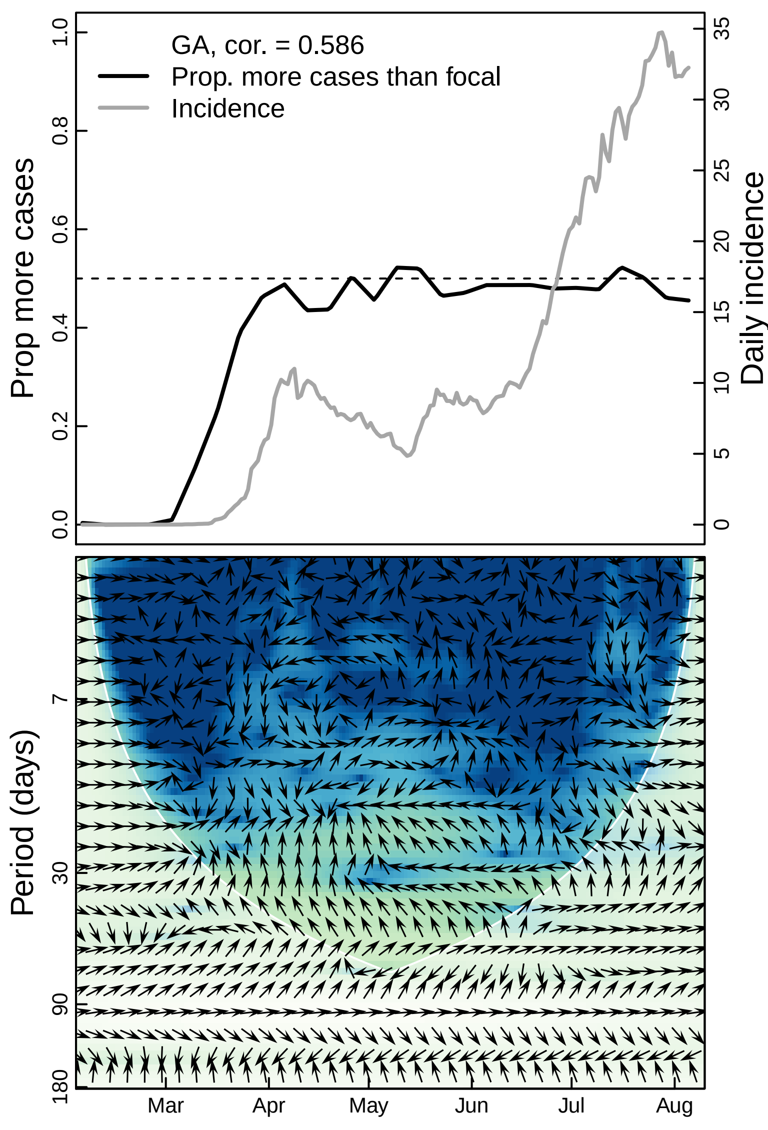

GA, cor. = 0.586
Prop. more cases than focal
Incidence
Prop more cases
Daily incidence
0.0
0.2
0.4
0.6
0.8
1.0
0
5
10
15
20
25
30
35
Period (days)
7
30
90
180
Mar
Apr
May
Jun
Jul
Aug

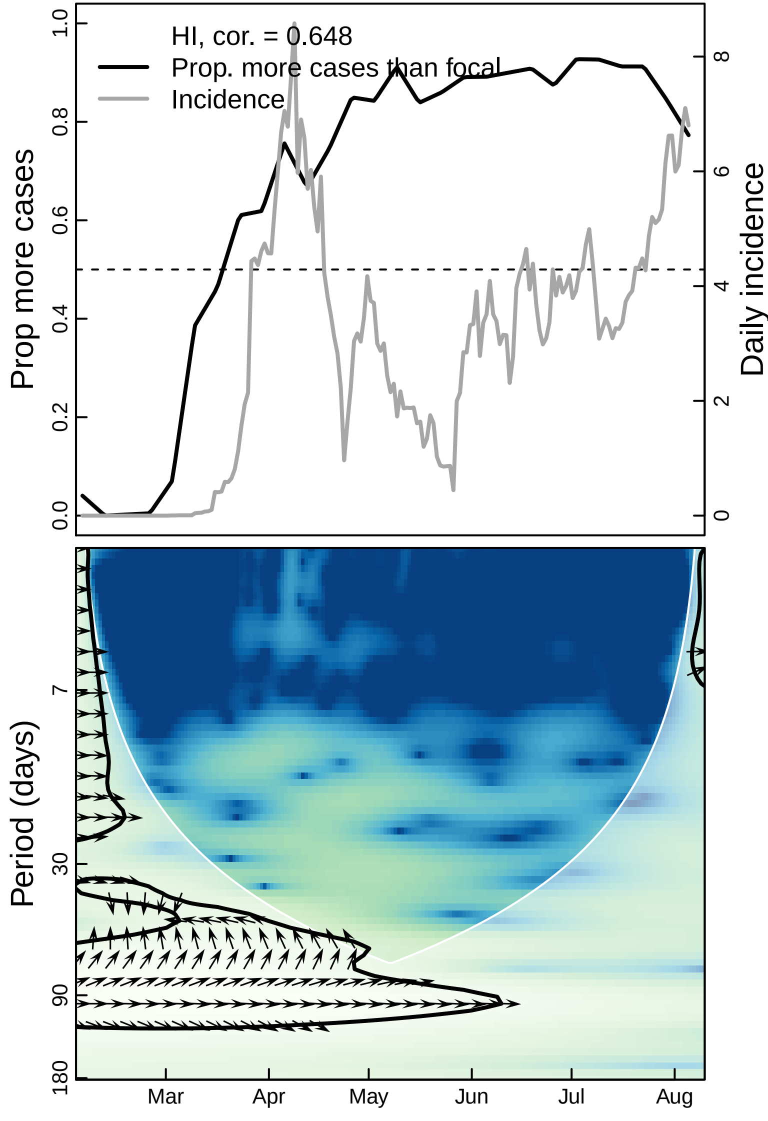
HI, cor. = 0.648
Prop. more cases than focal
Incidence
Prop more cases
Daily incidence
0.0
0.2
0.4
0.6
0.8
1.0
0
2
4
6
8
Period (days)
7
30
90
180
Mar
Apr
May
Jun
Jul
Aug

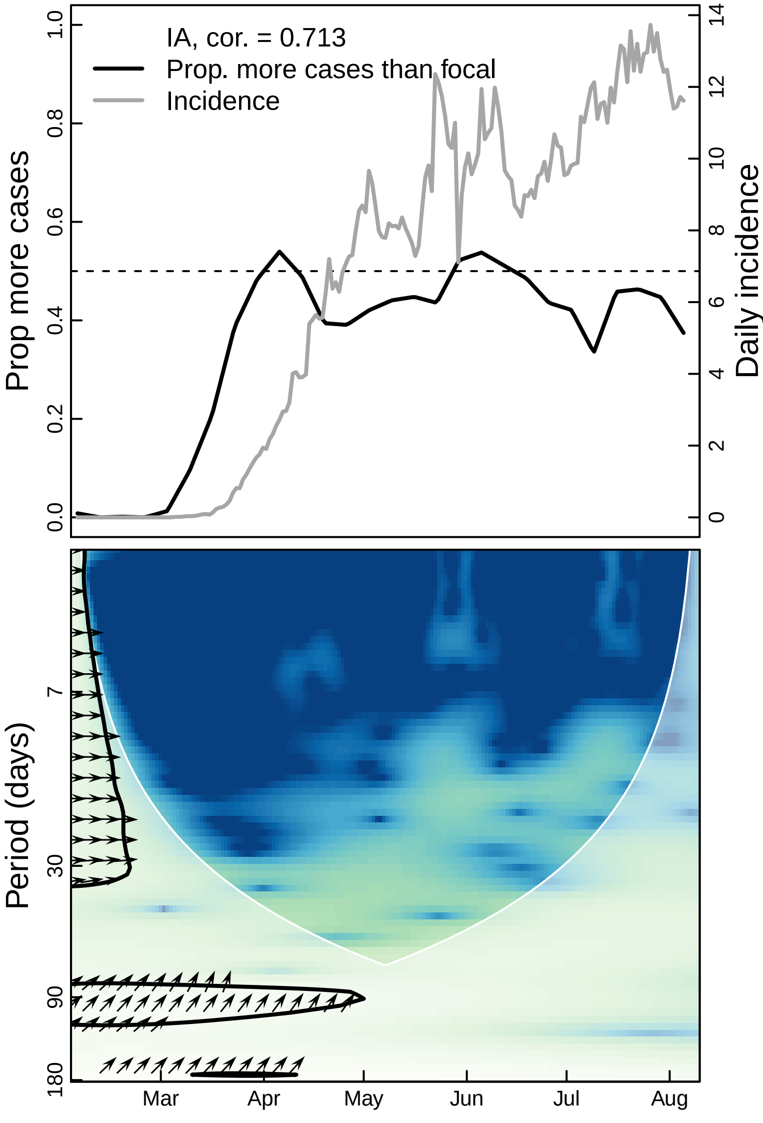

IA, cor. = 0.713
Prop. more cases than focal
Incidence
Prop more cases
Daily incidence
0.0
0.2
0.4
0.6
0.8
1.0
0
2
4
6
8
10
12
14
Period (days)
7
30
90
180
Mar
Apr
May
Jun
Jul
Aug

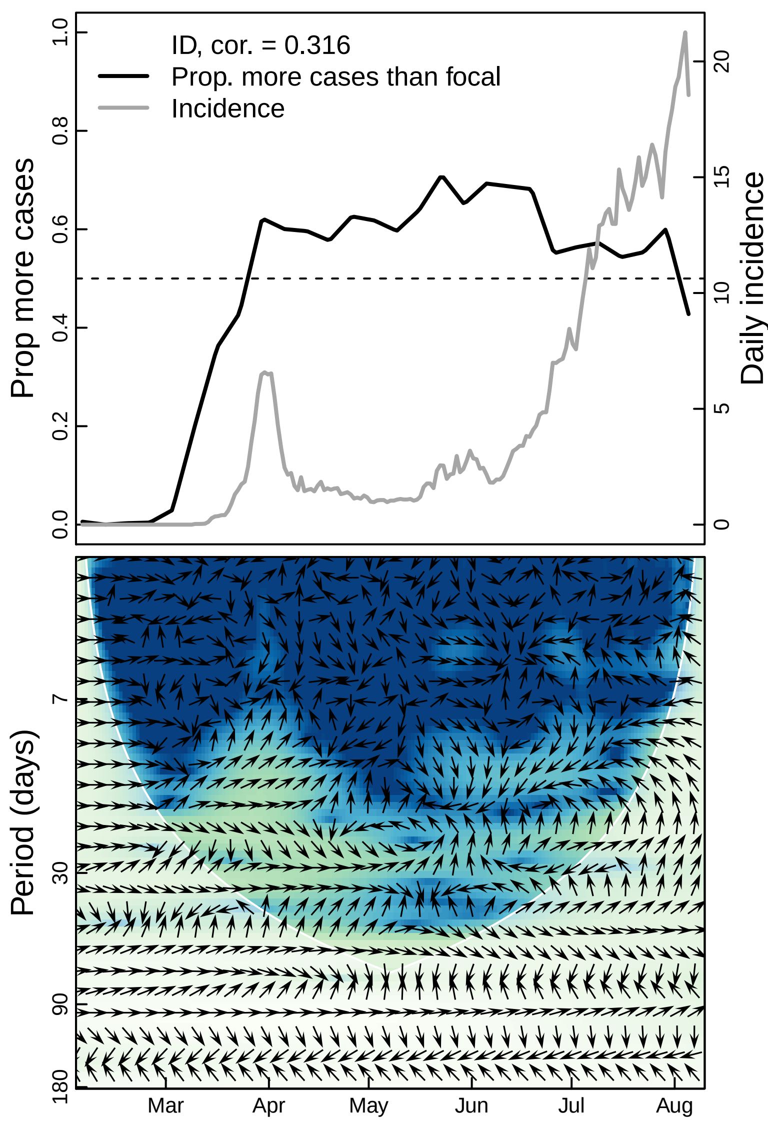

ID, cor. = 0.316
Prop. more cases than focal
Incidence
Prop more cases
Daily incidence
0.0
0.2
0.4
0.6
0.8
1.0
0
5
10
15
20
Period (days)
7
30
90
180
Mar
Apr
May
Jun
Jul
Aug

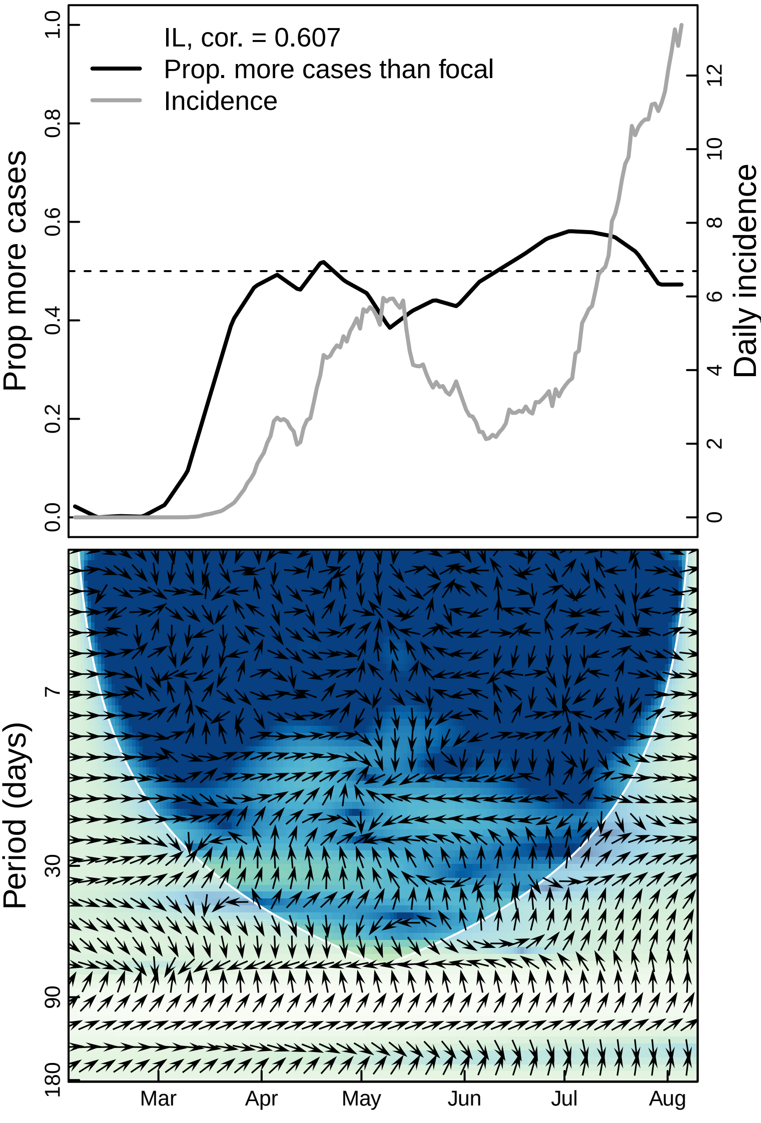

IL, cor. = 0.607
Prop. more cases than focal
Incidence
Prop more cases
Daily incidence
0.0
0.2
0.4
0.6
0.8
1.0
0
2
4
6
8
10
12
Period (days)
7
30
90
180
Mar
Apr
May
Jun
Jul
Aug

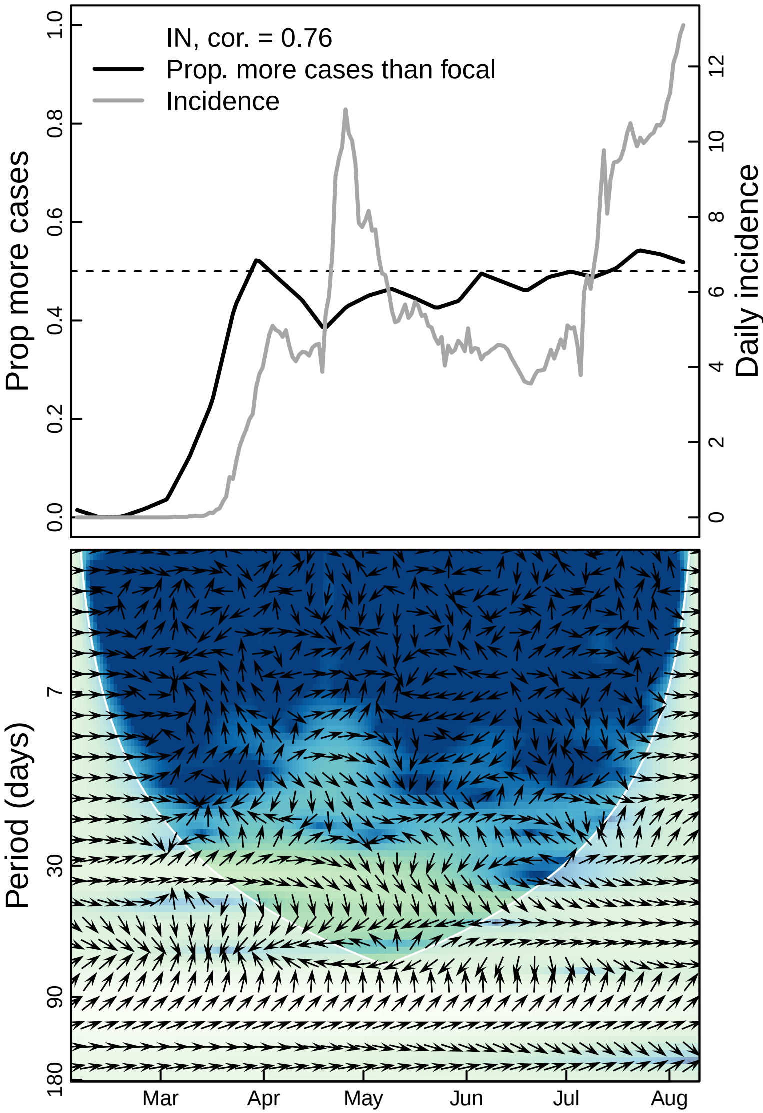
IN, cor. = 0.76
Prop. more cases than focal
Incidence
Prop more cases
Daily incidence
1.0
0.8
0.6
0.4
0.2
0.0
12
10
8
6
4
2
0
Period (days)
7
30
90
180
Mar
Apr
May
Jun
Jul
Aug

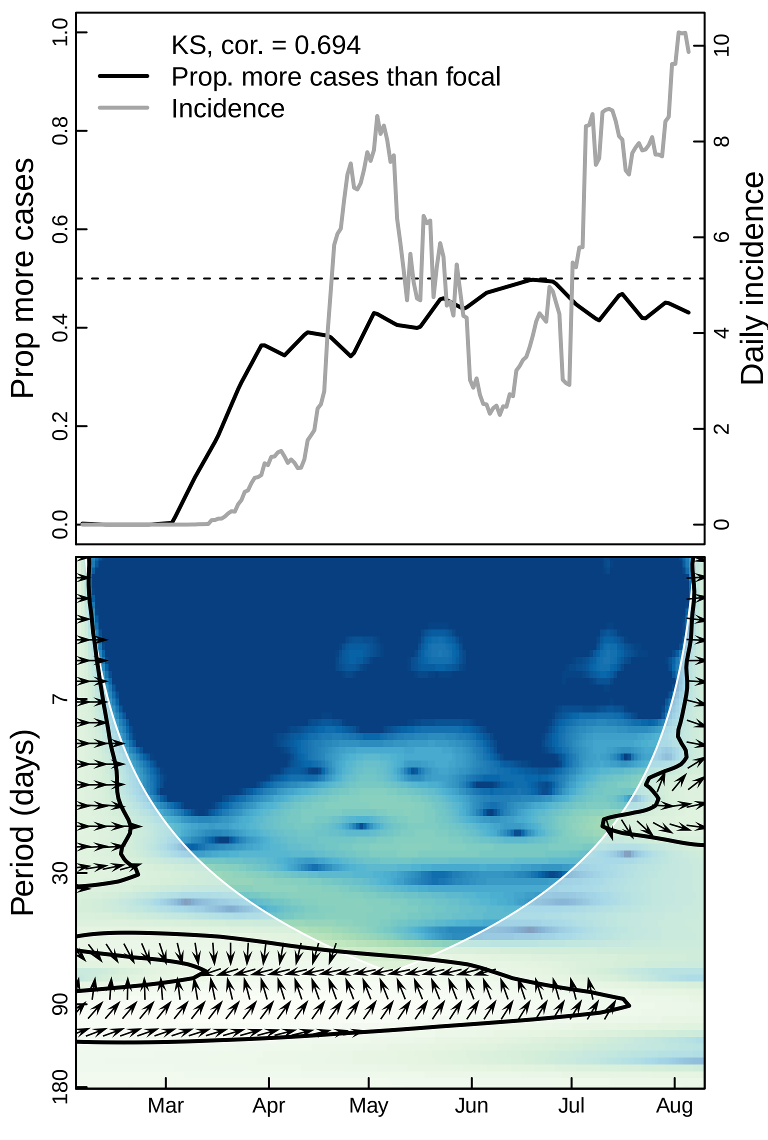

KS, cor. = 0.694
Prop. more cases than focal
Incidence
Prop more cases
Daily incidence
0.0
0.2
0.4
0.6
0.8
1.0
0
2
4
6
8
10
Period (days)
7
30
90
180
Mar
Apr
May
Jun
Jul
Aug

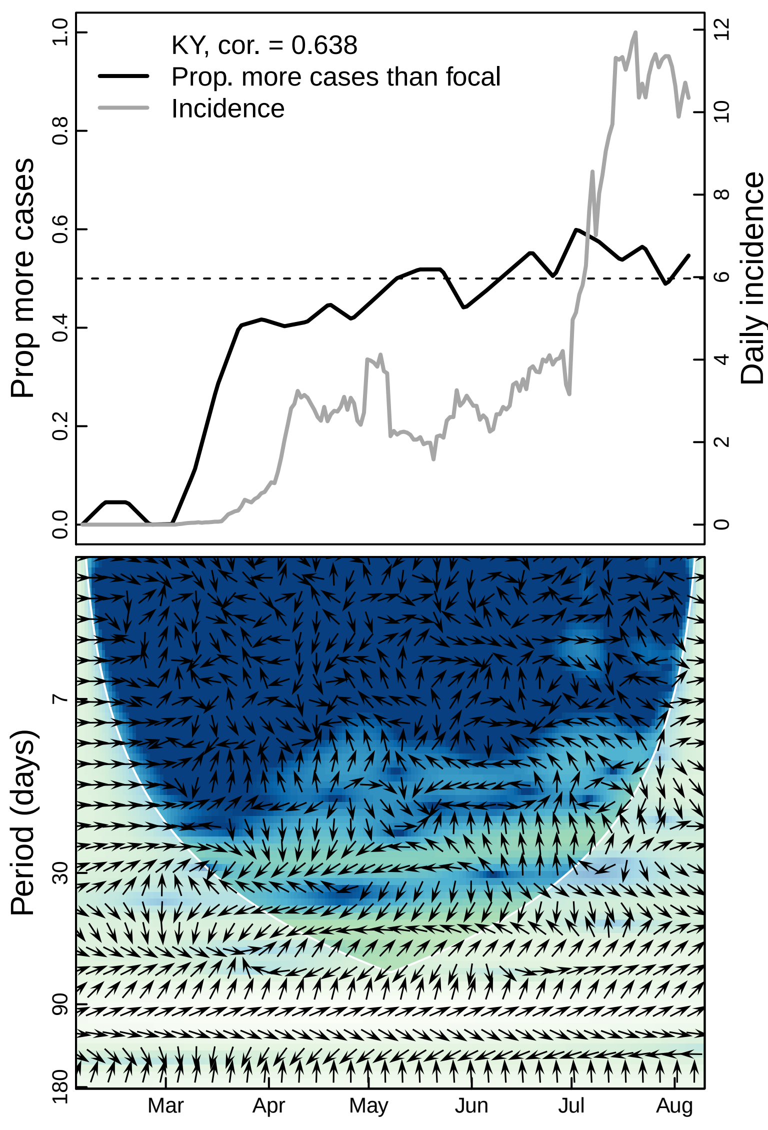
KY, cor. = 0.638
Prop. more cases than focal
Incidence
Prop more cases
Daily incidence
0.0
0.2
0.4
0.6
0.8
1.0
0
2
4
6
8
10
12
Period (days)
7
30
90
180
Mar
Apr
May
Jun
Jul
Aug

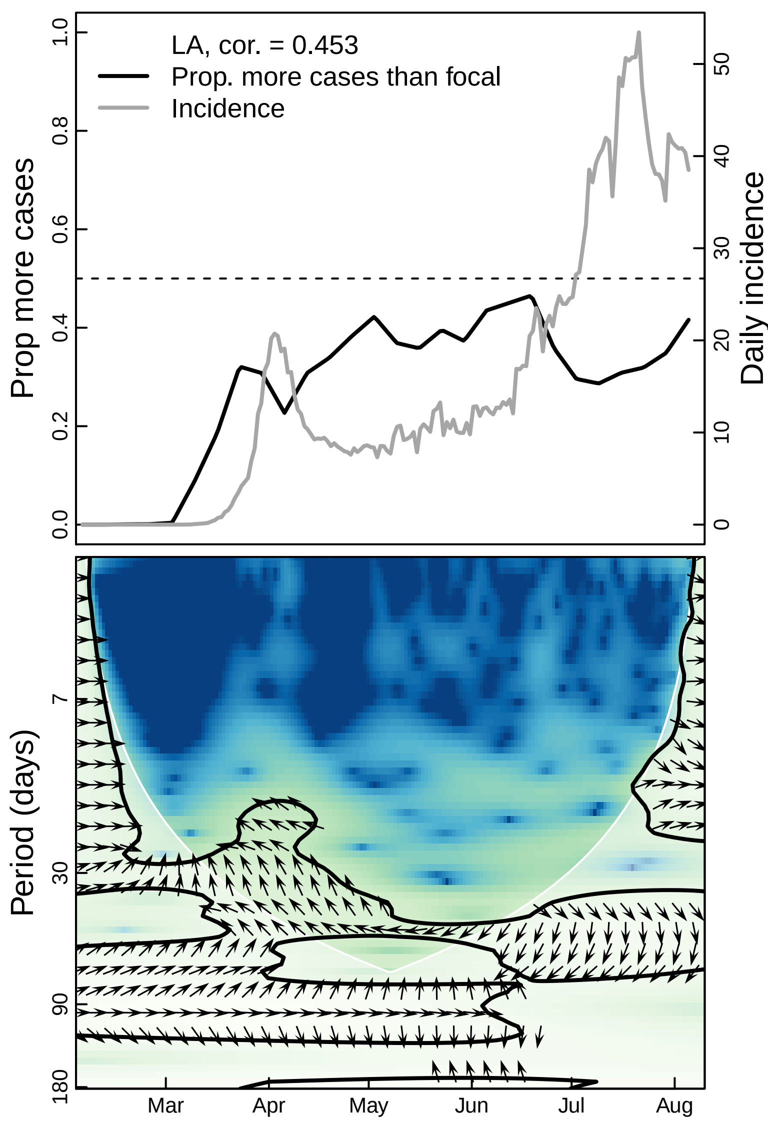

LA, cor. = 0.453
Prop. more cases than focal
Incidence
Prop more cases
Daily incidence
0.0
0.2
0.4
0.6
0.8
1.0
0
10
20
30
40
50
Period (days)
7
30
90
180
Mar
Apr
May
Jun
Jul
Aug

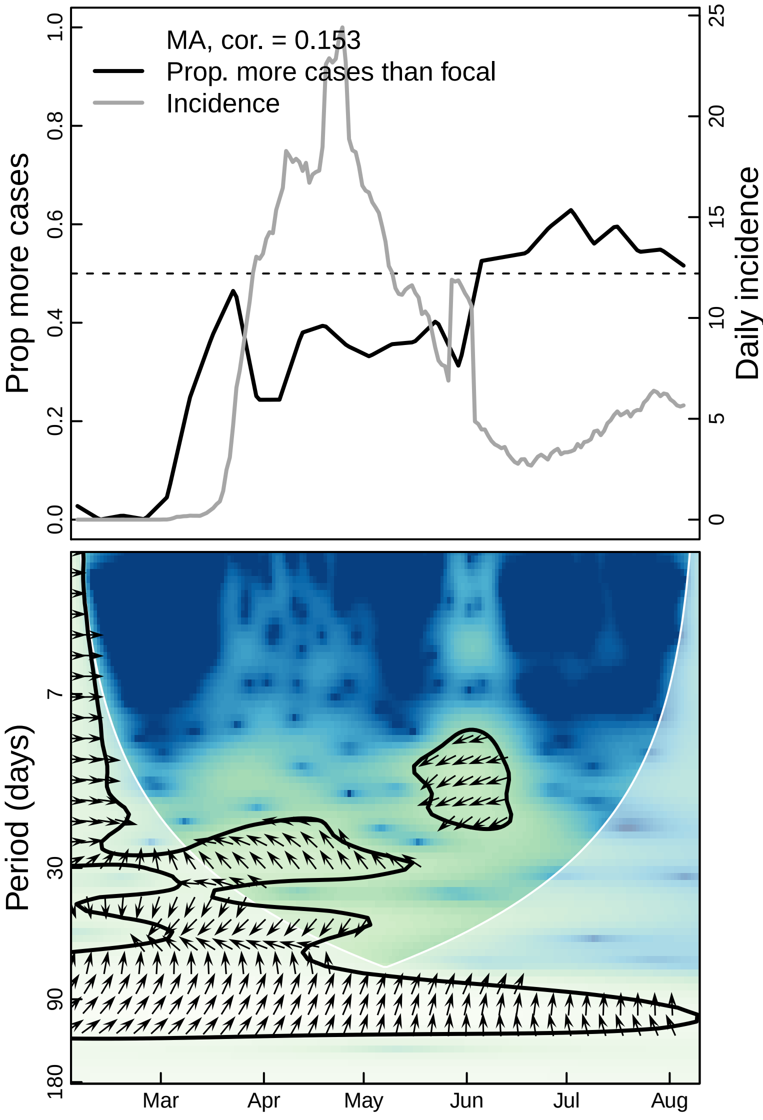
MA, cor. = 0.153
Prop. more cases than focal
Incidence
Prop more cases
Daily incidence
0.0
0.2
0.4
0.6
0.8
1.0
0
5
10
15
20
25
Period (days)
7
30
90
180
Mar
Apr
May
Jun
Jul
Aug

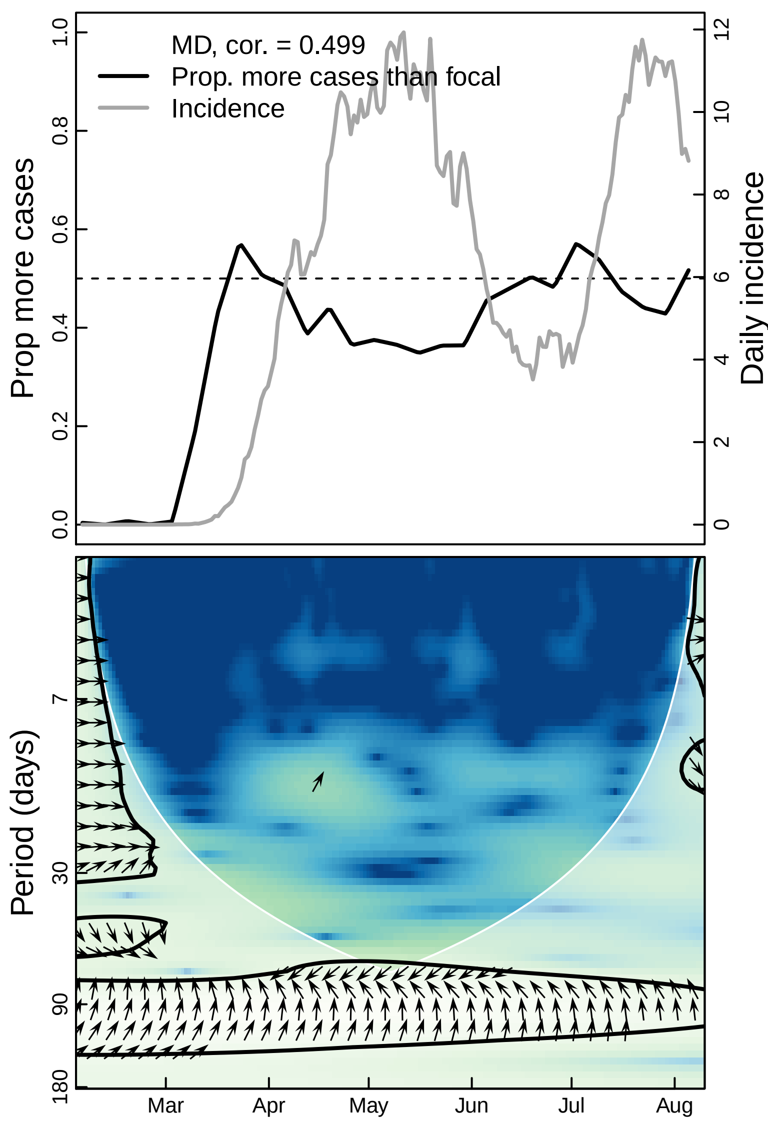

MD, cor. = 0.499
Prop. more cases than focal
Incidence
Prop more cases
Daily incidence
0.0
0.2
0.4
0.6
0.8
1.0
0
2
4
6
8
10
12
Period (days)
7
30
90
180
Mar
Apr
May
Jun
Jul
Aug

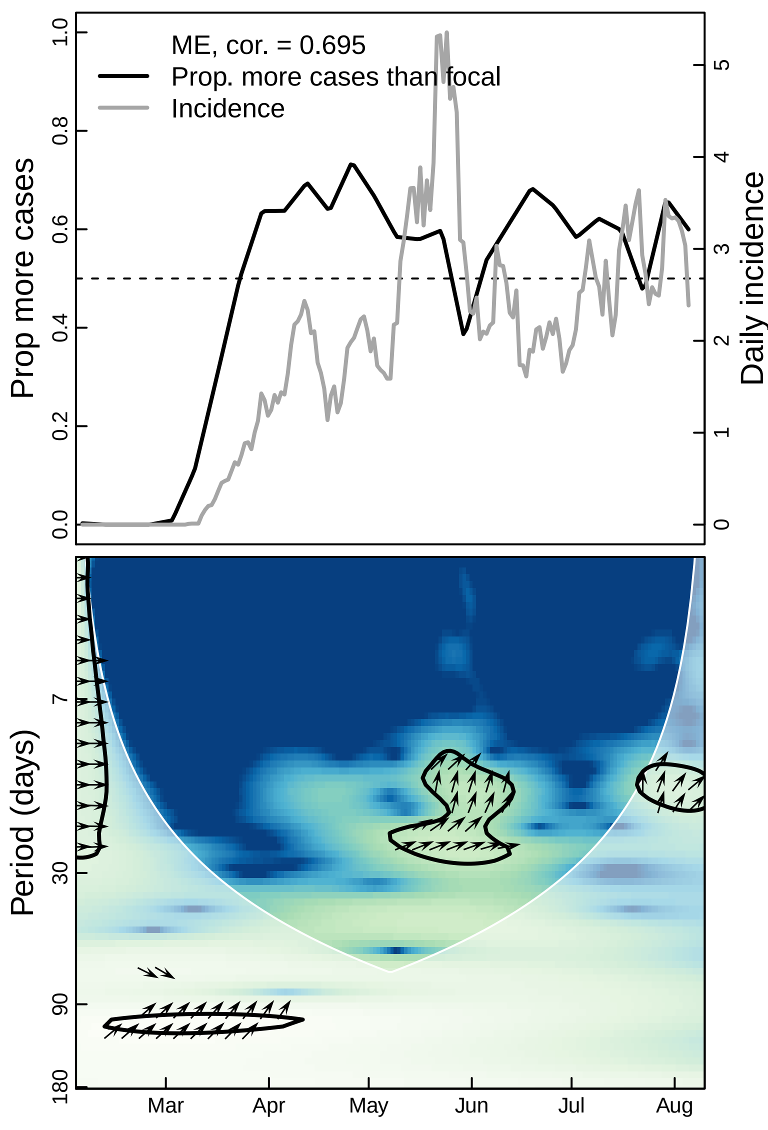
ME, cor. = 0.695
Prop. more cases than focal
Incidence
Prop more cases
Daily incidence
0.0
0.2
0.4
0.6
0.8
1.0
0
1
2
3
4
5
Period (days)
7
30
90
180
Mar
Apr
May
Jun
Jul
Aug

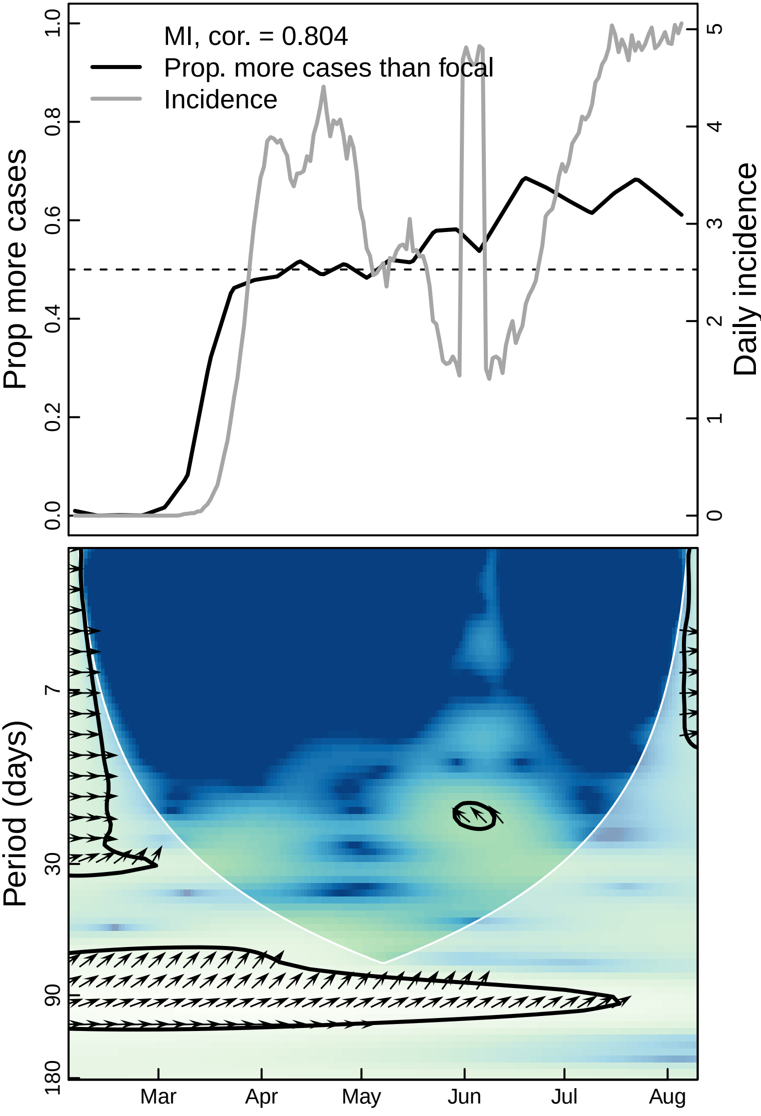

MI, cor. = 0.804
Prop. more cases than focal
Incidence
Prop more cases
Daily incidence
0.0
0.2
0.4
0.6
0.8
1.0
0
1
2
3
4
5
Period (days)
7
30
90
180
Mar
Apr
May
Jun
Jul
Aug

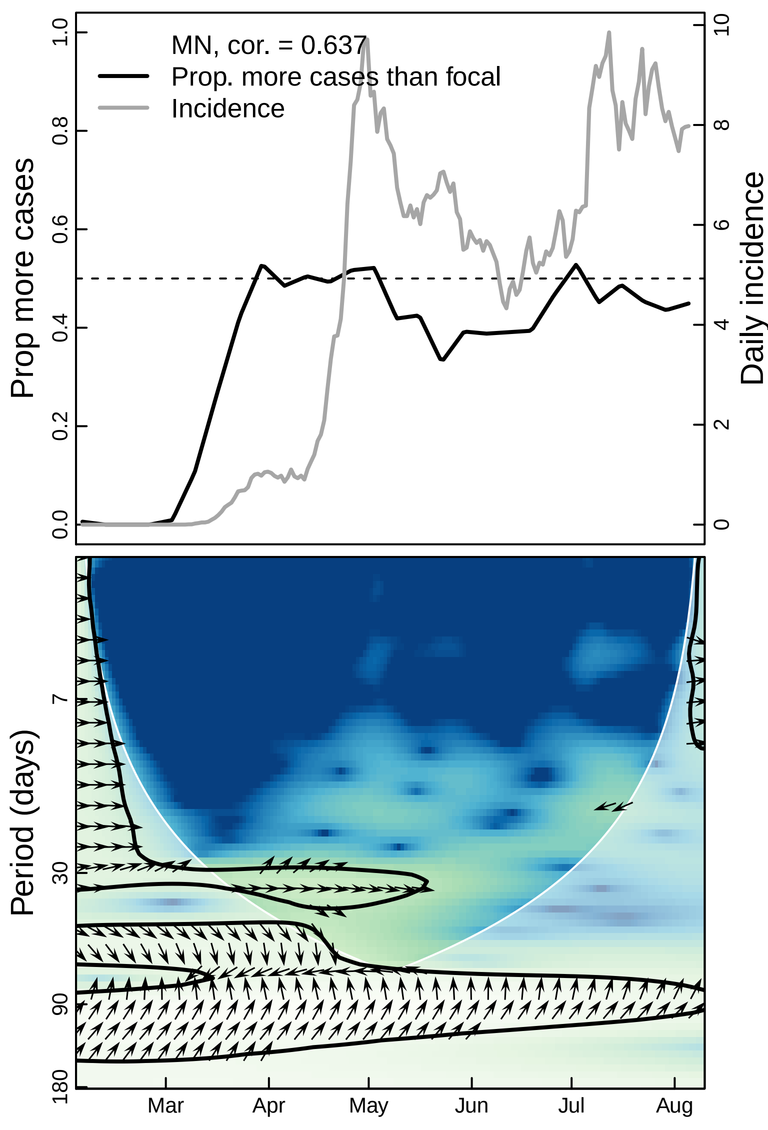
MN, cor. = 0.637
Prop. more cases than focal
Incidence
Prop more cases
Daily incidence
0.0
0.2
0.4
0.6
0.8
1.0
0
2
4
6
8
10
Period (days)
7
30
90
180
Mar
Apr
May
Jun
Jul
Aug

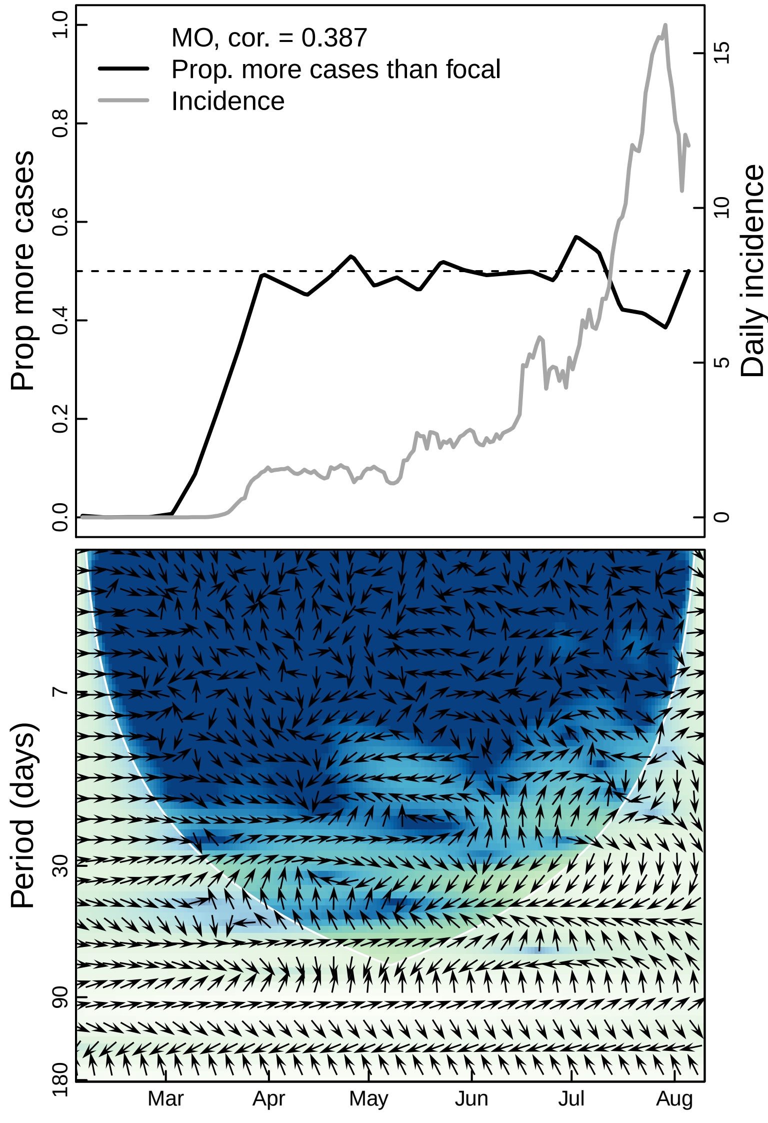

MO, cor. = 0.387
Prop. more cases than focal
Incidence
Prop more cases
Daily incidence
0.0
0.2
0.4
0.6
0.8
1.0
0
5
10
15
Period (days)
7
30
90
180
Mar
Apr
May
Jun
Jul
Aug

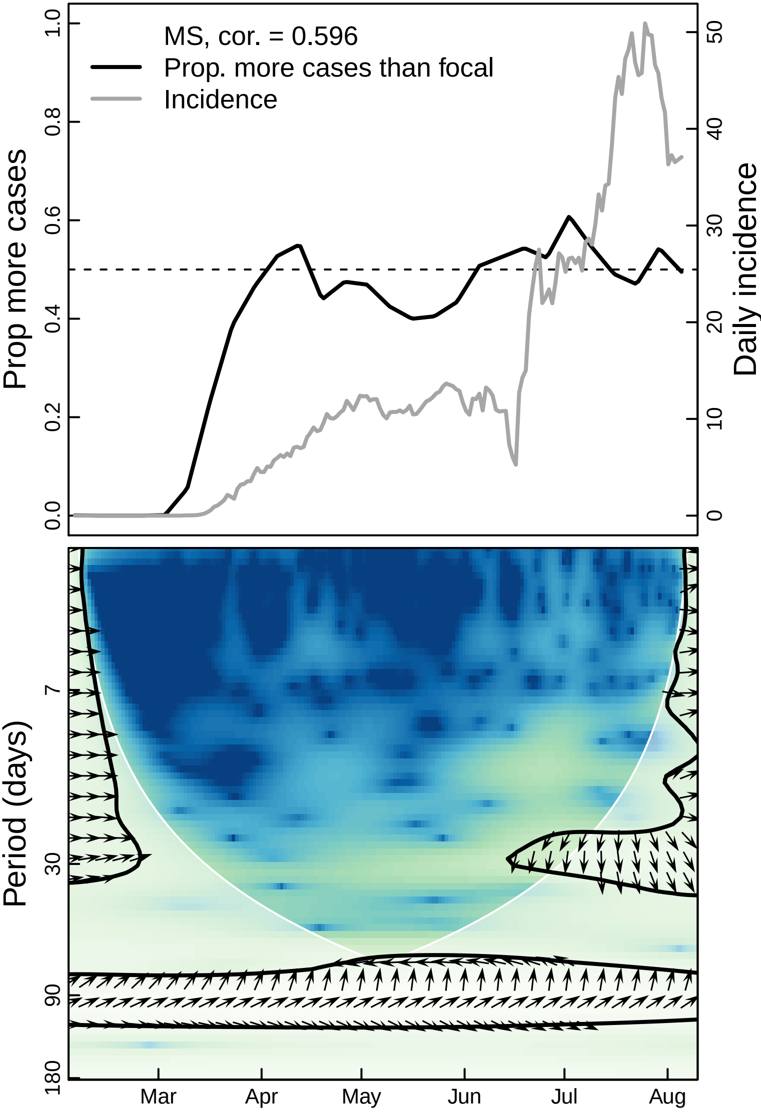

MS, cor. = 0.596
Prop. more cases than focal
Incidence
Prop more cases
Daily incidence
0.0
0.2
0.4
0.6
0.8
1.0
0
10
20
30
40
50
Period (days)
7
30
90
180
Mar
Apr
May
Jun
Jul
Aug

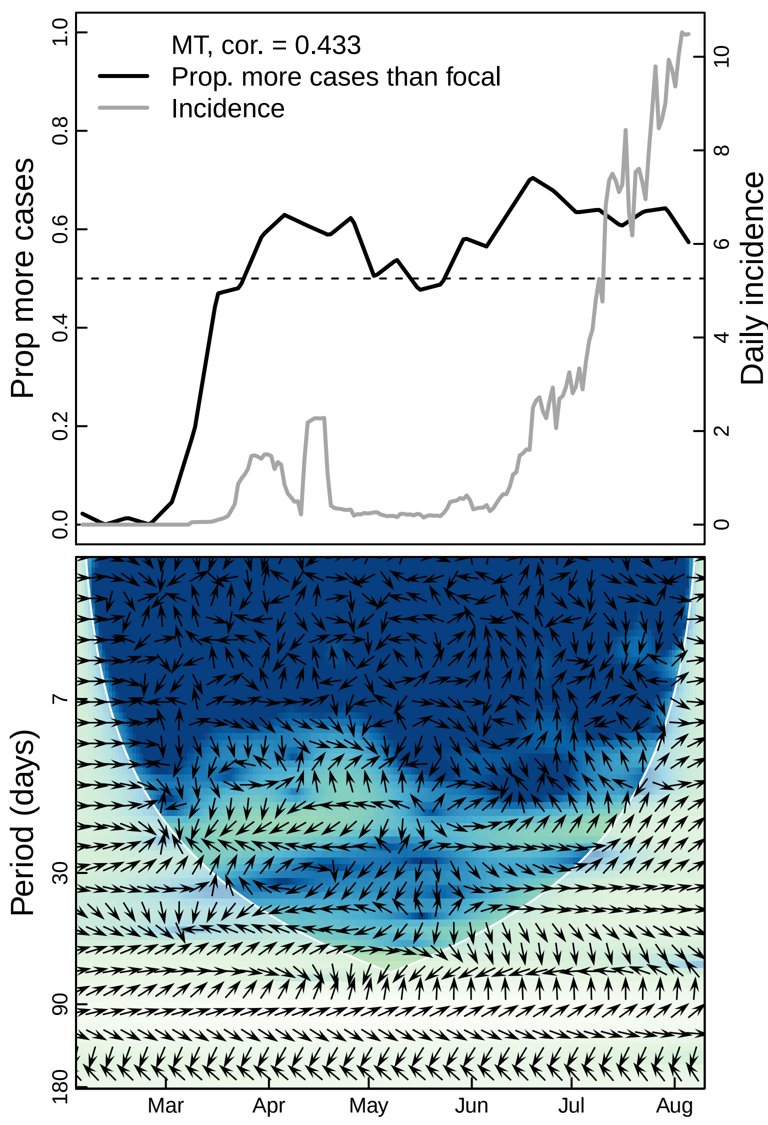

MT, cor. = 0.433
Prop. more cases than focal
Incidence
Prop more cases
Daily incidence
0.0
0.2
0.4
0.6
0.8
1.0
0
2
4
6
8
10
Period (days)
7
30
90
180
Mar
Apr
May
Jun
Jul
Aug

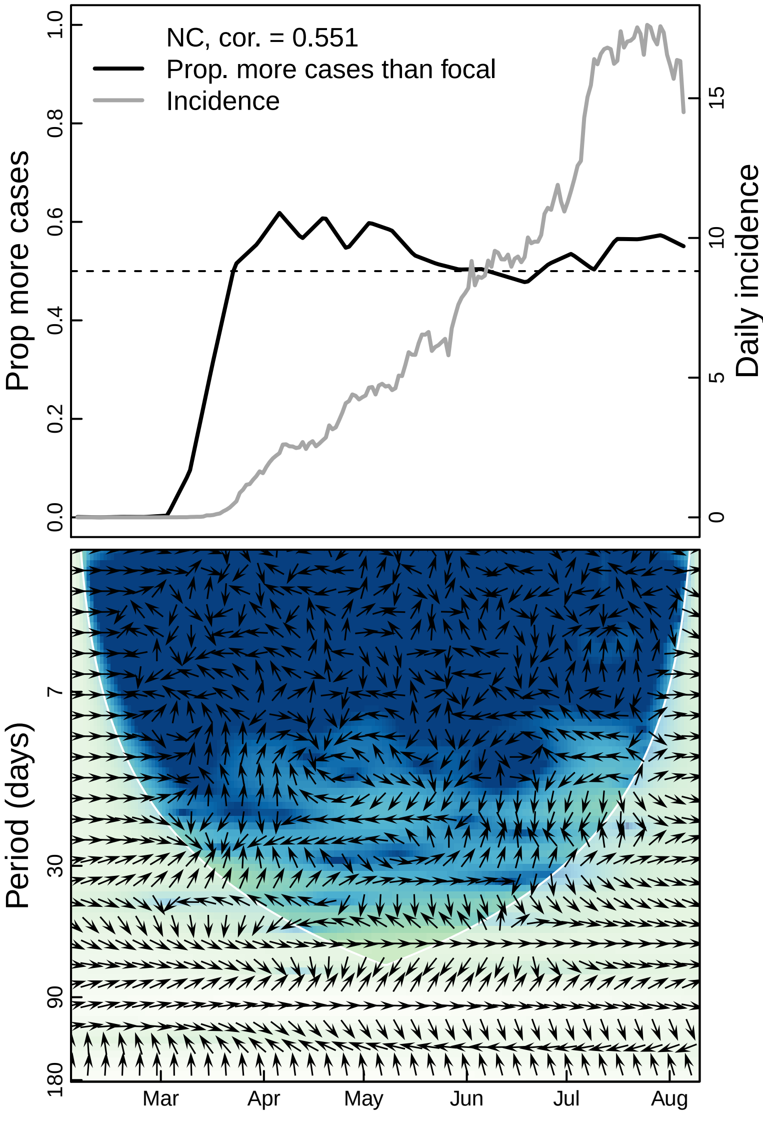

NC, cor. = 0.551
Prop. more cases than focal
Incidence
Prop more cases
Daily incidence
0.0
0.2
0.4
0.6
0.8
1.0
0
5
10
15
Period (days)
7
30
90
180
Mar
Apr
May
Jun
Jul
Aug

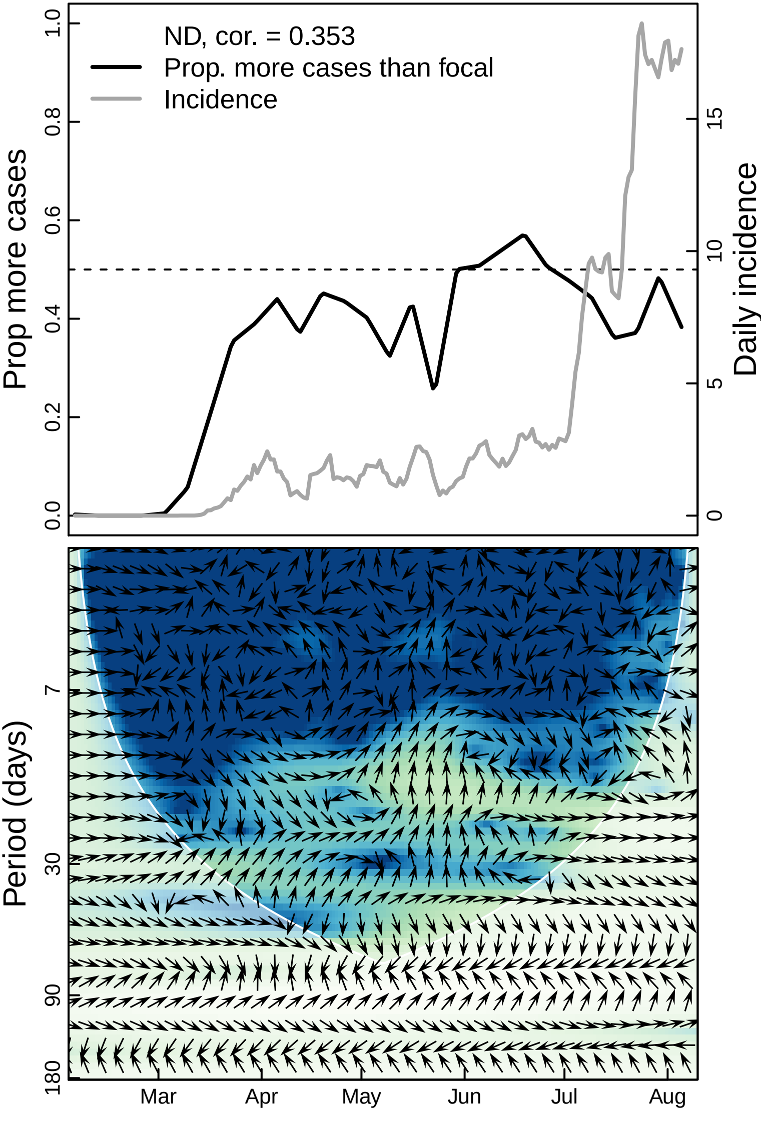

ND, cor. = 0.353
Prop. more cases than focal
Incidence
Prop more cases
Daily incidence
0.0
0.2
0.4
0.6
0.8
1.0
0
5
10
15
Period (days)
7
30
90
180
Mar
Apr
May
Jun
Jul
Aug

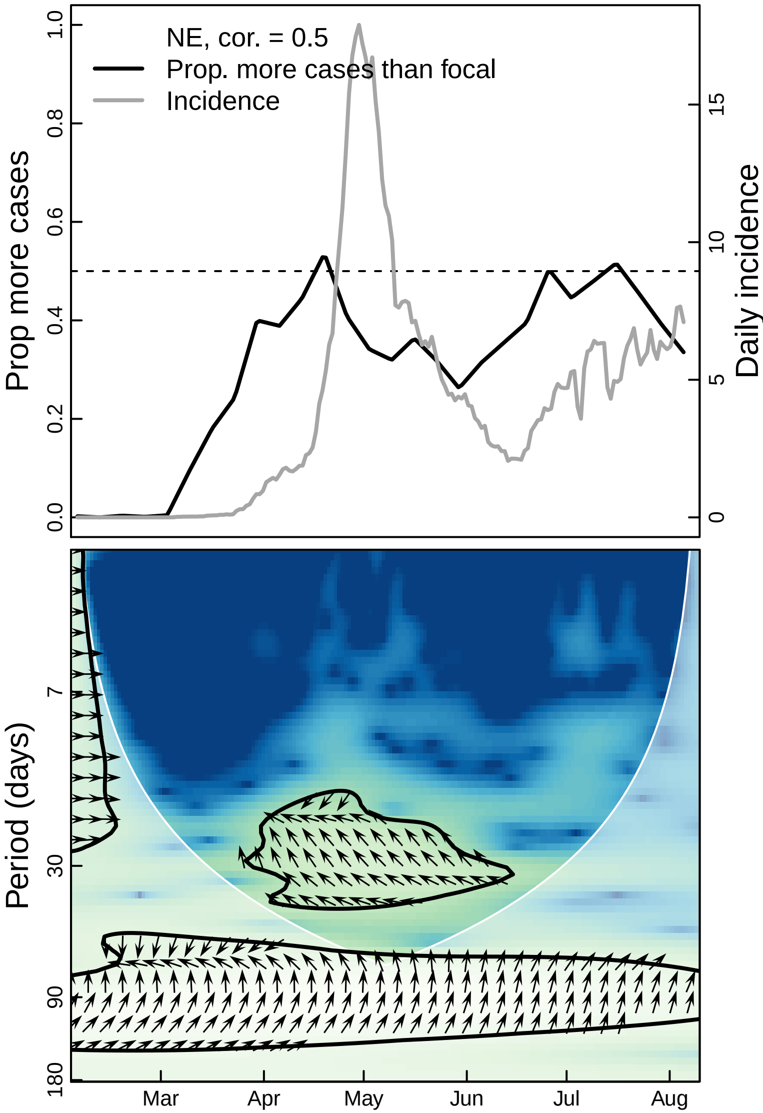
NE, cor. = 0.5
Prop. more cases than focal
Incidence
Prop more cases
Daily incidence
0.0
0.2
0.4
0.6
0.8
1.0
0
5
10
15
Period (days)
7
30
90
180
Mar
Apr
May
Jun
Jul
Aug

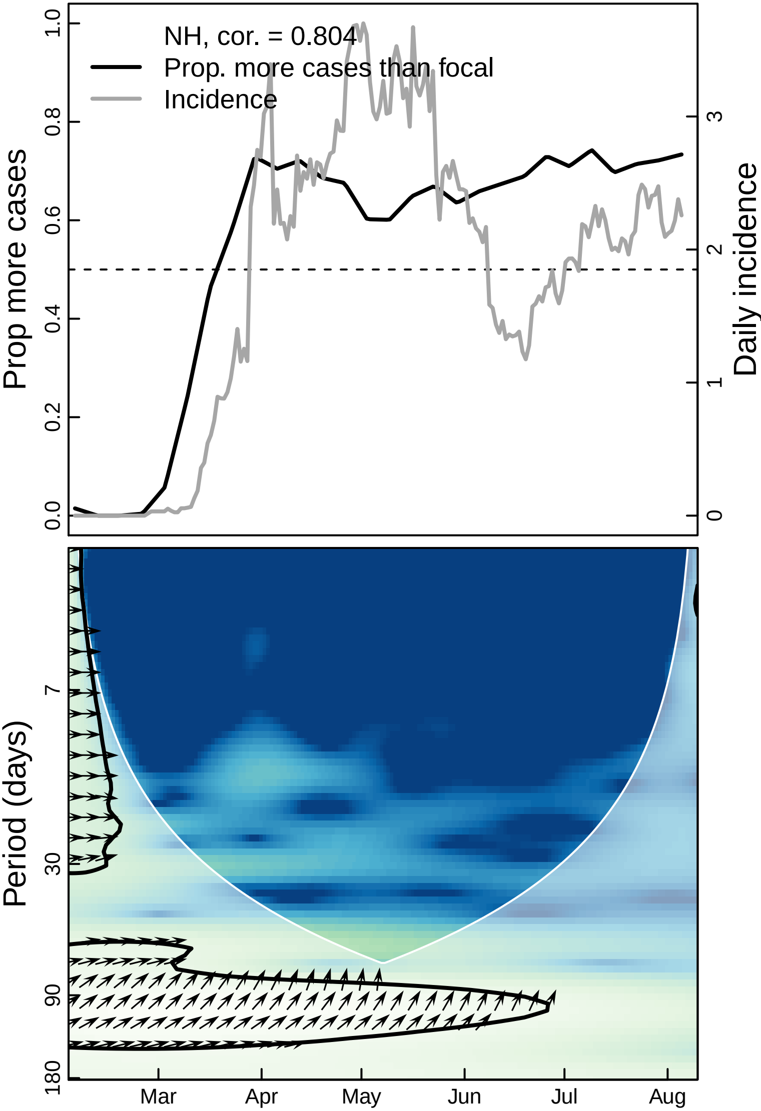

NH, cor. = 0.804
Prop. more cases than focal
Incidence
Prop more cases
Daily incidence
0.0
0.2
0.4
0.6
0.8
1.0
0
1
2
3
Period (days)
7
30
90
180
Mar
Apr
May
Jun
Jul
Aug

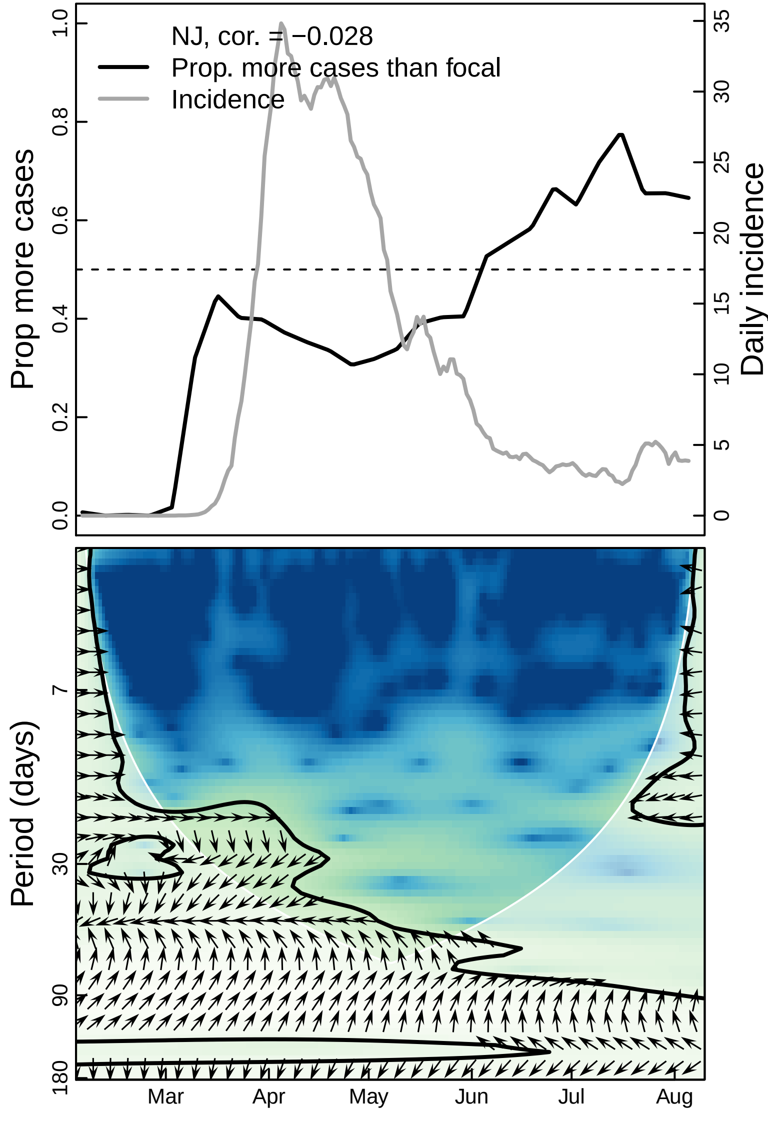

NJ, cor. = −0.028
Prop. more cases than focal
Incidence
Prop more cases
Daily incidence
0.0
0.2
0.4
0.6
0.8
1.0
0
5
10
15
20
25
30
35
Period (days)
7
30
90
180
Mar
Apr
May
Jun
Jul
Aug

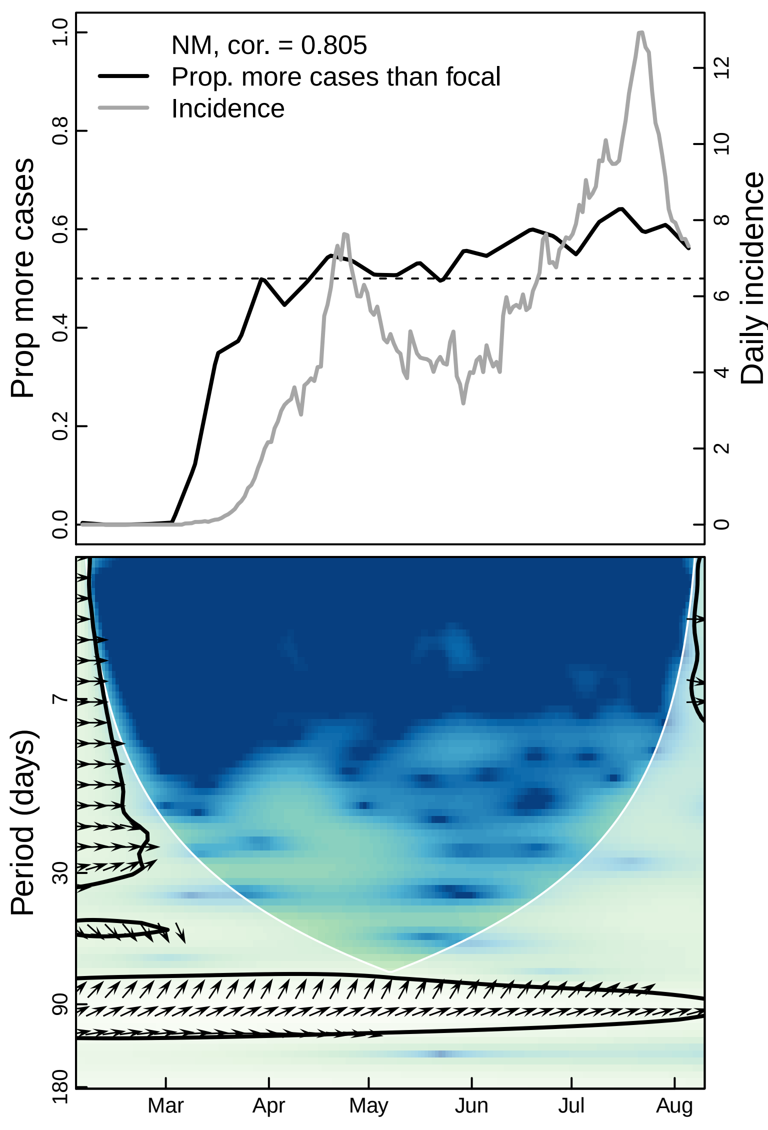

NM, cor. = 0.805
Prop. more cases than focal
Incidence
Prop more cases
Daily incidence
0.0
0.2
0.4
0.6
0.8
1.0
0
2
4
6
8
10
12
Period (days)
7
30
90
180
Mar
Apr
May
Jun
Jul
Aug

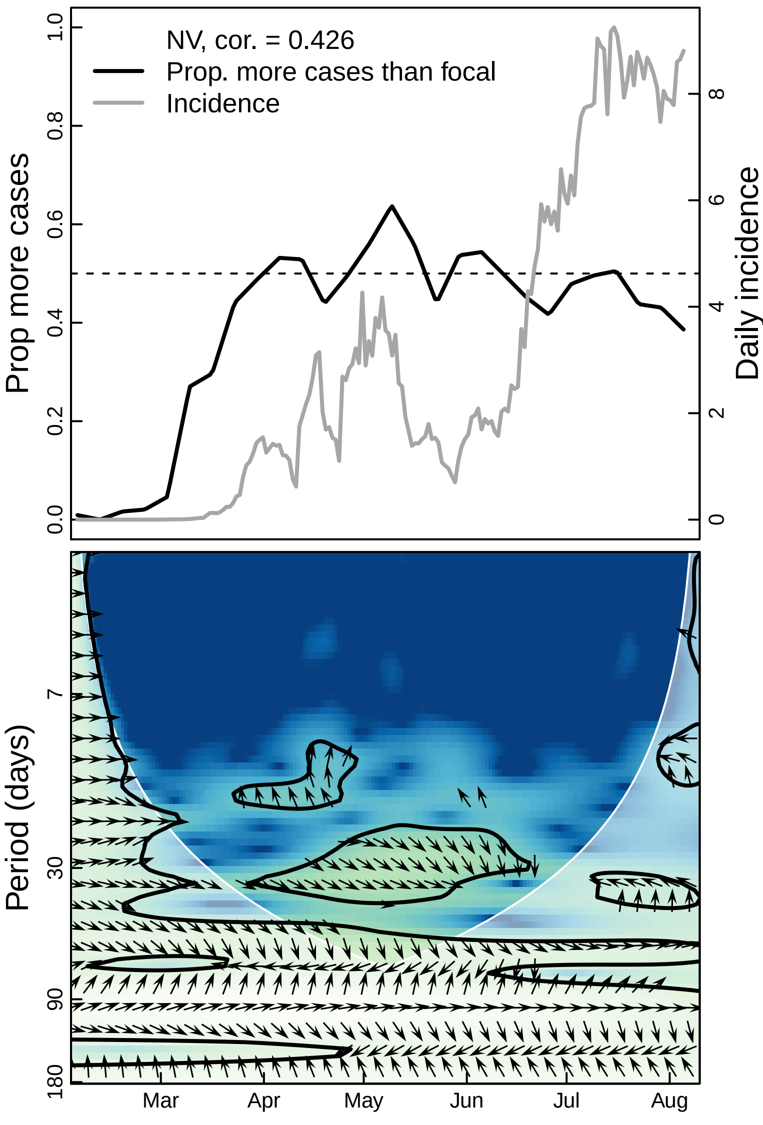

NV, cor. = 0.426
Prop. more cases than focal
Incidence
Prop more cases
Daily incidence
Period (days)
0.0
0.2
0.4
0.6
0.8
1.0
0
2
4
6
8
7
30
90
180
Mar
Apr
May
Jun
Jul
Aug

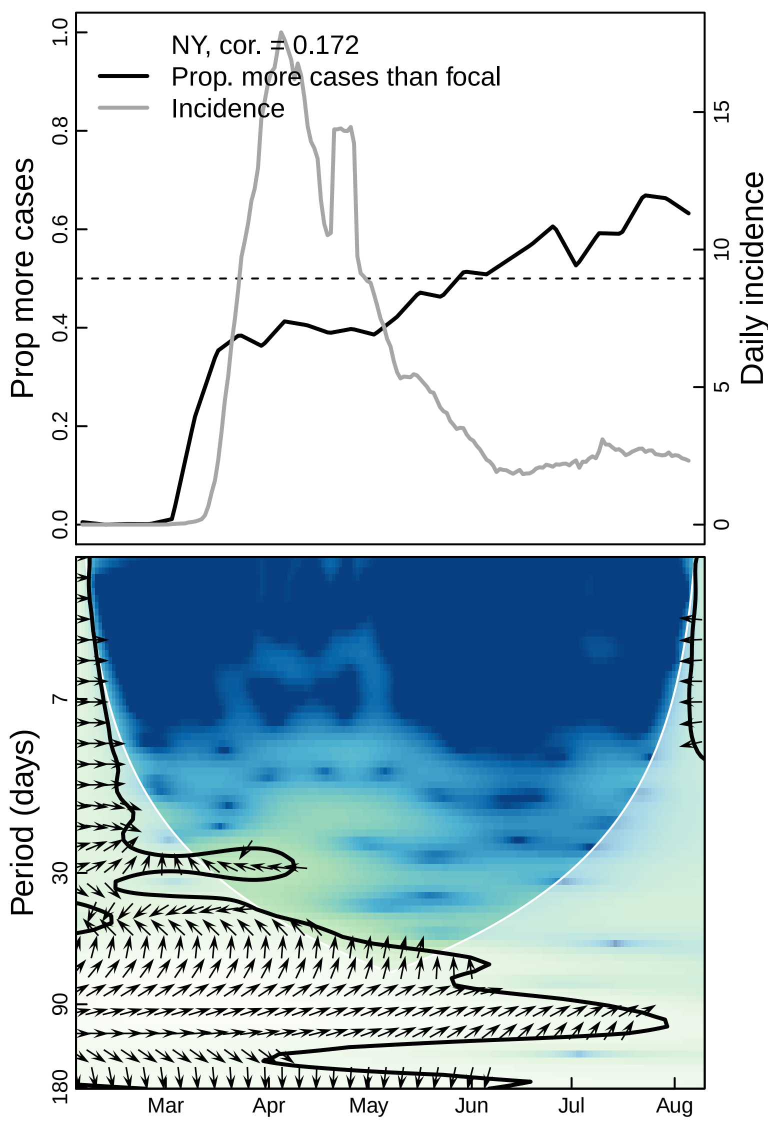

NY, cor. = 0.172
Prop. more cases than focal
Incidence
Prop more cases
Daily incidence
Period (days)
1.0
0.8
0.6
0.4
0.2
0.0
15
10
5
0
7
30
90
180
Mar
Apr
May
Jun
Jul
Aug

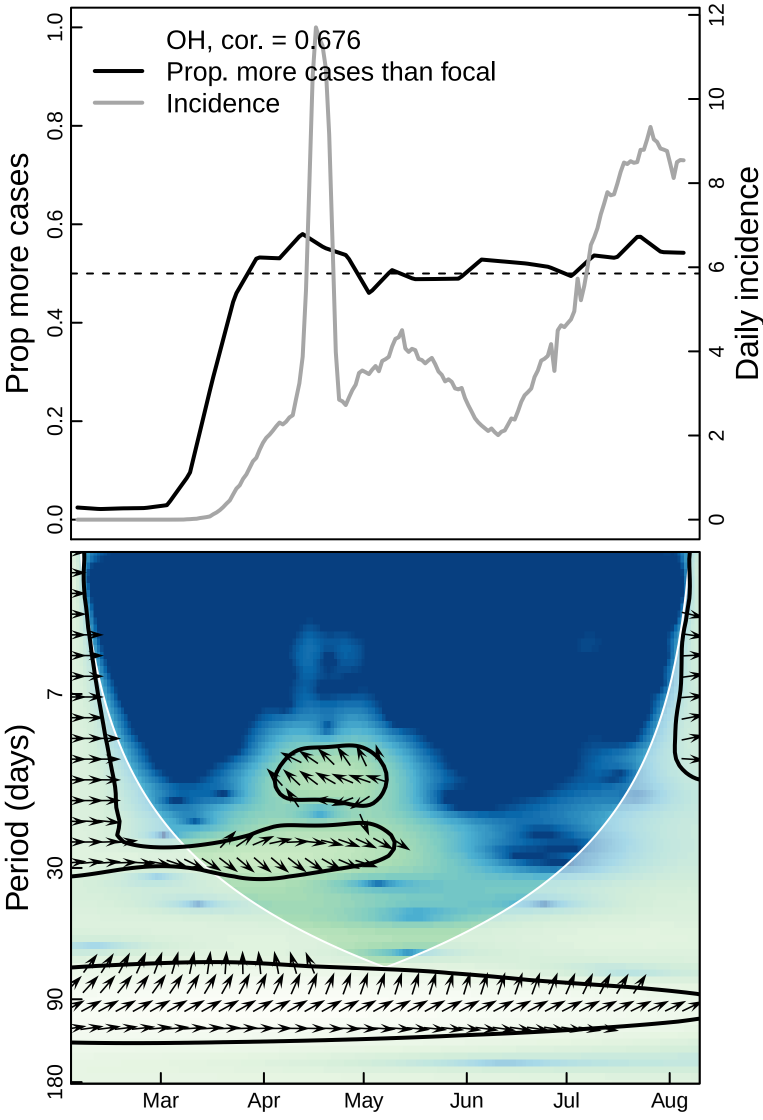

OH, cor. = 0.676
Prop. more cases than focal
Incidence
Prop more cases
Daily incidence
0.0
0.2
0.4
0.6
0.8
1.0
0
2
4
6
8
10
12
Period (days)
7
30
90
180
Mar
Apr
May
Jun
Jul
Aug

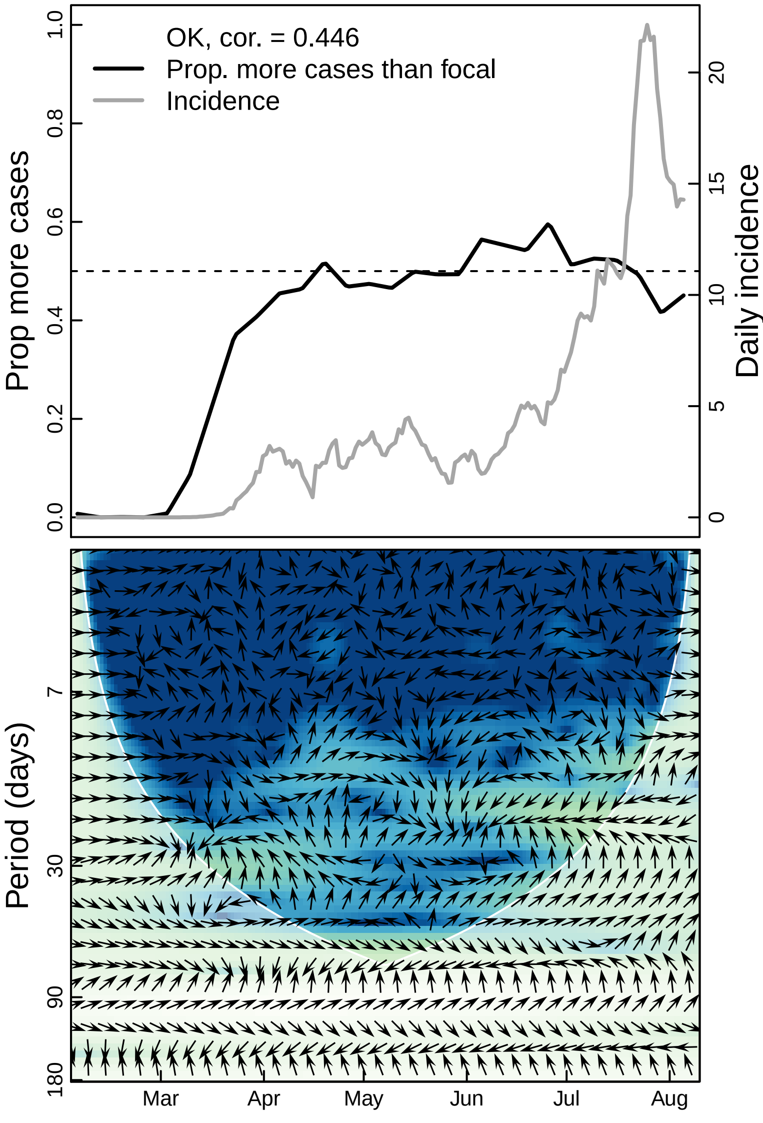
OK, cor. = 0.446
Prop. more cases than focal
Incidence
Prop more cases
Daily incidence
0.0
0.2
0.4
0.6
0.8
1.0
0
5
10
15
20
Period (days)
7
30
90
180
Mar
Apr
May
Jun
Jul
Aug

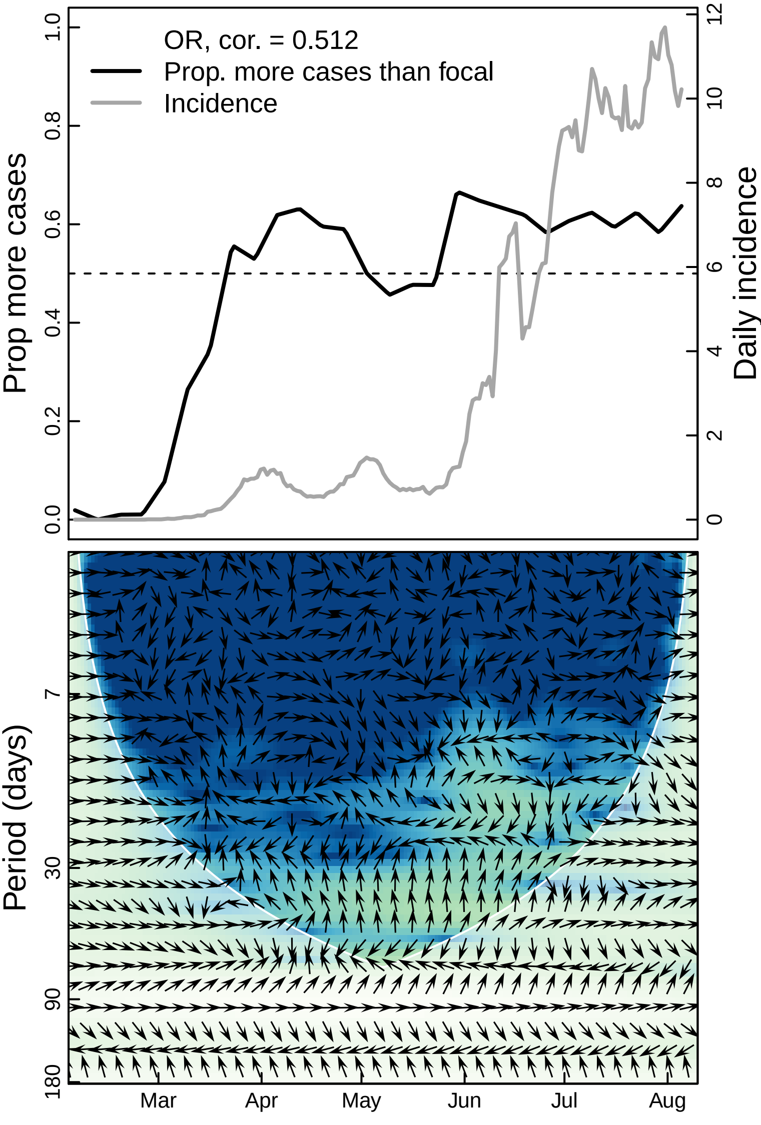
OR, cor. = 0.512
Prop. more cases than focal
Incidence
Prop more cases
Daily incidence
0.0
0.2
0.4
0.6
0.8
1.0
0
2
4
6
8
10
12
Period (days)
7
30
90
180
Mar
Apr
May
Jun
Jul
Aug

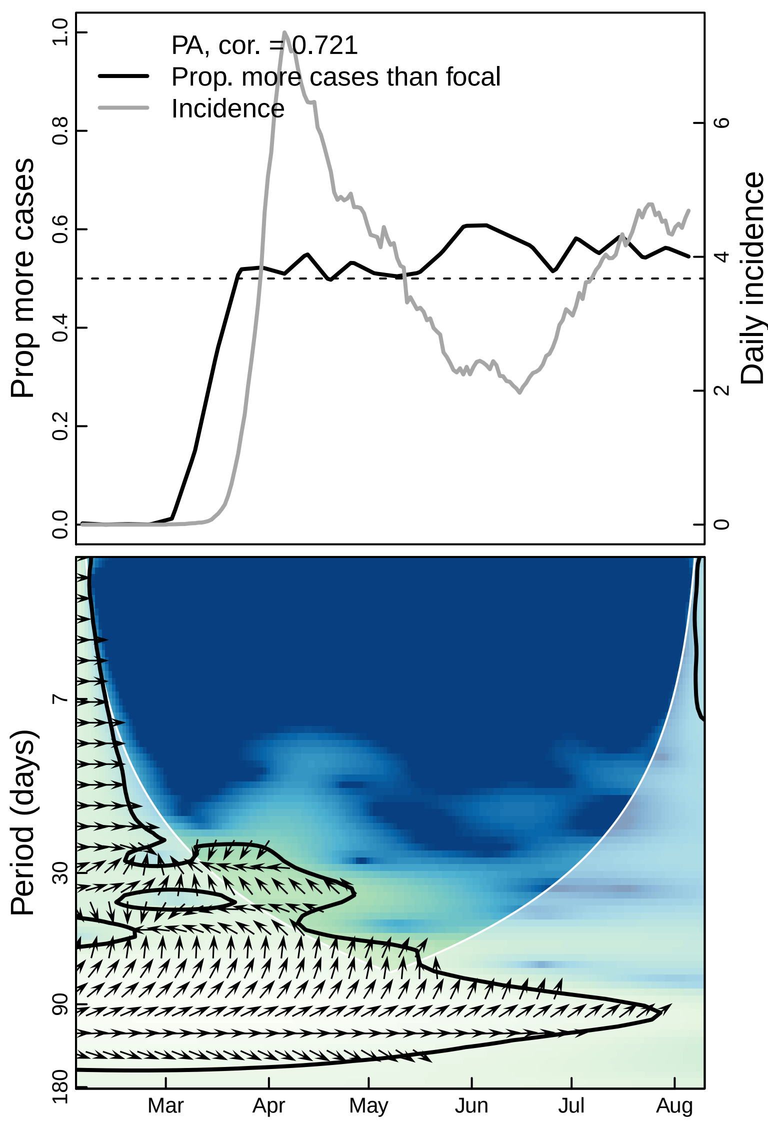

PA, cor. = 0.721
Prop. more cases than focal
Incidence
Prop more cases
Daily incidence
0.0
0.2
0.4
0.6
0.8
1.0
0
2
4
6
Period (days)
7
30
90
180
Mar
Apr
May
Jun
Jul
Aug

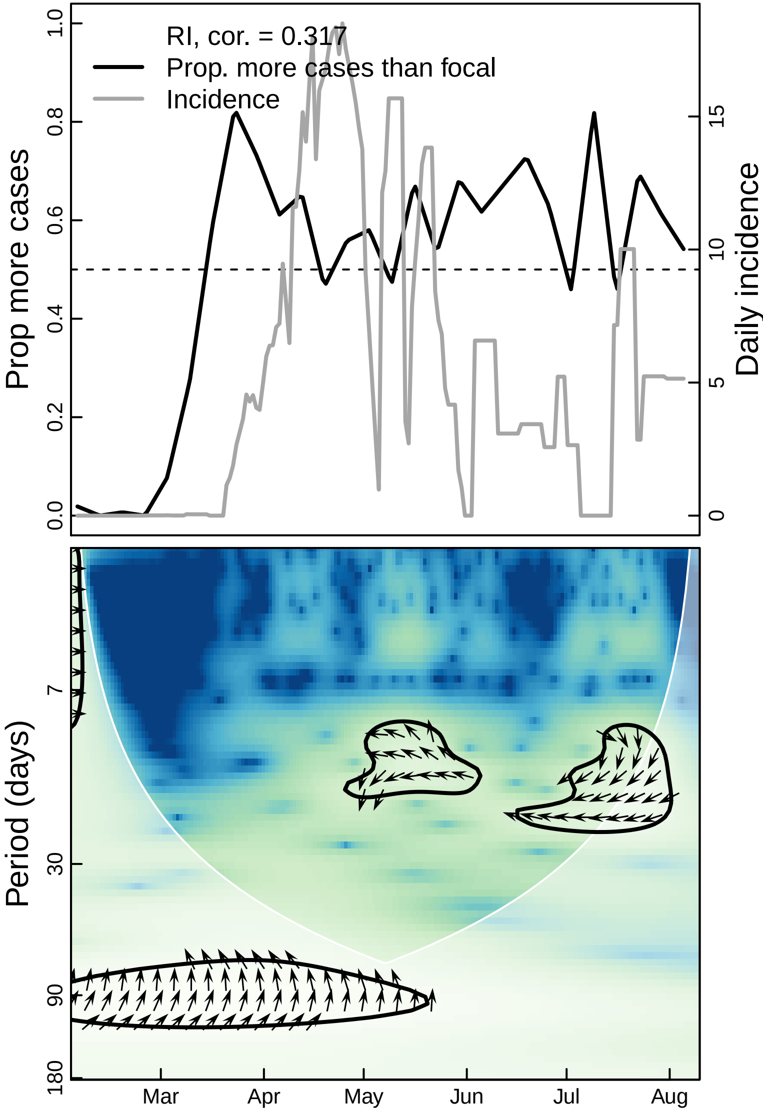

RI, cor. = 0.317
Prop. more cases than focal
Incidence
Prop more cases
Daily incidence
0.0
0.2
0.4
0.6
0.8
1.0
0
5
10
15
Period (days)
7
30
90
180
Mar
Apr
May
Jun
Jul
Aug

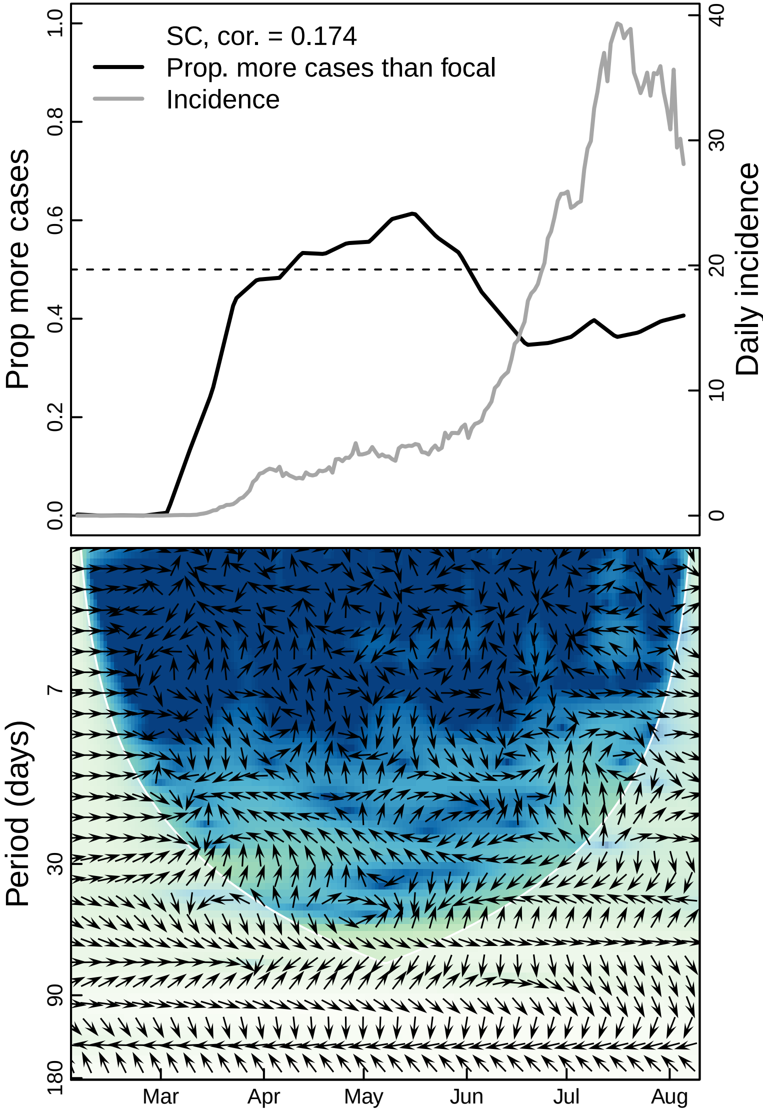

SC, cor. = 0.174
Prop. more cases than focal
Incidence
Prop more cases
Daily incidence
0.0
0.2
0.4
0.6
0.8
1.0
0
10
20
30
40
Period (days)
7
30
90
180
Mar
Apr
May
Jun
Jul
Aug

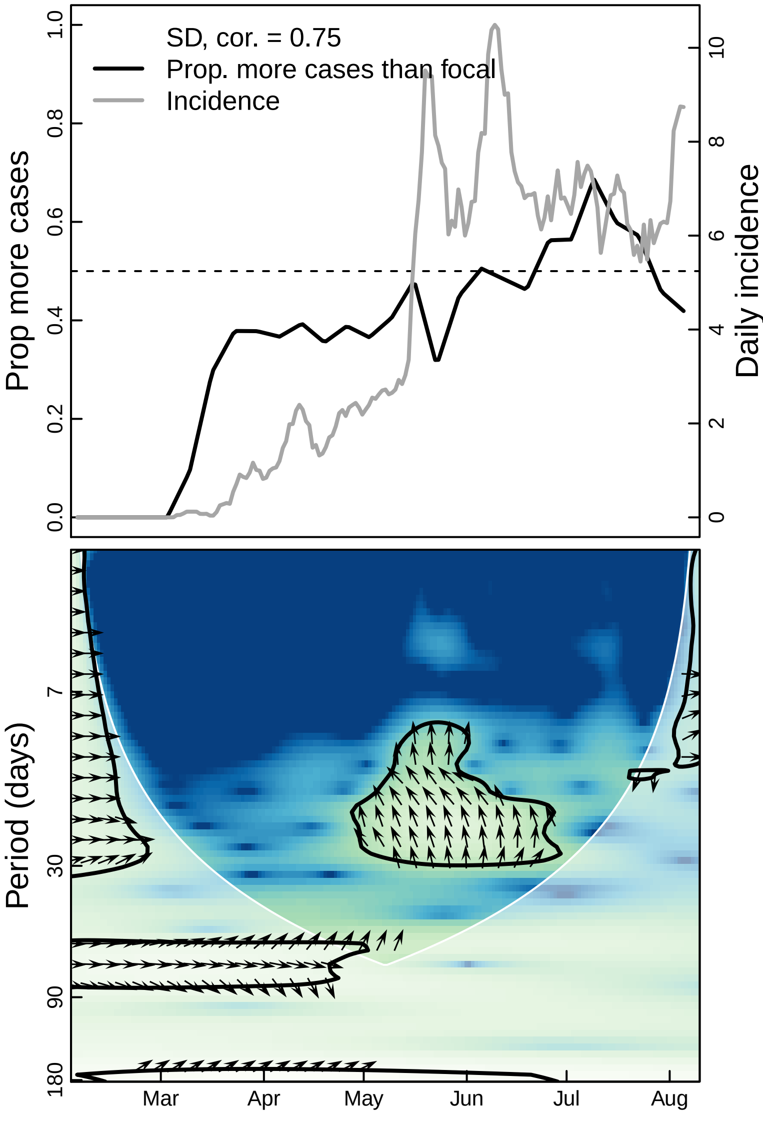
SD, cor. = 0.75
Prop. more cases than focal
Incidence
Prop more cases
Daily incidence
0.0
0.2
0.4
0.6
0.8
1.0
0
2
4
6
8
10
Period (days)
7
30
90
180
Mar
Apr
May
Jun
Jul
Aug

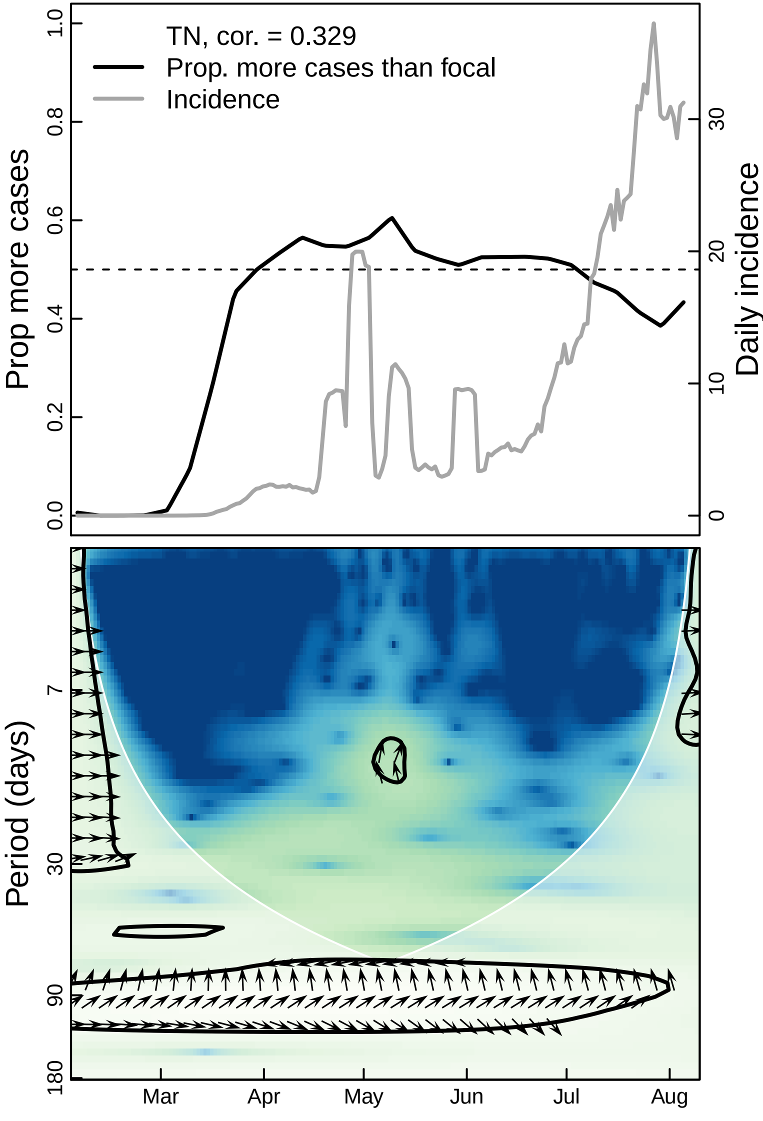

TN, cor. = 0.329
Prop. more cases than focal
Incidence
Prop more cases
Daily incidence
0.0
0.2
0.4
0.6
0.8
1.0
0
10
20
30
Period (days)
7
30
90
180
Mar
Apr
May
Jun
Jul
Aug

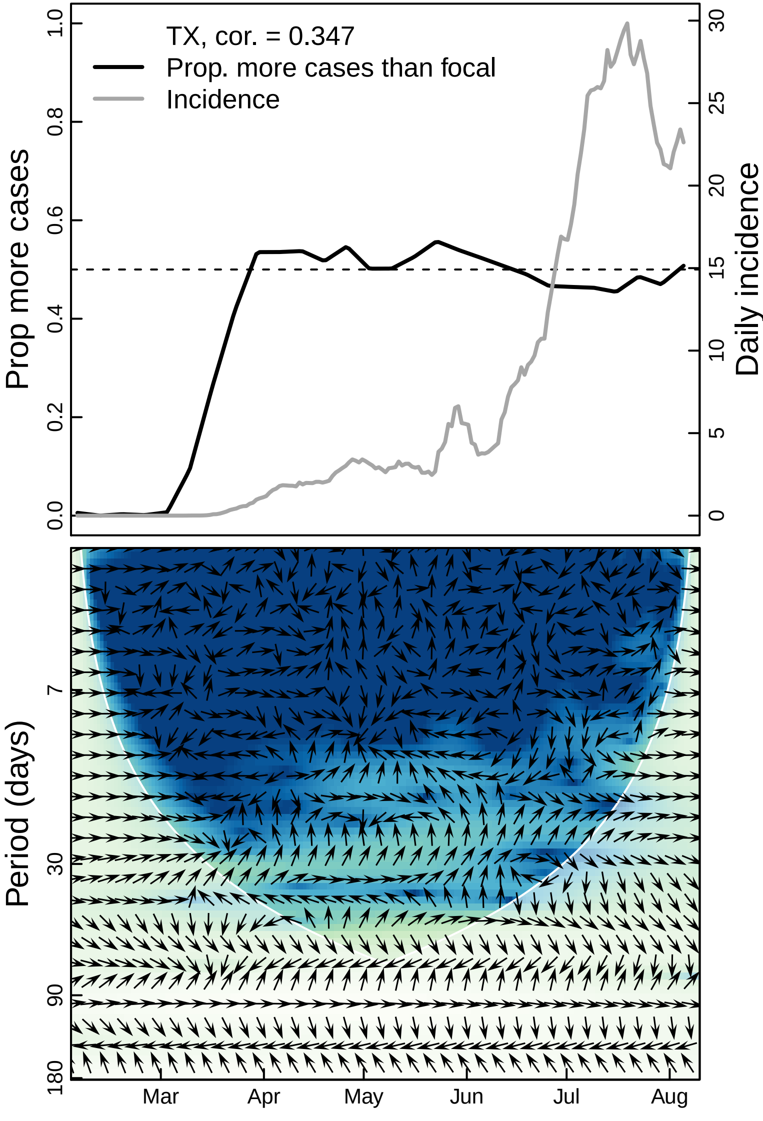

TX, cor. = 0.347
Prop. more cases than focal
Incidence
Prop more cases
Daily incidence
0.0
0.2
0.4
0.6
0.8
1.0
0
5
10
15
20
25
30
Period (days)
7
30
90
180
Mar
Apr
May
Jun
Jul
Aug

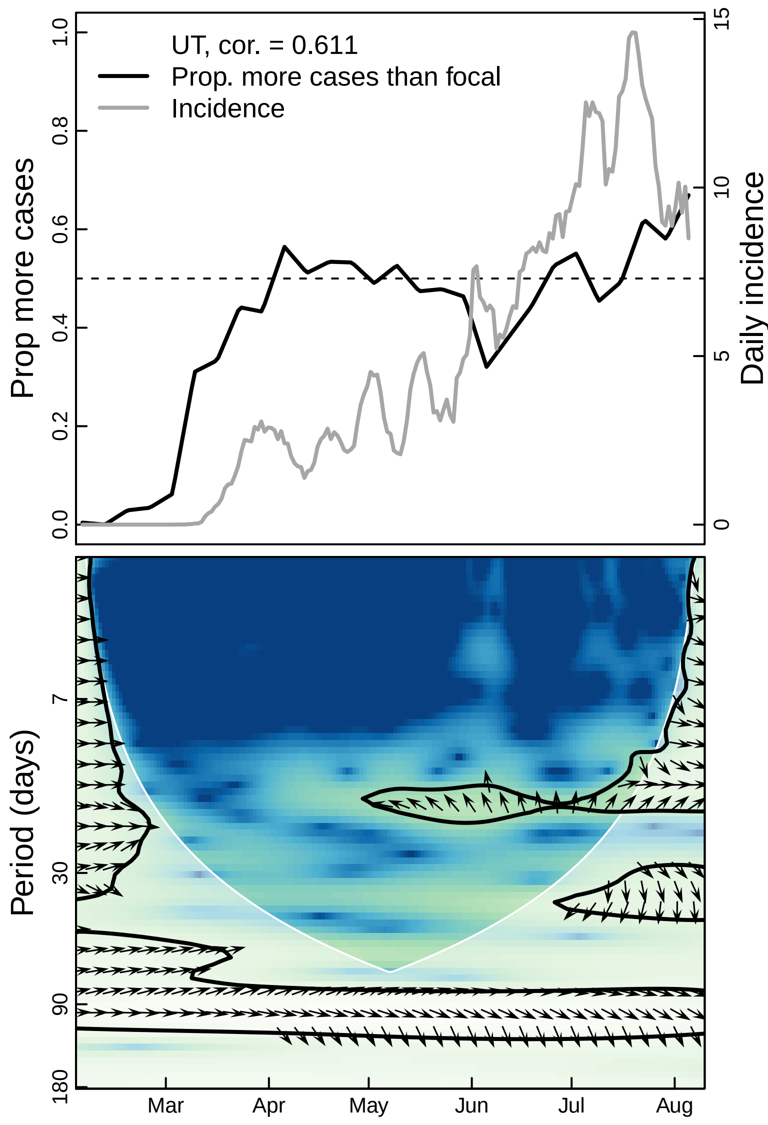

UT, cor. = 0.611
Prop. more cases than focal
Incidence
Prop more cases
Daily incidence
0.0
0.2
0.4
0.6
0.8
1.0
0
5
10
15
Period (days)
7
30
90
180
Mar
Apr
May
Jun
Jul
Aug

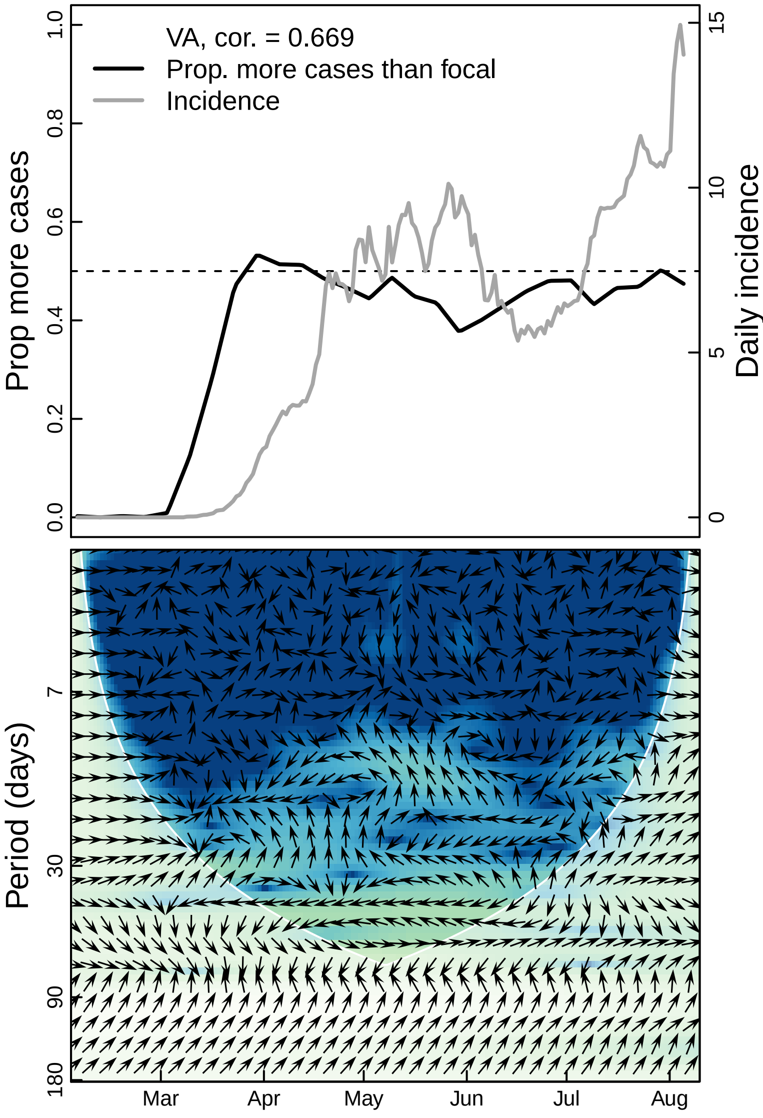

VA, cor. = 0.669
Prop. more cases than focal
Incidence
Prop more cases
Daily incidence
1.0
0.8
0.6
0.4
0.2
0.0
15
10
5
0
Period (days)
7
30
90
180
Mar
Apr
May
Jun
Jul
Aug

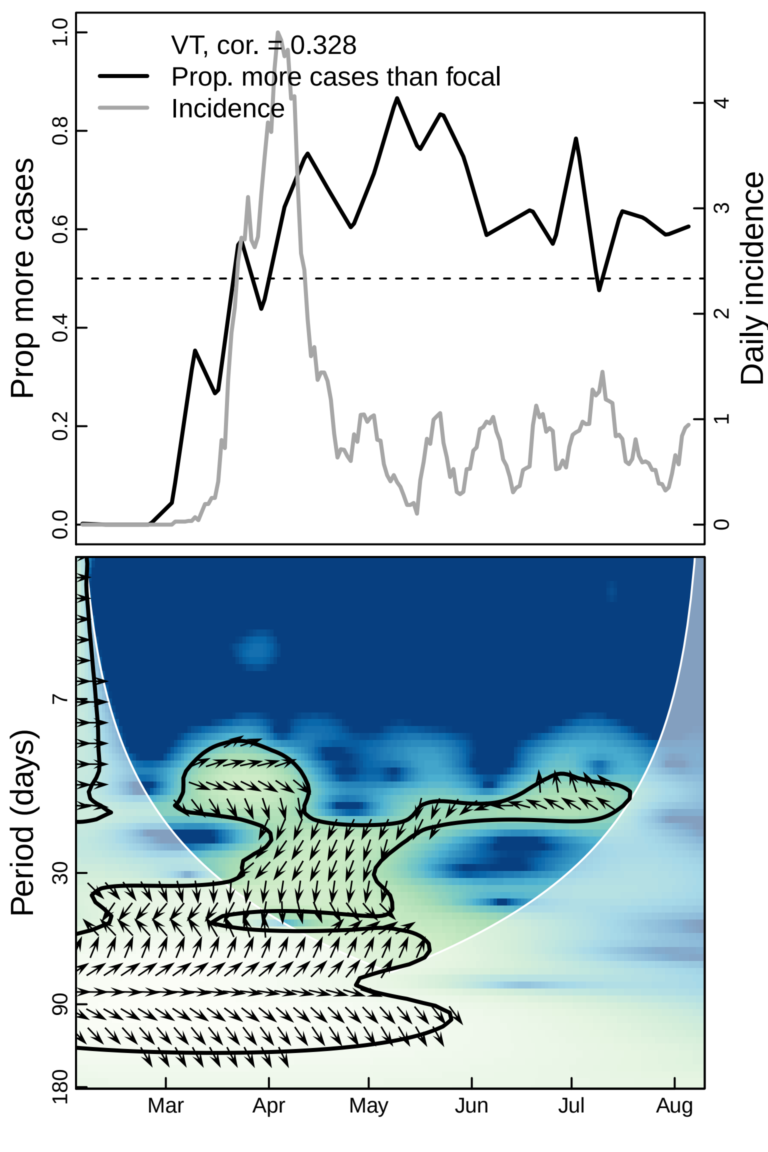
VT, cor. = 0.328
Prop. more cases than focal
Incidence
Prop more cases
Daily incidence
0.0
0.2
0.4
0.6
0.8
1.0
0
1
2
3
4
Period (days)
7
30
90
180
Mar
Apr
May
Jun
Jul
Aug

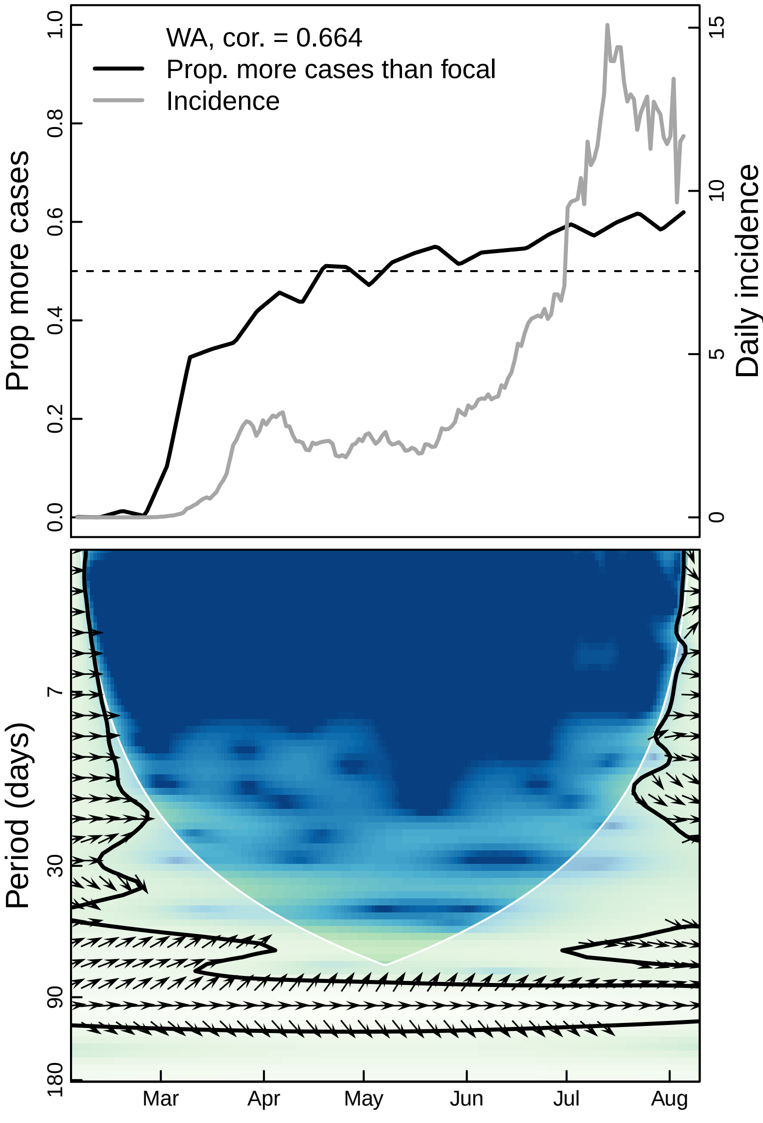

WA, cor. = 0.664
Prop. more cases than focal
Incidence
Prop more cases
Daily incidence
0.0
0.2
0.4
0.6
0.8
1.0
0
5
10
15
Period (days)
7
30
90
180
Mar
Apr
May
Jun
Jul
Aug

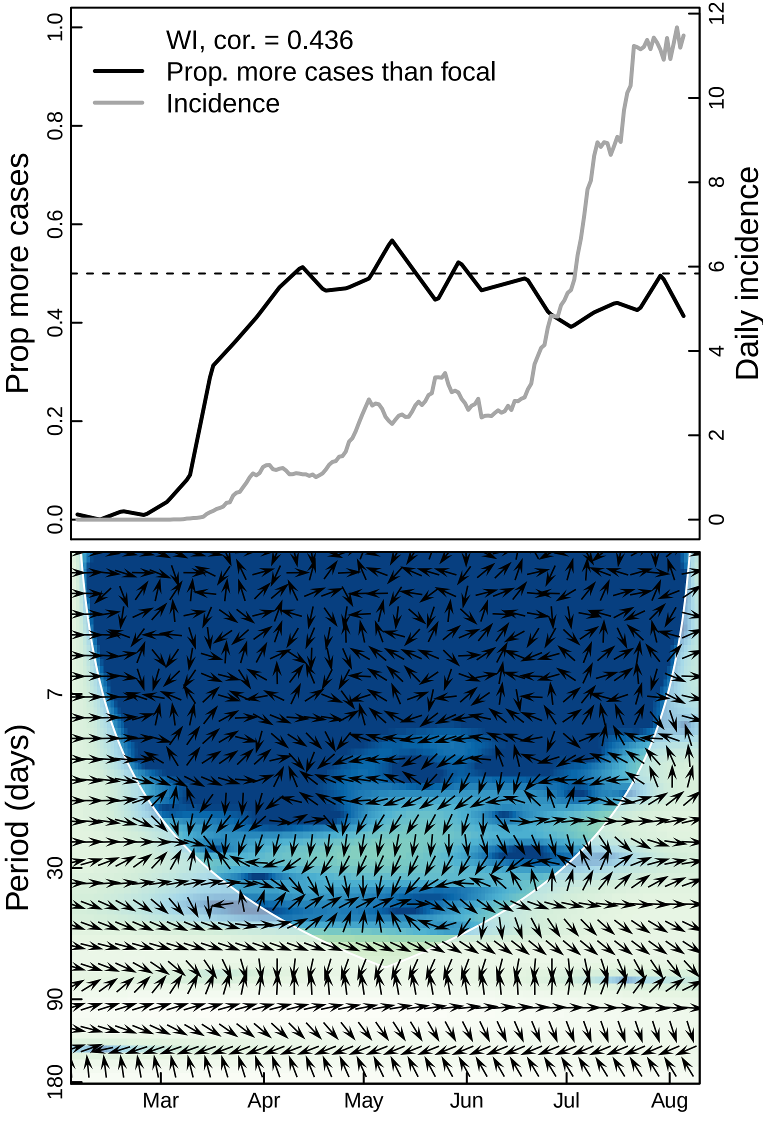
WI, cor. = 0.436
Prop. more cases than focal
Incidence
Prop more cases
Daily incidence
0.0
0.2
0.4
0.6
0.8
1.0
0
2
4
6
8
10
12
Period (days)
7
30
90
180
Mar
Apr
May
Jun
Jul
Aug

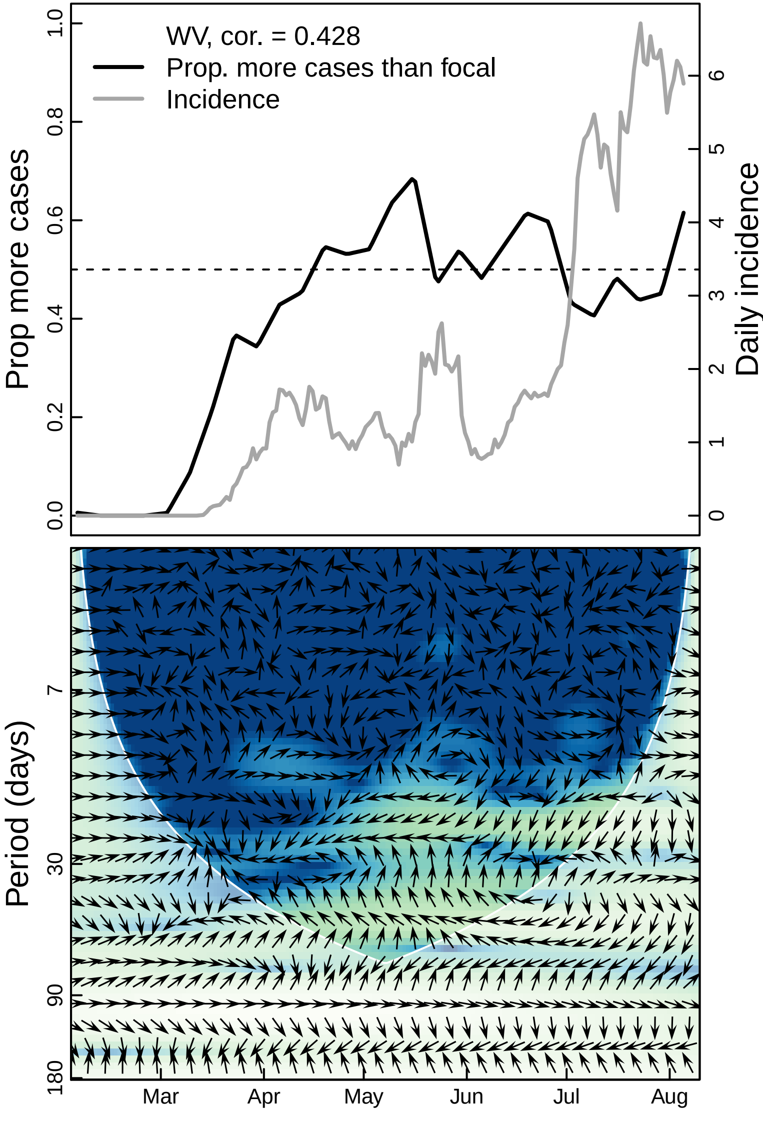
WV, cor. = 0.428
Prop. more cases than focal
Incidence
Prop more cases
Daily incidence
0.0
0.2
0.4
0.6
0.8
1.0
0
1
2
3
4
5
6
Period (days)
7
30
90
180
Mar
Apr
May
Jun
Jul
Aug

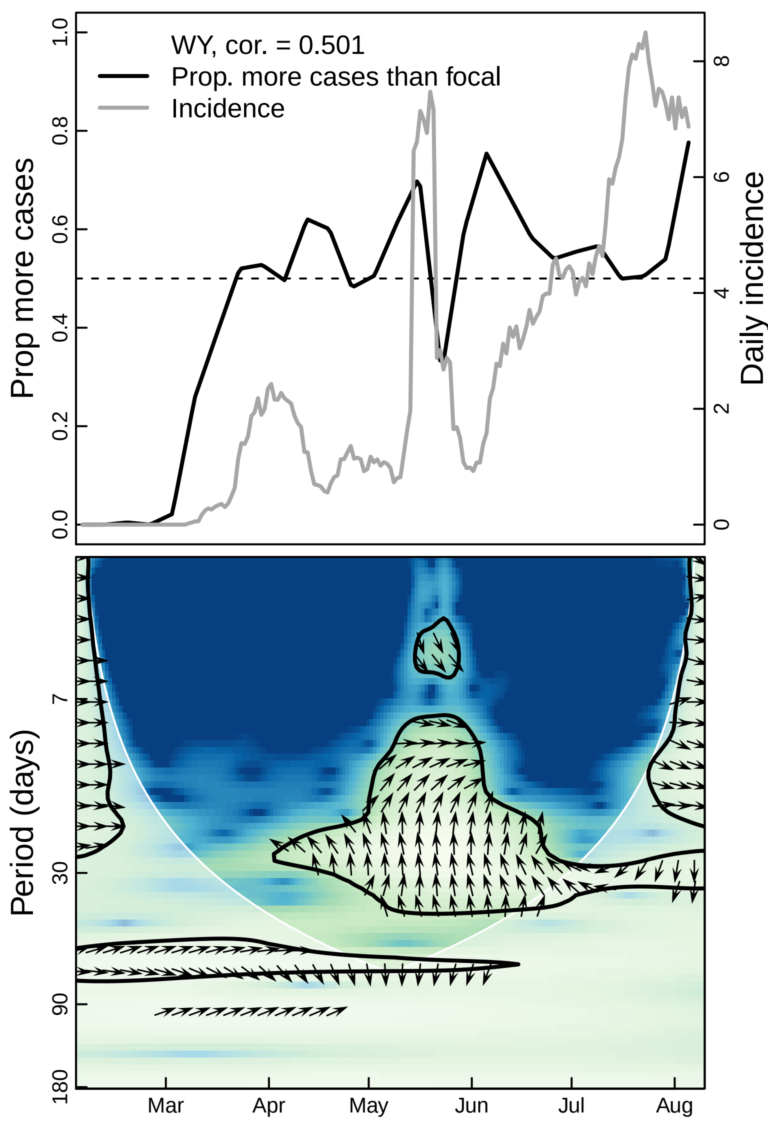

WY, cor. = 0.501
Prop. more cases than focal
Incidence
Prop more cases
Daily incidence
0.0
0.2
0.4
0.6
0.8
1.0
0
2
4
6
8
Period (days)
7
30
90
180
Mar
Apr
May
Jun
Jul
Aug
