## Supplementary material for "The unintended consequences of inconsistent pandemic control policies": Cross-wavelets - bars

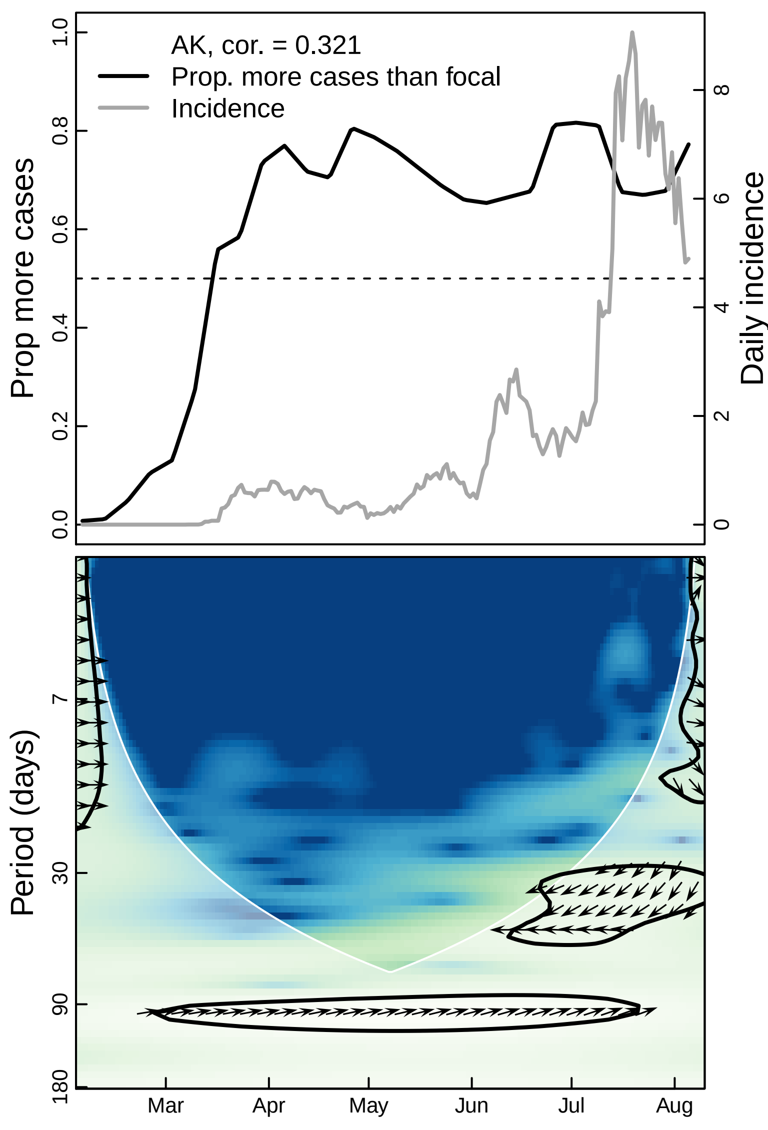
AK, cor. = 0.321
Prop. more cases than focal
Incidence
Prop more cases
Daily incidence
1.0
0.8
0.6
0.4
0.2
0.0
8
6
4
2
0
Period (days)
7
30
90
180
Mar
Apr
May
Jun
Jul
Aug

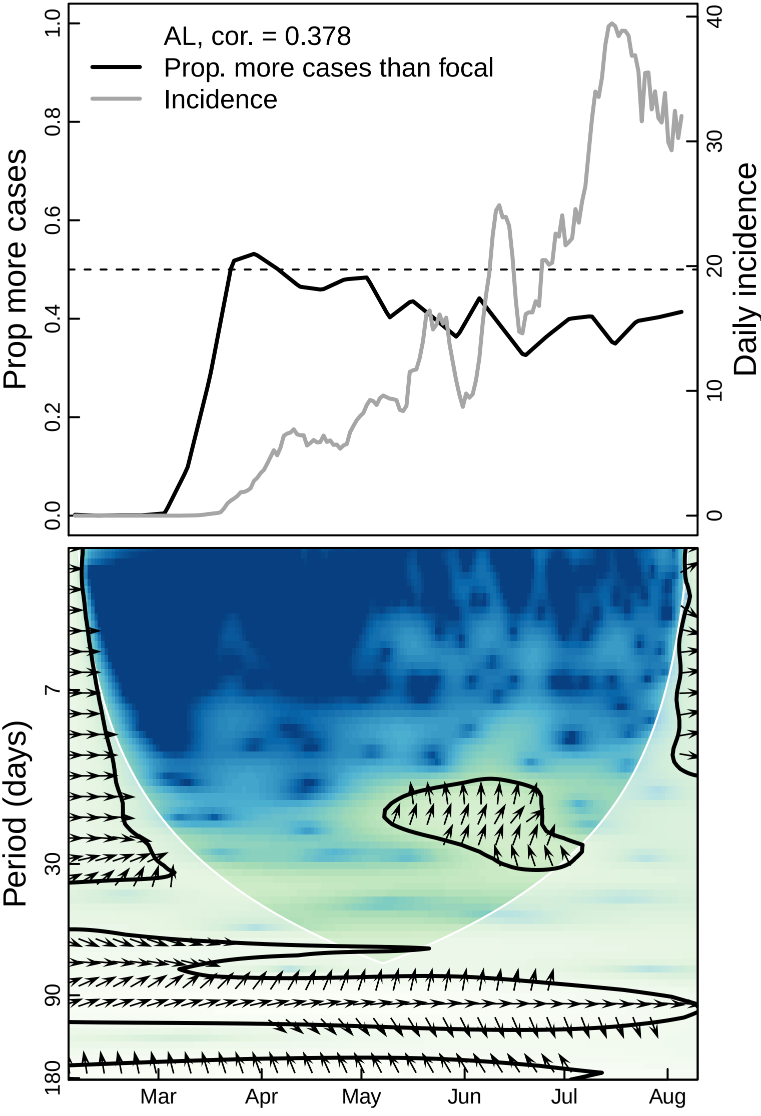
AL, cor. = 0.378
Prop. more cases than focal
Incidence
Prop more cases
Daily incidence
0.0
0.2
0.4
0.6
0.8
1.0
0
10
20
30
40
Period (days)
7
30
90
180
Mar
Apr
May
Jun
Jul
Aug

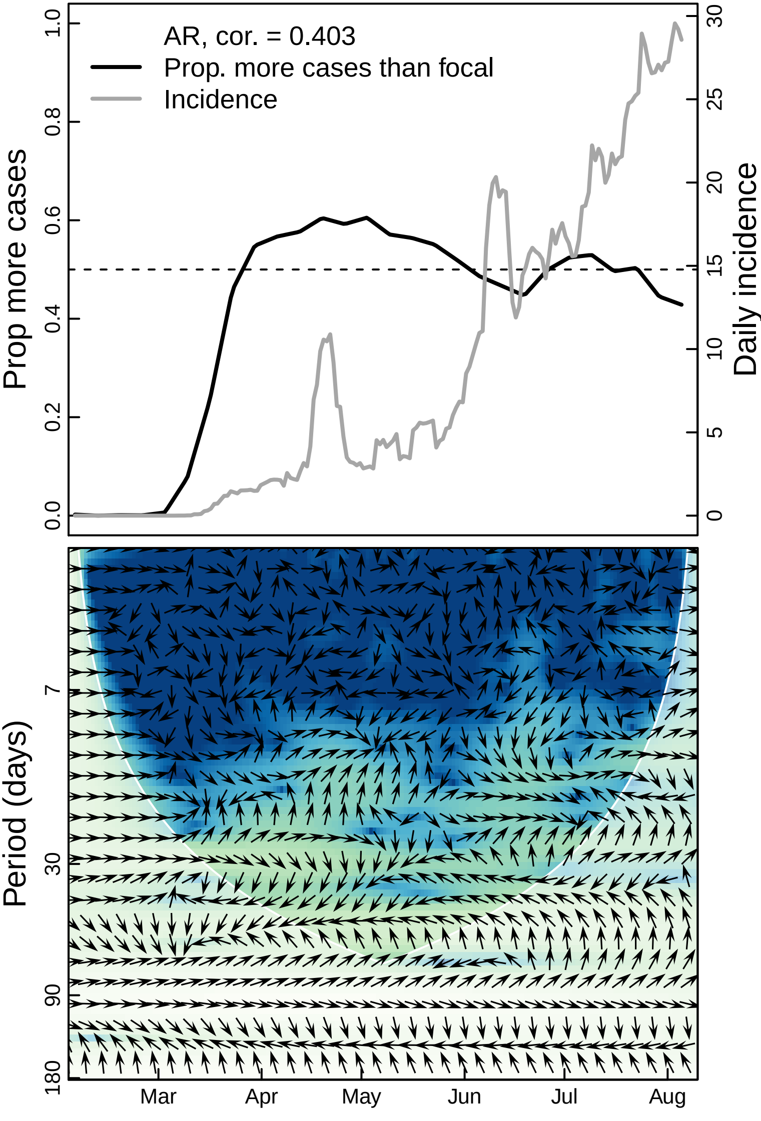

AR, cor. = 0.403
Prop. more cases than focal
Incidence
Prop more cases
Daily incidence
0.0
0.2
0.4
0.6
0.8
1.0
0
5
10
15
20
25
30
Period (days)
7
30
90
180
Mar
Apr
May
Jun
Jul
Aug

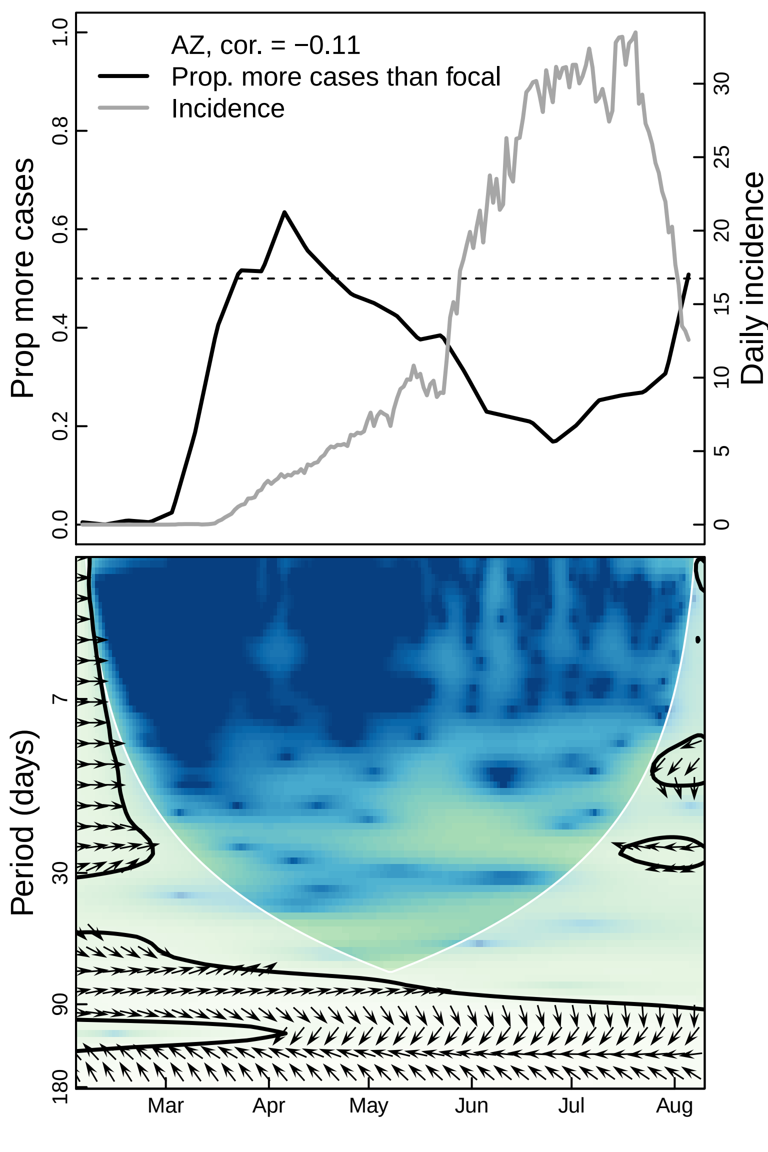

AZ, cor. = −0.11
Prop. more cases than focal
Incidence
Prop more cases
Daily incidence
1.0
0.8
0.6
0.4
0.2
0.0
30
25
20
15
10
5
0
Period (days)
7
30
90
180
Mar
Apr
May
Jun
Jul
Aug

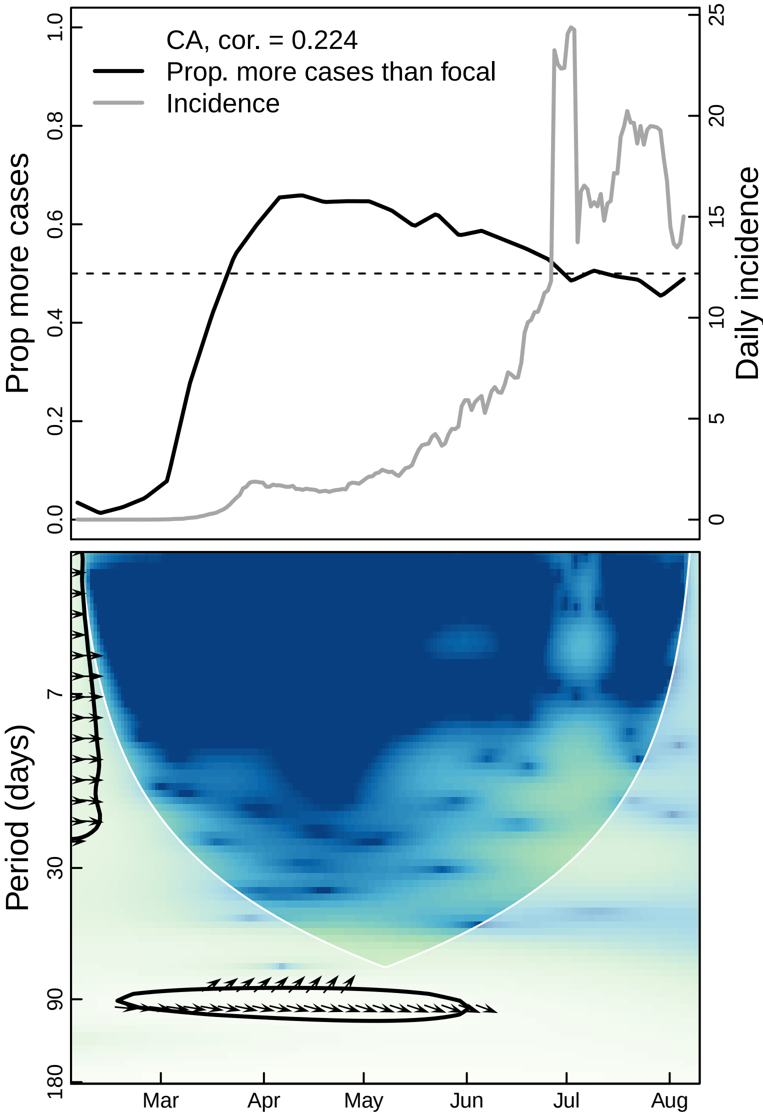

CA, cor. = 0.224
Prop. more cases than focal
Incidence
Prop more cases
Daily incidence
0.0
0.2
0.4
0.6
0.8
1.0
0
5
10
15
20
25
Period (days)
7
30
90
180
Mar
Apr
May
Jun
Jul
Aug

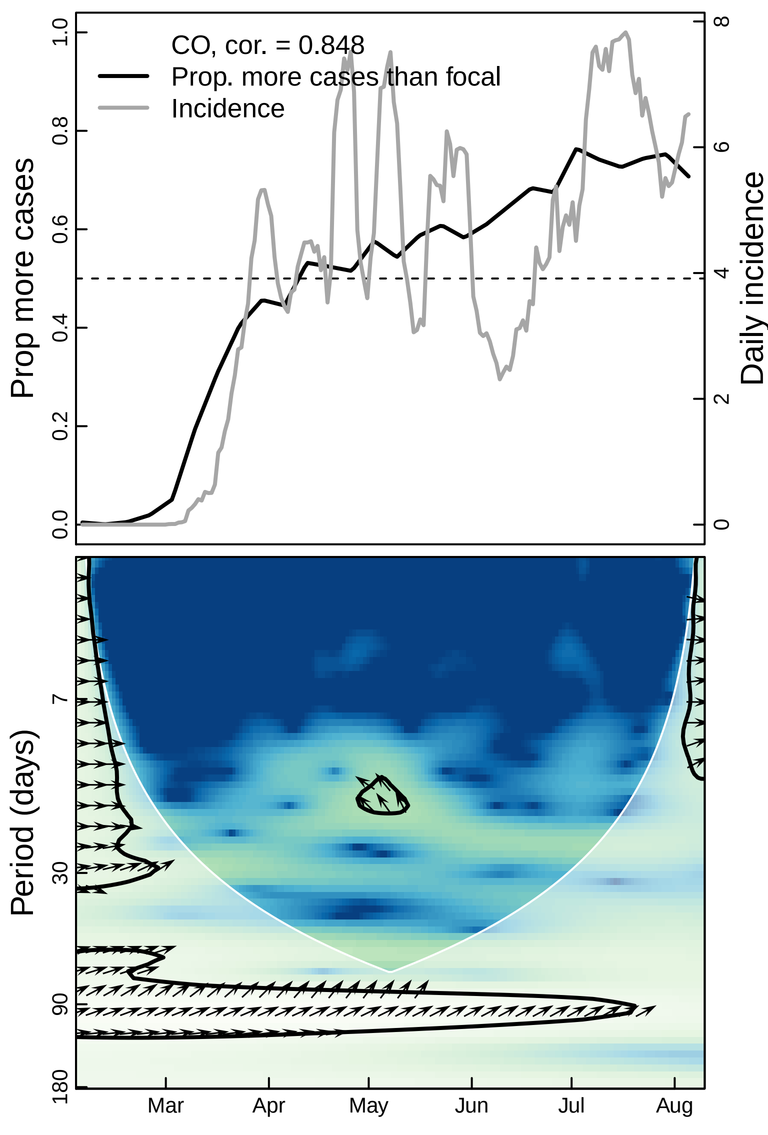

CO, cor. = 0.848
Prop. more cases than focal
Incidence
Prop more cases
Daily incidence
0.0
0.2
0.4
0.6
0.8
1.0
0
2
4
6
8
Period (days)
7
30
90
180
Mar
Apr
May
Jun
Jul
Aug

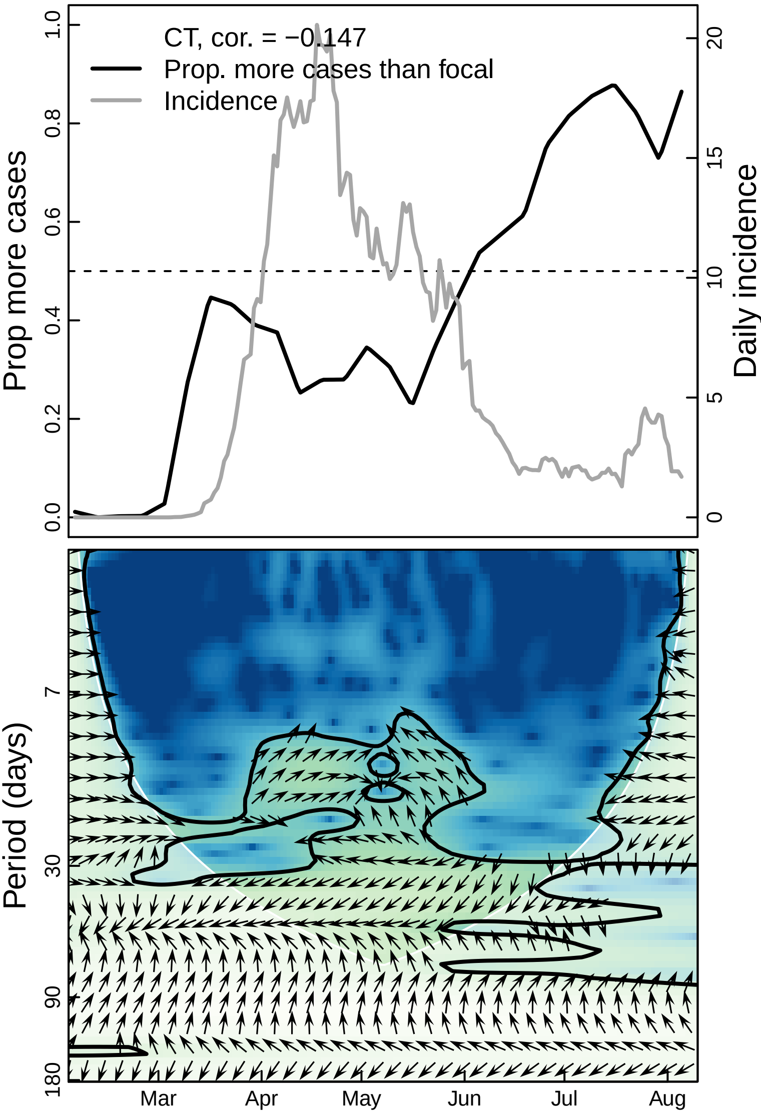

CT, cor. = −0.147
Prop. more cases than focal
Incidence
Prop more cases
Daily incidence
0.0
0.2
0.4
0.6
0.8
1.0
0
5
10
15
20
Period (days)
7
30
90
180
Mar
Apr
May
Jun
Jul
Aug

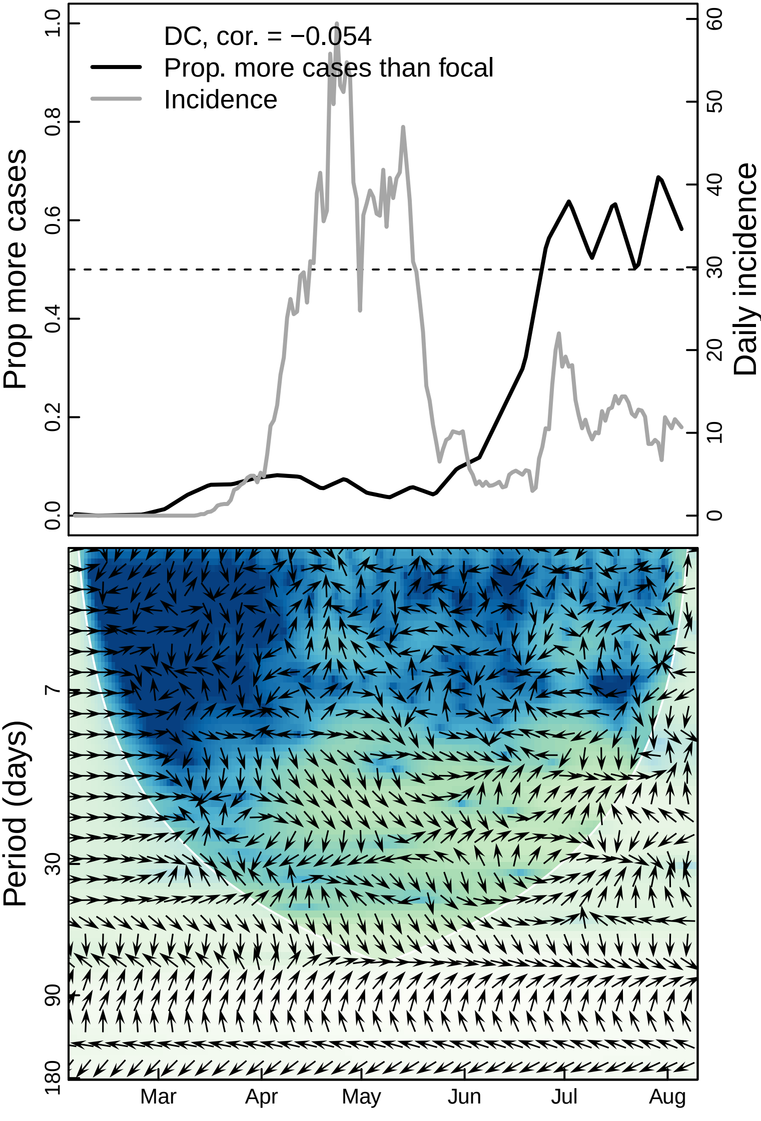
DC, cor. = −0.054
Prop. more cases than focal
Incidence
Prop more cases
Daily incidence
0.0
0.2
0.4
0.6
0.8
1.0
0
10
20
30
40
50
60
Period (days)
7
30
90
180
Mar
Apr
May
Jun
Jul
Aug

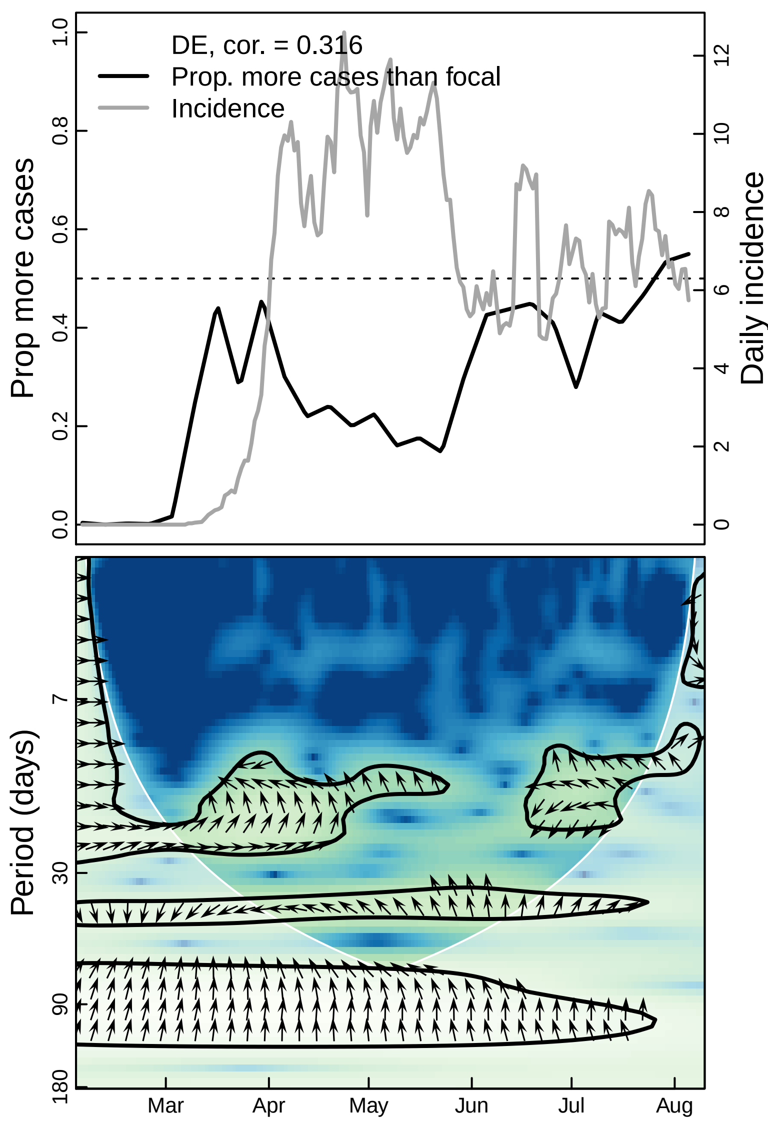

DE, cor. = 0.316
Prop. more cases than focal
Incidence
Prop more cases
Daily incidence
0.0
0.2
0.4
0.6
0.8
1.0
0
2
4
6
8
10
12
Period (days)
7
30
90
180
Mar
Apr
May
Jun
Jul
Aug

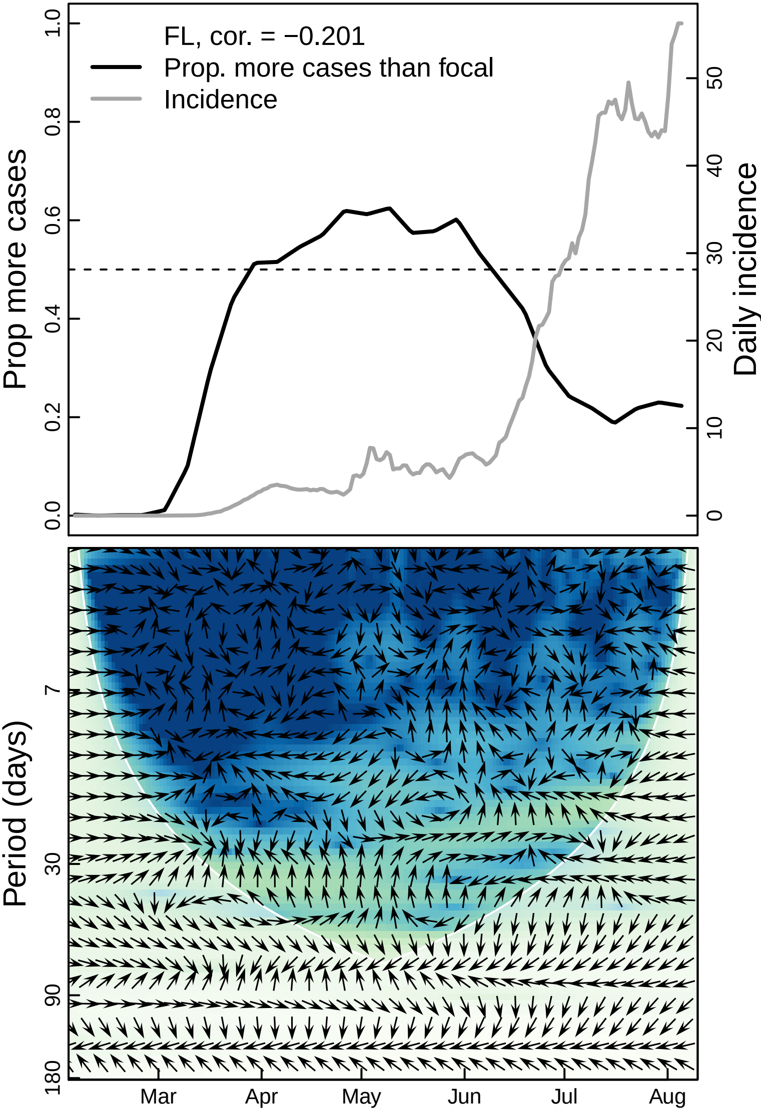
FL, cor. = −0.201
Prop. more cases than focal
Incidence
Prop more cases
Daily incidence
0.0
0.2
0.4
0.6
0.8
1.0
0
10
20
30
40
50
Period (days)
7
30
90
180
Mar
Apr
May
Jun
Jul
Aug

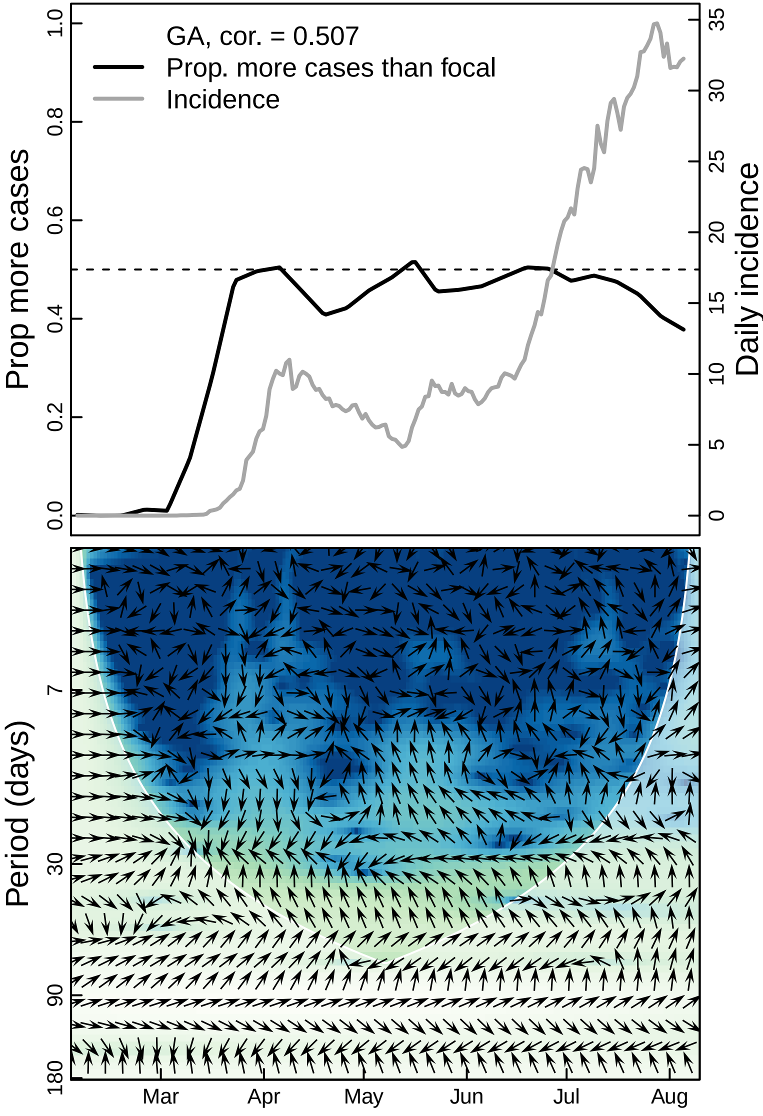

GA, cor. = 0.507
Prop. more cases than focal
Incidence
Prop more cases
Daily incidence
Period (days)
Mar
Apr
May
Jun
Jul
Aug

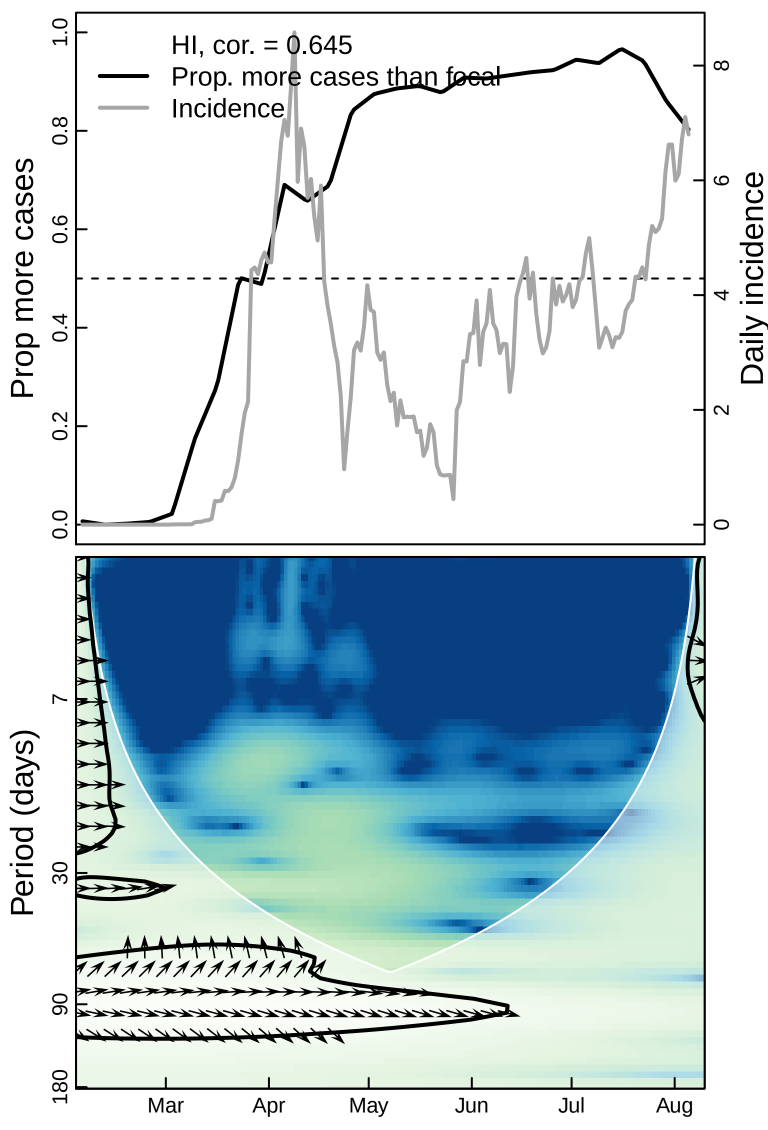

HI, cor. = 0.645
Prop. more cases than focal
Incidence
Prop more cases
Daily incidence
0.0
0.2
0.4
0.6
0.8
1.0
0
2
4
6
8
Period (days)
7
30
90
180
Mar
Apr
May
Jun
Jul
Aug

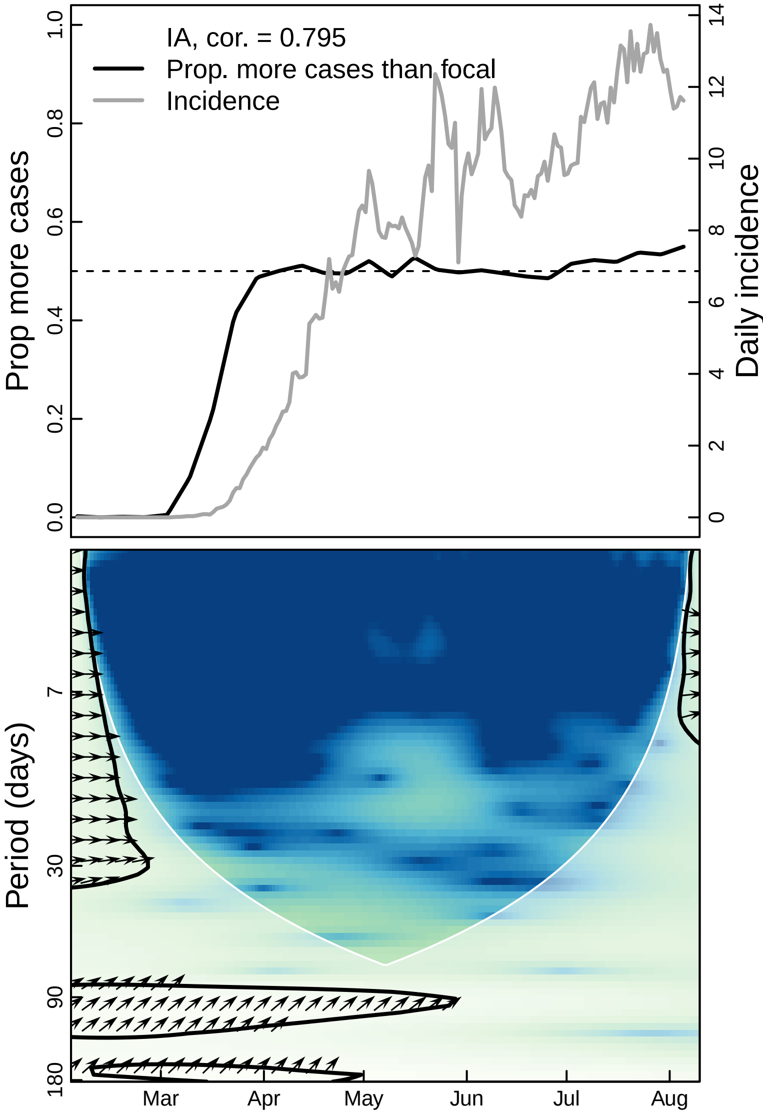

IA, cor. = 0.795
Prop. more cases than focal
Incidence
Prop more cases
Daily incidence
0.0
0.2
0.4
0.6
0.8
1.0
0
2
4
6
8
10
12
14
Period (days)
7
30
90
180
Mar
Apr
May
Jun
Jul
Aug

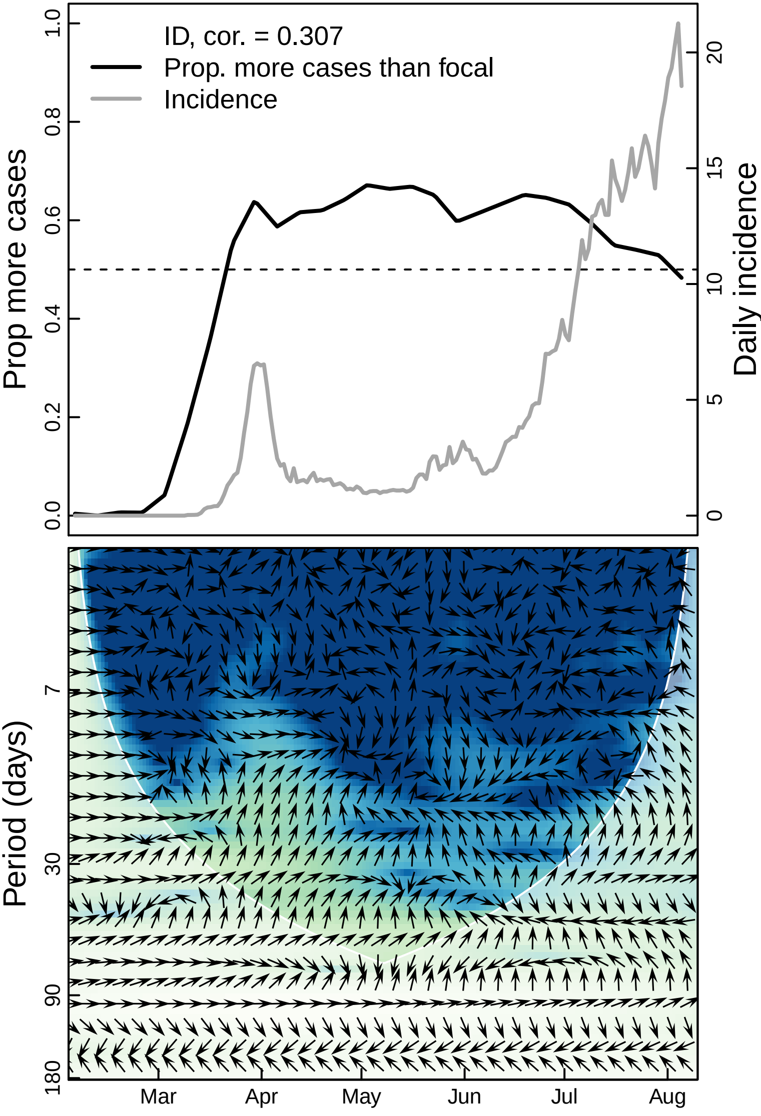

ID, cor. = 0.307
Prop. more cases than focal
Incidence
Prop more cases
Daily incidence
0.0
0.2
0.4
0.6
0.8
1.0
0
5
10
15
20
Period (days)
7
30
90
180
Mar
Apr
May
Jun
Jul
Aug

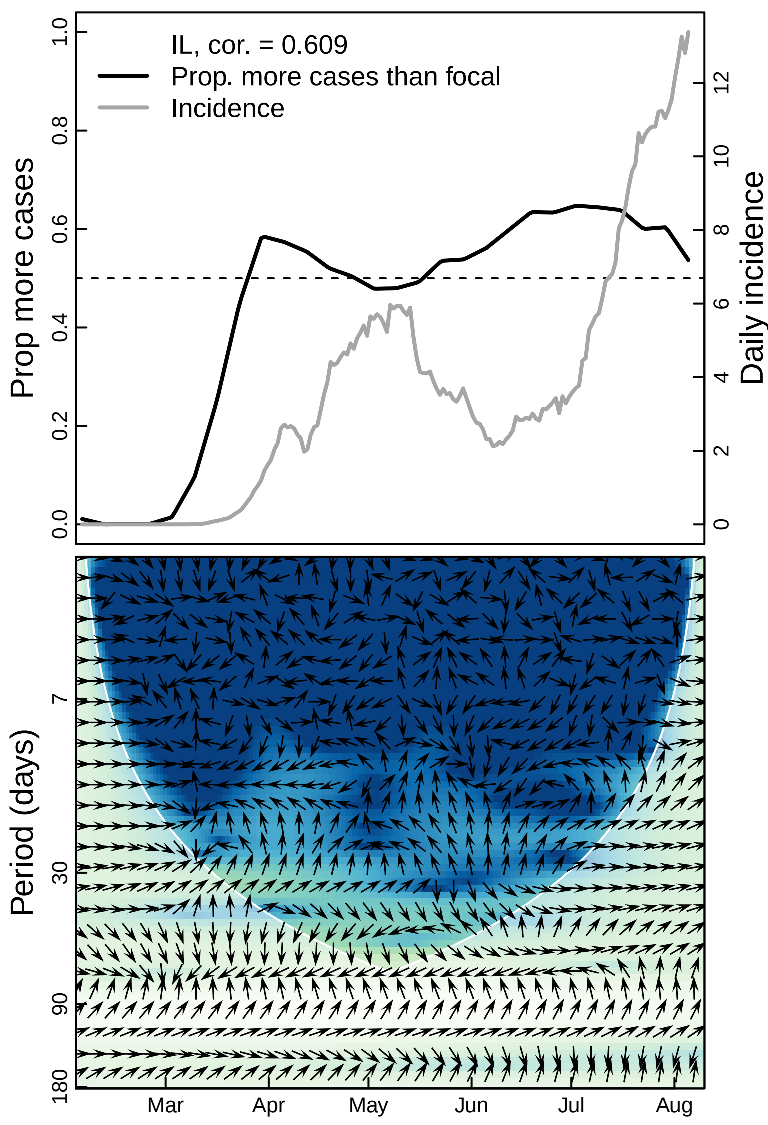

IL, cor. = 0.609
Prop. more cases than focal
Incidence
Prop more cases
Daily incidence
0.0
0.2
0.4
0.6
0.8
1.0
0
2
4
6
8
10
12
Period (days)
7
30
90
180
Mar
Apr
May
Jun
Jul
Aug

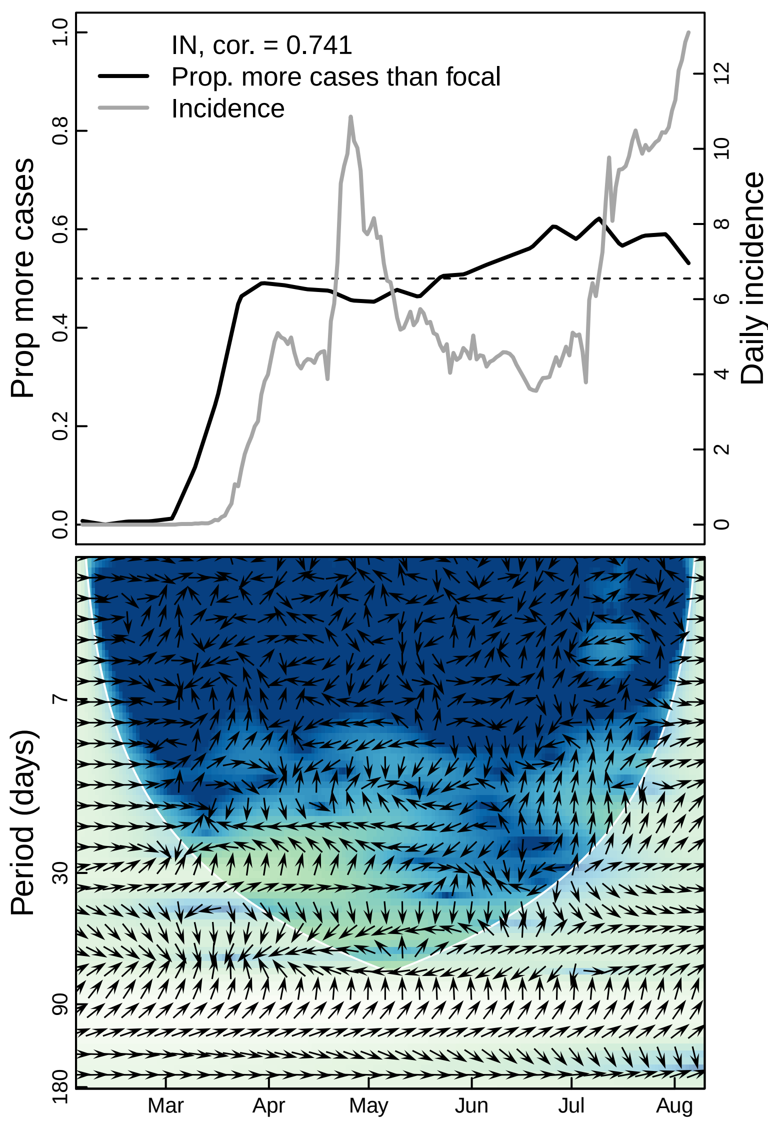

IN, cor. = 0.741
Prop. more cases than focal
Incidence
Prop more cases
Daily incidence
0.0
0.2
0.4
0.6
0.8
1.0
0
2
4
6
8
10
12
Period (days)
7
30
90
180
Mar
Apr
May
Jun
Jul
Aug

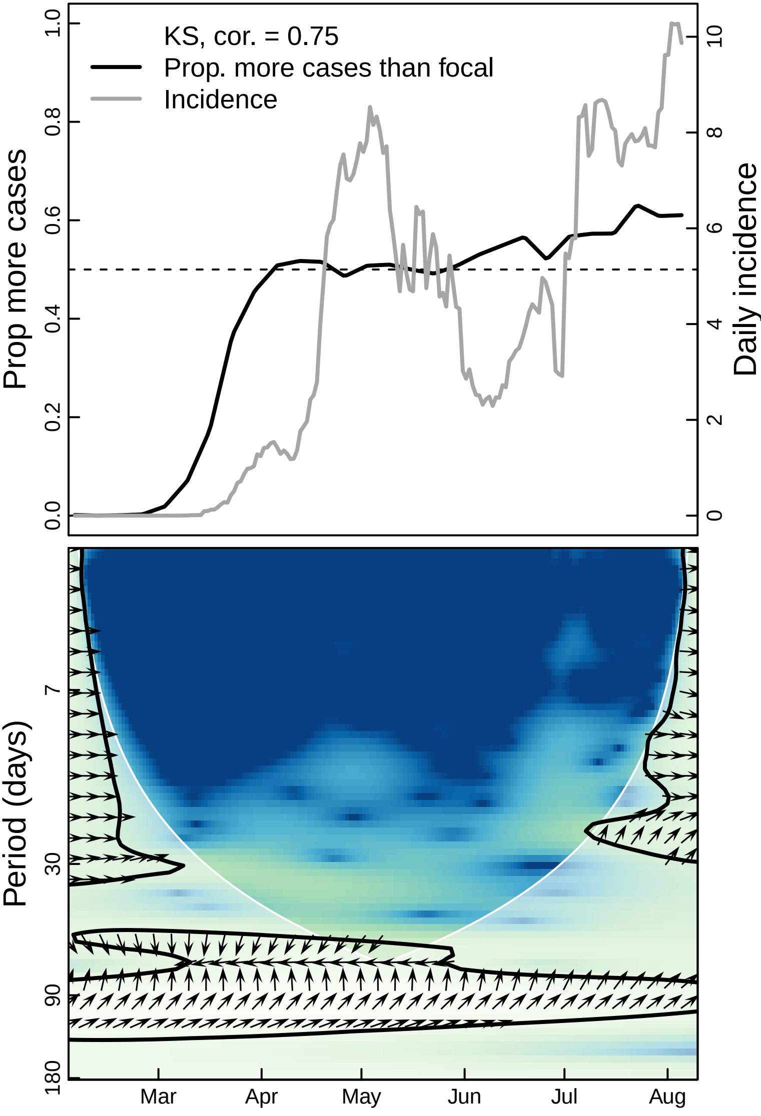
KS, cor. = 0.75
Prop. more cases than focal
Incidence
Prop more cases
Daily incidence
0.0
0.2
0.4
0.6
0.8
1.0
0
2
4
6
8
10
Period (days)
7
30
90
180
Mar
Apr
May
Jun
Jul
Aug

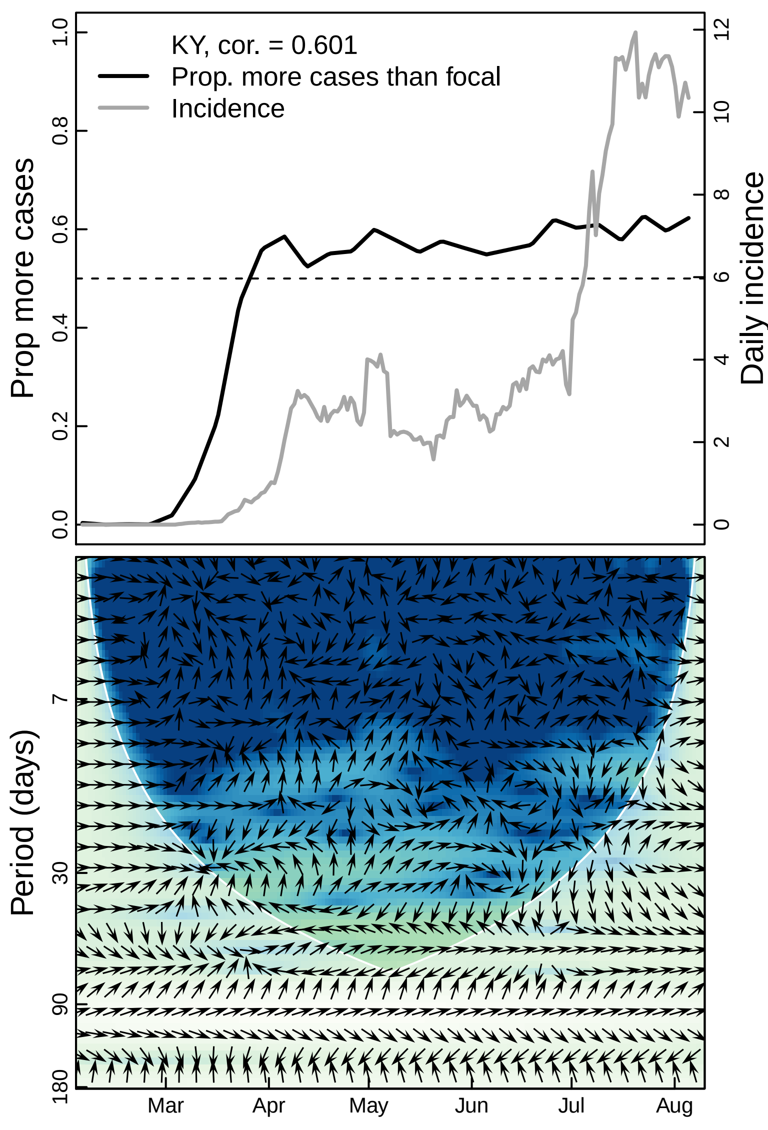

KY, cor. = 0.601
Prop. more cases than focal
Incidence
Prop more cases
Daily incidence
0.0
0.2
0.4
0.6
0.8
1.0
0
2
4
6
8
10
12
Period (days)
7
30
90
180
Mar
Apr
May
Jun
Jul
Aug

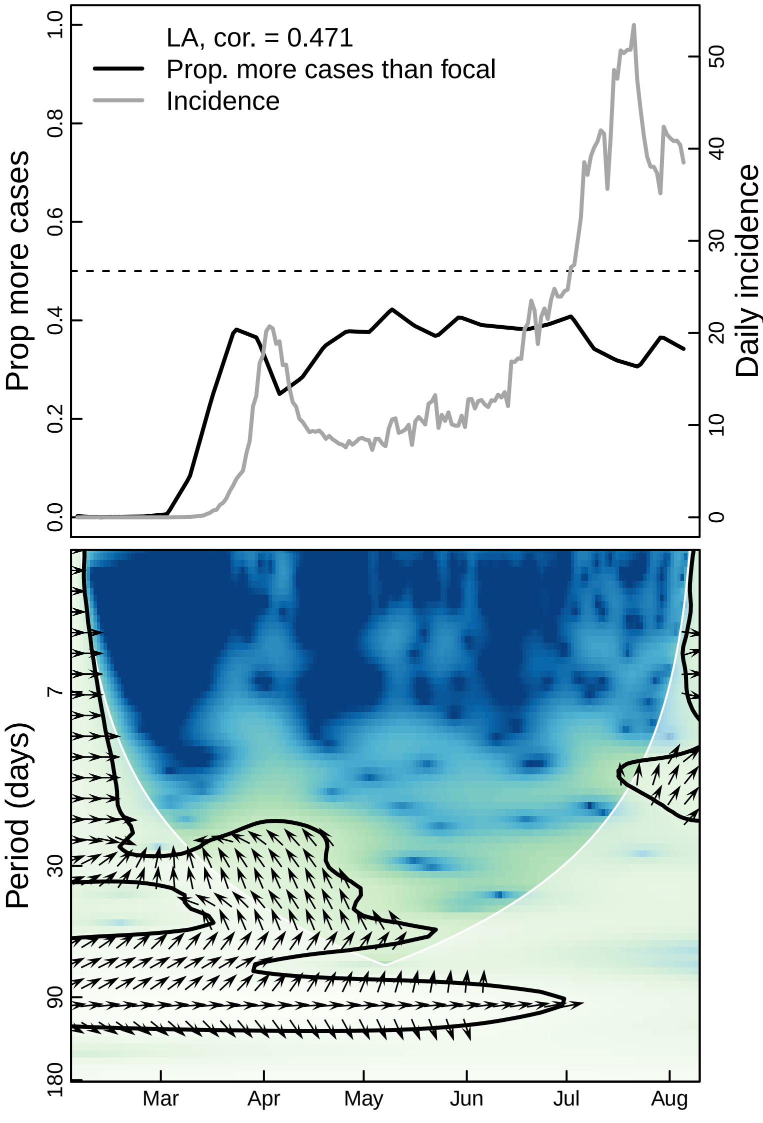

LA, cor. = 0.471
Prop. more cases than focal
Incidence
Prop more cases
Daily incidence
0.0
0.2
0.4
0.6
0.8
1.0
0
10
20
30
40
50
Period (days)
7
30
90
180
Mar
Apr
May
Jun
Jul
Aug

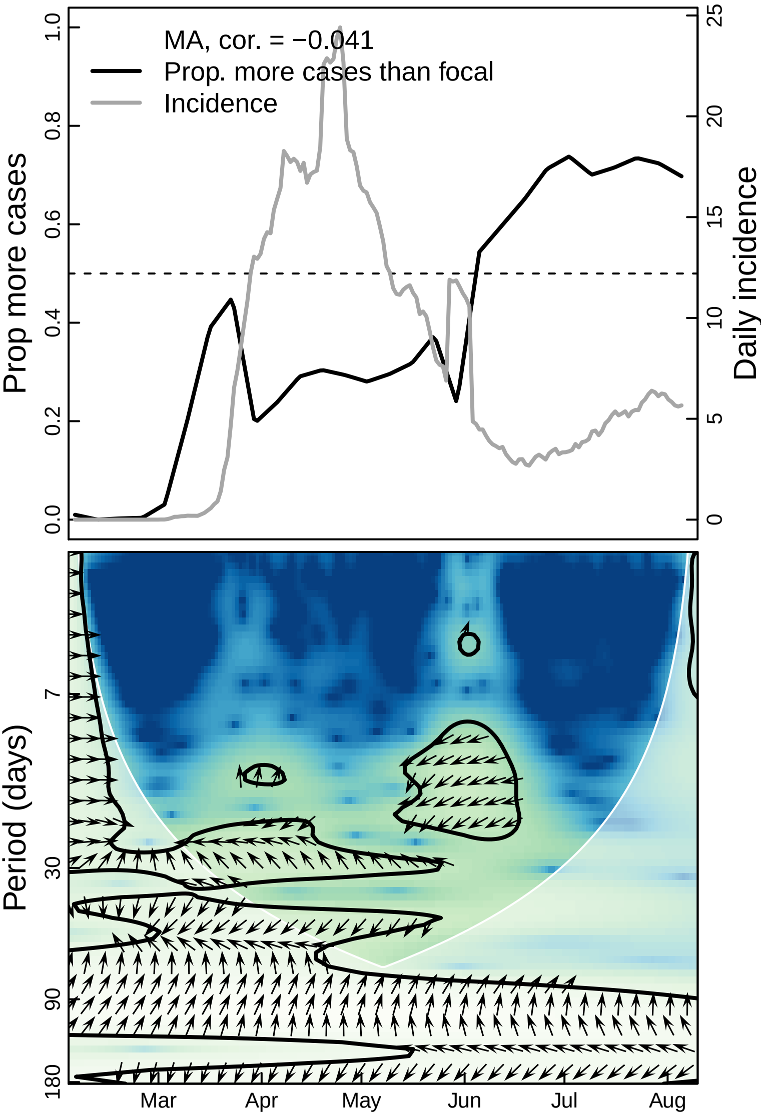

MA, cor. = −0.041
Prop. more cases than focal
Incidence
Prop more cases
Daily incidence
0.0
0.2
0.4
0.6
0.8
1.0
0
5
10
15
20
25
Period (days)
7
30
90
180
Mar
Apr
May
Jun
Jul
Aug

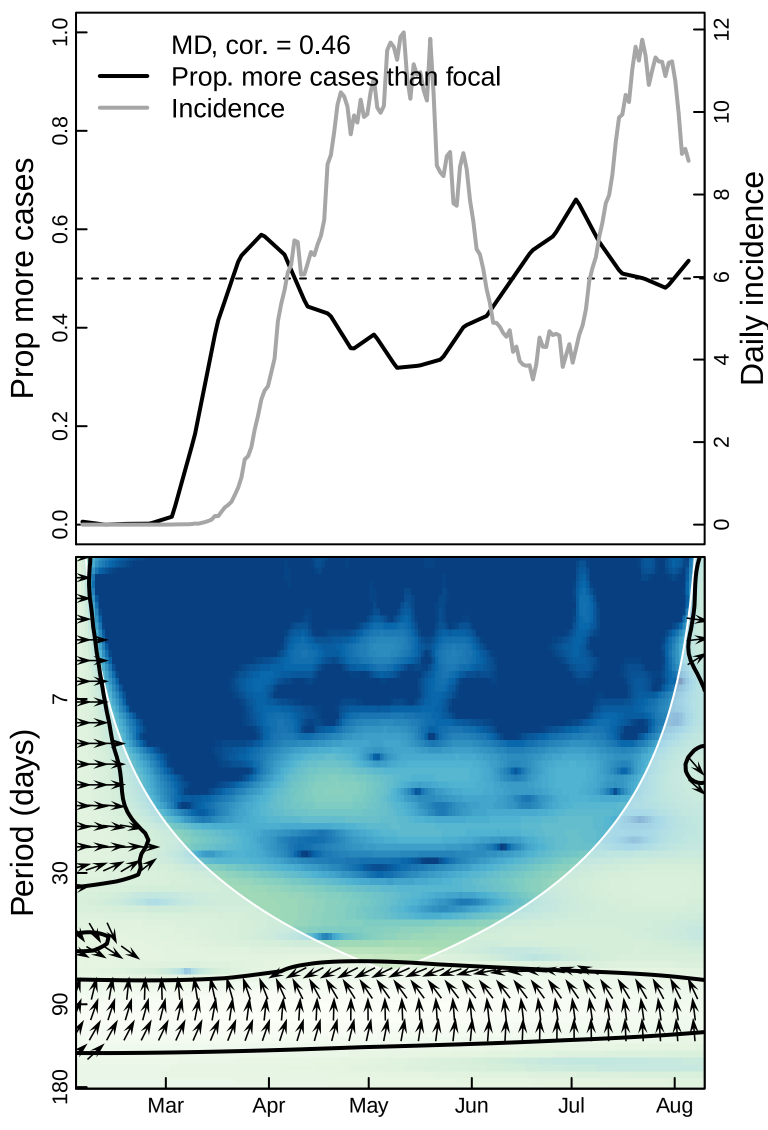

MD, cor. = 0.46
Prop. more cases than focal
Incidence
Prop more cases
Daily incidence
Period (days)
0.0
0.2
0.4
0.6
0.8
1.0
0
2
4
6
8
10
12
7
30
90
180
Mar
Apr
May
Jun
Jul
Aug

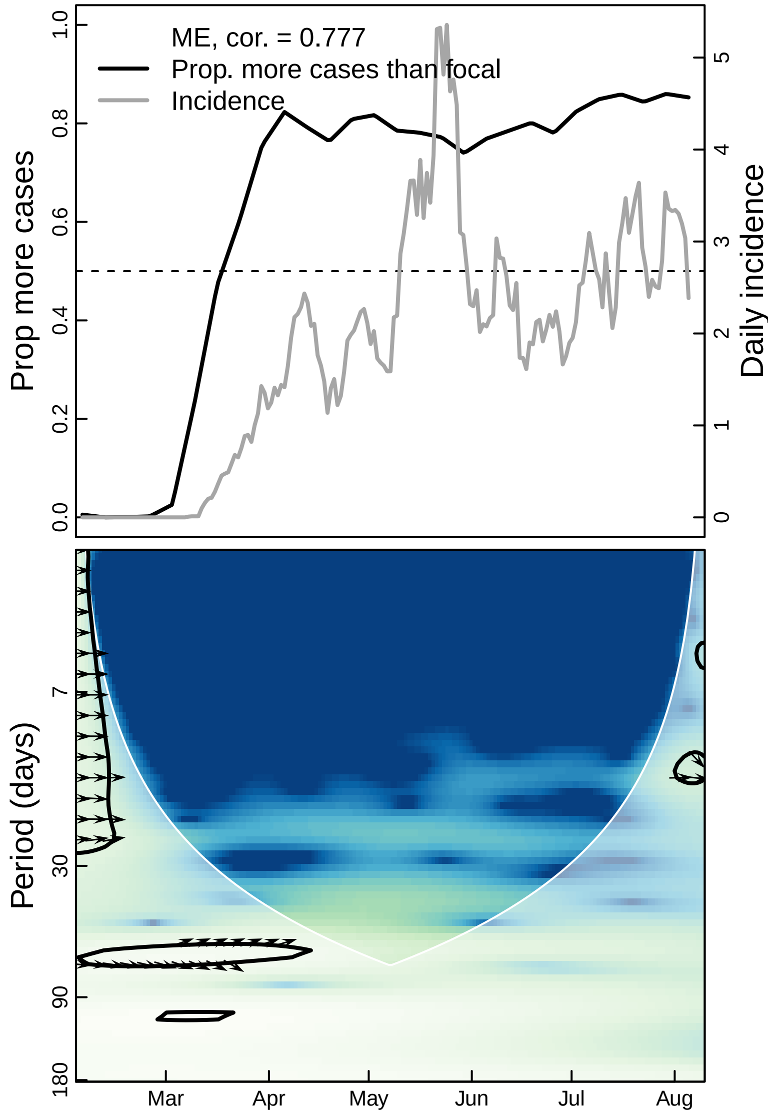
ME, cor. = 0.777
Prop. more cases than focal
Incidence
Prop more cases
Daily incidence
0.0
0.2
0.4
0.6
0.8
1.0
0
1
2
3
4
5
Period (days)
7
30
90
180
Mar
Apr
May
Jun
Jul
Aug

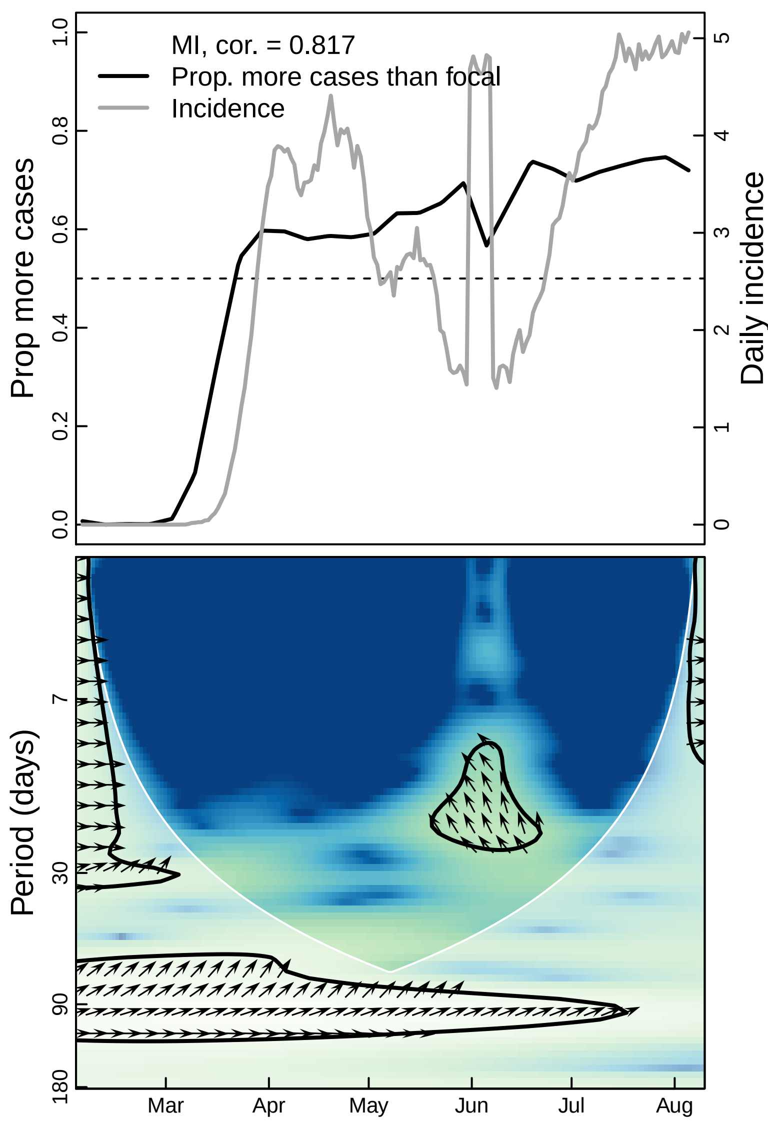
MI, cor. = 0.817
Prop. more cases than focal
Incidence
Prop more cases
Daily incidence
0.0
0.2
0.4
0.6
0.8
1.0
0
1
2
3
4
5
Period (days)
7
30
90
180
Mar
Apr
May
Jun
Jul
Aug

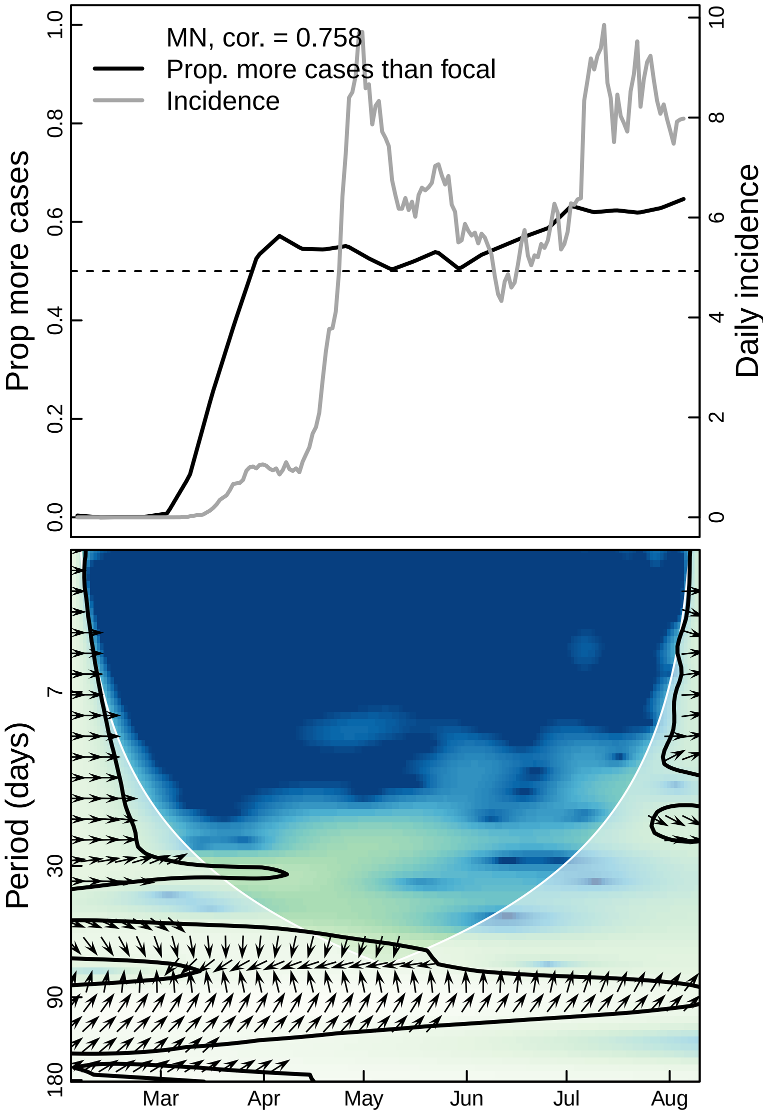
MN, cor. = 0.758
Prop. more cases than focal
Incidence
Prop more cases
Daily incidence
0.0
0.2
0.4
0.6
0.8
1.0
0
2
4
6
8
10
Period (days)
7
30
90
180
Mar
Apr
May
Jun
Jul
Aug

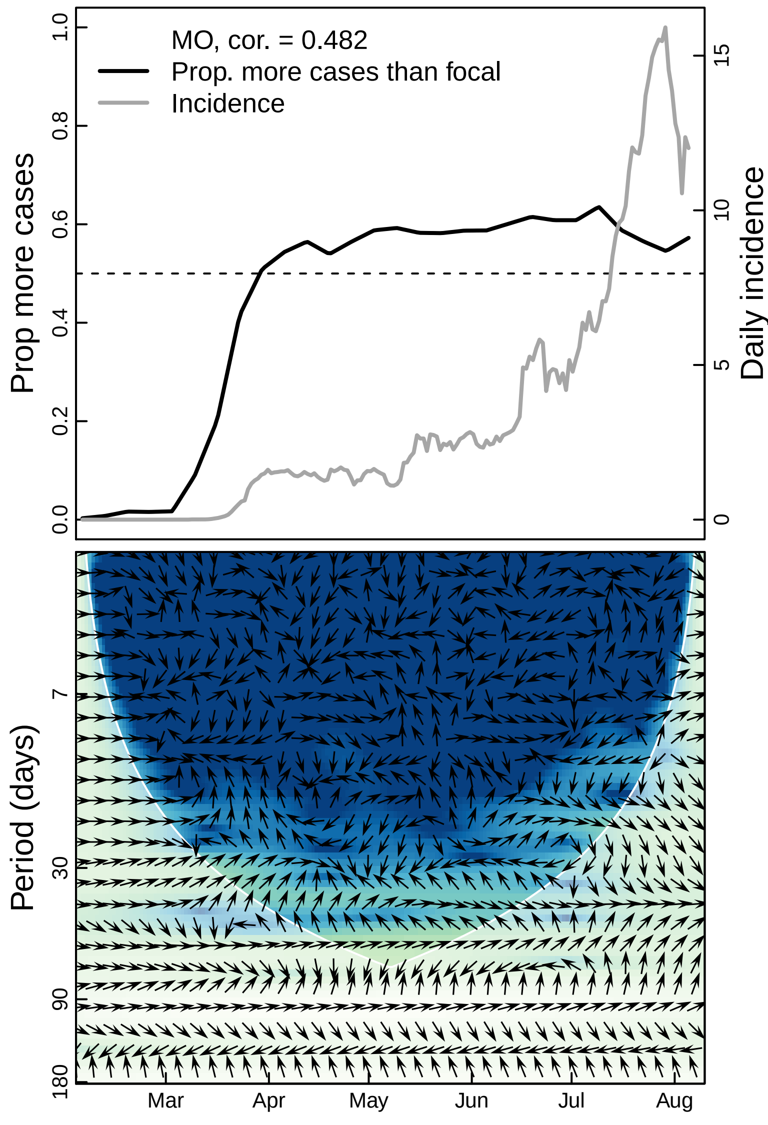
MO, cor. = 0.482
Prop. more cases than focal
Incidence
Prop more cases
Daily incidence
0.0
0.2
0.4
0.6
0.8
1.0
0
5
10
15
Period (days)
7
30
90
180
Mar
Apr
May
Jun
Jul
Aug

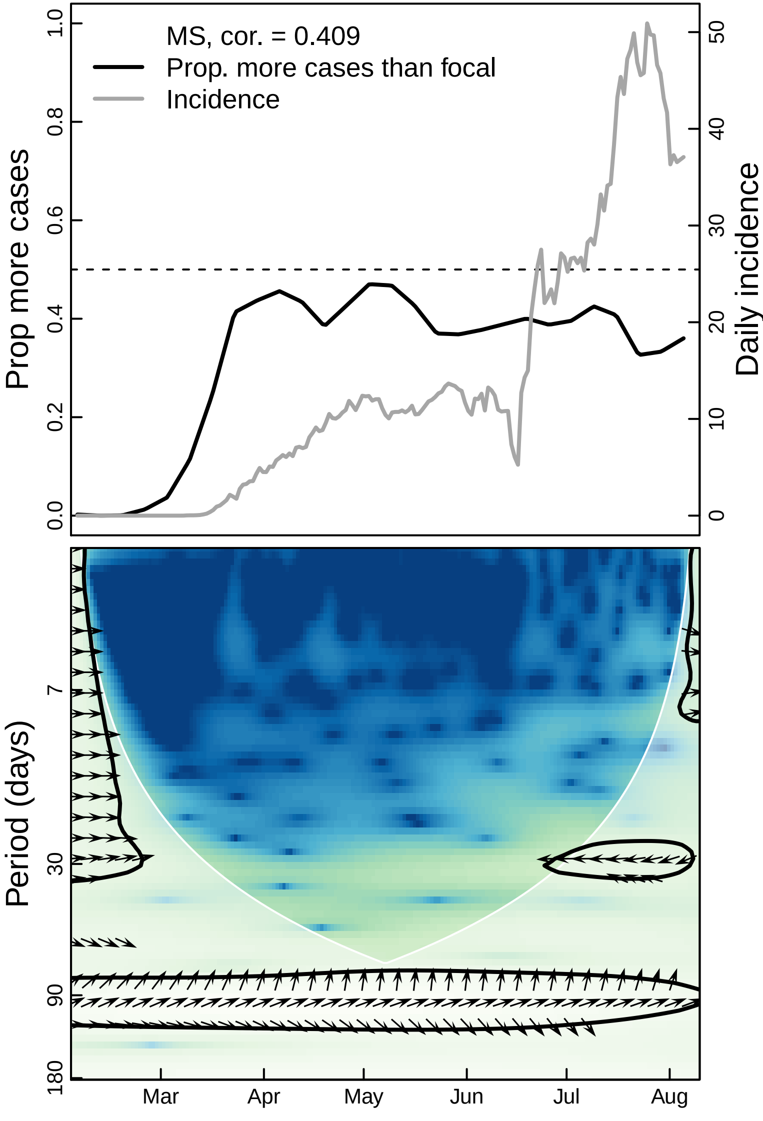

MS, cor. = 0.409
Prop. more cases than focal
Incidence
Prop more cases
Daily incidence
0.0
0.2
0.4
0.6
0.8
1.0
0
10
20
30
40
50
Period (days)
7
30
90
180
Mar
Apr
May
Jun
Jul
Aug

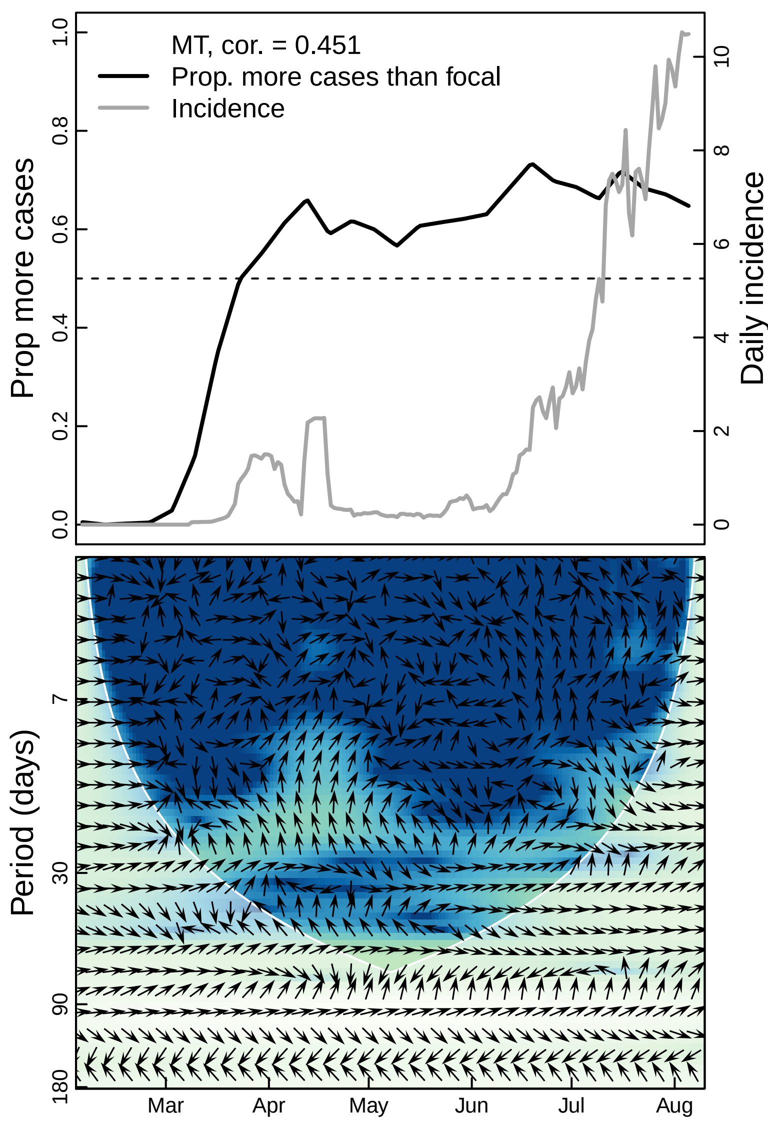
MT, cor. = 0.451
Prop. more cases than focal
Incidence
Prop more cases
Daily incidence
0.0
0.2
0.4
0.6
0.8
1.0
0
2
4
6
8
10
Period (days)
7
30
90
180
Mar
Apr
May
Jun
Jul
Aug

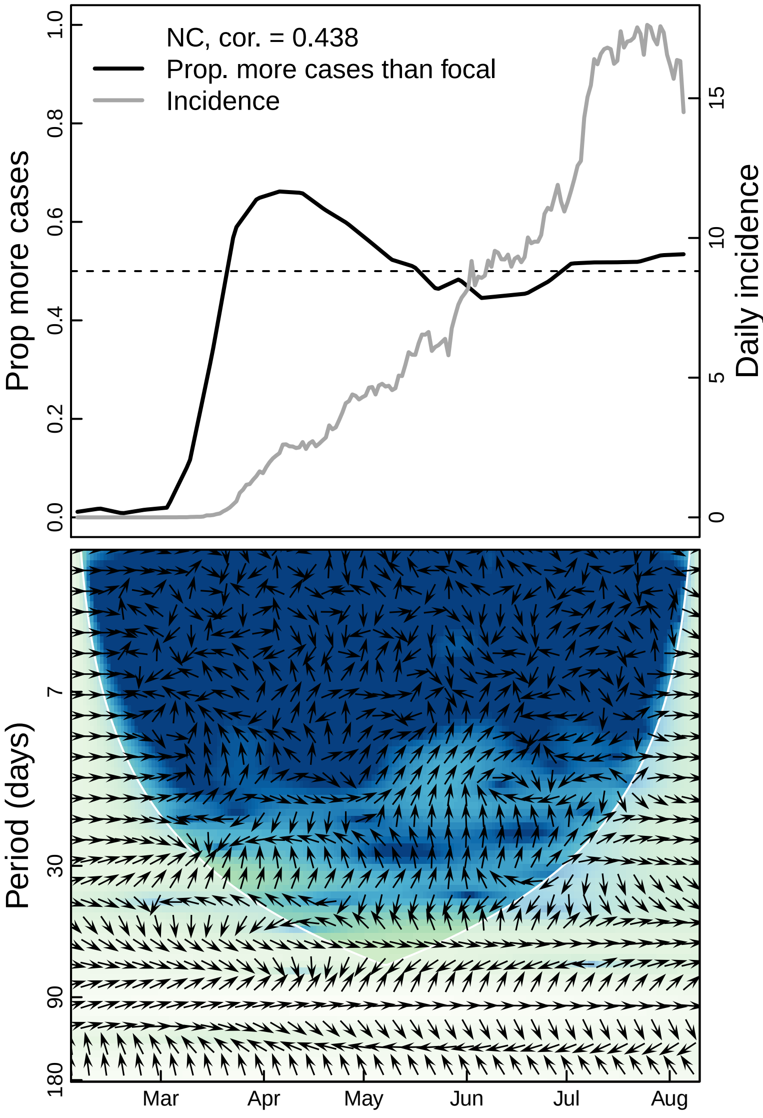

NC, cor. = 0.438
Prop. more cases than focal
Incidence
Prop more cases
Daily incidence
0.0
0.2
0.4
0.6
0.8
1.0
0
5
10
15
Period (days)
7
30
90
180
Mar
Apr
May
Jun
Jul
Aug

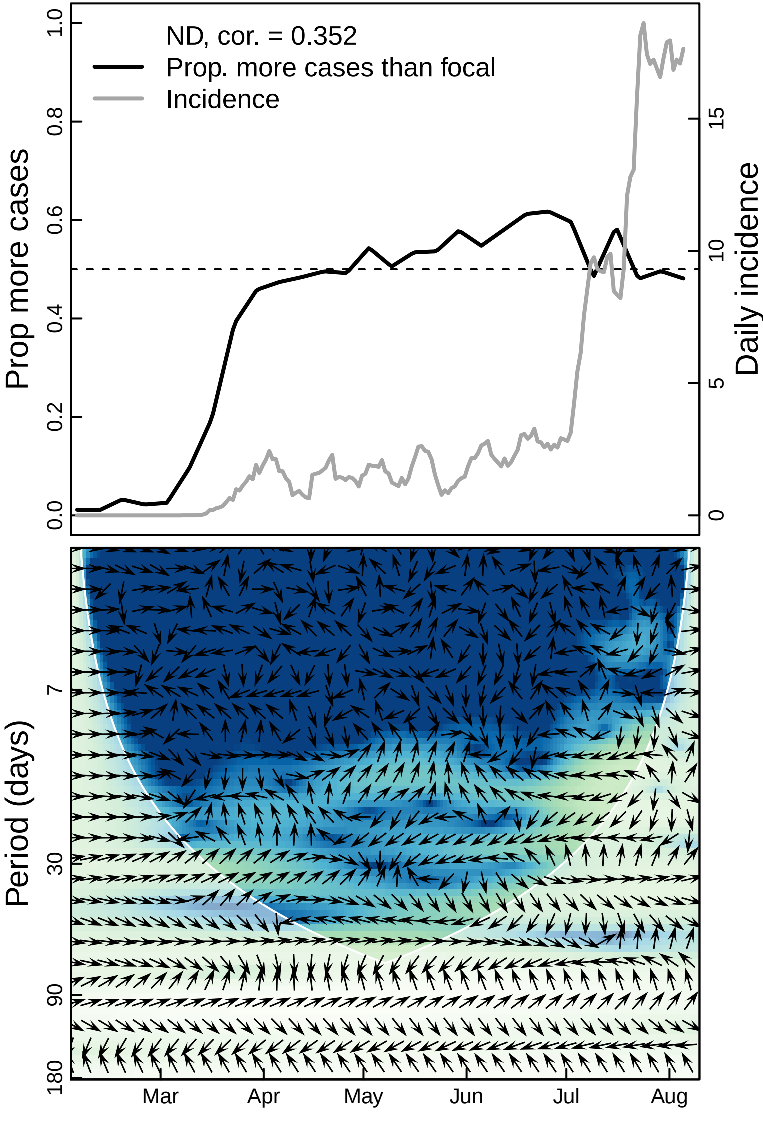

ND, cor. = 0.352
Prop. more cases than focal
Incidence
Prop more cases
Daily incidence
0.0
0.2
0.4
0.6
0.8
1.0
0
5
10
15
Period (days)
7
30
90
180
Mar
Apr
May
Jun
Jul
Aug

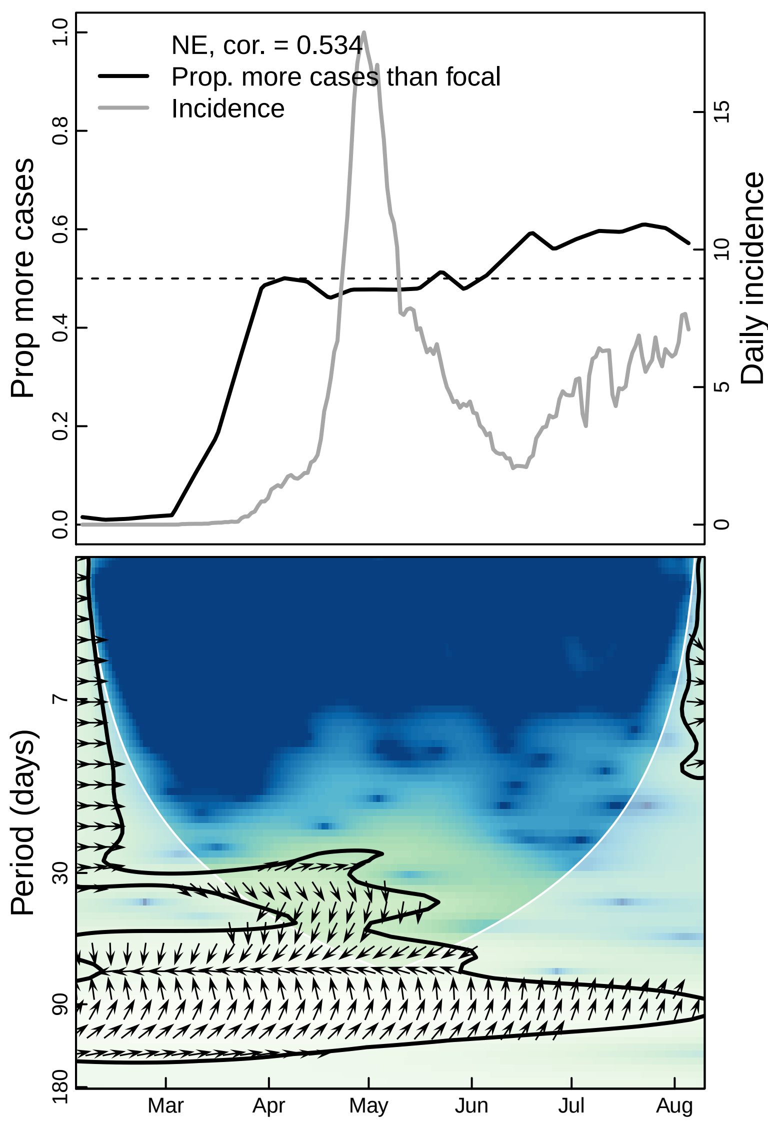

NE, cor. = 0.534
Prop. more cases than focal
Incidence
Prop more cases
Daily incidence
0.0
0.2
0.4
0.6
0.8
1.0
0
5
10
15
Period (days)
7
30
90
180
Mar
Apr
May
Jun
Jul
Aug

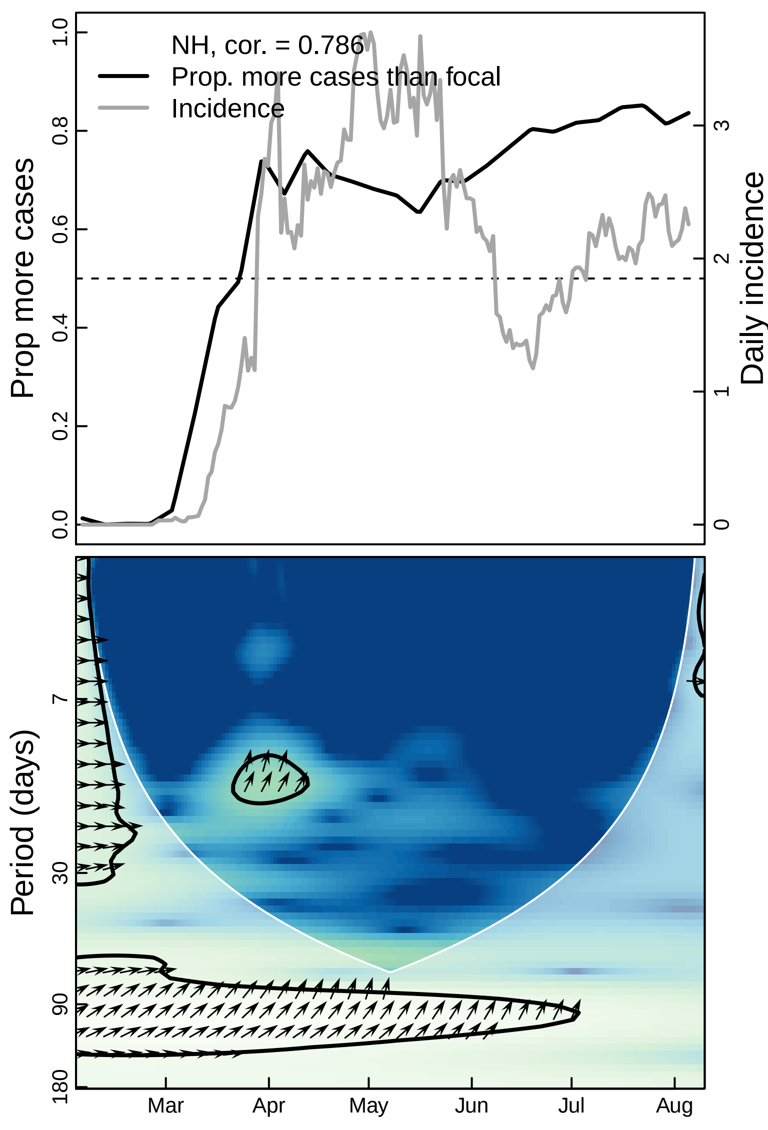
NH, cor. = 0.786
Prop. more cases than focal
Incidence
Prop more cases
Daily incidence
0.0
0.2
0.4
0.6
0.8
1.0
0
1
2
3
Period (days)
7
30
90
180
Mar
Apr
May
Jun
Jul
Aug

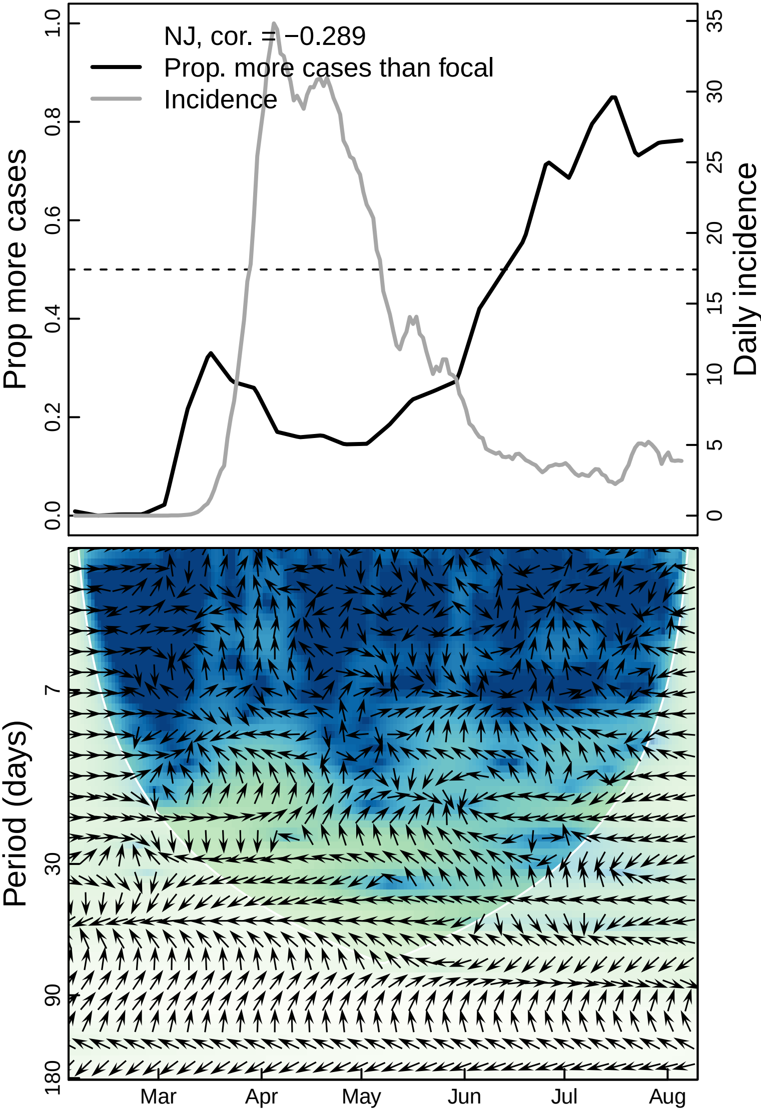
NJ, cor. = −0.289
Prop. more cases than focal
Incidence
Prop more cases
Daily incidence
0.0
0.2
0.4
0.6
0.8
1.0
0
5
10
15
20
25
30
35
Period (days)
7
30
90
180
Mar
Apr
May
Jun
Jul
Aug

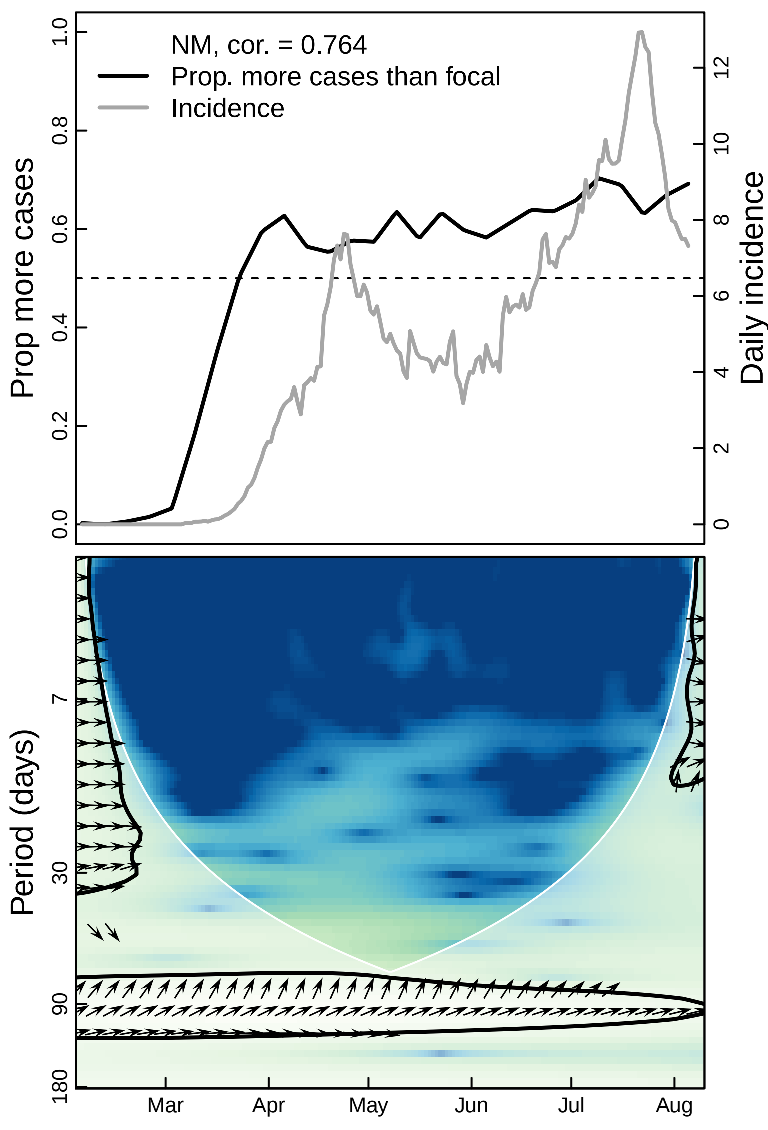

NM, cor. = 0.764
Prop. more cases than focal
Incidence
Prop more cases
Daily incidence
0.0
0.2
0.4
0.6
0.8
1.0
0
2
4
6
8
10
12
Period (days)
7
30
90
180
Mar
Apr
May
Jun
Jul
Aug

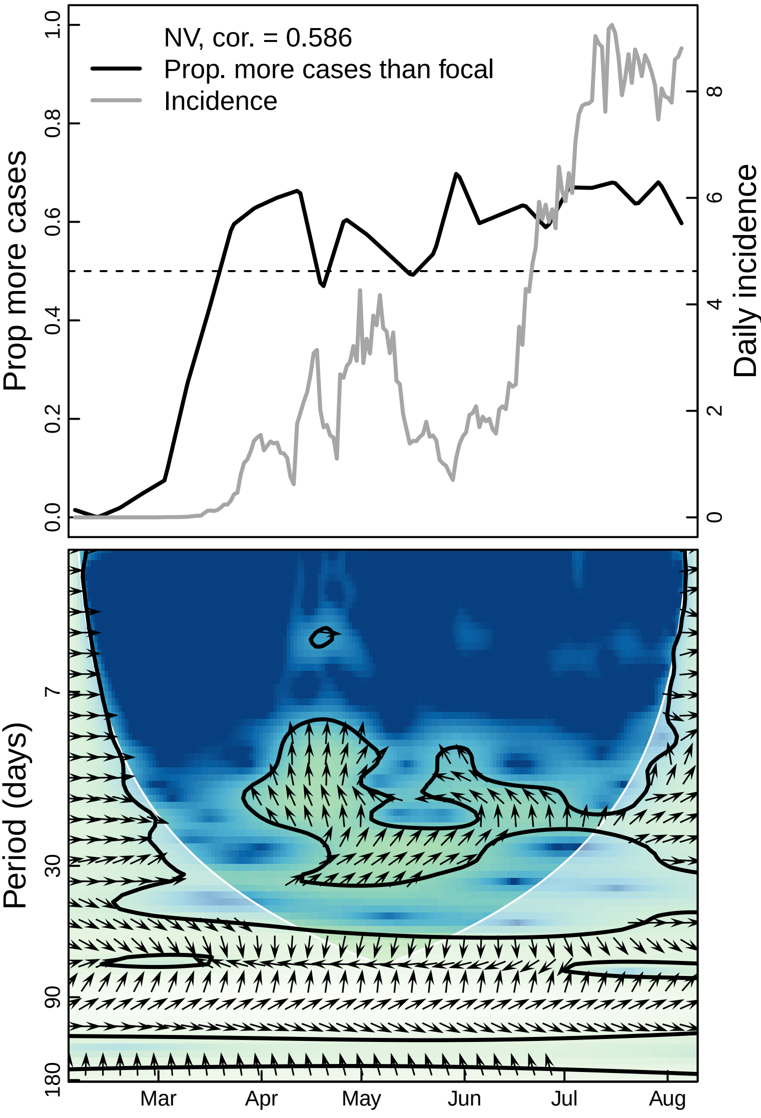

NV, cor. = 0.586
Prop. more cases than focal
Incidence
Prop more cases
Daily incidence
0.0
0.2
0.4
0.6
0.8
1.0
0
2
4
6
8
Period (days)
7
30
90
180
Mar
Apr
May
Jun
Jul
Aug

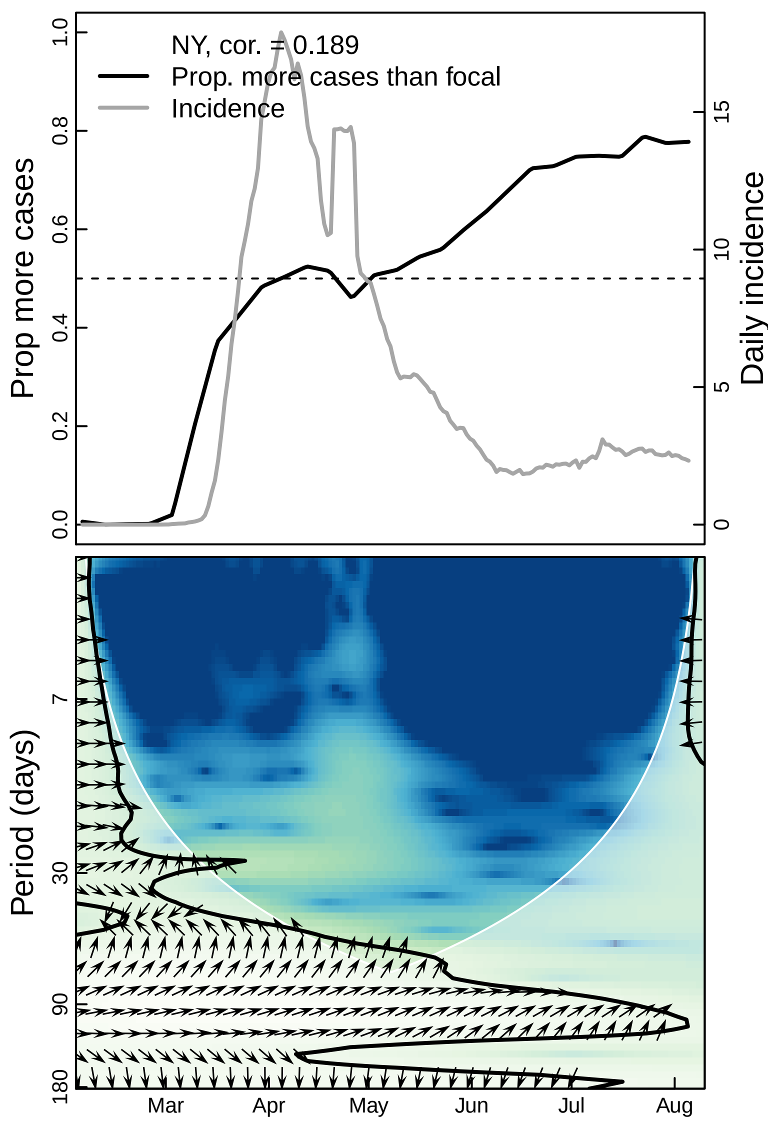
NY, cor. = 0.189
Prop. more cases than focal
Incidence
Prop more cases
Daily incidence
0.0
0.2
0.4
0.6
0.8
1.0
0
5
10
15
Period (days)
7
30
90
180
Mar
Apr
May
Jun
Jul
Aug

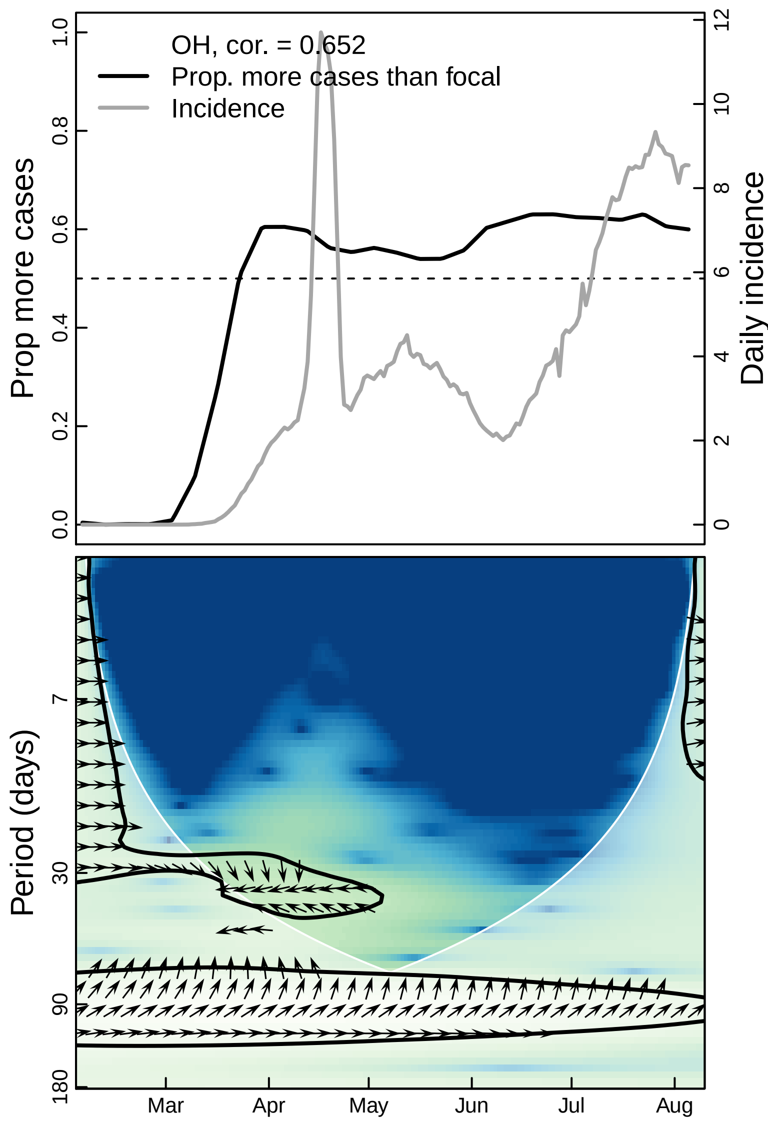
OH, cor. = 0.652
Prop. more cases than focal
Incidence
Prop more cases
Daily incidence
0.0
0.2
0.4
0.6
0.8
1.0
0
2
4
6
8
10
12
Period (days)
7
30
90
180
Mar
Apr
May
Jun
Jul
Aug

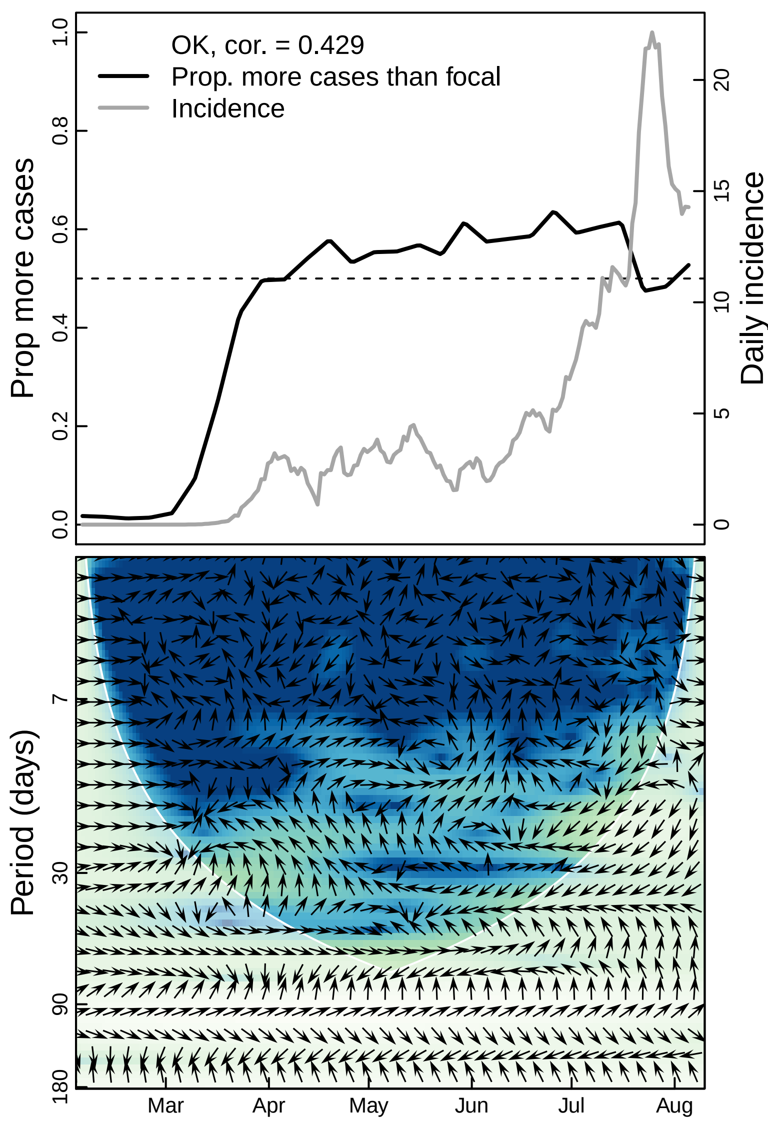

OK, cor. = 0.429
Prop. more cases than focal
Incidence
Prop more cases
Daily incidence
0.0
0.2
0.4
0.6
0.8
1.0
0
5
10
15
20
Period (days)
7
30
90
180
Mar
Apr
May
Jun
Jul
Aug

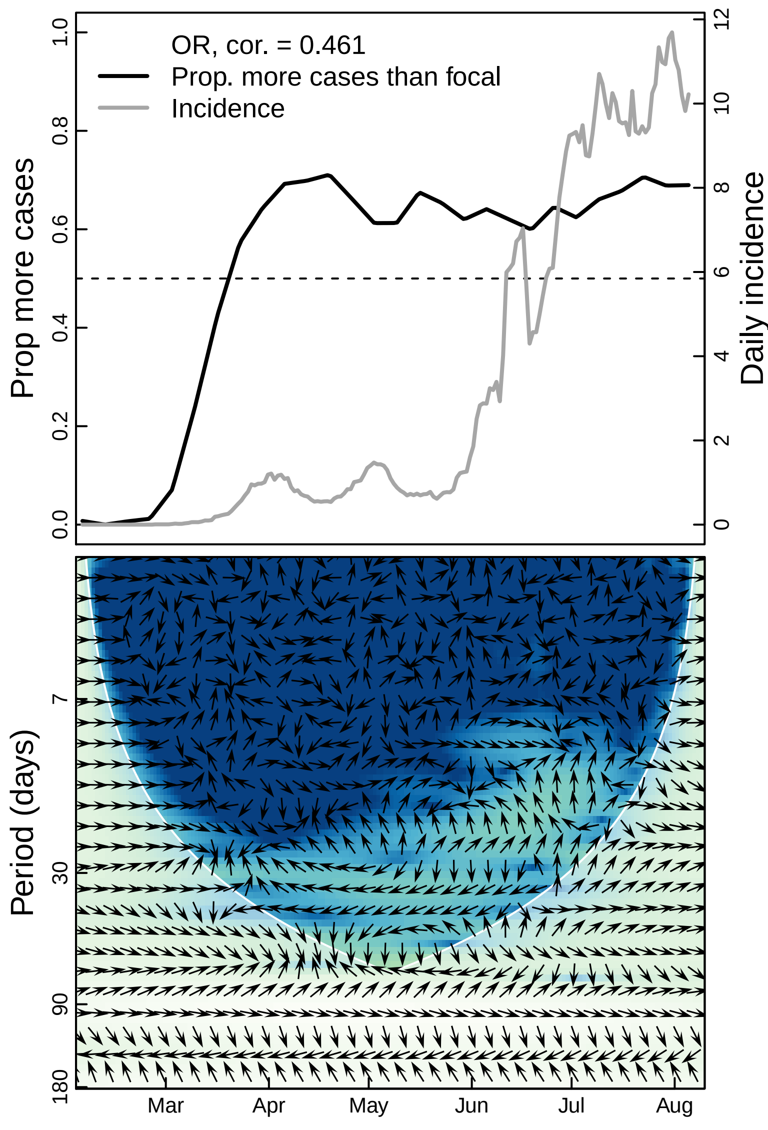
OR, cor. = 0.461
Prop. more cases than focal
Incidence
Prop more cases
Daily incidence
0.0
0.2
0.4
0.6
0.8
1.0
0
2
4
6
8
10
12
Period (days)
7
30
90
180
Mar
Apr
May
Jun
Jul
Aug

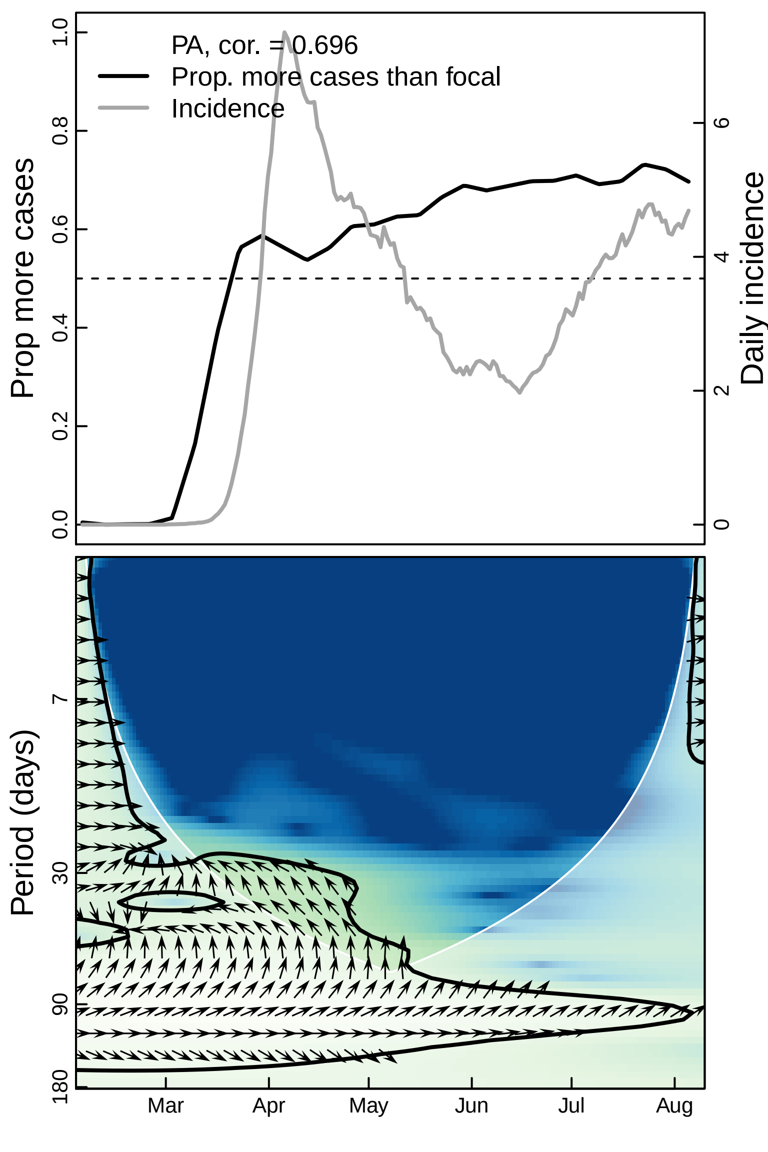

PA, cor. = 0.696
Prop. more cases than focal
Incidence
Prop more cases
Daily incidence
0.0
0.2
0.4
0.6
0.8
1.0
0
2
4
6
Period (days)
7
30
90
180
Mar
Apr
May
Jun
Jul
Aug

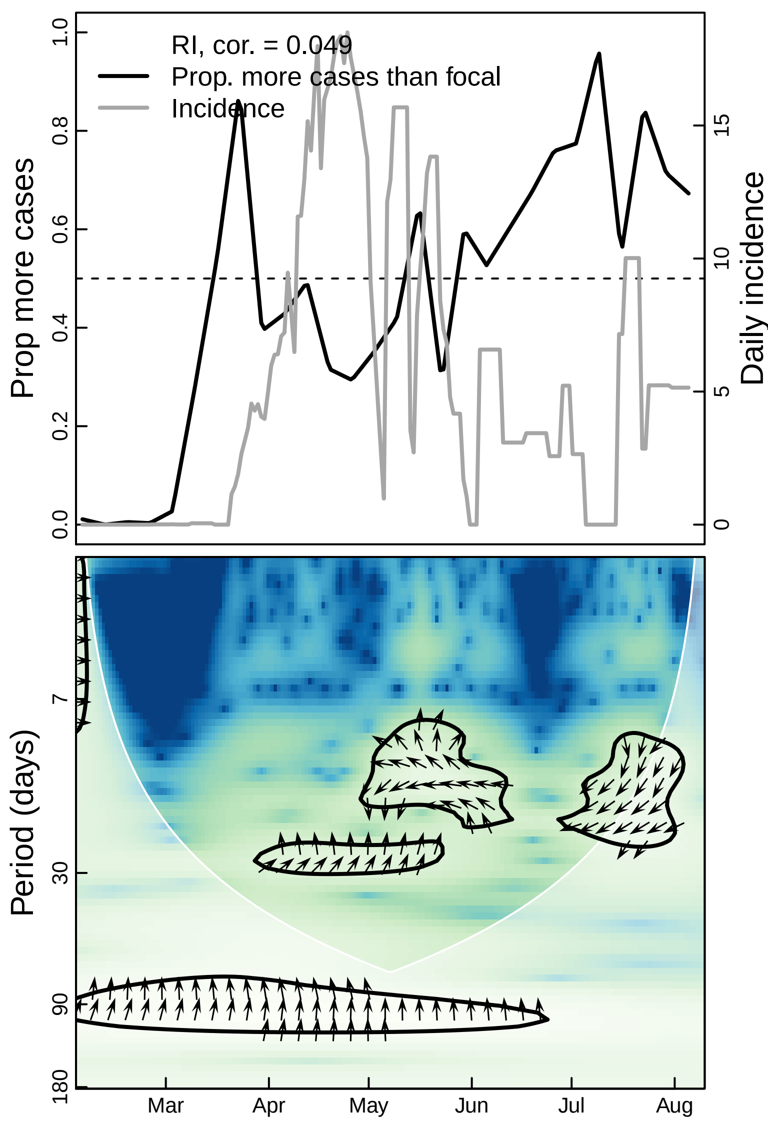
RI, cor. = 0.049
Prop. more cases than focal
Incidence
Prop more cases
Daily incidence
0.0
0.2
0.4
0.6
0.8
1.0
0
5
10
15
Period (days)
7
30
90
180
Mar
Apr
May
Jun
Jul
Aug

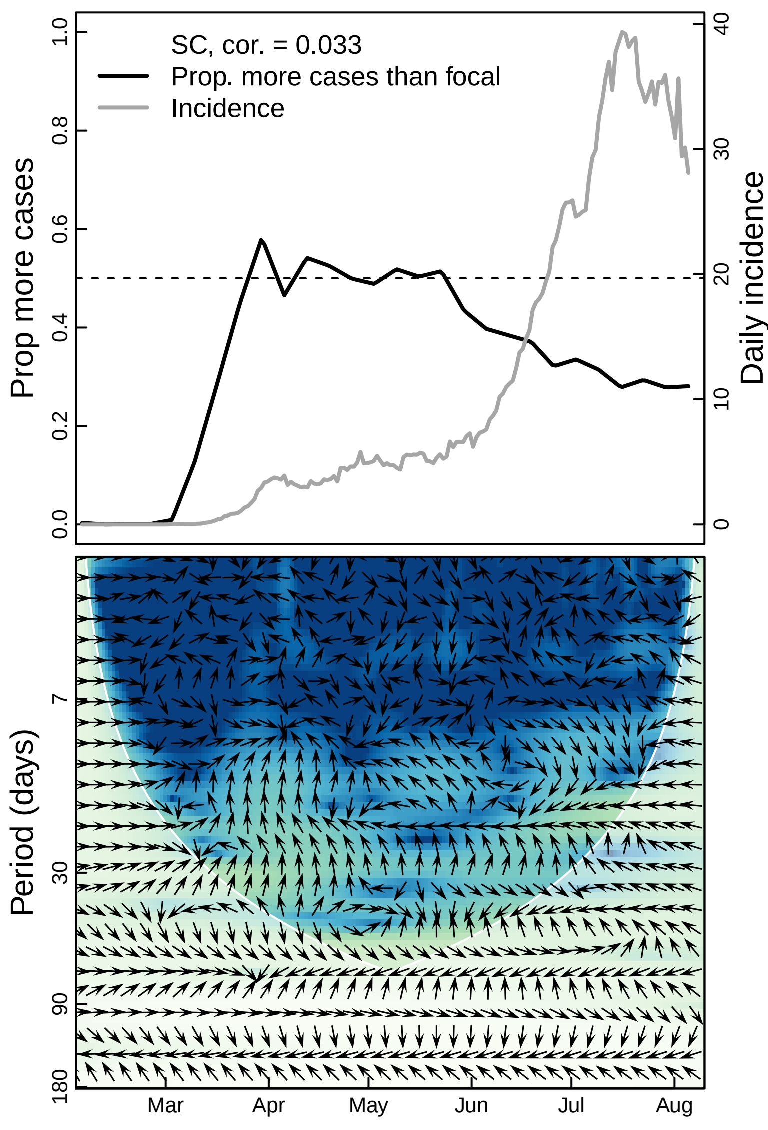

SC, cor. = 0.033
Prop. more cases than focal
Incidence
Prop more cases
Daily incidence
0.0
0.2
0.4
0.6
0.8
1.0
0
10
20
30
40
Period (days)
7
30
90
180
Mar
Apr
May
Jun
Jul
Aug

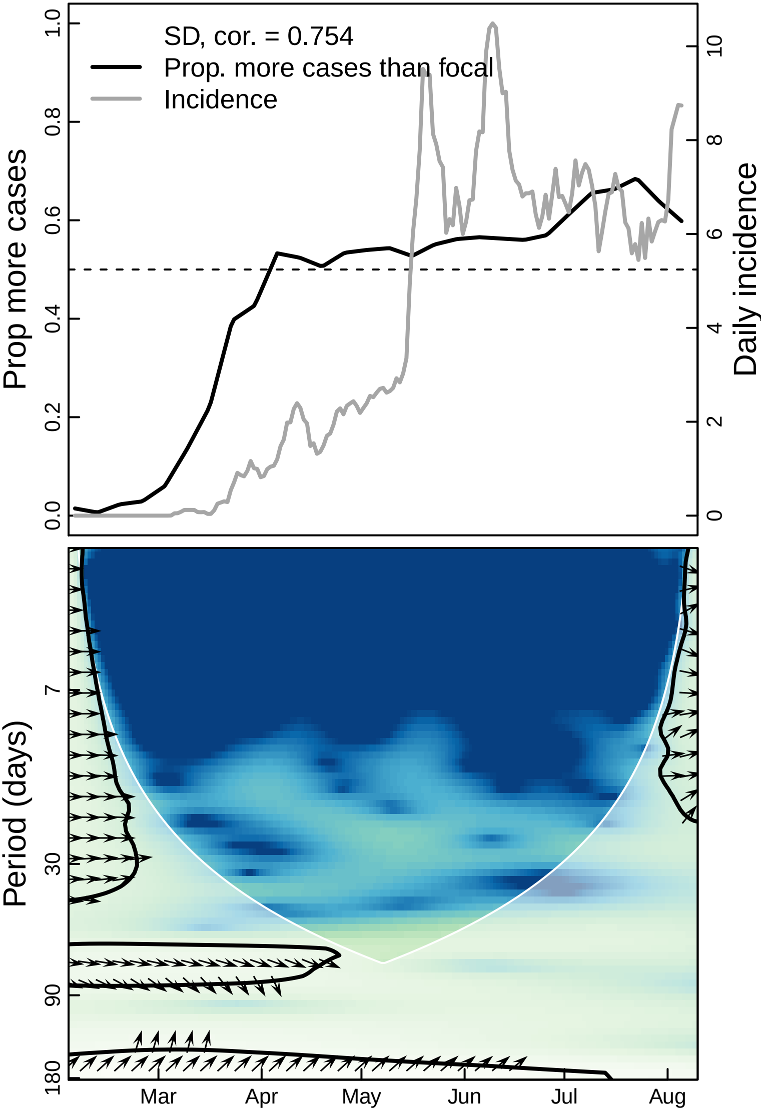

SD, cor. = 0.754
Prop. more cases than focal
Incidence
Prop more cases
Daily incidence
0.0
0.2
0.4
0.6
0.8
1.0
0
2
4
6
8
10
Period (days)
7
30
90
180
Mar
Apr
May
Jun
Jul
Aug

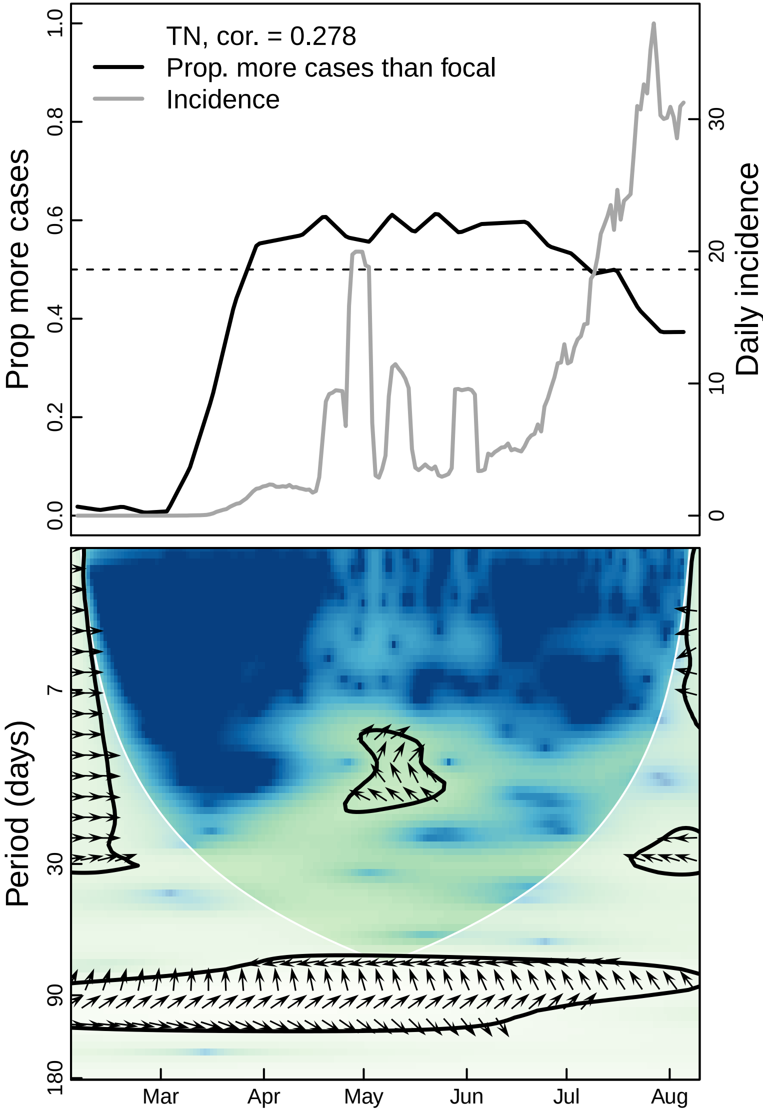

TN, cor. = 0.278
Prop. more cases than focal
Incidence
Prop more cases
Daily incidence
0.0
0.2
0.4
0.6
0.8
1.0
0
10
20
30
Period (days)
7
30
90
180
Mar
Apr
May
Jun
Jul
Aug

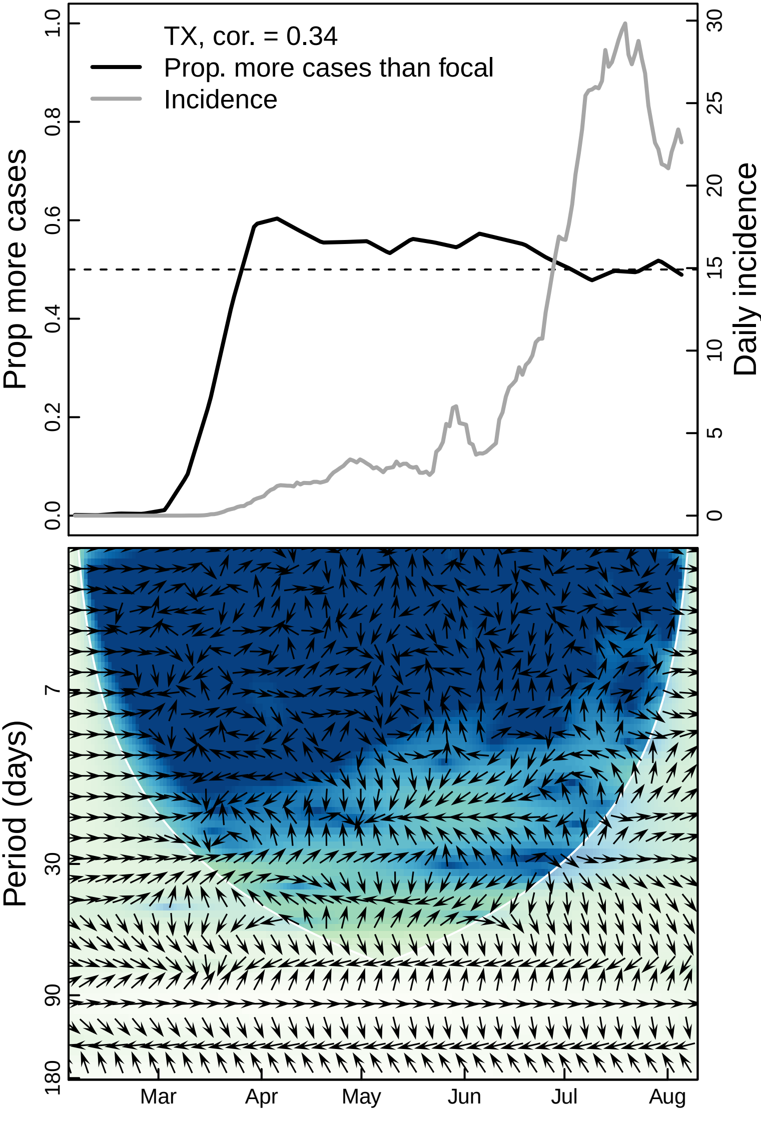

TX, cor. = 0.34
Prop. more cases than focal
Incidence
Prop more cases
Daily incidence
0.0
0.2
0.4
0.6
0.8
1.0
0
5
10
15
20
25
30
Period (days)
7
30
90
180
Mar
Apr
May
Jun
Jul
Aug

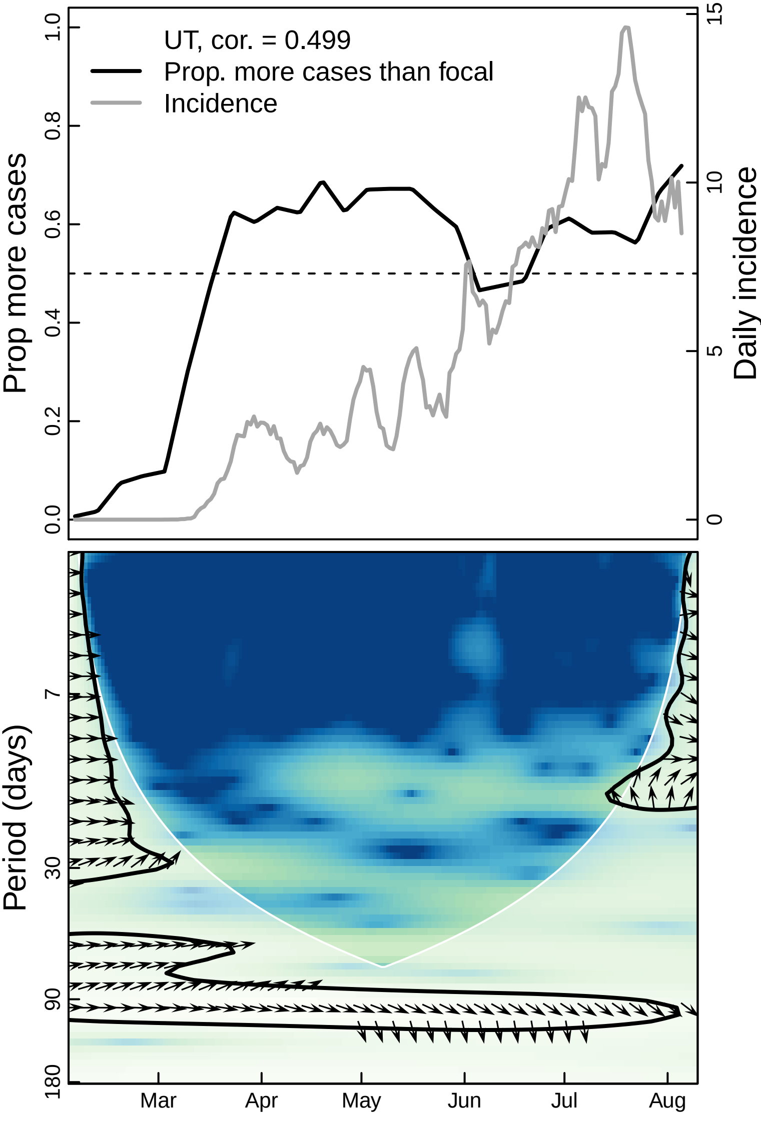
UT, cor. = 0.499
Prop. more cases than focal
Incidence
Prop more cases
Daily incidence
0.0
0.2
0.4
0.6
0.8
1.0
0
5
10
15
Period (days)
7
30
90
180
Mar
Apr
May
Jun
Jul
Aug

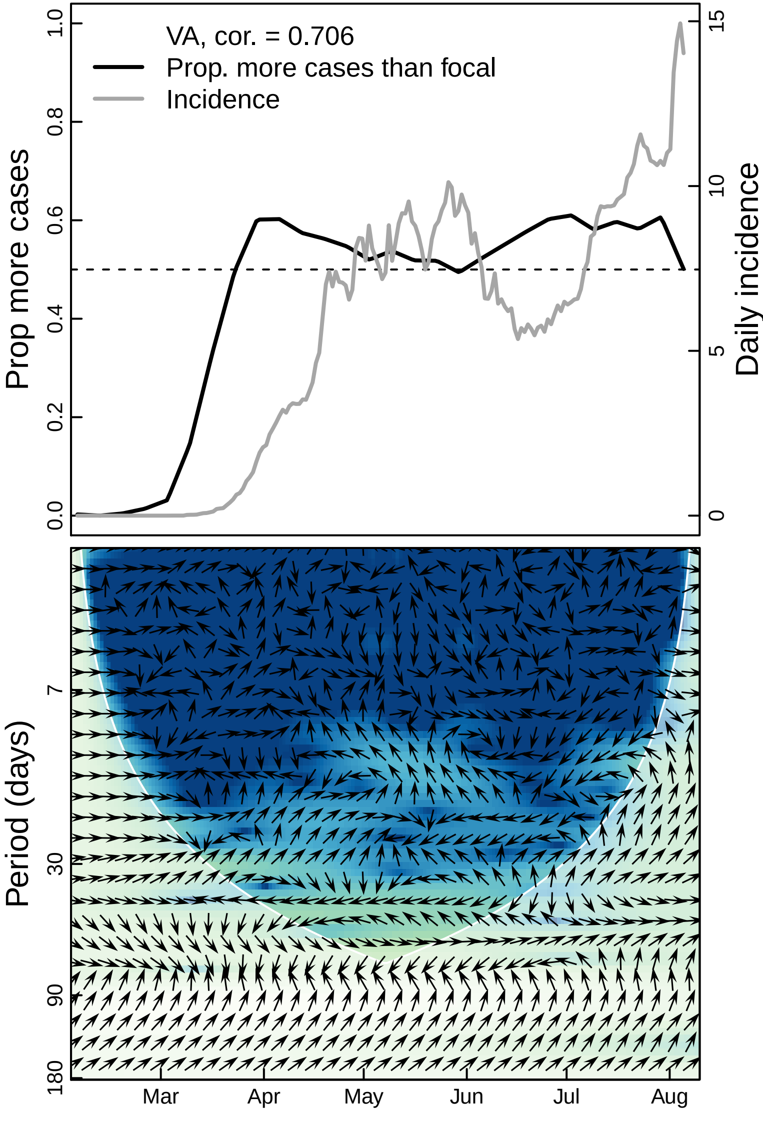

VA, cor. = 0.706
Prop. more cases than focal
Incidence
Prop more cases
Daily incidence
0.0
0.2
0.4
0.6
0.8
1.0
0
5
10
15
Period (days)
7
30
90
180
Mar
Apr
May
Jun
Jul
Aug

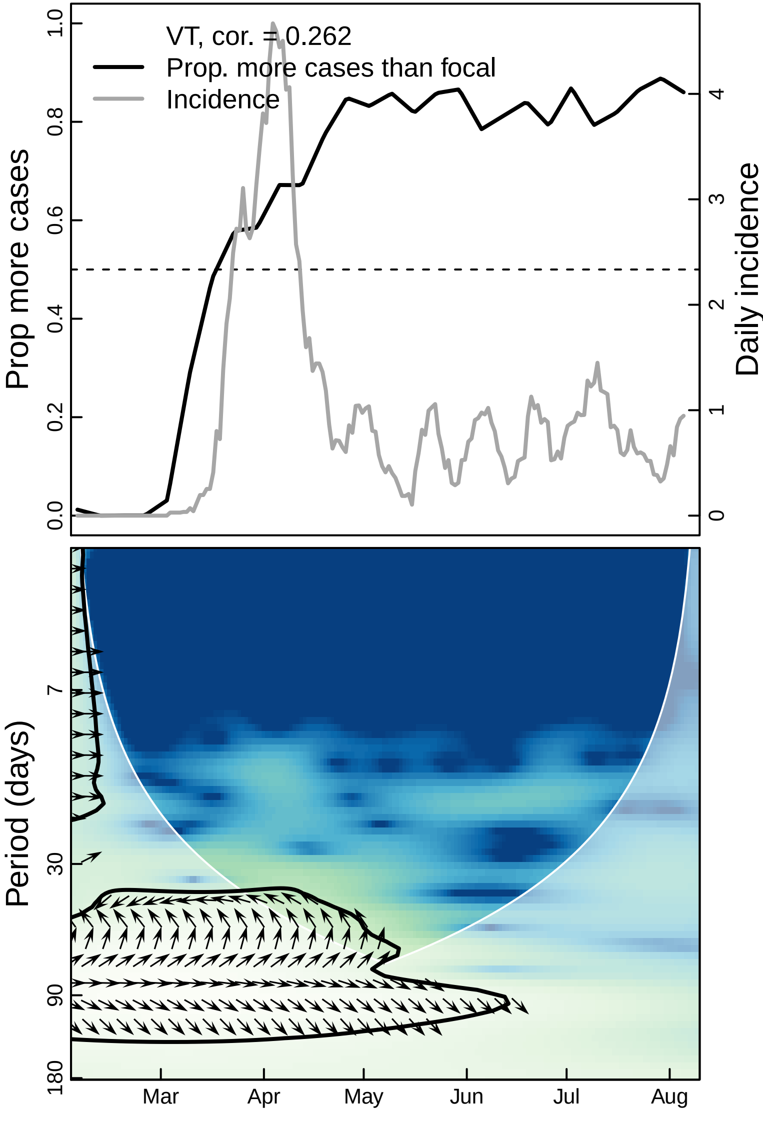

VT, cor. = 0.262
Prop. more cases than focal
Incidence
Prop more cases
Daily incidence
0.0
0.2
0.4
0.6
0.8
1.0
0
1
2
3
4
Period (days)
7
30
90
180
Mar
Apr
May
Jun
Jul
Aug

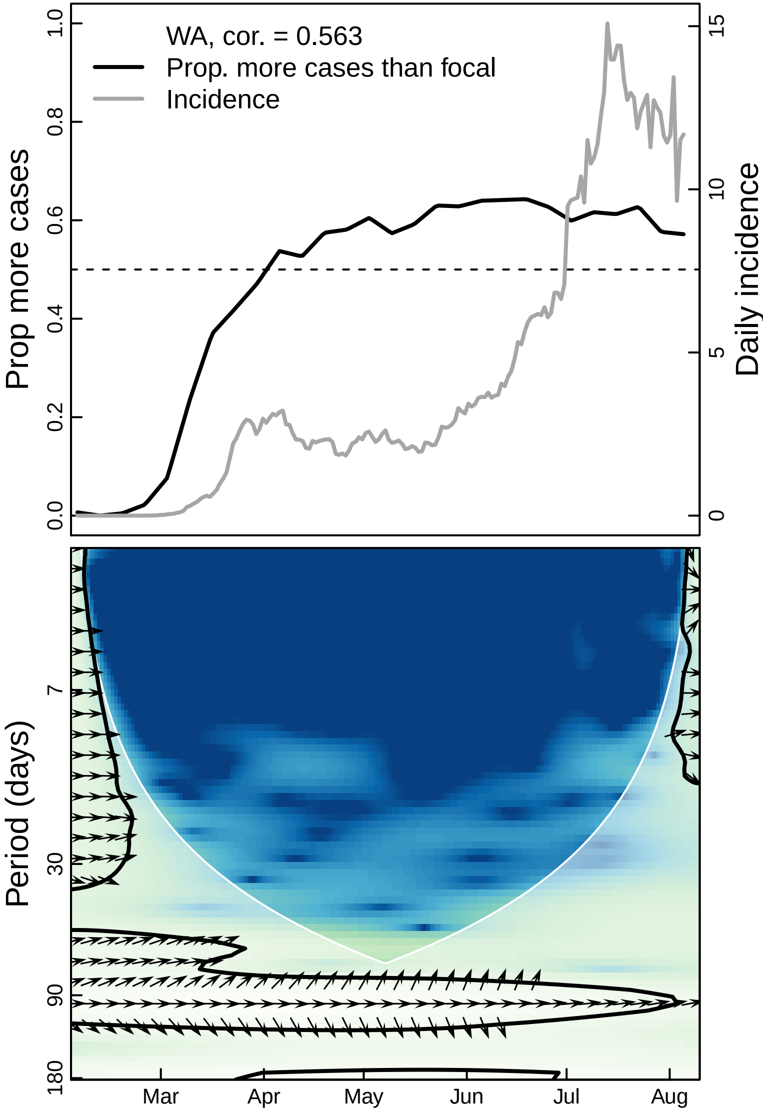

WA, cor. = 0.563
Prop. more cases than focal
Incidence
Prop more cases
Daily incidence
0.0
0.2
0.4
0.6
0.8
1.0
0
5
10
15
Period (days)
7
30
90
180
Mar
Apr
May
Jun
Jul
Aug

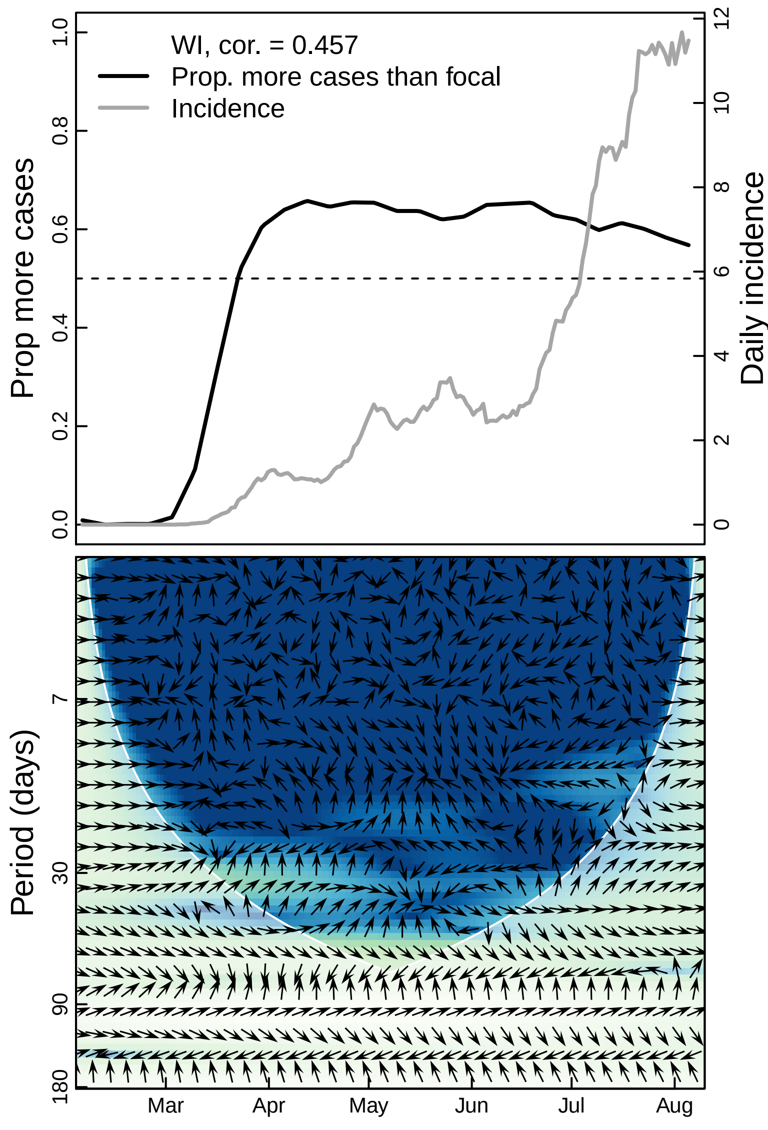
WI, cor. = 0.457
Prop. more cases than focal
Incidence
Prop more cases
Daily incidence
0.0
0.2
0.4
0.6
0.8
1.0
0
2
4
6
8
10
12
Period (days)
7
30
90
180
Mar
Apr
May
Jun
Jul
Aug

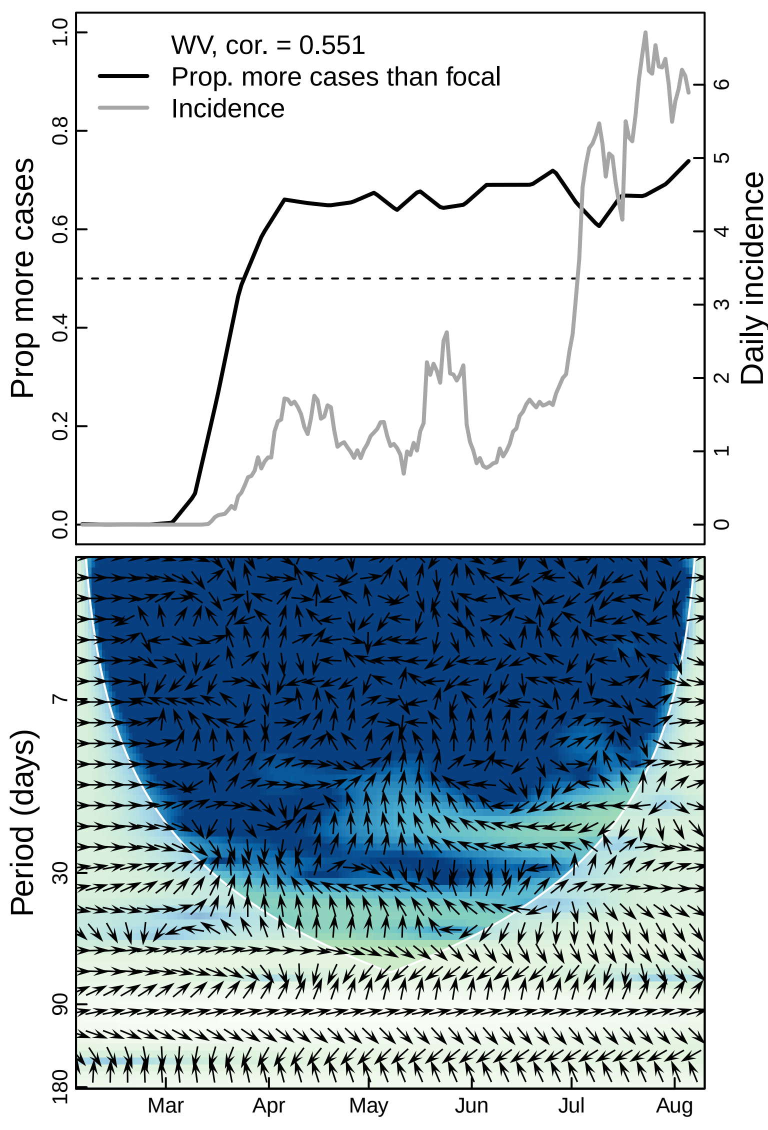
WV, cor. = 0.551
Prop. more cases than focal
Incidence
Prop more cases
Daily incidence
0.0
0.2
0.4
0.6
0.8
1.0
0
1
2
3
4
5
6
Period (days)
7
30
90
180
Mar
Apr
May
Jun
Jul
Aug

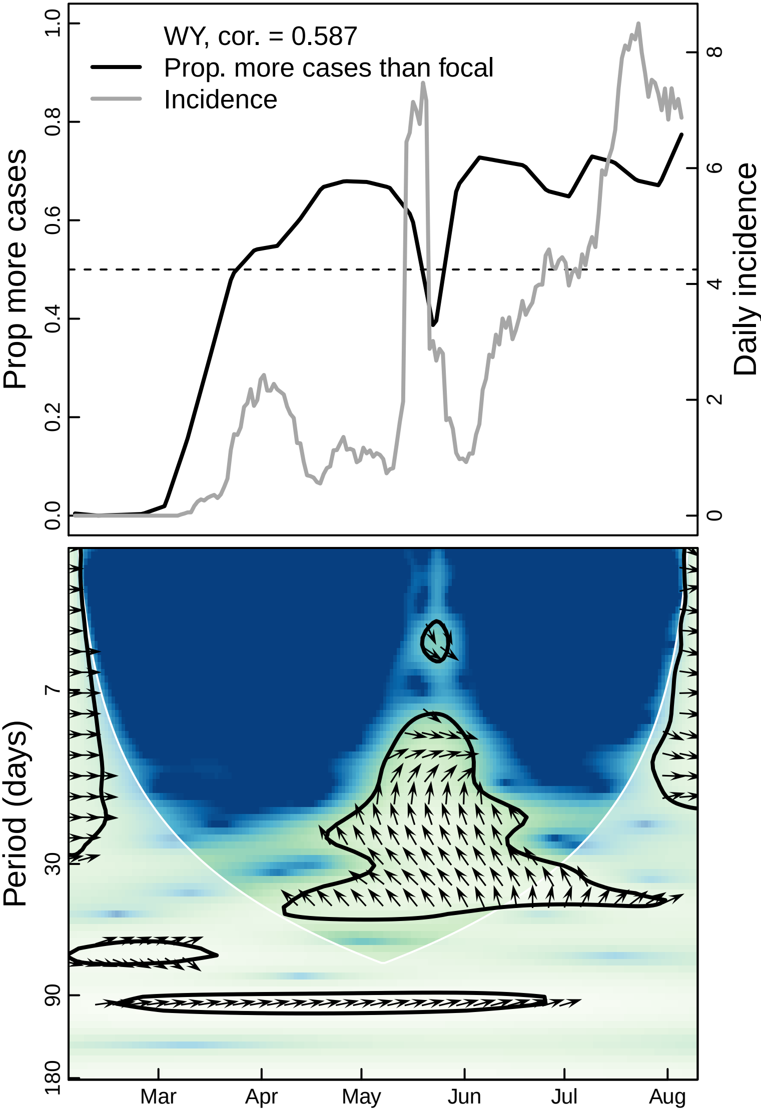

WY, cor. = 0.587
Prop. more cases than focal
Incidence
Prop more cases
Daily incidence
0.0
0.2
0.4
0.6
0.8
1.0
0
2
4
6
8
Period (days)
7
30
90
180
Mar
Apr
May
Jun
Jul
Aug
