## Supplementary material for "The unintended consequences of inconsistent pandemic control policies": Cross-wavelets - churches

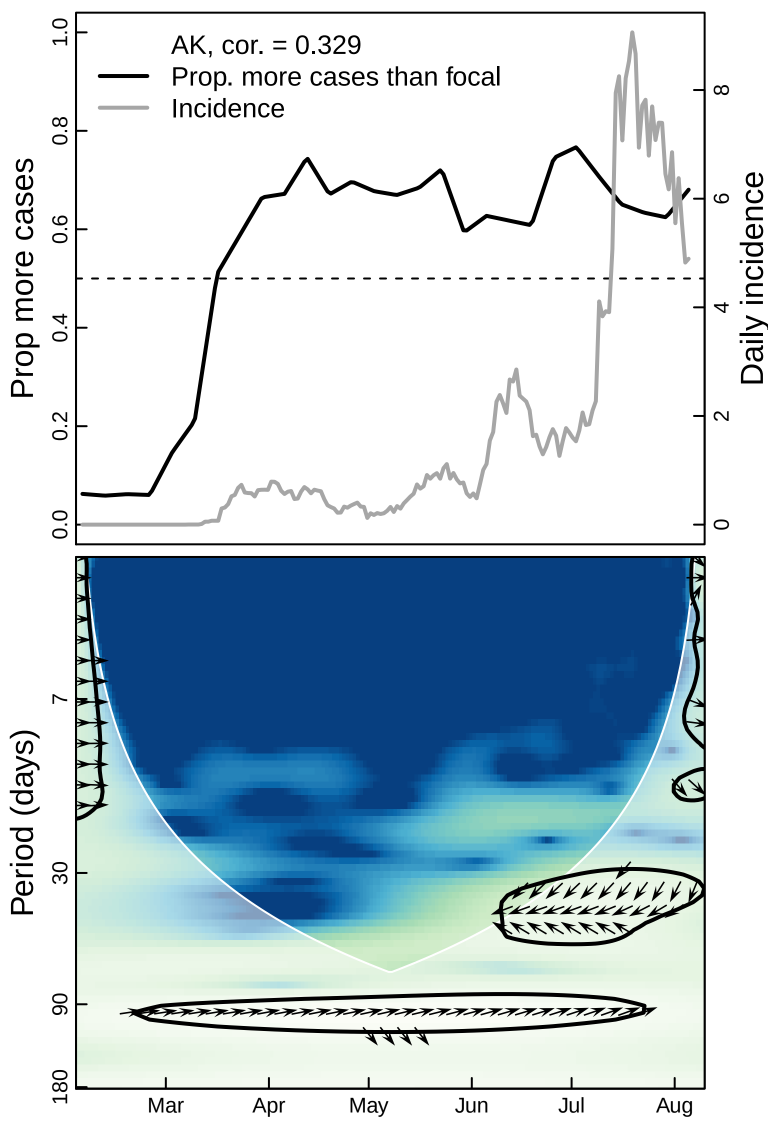

AK, cor. = 0.329
Prop. more cases than focal
Incidence
Prop more cases
Daily incidence
1.0
0.8
0.6
0.4
0.2
0.0
8
6
4
2
0
Period (days)
7
30
90
180
Mar
Apr
May
Jun
Jul
Aug

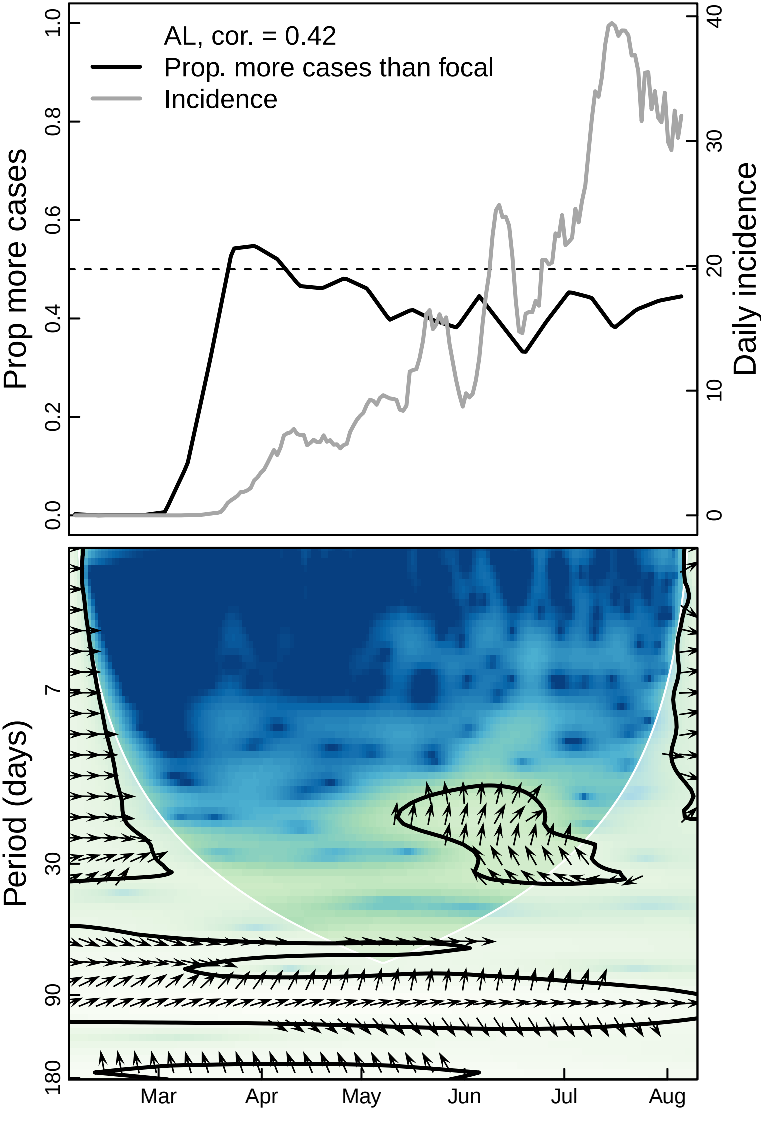

AL, cor. = 0.42
Prop. more cases than focal
Incidence
Prop more cases
Daily incidence
0.0
0.2
0.4
0.6
0.8
1.0
0
10
20
30
40
Period (days)
7
30
90
180
Mar
Apr
May
Jun
Jul
Aug

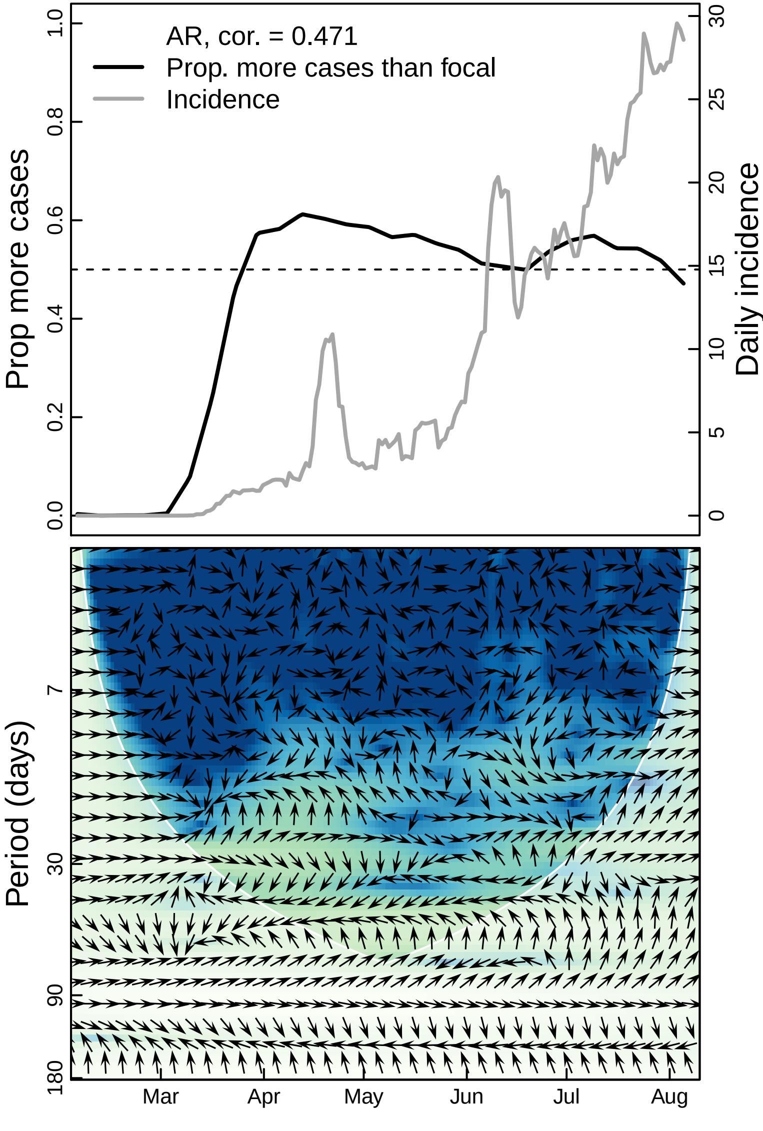

AR, cor. = 0.471
Prop. more cases than focal
Incidence
Prop more cases
Daily incidence
0.0
0.2
0.4
0.6
0.8
1.0
0
5
10
15
20
25
30
Period (days)
7
30
90
180
Mar
Apr
May
Jun
Jul
Aug

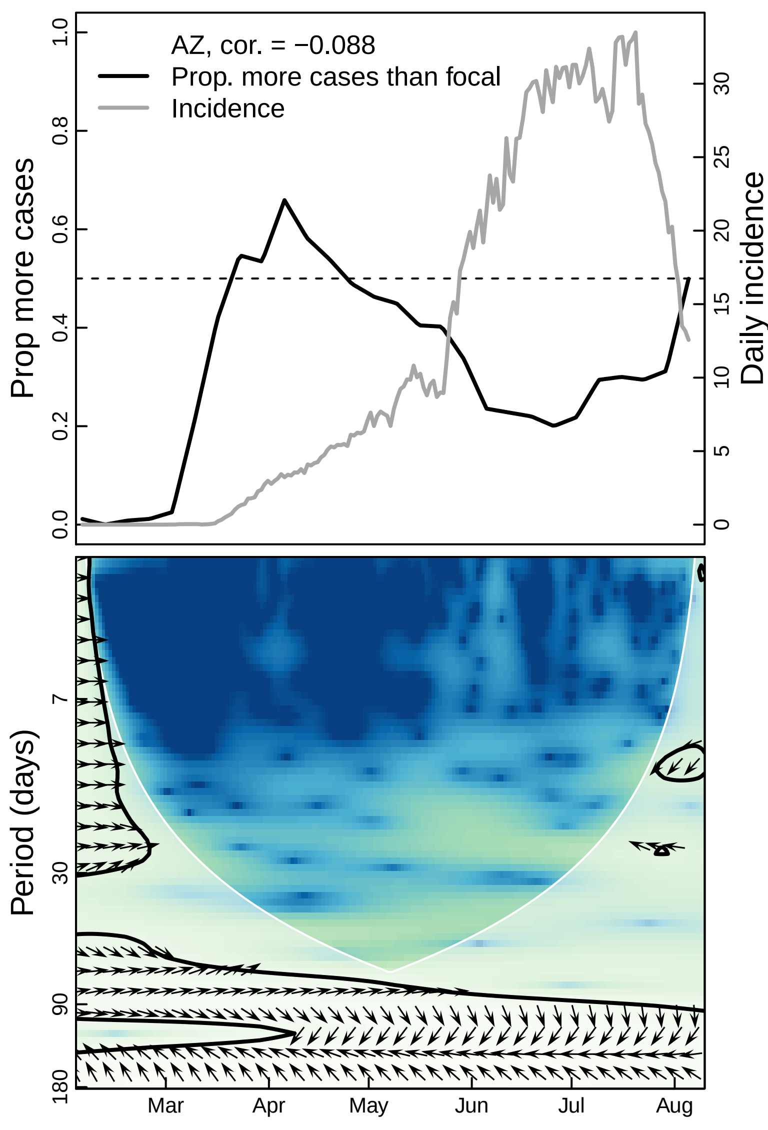

AZ, cor. = −0.088
Prop. more cases than focal
Incidence
Prop more cases
Daily incidence
0.0
0.2
0.4
0.6
0.8
1.0
0
5
10
15
20
25
30
Period (days)
7
30
90
180
Mar
Apr
May
Jun
Jul
Aug

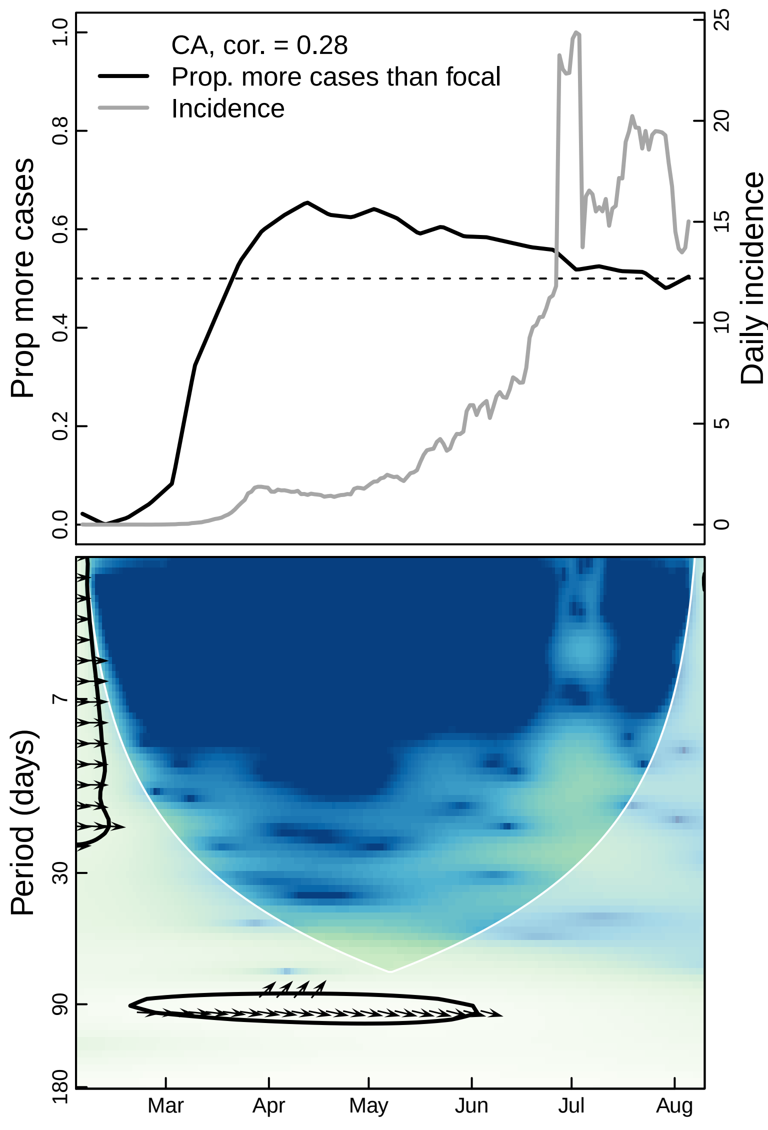

CA, cor. = 0.28
Prop. more cases than focal
Incidence
Prop more cases
Daily incidence
0.0
0.2
0.4
0.6
0.8
1.0
0
5
10
15
20
25
Period (days)
7
30
90
180
Mar
Apr
May
Jun
Jul
Aug

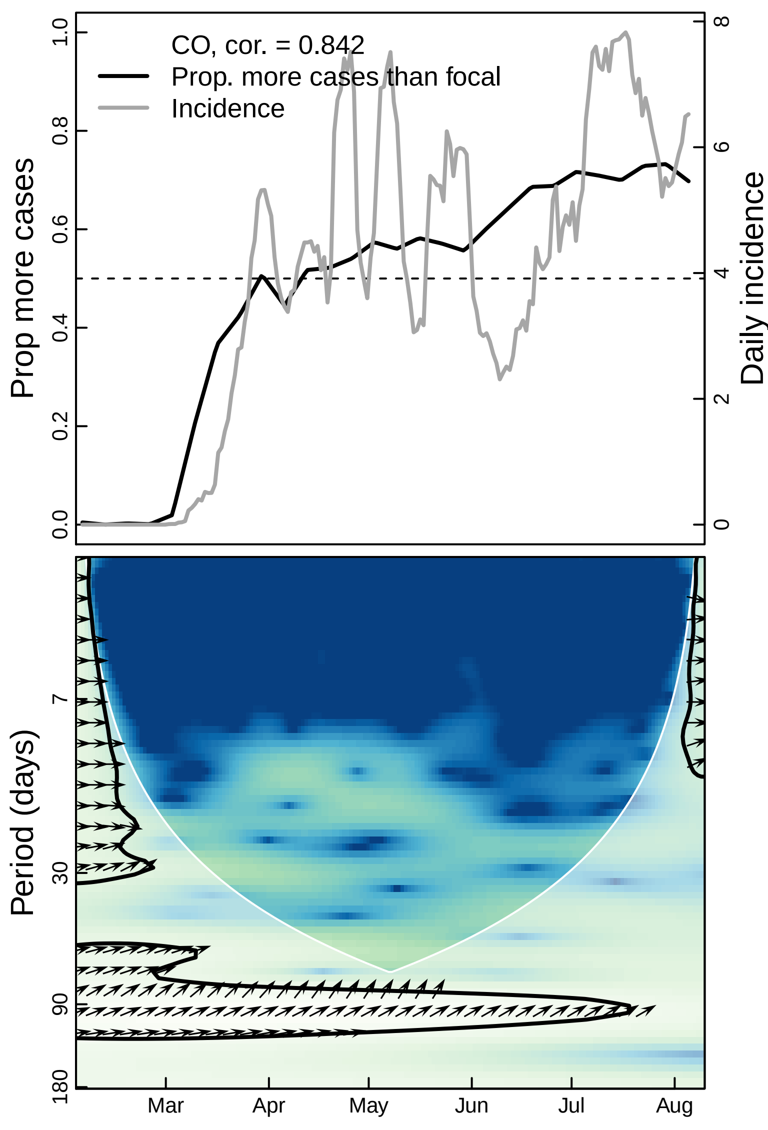

CO, cor. = 0.842
Prop. more cases than focal
Incidence
Prop more cases
Daily incidence
0.0
0.2
0.4
0.6
0.8
1.0
0
2
4
6
8
Period (days)
7
30
90
180
Mar
Apr
May
Jun
Jul
Aug

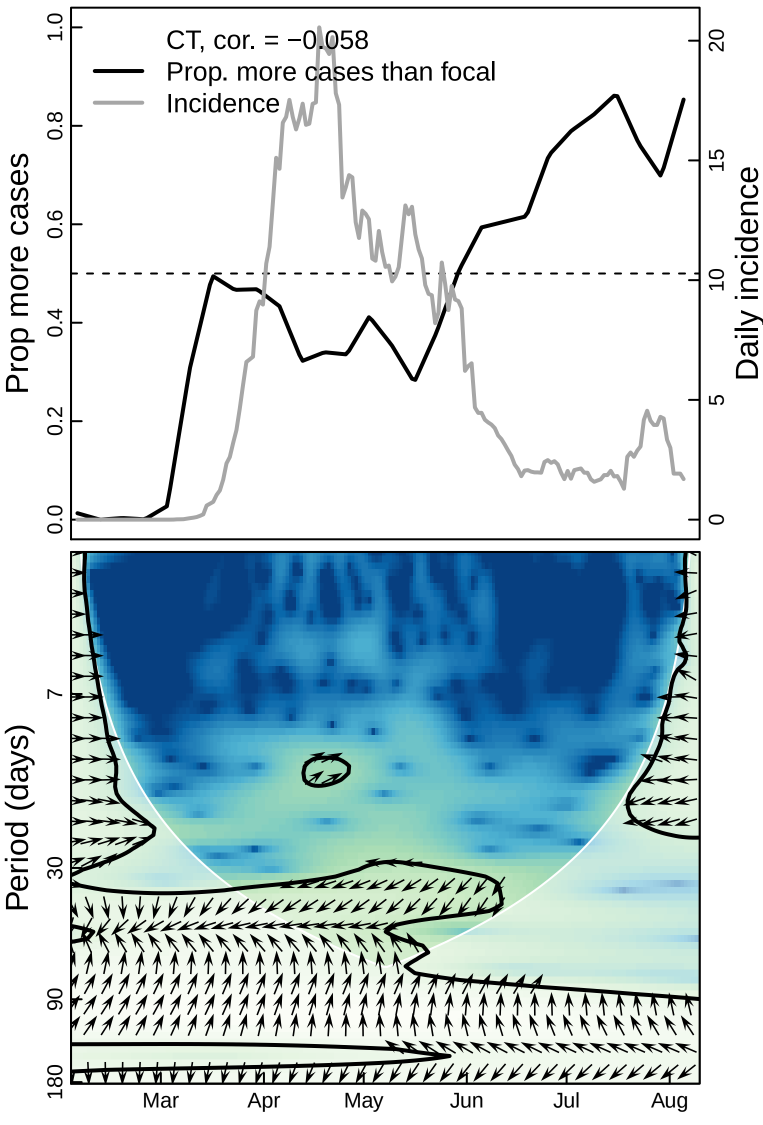
CT, cor. = −0.058
Prop. more cases than focal
Incidence
Prop more cases
Daily incidence
0.0
0.2
0.4
0.6
0.8
1.0
0
5
10
15
20
Period (days)
7
30
90
180
Mar
Apr
May
Jun
Jul
Aug

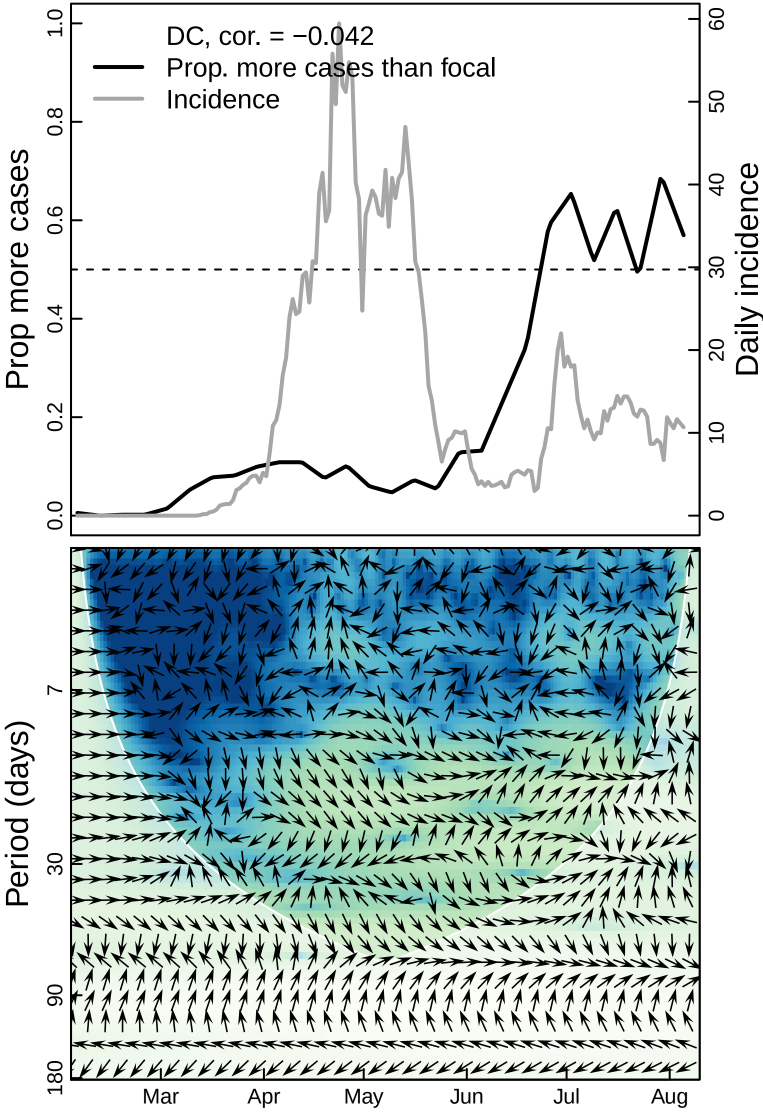

DC, cor. = −0.042
Prop. more cases than focal
Incidence
Prop more cases
Daily incidence
0.0
0.2
0.4
0.6
0.8
1.0
0
10
20
30
40
50
60
Period (days)
7
30
90
180
Mar
Apr
May
Jun
Jul
Aug

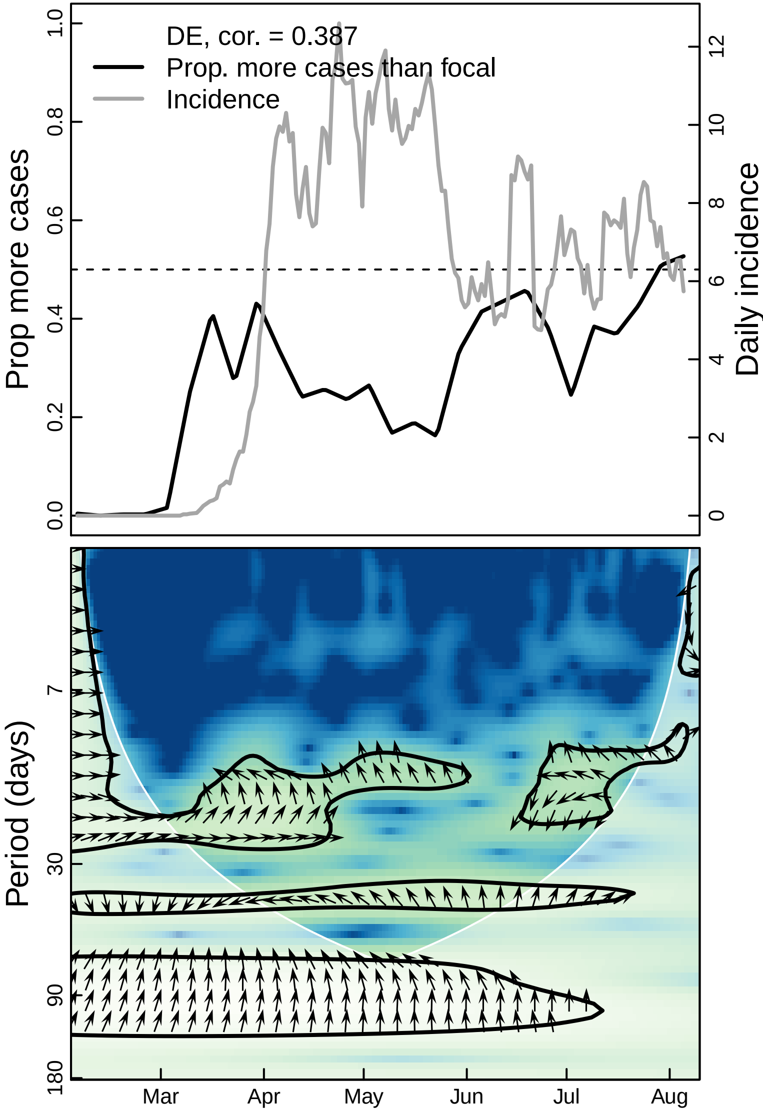

DE, cor. = 0.387
Prop. more cases than focal
Incidence
Prop more cases
Daily incidence
0.0
0.2
0.4
0.6
0.8
1.0
0
2
4
6
8
10
12
Period (days)
7
30
90
180
Mar
Apr
May
Jun
Jul
Aug

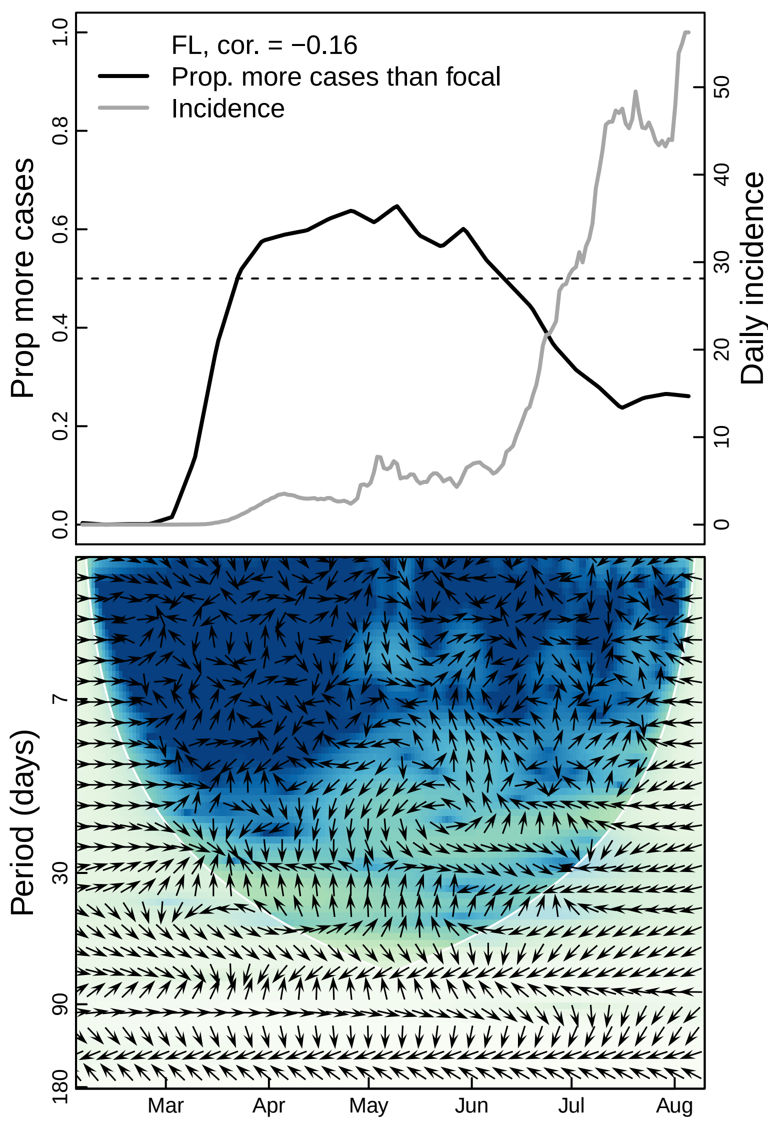
FL, cor. = −0.16
Prop. more cases than focal
Incidence
Prop more cases
Daily incidence
0.0
0.2
0.4
0.6
0.8
1.0
0
10
20
30
40
50
Period (days)
7
30
90
180
Mar
Apr
May
Jun
Jul
Aug

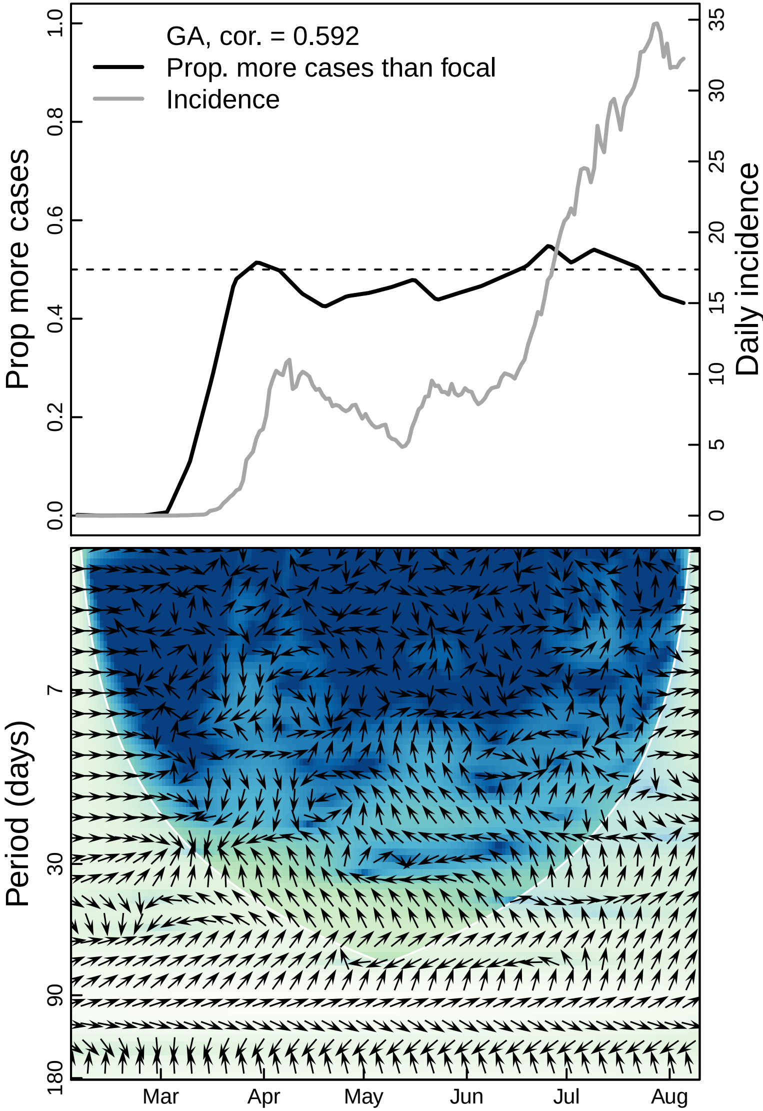

GA, cor. = 0.592
Prop. more cases than focal
Incidence
Prop more cases
Daily incidence
0.0
0.2
0.4
0.6
0.8
1.0
0
5
10
15
20
25
30
35
Period (days)
7
30
90
180
Mar
Apr
May
Jun
Jul
Aug

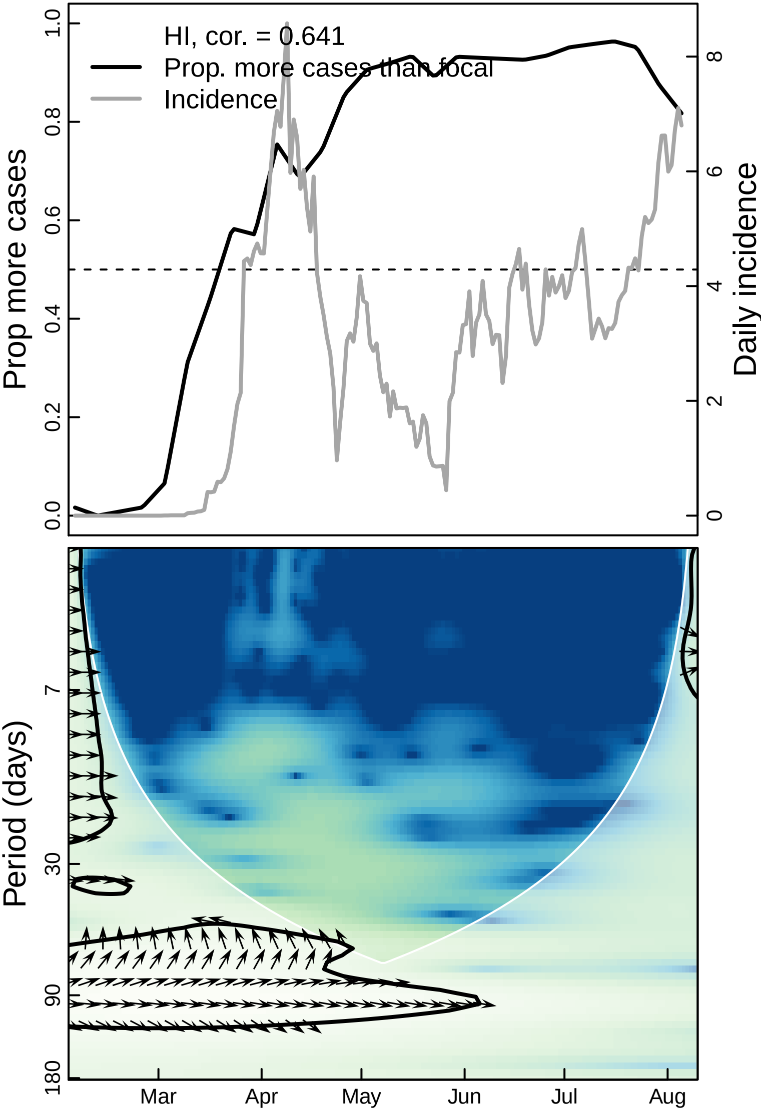

HI, cor. = 0.641
Prop. more cases than focal
Incidence
Prop more cases
Daily incidence
0.0
0.2
0.4
0.6
0.8
1.0
0
2
4
6
8
Period (days)
7
30
90
180
Mar
Apr
May
Jun
Jul
Aug

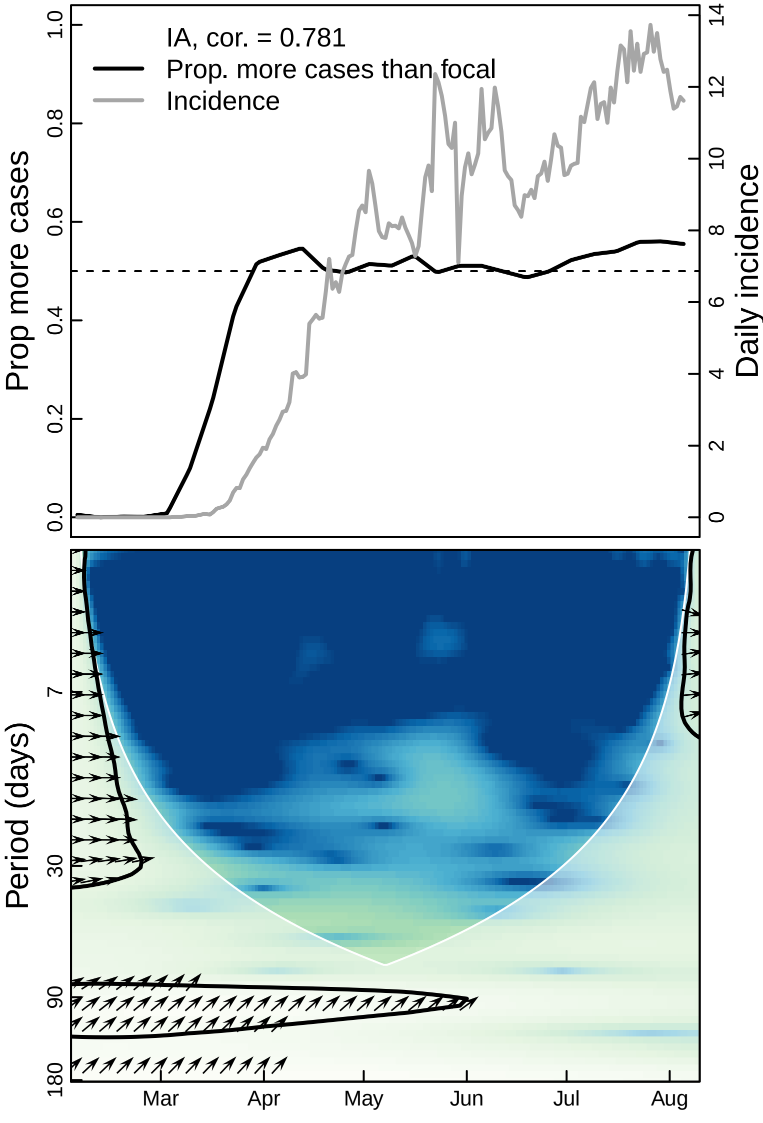

IA, cor. = 0.781
Prop. more cases than focal
Incidence
Prop more cases
Daily incidence
0.0
0.2
0.4
0.6
0.8
1.0
0
2
4
6
8
10
12
14
Period (days)
7
30
90
180
Mar
Apr
May
Jun
Jul
Aug

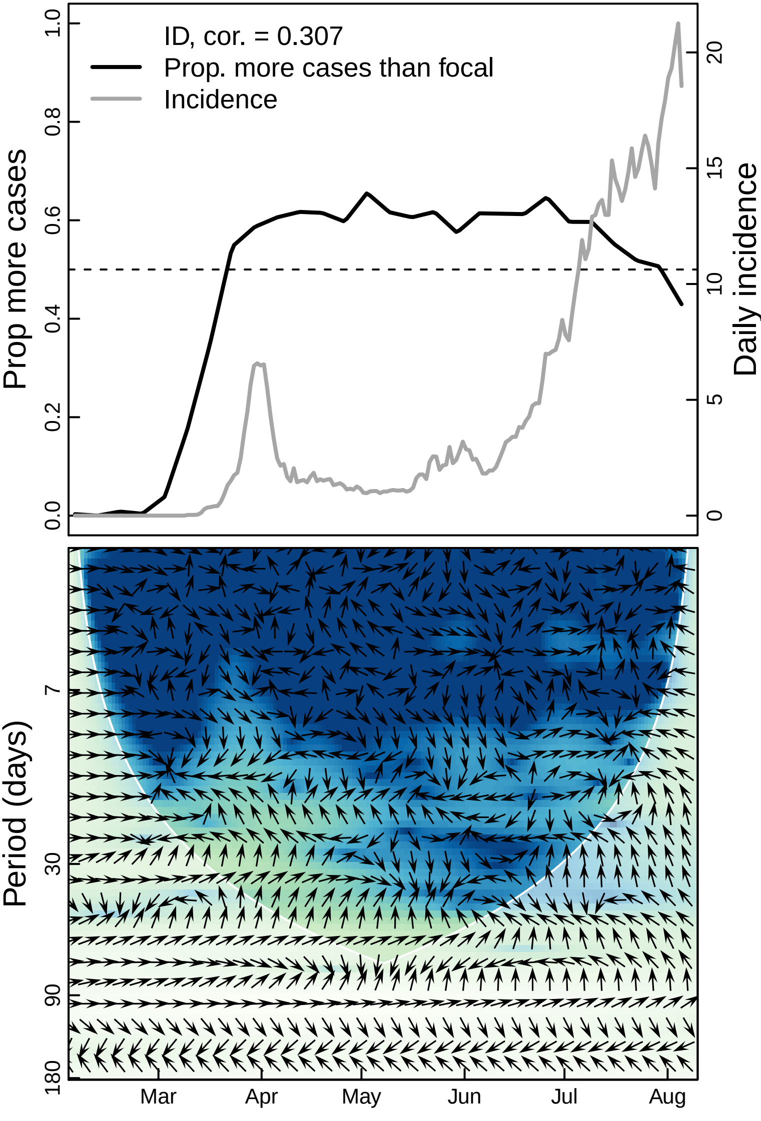

ID, cor. = 0.307
Prop. more cases than focal
Incidence
Prop more cases
Daily incidence
0.0
0.2
0.4
0.6
0.8
1.0
0
5
10
15
20
Period (days)
7
30
90
180
Mar
Apr
May
Jun
Jul
Aug

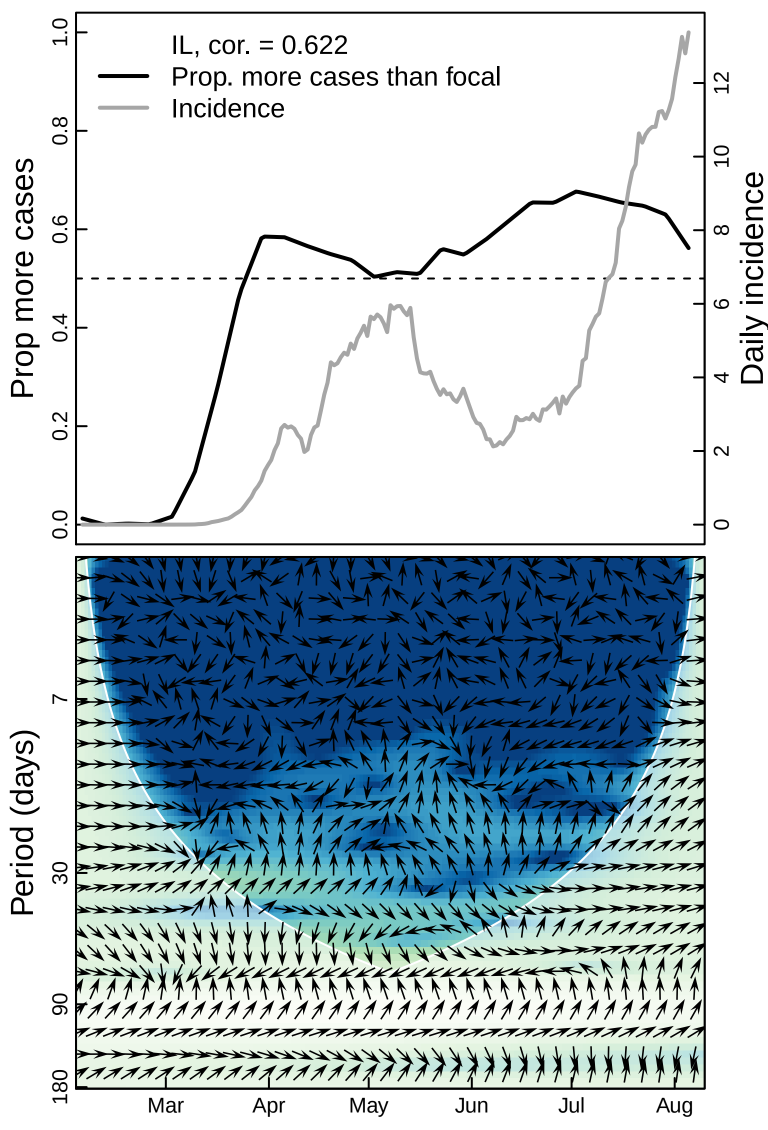

IL, cor. = 0.622
Prop. more cases than focal
Incidence
Prop more cases
Daily incidence
0.0
0.2
0.4
0.6
0.8
1.0
0
2
4
6
8
10
12
Period (days)
7
30
90
180
Mar
Apr
May
Jun
Jul
Aug

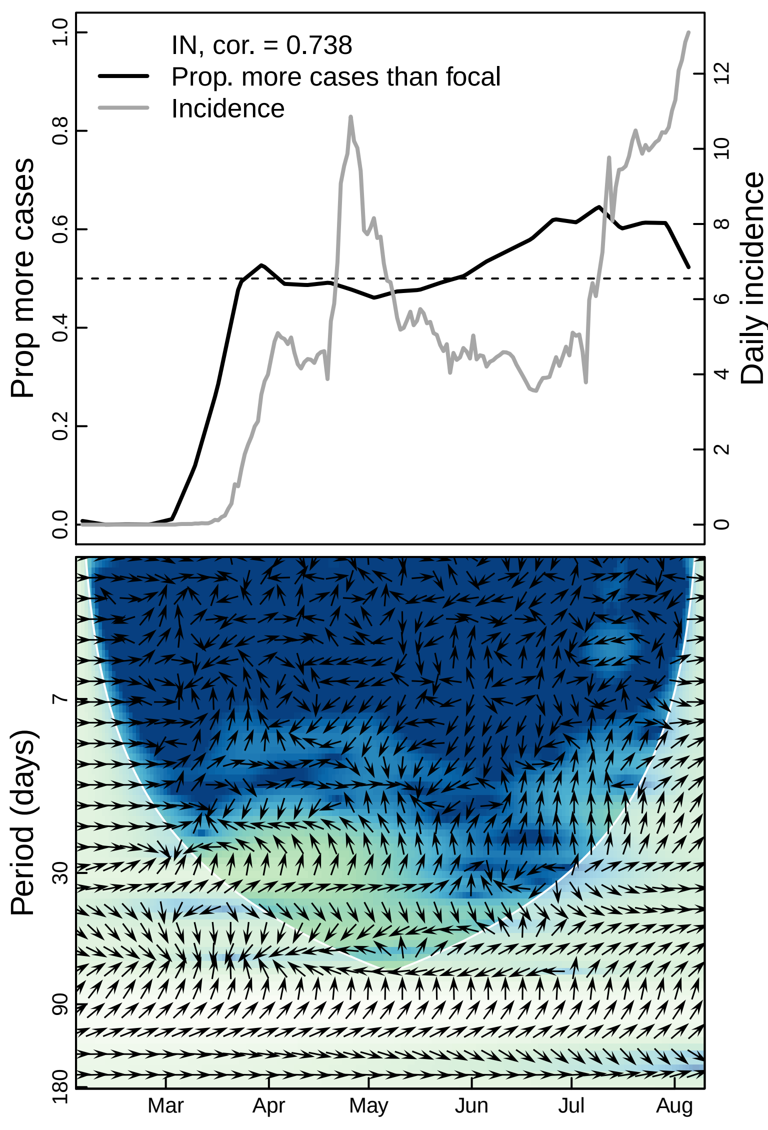

IN, cor. = 0.738
Prop. more cases than focal
Incidence
Prop more cases
Daily incidence
0.0
0.2
0.4
0.6
0.8
1.0
0
2
4
6
8
10
12
Period (days)
7
30
90
180
Mar
Apr
May
Jun
Jul
Aug

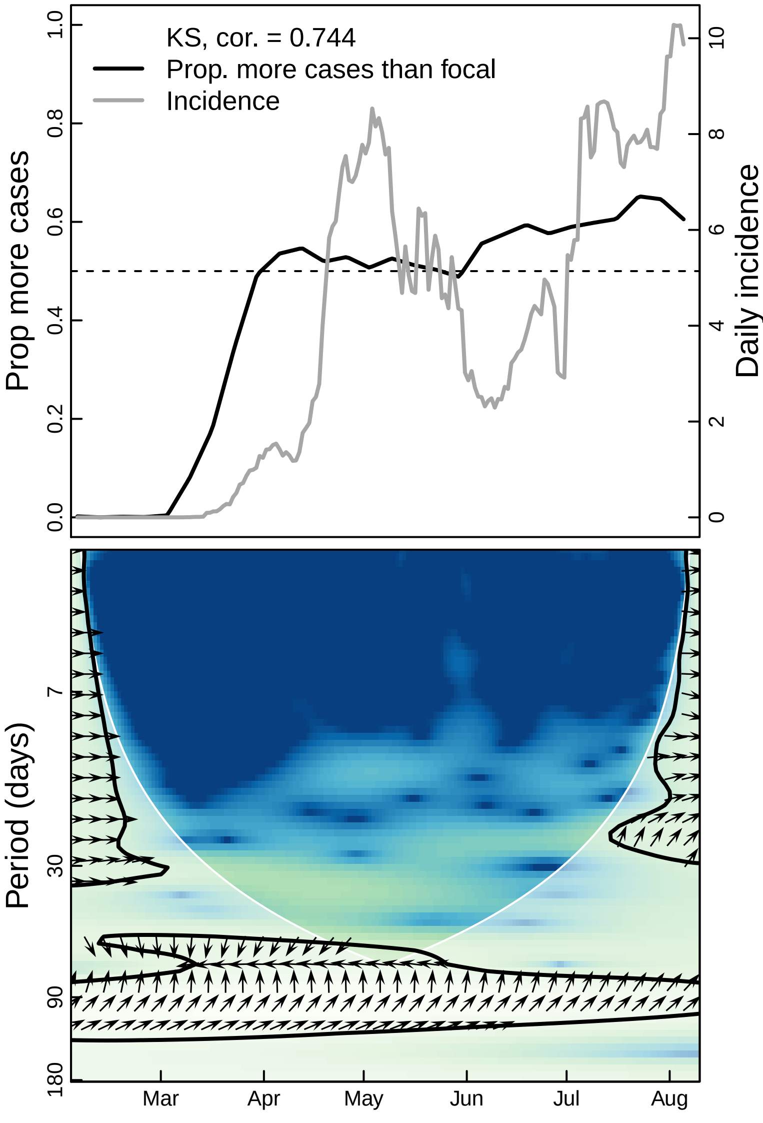

KS, cor. = 0.744
Prop. more cases than focal
Incidence
Prop more cases
Daily incidence
0.0
0.2
0.4
0.6
0.8
1.0
0
2
4
6
8
10
Period (days)
7
30
90
180
Mar
Apr
May
Jun
Jul
Aug

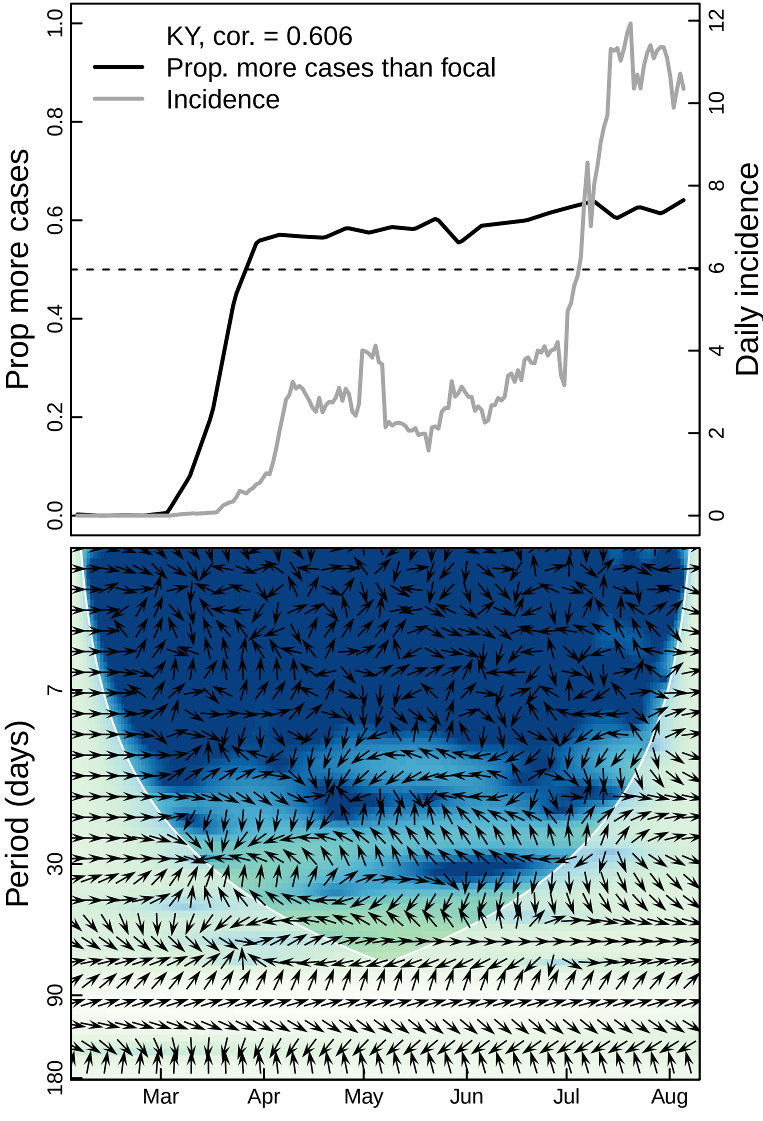
KY, cor. = 0.606
Prop. more cases than focal
Incidence
Prop more cases
Daily incidence
0.0
0.2
0.4
0.6
0.8
1.0
0
2
4
6
8
10
12
Period (days)
7
30
90
180
Mar
Apr
May
Jun
Jul
Aug

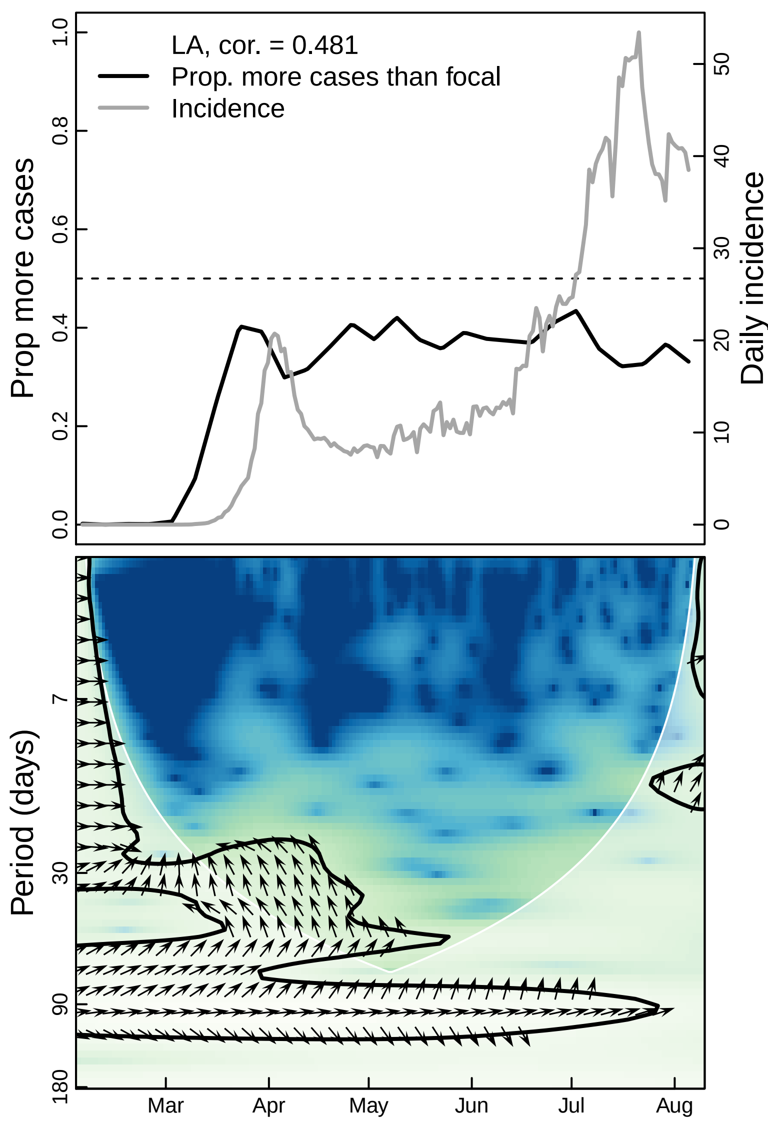
LA, cor. = 0.481
Prop. more cases than focal
Incidence
Prop more cases
Daily incidence
0.0
0.2
0.4
0.6
0.8
1.0
0
10
20
30
40
50
Period (days)
7
30
90
180
Mar
Apr
May
Jun
Jul
Aug

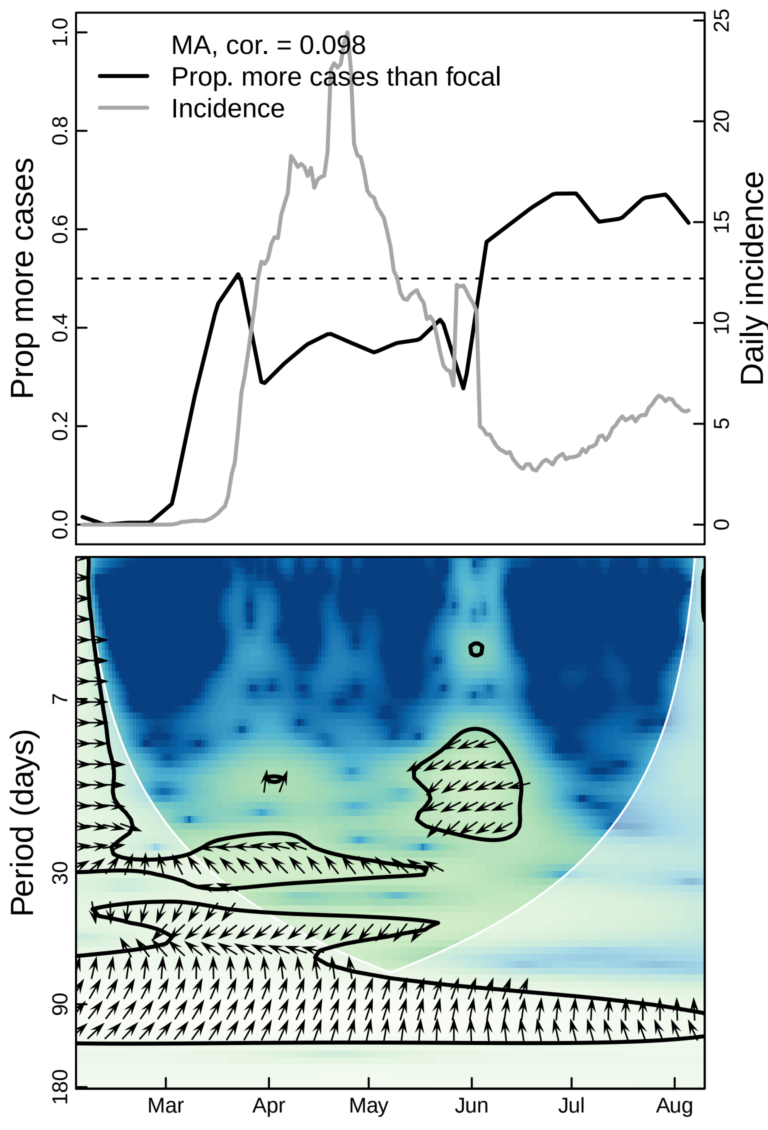

MA, cor. = 0.098
Prop. more cases than focal
Incidence
Prop more cases
Daily incidence
0.0
0.2
0.4
0.6
0.8
1.0
0
5
10
15
20
25
Period (days)
7
30
90
180
Mar
Apr
May
Jun
Jul
Aug

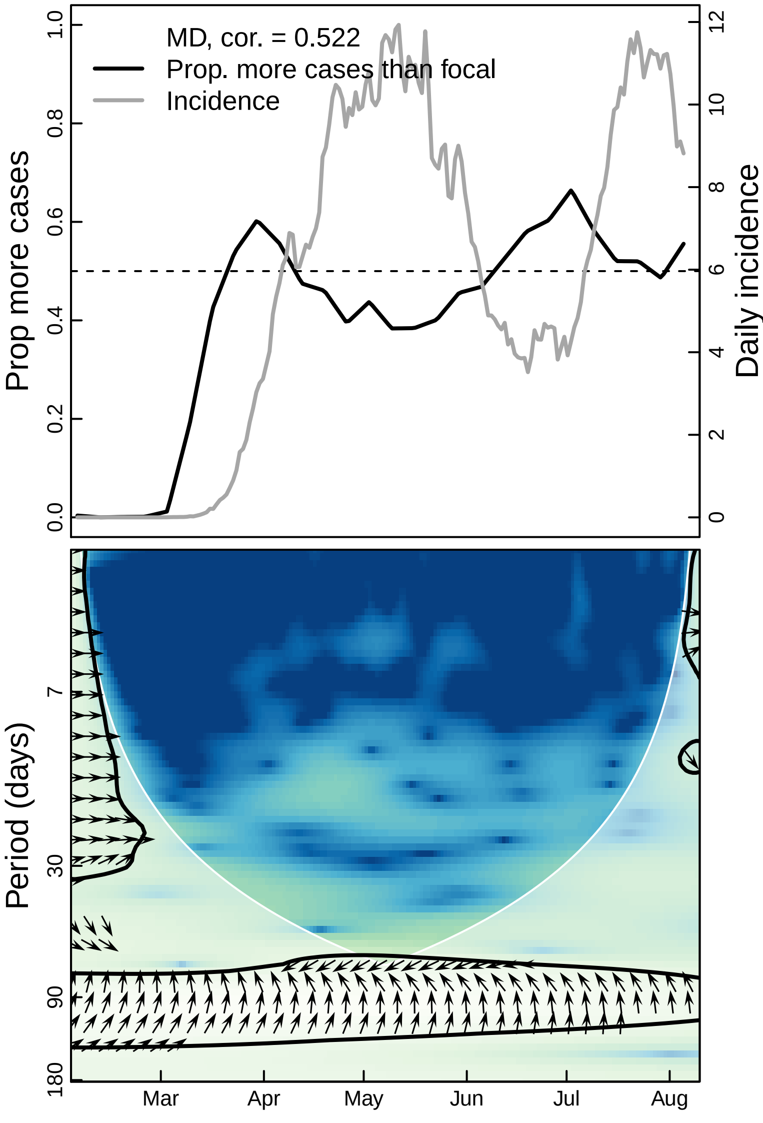

MD, cor. = 0.522
Prop. more cases than focal
Incidence
Prop more cases
Daily incidence
0.0
0.2
0.4
0.6
0.8
1.0
0
2
4
6
8
10
12
Period (days)
7
30
90
180
Mar
Apr
May
Jun
Jul
Aug

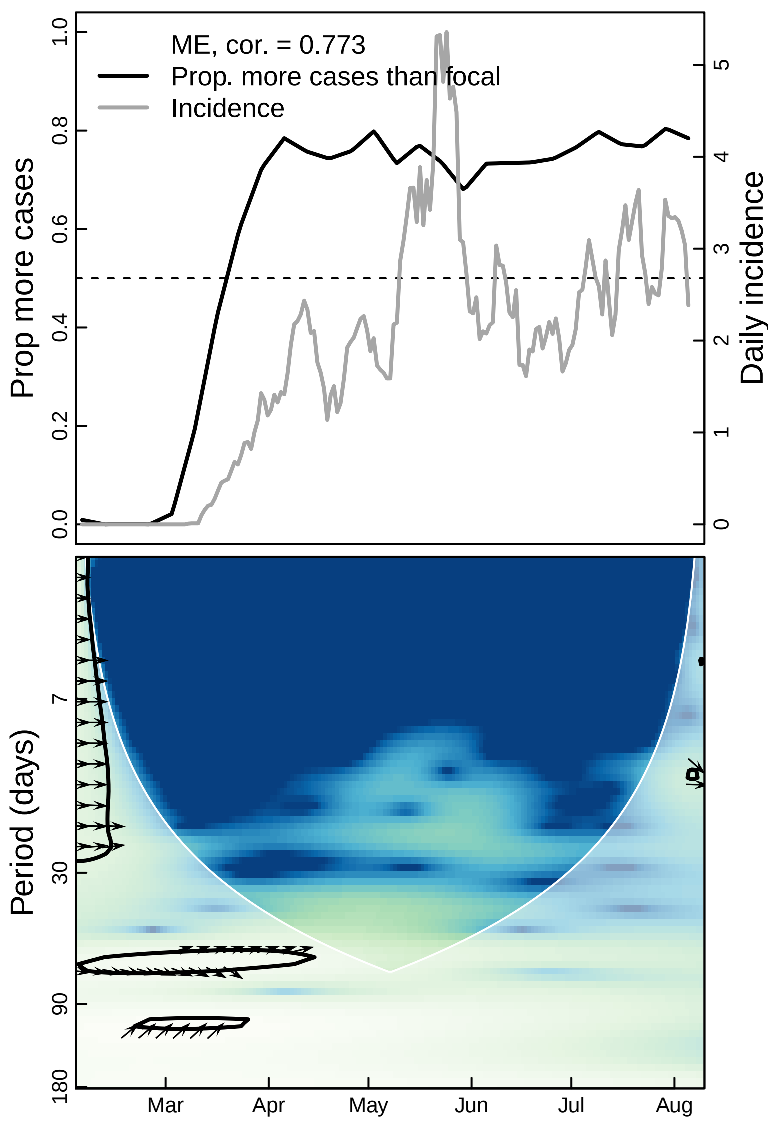

ME, cor. = 0.773
Prop. more cases than focal
Incidence
Prop more cases
Daily incidence
0.0
0.2
0.4
0.6
0.8
1.0
0
1
2
3
4
5
Period (days)
7
30
90
180
Mar
Apr
May
Jun
Jul
Aug

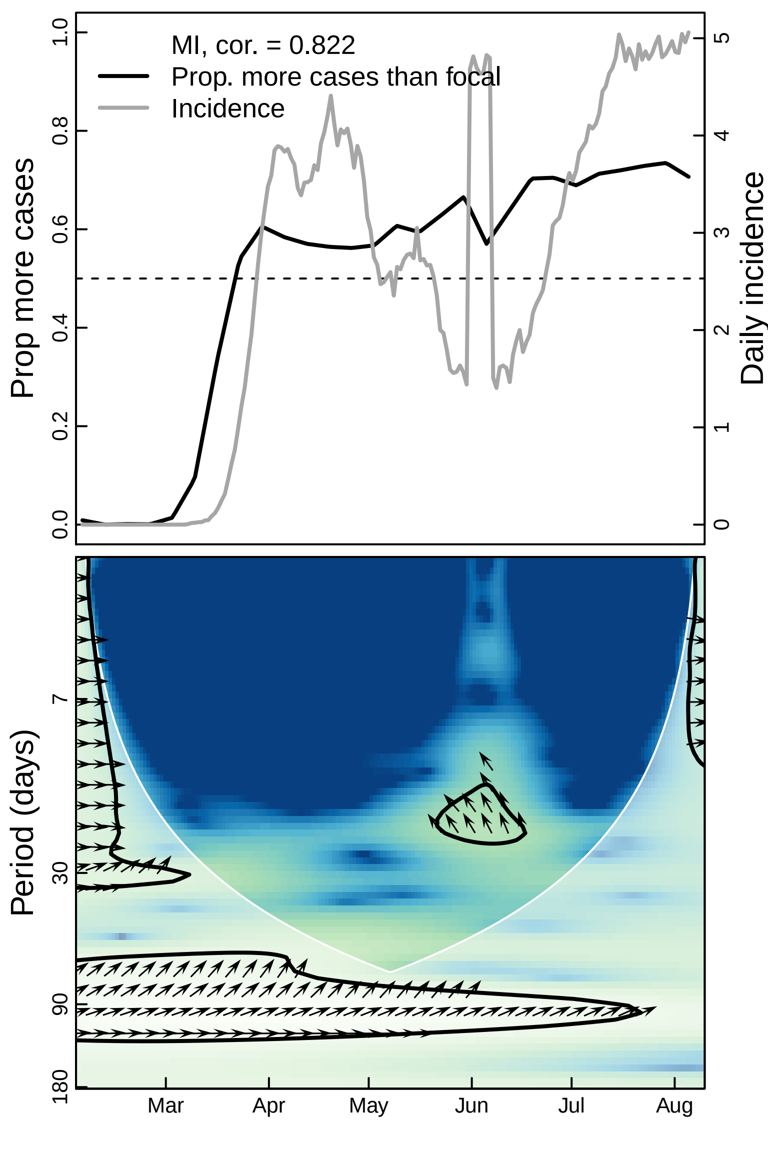

MI, cor. = 0.822
Prop. more cases than focal
Incidence
Prop more cases
Daily incidence
0.0
0.2
0.4
0.6
0.8
1.0
0
1
2
3
4
5
Period (days)
7
30
90
180
Mar
Apr
May
Jun
Jul
Aug

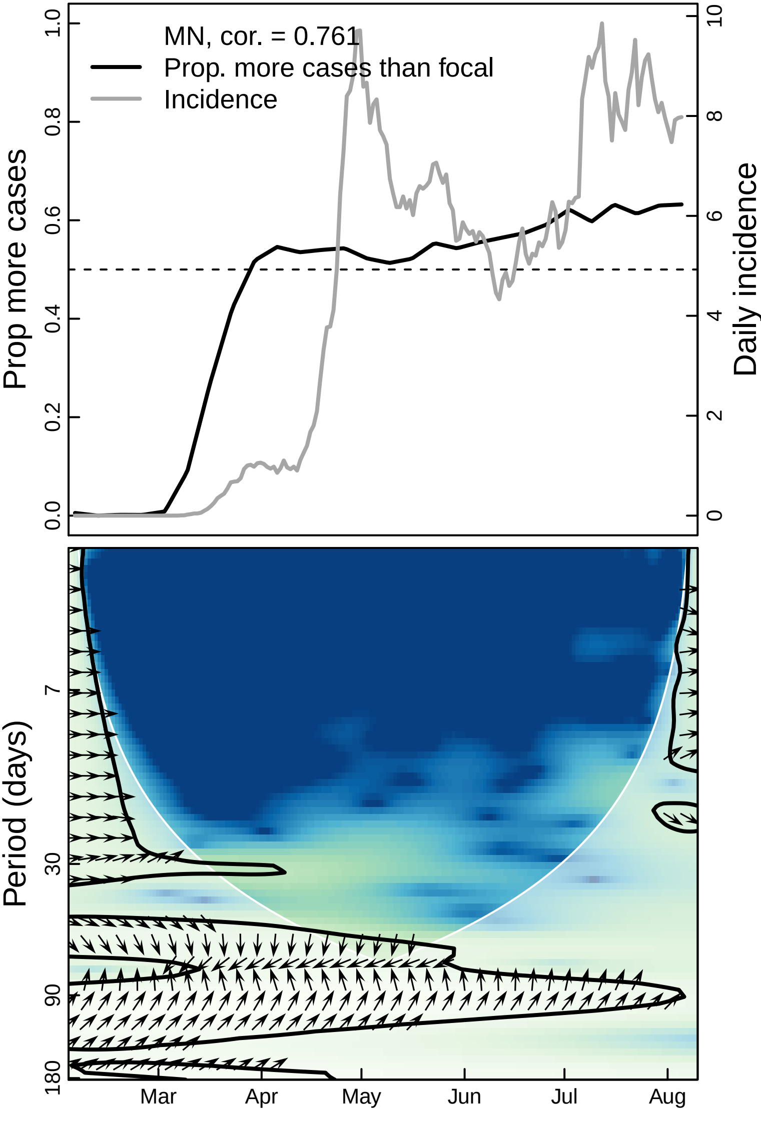

MN, cor. = 0.761
Prop. more cases than focal
Incidence
Prop more cases
Daily incidence
0.0
0.2
0.4
0.6
0.8
1.0
0
2
4
6
8
10
Period (days)
7
30
90
180
Mar
Apr
May
Jun
Jul
Aug

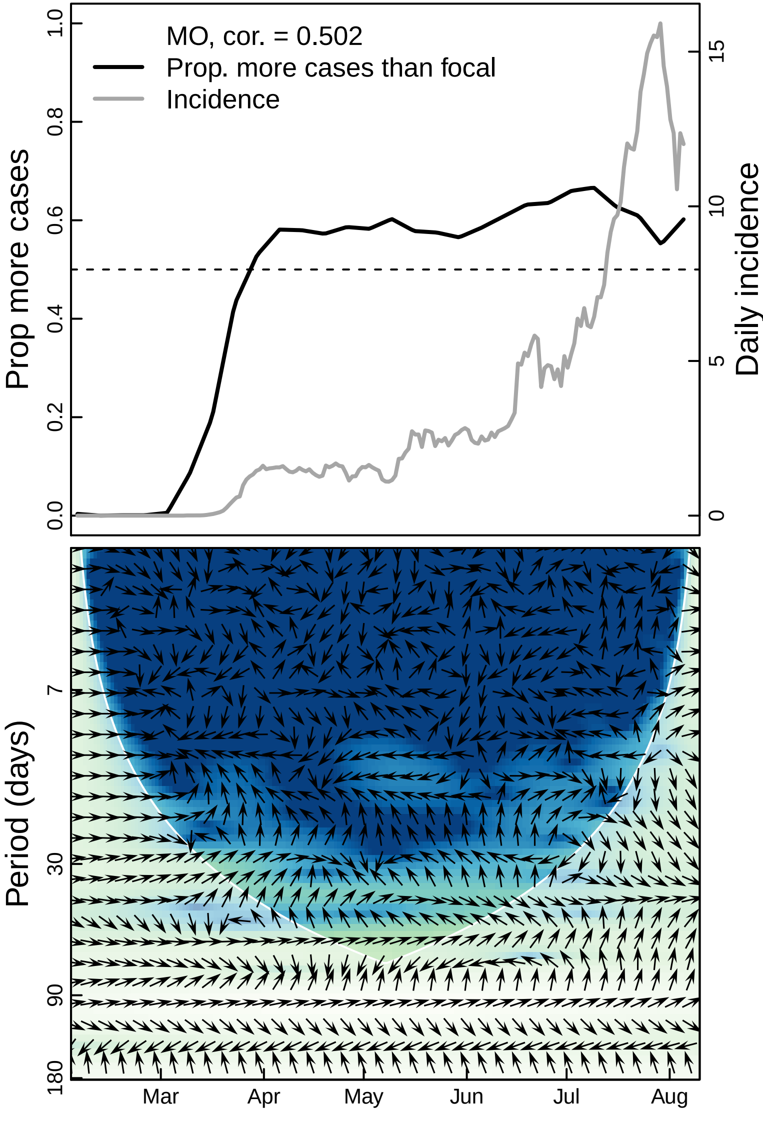

MO, cor. = 0.502
Prop. more cases than focal
Incidence
Prop more cases
Daily incidence
0.0
0.2
0.4
0.6
0.8
1.0
0
5
10
15
Period (days)
7
30
90
180
Mar
Apr
May
Jun
Jul
Aug

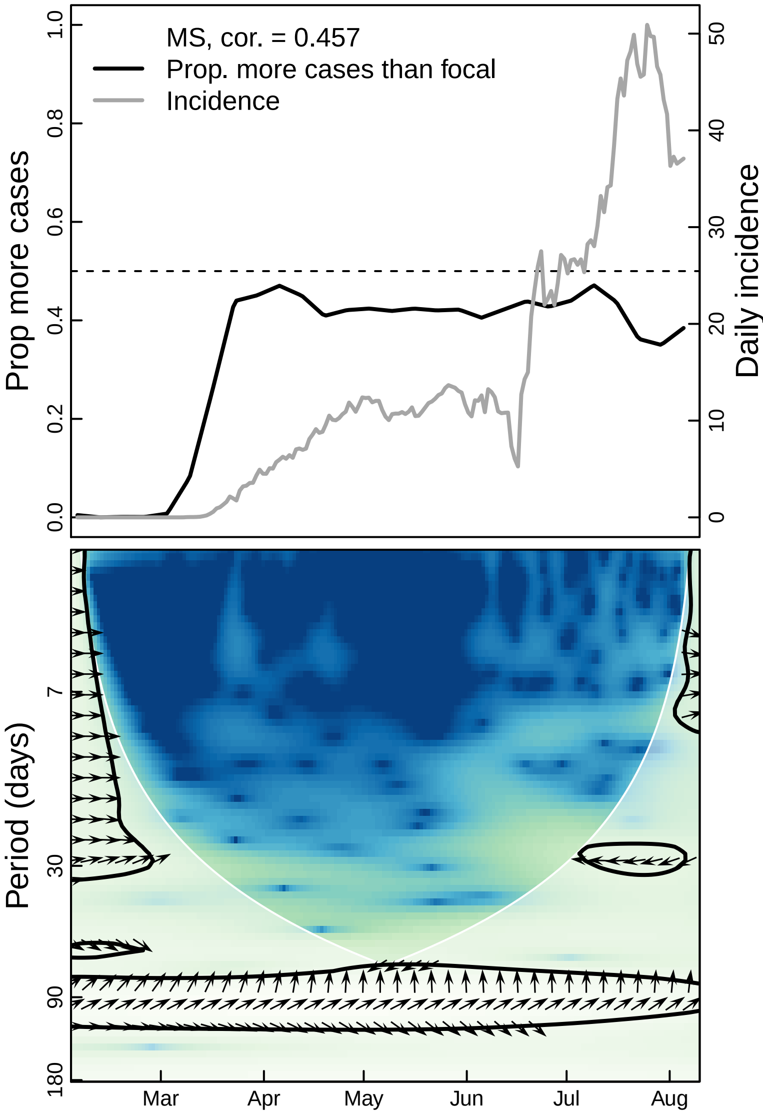

MS, cor. = 0.457
Prop. more cases than focal
Incidence
Prop more cases
Daily incidence
0.0
0.2
0.4
0.6
0.8
1.0
0
10
20
30
40
50
Period (days)
7
30
90
180
Mar
Apr
May
Jun
Jul
Aug

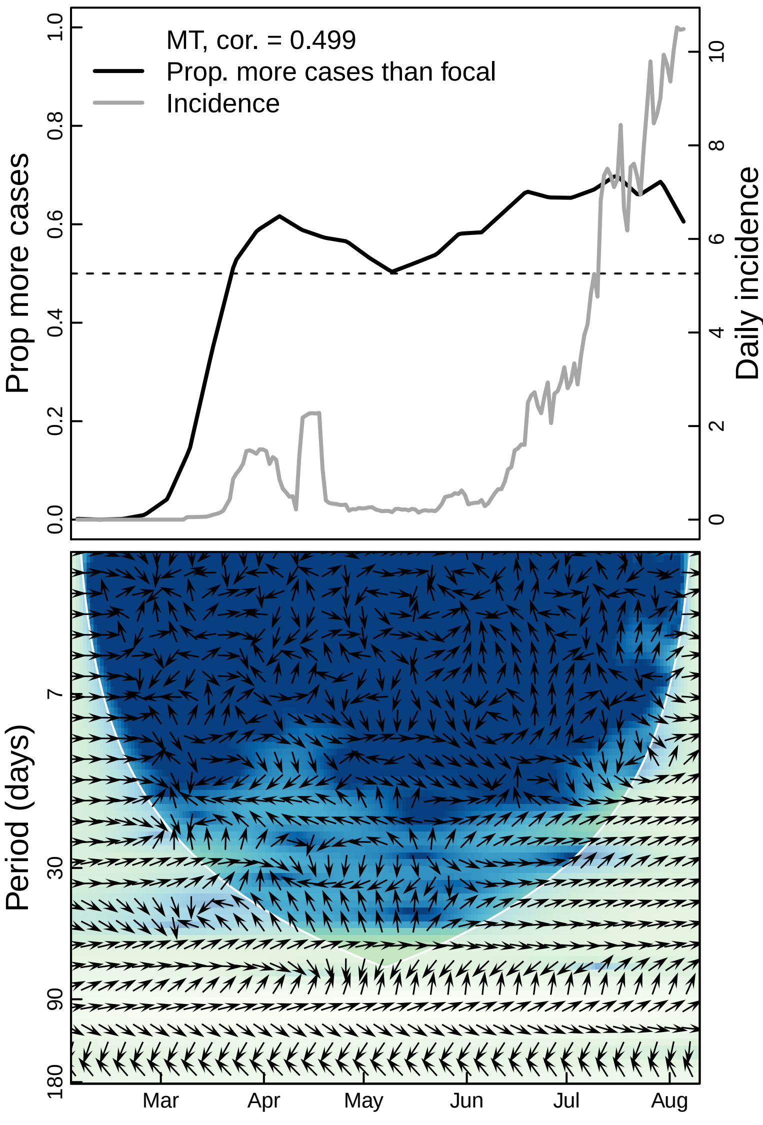

MT, cor. = 0.499
Prop. more cases than focal
Incidence
Prop more cases
Daily incidence
0.0
0.2
0.4
0.6
0.8
1.0
0
2
4
6
8
10
Period (days)
7
30
90
180
Mar
Apr
May
Jun
Jul
Aug

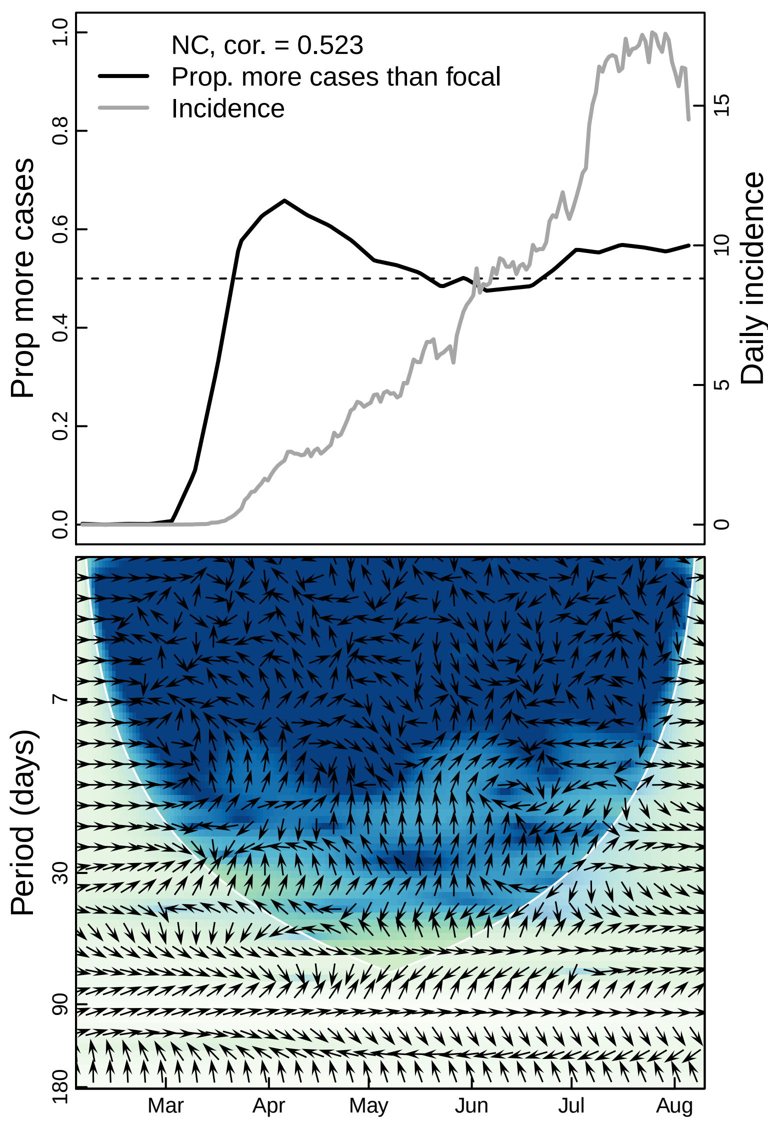

NC, cor. = 0.523
Prop. more cases than focal
Incidence
Prop more cases
Daily incidence
0.0
0.2
0.4
0.6
0.8
1.0
0
5
10
15
Period (days)
7
30
90
180
Mar
Apr
May
Jun
Jul
Aug

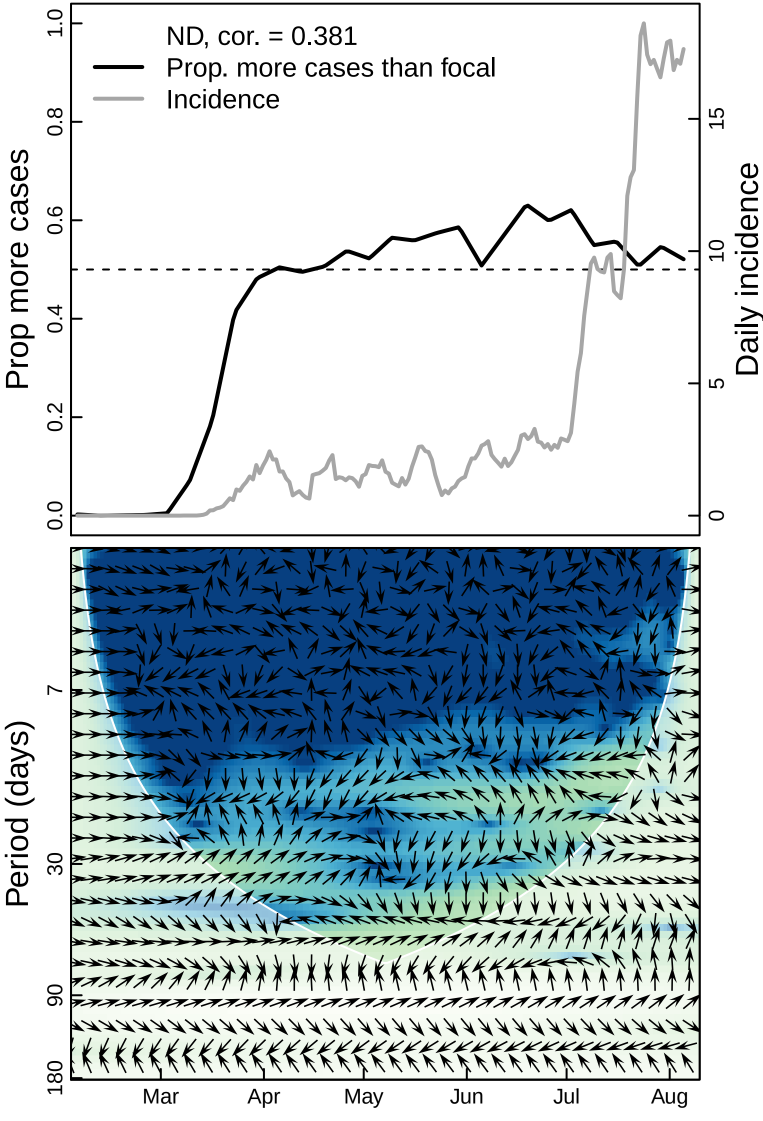
ND, cor. = 0.381
Prop. more cases than focal
Incidence
Prop more cases
Daily incidence
0.0
0.2
0.4
0.6
0.8
1.0
0
5
10
15
Period (days)
7
30
90
180
Mar
Apr
May
Jun
Jul
Aug

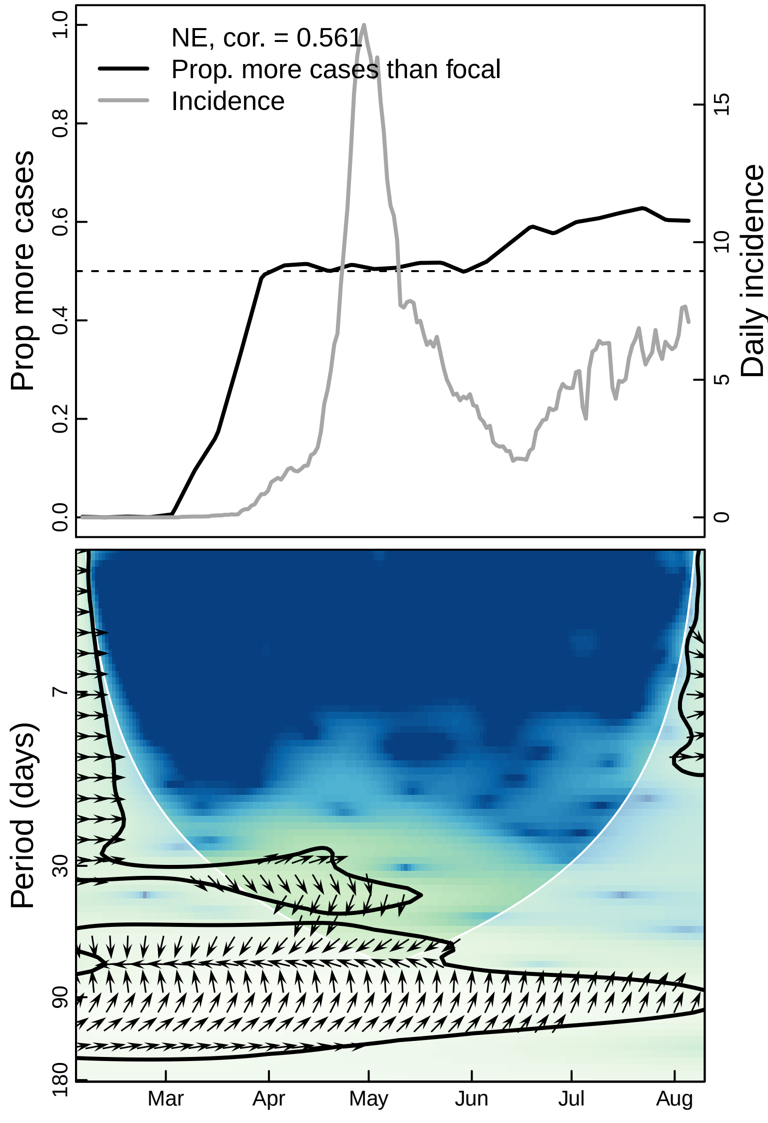

NE, cor. = 0.561
Prop. more cases than focal
Incidence
Prop more cases
Daily incidence
0.0
0.2
0.4
0.6
0.8
1.0
0
5
10
15
Period (days)
7
30
90
180
Mar
Apr
May
Jun
Jul
Aug

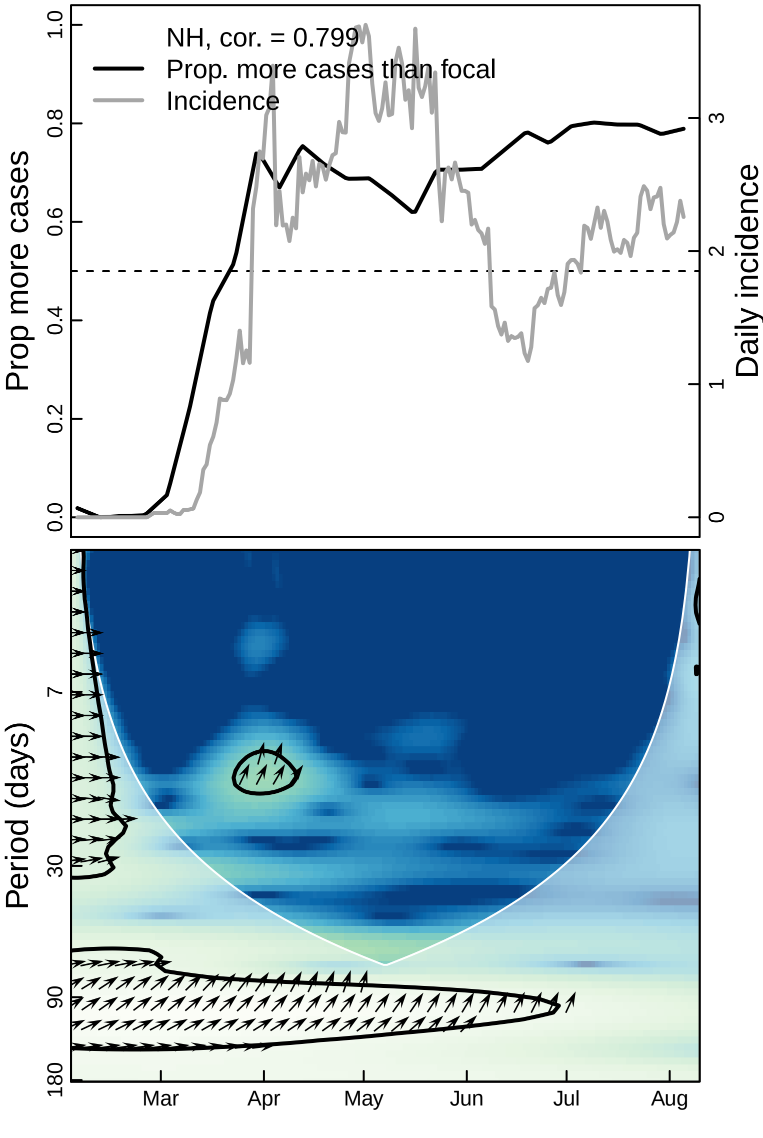
NH, cor. = 0.799
Prop. more cases than focal
Incidence
Prop more cases
Daily incidence
Period (days)
0.0
0.2
0.4
0.6
0.8
1.0
0
1
2
3
7
30
90
180
Mar
Apr
May
Jun
Jul
Aug

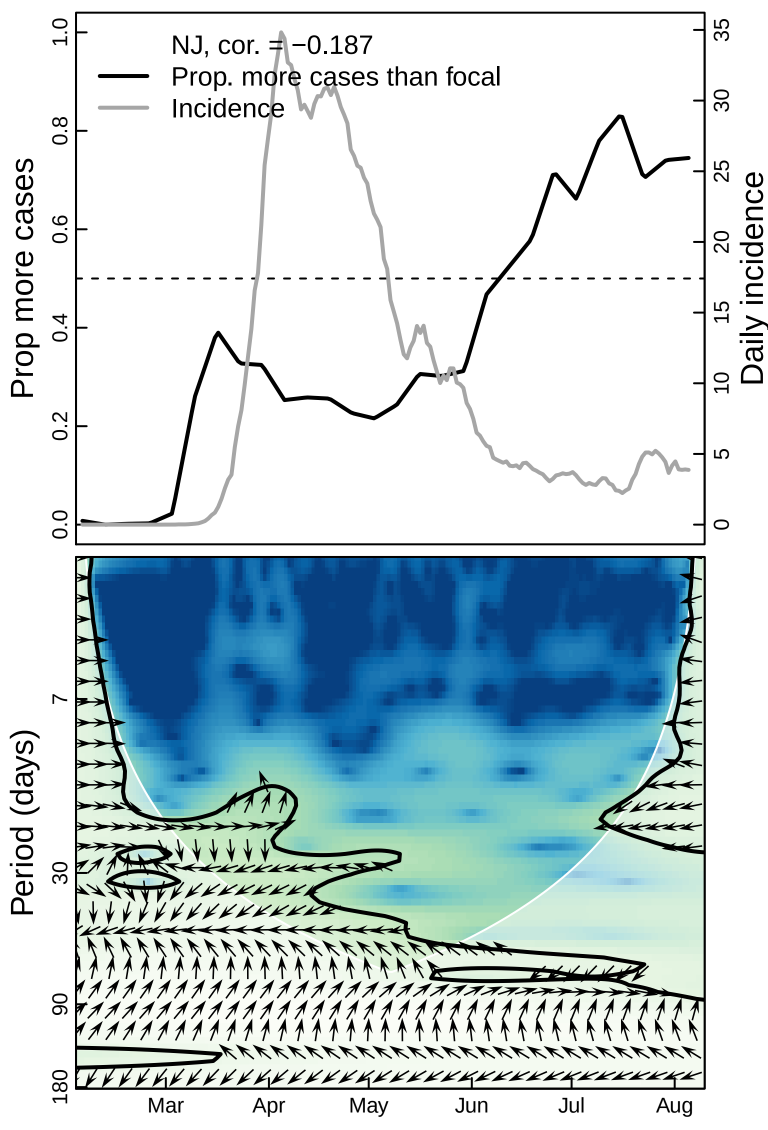

NJ, cor. = −0.187
Prop. more cases than focal
Incidence
Prop more cases
Daily incidence
Period (days)
Mar
Apr
May
Jun
Jul
Aug

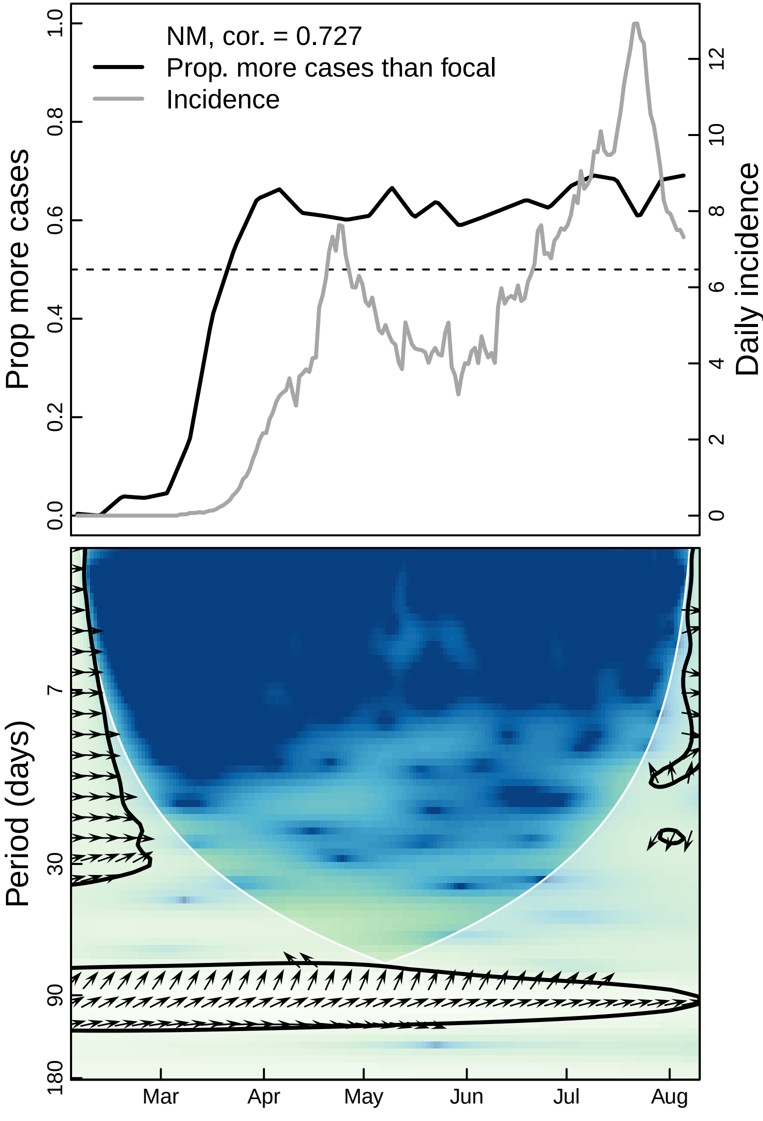

NM, cor. = 0.727
Prop. more cases than focal
Incidence
Prop more cases
Daily incidence
0.0
0.2
0.4
0.6
0.8
1.0
0
2
4
6
8
10
12
Period (days)
7
30
90
180
Mar
Apr
May
Jun
Jul
Aug

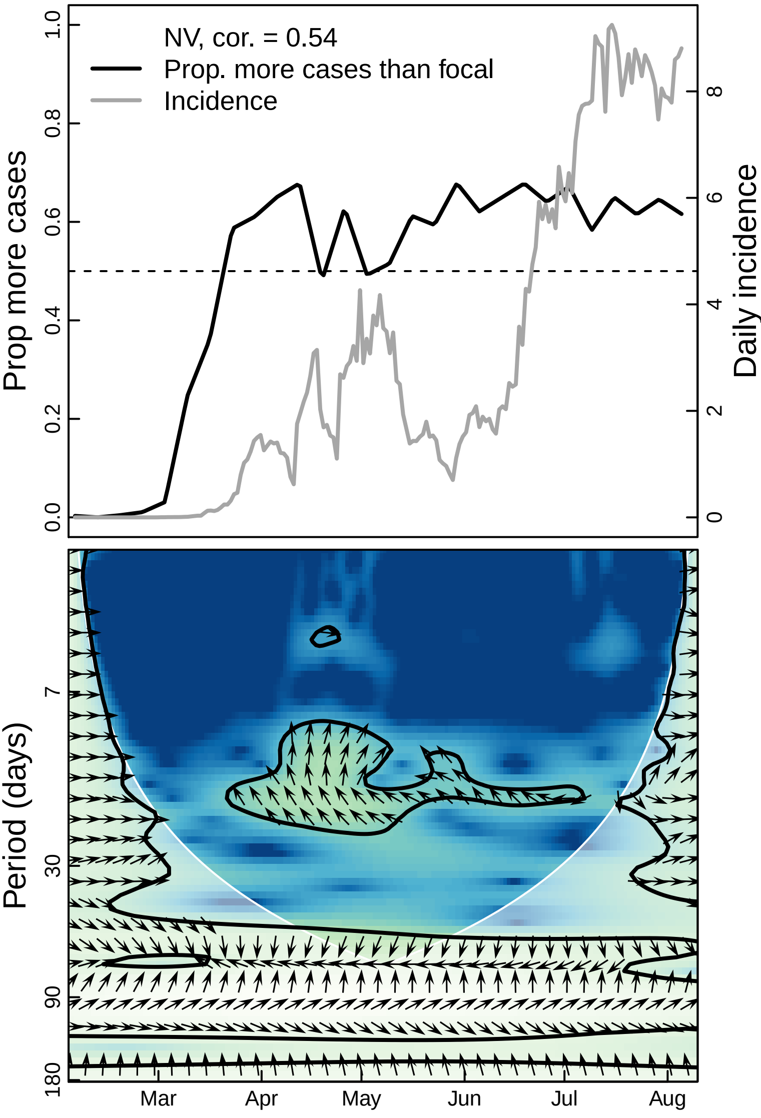

NV, cor. = 0.54
Prop. more cases than focal
Incidence
Prop more cases
Daily incidence
0.0
0.2
0.4
0.6
0.8
1.0
0
2
4
6
8
Period (days)
7
30
90
180
Mar
Apr
May
Jun
Jul
Aug

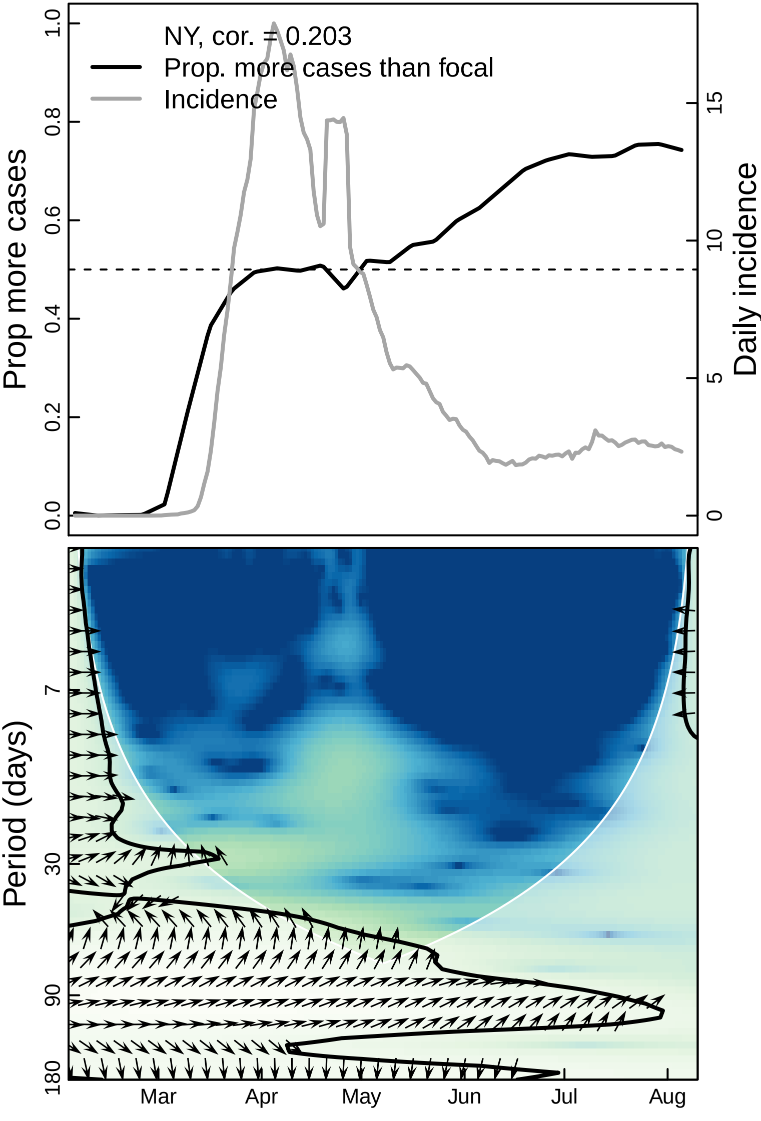
NY, cor. = 0.203
Prop. more cases than focal
Incidence
Prop more cases
Daily incidence
0.0
0.2
0.4
0.6
0.8
1.0
0
5
10
15
Period (days)
7
30
90
180
Mar
Apr
May
Jun
Jul
Aug

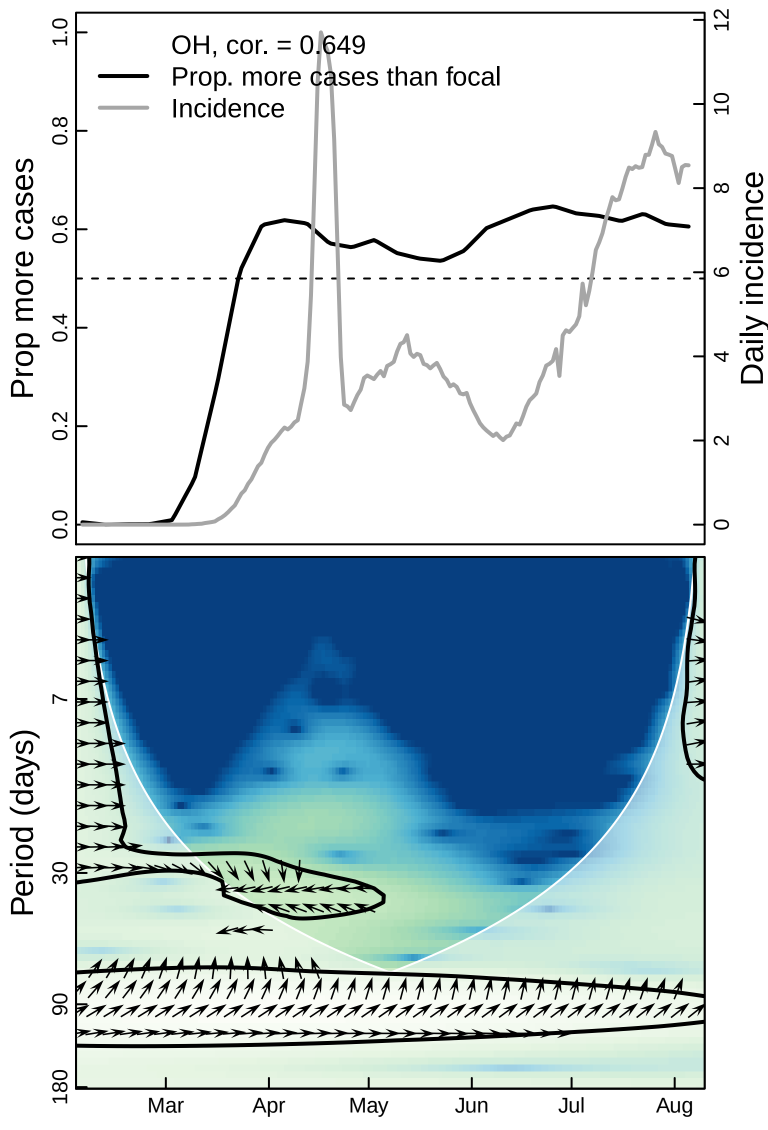

OH, cor. = 0.649
Prop. more cases than focal
Incidence
Prop more cases
Daily incidence
0.0
0.2
0.4
0.6
0.8
1.0
0
2
4
6
8
10
12
Period (days)
7
30
90
180
Mar
Apr
May
Jun
Jul
Aug

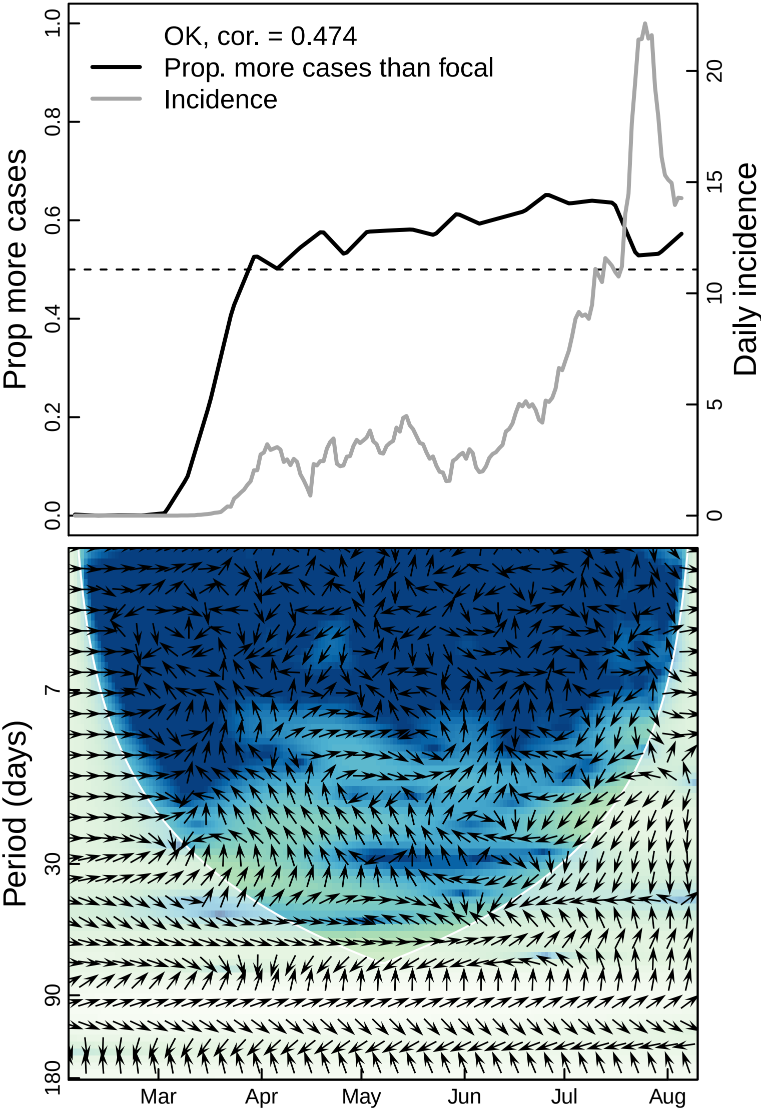
OK, cor. = 0.474
Prop. more cases than focal
Incidence
Prop more cases
Daily incidence
0.0
0.2
0.4
0.6
0.8
1.0
0
5
10
15
20
Period (days)
7
30
90
180
Mar
Apr
May
Jun
Jul
Aug

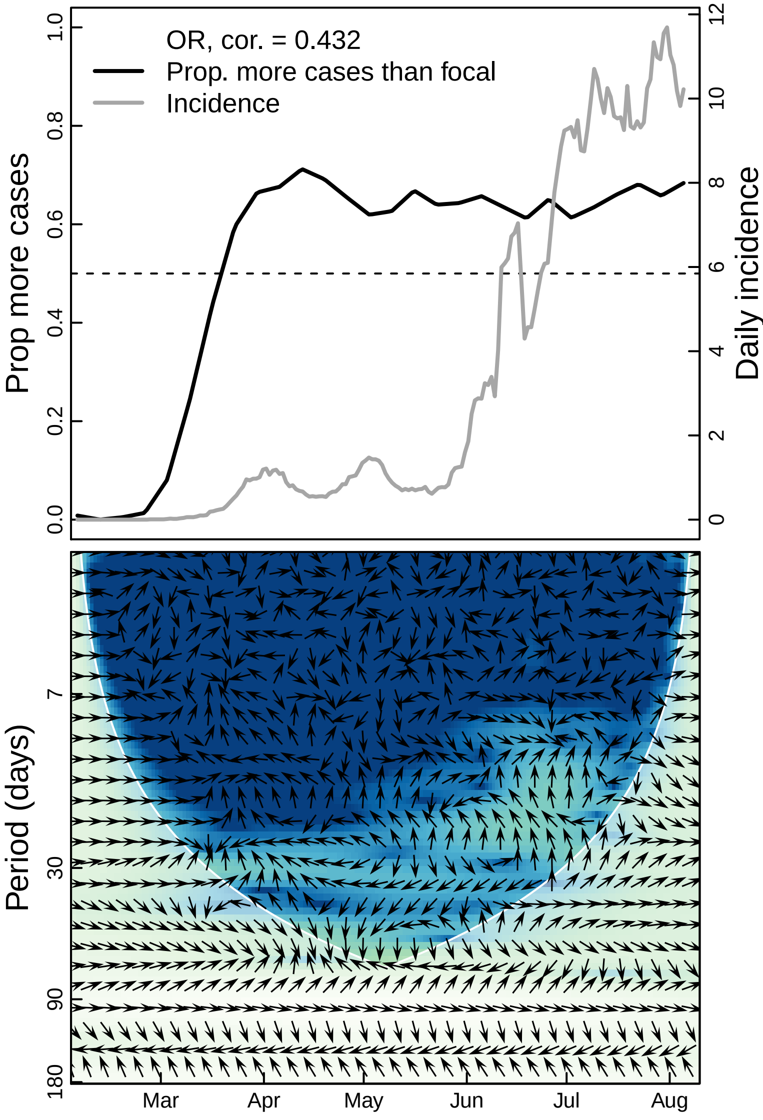

OR, cor. = 0.432
Prop. more cases than focal
Incidence
Prop more cases
Daily incidence
0.0
0.2
0.4
0.6
0.8
1.0
0
2
4
6
8
10
12
Period (days)
7
30
90
180
Mar
Apr
May
Jun
Jul
Aug

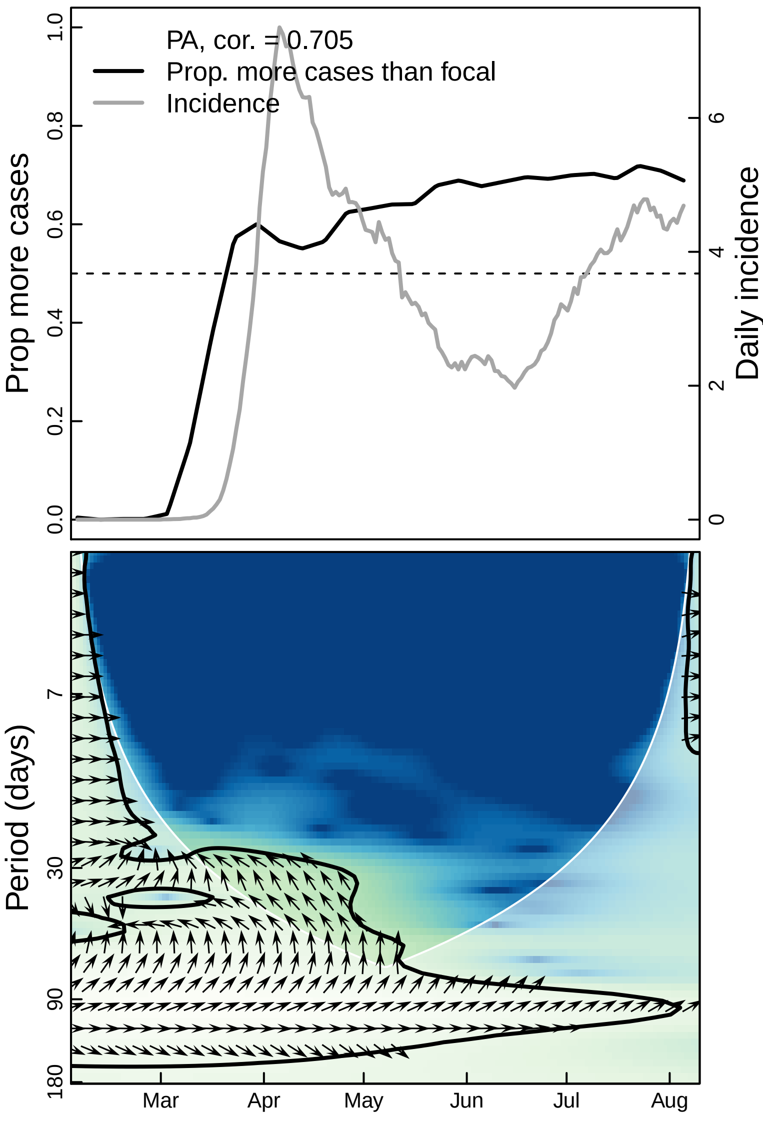

PA, cor. = 0.705
Prop. more cases than focal
Incidence
Prop more cases
Daily incidence
0.0
0.2
0.4
0.6
0.8
1.0
0
2
4
6
Period (days)
7
30
90
180
Mar
Apr
May
Jun
Jul
Aug

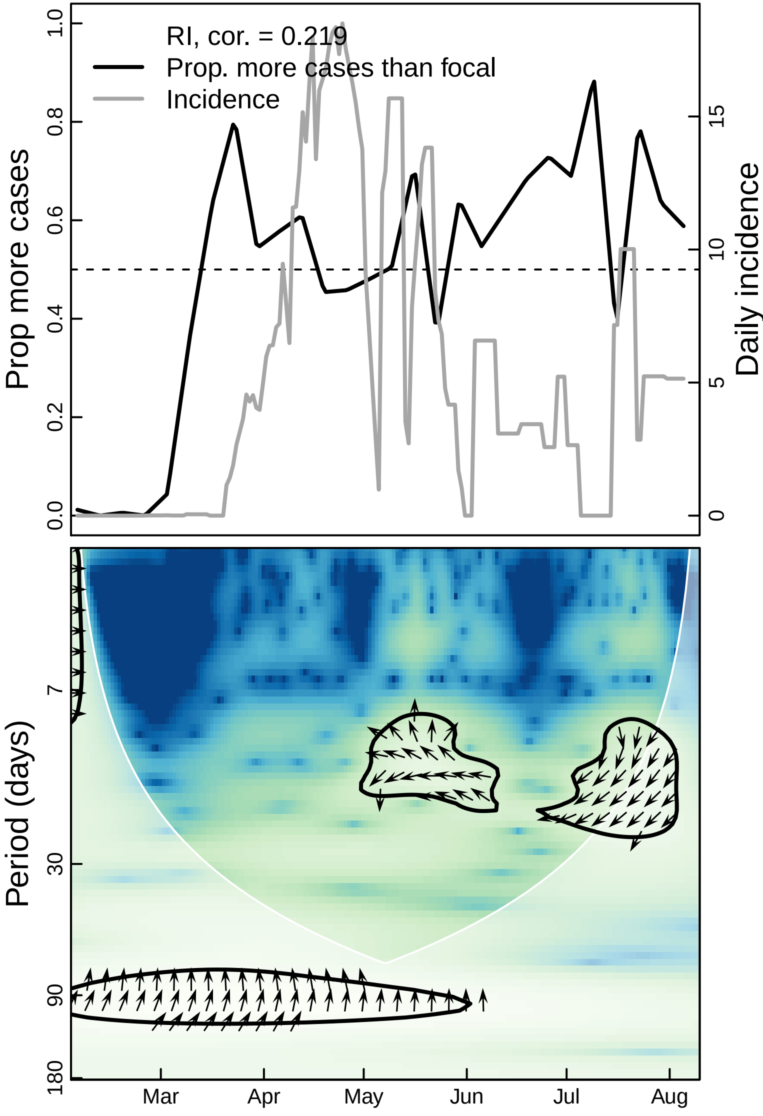

RI, cor. = 0.219
Prop. more cases than focal
Incidence
Prop more cases
Daily incidence
0.0
0.2
0.4
0.6
0.8
1.0
0
5
10
15
Period (days)
7
30
90
180
Mar
Apr
May
Jun
Jul
Aug

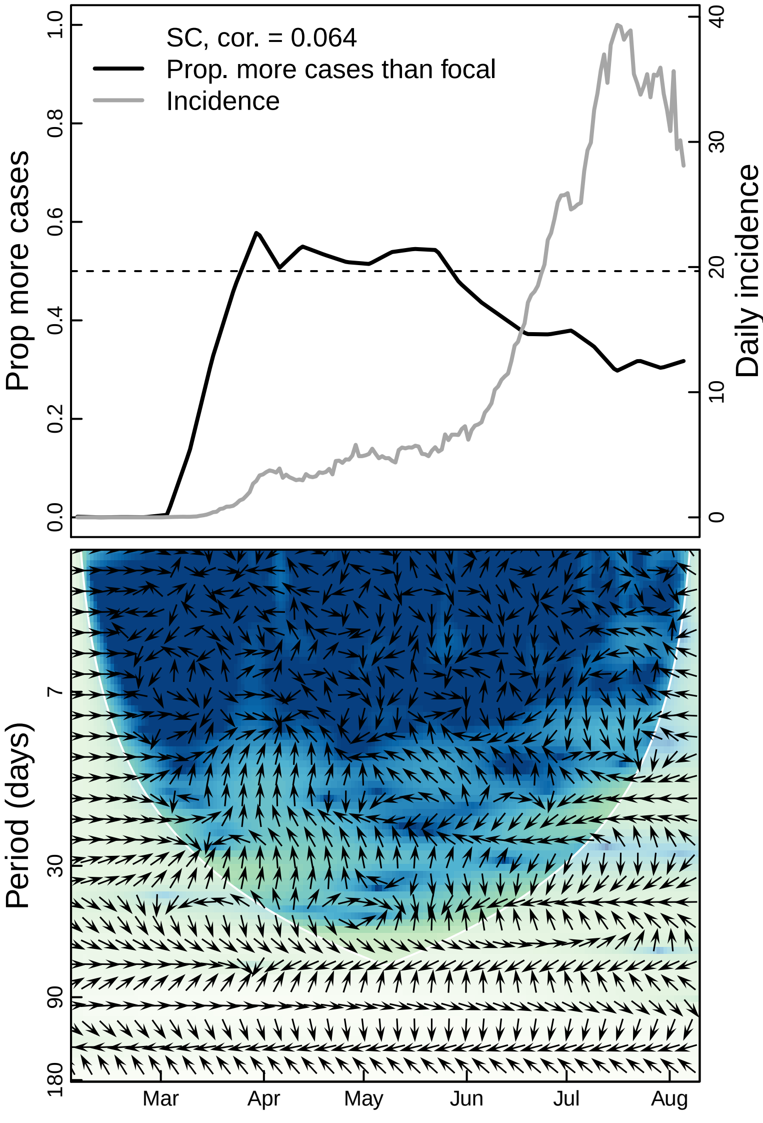

SC, cor. = 0.064
Prop. more cases than focal
Incidence
Prop more cases
Daily incidence
0.0
0.2
0.4
0.6
0.8
1.0
0
10
20
30
40
Period (days)
7
30
90
180
Mar
Apr
May
Jun
Jul
Aug

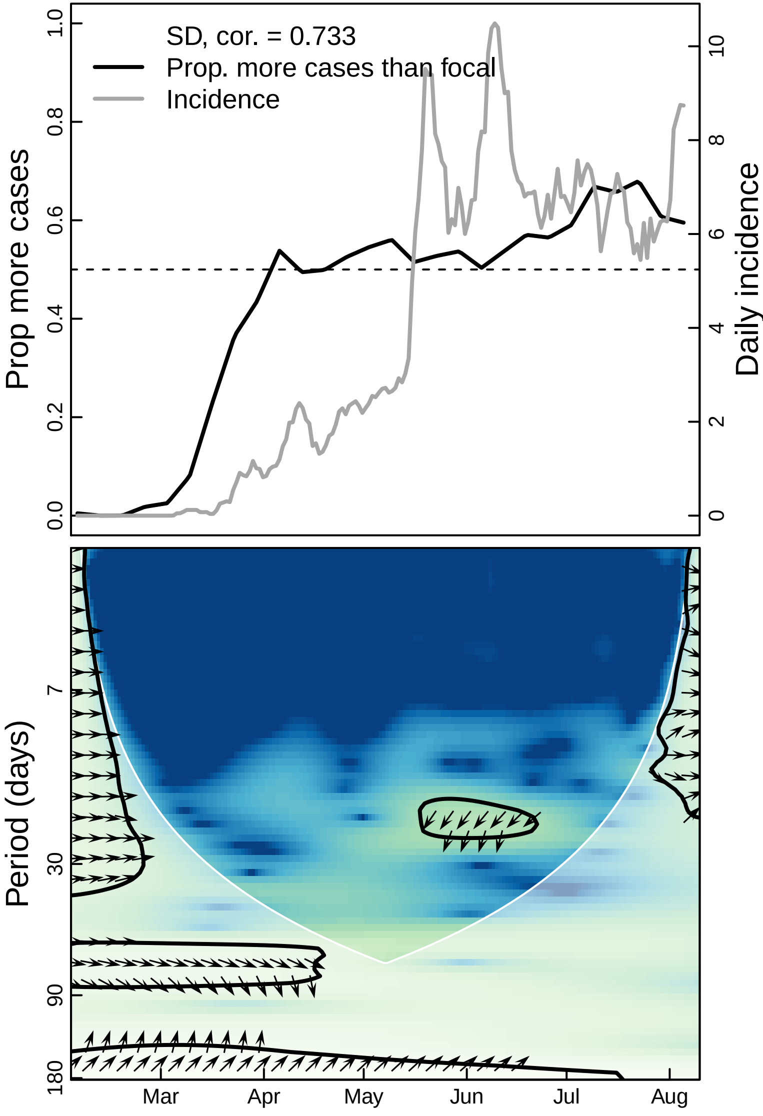

SD, cor. = 0.733
Prop. more cases than focal
Incidence
Prop more cases
Daily incidence
0.0
0.2
0.4
0.6
0.8
1.0
0
2
4
6
8
10
Period (days)
7
30
90
180
Mar
Apr
May
Jun
Jul
Aug

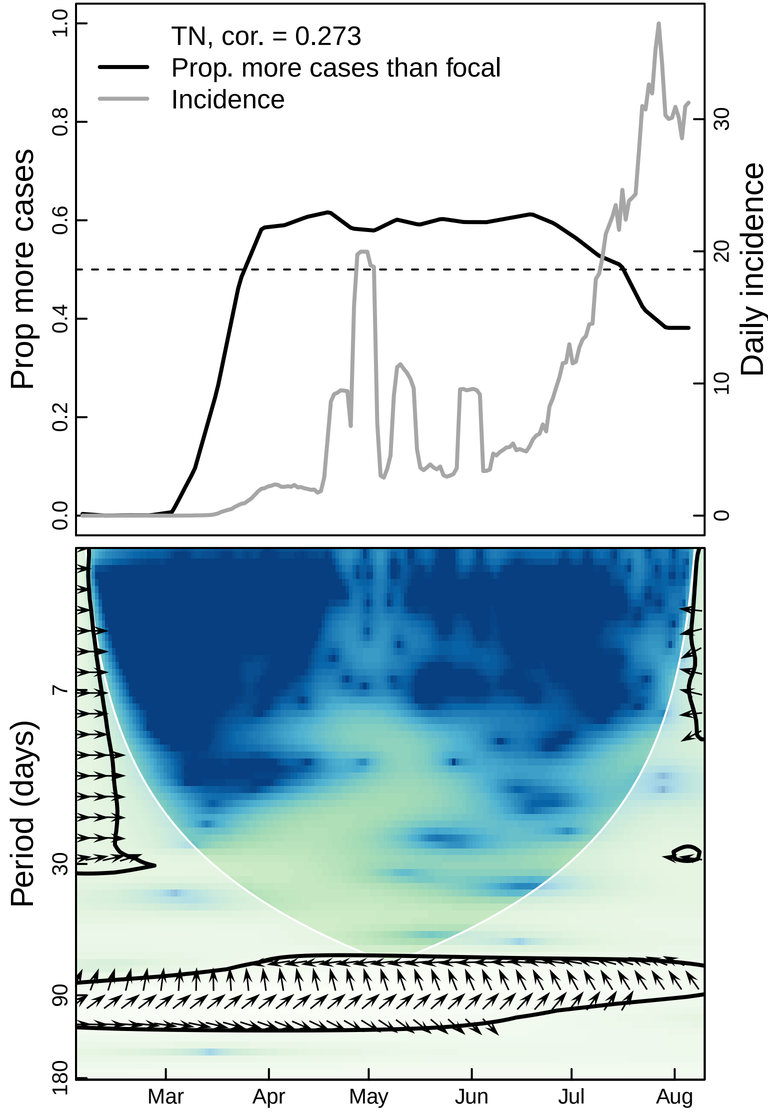
TN, cor. = 0.273
Prop. more cases than focal
Incidence
Prop more cases
Daily incidence
0.0
0.2
0.4
0.6
0.8
1.0
0
10
20
30
Period (days)
7
30
90
180
Mar
Apr
May
Jun
Jul
Aug

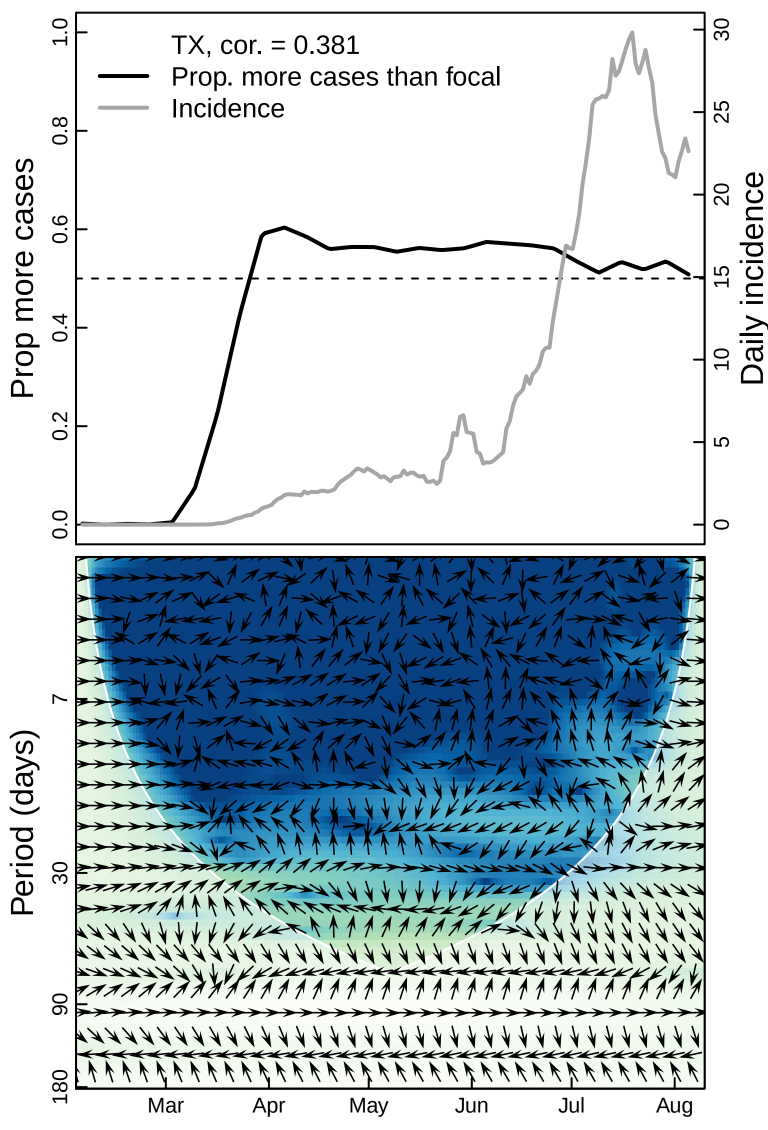

TX, cor. = 0.381
Prop. more cases than focal
Incidence
Prop more cases
0.0
0.2
0.4
0.6
0.8
1.0
Daily incidence
0
5
10
15
20
25
30
Period (days)
7
30
90
180
Mar
Apr
May
Jun
Jul
Aug

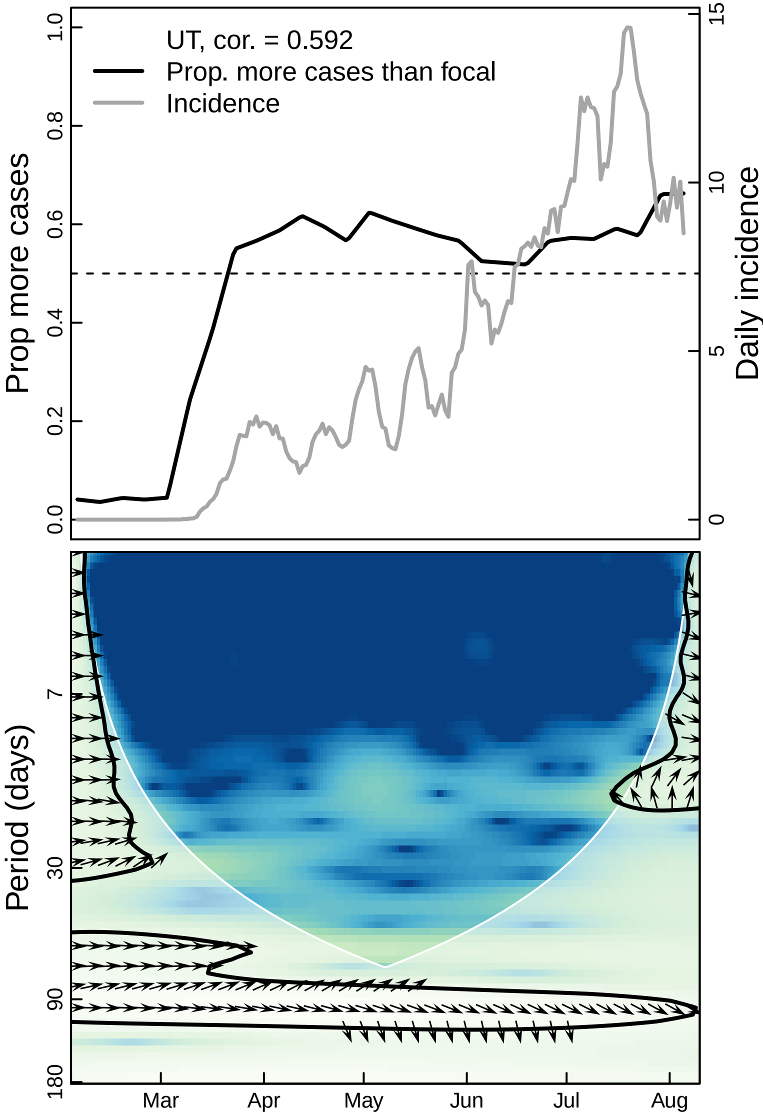
UT, cor. = 0.592
Prop. more cases than focal
Incidence
Prop more cases
Daily incidence
1.0
0.8
0.6
0.4
0.2
0.0
15
10
5
0
Period (days)
7
30
90
180
Mar
Apr
May
Jun
Jul
Aug

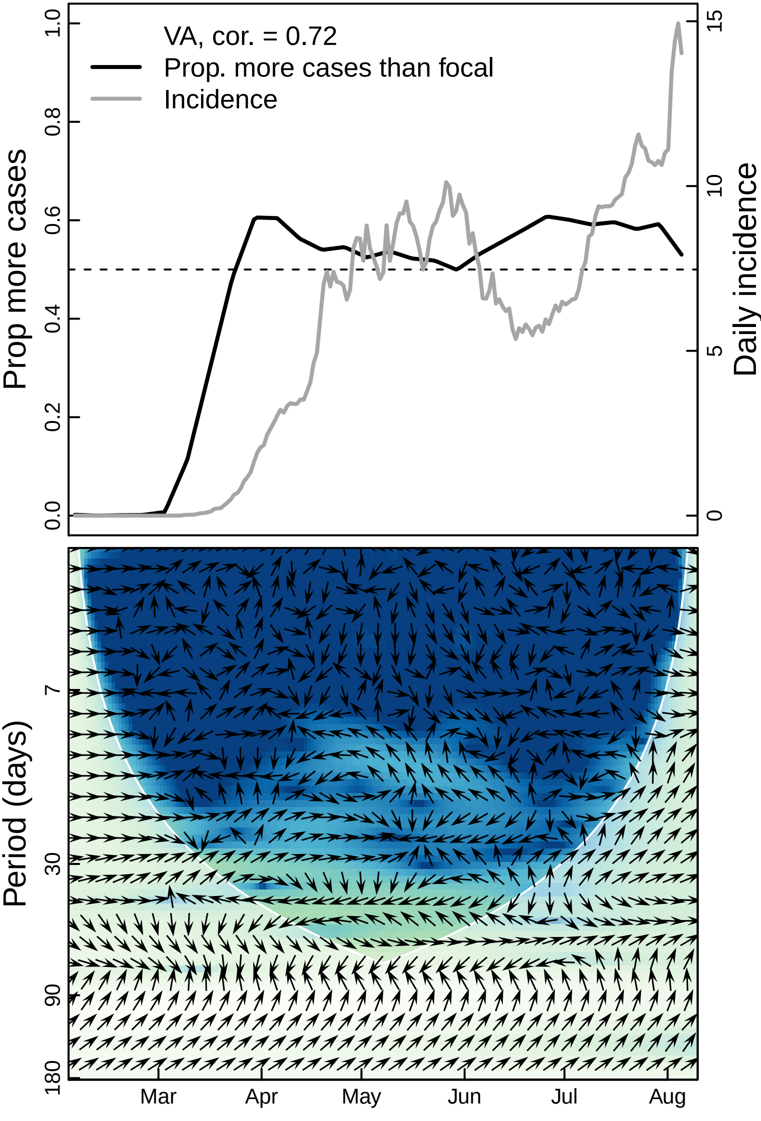

VA, cor. = 0.72
Prop. more cases than focal
Incidence
Prop more cases
Daily incidence
1.0
0.8
0.6
0.4
0.2
0.0
15
10
5
0
Period (days)
7
30
90
180
Mar
Apr
May
Jun
Jul
Aug

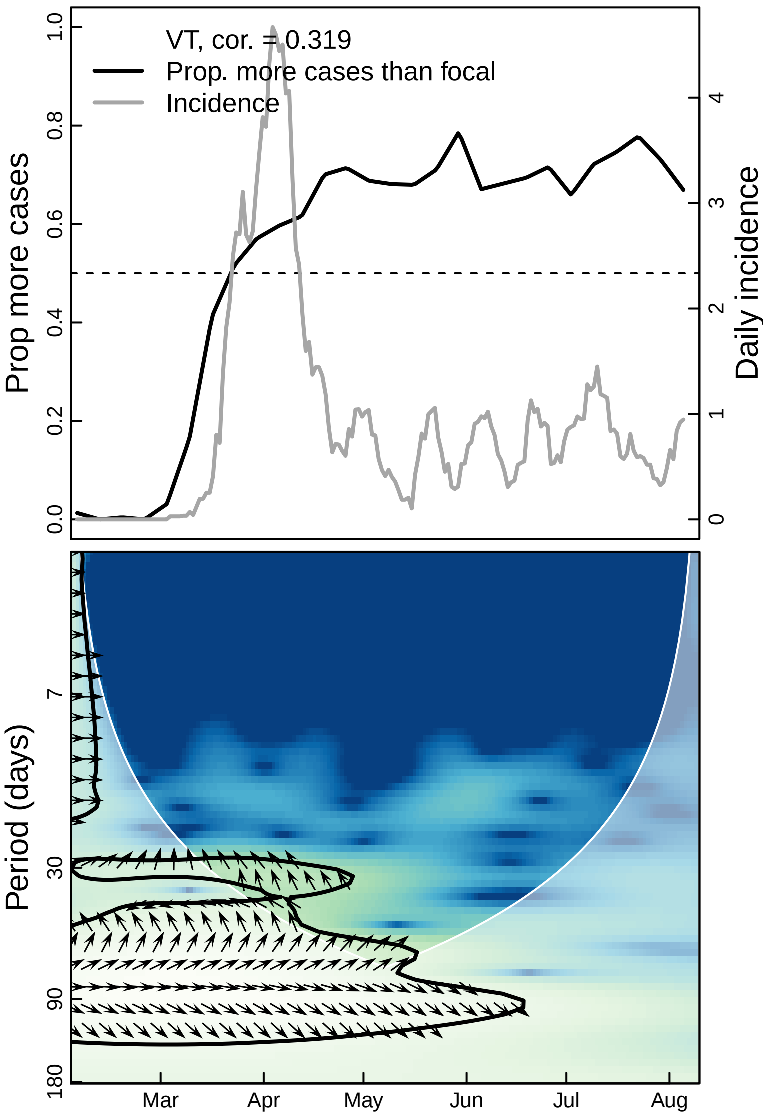

VT, cor. = 0.319
Prop. more cases than focal
Incidence
Prop more cases
Daily incidence
0.0
0.2
0.4
0.6
0.8
1.0
0
1
2
3
4
Period (days)
7
30
90
180
Mar
Apr
May
Jun
Jul
Aug

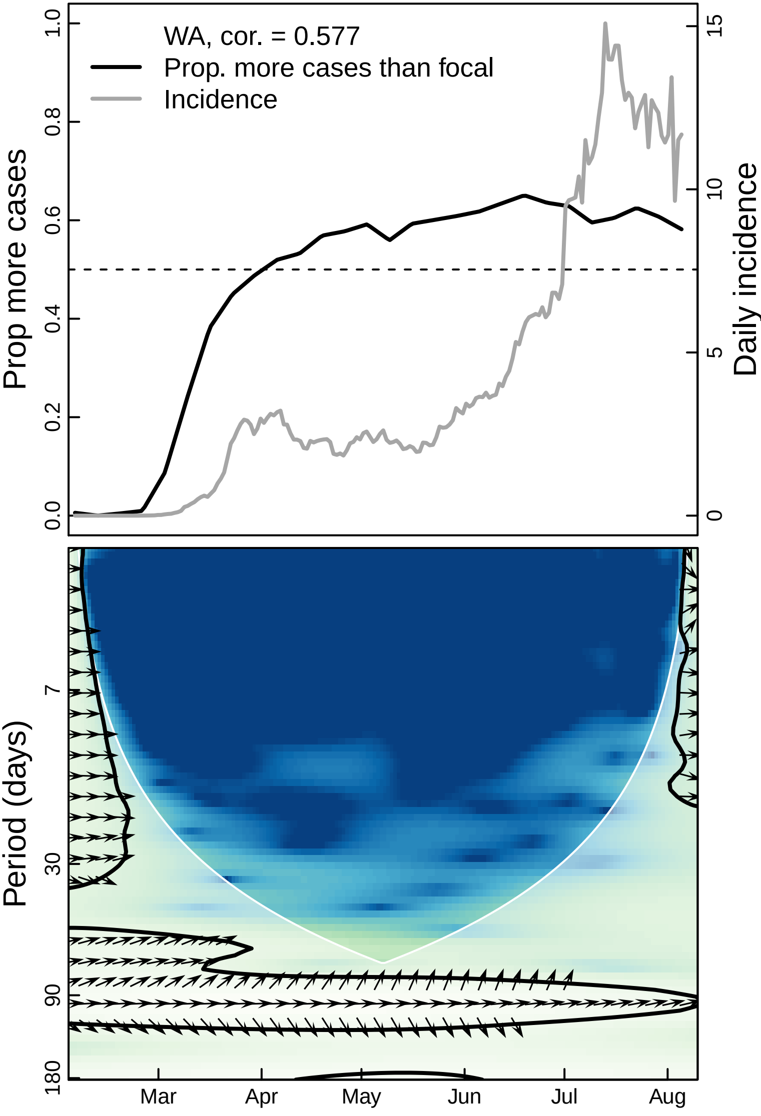

WA, cor. = 0.577
Prop. more cases than focal
Incidence
Prop more cases
Daily incidence
0.0
0.2
0.4
0.6
0.8
1.0
0
5
10
15
Period (days)
7
30
90
180
Mar
Apr
May
Jun
Jul
Aug

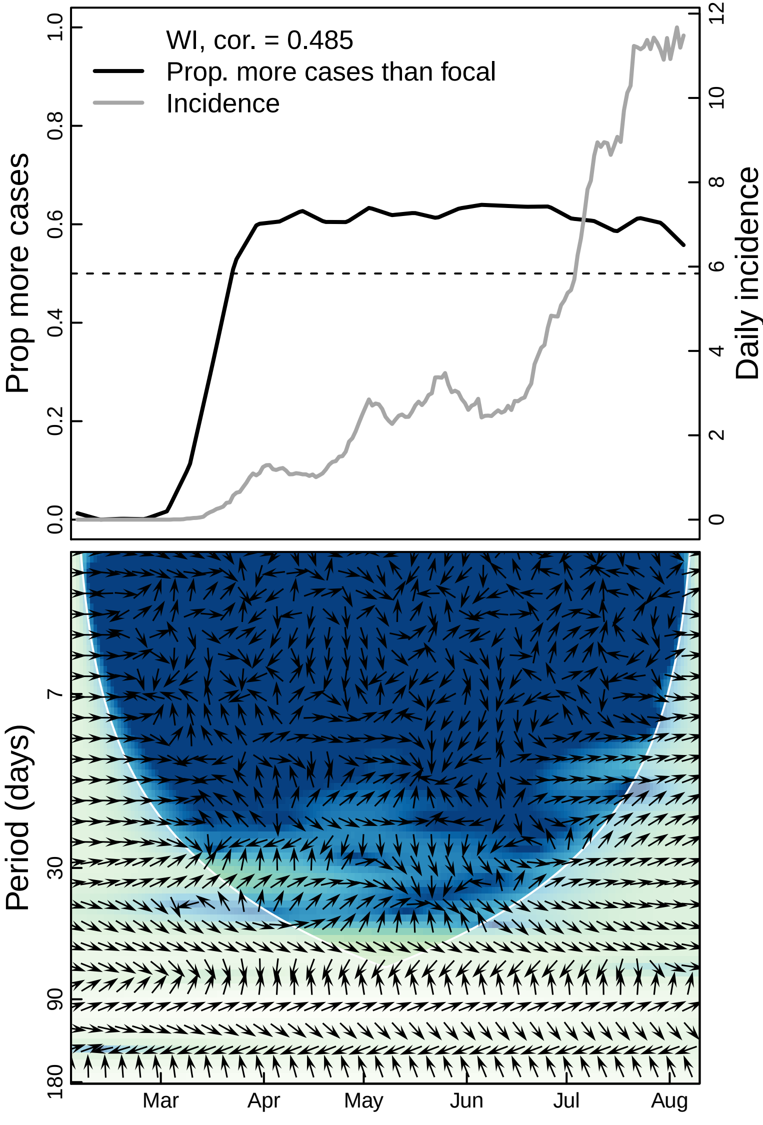

WI, cor. = 0.485
Prop. more cases than focal
Incidence
Prop more cases
Daily incidence
0.0
0.2
0.4
0.6
0.8
1.0
0
2
4
6
8
10
12
Period (days)
7
30
90
180
Mar
Apr
May
Jun
Jul
Aug

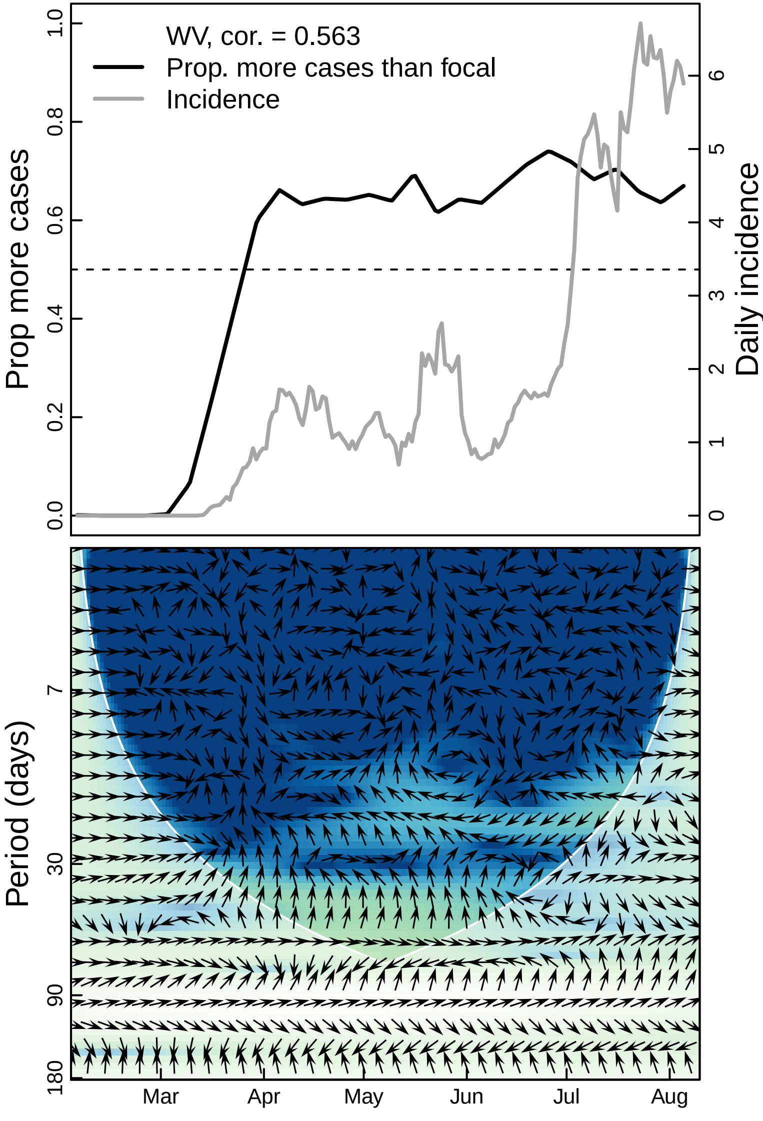
WV, cor. = 0.563
Prop. more cases than focal
Incidence
Prop more cases
Daily incidence
0.0
0.2
0.4
0.6
0.8
1.0
0
1
2
3
4
5
6
Period (days)
7
30
90
180
Mar
Apr
May
Jun
Jul
Aug

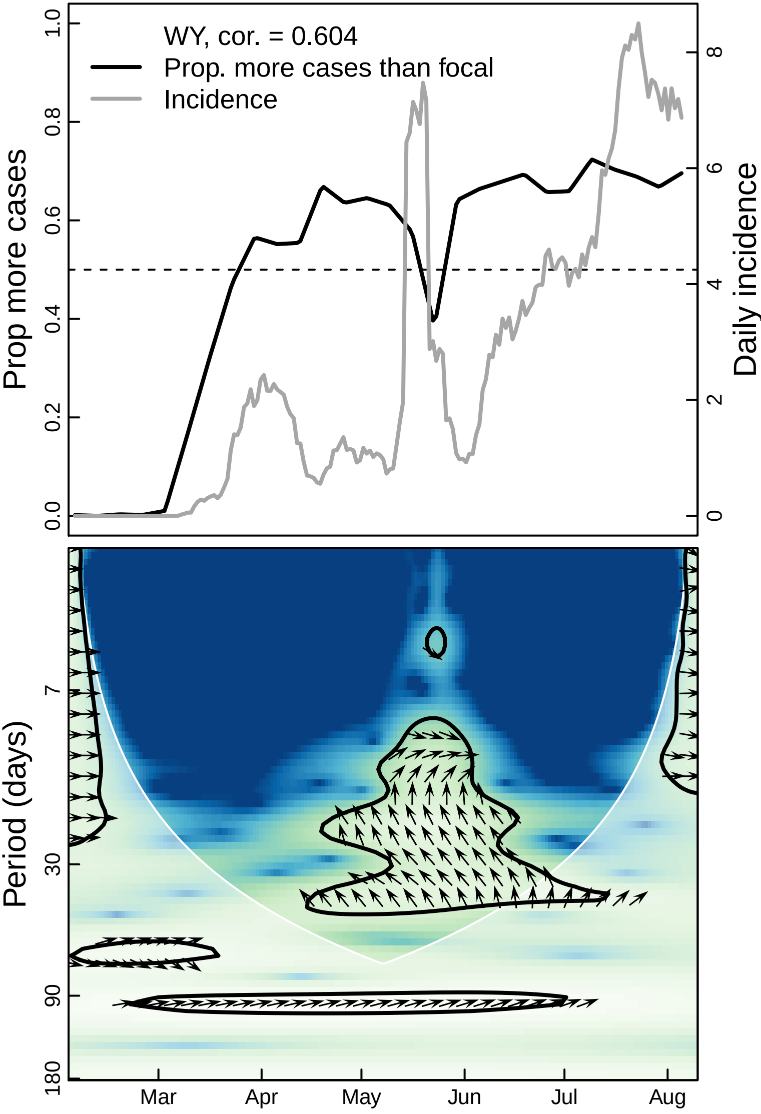

WY, cor. = 0.604
Prop. more cases than focal
Incidence
Prop more cases
Daily incidence
0.0
0.2
0.4
0.6
0.8
1.0
0
2
4
6
8
Period (days)
7
30
90
180
Mar
Apr
May
Jun
Jul
Aug
